# Inpatient Mortality and Risk Factors Among Hospitalized Medicare Beneficiaries Ages 65 and Older with COVID-19 Treated with COVID-19 Convalescent Plasma (CCP), During 2020-2021

**DOI:** 10.64898/2026.10.22.26365943

**Authors:** Mikhail Menis, Nathan Duma, Sarah Barney, Yoganand Chillarige, Whitney R. Steele, Stephen H. Chang, Carlos H. Villa, Anne F. Eder, Barbee I. Whitaker

## Abstract

**Background:** COVID-19 has had a substantial burden on older adults aged 65+. This study’s objective was to evaluate inpatient mortality among hospitalized Fee-for-Service (FFS) Medicare beneficiaries ages 65+ with COVID-19 treated with COVID-19 convalescent plasma (CCP) during August 2020-December 2021: overall, monthly, and by patient and hospital characteristics.

**Study Design and Methods:** Our retrospective claims-based study utilized large Centers for Medicare & Medicaid Services administrative databases to assess inpatient COVID-19 stays with recorded CCP transfusions among FFS Medicare beneficiaries ages 65+. CCP transfusions were identified by recorded procedure codes, and inpatient mortality via the patient status discharge code. Crude inpatient mortality rates (per 100 CCP transfusion stays with COVID-19) and corresponding 95% confidence intervals were evaluated across a range of patient and hospital characteristics. Multivariate logistic regression analyses were used to assess potential risk factors.

**Results:** The overall crude inpatient mortality was 25.5% (95%CI:25.2%-25.8%), declining from 30.7% (95%CI:29.1%-32.3%) in August 2020 to 7.4% (95%CI:3.8%-15.0%) in December 2021, p<0.001. The study identified significantly higher adjusted odds of inpatient mortality for each year of advancing age (1.06; 95%CI:1.06-1.07), immunocompromised (IC) vs. non-IC (1.14; 95%CI:1.05-1.24), males vs. females (1.20; 95%CI:1.15-1.25), Medicaid-eligible vs. not (1.13; 95%CI:1.07-1.20), medium (1.14; 95%CI:1.06-1.22) and large (1.09; 95%CI:1.01-1.19) compared to small hospitals, with ICU/CCU admission vs. not (2.71; 95%CI:2.57-2.85), mechanical ventilation<96 hours (13.11; 95%CI:12.11-14.20) and ≥96 hours (11.59; 95%CI:10.89-12.34) vs. not, among others.

**Discussion:** The findings highlight importance of patient and hospital characteristics, including demographics, comorbidities, socioeconomic status, and COVID-19 severity, for inpatient mortality among Medicare beneficiaries receiving CCP.

## INTRODUCTION

COVID-19 has had a substantial burden on older adults aged 65+ and immunocompromised (IC) persons during the public health emergency, with most of the COVID-19 deaths in the United States occurring in the hospital or other inpatient healthcare settings (>65%) and among persons ages 65+ (>75%) [1–4]. The literature [1, 5–7] identified various risk factors for severe COVID-19 outcomes, including but not limited to advancing age, IC status, and underlying comorbidities. The investigation by Senefeld et al. [8] evaluated crude 30-day mortality by patient and hospital characteristics among hospitalized subjects ages 18+ enrolled in the Expanded Access Program (EAP) for COVID-19 convalescent plasma (CCP) during April-August 2020. It suggested multiple factors that could be associated with higher risk of mortality including demographics, comorbidities, COVID-19 severity, residence, concurrent therapies, and hospital characteristics (e.g. type, location). Hospitalized patients during the early pandemic received a number of investigational treatments, including CCP.

CCP became available for COVID-19 treatment in March 2020, first under individual patient emergency Investigational New Drug applications (eINDs), then under the national EAP protocol. [8,9] In August 2020, CCP received an Emergency Use Authorization (EUA) by U.S. FDA for the treatment of hospitalized patients with COVID-19. [10] To our knowledge, there are no published population-based observational studies beyond the EAP period that evaluated inpatient mortality and risk factors among inpatient U.S. Medicare Fee-for-Service (FFS) beneficiaries aged 65+ with COVID-19 treated with CCP, who are likely at high risk for severe COVID-19 outcomes [1,5,8,11,12]. Our study, therefore, characterized CCP recipients; evaluated overall, annual, and monthly crude inpatient mortality rates; and ascertained adjusted hospital (e.g., type and size) and patient risk factors (e.g., demographics, underlying health conditions, socioeconomic factors, IC status, residence, COVID-19 severity) for inpatient mortality among CCP-treated inpatient FFS Medicare beneficiaries aged 65+ with COVID-19 during August 2020-December 2021.

## MATERIALS AND METHODS

### Study Design

This retrospective single-arm cohort claims-based study utilized large Centers for Medicare & Medicaid Services (CMS) administrative databases. The study retrospectively evaluated inpatient COVID-19 stays with recorded CCP transfusions among FFS Medicare beneficiaries ages 65+, who were continuously enrolled in Part A (hospital insurance) and Part B (medical insurance) within 3 months prior to the index date (i.e., admission date), during August 1, 2020, through December 31, 2021. Stays with both CCP and other blood component transfusions were excluded to avoid confounding due to a much smaller sample size (about 6% of all CCP users) and presence of very different risk factors for inpatient mortality (e.g., indications and quantity of other blood components). ICD-10-CM diagnosis code U07.1 identified inpatient stays with COVID-19; and ICD-10-PCS procedure codes XW13325 and XW14325 were used to identify CCP transfusions. Inpatient (IP) mortality during hospitalization was assessed via the patient status discharge code in the inpatient Medicare claims data. The IC status was evaluated based on at least two recorded diagnosis or procedure codes for malignancies, chemotherapy, radiation, or transplantation within 3 months prior to or on the index date. (See Supplementary Appendix Table).

### Analytic plan

The study characterized hospitalized beneficiaries with COVID-19 receiving CCP and assessed corresponding crude inpatient mortality rates per 100 inpatient stays: overall, annually, and by hospital admission months, hospital ownership and size, patient demographics, IC status, nursing home residence status, U.S. Census Regions [13], urban vs. rural residence, Area Deprivation Index (ADI)[14] (with higher ADI scores indicating higher levels of neighborhood disadvantage), emergency room (ER) admission, Medicaid eligibility, COVID-19 as a reason for admission, time since most recent COVID-19 diagnosis recorded within 30 days prior to admission, Charlson Comorbidity Index (CCI) scores [15,16], and by histories of underlying health conditions (e.g., Elixhauser comorbidities[16,17]), thrombotic events [TE], and end-stage-renal disease [ESRD]. We used ICD-10-CM diagnosis codes, ICD-10-PCS procedure codes, and revenue codes recorded on claims to identify evaluated characteristics. Inpatient mortality was also assessed by the following COVID-19 severity measures: TE occurrence during stay, mechanical ventilation [MV], intensive care unit or coronary care unit [ICU/CCU] admission, length of stay [LOS], and the use of immunoglobulins (IVIGs and hyperimmunoglobulins), tocilizumab, baricitinib and remdesivir. Subgroup analyses were performed by calendar year, IC status, demographics, time since most recent COVID-19, Medicaid eligibility status, and by four U.S. Census regions [13]. The overall and subgroup multivariate logistic regression analyses were used to assess the adjusted effects of potential hospital and patient risk factors on inpatient mortality among beneficiaries receiving CCP.

### Statistical Analysis

Crude mortality rates (based on observed number of inpatient deaths divided by the number of CCP transfusion stays with COVID-19) and corresponding 95% confidence intervals (CIs) were evaluated across a range of patient and hospital characteristics. 95% CIs were estimated using binomial proportions via the Wilson method. The Cochran-Armitage trend test [18,19] was used to assess monthly crude inpatient mortality trends, overall and by IC status, and respectively by regions, during the study period.

The study conducted multivariate logistic regression analyses to estimate adjusted odds ratios (ORs) and 95% CIs for inpatient mortality comparisons by characteristics evaluated. All the non-history characteristics ascertained during inpatient stay were *a priori* retained in the final regression models along with statistically significant 90-day history covariates as identified in the univariate logistic regression modeling. Forward stepwise selection was then used to include remaining history variables at p<0.05, overall and for each subgroup analysis separately. Multivariable regression splines (MVRS) were created to assess adjusted inpatient mortality odds by admission months, age, ADI, CCI, and time since most recent COVID-19 diagnosis, which were all treated as continuous variables. Pearson’s Chi-Square Goodness-of-Fit, Deviance Goodness-of-Fit, and Area Under the ROC Curve (AUC) tests were performed to evaluate model fit and predictive ability of the final regression models. Additionally, the study ascertained variance inflation factors (VIFs) to investigate potential multicollinearity (i.e., correlations between the covariates) in the models. All analyses were performed in SAS 9.4 (SAS Institute Inc., Cary, NC, USA). The study used p<0.05 as *a priori* level of significance.

## RESULTS

### CCP Recipient Characteristics and Crude Inpatient Mortality

Table 1 shows patient and hospital characteristics for CCP transfusion hospital stays with COVID-19 and corresponding crude inpatient mortality rates and adjusted ORs for comparisons in the overall multivariate regression analysis. Of 81,751 COVID-19 CCP transfusion stays, 20,876 (25.5%; 95%CI:25.2%-25.8%) recorded inpatient mortality. The majority of CCP recipients were: community-dwelling (91.1%), non-IC (90.0%), males (54.3%), non-Medicaid-eligible (79.4%), admitted in 2020 (64.9%) and through emergency room (89.2%); and with: COVID-19 as a reason for admission (52.2%), recorded use of remdesivir (75.6%), ICU/CCU admission (59.2%), LOS≥7 days (67.9%), residence in the urban areas (68.1%) and in the South (54.0%). CCP recipients were also mostly in non-profit (66.8%) and medium-size hospitals (64.2%). The crude inpatient mortality rates (%) varied by patient and hospital characteristics with among the highest mortality rates per 100 stays identified for those with ICU/CCU admission (36.6; 95%CI:36.2-37.1), thrombosis during hospitalization (41.8; 95%CI: 40.9-42.7), CCI score≥5 (33.6; 95%CI:32.8-34.5), MV<96 hours (74.5; 95%CI:73.2-75.7), and ≥96 hours (73.6; 95%CI:72.7-74.5). Among the lowest mortality rates were identified for non-ICU/CCU stays (9.4; 95%CI:9.1-9.7), LOS 5-6 days (12.5; 95%CI:12.0-13.0) and non-MV stays (15.3; 95%CI:15.0-15.6).

**Table 1.**
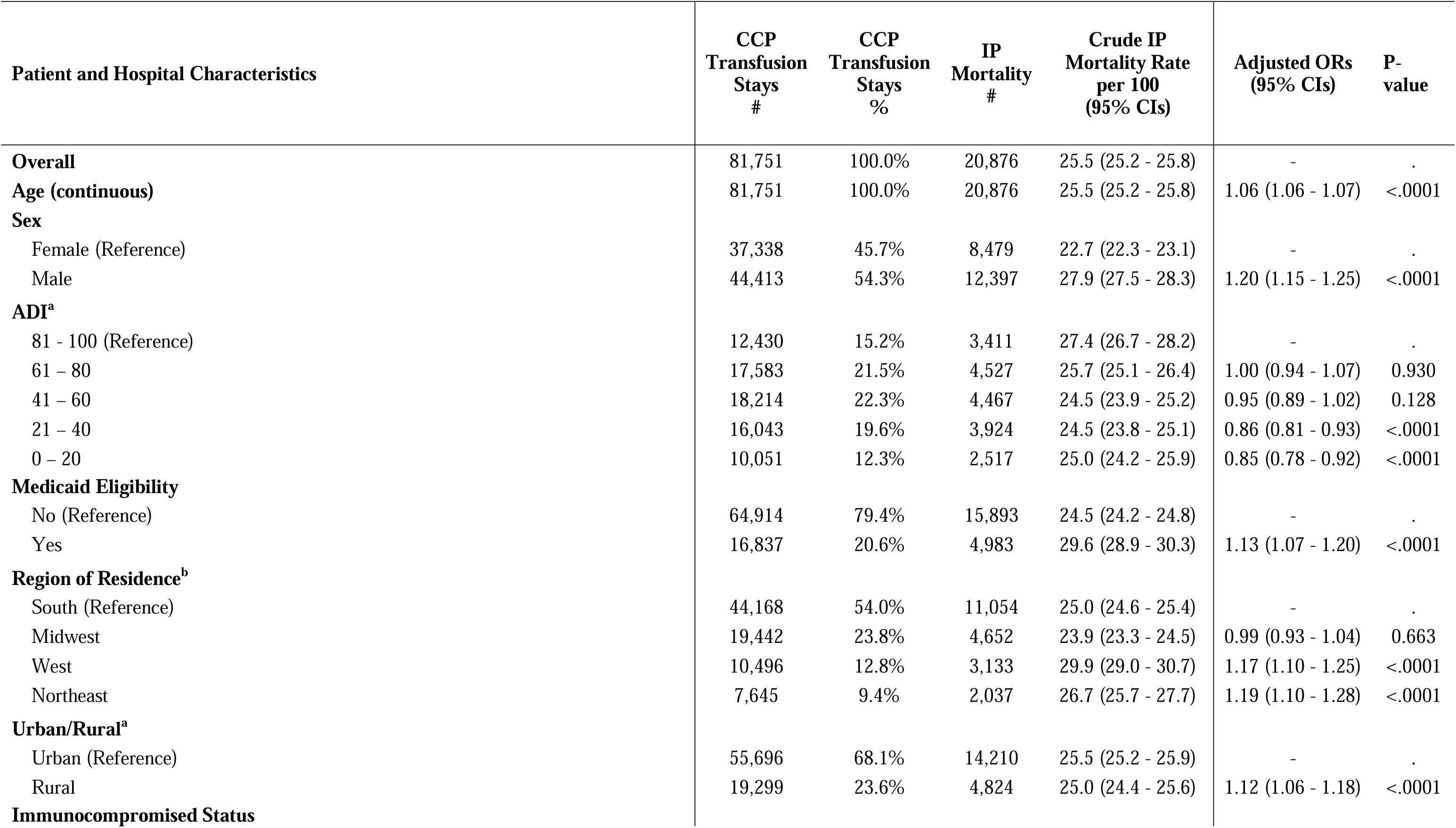
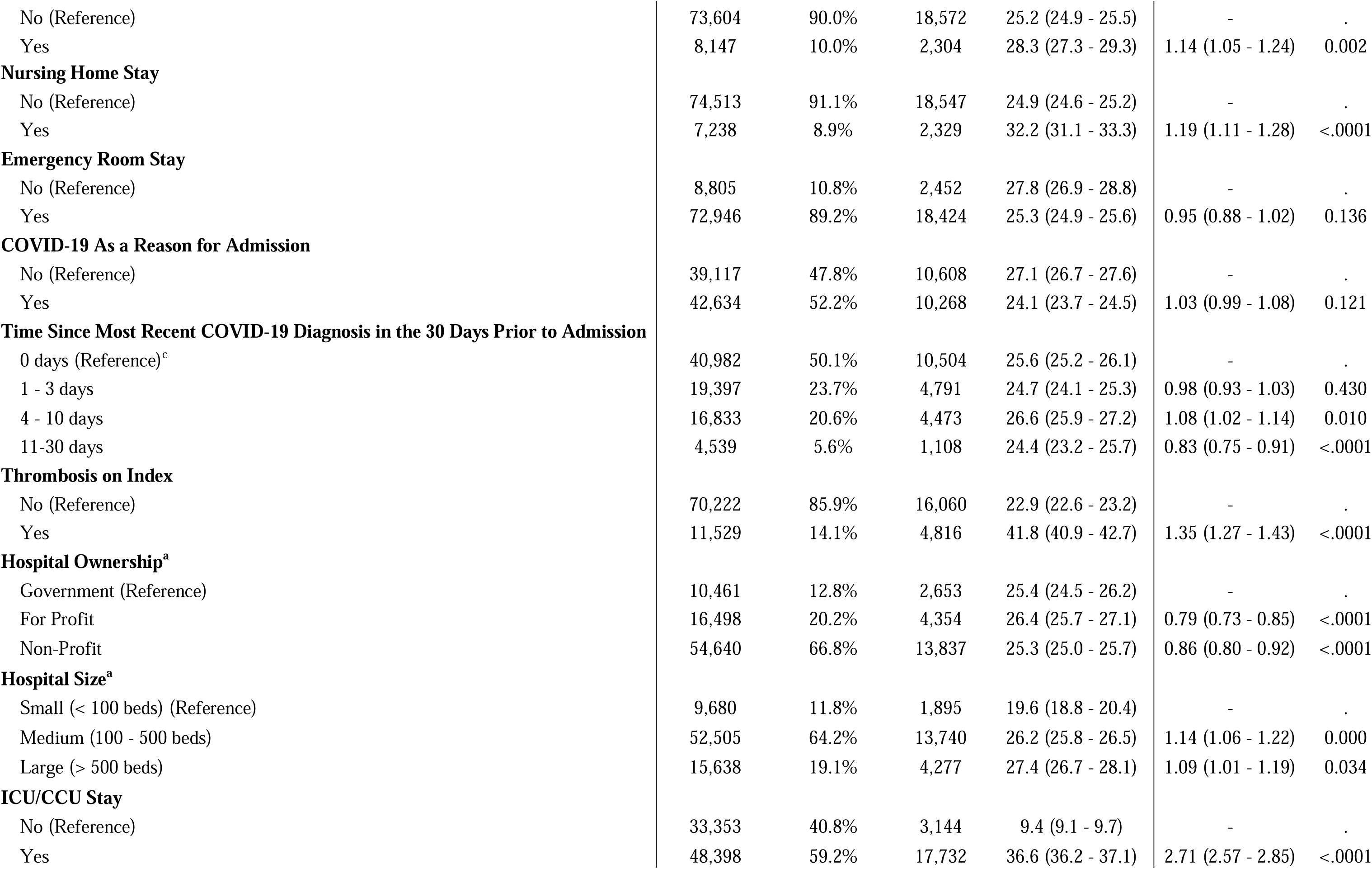
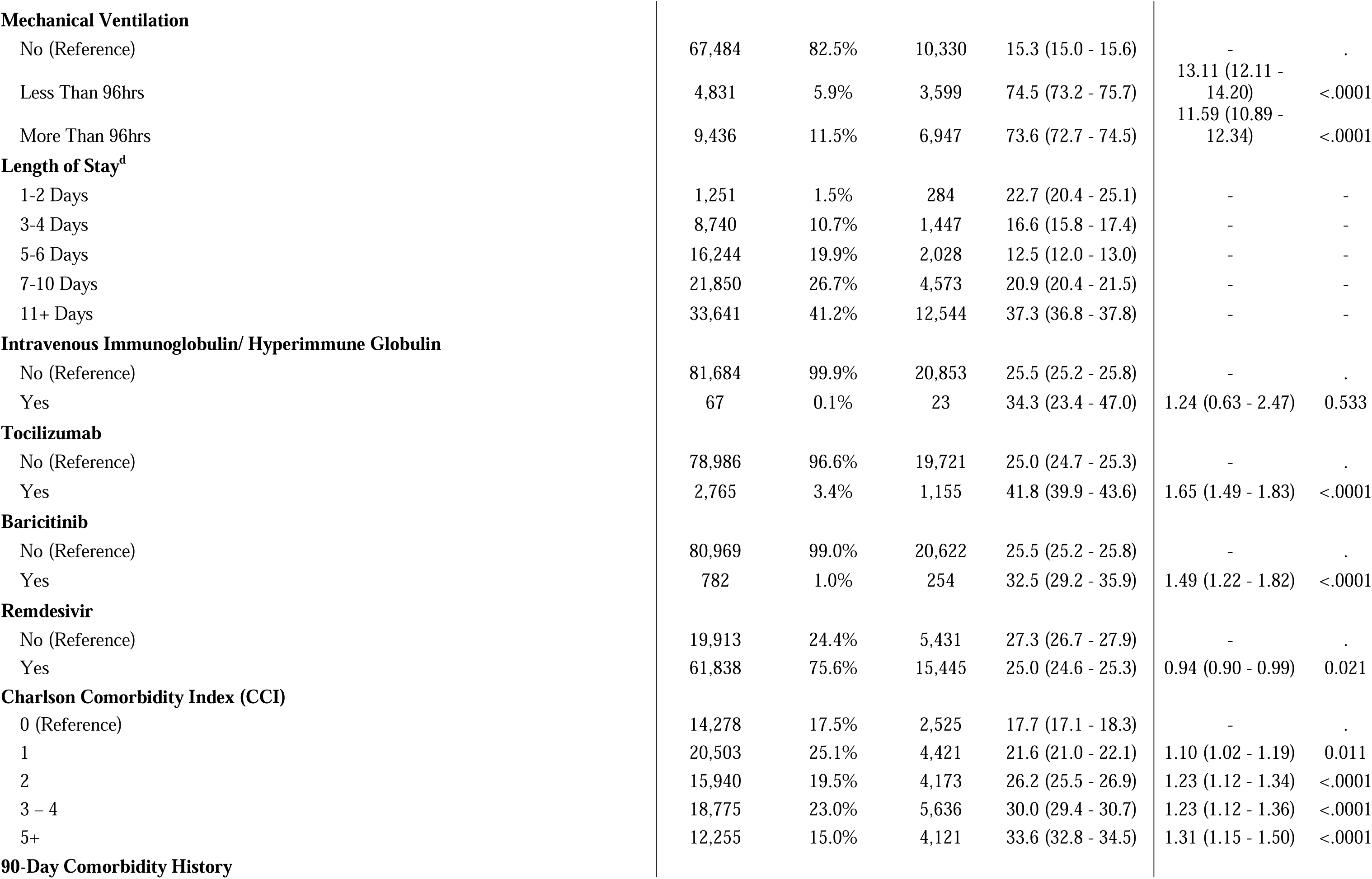
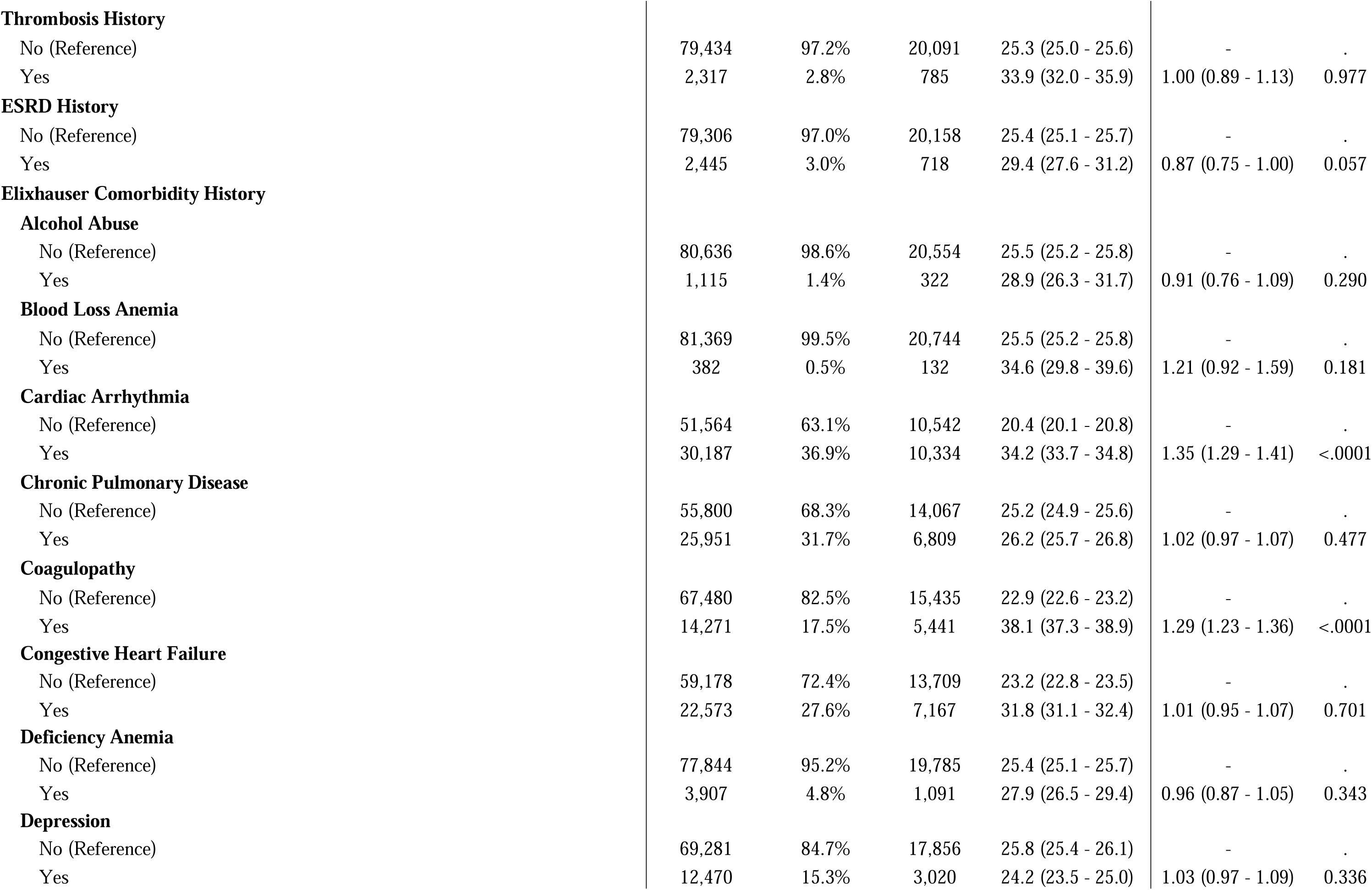
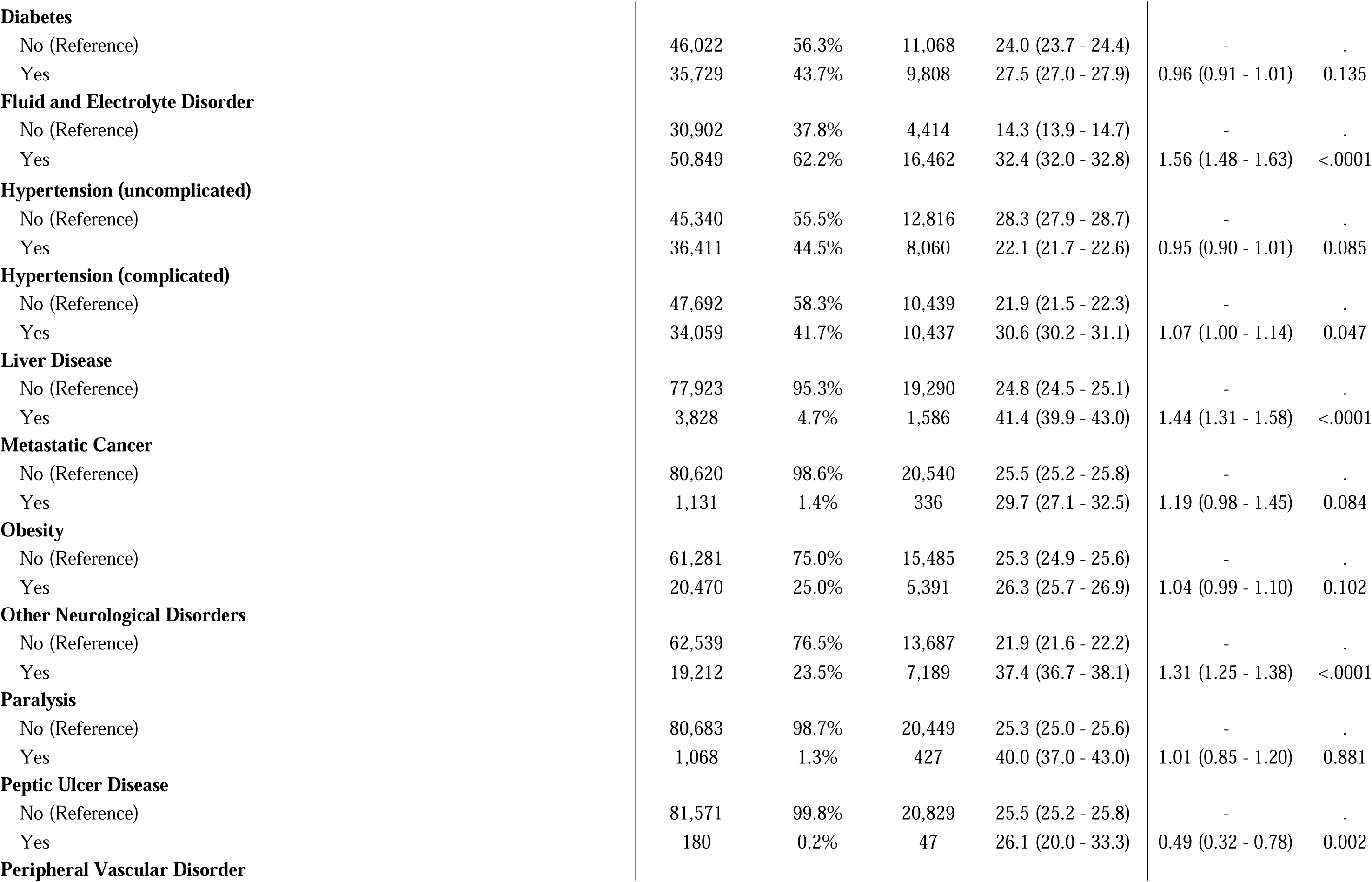
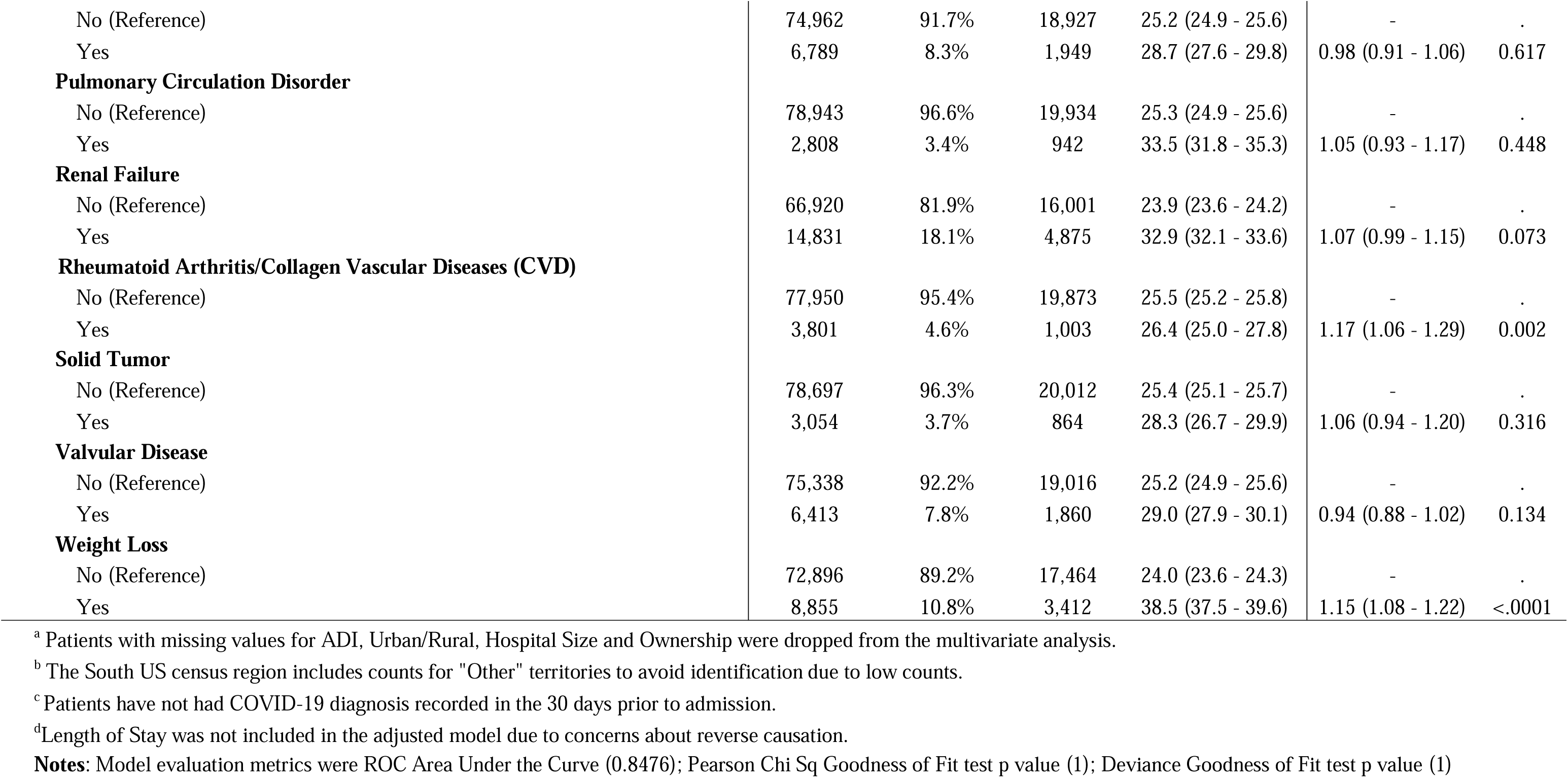
Inpatient (IP) Mortality Rates and Corresponding Adjusted Odds Ratios (ORs) by Patient and Hospital characteristics Among Recipients of COVID-19 Convalescent Plasma (CCP)

| Patient and Hospital Characteristics | CCP<br>Transfusion<br>Stays<br># | CCP<br>Transfusion<br>Stays<br>% | IP<br>Mortality<br># | Crude IP<br>Mortality Rate<br>per 100<br>(95% CIs) | Adjusted ORs<br>(95% CIs) | P-value |
| --- | --- | --- | --- | --- | --- | --- |
| **Overall** | 81,751 | 100.0% | 20,876 | 25.5 (25.2 - 25.8) | - | . |
| **Age (continuous)** | 81,751 | 100.0% | 20,876 | 25.5 (25.2 - 25.8) | 1.06 (1.06 - 1.07) | <.0001 |
| **Sex** |  |  |  |  |  |  |
| Female (Reference) | 37,338 | 45.7% | 8,479 | 22.7 (22.3 - 23.1) | - | . |
| Male | 44,413 | 54.3% | 12,397 | 27.9 (27.5 - 28.3) | 1.20 (1.15 - 1.25) | <.0001 |
| **ADI<sup>a</sup>** |  |  |  |  |  |  |
| 81 - 100 (Reference) | 12,430 | 15.2% | 3,411 | 27.4 (26.7 - 28.2) | - | . |
| 61 – 80 | 17,583 | 21.5% | 4,527 | 25.7 (25.1 - 26.4) | 1.00 (0.94 - 1.07) | 0.930 |
| 41 – 60 | 18,214 | 22.3% | 4,467 | 24.5 (23.9 - 25.2) | 0.95 (0.89 - 1.02) | 0.128 |
| 21 – 40 | 16,043 | 19.6% | 3,924 | 24.5 (23.8 - 25.1) | 0.86 (0.81 - 0.93) | <.0001 |
| 0 – 20 | 10,051 | 12.3% | 2,517 | 25.0 (24.2 - 25.9) | 0.85 (0.78 - 0.92) | <.0001 |
| **Medicaid Eligibility** |  |  |  |  |  |  |
| No (Reference) | 64,914 | 79.4% | 15,893 | 24.5 (24.2 - 24.8) | - | . |
| Yes | 16,837 | 20.6% | 4,983 | 29.6 (28.9 - 30.3) | 1.13 (1.07 - 1.20) | <.0001 |
| **Region of Residence<sup>b</sup>** |  |  |  |  |  |  |
| South (Reference) | 44,168 | 54.0% | 11,054 | 25.0 (24.6 - 25.4) | - | . |
| Midwest | 19,442 | 23.8% | 4,652 | 23.9 (23.3 - 24.5) | 0.99 (0.93 - 1.04) | 0.663 |
| West | 10,496 | 12.8% | 3,133 | 29.9 (29.0 - 30.7) | 1.17 (1.10 - 1.25) | <.0001 |
| Northeast | 7,645 | 9.4% | 2,037 | 26.7 (25.7 - 27.7) | 1.19 (1.10 - 1.28) | <.0001 |
| **Urban/Rural<sup>a</sup>** |  |  |  |  |  |  |
| Urban (Reference) | 55,696 | 68.1% | 14,210 | 25.5 (25.2 - 25.9) | - | . |
| Rural | 19,299 | 23.6% | 4,824 | 25.0 (24.4 - 25.6) | 1.12 (1.06 - 1.18) | <.0001 |
| **Immunocompromised Status** |  |  |  |  |  |  |
| No (Reference) | 73,604 | 90.0% | 18,572 | 25.2 (24.9 - 25.5) | - | . |
| Yes | 8,147 | 10.0% | 2,304 | 28.3 (27.3 - 29.3) | 1.14 (1.05 - 1.24) | 0.002 |
| **Nursing Home Stay** |  |  |  |  |  |  |
| No (Reference) | 74,513 | 91.1% | 18,547 | 24.9 (24.6 - 25.2) | - | . |
| Yes | 7,238 | 8.9% | 2,329 | 32.2 (31.1 - 33.3) | 1.19 (1.11 - 1.28) | <.0001 |
| **Emergency Room Stay** |  |  |  |  |  |  |
| No (Reference) | 8,805 | 10.8% | 2,452 | 27.8 (26.9 - 28.8) | - | . |
| Yes | 72,946 | 89.2% | 18,424 | 25.3 (24.9 - 25.6) | 0.95 (0.88 - 1.02) | 0.136 |
| **COVID-19 As a Reason for Admission** |  |  |  |  |  |  |
| No (Reference) | 39,117 | 47.8% | 10,608 | 27.1 (26.7 - 27.6) | - | . |
| Yes | 42,634 | 52.2% | 10,268 | 24.1 (23.7 - 24.5) | 1.03 (0.99 - 1.08) | 0.121 |
| **Time Since Most Recent COVID-19 Diagnosis in the 30 Days Prior to Admission** |  |  |  |  |  |  |
| 0 days (Reference)<sup>c</sup> | 40,982 | 50.1% | 10,504 | 25.6 (25.2 - 26.1) | - | . |
| 1 - 3 days | 19,397 | 23.7% | 4,791 | 24.7 (24.1 - 25.3) | 0.98 (0.93 - 1.03) | 0.430 |
| 4 - 10 days | 16,833 | 20.6% | 4,473 | 26.6 (25.9 - 27.2) | 1.08 (1.02 - 1.14) | 0.010 |
| 11-30 days | 4,539 | 5.6% | 1,108 | 24.4 (23.2 - 25.7) | 0.83 (0.75 - 0.91) | <.0001 |
| **Thrombosis on Index** |  |  |  |  |  |  |
| No (Reference) | 70,222 | 85.9% | 16,060 | 22.9 (22.6 - 23.2) | - | . |
| Yes | 11,529 | 14.1% | 4,816 | 41.8 (40.9 - 42.7) | 1.35 (1.27 - 1.43) | <.0001 |
| **Hospital Ownership<sup>a</sup>** |  |  |  |  |  |  |
| Government (Reference) | 10,461 | 12.8% | 2,653 | 25.4 (24.5 - 26.2) | - | . |
| For Profit | 16,498 | 20.2% | 4,354 | 26.4 (25.7 - 27.1) | 0.79 (0.73 - 0.85) | <.0001 |
| Non-Profit | 54,640 | 66.8% | 13,837 | 25.3 (25.0 - 25.7) | 0.86 (0.80 - 0.92) | <.0001 |
| **Hospital Size<sup>a</sup>** |  |  |  |  |  |  |
| Small (< 100 beds) (Reference) | 9,680 | 11.8% | 1,895 | 19.6 (18.8 - 20.4) | - | . |
| Medium (100 - 500 beds) | 52,505 | 64.2% | 13,740 | 26.2 (25.8 - 26.5) | 1.14 (1.06 - 1.22) | 0.000 |
| Large (> 500 beds) | 15,638 | 19.1% | 4,277 | 27.4 (26.7 - 28.1) | 1.09 (1.01 - 1.19) | 0.034 |
| **ICU/CCU Stay** |  |  |  |  |  |  |
| No (Reference) | 33,353 | 40.8% | 3,144 | 9.4 (9.1 - 9.7) | - | . |
| Yes | 48,398 | 59.2% | 17,732 | 36.6 (36.2 - 37.1) | 2.71 (2.57 - 2.85) | <.0001 |
| **Mechanical Ventilation** |  |  |  |  |  |  |
| No (Reference) | 67,484 | 82.5% | 10,330 | 15.3 (15.0 - 15.6) | - | . |
| Less Than 96hrs | 4,831 | 5.9% | 3,599 | 74.5 (73.2 - 75.7) | 13.11 (12.11 - 14.20) | <.0001 |
| More Than 96hrs | 9,436 | 11.5% | 6,947 | 73.6 (72.7 - 74.5) | 11.59 (10.89 - 12.34) | <.0001 |
| **Length of Stay<sup>d</sup>** |  |  |  |  |  |  |
| 1-2 Days | 1,251 | 1.5% | 284 | 22.7 (20.4 - 25.1) | - | - |
| 3-4 Days | 8,740 | 10.7% | 1,447 | 16.6 (15.8 - 17.4) | - | - |
| 5-6 Days | 16,244 | 19.9% | 2,028 | 12.5 (12.0 - 13.0) | - | - |
| 7-10 Days | 21,850 | 26.7% | 4,573 | 20.9 (20.4 - 21.5) | - | - |
| 11+ Days | 33,641 | 41.2% | 12,544 | 37.3 (36.8 - 37.8) | - | - |
| **Intravenous Immunoglobulin/ Hyperimmune Globulin** |  |  |  |  |  |  |
| No (Reference) | 81,684 | 99.9% | 20,853 | 25.5 (25.2 - 25.8) | - | . |
| Yes | 67 | 0.1% | 23 | 34.3 (23.4 - 47.0) | 1.24 (0.63 - 2.47) | 0.533 |
| **Tocilizumab** |  |  |  |  |  |  |
| No (Reference) | 78,986 | 96.6% | 19,721 | 25.0 (24.7 - 25.3) | - | . |
| Yes | 2,765 | 3.4% | 1,155 | 41.8 (39.9 - 43.6) | 1.65 (1.49 - 1.83) | <.0001 |
| **Baricitinib** |  |  |  |  |  |  |
| No (Reference) | 80,969 | 99.0% | 20,622 | 25.5 (25.2 - 25.8) | - | . |
| Yes | 782 | 1.0% | 254 | 32.5 (29.2 - 35.9) | 1.49 (1.22 - 1.82) | <.0001 |
| **Remdesivir** |  |  |  |  |  |  |
| No (Reference) | 19,913 | 24.4% | 5,431 | 27.3 (26.7 - 27.9) | - | . |
| Yes | 61,838 | 75.6% | 15,445 | 25.0 (24.6 - 25.3) | 0.94 (0.90 - 0.99) | 0.021 |
| **Charlson Comorbidity Index (CCI)** |  |  |  |  |  |  |
| 0 (Reference) | 14,278 | 17.5% | 2,525 | 17.7 (17.1 - 18.3) | - | . |
| 1 | 20,503 | 25.1% | 4,421 | 21.6 (21.0 - 22.1) | 1.10 (1.02 - 1.19) | 0.011 |
| 2 | 15,940 | 19.5% | 4,173 | 26.2 (25.5 - 26.9) | 1.23 (1.12 - 1.34) | <.0001 |
| 3 – 4 | 18,775 | 23.0% | 5,636 | 30.0 (29.4 - 30.7) | 1.23 (1.12 - 1.36) | <.0001 |
| 5+ | 12,255 | 15.0% | 4,121 | 33.6 (32.8 - 34.5) | 1.31 (1.15 - 1.50) | <.0001 |
| **90-Day Comorbidity History** |  |  |  |  |  |  |
| **Thrombosis History** |  |  |  |  |  |  |
| No (Reference) | 79,434 | 97.2% | 20,091 | 25.3 (25.0 - 25.6) | - | . |
| Yes | 2,317 | 2.8% | 785 | 33.9 (32.0 - 35.9) | 1.00 (0.89 - 1.13) | 0.977 |
| **ESRD History** |  |  |  |  |  |  |
| No (Reference) | 79,306 | 97.0% | 20,158 | 25.4 (25.1 - 25.7) | - | . |
| Yes | 2,445 | 3.0% | 718 | 29.4 (27.6 - 31.2) | 0.87 (0.75 - 1.00) | 0.057 |
| **Elixhauser Comorbidity History** |  |  |  |  |  |  |
| **Alcohol Abuse** |  |  |  |  |  |  |
| No (Reference) | 80,636 | 98.6% | 20,554 | 25.5 (25.2 - 25.8) | - | . |
| Yes | 1,115 | 1.4% | 322 | 28.9 (26.3 - 31.7) | 0.91 (0.76 - 1.09) | 0.290 |
| **Blood Loss Anemia** |  |  |  |  |  |  |
| No (Reference) | 81,369 | 99.5% | 20,744 | 25.5 (25.2 - 25.8) | - | . |
| Yes | 382 | 0.5% | 132 | 34.6 (29.8 - 39.6) | 1.21 (0.92 - 1.59) | 0.181 |
| **Cardiac Arrhythmia** |  |  |  |  |  |  |
| No (Reference) | 51,564 | 63.1% | 10,542 | 20.4 (20.1 - 20.8) | - | . |
| Yes | 30,187 | 36.9% | 10,334 | 34.2 (33.7 - 34.8) | 1.35 (1.29 - 1.41) | <.0001 |
| **Chronic Pulmonary Disease** |  |  |  |  |  |  |
| No (Reference) | 55,800 | 68.3% | 14,067 | 25.2 (24.9 - 25.6) | - | . |
| Yes | 25,951 | 31.7% | 6,809 | 26.2 (25.7 - 26.8) | 1.02 (0.97 - 1.07) | 0.477 |
| **Coagulopathy** |  |  |  |  |  |  |
| No (Reference) | 67,480 | 82.5% | 15,435 | 22.9 (22.6 - 23.2) | - | . |
| Yes | 14,271 | 17.5% | 5,441 | 38.1 (37.3 - 38.9) | 1.29 (1.23 - 1.36) | <.0001 |
| **Congestive Heart Failure** |  |  |  |  |  |  |
| No (Reference) | 59,178 | 72.4% | 13,709 | 23.2 (22.8 - 23.5) | - | . |
| Yes | 22,573 | 27.6% | 7,167 | 31.8 (31.1 - 32.4) | 1.01 (0.95 - 1.07) | 0.701 |
| **Deficiency Anemia** |  |  |  |  |  |  |
| No (Reference) | 77,844 | 95.2% | 19,785 | 25.4 (25.1 - 25.7) | - | . |
| Yes | 3,907 | 4.8% | 1,091 | 27.9 (26.5 - 29.4) | 0.96 (0.87 - 1.05) | 0.343 |
| **Depression** |  |  |  |  |  |  |
| No (Reference) | 69,281 | 84.7% | 17,856 | 25.8 (25.4 - 26.1) | - | . |
| Yes | 12,470 | 15.3% | 3,020 | 24.2 (23.5 - 25.0) | 1.03 (0.97 - 1.09) | 0.336 |
| **Diabetes** |  |  |  |  |  |  |
| No (Reference) | 46,022 | 56.3% | 11,068 | 24.0 (23.7 - 24.4) | - | . |
| Yes | 35,729 | 43.7% | 9,808 | 27.5 (27.0 - 27.9) | 0.96 (0.91 - 1.01) | 0.135 |
| **Fluid and Electrolyte Disorder** |  |  |  |  |  |  |
| No (Reference) | 30,902 | 37.8% | 4,414 | 14.3 (13.9 - 14.7) | - | . |
| Yes | 50,849 | 62.2% | 16,462 | 32.4 (32.0 - 32.8) | 1.56 (1.48 - 1.63) | <.0001 |
| **Hypertension (uncomplicated)** |  |  |  |  |  |  |
| No (Reference) | 45,340 | 55.5% | 12,816 | 28.3 (27.9 - 28.7) | - | . |
| Yes | 36,411 | 44.5% | 8,060 | 22.1 (21.7 - 22.6) | 0.95 (0.90 - 1.01) | 0.085 |
| **Hypertension (complicated)** |  |  |  |  |  |  |
| No (Reference) | 47,692 | 58.3% | 10,439 | 21.9 (21.5 - 22.3) | - | . |
| Yes | 34,059 | 41.7% | 10,437 | 30.6 (30.2 - 31.1) | 1.07 (1.00 - 1.14) | 0.047 |
| **Liver Disease** |  |  |  |  |  |  |
| No (Reference) | 77,923 | 95.3% | 19,290 | 24.8 (24.5 - 25.1) | - | . |
| Yes | 3,828 | 4.7% | 1,586 | 41.4 (39.9 - 43.0) | 1.44 (1.31 - 1.58) | <.0001 |
| **Metastatic Cancer** |  |  |  |  |  |  |
| No (Reference) | 80,620 | 98.6% | 20,540 | 25.5 (25.2 - 25.8) | - | . |
| Yes | 1,131 | 1.4% | 336 | 29.7 (27.1 - 32.5) | 1.19 (0.98 - 1.45) | 0.084 |
| **Obesity** |  |  |  |  |  |  |
| No (Reference) | 61,281 | 75.0% | 15,485 | 25.3 (24.9 - 25.6) | - | . |
| Yes | 20,470 | 25.0% | 5,391 | 26.3 (25.7 - 26.9) | 1.04 (0.99 - 1.10) | 0.102 |
| **Other Neurological Disorders** |  |  |  |  |  |  |
| No (Reference) | 62,539 | 76.5% | 13,687 | 21.9 (21.6 - 22.2) | - | . |
| Yes | 19,212 | 23.5% | 7,189 | 37.4 (36.7 - 38.1) | 1.31 (1.25 - 1.38) | <.0001 |
| **Paralysis** |  |  |  |  |  |  |
| No (Reference) | 80,683 | 98.7% | 20,449 | 25.3 (25.0 - 25.6) | - | . |
| Yes | 1,068 | 1.3% | 427 | 40.0 (37.0 - 43.0) | 1.01 (0.85 - 1.20) | 0.881 |
| **Peptic Ulcer Disease** |  |  |  |  |  |  |
| No (Reference) | 81,571 | 99.8% | 20,829 | 25.5 (25.2 - 25.8) | - | . |
| Yes | 180 | 0.2% | 47 | 26.1 (20.0 - 33.3) | 0.49 (0.32 - 0.78) | 0.002 |
| **Peripheral Vascular Disorder** |  |  |  |  |  |  |
| No (Reference) | 74,962 | 91.7% | 18,927 | 25.2 (24.9 - 25.6) | - | . |
| Yes | 6,789 | 8.3% | 1,949 | 28.7 (27.6 - 29.8) | 0.98 (0.91 - 1.06) | 0.617 |
| **Pulmonary Circulation Disorder** |  |  |  |  |  |  |
| No (Reference) | 78,943 | 96.6% | 19,934 | 25.3 (24.9 - 25.6) | - | . |
| Yes | 2,808 | 3.4% | 942 | 33.5 (31.8 - 35.3) | 1.05 (0.93 - 1.17) | 0.448 |
| **Renal Failure** |  |  |  |  |  |  |
| No (Reference) | 66,920 | 81.9% | 16,001 | 23.9 (23.6 - 24.2) | - | . |
| Yes | 14,831 | 18.1% | 4,875 | 32.9 (32.1 - 33.6) | 1.07 (0.99 - 1.15) | 0.073 |
| **Rheumatoid Arthritis/Collagen Vascular Diseases (CVD)** |  |  |  |  |  |  |
| No (Reference) | 77,950 | 95.4% | 19,873 | 25.5 (25.2 - 25.8) | - | . |
| Yes | 3,801 | 4.6% | 1,003 | 26.4 (25.0 - 27.8) | 1.17 (1.06 - 1.29) | 0.002 |
| **Solid Tumor** |  |  |  |  |  |  |
| No (Reference) | 78,697 | 96.3% | 20,012 | 25.4 (25.1 - 25.7) | - | . |
| Yes | 3,054 | 3.7% | 864 | 28.3 (26.7 - 29.9) | 1.06 (0.94 - 1.20) | 0.316 |
| **Valvular Disease** |  |  |  |  |  |  |
| No (Reference) | 75,338 | 92.2% | 19,016 | 25.2 (24.9 - 25.6) | - | . |
| Yes | 6,413 | 7.8% | 1,860 | 29.0 (27.9 - 30.1) | 0.94 (0.88 - 1.02) | 0.134 |
| **Weight Loss** |  |  |  |  |  |  |
| No (Reference) | 72,896 | 89.2% | 17,464 | 24.0 (23.6 - 24.3) | - | . |
| Yes | 8,855 | 10.8% | 3,412 | 38.5 (37.5 - 39.6) | 1.15 (1.08 - 1.22) | <.0001 |

<sup>a</sup> Patients with missing values for ADI, Urban/Rural, Hospital Size and Ownership were dropped from the multivariate analysis.
<sup>b</sup> The South US census region includes counts for "Other" territories to avoid identification due to low counts.
<sup>c</sup> Patients have not had COVID-19 diagnosis recorded in the 30 days prior to admission.
<sup>d</sup> Length of Stay was not included in the adjusted model due to concerns about reverse causation.
**Notes:** Model evaluation metrics were ROC Area Under the Curve (0.8476); Pearson Chi Sq Goodness of Fit test p value (1); Deviance Goodness of Fit test p value (1)

Figure 1 shows crude IP mortality by admission months among CCP recipients during the study period, overall and by IC status, with significantly declining mortality (p<0.001) overall: from 30.7% (95%CI:29.1%-32.3%) in August 2020 to 7.4% (95%CI:3.8%-15.0%) in December 2021; and for non-IC: from 31.1% (95%CI:29.4%-32.8%) to 8.6% (95%CI:4.4%-17.2%), respectively. Also, significantly declining IP mortality (p<0.001) was identified in the pre-Delta variant circulation period (August 2020-May 2021) overall and for non-IC status; and, in contrast, an increasing mortality trend (p=0.02) occurred during Delta circulation period (June 2021-November 2021) among non-IC. Among IC, there were no significant changes in the inpatient mortality rates during study period. Supplementary Figures 1 through 4 show monthly IP mortality rates by U.S. regions, with significantly declining trends (p<0.001) in the South and the Midwest, overall and for non-IC; and non-significant changes in monthly mortality in the West and Northeast, overall and by IC status.

**Figure 1:**
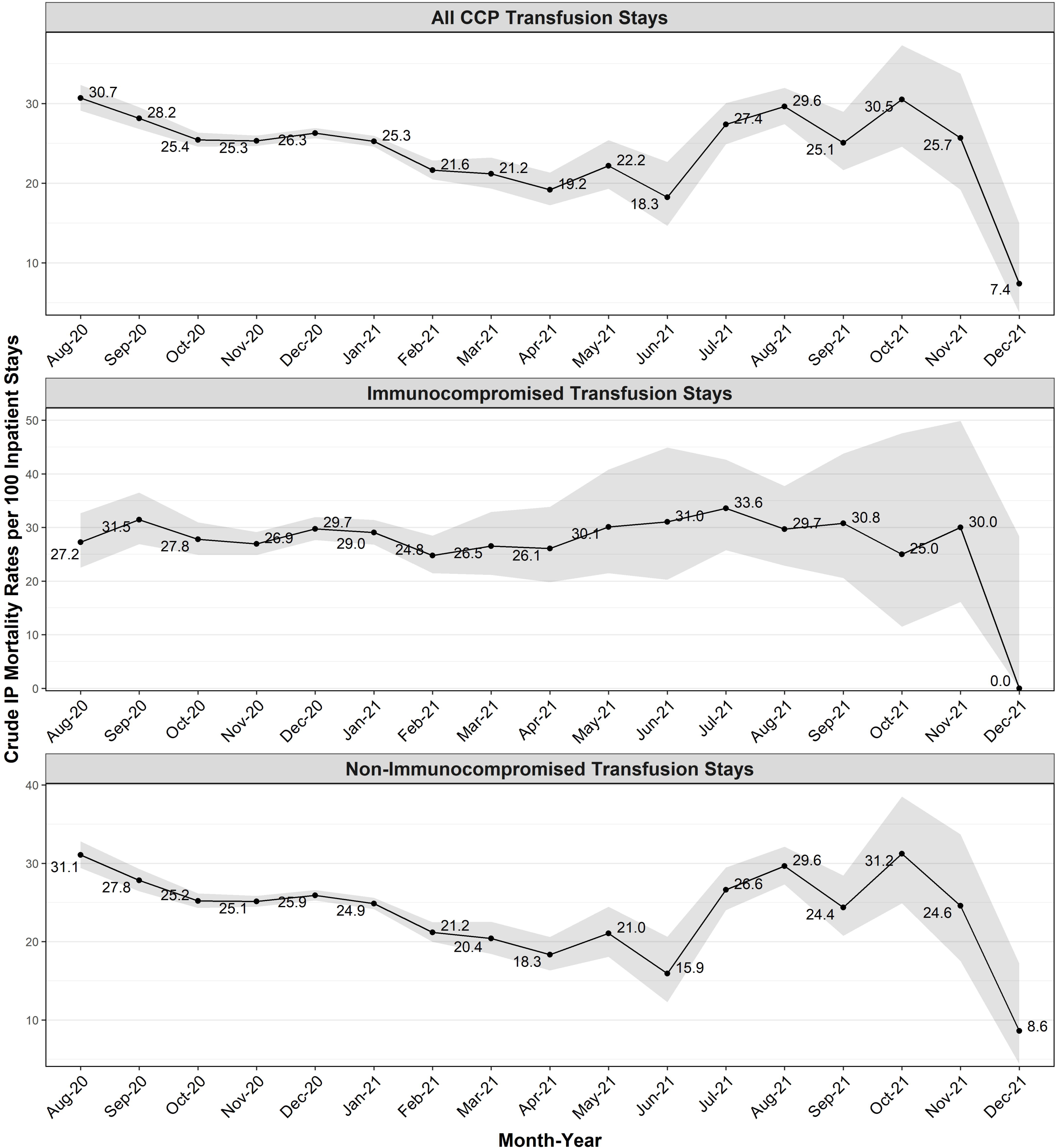
Monthly Inpatient (IP) Mortality Rates among Inpatient COVID-19 Convalescent Plasma (CCP) Recipients, Overall and by IC Status. **Alt text:** Figure 1 shows crude monthly inpatient mortality rates per 100 inpatient COVID-19 stays with convalescent plasma transfusion, overall and by immunocompromised status. It shows significantly declining rates overall and for non-immunocompromised.

### Factors Associated with Inpatient Mortality

The overall and subgroup multivariable regression analyses showed generally good model fit, good predictive ability, and small VIFs suggesting either no or low potential correlations between covariates in the final models. The overall adjusted analysis (Table 1) showed significantly higher inpatient mortality odds for: IC vs. non-IC (1.14; 95%CI:1.05-1.24); males vs. females (1.20; 95%CI:1.15-1.25); Medicaid-eligible (1.13; 95%CI:1.07-1.20); nursing home- vs. community-dwelling patients (1.19; 95%CI:1.11-1.28); West vs. South (1.17; 95%CI:1.10-1.25); Northeast vs. South (1.19; 95%CI:1.10-1.28); rural vs. urban residents (1.12; 95%CI:1.06-1.18); thrombosis during hospital stay (1.35; 95%CI:1.27-1.43); ICU/CCU admissions (2.71; 95%CI:2.57-2.85); MV<96 hours (13.11; 95%CI:12.11-14.20); and MV≥96 hours (11.59; 95%CI:10.89-12.34). Significantly higher odds of IP mortality were identified for CCI 5+ vs. 0 (1.31; 95%CI:1.15-1.50) and for CCI 1-4 vs. 0 (p<0.05). Significantly lower IP mortality odds were identified with ADI 0-20 (0.85; 95%CI:0.78-0.92) and ADI 21-40 (0.86; 95%CI:0.81-0.93) when compared to ADI 81-100. Significantly higher mortality was identified with increasing age (1.06; 95%CI:1.06-1.07).

In the overall adjusted analysis, study showed significantly increasing inpatient mortality odds with higher CCI (1.038; 95%CI:1.016-1.060) and ADI (1.002; 95%CI:1.002-1.003), treated as continuous variables (data not shown). Supplementary Figures 5-9 show MVRS graphs of adjusted inpatient mortality odds by age, ADI, CCI scores, time since recent COVID-19, and by admission months treated as continuous variables. The odds of inpatient mortality increased linearly with higher age (Supplementary Figure 5), with higher ADI up to ADI>60, and with higher CCI scores up to CCI>6 (Supplementary Figures 6 and 7). Inpatient mortality varied non-linearly with time since most recent COVID-19 diagnosis (Supplementary Figure 8). Supplementary Figure 9 shows that the adjusted odds of mortality peaked twice, around December 2020 and August 2021 (the Delta variant circulation).

Overall, histories of specific conditions showed significantly higher adjusted odds of IP mortality, including cardiac arrhythmia (1.35, 95%CI:1.29-1.41), fluid and electrolyte disorders (1.56, 95%CI:1.48-1.63), liver disease (1.44, 95%CI:1.31-1.58), other neurologic disorders (1.31, 95%CI:1.25-1.38), and rheumatoid arthritis/collagen vascular diseases (CVD) (1.17, 95%CI:1.06-1.29), among others. Medium (1.14; 95%CI:1.06-1.22) and large (1.09; 95%CI:1.01-1.19) hospitals showed higher mortality odds as compared to small hospitals. Lower adjusted odds of IP mortality were identified for remdesivir users (0.94; 95%CI:0.90-0.99) and for the for-profit (0.79; 95%CI:0.73-0.85) and non-for-profit (0.86; 95%CI:0.80-0.92) hospitals as compared to government hospitals. Higher mortality odds were identified for CCP stays with COVID-19 diagnosis recorded within 4-10 days prior to admission (1.08; 95%CI:1.02-1.14); and lower mortality, with COVID-19 diagnosis recorded within 11-30 days prior to admission (0.83; 95%CI:0.75-0.91) as compared to stays with no COVID-19 within 30 days prior to admission. (Table 1)

Supplementary Tables 1-17 show CCP-recipient characteristics with corresponding crude inpatient mortality and adjusted ORs by pre-specified subgroups (e.g., calendar year, IC status, residence regions, demographics, time since the most recent COVID-19 diagnosis, Medicaid eligibility). Risk factors for IP mortality were generally consistent for all the subgroups, with some differences identified. For 2021 but not for 2020 (Supplementary Tables 1 and 2), the mortality odds were significantly lower for remdesivir recipients (0.87; 95%CI 0.80-0.95) and for emergency room admission stays (0.87; 95%CI 0.76-1.00) and were not different for rural vs urban residents (1.04; 95%CI 0.94-1.14). In 2021 vs. 2020, CCP recipients had the following rates: crude IP mortality (24.2% vs. 26.2%), MV (16.1% vs. 18.2%), and ICU/CCU admission (57.2% vs. 60.3%). Among IC (Supplementary Table 3), higher odds of IP mortality were identified with history of metastatic cancer (1.36; 95%CI 1.02-1.82) and radiation (1.43; 95%CI 1.09-1.88). For IC, the use of remdesivir (1.03; 95%CI 0.89-1.20) as well as some other characteristics (e.g. Medicaid eligibility, rural vs. urban residency) did not have significant impact on mortality. In contrast, among non-IC (Supplementary Table 4), remdesivir users had lower IP mortality odds (0.93; 95%CI 0.88-0.98). For specific regions [13] (Supplementary Tables 5-8), highest crude inpatient mortality was identified for West Pacific (33.5%) and lowest for Northeast New England (22.6%). In the South (Supplementary Table 5), significantly higher adjusted odds of inpatient mortality were seen for East South Central vs. South Atlantic residents (1.09; 95%CI 1.01-1.19); in the Midwest (Supplementary Table 6): for West North Central vs. East North Central (1.10; 95%CI 1.01-1.21); in the West (Supplementary Table 7): for Pacific vs. Mountain (1.31; 95%CI 1.15-1.50); and in the Northeast (Supplementary Table 8): for Middle Atlantic vs. New England residents (1.26; 95%CI 1.05-1.52).

## DISCUSSION

Our population-based retrospective nationwide inferential study on inpatient mortality and risk factors among inpatient Medicare beneficiaries ages 65+ who received CCP for treatment of COVID-19 shows variation of mortality over time and by patient and hospital characteristics. The study findings show significant associations of inpatient mortality with demographics, underlying comorbidities, socioeconomic status, residence, COVID-19 severity, hospital size and ownership. Overall, our study is consistent with published literature [8,11,12] and shows that inpatient CCP recipients ages 65+ had severe COVID-19 (i.e., high inpatient mortality, ICU/CCU admission, MV, and long LOS) during the study period. The findings identified a significantly declining monthly inpatient mortality overall and for non-IC recipients during August 2020-December 2021 but not among IC, which could be related in-part to greater IC patient severity. For the year 2021 vs. 2020, CCP recipients had lower rates of inpatient mortality, mechanical ventilation, ICU/CCU admission and shorter lengths of stay (data not shown), further supporting declining risk of severe COVID-19 outcomes over time [8,20]. The declining mortality among CCP recipients coincided with the increasing availability of vaccination and other COVID-19 treatments (e.g., monoclonal antibodies), infection-acquired immunity, recommendations for timing CCP administration early in the COVID-19 infection as well as with the emergence of the less severe Omicron variant at the end of 2021. [10,20–26]

Among CCP recipients with COVID-19, the study shows higher adjusted inpatient mortality odds for: IC vs. non-IC, males vs. females, Medicaid- vs. non-Medicaid-eligible; and with: increasing age, higher ADI, ICU/CCU admissions, mechanical ventilation, thrombosis during stay, higher CCI scores, and histories of cardiac arrhythmia, other neurological disorders, rheumatoid arthritis/CVD, and coagulopathy, among others; as well as for: nursing home- vs. community-dwelling beneficiaries, rural vs. urban residents, residents of Northeast and West vs. South, and for medium- and large- vs. small-size hospitals. Many of the study findings were consistent with Senefeld et al. [8] and Perichon et al. [27]. Inpatient mortality risk was found to be lower for CCP recipients in the least disadvantaged neighborhoods, at for-profit and non-profit vs. government-owned hospitals, with remdesivir administration, and with the most recent COVID-19 diagnosis recorded 11-30 days prior to inpatient admission (i.e., non-recent COVID- 19). Compared to the EAP cohort in a study by Senefeld et al. [8], CCP recipients in our study were more likely to be older and with history of pulmonary diseases, diabetes, and remdesivir administration, among other differences. Despite the differences, CCP recipients in both studies had severe COVID-19 infections with similar mortality rates, but with underlying comorbidities more likely to be present (i.e. higher pre-existing risk) in the Medicare beneficiaries aged 65+ whom we studied. Based on the CCP treatment benefit index (TBI) developed by Park et al. [28], inpatient CCP recipients ages 65+ depending on COVID-19 stage (i.e. severity) may be hypothesized to have received treatment benefit with CCP administration, which is supported by the literature [29,30] and needs further confirmation as our study was not designed to evaluate CCP effectiveness.

The study findings showed some variations in risk factors for inpatient mortality by specific population subgroups we evaluated. For IC, significantly higher mortality risk was identified for those with metastatic cancer and radiation, with a non-significant impact by hospital characteristics and remdesivir use, suggesting effects of more severe underlying conditions. Also, for IC, no significant differences in mortality risk were identified for Medicaid-eligible vs. not, rural vs. urban residence, and by area deprivation index, suggesting less impact of socioeconomic status and residence. For CCP recipients ages 65-79, significantly higher mortality was identified in beneficiaries with COVID-19 recorded as a reason for admission and with history of metastatic cancer. For hospitalizations with recent COVID-19 (i.e., recorded within 1-10 days prior to admission), higher mortality was identified in beneficiaries with COVID-19 recorded as a reason for admission and with histories of renal failure and obesity, among others. For hospitalizations with non-recent COVID-19 (i.e., within 11-30 days prior to admission) vs. recent COVID-19, no significant differences in mortality risk were identified by Medicaid eligibility status, rural vs. urban residence, and by IC status, additionally suggesting the importance of recency of COVID-19 infection. For Medicaid-eligible beneficiaries, IC status was not associated with inpatient mortality further suggesting the socioeconomic status to be amongst the major determinants of severe outcomes in COVID-19 patients. For 2021 vs. 2020, no association with inpatient mortality was identified for rural vs. urban residence and by hospital size, whereas a significantly lower mortality risk was found with emergency room admissions and with remdesivir administrations, suggesting effects of better quality of care for COVID-19 and of lower COVID-19 severity.

Our retrospective study used administrative claims databases, and thus limitations include potential under- or mis-recording of CCP transfusion and COVID-19 diagnosis; as well as lack of clinical details to validate procedure and diagnosis codes. It was based on CCP transfusion coding availability starting from August 2020. The study findings are not generalizable to the U.S. population less than 65 years of age, and small counts for some groups may have led to spurious associations or false-negative results. Exclusion of stays with other blood component transfusions may have resulted in less severe CCP recipients evaluated thus potentially affecting generalizability of study results. Although our study adjusted for possible confounding covariates, an unmeasured confounding could still be present due to lack of full information in the claims data on clinical COVID-19 infection severity measures, COVID-19 treatment prescribing practices, among others, which could have affected study results. The study included community-dwelling and nursing home elderly, who typically have different characteristics (e.g., demographics, frailty), and thus future investigations are needed to study them separately. Also, the study did not evaluate the effects of vaccination history among inpatient CCP recipients in 2021, which could have influenced the magnitude of results. Further analyses are needed to evaluate CCP utilization and risk factors among the inpatient U.S. elderly. Finally, causality cannot be established, and thus future investigations are needed to confirm the findings.

In summary, our investigation of real-world data describes the population of Medicare beneficiaries ages 65+ who received CCP for treatment of COVID-19. The study demonstrates the variation of inpatient mortality by characteristics evaluated. The findings show among the highest inpatient mortality risk in beneficiaries requiring mechanical ventilation or ICU/CCU admission. The study identified a consistently higher inpatient mortality risk with advancing age, and for: males, socioeconomically disadvantaged beneficiaries (i.e., for Medicaid-eligible and with higher ADI), immunocompromised beneficiaries, and those residing in the rural areas, nursing homes, and in the West and Northeast. It also shows the importance of underlying comorbidities. Finally, consistent with other reports [31,32], we found that most of the CCP use in the elderly occurred during 2020, and factors influencing CCP use need investigation. Future investigations among U.S. older adults may also be needed to evaluate real-world effectiveness of CCP in preventing inpatient mortality for various patient subgroups (e.g., ages 65-79 vs. 80+, IC vs. non-IC, and by COVID-19 severity). Overall, the study provides important findings on the risk factors for mortality among inpatient elderly treated with CCP for COVID-19. The study findings can help inform potential future evaluation of real-world CCP effectiveness in different high-risk populations as well as development and implementation of prevention strategies that help to improve the survival of older COVID-19 patients.

## Acknowledgments

Our colleague and friend, Jeffrey A. Kelman, MD, passed away on February 8, 2024. Dr. Kelman was the Chief Medical Officer for the Center for Medicare at the Centers for Medicare and Medicaid Services. This study and the many others we have performed using Medicare data, on the use and safety of blood components and products, were possible thanks to the advocacy, dedication, and scientific acumen of Dr. Kelman. We would like to honor his extraordinary scientific contributions to public health.

## Notes

### Author Contributions

Nathan Duma, Sarah Barney, and Yoganand Chillarige, contributed to study design, data collection and analysis, results interpretation, manuscript writing and review. Mikhail Menis, Stephen H. Chang, Whitney R. Steele, Carlos H. Villa, Anne F. Eder, and Barbee I. Whitaker contributed to study design, results interpretation, manuscript writing and review.

### Disclaimer

The views expressed in this manuscript are those of the authors and do not reflect official policy of the FDA or the United States Government and do not bind or obligate FDA.

### Data availability

Data not publicly available. An individual wanting access to CMS data would have to file their own Data Use Agreement (DUA) with the Centers for Medicare and Medicaid Services and gain approval to have access. In order to request the data, the readers may contact ResDAC (Research Data Assistance Center) at http://www.resdac.org

### Institutional review board and patient informed consent

This study is a public health surveillance activity conducted and authorized by a public health authority and necessary to allow FDA to identify, monitor, assess or investigate potential public health signals, onsets of disease outbreaks, or conditions of public health importance under CFR 46.102(k) and 45 CFR 46.102(l)(2).The use of Medicare administrative data was approved by the Centers for Medicare and Medicaid Services (CMS) privacy board under a data use agreement; and institutional review board and patient informed consent were not required.

