## Supplementary for "Inpatient Mortality and Risk Factors Among Hospitalized Medicare Beneficiaries Ages 65 and Older with COVID-19 Treated with COVID-19 Convalescent Plasma (CCP), During 2020-2021"

| **Supplementary Table 1.** Inpatient (IP) Mortality Rates and Corresponding Adjusted Odds Ratios (ORs) by Patient and Hospital Characteristics Among Recipients of COVID-19 Convalescent Plasma (CCP) in 2020 | | | | | | |  |
| --- | --- | --- | --- | --- | --- | --- | --- |
| **Patient and Hospital Characteristics** | **CCP Transfusion Stays #** | **CCP Transfusion Stays  %** | **IP Mortality  #** | **Crude IP Mortality Rate per 100  (95% CIs)** | **Adjusted ORs (95% CIs)** | | **P-value** |
| **Overall** | 53,076 | 100.0% | 13,928 | 26.2 (25.9 - 26.6) | - | | . |
| **Age (continuous)** | 53,076 | 100.0% | 13,928 | 26.2 (25.9 - 26.6) | 1.06 (1.06 - 1.07) | | <.0001 |
| **Sex** |  |  |  |  |  | |  |
| Female (Reference) | 23,823 | 44.9% | 5,553 | 23.3 (22.8 - 23.9) | - | | . |
| Male | 29,253 | 55.1% | 8,375 | 28.6 (28.1 - 29.2) | 1.19 (1.13 - 1.26) | | <.0001 |
| **ADI^a^** |  |  |  |  |  | |  |
| 81 - 100 (Reference) | 8,031 | 15.1% | 2,288 | 28.5 (27.5 - 29.5) | - | | . |
| 61 – 80 | 11,427 | 21.5% | 3,024 | 26.5 (25.7 - 27.3) | 0.97 (0.90 - 1.06) | | 0.508 |
| 41 – 60 | 12,007 | 22.6% | 3,003 | 25.0 (24.2 - 25.8) | 0.92 (0.85 - 1.00) | | 0.043 |
| 21 – 40 | 10,390 | 19.6% | 2,605 | 25.1 (24.2 - 25.9) | 0.86 (0.79 - 0.94) | | 0.001 |
| 0 – 20 | 6,223 | 11.7% | 1,583 | 25.4 (24.4 - 26.5) | 0.83 (0.75 - 0.92) | | 0.000 |
| **Medicaid Eligibility** |  |  |  |  |  | |  |
| No (Reference) | 41,879 | 78.9% | 10,535 | 25.2 (24.7 - 25.6) | - | | . |
| Yes | 11,197 | 21.1% | 3,393 | 30.3 (29.5 - 31.2) | 1.15 (1.07 - 1.23) | | 0.000 |
| **Region of Residence^b^** |  |  |  |  |  | |  |
| South (Reference) | 26,429 | 49.8% | 6,841 | 25.9 (25.4 - 26.4) | - | | . |
| Midwest | 15,211 | 28.7% | 3,757 | 24.7 (24.0 - 25.4) | 1.04 (0.98 - 1.11) | | 0.203 |
| West | 6,971 | 13.1% | 2,117 | 30.4 (29.3 - 31.5) | 1.20 (1.10 - 1.30) | | <.0001 |
| Northeast | 4,465 | 8.4% | 1,213 | 27.2 (25.9 - 28.5) | 1.15 (1.04 - 1.27) | | 0.005 |
| **Urban/Rural^a^** |  |  |  |  |  | |  |
| Urban (Reference) | 35,391 | 66.7% | 9,245 | 26.1 (25.7 - 26.6) | - | | . |
| Rural | 13,136 | 24.7% | 3,386 | 25.8 (25.0 - 26.5) | 1.16 (1.09 - 1.24) | | <.0001 |
| **Immunocompromised Status** |  |  |  |  |  | |  |
| No (Reference) | 48,030 | 90.5% | 12,493 | 26.0 (25.6 - 26.4) | - | | . |
| Yes | 5,046 | 9.5% | 1,435 | 28.4 (27.2 - 29.7) | 1.10 (0.99 - 1.22) | | 0.083 |
| **Nursing Home Stay** |  |  |  |  |  | |  |
| No (Reference) | 47,616 | 89.7% | 12,143 | 25.5 (25.1 - 25.9) | - | | . |
| Yes | 5,460 | 10.3% | 1,785 | 32.7 (31.5 - 34.0) | 1.18 (1.08 - 1.29) | | 0.000 |
| **Emergency Room Stay** |  |  |  |  |  | |  |
| No (Reference) | 6,476 | 12.2% | 1,840 | 28.4 (27.3 - 29.5) | - | | . |
| Yes | 46,600 | 87.8% | 12,088 | 25.9 (25.5 - 26.3) | 0.98 (0.90 - 1.07) | | 0.640 |
| **COVID-19 As a Reason for Admission** |  |  |  |  |  | |  |
| No (Reference) | 25,541 | 48.1% | 7,138 | 27.9 (27.4 - 28.5) | - | | . |
| Yes | 27,535 | 51.9% | 6,790 | 24.7 (24.2 - 25.2) | 1.04 (0.99 - 1.10) | | 0.142 |
| **Time Since Most Recent COVID-19 Diagnosis in the 30 Days Prior to Admission** |  |  |  |  |  | |  |
| 0 days (Reference)^c^ | 26,363 | 49.7% | 6,978 | 26.5 (25.9 - 27.0) | - | | . |
| 1 - 3 days | 12,592 | 23.7% | 3,174 | 25.2 (24.5 - 26.0) | 0.96 (0.90 - 1.02) | | 0.204 |
| 4 - 10 days | 11,263 | 21.2% | 3,062 | 27.2 (26.4 - 28.0) | 1.06 (0.99 - 1.14) | | 0.073 |
| 11-30 days | 2,858 | 5.4% | 714 | 25.0 (23.4 - 26.6) | 0.84 (0.75 - 0.94) | | 0.003 |
| **Thrombosis on Index** |  |  |  |  |  | |  |
| No (Reference) | 45,704 | 86.1% | 10,803 | 23.6 (23.3 - 24.0) | - | | . |
| Yes | 7,372 | 13.9% | 3,125 | 42.4 (41.3 - 43.5) | 1.33 (1.24 - 1.44) | | <.0001 |
| **Hospital Ownership^a^** |  |  |  |  |  | |  |
| Government (Reference) | 6,517 | 12.3% | 1,739 | 26.7 (25.6 - 27.8) | - | | . |
| For Profit | 9,534 | 18.0% | 2,571 | 27.0 (26.1 - 27.9) | 0.76 (0.69 - 0.83) | | <.0001 |
| Non-Profit | 36,953 | 69.6% | 9,602 | 26.0 (25.5 - 26.4) | 0.82 (0.76 - 0.89) | | <.0001 |
| **Hospital Size^a^** |  |  |  |  |  | |  |
| Small (< 100 beds) (Reference) | 6,143 | 11.6% | 1,205 | 19.6 (18.6 - 20.6) | - | | . |
| Medium (100 - 500 beds) | 34,043 | 64.1% | 9,172 | 26.9 (26.5 - 27.4) | 1.18 (1.08 - 1.29) | | 0.000 |
| Large (> 500 beds) | 10,323 | 19.4% | 2,899 | 28.1 (27.2 - 29.0) | 1.14 (1.03 - 1.27) | | 0.010 |
| **ICU/CCU Stay** |  |  |  |  |  | |  |
| No (Reference) | 21,071 | 39.7% | 2,022 | 9.6 (9.2 - 10.0) | - | | . |
| Yes | 32,005 | 60.3% | 11,906 | 37.2 (36.7 - 37.7) | 2.74 (2.57 - 2.92) | | <.0001 |
| **Mechanical Ventilation** |  |  |  |  |  | |  |
| No (Reference) | 43,415 | 81.8% | 6,850 | 15.8 (15.4 - 16.1) | - | | . |
| Less Than 96hrs | 3,066 | 5.8% | 2,243 | 73.2 (71.5 - 74.7) | 11.72 (10.63 - 12.93) | | <.0001 |
| More Than 96hrs | 6,595 | 12.4% | 4,835 | 73.3 (72.2 - 74.4) | 11.33 (10.51 - 12.23) | | <.0001 |
| **Tocilizumab** |  |  |  |  |  | |  |
| No (Reference) | 51,758 | 97.5% | 13,371 | 25.8 (25.5 - 26.2) | - | | . |
| Yes | 1,318 | 2.5% | 557 | 42.3 (39.6 - 45.0) | 1.59 (1.37 - 1.85) | | <.0001 |
| **Baricitinib** |  |  |  |  |  | |  |
| No (Reference) | 52,993 | 99.8% | 13,896 | 26.2 (25.8 - 26.6) | - | | . |
| Yes | 83 | 0.2% | 32 | 38.6 (28.3 - 49.9) | 1.29 (0.72 - 2.30) | | 0.387 |
| **Remdesivir** |  |  |  |  |  | |  |
| No (Reference) | 13,473 | 25.4% | 3,680 | 27.3 (26.6 - 28.1) | - | | . |
| Yes | 39,603 | 74.6% | 10,248 | 25.9 (25.4 - 26.3) | 0.98 (0.93 - 1.05) | | 0.610 |
| **Charlson Comorbidity Index (CCI)** |  |  |  |  |  | |  |
| 0 (Reference) | 8,986 | 16.9% | 1,649 | 18.4 (17.6 - 19.2) | - | | . |
| 1 | 13,251 | 25.0% | 2,924 | 22.1 (21.4 - 22.8) | 1.12 (1.02 - 1.23) | | 0.016 |
| 2 | 10,287 | 19.4% | 2,721 | 26.5 (25.6 - 27.3) | 1.22 (1.09 - 1.36) | | 0.000 |
| 3 – 4 | 12,364 | 23.3% | 3,826 | 30.9 (30.1 - 31.8) | 1.30 (1.15 - 1.47) | | <.0001 |
| 5+ | 8,188 | 15.4% | 2,808 | 34.3 (33.3 - 35.3) | 1.38 (1.18 - 1.62) | | <.0001 |
| **90-Day Comorbidity History** |  |  |  |  |  | |  |
| **Thrombosis History** |  |  |  |  |  | |  |
| No (Reference) | 51,555 | 97.1% | 13,392 | 26.0 (25.6 - 26.4) | - | | . |
| Yes | 1,521 | 2.9% | 536 | 35.2 (32.8 - 37.7) | 1.07 (0.92 - 1.23) | | 0.392 |
| **ESRD History** |  |  |  |  |  | |  |
| No (Reference) | 51,470 | 97.0% | 13,460 | 26.2 (25.8 - 26.5) | - | | . |
| Yes | 1,606 | 3.0% | 468 | 29.1 (26.9 - 31.4) | 0.88 (0.73 - 1.06) | | 0.174 |
| **Elixhauser Comorbidity History** |  |  |  |  |  | |  |
| **Blood Loss Anemia** |  |  |  |  |  | |  |
| No (Reference) | 52,826 | 99.5% | 13,843 | 26.2 (25.8 - 26.6) | - | | . |
| Yes | 250 | 0.5% | 85 | 34.0 (28.2 - 40.3) | 1.17 (0.83 - 1.65) | | 0.369 |
| **Cardiac Arrhythmia** |  |  |  |  |  | |  |
| No (Reference) | 33,267 | 62.7% | 7,001 | 21.1 (20.6 - 21.5) | - | | . |
| Yes | 19,809 | 37.3% | 6,927 | 35.0 (34.3 - 35.6) | 1.35 (1.28 - 1.43) | | <.0001 |
| **Chronic Pulmonary Disease** |  |  |  |  |  | |  |
| No (Reference) | 35,975 | 67.8% | 9,331 | 25.9 (25.5 - 26.4) | - | | . |
| Yes | 17,101 | 32.2% | 4,597 | 26.9 (26.2 - 27.6) | 0.99 (0.93 - 1.06) | | 0.820 |
| **Coagulopathy** |  |  |  |  |  | |  |
| No (Reference) | 43,884 | 82.7% | 10,395 | 23.7 (23.3 - 24.1) | - | | . |
| Yes | 9,192 | 17.3% | 3,533 | 38.4 (37.4 - 39.4) | 1.26 (1.18 - 1.34) | | <.0001 |
| **Congestive Heart Failure** |  |  |  |  |  | |  |
| No (Reference) | 38,014 | 71.6% | 9,019 | 23.7 (23.3 - 24.2) | - | | . |
| Yes | 15,062 | 28.4% | 4,909 | 32.6 (31.8 - 33.3) | 1.06 (0.98 - 1.14) | | 0.130 |
| **Deficiency Anemia** |  |  |  |  |  | |  |
| No (Reference) | 50,558 | 95.3% | 13,222 | 26.2 (25.8 - 26.5) | - | | . |
| Yes | 2,518 | 4.7% | 706 | 28.0 (26.3 - 29.8) | 0.95 (0.84 - 1.06) | | 0.348 |
| **Depression** |  |  |  |  |  | |  |
| No (Reference) | 44,689 | 84.2% | 11,838 | 26.5 (26.1 - 26.9) | - | | . |
| Yes | 8,387 | 15.8% | 2,090 | 24.9 (24.0 - 25.9) | 1.01 (0.94 - 1.08) | | 0.819 |
| **Diabetes** |  |  |  |  |  | |  |
| No (Reference) | 29,613 | 55.8% | 7,303 | 24.7 (24.2 - 25.2) | - | | . |
| Yes | 23,463 | 44.2% | 6,625 | 28.2 (27.7 - 28.8) | 0.95 (0.89 - 1.02) | | 0.172 |
| **Fluid and Electrolyte Disorder** |  |  |  |  |  | |  |
| No (Reference) | 20,008 | 37.7% | 2,927 | 14.6 (14.1 - 15.1) | - | | . |
| Yes | 33,068 | 62.3% | 11,001 | 33.3 (32.8 - 33.8) | 1.58 (1.49 - 1.68) | | <.0001 |
| **Hypertension (uncomplicated)** |  |  |  |  |  | |  |
| No (Reference) | 29,673 | 55.9% | 8,596 | 29.0 (28.5 - 29.5) | - | | . |
| Yes | 23,403 | 44.1% | 5,332 | 22.8 (22.2 - 23.3) | 0.96 (0.90 - 1.03) | | 0.286 |
| **Hypertension (complicated)** |  |  |  |  |  | |  |
| No (Reference) | 30,638 | 57.7% | 6,924 | 22.6 (22.1 - 23.1) | - | | . |
| Yes | 22,438 | 42.3% | 7,004 | 31.2 (30.6 - 31.8) | 1.05 (0.96 - 1.13) | | 0.288 |
| **Liver Disease** |  |  |  |  |  | |  |
| No (Reference) | 50,652 | 95.4% | 12,896 | 25.5 (25.1 - 25.8) | - | | . |
| Yes | 2,424 | 4.6% | 1,032 | 42.6 (40.6 - 44.6) | 1.44 (1.28 - 1.61) | | <.0001 |
| **Metastatic Cancer** |  |  |  |  |  | |  |
| No (Reference) | 52,371 | 98.7% | 13,702 | 26.2 (25.8 - 26.5) | - | | . |
| Yes | 705 | 1.3% | 226 | 32.1 (28.7 - 35.7) | 1.37 (1.07 - 1.75) | | 0.012 |
| **Other Neurological Disorders** |  |  |  |  |  | |  |
| No (Reference) | 40,183 | 75.7% | 9,006 | 22.4 (22.0 - 22.8) | - | | . |
| Yes | 12,893 | 24.3% | 4,922 | 38.2 (37.3 - 39.0) | 1.29 (1.22 - 1.37) | | <.0001 |
| **Paralysis** |  |  |  |  |  | |  |
| No (Reference) | 52,342 | 98.6% | 13,643 | 26.1 (25.7 - 26.4) | - | | . |
| Yes | 734 | 1.4% | 285 | 38.8 (35.3 - 42.5) | 0.88 (0.72 - 1.08) | | 0.232 |
| **Peptic Ulcer Disease** |  |  |  |  |  | |  |
| No (Reference) | 52,962 | 99.8% | 13,901 | 26.2 (25.9 - 26.6) | - | | . |
| Yes | 114 | 0.2% | 27 | 23.7 (16.4 - 32.7) | 0.38 (0.21 - 0.66) | | 0.001 |
| **Peripheral Vascular Disorder** |  |  |  |  |  | |  |
| No (Reference) | 48,611 | 91.6% | 12,618 | 26.0 (25.6 - 26.3) | - | | . |
| Yes | 4,465 | 8.4% | 1,310 | 29.3 (28.0 - 30.7) | 0.98 (0.90 - 1.08) | | 0.737 |
| **Pulmonary Circulation Disorder** |  |  |  |  |  | |  |
| No (Reference) | 51,337 | 96.7% | 13,348 | 26.0 (25.6 - 26.4) | - | | . |
| Yes | 1,739 | 3.3% | 580 | 33.4 (31.1 - 35.6) | 1.02 (0.88 - 1.18) | | 0.811 |
| **Renal Failure** |  |  |  |  |  | |  |
| No (Reference) | 43,054 | 81.1% | 10,588 | 24.6 (24.2 - 25.0) | - | | . |
| Yes | 10,022 | 18.9% | 3,340 | 33.3 (32.4 - 34.3) | 1.05 (0.96 - 1.14) | | 0.287 |
| **Solid Tumor** |  |  |  |  |  | |  |
| No (Reference) | 51,110 | 96.3% | 13,354 | 26.1 (25.7 - 26.5) | - | | . |
| Yes | 1,966 | 3.7% | 574 | 29.2 (27.2 - 31.3) | 1.01 (0.87 - 1.18) | | 0.866 |
| **Valvular Disease** |  |  |  |  |  | |  |
| No (Reference) | 48,913 | 92.2% | 12,677 | 25.9 (25.5 - 26.3) | - | | . |
| Yes | 4,163 | 7.8% | 1,251 | 30.1 (28.7 - 31.5) | 0.96 (0.87 - 1.05) | | 0.352 |
| **Weight Loss** |  |  |  |  |  | |  |
| No (Reference) | 47,247 | 89.0% | 11,665 | 24.7 (24.3 - 25.1) | - | | . |
| Yes | 5,829 | 11.0% | 2,263 | 38.8 (37.6 - 40.1) | 1.10 (1.02 - 1.19) | | 0.015 |
| ^a^ Patients with missing values for ADI, Urban/Rural, Hospital Size and Ownership were dropped from the multivariate analysis. | | | | | |  |  |
| ^b^ The South US census region includes counts for "Other" territories to avoid identification due to low counts. | | | |  |  | |  |
| ^c^ Patients have not had COVID-19 diagnosis recorded in the 30 days prior to admission. | | | |  |  | |  |
| **Notes:** Model evaluation metrics were ROC Area Under the Curve (0.8466); Pearson Chi Sq Goodness of Fit test p value (1); Deviance Goodness of Fit test p value (1). | | | | | | |  |
| Intravenous Immunoglobulin/Hyperimmune Globulin Group has been omitted to avoid identification due to low counts. | | | | | |  |  |

| **Supplementary Table 2.** Inpatient (IP) Mortality Rates and Corresponding Adjusted Odds Ratios (ORs) by Patient and Hospital Characteristics Among Recipients of COVID-19 Convalescent Plasma (CCP) in 2021 | | | | |  | |  |
| --- | --- | --- | --- | --- | --- | --- | --- |
| **Patient and Hospital Characteristics** | **CCP Transfusion Stays #** | **CCP Transfusion Stays  %** | **IP Mortality  #** | **Crude IP Mortality Rate per 100  (95% CIs)** | | **Adjusted ORs (95% CIs)** | **P-value** |
| **Overall** | 28,675 | 100.0% | 6,948 | 24.2 (23.7 - 24.7) | | - | . |
| **Age (continuous)** | 28,675 | 100.0% | 6,948 | 24.2 (23.7 - 24.7) | | 1.06 (1.05 - 1.06) | <.0001 |
| **Sex** |  |  |  |  | |  |  |
| Female (Reference) | 13,515 | 47.1% | 2,926 | 21.7 (21.0 - 22.4) | | - | . |
| Male | 15,160 | 52.9% | 4,022 | 26.5 (25.8 - 27.2) | | 1.19 (1.11 - 1.29) | <.0001 |
| **ADI^a^** |  |  |  |  | |  |  |
| 81 - 100 (Reference) | 4,399 | 15.3% | 1,123 | 25.5 (24.3 - 26.8) | | - | . |
| 61 – 80 | 6,156 | 21.5% | 1,503 | 24.4 (23.4 - 25.5) | | 1.06 (0.94 - 1.19) | 0.333 |
| 41 – 60 | 6,207 | 21.6% | 1,464 | 23.6 (22.5 - 24.7) | | 1.01 (0.90 - 1.13) | 0.883 |
| 21 – 40 | 5,653 | 19.7% | 1,319 | 23.3 (22.2 - 24.5) | | 0.86 (0.76 - 0.98) | 0.020 |
| 0 – 20 | 3,828 | 13.3% | 934 | 24.4 (23.1 - 25.8) | | 0.89 (0.77 - 1.02) | 0.084 |
| **Medicaid Eligibility** |  |  |  |  | |  |  |
| No (Reference) | 23,035 | 80.3% | 5,358 | 23.3 (22.7 - 23.8) | | - | . |
| Yes | 5,640 | 19.7% | 1,590 | 28.2 (27.0 - 29.4) | | 1.10 (0.99 - 1.21) | 0.063 |
| **Region of Residence^b^** |  |  |  |  | |  |  |
| South (Reference) | 17,739 | 61.9% | 4,213 | 23.7 (23.1 - 24.4) | | - | . |
| Midwest | 4,231 | 14.8% | 895 | 21.2 (19.9 - 22.4) | | 0.82 (0.73 - 0.92) | 0.001 |
| West | 3,525 | 12.3% | 1,016 | 28.8 (27.3 - 30.4) | | 1.13 (1.01 - 1.27) | 0.034 |
| Northeast | 3,180 | 11.1% | 824 | 25.9 (24.4 - 27.5) | | 1.26 (1.12 - 1.42) | 0.000 |
| **Urban/Rural^a^** |  |  |  |  | |  |  |
| Urban (Reference) | 20,305 | 70.8% | 4,965 | 24.5 (23.9 - 25.1) | | - | . |
| Rural | 6,163 | 21.5% | 1,438 | 23.3 (22.3 - 24.4) | | 1.03 (0.94 - 1.14) | 0.488 |
| **Immunocompromised Status** |  |  |  |  | |  |  |
| No (Reference) | 25,574 | 89.2% | 6,079 | 23.8 (23.3 - 24.3) | | - | . |
| Yes | 3,101 | 10.8% | 869 | 28.0 (26.5 - 29.6) | | 1.24 (1.08 - 1.42) | 0.002 |
| **Nursing Home Stay** |  |  |  |  | |  |  |
| No (Reference) | 26,897 | 93.8% | 6,404 | 23.8 (23.3 - 24.3) | | - | . |
| Yes | 1,778 | 6.2% | 544 | 30.6 (28.5 - 32.8) | | 1.21 (1.05 - 1.40) | 0.007 |
| **Emergency Room Stay** |  |  |  |  | |  |  |
| No (Reference) | 2,329 | 8.1% | 612 | 26.3 (24.5 - 28.1) | | - | . |
| Yes | 26,346 | 91.9% | 6,336 | 24.0 (23.5 - 24.6) | | 0.87 (0.76 - 1.00) | 0.049 |
| **COVID-19 As a Reason for Admission** |  |  |  |  | |  |  |
| No (Reference) | 13,576 | 47.3% | 3,470 | 25.6 (24.8 - 26.3) | | - | . |
| Yes | 15,099 | 52.7% | 3,478 | 23.0 (22.4 - 23.7) | | 1.02 (0.95 - 1.10) | 0.515 |
| **Time Since Most Recent COVID-19 Diagnosis in the 30 Days Prior to Admission** |  |  |  |  | |  |  |
| 0 days (Reference)^c^ | 14,619 | 51.0% | 3,526 | 24.1 (23.4 - 24.8) | | - | . |
| 1 - 3 days | 6,805 | 23.7% | 1,617 | 23.8 (22.8 - 24.8) | | 1.02 (0.93 - 1.12) | 0.668 |
| 4 - 10 days | 5,570 | 19.4% | 1,411 | 25.3 (24.2 - 26.5) | | 1.10 (1.00 - 1.21) | 0.060 |
| 11-30 days | 1,681 | 5.9% | 394 | 23.4 (21.4 - 25.6) | | 0.79 (0.67 - 0.93) | 0.004 |
| **Thrombosis on Index** |  |  |  |  | |  |  |
| No (Reference) | 24,518 | 85.5% | 5,257 | 21.4 (20.9 - 22.0) | | - | . |
| Yes | 4,157 | 14.5% | 1,691 | 40.7 (39.2 - 42.2) | | 1.35 (1.22 - 1.50) | <.0001 |
| **Hospital Ownership^a^** |  |  |  |  | |  |  |
| Government (Reference) | 3,944 | 13.8% | 914 | 23.2 (21.9 - 24.5) | | - | . |
| For Profit | 6,964 | 24.3% | 1,783 | 25.6 (24.6 - 26.6) | | 0.83 (0.74 - 0.94) | 0.003 |
| Non-Profit | 17,687 | 61.7% | 4,235 | 23.9 (23.3 - 24.6) | | 0.92 (0.82 - 1.02) | 0.129 |
| **Hospital Size^a^** |  |  |  |  | |  |  |
| Small (< 100 beds) (Reference) | 3,537 | 12.3% | 690 | 19.5 (18.2 - 20.9) | | - | . |
| Medium (100 - 500 beds) | 18,462 | 64.4% | 4,568 | 24.7 (24.1 - 25.4) | | 1.07 (0.95 - 1.21) | 0.276 |
| Large (> 500 beds) | 5,315 | 18.5% | 1,378 | 25.9 (24.8 - 27.1) | | 1.00 (0.87 - 1.16) | 0.947 |
| **ICU/CCU Stay** |  |  |  |  | |  |  |
| No (Reference) | 12,282 | 42.8% | 1,122 | 9.1 (8.6 - 9.7) | | - | . |
| Yes | 16,393 | 57.2% | 5,826 | 35.5 (34.8 - 36.3) | | 2.66 (2.44 - 2.90) | <.0001 |
| **Mechanical Ventilation** |  |  |  |  | |  |  |
| No (Reference) | 24,069 | 83.9% | 3,480 | 14.5 (14.0 - 14.9) | | - | . |
| Less Than 96hrs | 1,765 | 6.2% | 1,356 | 76.8 (74.8 - 78.8) | | 16.51 (14.38 - 18.95) | <.0001 |
| More Than 96hrs | 2,841 | 9.9% | 2,112 | 74.3 (72.7 - 75.9) | | 12.48 (11.15 - 13.97) | <.0001 |
| **Tocilizumab** |  |  |  |  | |  |  |
| No (Reference) | 27,228 | 95.0% | 6,350 | 23.3 (22.8 - 23.8) | | - | . |
| Yes | 1,447 | 5.0% | 598 | 41.3 (38.8 - 43.9) | | 1.72 (1.48 - 1.99) | <.0001 |
| **Baricitinib** |  |  |  |  | |  |  |
| No (Reference) | 27,976 | 97.6% | 6,726 | 24.0 (23.5 - 24.5) | | - | . |
| Yes | 699 | 2.4% | 222 | 31.8 (28.3 - 35.4) | | 1.53 (1.23 - 1.91) | 0.000 |
| **Remdesivir** |  |  |  |  | |  |  |
| No (Reference) | 6,440 | 22.5% | 1,751 | 27.2 (26.1 - 28.3) | | - | . |
| Yes | 22,235 | 77.5% | 5,197 | 23.4 (22.8 - 23.9) | | 0.87 (0.80 - 0.95) | 0.002 |
| **Charlson Comorbidity Index (CCI)** |  |  |  |  | |  |  |
| 0 (Reference) | 5,292 | 18.5% | 876 | 16.6 (15.6 - 17.6) | | - | . |
| 1 | 7,252 | 25.3% | 1,497 | 20.6 (19.7 - 21.6) | | 1.08 (0.95 - 1.23) | 0.230 |
| 2 | 5,653 | 19.7% | 1,452 | 25.7 (24.6 - 26.8) | | 1.27 (1.10 - 1.47) | 0.001 |
| 3 – 4 | 6,411 | 22.4% | 1,810 | 28.2 (27.1 - 29.4) | | 1.15 (0.97 - 1.36) | 0.097 |
| 5+ | 4,067 | 14.2% | 1,313 | 32.3 (30.9 - 33.7) | | 1.24 (1.00 - 1.54) | 0.051 |
| **90-Day Comorbidity History** |  |  |  |  | |  |  |
| **Thrombosis History** |  |  |  |  | |  |  |
| No (Reference) | 27,879 | 97.2% | 6,699 | 24.0 (23.5 - 24.5) | | - | . |
| Yes | 796 | 2.8% | 249 | 31.3 (28.1 - 34.6) | | 0.89 (0.72 - 1.09) | 0.263 |
| **ESRD History** |  |  |  |  | |  |  |
| No (Reference) | 27,836 | 97.1% | 6,698 | 24.1 (23.6 - 24.6) | | - | . |
| Yes | 839 | 2.9% | 250 | 29.8 (26.7 - 33.0) | | 0.83 (0.65 - 1.06) | 0.141 |
| **Elixhauser Comorbidity History** |  |  |  |  | |  |  |
| **Blood Loss Anemia** |  |  |  |  | |  |  |
| No (Reference) | 28,543 | 99.5% | 6,901 | 24.2 (23.7 - 24.7) | | - | . |
| Yes | 132 | 0.5% | 47 | 35.6 (27.6 - 44.5) | | 1.23 (0.78 - 1.95) | 0.379 |
| **Cardiac Arrhythmia** |  |  |  |  | |  |  |
| No (Reference) | 18,297 | 63.8% | 3,541 | 19.4 (18.8 - 19.9) | | - | . |
| Yes | 10,378 | 36.2% | 3,407 | 32.8 (31.9 - 33.7) | | 1.34 (1.25 - 1.45) | <.0001 |
| **Chronic Pulmonary Disease** |  |  |  |  | |  |  |
| No (Reference) | 19,825 | 69.1% | 4,736 | 23.9 (23.3 - 24.5) | | - | . |
| Yes | 8,850 | 30.9% | 2,212 | 25.0 (24.1 - 25.9) | | 1.07 (0.99 - 1.17) | 0.101 |
| **Coagulopathy** |  |  |  |  | |  |  |
| No (Reference) | 23,596 | 82.3% | 5,040 | 21.4 (20.8 - 21.9) | | - | . |
| Yes | 5,079 | 17.7% | 1,908 | 37.6 (36.2 - 38.9) | | 1.36 (1.25 - 1.49) | <.0001 |
| **Congestive Heart Failure** |  |  |  |  | |  |  |
| No (Reference) | 21,164 | 73.8% | 4,690 | 22.2 (21.6 - 22.7) | | - | . |
| Yes | 7,511 | 26.2% | 2,258 | 30.1 (29.0 - 31.1) | | 0.93 (0.83 - 1.03) | 0.156 |
| **Deficiency Anemia** |  |  |  |  | |  |  |
| No (Reference) | 27,286 | 95.2% | 6,563 | 24.1 (23.5 - 24.6) | | - | . |
| Yes | 1,389 | 4.8% | 385 | 27.7 (25.4 - 30.2) | | 0.96 (0.82 - 1.13) | 0.631 |
| **Depression** |  |  |  |  | |  |  |
| No (Reference) | 24,592 | 85.8% | 6,018 | 24.5 (23.9 - 25.0) | | - | . |
| Yes | 4,083 | 14.2% | 930 | 22.8 (21.5 - 24.1) | | 1.08 (0.97 - 1.19) | 0.168 |
| **Diabetes** |  |  |  |  | |  |  |
| No (Reference) | 16,409 | 57.2% | 3,765 | 22.9 (22.3 - 23.6) | | - | . |
| Yes | 12,266 | 42.8% | 3,183 | 25.9 (25.2 - 26.7) | | 0.96 (0.88 - 1.06) | 0.423 |
| **Fluid and Electrolyte Disorder** |  |  |  |  | |  |  |
| No (Reference) | 10,894 | 38.0% | 1,487 | 13.6 (13.0 - 14.3) | | - | . |
| Yes | 17,781 | 62.0% | 5,461 | 30.7 (30.0 - 31.4) | | 1.51 (1.39 - 1.64) | <.0001 |
| **Hypertension (uncomplicated)** |  |  |  |  | |  |  |
| No (Reference) | 15,667 | 54.6% | 4,220 | 26.9 (26.2 - 27.6) | | - | . |
| Yes | 13,008 | 45.4% | 2,728 | 21.0 (20.3 - 21.7) | | 0.94 (0.85 - 1.03) | 0.189 |
| **Hypertension (complicated)** |  |  |  |  | |  |  |
| No (Reference) | 17,054 | 59.5% | 3,515 | 20.6 (20.0 - 21.2) | | - | . |
| Yes | 11,621 | 40.5% | 3,433 | 29.5 (28.7 - 30.4) | | 1.11 (0.99 - 1.25) | 0.064 |
| **Liver Disease** |  |  |  |  | |  |  |
| No (Reference) | 27,271 | 95.1% | 6,394 | 23.4 (22.9 - 24.0) | | - | . |
| Yes | 1,404 | 4.9% | 554 | 39.5 (36.9 - 42.1) | | 1.42 (1.21 - 1.66) | <.0001 |
| **Obesity** |  |  |  |  | |  |  |
| No (Reference) | 21,492 | 75.0% | 5,100 | 23.7 (23.2 - 24.3) | | - | . |
| Yes | 7,183 | 25.0% | 1,848 | 25.7 (24.7 - 26.8) | | 1.06 (0.97 - 1.16) | 0.208 |
| **Other Neurological Disorders** |  |  |  |  | |  |  |
| No (Reference) | 22,356 | 78.0% | 4,681 | 20.9 (20.4 - 21.5) | | - | . |
| Yes | 6,319 | 22.0% | 2,267 | 35.9 (34.7 - 37.1) | | 1.34 (1.24 - 1.46) | <.0001 |
| **Paralysis** |  |  |  |  | |  |  |
| No (Reference) | 28,341 | 98.8% | 6,806 | 24.0 (23.5 - 24.5) | | - | . |
| Yes | 334 | 1.2% | 142 | 42.5 (37.2 - 48.0) | | 1.40 (1.03 - 1.90) | 0.033 |
| **Peripheral Vascular Disorder** |  |  |  |  | |  |  |
| No (Reference) | 26,351 | 91.9% | 6,309 | 23.9 (23.4 - 24.5) | | - | . |
| Yes | 2,324 | 8.1% | 639 | 27.5 (25.7 - 29.4) | | 0.97 (0.86 - 1.11) | 0.697 |
| **Pulmonary Circulation Disorder** |  |  |  |  | |  |  |
| No (Reference) | 27,606 | 96.3% | 6,586 | 23.9 (23.4 - 24.4) | | - | . |
| Yes | 1,069 | 3.7% | 362 | 33.9 (31.0 - 36.8) | | 1.11 (0.92 - 1.35) | 0.267 |
| **Renal Failure** |  |  |  |  | |  |  |
| No (Reference) | 23,866 | 83.2% | 5,413 | 22.7 (22.2 - 23.2) | | - | . |
| Yes | 4,809 | 16.8% | 1,535 | 31.9 (30.6 - 33.3) | | 1.10 (0.97 - 1.24) | 0.152 |
| **Rheumatoid Arthritis/Collagen Vascular Diseases (CVD)** |  |  |  |  | |  |  |
| No (Reference) | 27,360 | 95.4% | 6,613 | 24.2 (23.7 - 24.7) | | - | . |
| Yes | 1,315 | 4.6% | 335 | 25.5 (23.2 - 27.9) | | 1.26 (1.07 - 1.49) | 0.006 |
| **Valvular Disease** |  |  |  |  | |  |  |
| No (Reference) | 26,425 | 92.2% | 6,339 | 24.0 (23.5 - 24.5) | | - | . |
| Yes | 2,250 | 7.8% | 609 | 27.1 (25.2 - 29.0) | | 0.93 (0.82 - 1.06) | 0.263 |
| **Weight Loss** |  |  |  |  | |  |  |
| No (Reference) | 25,649 | 89.4% | 5,799 | 22.6 (22.1 - 23.1) | | - | . |
| Yes | 3,026 | 10.6% | 1,149 | 38.0 (36.2 - 39.7) | | 1.24 (1.11 - 1.38) | <.0001 |
| ^a^ Patients with missing values for ADI, Urban/Rural, Hospital Size and Ownership were dropped from the multivariate analysis. | | | | |  | |  |
| ^b^ The South US census region includes counts for "Other" territories to avoid identification due to low counts. | | | | | | | |
| ^c^ Patients have not had COVID-19 diagnosis recorded in the 30 days prior to admission. | | | | | | | |
| **Notes**: Model evaluation metrics were ROC Area Under the Curve (0.8506); Pearson Chi Sq Goodness of Fit test p value (1); Deviance Goodness of Fit test p value (1). | | | | | | |  |
| Intravenous Immunoglobulin/Hyperimmune Globulin Group has been omitted to avoid identification due to low counts. | | | | |  | |  |

| **Supplementary Table 3.** Inpatient (IP) Mortality Rates and Corresponding Adjusted Odds Ratios (ORs) by Patient and Hospital Characteristics Among Immunocompromised Recipients of COVID-19 Convalescent Plasma (CCP) | | | | | |  |
| --- | --- | --- | --- | --- | --- | --- |
| **Patient and Hospital Characteristics** | **CCP Transfusion Stays #** | **CCP Transfusion Stays  %** | **IP Mortality  #** | **Crude IP Mortality Rate per 100  (95% CIs)** | **Adjusted ORs (95% CIs)** | **P-value** |
| **Overall** | 8,147 | 100.0% | 2,304 | 28.3 (27.3 - 29.3) | - | . |
| **Age (continuous)** | 8,147 | 100.0% | 2,304 | 28.3 (27.3 - 29.3) | 1.06 (1.05 - 1.07) | <.0001 |
| **Sex** |  |  |  |  |  |  |
| Female (Reference) | 3,538 | 43.4% | 922 | 26.1 (24.6 - 27.5) | - | . |
| Male | 4,609 | 56.6% | 1,382 | 30.0 (28.7 - 31.3) | 1.13 (0.98 - 1.29) | 0.082 |
| **ADI^a^** |  |  |  |  |  |  |
| 81 - 100 (Reference) | 1,333 | 16.4% | 400 | 30.0 (27.6 - 32.6) | - | . |
| 61 – 80 | 1,638 | 20.1% | 460 | 28.1 (25.9 - 30.3) | 0.92 (0.75 - 1.13) | 0.409 |
| 41 – 60 | 1,825 | 22.4% | 492 | 27.0 (24.9 - 29.1) | 0.85 (0.69 - 1.05) | 0.138 |
| 21 – 40 | 1,633 | 20.0% | 436 | 26.7 (24.6 - 28.9) | 0.83 (0.66 - 1.03) | 0.086 |
| 0 – 20 | 1,064 | 13.1% | 315 | 29.6 (26.9 - 32.5) | 0.95 (0.74 - 1.21) | 0.672 |
| **Medicaid Eligibility** |  |  |  |  |  |  |
| No (Reference) | 6,272 | 77.0% | 1,730 | 27.6 (26.5 - 28.7) | - | . |
| Yes | 1,875 | 23.0% | 574 | 30.6 (28.5 - 32.8) | 1.03 (0.86 - 1.22) | 0.761 |
| **Region of Residence^b^** |  |  |  |  |  |  |
| South (Reference) | 4,169 | 51.2% | 1,141 | 27.4 (26.0 - 28.8) | - | . |
| Midwest | 1,933 | 23.7% | 539 | 27.9 (25.9 - 30.0) | 1.06 (0.89 - 1.26) | 0.505 |
| West | 1,206 | 14.8% | 384 | 31.8 (29.2 - 34.6) | 1.10 (0.90 - 1.34) | 0.336 |
| Northeast | 839 | 10.3% | 240 | 28.6 (25.6 - 31.8) | 1.24 (0.98 - 1.56) | 0.078 |
| **Urban/Rural^a^** |  |  |  |  |  |  |
| Urban (Reference) | 5,839 | 71.7% | 1,632 | 27.9 (26.8 - 29.1) | - | . |
| Rural | 1,720 | 21.1% | 488 | 28.4 (26.3 - 30.6) | 1.09 (0.91 - 1.29) | 0.354 |
| **Nursing Home Stay** |  |  |  |  |  |  |
| No (Reference) | 7,400 | 90.8% | 2,050 | 27.7 (26.7 - 28.7) | - | . |
| Yes | 747 | 9.2% | 254 | 34.0 (30.6 - 37.5) | 1.26 (1.01 - 1.57) | 0.040 |
| **Emergency Room Stay** |  |  |  |  |  |  |
| No (Reference) | 969 | 11.9% | 316 | 32.6 (29.7 - 35.7) | - | . |
| Yes | 7,178 | 88.1% | 1,988 | 27.7 (26.7 - 28.7) | 0.91 (0.73 - 1.13) | 0.371 |
| **COVID-19 As a Reason for Admission** |  |  |  |  |  |  |
| No (Reference) | 4,219 | 51.8% | 1,214 | 28.8 (27.4 - 30.2) | - | . |
| Yes | 3,928 | 48.2% | 1,090 | 27.8 (26.4 - 29.2) | 1.10 (0.96 - 1.25) | 0.169 |
| **Time Since Most Recent COVID-19 Diagnosis in the 30 Days Prior to Admission** |  |  |  |  |  |  |
| 0 days (Reference)^c^ | 3,965 | 48.7% | 1,135 | 28.6 (27.2 - 30.1) | - | . |
| 1 - 3 days | 1,968 | 24.2% | 559 | 28.4 (26.4 - 30.5) | 1.03 (0.88 - 1.21) | 0.733 |
| 4 - 10 days | 1,457 | 17.9% | 420 | 28.8 (26.5 - 31.2) | 1.06 (0.88 - 1.26) | 0.549 |
| 11-30 days | 757 | 9.3% | 190 | 25.1 (22.1 - 28.4) | 0.78 (0.61 - 1.00) | 0.047 |
| **Thrombosis on Index** |  |  |  |  |  |  |
| No (Reference) | 6,917 | 84.9% | 1,759 | 25.4 (24.4 - 26.5) | - | . |
| Yes | 1,230 | 15.1% | 545 | 44.3 (41.5 - 47.1) | 1.33 (1.11 - 1.59) | 0.002 |
| **Hospital Ownership^a^** |  |  |  |  |  |  |
| Government (Reference) | 985 | 12.1% | 255 | 25.9 (23.2 - 28.8) | - | . |
| For Profit | 1,589 | 19.5% | 489 | 30.8 (28.5 - 33.1) | 1.04 (0.82 - 1.33) | 0.732 |
| Non-Profit | 5,497 | 67.5% | 1,543 | 28.1 (26.9 - 29.3) | 1.02 (0.82 - 1.26) | 0.859 |
| **Hospital Size^a^** |  |  |  |  |  |  |
| Small (< 100 beds) (Reference) | 616 | 7.6% | 141 | 22.9 (19.7 - 26.5) | - | . |
| Medium (100 - 500 beds) | 5,101 | 62.6% | 1,460 | 28.6 (27.4 - 29.9) | 1.03 (0.80 - 1.34) | 0.807 |
| Large (> 500 beds) | 2,014 | 24.7% | 604 | 30.0 (28.0 - 32.1) | 0.97 (0.73 - 1.29) | 0.825 |
| **ICU/CCU Stay** |  |  |  |  |  |  |
| No (Reference) | 3,152 | 38.7% | 332 | 10.5 (9.5 - 11.7) | - | . |
| Yes | 4,995 | 61.3% | 1,972 | 39.5 (38.1 - 40.9) | 2.79 (2.38 - 3.26) | <.0001 |
| **Mechanical Ventilation** |  |  |  |  |  |  |
| No (Reference) | 6,675 | 81.9% | 1,132 | 17.0 (16.1 - 17.9) | - | . |
| Less Than 96hrs | 648 | 8.0% | 520 | 80.2 (76.9 - 83.2) | 16.44 (13.03 - 20.74) | <.0001 |
| More Than 96hrs | 824 | 10.1% | 652 | 79.1 (76.2 - 81.8) | 13.33 (10.78 - 16.47) | <.0001 |
| **Intravenous Immunoglobulin/Hyperimmune Globulin** |  |  |  |  |  |  |
| No (Reference) | 8,109 | 99.5% | 2,293 | 28.3 (27.3 - 29.3) | - | . |
| Yes | 38 | 0.5% | 11 | 28.9 (16.0 - 46.1) | 1.08 (0.42 - 2.77) | 0.870 |
| **Tocilizumab** |  |  |  |  |  |  |
| No (Reference) | 7,899 | 97.0% | 2,182 | 27.6 (26.6 - 28.6) | - | . |
| Yes | 248 | 3.0% | 122 | 49.2 (42.8 - 55.6) | 2.19 (1.55 - 3.10) | <.0001 |
| **Baricitinib** |  |  |  |  |  |  |
| No (Reference) | 8,081 | 99.2% | 2,276 | 28.2 (27.2 - 29.2) | - | . |
| Yes | 66 | 0.8% | 28 | 42.4 (30.6 - 55.2) | 1.88 (0.97 - 3.63) | 0.061 |
| **Remdesivir** |  |  |  |  |  |  |
| No (Reference) | 3,052 | 37.5% | 893 | 29.3 (27.7 - 30.9) | - | . |
| Yes | 5,095 | 62.5% | 1,411 | 27.7 (26.5 - 28.9) | 1.03 (0.89 - 1.20) | 0.665 |
| **Charlson Comorbidity Index (CCI)** |  |  |  |  |  |  |
| 0 (Reference) | 324 | 4.0% | 67 | 20.7 (16.5 - 25.6) | - | . |
| 1 | 589 | 7.2% | 129 | 21.9 (18.7 - 25.5) | 0.75 (0.49 - 1.16) | 0.198 |
| 2 | 1,347 | 16.5% | 322 | 23.9 (21.7 - 26.3) | 0.96 (0.65 - 1.41) | 0.841 |
| 3 – 4 | 2,482 | 30.5% | 719 | 29.0 (27.2 - 30.8) | 1.03 (0.70 - 1.50) | 0.892 |
| 5+ | 3,405 | 41.8% | 1,067 | 31.3 (29.8 - 32.9) | 0.98 (0.65 - 1.48) | 0.922 |
| **90-Day Comorbidity History** |  |  |  |  |  |  |
| **Transplant Dialysis^d^** |  |  |  |  |  |  |
| No (Reference) | 4,650 | 57.1% | 1,273 | 27.4 (26.1 - 28.7) | - | . |
| Yes | 3,497 | 42.9% | 1,031 | 29.5 (28.0 - 31.0) | 0.96 (0.79 - 1.17) | 0.680 |
| **Radiation^d^** |  |  |  |  |  |  |
| No (Reference) | 7,664 | 94.1% | 2,144 | 28.0 (27.0 - 29.0) | - | . |
| Yes | 483 | 5.9% | 160 | 33.1 (29.0 - 37.6) | 1.43 (1.09 - 1.88) | 0.009 |
| **Thrombosis History** |  |  |  |  |  |  |
| No (Reference) | 7,712 | 94.7% | 2,149 | 27.9 (26.9 - 28.9) | - | . |
| Yes | 435 | 5.3% | 155 | 35.6 (31.2 - 40.4) | 0.96 (0.72 - 1.27) | 0.758 |
| **Elixhauser Comorbidity History** |  |  |  |  |  |  |
| **Cardiac Arrhythmia** |  |  |  |  |  |  |
| No (Reference) | 4,842 | 59.4% | 1,115 | 23.0 (21.9 - 24.2) | - | . |
| Yes | 3,305 | 40.6% | 1,189 | 36.0 (34.3 - 37.6) | 1.27 (1.11 - 1.46) | 0.000 |
| **Coagulopathy** |  |  |  |  |  |  |
| No (Reference) | 5,988 | 73.5% | 1,504 | 25.1 (24.0 - 26.2) | - | . |
| Yes | 2,159 | 26.5% | 800 | 37.1 (35.0 - 39.1) | 1.22 (1.06 - 1.41) | 0.006 |
| **Congestive Heart Failure** |  |  |  |  |  |  |
| No (Reference) | 5,101 | 62.6% | 1,317 | 25.8 (24.6 - 27.0) | - | . |
| Yes | 3,046 | 37.4% | 987 | 32.4 (30.7 - 34.1) | 1.08 (0.91 - 1.27) | 0.375 |
| **Diabetes** |  |  |  |  |  |  |
| No (Reference) | 3,967 | 48.7% | 1,059 | 26.7 (25.3 - 28.1) | - | . |
| Yes | 4,180 | 51.3% | 1,245 | 29.8 (28.4 - 31.2) | 0.94 (0.80 - 1.11) | 0.481 |
| **Fluid and Electrolyte Disorder** |  |  |  |  |  |  |
| No (Reference) | 2,632 | 32.3% | 477 | 18.1 (16.7 - 19.7) | - | . |
| Yes | 5,515 | 67.7% | 1,827 | 33.1 (31.9 - 34.4) | 1.32 (1.13 - 1.53) | 0.000 |
| **Hypertension (uncomplicated)** |  |  |  |  |  |  |
| No (Reference) | 5,738 | 70.4% | 1,692 | 29.5 (28.3 - 30.7) | - | . |
| Yes | 2,409 | 29.6% | 612 | 25.4 (23.7 - 27.2) | 0.97 (0.81 - 1.17) | 0.760 |
| **Hypertension (complicated)** |  |  |  |  |  |  |
| No (Reference) | 3,197 | 39.2% | 786 | 24.6 (23.1 - 26.1) | - | . |
| Yes | 4,950 | 60.8% | 1,518 | 30.7 (29.4 - 32.0) | 1.02 (0.83 - 1.25) | 0.880 |
| **Liver Disease** |  |  |  |  |  |  |
| No (Reference) | 7,534 | 92.5% | 2,040 | 27.1 (26.1 - 28.1) | - | . |
| Yes | 613 | 7.5% | 264 | 43.1 (39.1 - 47.1) | 1.52 (1.19 - 1.93) | 0.001 |
| **Metastatic Cancer** |  |  |  |  |  |  |
| No (Reference) | 7,537 | 92.5% | 2,109 | 28.0 (27.0 - 29.0) | - | . |
| Yes | 610 | 7.5% | 195 | 32.0 (28.3 - 35.9) | 1.36 (1.02 - 1.82) | 0.035 |
| **Other Neurological Disorders** |  |  |  |  |  |  |
| No (Reference) | 6,095 | 74.8% | 1,452 | 23.8 (22.8 - 24.9) | - | . |
| Yes | 2,052 | 25.2% | 852 | 41.5 (39.4 - 43.7) | 1.45 (1.25 - 1.68) | <.0001 |
| **Paralysis** |  |  |  |  |  |  |
| No (Reference) | 8,031 | 98.6% | 2,259 | 28.1 (27.1 - 29.1) | - | . |
| Yes | 116 | 1.4% | 45 | 38.8 (30.0 - 48.3) | 0.79 (0.46 - 1.33) | 0.370 |
| **Pulmonary Circulation Disorder** |  |  |  |  |  |  |
| No (Reference) | 7,903 | 97.0% | 2,198 | 27.8 (26.8 - 28.8) | - | . |
| Yes | 244 | 3.0% | 106 | 43.4 (37.2 - 49.9) | 1.56 (1.08 - 2.25) | 0.018 |
| **Renal Failure** |  |  |  |  |  |  |
| No (Reference) | 4,267 | 52.4% | 1,124 | 26.3 (25.0 - 27.7) | - | . |
| Yes | 3,880 | 47.6% | 1,180 | 30.4 (29.0 - 31.9) | 1.10 (0.88 - 1.37) | 0.389 |
| **Rheumatoid Arthritis/Collagen Vascular Diseases (CVD)** |  |  |  |  |  |  |
| No (Reference) | 7,510 | 92.2% | 2,123 | 28.3 (27.3 - 29.3) | - | . |
| Yes | 637 | 7.8% | 181 | 28.4 (25.0 - 32.1) | 1.37 (1.07 - 1.75) | 0.013 |
| **Valvular Disease** |  |  |  |  |  |  |
| No (Reference) | 7,261 | 89.1% | 2,028 | 27.9 (26.9 - 29.0) | - | . |
| Yes | 886 | 10.9% | 276 | 31.2 (28.1 - 34.3) | 0.91 (0.74 - 1.12) | 0.363 |
| **Weight Loss** |  |  |  |  |  |  |
| No (Reference) | 6,839 | 84.0% | 1,799 | 26.3 (25.3 - 27.4) | - | . |
| Yes | 1,308 | 16.1% | 505 | 38.6 (36.0 - 41.3) | 1.17 (0.98 - 1.38) | 0.077 |
| ^a^ Patients with missing values for ADI, Urban/Rural, Hospital Size and Ownership were dropped from the multivariate analysis. | | | | |  |  |
| ^b^ The South US census region includes counts for "Other" territories to avoid identification due to low counts. | | | | | | |
| ^c^ Patients have not had COVID-19 diagnosis recorded in the 30 days prior to admission. | | | | | | |
| ^d^ Analysis evaluates specific IC conditions and immune suppressing treatment groups selected in the model; Overall, IC CCP recipients include beneficiaries with Myelodysplastic Syndromes, Myeloproliferative disorders, Immunoproliferative diseases, other related conditions and/or immune suppressing treatments listed in Supplementary Appendix Table. | | | | | | |
| **Notes:** Model evaluation metrics were ROC Area Under the Curve (0.8456); Pearson Chi Sq Goodness of Fit test p value (0.9359); Deviance Goodness of Fit test p value (1). | | | | | |  |

| **Supplementary Table 4.** Inpatient (IP) Mortality Rates and Corresponding Adjusted Odds Ratios (ORs) by Patient and Hospital Characteristics Among Non-Immunocompromised Recipients of COVID-19 Convalescent Plasma (CCP) | | | | | |  |
| --- | --- | --- | --- | --- | --- | --- |
| **Patient and Hospital Characteristics** | **CCP Transfusion Stays #** | **CCP Transfusion Stays  %** | **IP Mortality  #** | **Crude IP Mortality Rate per 100  (95% CIs)** | **Adjusted ORs (95% CIs)** | **P-value** |
| **Overall** | 73,604 | 100.0% | 18,572 | 25.2 (24.9 - 25.5) | - | . |
| **Age (continuous)** | 73,604 | 100.0% | 18,572 | 25.2 (24.9 - 25.5) | 1.06 (1.06 - 1.07) | <.0001 |
| **Sex** |  |  |  |  |  |  |
| Female (Reference) | 33,800 | 45.9% | 7,557 | 22.4 (21.9 - 22.8) | - | . |
| Male | 39,804 | 54.1% | 11,015 | 27.7 (27.2 - 28.1) | 1.21 (1.16 - 1.27) | <.0001 |
| **ADI^a^** |  |  |  |  |  |  |
| 81 - 100 (Reference) | 11,097 | 15.1% | 3,011 | 27.1 (26.3 - 28.0) | - | . |
| 61 – 80 | 15,945 | 21.7% | 4,067 | 25.5 (24.8 - 26.2) | 1.01 (0.95 - 1.09) | 0.688 |
| 41 – 60 | 16,389 | 22.3% | 3,975 | 24.3 (23.6 - 24.9) | 0.96 (0.89 - 1.03) | 0.254 |
| 21 – 40 | 14,410 | 19.6% | 3,488 | 24.2 (23.5 - 24.9) | 0.87 (0.81 - 0.94) | 0.000 |
| 0 – 20 | 8,987 | 12.2% | 2,202 | 24.5 (23.6 - 25.4) | 0.84 (0.77 - 0.91) | <.0001 |
| **Medicaid Eligibility** |  |  |  |  |  |  |
| No (Reference) | 58,642 | 79.7% | 14,163 | 24.2 (23.8 - 24.5) | - | . |
| Yes | 14,962 | 20.3% | 4,409 | 29.5 (28.7 - 30.2) | 1.14 (1.08 - 1.22) | <.0001 |
| **Region of Residence^b^** |  |  |  |  |  |  |
| South (Reference) | 39,999 | 54.3% | 9,913 | 24.8 (24.4 - 25.2) | - | . |
| Midwest | 17,509 | 23.8% | 4,113 | 23.5 (22.9 - 24.1) | 0.98 (0.92 - 1.04) | 0.494 |
| West | 9,290 | 12.6% | 2,749 | 29.6 (28.7 - 30.5) | 1.18 (1.10 - 1.27) | <.0001 |
| Northeast | 6,806 | 9.2% | 1,797 | 26.4 (25.4 - 27.5) | 1.18 (1.09 - 1.28) | <.0001 |
| **Urban/Rural^a^** |  |  |  |  |  |  |
| Urban (Reference) | 49,857 | 67.7% | 12,578 | 25.2 (24.8 - 25.6) | - | . |
| Rural | 17,579 | 23.9% | 4,336 | 24.7 (24.0 - 25.3) | 1.12 (1.06 - 1.19) | <.0001 |
| **Nursing Home Stay** |  |  |  |  |  |  |
| No (Reference) | 67,113 | 91.2% | 16,497 | 24.6 (24.3 - 24.9) | - | . |
| Yes | 6,491 | 8.8% | 2,075 | 32.0 (30.8 - 33.1) | 1.18 (1.09 - 1.28) | <.0001 |
| **Emergency Room Stay** |  |  |  |  |  |  |
| No (Reference) | 7,836 | 10.6% | 2,136 | 27.3 (26.3 - 28.3) | - | . |
| Yes | 65,768 | 89.4% | 16,436 | 25.0 (24.7 - 25.3) | 0.95 (0.88 - 1.03) | 0.187 |
| **COVID-19 As a Reason for Admission** |  |  |  |  |  |  |
| No (Reference) | 34,898 | 47.4% | 9,394 | 26.9 (26.5 - 27.4) | - | . |
| Yes | 38,706 | 52.6% | 9,178 | 23.7 (23.3 - 24.1) | 1.03 (0.98 - 1.07) | 0.253 |
| **Time Since Most Recent COVID-19 Diagnosis in the 30 Days Prior to Admission** |  |  |  |  |  |  |
| 0 days (Reference)^c^ | 37,017 | 50.3% | 9,369 | 25.3 (24.9 - 25.8) | - | . |
| 1 - 3 days | 17,429 | 23.7% | 4,232 | 24.3 (23.6 - 24.9) | 0.97 (0.92 - 1.03) | 0.348 |
| 4 - 10 days | 15,376 | 20.9% | 4,053 | 26.4 (25.7 - 27.1) | 1.08 (1.02 - 1.14) | 0.011 |
| 11-30 days | 3,782 | 5.1% | 918 | 24.3 (22.9 - 25.7) | 0.84 (0.76 - 0.93) | 0.001 |
| **Thrombosis on Index** |  |  |  |  |  |  |
| No (Reference) | 63,305 | 86.0% | 14,301 | 22.6 (22.3 - 22.9) | - | . |
| Yes | 10,299 | 14.0% | 4,271 | 41.5 (40.5 - 42.4) | 1.35 (1.26 - 1.44) | <.0001 |
| **Hospital Ownership^a^** |  |  |  |  |  |  |
| Government (Reference) | 9,476 | 12.9% | 2,398 | 25.3 (24.4 - 26.2) | - | . |
| For Profit | 14,909 | 20.3% | 3,865 | 25.9 (25.2 - 26.6) | 0.76 (0.71 - 0.83) | <.0001 |
| Non-Profit | 49,143 | 66.8% | 12,294 | 25.0 (24.6 - 25.4) | 0.84 (0.79 - 0.90) | <.0001 |
| **Hospital Size^a^** |  |  |  |  |  |  |
| Small (< 100 beds) (Reference) | 9,064 | 12.3% | 1,754 | 19.4 (18.5 - 20.2) | - | . |
| Medium (100 - 500 beds) | 47,404 | 64.4% | 12,280 | 25.9 (25.5 - 26.3) | 1.15 (1.07 - 1.24) | 0.000 |
| Large (> 500 beds) | 13,624 | 18.5% | 3,673 | 27.0 (26.2 - 27.7) | 1.11 (1.02 - 1.21) | 0.021 |
| **ICU/CCU Stay** |  |  |  |  |  |  |
| No (Reference) | 30,201 | 41.0% | 2,812 | 9.3 (9.0 - 9.6) | - | . |
| Yes | 43,403 | 59.0% | 15,760 | 36.3 (35.9 - 36.8) | 2.70 (2.56 - 2.85) | <.0001 |
| **Mechanical Ventilation** |  |  |  |  |  |  |
| No (Reference) | 60,809 | 82.6% | 9,198 | 15.1 (14.8 - 15.4) | - | . |
| Less Than 96hrs | 4,183 | 5.7% | 3,079 | 73.6 (72.2 - 74.9) | 12.77 (11.73 - 13.90) | <.0001 |
| More Than 96hrs | 8,612 | 11.7% | 6,295 | 73.1 (72.1 - 74.0) | 11.44 (10.71 - 12.22) | <.0001 |
| **Intravenous Immunoglobulin/Hyperimmune Globulin** |  |  |  |  |  |  |
| No (Reference) | 73,575 | 100.0% | 18,560 | 25.2 (24.9 - 25.5) | - | . |
| Yes | 29 | 0.0% | 12 | 41.4 (24.1 - 60.9) | 1.44 (0.51 - 4.13) | 0.492 |
| **Tocilizumab** |  |  |  |  |  |  |
| No (Reference) | 71,087 | 96.6% | 17,539 | 24.7 (24.4 - 25.0) | - | . |
| Yes | 2,517 | 3.4% | 1,033 | 41.0 (39.1 - 43.0) | 1.60 (1.44 - 1.79) | <.0001 |
| **Baricitinib** |  |  |  |  |  |  |
| No (Reference) | 72,888 | 99.0% | 18,346 | 25.2 (24.9 - 25.5) | - | . |
| Yes | 716 | 1.0% | 226 | 31.6 (28.2 - 35.1) | 1.46 (1.18 - 1.80) | 0.000 |
| **Remdesivir** |  |  |  |  |  |  |
| No (Reference) | 16,861 | 22.9% | 4,538 | 26.9 (26.2 - 27.6) | - | . |
| Yes | 56,743 | 77.1% | 14,034 | 24.7 (24.4 - 25.1) | 0.93 (0.88 - 0.98) | 0.010 |
| **Charlson Comorbidity Index (CCI)** |  |  |  |  |  |  |
| 0 (Reference) | 13,954 | 19.0% | 2,458 | 17.6 (17.0 - 18.3) | - | . |
| 1 | 19,914 | 27.1% | 4,292 | 21.6 (21.0 - 22.1) | 1.12 (1.04 - 1.21) | 0.004 |
| 2 | 14,593 | 19.8% | 3,851 | 26.4 (25.7 - 27.1) | 1.25 (1.14 - 1.37) | <.0001 |
| 3 – 4 | 16,293 | 22.1% | 4,917 | 30.2 (29.5 - 30.9) | 1.24 (1.12 - 1.39) | <.0001 |
| 5+ | 8,850 | 12.0% | 3,054 | 34.5 (33.5 - 35.5) | 1.37 (1.19 - 1.58) | <.0001 |
| **90-Day Comorbidity History** |  |  |  |  |  |  |
| **Thrombosis History** |  |  |  |  |  |  |
| No (Reference) | 71,722 | 97.4% | 17,942 | 25.0 (24.7 - 25.3) | - | . |
| Yes | 1,882 | 2.6% | 630 | 33.5 (31.4 - 35.7) | 1.00 (0.88 - 1.15) | 0.958 |
| **Elixhauser Comorbidity History** |  |  |  |  |  |  |
| **Alcohol Abuse** |  |  |  |  |  |  |
| No (Reference) | 72,589 | 98.6% | 18,277 | 25.2 (24.9 - 25.5) | - | . |
| Yes | 1,015 | 1.4% | 295 | 29.1 (26.3 - 32.0) | 0.93 (0.77 - 1.12) | 0.449 |
| **Blood Loss Anemia** |  |  |  |  |  |  |
| No (Reference) | 73,294 | 99.6% | 18,464 | 25.2 (24.9 - 25.5) | - | . |
| Yes | 310 | 0.4% | 108 | 34.8 (29.6 - 40.5) | 1.14 (0.84 - 1.56) | 0.390 |
| **Cardiac Arrhythmia** |  |  |  |  |  |  |
| No (Reference) | 46,722 | 63.5% | 9,427 | 20.2 (19.8 - 20.5) | - | . |
| Yes | 26,882 | 36.5% | 9,145 | 34.0 (33.5 - 34.6) | 1.36 (1.29 - 1.42) | <.0001 |
| **Chronic Pulmonary Disease** |  |  |  |  |  |  |
| No (Reference) | 50,280 | 68.3% | 12,540 | 24.9 (24.6 - 25.3) | - | . |
| Yes | 23,324 | 31.7% | 6,032 | 25.9 (25.3 - 26.4) | 1.00 (0.95 - 1.06) | 0.980 |
| **Coagulopathy** |  |  |  |  |  |  |
| No (Reference) | 61,492 | 83.5% | 13,931 | 22.7 (22.3 - 23.0) | - | . |
| Yes | 12,112 | 16.5% | 4,641 | 38.3 (37.5 - 39.2) | 1.31 (1.24 - 1.38) | <.0001 |
| **Congestive Heart Failure** |  |  |  |  |  |  |
| No (Reference) | 54,077 | 73.5% | 12,392 | 22.9 (22.6 - 23.3) | - | . |
| Yes | 19,527 | 26.5% | 6,180 | 31.6 (31.0 - 32.3) | 1.00 (0.94 - 1.07) | 0.894 |
| **Deficiency Anemia** |  |  |  |  |  |  |
| No (Reference) | 70,340 | 95.6% | 17,660 | 25.1 (24.8 - 25.4) | - | . |
| Yes | 3,264 | 4.4% | 912 | 27.9 (26.4 - 29.5) | 0.98 (0.89 - 1.09) | 0.746 |
| **Depression** |  |  |  |  |  |  |
| No (Reference) | 62,472 | 84.9% | 15,924 | 25.5 (25.1 - 25.8) | - | . |
| Yes | 11,132 | 15.1% | 2,648 | 23.8 (23.0 - 24.6) | 1.03 (0.97 - 1.09) | 0.385 |
| **Diabetes** |  |  |  |  |  |  |
| No (Reference) | 42,055 | 57.1% | 10,009 | 23.8 (23.4 - 24.2) | - | . |
| Yes | 31,549 | 42.9% | 8,563 | 27.1 (26.7 - 27.6) | 0.95 (0.90 - 1.01) | 0.130 |
| **Fluid and Electrolyte Disorder** |  |  |  |  |  |  |
| No (Reference) | 28,270 | 38.4% | 3,937 | 13.9 (13.5 - 14.3) | - | . |
| Yes | 45,334 | 61.6% | 14,635 | 32.3 (31.9 - 32.7) | 1.58 (1.51 - 1.67) | <.0001 |
| **Hypertension (uncomplicated)** |  |  |  |  |  |  |
| No (Reference) | 39,602 | 53.8% | 11,124 | 28.1 (27.6 - 28.5) | - | . |
| Yes | 34,002 | 46.2% | 7,448 | 21.9 (21.5 - 22.3) | 0.95 (0.90 - 1.01) | 0.101 |
| **Hypertension (complicated)** |  |  |  |  |  |  |
| No (Reference) | 44,495 | 60.5% | 9,653 | 21.7 (21.3 - 22.1) | - | . |
| Yes | 29,109 | 39.5% | 8,919 | 30.6 (30.1 - 31.2) | 1.08 (1.00 - 1.15) | 0.041 |
| **Liver Disease** |  |  |  |  |  |  |
| No (Reference) | 70,389 | 95.6% | 17,250 | 24.5 (24.2 - 24.8) | - | . |
| Yes | 3,215 | 4.4% | 1,322 | 41.1 (39.4 - 42.8) | 1.42 (1.28 - 1.57) | <.0001 |
| **Obesity** |  |  |  |  |  |  |
| No (Reference) | 54,921 | 74.6% | 13,713 | 25.0 (24.6 - 25.3) | - | . |
| Yes | 18,683 | 25.4% | 4,859 | 26.0 (25.4 - 26.6) | 1.04 (0.98 - 1.10) | 0.174 |
| **Other Neurological Disorders** |  |  |  |  |  |  |
| No (Reference) | 56,444 | 76.7% | 12,235 | 21.7 (21.3 - 22.0) | - | . |
| Yes | 17,160 | 23.3% | 6,337 | 36.9 (36.2 - 37.7) | 1.29 (1.23 - 1.36) | <.0001 |
| **Paralysis** |  |  |  |  |  |  |
| No (Reference) | 72,652 | 98.7% | 18,190 | 25.0 (24.7 - 25.4) | - | . |
| Yes | 952 | 1.3% | 382 | 40.1 (37.0 - 43.3) | 1.04 (0.87 - 1.25) | 0.673 |
| **Peripheral Vascular Disorder** |  |  |  |  |  |  |
| No (Reference) | 67,702 | 92.0% | 16,889 | 24.9 (24.6 - 25.3) | - | . |
| Yes | 5,902 | 8.0% | 1,683 | 28.5 (27.4 - 29.7) | 0.98 (0.90 - 1.06) | 0.621 |
| **Pulmonary Circulation Disorder** |  |  |  |  |  |  |
| No (Reference) | 71,040 | 96.5% | 17,736 | 25.0 (24.7 - 25.3) | - | . |
| Yes | 2,564 | 3.5% | 836 | 32.6 (30.8 - 34.5) | 1.00 (0.89 - 1.14) | 0.938 |
| **Renal Failure** |  |  |  |  |  |  |
| No (Reference) | 62,653 | 85.1% | 14,877 | 23.7 (23.4 - 24.1) | - | . |
| Yes | 10,951 | 14.9% | 3,695 | 33.7 (32.9 - 34.6) | 1.06 (0.98 - 1.15) | 0.127 |
| **Rheumatoid Arthritis/Collagen Vascular Diseases (CVD)** |  |  |  |  |  |  |
| No (Reference) | 70,440 | 95.7% | 17,750 | 25.2 (24.9 - 25.5) | - | . |
| Yes | 3,164 | 4.3% | 822 | 26.0 (24.5 - 27.6) | 1.14 (1.02 - 1.27) | 0.017 |
| **Solid Tumor** |  |  |  |  |  |  |
| No (Reference) | 71,695 | 97.4% | 18,050 | 25.2 (24.9 - 25.5) | - | . |
| Yes | 1,909 | 2.6% | 522 | 27.3 (25.4 - 29.4) | 1.04 (0.91 - 1.20) | 0.571 |
| **Valvular Disease** |  |  |  |  |  |  |
| No (Reference) | 68,077 | 92.5% | 16,988 | 25.0 (24.6 - 25.3) | - | . |
| Yes | 5,527 | 7.5% | 1,584 | 28.7 (27.5 - 29.9) | 0.95 (0.88 - 1.03) | 0.239 |
| **Weight Loss** |  |  |  |  |  |  |
| No (Reference) | 66,057 | 89.7% | 15,665 | 23.7 (23.4 - 24.0) | - | . |
| Yes | 7,547 | 10.3% | 2,907 | 38.5 (37.4 - 39.6) | 1.14 (1.06 - 1.22) | 0.000 |
| ^a^ Patients with missing values for ADI, Urban/Rural, Hospital Size and Ownership were dropped from the multivariate analysis. | | | | | | |
| ^b^ The South US census region includes counts for "Other" territories to avoid identification due to low counts. | | | | | | |
| ^c^ Patients have not had COVID-19 diagnosis recorded in the 30 days prior to admission. | | | | | | |
| **Notes:** Model evaluation metrics were ROC Area Under the Curve (0.8481); Pearson Chi Sq Goodness of Fit test p value (1); Deviance Goodness of Fit test p value (1). | | | | | |  |
| Peptic Ulcer Disease Group has been omitted to avoid identification due to low counts. | | | | |  |  |

| **Supplementary Table 5.** Inpatient (IP) Mortality Rates and Corresponding Adjusted Odds Ratios (ORs) by Patient and Hospital Characteristics Among Recipients of COVID-19 Convalescent Plasma (CCP) Residing in the South US Census Region | | | | | |  |
| --- | --- | --- | --- | --- | --- | --- |
| **Patient and Hospital Characteristics** | **CCP Transfusion Stays #** | **CCP Transfusion Stays  %** | **IP Mortality  #** | **Crude IP Mortality Rate per 100  (95% CIs)** | **Adjusted ORs (95% CIs)** | **P-value** |
| **Overall** | 44,168 | 100.0% | 11,054 | 25.0 (24.6 - 25.4) | - | . |
| **Age (continuous)** | 44,168 | 100.0% | 11,054 | 25.0 (24.6 - 25.4) | 1.06 (1.06 - 1.06) | <.0001 |
| **Sex** |  |  |  |  |  |  |
| Female (Reference) | 20,694 | 46.9% | 4,537 | 21.9 (21.4 - 22.5) | - | . |
| Male | 23,474 | 53.1% | 6,517 | 27.8 (27.2 - 28.3) | 1.27 (1.19 - 1.35) | <.0001 |
| **ADI^a^** |  |  |  |  |  |  |
| 81 - 100 (Reference) | 8,209 | 18.6% | 2,206 | 26.9 (25.9 - 27.8) | - | . |
| 61 - 80 | 10,771 | 24.4% | 2,776 | 25.8 (25.0 - 26.6) | 1.01 (0.93 - 1.09) | 0.878 |
| 41 - 60 | 9,599 | 21.7% | 2,376 | 24.8 (23.9 - 25.6) | 0.98 (0.90 - 1.07) | 0.627 |
| 21 - 40 | 7,663 | 17.3% | 1,747 | 22.8 (21.9 - 23.8) | 0.84 (0.76 - 0.92) | 0.000 |
| 0 - 20 | 3,540 | 8.0% | 777 | 21.9 (20.6 - 23.4) | 0.84 (0.74 - 0.95) | 0.005 |
| **Medicaid Eligibility** |  |  |  |  |  |  |
| No (Reference) | 35,704 | 80.8% | 8,709 | 24.4 (23.9 - 24.8) | - | . |
| Yes | 8,464 | 19.2% | 2,345 | 27.7 (26.8 - 28.7) | 1.10 (1.02 - 1.20) | 0.019 |
| **Sub-Region of Residence** |  |  |  |  |  |  |
| South: South Atlantic (Reference) | 20,207 | 45.8% | 5,036 | 24.9 (24.3 - 25.5) | - | . |
| South: East South Central | 8,181 | 18.5% | 2,200 | 26.9 (25.9 - 27.9) | 1.09 (1.01 - 1.19) | 0.028 |
| South: West South Central | 15,736 | 35.6% | 3,806 | 24.2 (23.5 - 24.9) | 0.94 (0.88 - 1.00) | 0.060 |
| **Urban/Rural^a^** |  |  |  |  |  |  |
| Urban (Reference) | 28,781 | 65.2% | 7,055 | 24.5 (24.0 - 25.0) | - | . |
| Rural | 11,368 | 25.7% | 2,920 | 25.7 (24.9 - 26.5) | 1.07 (0.99 - 1.15) | 0.078 |
| **Immunocompromised Status** |  |  |  |  |  |  |
| No (Reference) | 39,999 | 90.6% | 9,913 | 24.8 (24.4 - 25.2) | - | . |
| Yes | 4,169 | 9.4% | 1,141 | 27.4 (26.0 - 28.8) | 1.16 (1.03 - 1.31) | 0.014 |
| **Nursing Home Stay** |  |  |  |  |  |  |
| No (Reference) | 40,181 | 91.0% | 9,834 | 24.5 (24.1 - 24.9) | - | . |
| Yes | 3,987 | 9.0% | 1,220 | 30.6 (29.2 - 32.1) | 1.11 (1.01 - 1.24) | 0.038 |
| **Emergency Room Stay** |  |  |  |  |  |  |
| No (Reference) | 3,855 | 8.7% | 1,089 | 28.2 (26.8 - 29.7) | - | . |
| Yes | 40,313 | 91.3% | 9,965 | 24.7 (24.3 - 25.1) | 0.99 (0.89 - 1.10) | 0.881 |
| **COVID-19 As a Reason for Admission** |  |  |  |  |  |  |
| No (Reference) | 21,013 | 47.6% | 5,587 | 26.6 (26.0 - 27.2) | - | . |
| Yes | 23,155 | 52.4% | 5,467 | 23.6 (23.1 - 24.2) | 1.02 (0.96 - 1.08) | 0.581 |
| **Time Since Most Recent COVID-19 Diagnosis in the 30 Days Prior to Admission** |  |  |  |  |  |  |
| 0 days (Reference)^b^ | 21,329 | 48.3% | 5,310 | 24.9 (24.3 - 25.5) | - | . |
| 1 - 3 days | 10,276 | 23.3% | 2,512 | 24.4 (23.6 - 25.3) | 1.01 (0.94 - 1.08) | 0.836 |
| 4 - 10 days | 9,797 | 22.2% | 2,587 | 26.4 (25.5 - 27.3) | 1.13 (1.05 - 1.22) | 0.001 |
| 11-30 days | 2,766 | 6.3% | 645 | 23.3 (21.8 - 24.9) | 0.85 (0.75 - 0.96) | 0.010 |
| **Thrombosis on Index** |  |  |  |  |  |  |
| No (Reference) | 38,301 | 86.7% | 8,587 | 22.4 (22.0 - 22.8) | - | . |
| Yes | 5,867 | 13.3% | 2,467 | 42.0 (40.8 - 43.3) | 1.35 (1.24 - 1.48) | <.0001 |
| **Hospital Ownership^a^** |  |  |  |  |  |  |
| Government (Reference) | 7,680 | 17.4% | 1,983 | 25.8 (24.8 – 26.8) | - | . |
| For Profit | 11,907 | 27.0% | 2,983 | 25.1 (24.3 – 25.8) | 0.78 (0.71 - 0.85) | <.0001 |
| Non-Profit | 24,493 | 55.5% | 6,071 | 24.8 (24.2 – 25.3) | 0.88 (0.81 – 0.95) | 0.001 |
| **Hospital Size^a^** |  |  |  |  |  |  |
| Small (< 100 beds) (Reference) | 5,426 | 12.3% | 1,132 | 20.9 (19.8 - 22.0) | - | . |
| Medium (100 - 500 beds) | 27,679 | 62.7% | 7,026 | 25.4 (24.9 - 25.9) | 1.07 (0.97 - 1.18) | 0.153 |
| Large (> 500 beds) | 8,806 | 19.9% | 2,352 | 26.7 (25.8 - 27.6) | 1.05 (0.94 - 1.17) | 0.415 |
| **ICU/CCU Stay** |  |  |  |  |  |  |
| No (Reference) | 17,629 | 39.9% | 1,584 | 9.0 (8.6 - 9.4) | - | . |
| Yes | 26,539 | 60.1% | 9,470 | 35.7 (35.1 - 36.3) | 2.69 (2.50 - 2.88) | <.0001 |
| **Mechanical Ventilation** |  |  |  |  |  |  |
| No (Reference) | 36,507 | 82.7% | 5,370 | 14.7 (14.3 - 15.1) | - | . |
| Less Than 96hrs | 2,717 | 6.2% | 2,054 | 75.6 (73.9 - 77.2) | 14.41 (12.92 - 16.06) | <.0001 |
| More Than 96hrs | 4,944 | 11.2% | 3,630 | 73.4 (72.2 - 74.6) | 12.19 (11.17 - 13.30) | <.0001 |
| **Tocilizumab** |  |  |  |  |  |  |
| No (Reference) | 42,639 | 96.5% | 10,426 | 24.5 (24.0 - 24.9) | - | . |
| Yes | 1,529 | 3.5% | 628 | 41.1 (38.6 - 43.6) | 1.76 (1.53 - 2.04) | <.0001 |
| **Baricitinib** |  |  |  |  |  |  |
| No (Reference) | 43,679 | 98.9% | 10,895 | 24.9 (24.5 - 25.4) | - | . |
| Yes | 489 | 1.1% | 159 | 32.5 (28.4 - 36.9) | 1.67 (1.30 - 2.14) | <.0001 |
| **Remdesivir** |  |  |  |  |  |  |
| No (Reference) | 11,727 | 26.6% | 3,067 | 26.2 (25.4 - 27.0) | - | . |
| Yes | 32,441 | 73.4% | 7,987 | 24.6 (24.2 - 25.1) | 0.97 (0.91 - 1.04) | 0.412 |
| **Charlson Comorbidity Index (CCI)** |  |  |  |  |  |  |
| 0 (Reference) | 7,925 | 17.9% | 1,371 | 17.3 (16.5 - 18.2) | - | . |
| 1 | 11,181 | 25.3% | 2,353 | 21.0 (20.3 - 21.8) | 1.09 (0.98 - 1.21) | 0.113 |
| 2 | 8,689 | 19.7% | 2,241 | 25.8 (24.9 - 26.7) | 1.20 (1.06 - 1.35) | 0.003 |
| 3 – 4 | 9,869 | 22.3% | 2,903 | 29.4 (28.5 - 30.3) | 1.19 (1.04 - 1.37) | 0.012 |
| 5+ | 6,504 | 14.7% | 2,186 | 33.6 (32.5 - 34.8) | 1.34 (1.12 - 1.60) | 0.002 |
| **90-Day Comorbidity History** |  |  |  |  |  |  |
| **Thrombosis History** |  |  |  |  |  |  |
| No (Reference) | 42,930 | 97.2% | 10,640 | 24.8 (24.4 - 25.2) | - | . |
| Yes | 1,238 | 2.8% | 414 | 33.4 (30.8 - 36.2) | 1.04 (0.88 - 1.22) | 0.652 |
| **ESRD History** |  |  |  |  |  |  |
| No (Reference) | 42,866 | 97.1% | 10,696 | 25.0 (24.5 - 25.4) | - | . |
| Yes | 1,302 | 2.9% | 358 | 27.5 (25.1 - 30.0) | 0.78 (0.63 - 0.96) | 0.018 |
| **Elixhauser Comorbidity History** |  |  |  |  |  |  |
| **Blood Loss Anemia** |  |  |  |  |  |  |
| No (Reference) | 43,977 | 99.6% | 10,992 | 25.0 (24.6 - 25.4) | - | . |
| Yes | 191 | 0.4% | 62 | 32.5 (26.0 - 39.7) | 0.88 (0.58 - 1.33) | 0.537 |
| **Cardiac Arrhythmia** |  |  |  |  |  |  |
| No (Reference) | 28,101 | 63.6% | 5,602 | 19.9 (19.5 - 20.4) | - | . |
| Yes | 16,067 | 36.4% | 5,452 | 33.9 (33.2 - 34.7) | 1.36 (1.28 - 1.45) | <.0001 |
| **Chronic Pulmonary Disease** |  |  |  |  |  |  |
| No (Reference) | 30,359 | 68.7% | 7,445 | 24.5 (24.0 - 25.0) | - | . |
| Yes | 13,809 | 31.3% | 3,609 | 26.1 (25.4 - 26.9) | 1.02 (0.96 - 1.10) | 0.501 |
| **Coagulopathy** |  |  |  |  |  |  |
| No (Reference) | 36,909 | 83.6% | 8,290 | 22.5 (22.0 - 22.9) | - | . |
| Yes | 7,259 | 16.4% | 2,764 | 38.1 (37.0 - 39.2) | 1.30 (1.21 - 1.40) | <.0001 |
| **Congestive Heart Failure** |  |  |  |  |  |  |
| No (Reference) | 32,161 | 72.8% | 7,266 | 22.6 (22.1 - 23.1) | - | . |
| Yes | 12,007 | 27.2% | 3,788 | 31.5 (30.7 - 32.4) | 1.03 (0.95 - 1.12) | 0.442 |
| **Deficiency Anemia** |  |  |  |  |  |  |
| No (Reference) | 42,113 | 95.3% | 10,465 | 24.8 (24.4 - 25.3) | - | . |
| Yes | 2,055 | 4.7% | 589 | 28.7 (26.7 - 30.7) | 1.04 (0.91 - 1.18) | 0.596 |
| **Depression** |  |  |  |  |  |  |
| No (Reference) | 37,590 | 85.1% | 9,482 | 25.2 (24.8 - 25.7) | - | . |
| Yes | 6,578 | 14.9% | 1,572 | 23.9 (22.9 - 25.0) | 1.00 (0.92 - 1.09) | 0.968 |
| **Diabetes** |  |  |  |  |  |  |
| No (Reference) | 24,641 | 55.8% | 5,794 | 23.5 (23.0 - 24.0) | - | . |
| Yes | 19,527 | 44.2% | 5,260 | 26.9 (26.3 - 27.6) | 0.95 (0.88 - 1.03) | 0.231 |
| **Fluid and Electrolyte Disorder** |  |  |  |  |  |  |
| No (Reference) | 16,809 | 38.1% | 2,357 | 14.0 (13.5 - 14.6) | - | . |
| Yes | 27,359 | 61.9% | 8,697 | 31.8 (31.2 - 32.3) | 1.53 (1.44 - 1.64) | <.0001 |
| **Hypertension (uncomplicated)** |  |  |  |  |  |  |
| No (Reference) | 23,757 | 53.8% | 6,617 | 27.9 (27.3 - 28.4) | - | . |
| Yes | 20,411 | 46.2% | 4,437 | 21.7 (21.2 - 22.3) | 0.96 (0.89 - 1.04) | 0.325 |
| **Hypertension (complicated)** |  |  |  |  |  |  |
| No (Reference) | 25,883 | 58.6% | 5,520 | 21.3 (20.8 - 21.8) | - | . |
| Yes | 18,285 | 41.4% | 5,534 | 30.3 (29.6 - 30.9) | 1.07 (0.98 - 1.18) | 0.150 |
| **Liver Disease** |  |  |  |  |  |  |
| No (Reference) | 42,154 | 95.4% | 10,232 | 24.3 (23.9 - 24.7) | - | . |
| Yes | 2,014 | 4.6% | 822 | 40.8 (38.7 - 43.0) | 1.42 (1.25 - 1.62) | <.0001 |
| **Obesity** |  |  |  |  |  |  |
| No (Reference) | 33,367 | 75.5% | 8,176 | 24.5 (24.0 - 25.0) | - | . |
| Yes | 10,801 | 24.5% | 2,878 | 26.6 (25.8 - 27.5) | 1.04 (0.97 - 1.11) | 0.310 |
| **Other Neurological Disorders** |  |  |  |  |  |  |
| No (Reference) | 33,720 | 76.3% | 7,194 | 21.3 (20.9 - 21.8) | - | . |
| Yes | 10,448 | 23.7% | 3,860 | 36.9 (36.0 - 37.9) | 1.36 (1.27 - 1.45) | <.0001 |
| **Paralysis** |  |  |  |  |  |  |
| No (Reference) | 43,599 | 98.7% | 10,838 | 24.9 (24.5 - 25.3) | - | . |
| Yes | 569 | 1.3% | 216 | 38.0 (34.0 - 42.1) | 0.98 (0.77 - 1.24) | 0.863 |
| **Peripheral Vascular Disorder** |  |  |  |  |  |  |
| No (Reference) | 40,721 | 92.2% | 10,060 | 24.7 (24.3 - 25.1) | - | . |
| Yes | 3,447 | 7.8% | 994 | 28.8 (27.3 - 30.4) | 0.98 (0.88 - 1.09) | 0.746 |
| **Pulmonary Circulation Disorder** |  |  |  |  |  |  |
| No (Reference) | 42,759 | 96.8% | 10,587 | 24.8 (24.4 - 25.2) | - | . |
| Yes | 1,409 | 3.2% | 467 | 33.1 (30.7 - 35.7) | 1.01 (0.85 - 1.19) | 0.925 |
| **Renal Failure** |  |  |  |  |  |  |
| No (Reference) | 36,167 | 81.9% | 8,486 | 23.5 (23.0 - 23.9) | - | . |
| Yes | 8,001 | 18.1% | 2,568 | 32.1 (31.1 - 33.1) | 1.06 (0.96 - 1.17) | 0.253 |
| **Rheumatoid Arthritis/Collagen Vascular Diseases (CVD)** |  |  |  |  |  |  |
| No (Reference) | 42,145 | 95.4% | 10,534 | 25.0 (24.6 - 25.4) | - | . |
| Yes | 2,023 | 4.6% | 520 | 25.7 (23.8 - 27.7) | 1.20 (1.05 - 1.37) | 0.009 |
| **Solid Tumor** |  |  |  |  |  |  |
| No (Reference) | 42,620 | 96.5% | 10,628 | 24.9 (24.5 - 25.4) | - | . |
| Yes | 1,548 | 3.5% | 426 | 27.5 (25.3 - 29.8) | 1.11 (0.94 - 1.30) | 0.209 |
| **Valvular Disease** |  |  |  |  |  |  |
| No (Reference) | 41,010 | 92.9% | 10,115 | 24.7 (24.2 - 25.1) | - | . |
| Yes | 3,158 | 7.1% | 939 | 29.7 (28.1 - 31.4) | 0.97 (0.87 - 1.08) | 0.576 |
| **Weight Loss** |  |  |  |  |  |  |
| No (Reference) | 39,614 | 89.7% | 9,326 | 23.5 (23.1 - 24.0) | - | . |
| Yes | 4,554 | 10.3% | 1,728 | 37.9 (36.5 - 39.4) | 1.17 (1.07 - 1.28) | 0.000 |
| ^a^ Patients with missing values for ADI, Urban/Rural, Hospital Size and Ownership were dropped from the multivariate analysis. | | | | | | |
| ^b^ Patients have not had COVID-19 diagnosis recorded in the 30 days prior to admission. | | | | | | |
| **Notes:** Model evaluation metrics were ROC Area Under the Curve (0.8504); Pearson Chi Sq Goodness of Fit test p value (1); Deviance Goodness of Fit test p value (1). | | | | | |  |
| Intravenous Immunoglobulin/Hyperimmune Globulin and Peptic Ulcer Disease Groups have been omitted to avoid identification due to low counts. | | | | | | |

| **Supplementary Table 6.** Inpatient (IP) Mortality Rates and Corresponding Adjusted Odds Ratios (ORs) by Patient and Hospital Characteristics Among Recipients of COVID-19 Convalescent Plasma (CCP) Residing in the Midwest US Census Region | | | | | | |
| --- | --- | --- | --- | --- | --- | --- |
| **Patient and Hospital Characteristics** | **CCP Transfusion Stays #** | **CCP Transfusion Stays  %** | **IP Mortality  #** | **Crude IP Mortality Rate per 100  (95% CIs)** | **Adjusted ORs (95% CIs)** | **P-value** |
| **Overall** | 19,442 | 100.0% | 4,652 | 23.9 (23.3 - 24.5) | - | . |
| **Age (continuous)** | 19,442 | 100.0% | 4,652 | 23.9 (23.3 - 24.5) | 1.06 (1.06 - 1.07) | <.0001 |
| **Sex** |  |  |  |  |  |  |
| Female (Reference) | 8,515 | 43.8% | 1,841 | 21.6 (20.8 - 22.5) | - | . |
| Male | 10,927 | 56.2% | 2,811 | 25.7 (24.9 - 26.6) | 1.10 (1.00 - 1.20) | 0.044 |
| **ADI^a^** |  |  |  |  |  |  |
| 81 - 100 (Reference) | 2,579 | 13.3% | 678 | 26.3 (24.6 - 28.0) | - | . |
| 61 - 80 | 4,939 | 25.4% | 1,198 | 24.3 (23.1 - 25.5) | 0.95 (0.83 - 1.09) | 0.447 |
| 41 - 60 | 5,440 | 28.0% | 1,226 | 22.5 (21.4 - 23.7) | 0.88 (0.77 - 1.01) | 0.079 |
| 21 - 40 | 3,720 | 19.1% | 886 | 23.8 (22.5 - 25.2) | 0.95 (0.82 - 1.10) | 0.505 |
| 0 - 20 | 1,208 | 6.2% | 268 | 22.2 (19.9 - 24.7) | 0.78 (0.64 - 0.96) | 0.021 |
| **Medicaid Eligibility** |  |  |  |  |  |  |
| No (Reference) | 16,409 | 84.4% | 3,853 | 23.5 (22.8 - 24.1) | - | . |
| Yes | 3,033 | 15.6% | 799 | 26.3 (24.8 - 28.0) | 1.04 (0.91 - 1.18) | 0.582 |
| **Sub-Region of Residence** |  |  |  |  |  |  |
| Midwest: East North Central (Reference) | 11,599 | 59.7% | 2,810 | 24.2 (23.5 - 25.0) | - | . |
| Midwest: West North Central | 7,843 | 40.3% | 1,842 | 23.5 (22.6 - 24.4) | 1.10 (1.01 - 1.21) | 0.036 |
| **Urban/Rural^a^** |  |  |  |  |  |  |
| Urban (Reference) | 11,539 | 59.4% | 2,775 | 24.0 (23.3 - 24.8) | - | . |
| Rural | 6,501 | 33.4% | 1,528 | 23.5 (22.5 - 24.6) | 1.18 (1.06 - 1.31) | 0.002 |
| **Immunocompromised Status** |  |  |  |  |  |  |
| No (Reference) | 17,509 | 90.1% | 4,113 | 23.5 (22.9 - 24.1) | - | . |
| Yes | 1,933 | 9.9% | 539 | 27.9 (25.9 - 30.0) | 1.12 (0.94 - 1.33) | 0.197 |
| **Nursing Home Stay** |  |  |  |  |  |  |
| No (Reference) | 17,715 | 91.1% | 4,106 | 23.2 (22.6 - 23.8) | - | . |
| Yes | 1,727 | 8.9% | 546 | 31.6 (29.4 - 33.9) | 1.40 (1.21 - 1.63) | <.0001 |
| **Emergency Room Stay** |  |  |  |  |  |  |
| No (Reference) | 3,689 | 19.0% | 983 | 26.6 (25.2 - 28.1) | - | . |
| Yes | 15,753 | 81.0% | 3,669 | 23.3 (22.6 - 24.0) | 0.89 (0.79 - 1.00) | 0.052 |
| **COVID-19 As a Reason for Admission** |  |  |  |  |  |  |
| No (Reference) | 8,839 | 45.5% | 2,229 | 25.2 (24.3 - 26.1) | - | . |
| Yes | 10,603 | 54.5% | 2,423 | 22.9 (22.1 - 23.7) | 1.08 (0.99 - 1.18) | 0.097 |
| **Time Since Most Recent COVID-19 Diagnosis in the 30 Days Prior to Admission** |  |  |  |  |  |  |
| 0 days (Reference)^b^ | 9,630 | 49.5% | 2,271 | 23.6 (22.7 - 24.4) | - | . |
| 1 - 3 days | 4,940 | 25.4% | 1,134 | 23.0 (21.8 - 24.2) | 0.94 (0.85 - 1.05) | 0.262 |
| 4 - 10 days | 4,029 | 20.7% | 1,059 | 26.3 (24.9 - 27.7) | 1.07 (0.96 - 1.20) | 0.228 |
| 11-30 days | 843 | 4.3% | 188 | 22.3 (19.6 - 25.3) | 0.76 (0.61 - 0.94) | 0.013 |
| **Thrombosis on Index** |  |  |  |  |  |  |
| No (Reference) | 16,492 | 84.8% | 3,529 | 21.4 (20.8 - 22.0) | - | . |
| Yes | 2,950 | 15.2% | 1,123 | 38.1 (36.3 - 39.9) | 1.27 (1.12 - 1.44) | 0.000 |
| **Hospital Ownership^a^** |  |  |  |  |  |  |
| Government (Reference) | 1,480 | 7.6% | 288 | 19.5 (17.5 - 21.6) | - | . |
| For Profit | 1,104 | 5.7% | 267 | 24.2 (21.7 - 26.8) | 0.86 (0.66 - 1.11) | 0.238 |
| Non-Profit | 16,847 | 86.7% | 4,095 | 24.3 (23.7 - 25.0) | 0.94 (0.78 - 1.13) | 0.513 |
| **Hospital Size^a^** |  |  |  |  |  |  |
| Small (< 100 beds) (Reference) | 2,997 | 15.4% | 489 | 16.3 (15.0 - 17.7) | - | . |
| Medium (100 - 500 beds) | 12,209 | 62.8% | 3,017 | 24.7 (24.0 - 25.5) | 1.16 (1.01 - 1.33) | 0.037 |
| Large (> 500 beds) | 3,106 | 16.0% | 877 | 28.2 (26.7 - 29.9) | 1.19 (1.01 - 1.41) | 0.036 |
| **ICU/CCU Stay** |  |  |  |  |  |  |
| No (Reference) | 8,737 | 44.9% | 733 | 8.4 (7.8 - 9.0) | - | . |
| Yes | 10,705 | 55.1% | 3,919 | 36.6 (35.7 - 37.5) | 3.30 (2.97 - 3.67) | <.0001 |
| **Mechanical Ventilation** |  |  |  |  |  |  |
| No (Reference) | 16,303 | 83.9% | 2,444 | 15.0 (14.4 - 15.6) | - | . |
| Less Than 96hrs | 1,010 | 5.2% | 676 | 66.9 (63.9 - 69.8) | 8.82 (7.49 - 10.39) | <.0001 |
| More Than 96hrs | 2,129 | 11.0% | 1,532 | 72.0 (70.0 - 73.8) | 9.98 (8.76 - 11.36) | <.0001 |
| **Tocilizumab** |  |  |  |  |  |  |
| No (Reference) | 18,795 | 96.7% | 4,391 | 23.4 (22.8 - 24.0) | - | . |
| Yes | 647 | 3.3% | 261 | 40.3 (36.6 - 44.2) | 1.48 (1.20 - 1.82) | 0.000 |
| **Baricitinib** |  |  |  |  |  |  |
| No (Reference) | 19,334 | 99.4% | 4,619 | 23.9 (23.3 - 24.5) | - | . |
| Yes | 108 | 0.6% | 33 | 30.6 (22.2 - 40.3) | 1.27 (0.71 - 2.27) | 0.429 |
| **Remdesivir** |  |  |  |  |  |  |
| No (Reference) | 4,173 | 21.5% | 1,051 | 25.2 (23.9 - 26.5) | - | . |
| Yes | 15,269 | 78.5% | 3,601 | 23.6 (22.9 - 24.3) | 0.92 (0.82 - 1.02) | 0.113 |
| **Charlson Comorbidity Index (CCI)** |  |  |  |  |  |  |
| 0 (Reference) | 3,233 | 16.6% | 544 | 16.8 (15.6 - 18.2) | - | . |
| 1 | 4,691 | 24.1% | 898 | 19.1 (18.0 - 20.3) | 1.06 (0.91 - 1.24) | 0.451 |
| 2 | 3,749 | 19.3% | 897 | 23.9 (22.6 - 25.3) | 1.31 (1.11 - 1.56) | 0.002 |
| 3 – 4 | 4,762 | 24.5% | 1,368 | 28.7 (27.4 - 30.0) | 1.39 (1.15 - 1.67) | 0.001 |
| 5+ | 3,007 | 15.5% | 945 | 31.4 (29.8 - 33.1) | 1.44 (1.13 - 1.85) | 0.004 |
| **90-Day Comorbidity Index** |  |  |  |  |  |  |
| **Thrombosis History** |  |  |  |  |  |  |
| No (Reference) | 18,876 | 97.1% | 4,457 | 23.6 (23.0 - 24.2) | - | . |
| Yes | 566 | 2.9% | 195 | 34.5 (30.6 - 38.6) | 1.12 (0.88 - 1.42) | 0.349 |
| **ESRD History** |  |  |  |  |  |  |
| No (Reference) | 18,946 | 97.4% | 4,511 | 23.8 (23.2 - 24.4) | - | . |
| Yes | 496 | 2.6% | 141 | 28.4 (24.5 - 32.7) | 1.03 (0.75 - 1.40) | 0.863 |
| **Elixhauser Comorbidity History** |  |  |  |  |  |  |
| **Blood Loss Anemia** |  |  |  |  |  |  |
| No (Reference) | 19,345 | 99.5% | 4,616 | 23.9 (23.3 - 24.5) | - | . |
| Yes | 97 | 0.5% | 36 | 37.1 (27.7 - 47.6) | 1.67 (0.99 - 2.82) | 0.056 |
| **Cardiac Arrhythmia** |  |  |  |  |  |  |
| No (Reference) | 12,072 | 62.1% | 2,329 | 19.3 (18.6 - 20.0) | - | . |
| Yes | 7,370 | 37.9% | 2,323 | 31.5 (30.5 - 32.6) | 1.28 (1.17 - 1.40) | <.0001 |
| **Coagulopathy** |  |  |  |  |  |  |
| No (Reference) | 16,076 | 82.7% | 3,409 | 21.2 (20.6 - 21.8) | - | . |
| Yes | 3,366 | 17.3% | 1,243 | 36.9 (35.3 - 38.6) | 1.39 (1.25 - 1.55) | <.0001 |
| **Congestive Heart Failure** |  |  |  |  |  |  |
| No (Reference) | 13,658 | 70.2% | 2,947 | 21.6 (20.9 - 22.3) | - | . |
| Yes | 5,784 | 29.8% | 1,705 | 29.5 (28.3 - 30.7) | 1.00 (0.89 - 1.13) | 0.993 |
| **Diabetes** |  |  |  |  |  |  |
| No (Reference) | 11,104 | 57.1% | 2,502 | 22.5 (21.8 - 23.3) | - | . |
| Yes | 8,338 | 42.9% | 2,150 | 25.8 (24.9 - 26.7) | 0.96 (0.86 - 1.07) | 0.434 |
| **Fluid and Electrolyte Disorder** |  |  |  |  |  |  |
| No (Reference) | 7,688 | 39.5% | 1,074 | 14.0 (13.2 - 14.8) | - | . |
| Yes | 11,754 | 60.5% | 3,578 | 30.4 (29.6 - 31.3) | 1.45 (1.32 - 1.60) | <.0001 |
| **Hypertension (uncomplicated)** |  |  |  |  |  |  |
| No (Reference) | 11,416 | 58.7% | 3,007 | 26.3 (25.5 - 27.2) | - | . |
| Yes | 8,026 | 41.3% | 1,645 | 20.5 (19.6 - 21.4) | 0.99 (0.88 - 1.11) | 0.863 |
| **Hypertension (complicated)** |  |  |  |  |  |  |
| No (Reference) | 10,941 | 56.3% | 2,219 | 20.3 (19.5 - 21.0) | - | . |
| Yes | 8,501 | 43.7% | 2,433 | 28.6 (27.7 - 29.6) | 1.11 (0.97 - 1.27) | 0.120 |
| **Liver Disease** |  |  |  |  |  |  |
| No (Reference) | 18,621 | 95.8% | 4,322 | 23.2 (22.6 - 23.8) | - | . |
| Yes | 821 | 4.2% | 330 | 40.2 (36.8 - 43.6) | 1.36 (1.11 - 1.66) | 0.003 |
| **Metastatic Cancer** |  |  |  |  |  |  |
| No (Reference) | 19,132 | 98.4% | 4,559 | 23.8 (23.2 - 24.4) | - | . |
| Yes | 310 | 1.6% | 93 | 30.0 (25.0 - 35.5) | 1.22 (0.84 - 1.77) | 0.307 |
| **Other Neurological Disorders** |  |  |  |  |  |  |
| No (Reference) | 15,217 | 78.3% | 3,147 | 20.7 (20.0 - 21.3) | - | . |
| Yes | 4,225 | 21.7% | 1,505 | 35.6 (34.2 - 37.1) | 1.23 (1.11 - 1.36) | <.0001 |
| **Paralysis** |  |  |  |  |  |  |
| No (Reference) | 19,200 | 98.8% | 4,556 | 23.7 (23.1 - 24.3) | - | . |
| Yes | 242 | 1.2% | 96 | 39.7 (33.5 - 46.2) | 1.06 (0.74 - 1.52) | 0.746 |
| **Peripheral Vascular Disorder** |  |  |  |  |  |  |
| No (Reference) | 17,644 | 90.8% | 4,174 | 23.7 (23.0 - 24.3) | - | . |
| Yes | 1,798 | 9.2% | 478 | 26.6 (24.6 - 28.7) | 0.90 (0.77 - 1.04) | 0.149 |
| **Pulmonary Circulation Disorder** |  |  |  |  |  |  |
| No (Reference) | 18,621 | 95.8% | 4,394 | 23.6 (23.0 - 24.2) | - | . |
| Yes | 821 | 4.2% | 258 | 31.4 (28.3 - 34.7) | 1.09 (0.87 - 1.35) | 0.459 |
| **Renal Failure** |  |  |  |  |  |  |
| No (Reference) | 16,054 | 82.6% | 3,594 | 22.4 (21.7 - 23.0) | - | . |
| Yes | 3,388 | 17.4% | 1,058 | 31.2 (29.7 - 32.8) | 1.00 (0.87 - 1.16) | 0.956 |
| **Solid Tumor** |  |  |  |  |  |  |
| No (Reference) | 18,665 | 96.0% | 4,430 | 23.7 (23.1 - 24.4) | - | . |
| Yes | 777 | 4.0% | 222 | 28.6 (25.4 - 31.9) | 1.10 (0.86 - 1.39) | 0.446 |
| **Valvular Disease** |  |  |  |  |  |  |
| No (Reference) | 17,769 | 91.4% | 4,211 | 23.7 (23.1 - 24.3) | - | . |
| Yes | 1,673 | 8.6% | 441 | 26.4 (24.3 - 28.6) | 0.89 (0.77 - 1.03) | 0.120 |
| **Weight Loss** |  |  |  |  |  |  |
| No (Reference) | 17,361 | 89.3% | 3,879 | 22.3 (21.7 - 23.0) | - | . |
| Yes | 2,081 | 10.7% | 773 | 37.1 (35.1 - 39.3) | 1.08 (0.95 - 1.23) | 0.238 |
| ^a^ Patients with missing values for ADI, Urban/Rural, Hospital Size and Ownership were dropped from the multivariate analysis. | | | | | | |
| ^b^ Patients have not had COVID-19 diagnosis recorded in the 30 days prior to admission. | | | | | | |
| **Notes:** Model evaluation metrics were ROC Area Under the Curve (0.8438); Pearson Chi Sq Goodness of Fit test p value (1); Deviance Goodness of Fit test p value (1). | | | | | | |
| Intravenous Immunoglobulin/Hyperimmune Globulin Group has been omitted to avoid identification due to low counts. | | | | | | |

| **Supplementary Table 7.** Inpatient (IP) Mortality Rates and Corresponding Adjusted Odds Ratios (ORs) by Patient and Hospital Characteristics Among Recipients of COVID-19 Convalescent Plasma (CCP) Residing in the West US Census Region | | | | | |  |
| --- | --- | --- | --- | --- | --- | --- |
| **Patient and Hospital Characteristics** | **CCP Transfusion Stays #** | **CCP Transfusion Stays  %** | **IP Mortality  #** | **Crude IP Mortality Rate per 100  (95% CIs)** | **Adjusted ORs  (95% CIs)** | **P-value** |
| **Overall** | 10,496 | 100.0% | 3,133 | 29.8 (29.0 - 30.7) | - | . |
| **Age (continuous)** | 10,496 | 100.0% | 3,133 | 29.8 (29.0 - 30.7) | 1.06 (1.06 - 1.07) | <.0001 |
| **Sex** |  |  |  |  |  |  |
| Female (Reference) | 4,628 | 44.1% | 1,258 | 27.2 (25.9 - 28.5) | - | . |
| Male | 5,868 | 55.9% | 1,875 | 32.0 (30.8 - 33.2) | 1.14 (1.01 - 1.28) | 0.033 |
| **ADI^a^** |  |  |  |  |  |  |
| 81 - 100 (Reference) | 939 | 8.9% | 324 | 34.5 (31.5 - 37.7) | - | . |
| 61 - 80 | 1,185 | 11.3% | 343 | 28.9 (26.4 - 31.6) | 0.89 (0.71 - 1.13) | 0.350 |
| 41 - 60 | 1,812 | 17.3% | 497 | 27.4 (25.4 - 29.6) | 0.86 (0.69 - 1.07) | 0.170 |
| 21 - 40 | 2,700 | 25.7% | 793 | 29.4 (27.7 - 31.1) | 0.80 (0.65 - 0.98) | 0.028 |
| 0 - 20 | 2,805 | 26.7% | 821 | 29.3 (27.6 - 31.0) | 0.72 (0.58 - 0.89) | 0.002 |
| **Medicaid Eligibility** |  |  |  |  |  |  |
| No (Reference) | 7,017 | 66.9% | 1,856 | 26.5 (25.4 - 27.5) | - | . |
| Yes | 3,479 | 33.1% | 1,277 | 36.7 (35.1 - 38.3) | 1.11 (0.96 - 1.28) | 0.147 |
| **Sub-Region of Residence** |  |  |  |  |  |  |
| West: Mountain (Reference) | 4,847 | 46.2% | 1,240 | 25.6 (24.4 - 26.8) | - | . |
| West: Pacific | 5,649 | 53.8% | 1,893 | 33.5 (32.3 - 34.8) | 1.31 (1.15 - 1.50) | <.0001 |
| **Urban/Rural^a^** |  |  |  |  |  |  |
| Urban (Reference) | 8,709 | 83.0% | 2,600 | 29.9 (28.9 - 30.8) | - | . |
| Rural | 790 | 7.5% | 201 | 25.4 (22.5 - 28.7) | 1.13 (0.90 - 1.44) | 0.296 |
| **Immunocompromised Status** |  |  |  |  |  |  |
| No (Reference) | 9,290 | 88.5% | 2,749 | 29.6 (28.7 - 30.5) | - | . |
| Yes | 1,206 | 11.5% | 384 | 31.8 (29.2 - 34.6) | 1.10 (0.88 - 1.37) | 0.394 |
| **Nursing Home Stay** |  |  |  |  |  |  |
| No (Reference) | 9,631 | 91.8% | 2,809 | 29.2 (28.3 - 30.1) | - | . |
| Yes | 865 | 8.2% | 324 | 37.5 (34.2 - 40.8) | 1.06 (0.87 - 1.29) | 0.569 |
| **Emergency Room Stay** |  |  |  |  |  |  |
| No (Reference) | 837 | 8.0% | 239 | 28.6 (25.5 - 31.8) | - | . |
| Yes | 9,659 | 92.0% | 2,894 | 30.0 (29.1 - 30.9) | 1.26 (0.97 - 1.63) | 0.084 |
| **COVID-19 As a Reason for Admission** |  |  |  |  |  |  |
| No (Reference) | 5,404 | 51.5% | 1,766 | 32.7 (31.4 - 34.0) | - | . |
| Yes | 5,092 | 48.5% | 1,367 | 26.8 (25.6 - 28.1) | 0.96 (0.85 - 1.07) | 0.441 |
| **Time Since Most Recent COVID-19 Diagnosis in the 30 Days Prior to Admission** |  |  |  |  |  |  |
| 0 days (Reference)^b^ | 5,761 | 54.9% | 1,754 | 30.4 (29.3 - 31.7) | - | . |
| 1 - 3 days | 2,407 | 22.9% | 686 | 28.5 (26.7 - 30.4) | 1.02 (0.88 - 1.17) | 0.818 |
| 4 - 10 days | 1,770 | 16.9% | 507 | 28.6 (26.6 - 30.8) | 0.97 (0.83 - 1.14) | 0.741 |
| 11-30 days | 558 | 5.3% | 186 | 33.3 (29.5 - 37.4) | 1.01 (0.78 - 1.30) | 0.957 |
| **Thrombosis on Index** |  |  |  |  |  |  |
| No (Reference) | 8,897 | 84.8% | 2,392 | 26.9 (26.0 - 27.8) | - | . |
| Yes | 1,599 | 15.2% | 741 | 46.3 (43.9 - 48.8) | 1.24 (1.05 - 1.45) | 0.010 |
| **Hospital Ownership^a^** |  |  |  |  |  |  |
| Government (Reference) | 973 | 9.3% | 283 | 29.1 (26.3 - 32.1) | - | . |
| For Profit | 3,055 | 29.1% | 990 | 32.4 (30.8 - 34.1) | 0.94 (0.74 - 1.18) | 0.579 |
| Non-Profit | 6,459 | 61.5% | 1,859 | 28.8 (27.7 - 29.9) | 0.85 (0.69 - 1.06) | 0.154 |
| **Hospital Size^a^** |  |  |  |  |  |  |
| Small (< 100 beds) (Reference) | 831 | 7.9% | 180 | 21.7 (18.9 - 24.7) | - | . |
| Medium (100 - 500 beds) | 8,209 | 78.2% | 2,482 | 30.2 (29.2 - 31.2) | 1.18 (0.93 - 1.49) | 0.173 |
| Large (> 500 beds) | 1,135 | 10.8% | 378 | 33.3 (30.6 - 36.1) | 1.24 (0.93 - 1.64) | 0.145 |
| **ICU/CCU Stay** |  |  |  |  |  |  |
| No (Reference) | 3,340 | 31.8% | 356 | 10.7 (9.6 - 11.8) | - | . |
| Yes | 7,156 | 68.2% | 2,777 | 38.8 (37.7 - 39.9) | 2.52 (2.17 - 2.92) | <.0001 |
| **Mechanical Ventilation** |  |  |  |  |  |  |
| No (Reference) | 8,331 | 79.4% | 1,480 | 17.8 (17.0 - 18.6) | - | . |
| Less Than 96hrs | 657 | 6.3% | 526 | 80.1 (76.8 - 83.0) | 15.90 (12.64 - 20.01) | <.0001 |
| More Than 96hrs | 1,508 | 14.4% | 1,127 | 74.7 (72.4 - 76.9) | 11.79 (10.02 - 13.86) | <.0001 |
| **Tocilizumab** |  |  |  |  |  |  |
| No (Reference) | 10,270 | 97.8% | 3,036 | 29.6 (28.7 - 30.5) | - | . |
| Yes | 226 | 2.2% | 97 | 42.9 (36.4 - 49.7) | 1.02 (0.71 - 1.47) | 0.898 |
| **Baricitinib** |  |  |  |  |  |  |
| No (Reference) | 10,384 | 98.9% | 3,095 | 29.8 (28.9 - 30.7) | - | . |
| Yes | 112 | 1.1% | 38 | 33.9 (25.4 - 43.6) | 1.03 (0.60 - 1.78) | 0.903 |
| **Remdesivir** |  |  |  |  |  |  |
| No (Reference) | 2,466 | 23.5% | 843 | 34.2 (32.3 - 36.1) | - | . |
| Yes | 8,030 | 76.5% | 2,290 | 28.5 (27.5 - 29.5) | 0.93 (0.81 - 1.07) | 0.317 |
| **Charlson Comorbidity Index (CCI)** |  |  |  |  |  |  |
| 0 (Reference) | 1,816 | 17.3% | 369 | 20.3 (18.5 - 22.3) | - | . |
| 1 | 2,764 | 26.3% | 745 | 27.0 (25.3 - 28.7) | 1.21 (0.99 - 1.47) | 0.064 |
| 2 | 1,993 | 19.0% | 606 | 30.4 (28.4 - 32.5) | 1.16 (0.93 - 1.45) | 0.188 |
| 3 – 4 | 2,344 | 22.3% | 818 | 34.9 (33.0 - 36.9) | 1.21 (0.95 - 1.54) | 0.121 |
| 5+ | 1,579 | 15.0% | 595 | 37.7 (35.3 - 40.1) | 1.18 (0.87 - 1.60) | 0.297 |
| **90-Day Comorbidity History** |  |  |  |  |  |  |
| **Thrombosis History** |  |  |  |  |  |  |
| No (Reference) | 10,213 | 97.3% | 3,040 | 29.8 (28.9 - 30.7) | - | . |
| Yes | 283 | 2.7% | 93 | 32.9 (27.5 - 38.7) | 0.69 (0.48 - 0.98) | 0.036 |
| **ESRD History** |  |  |  |  |  |  |
| No (Reference) | 10,062 | 95.9% | 2,976 | 29.6 (28.7 - 30.5) | - | . |
| Yes | 434 | 4.1% | 157 | 36.2 (31.7 - 40.9) | 0.93 (0.66 - 1.33) | 0.710 |
| **Elixhauser Comorbidity History** |  |  |  |  |  |  |
| **Cardiac Arrhythmia** |  |  |  |  |  |  |
| No (Reference) | 6,736 | 64.2% | 1,641 | 24.4 (23.3 - 25.4) | - | . |
| Yes | 3,760 | 35.8% | 1,492 | 39.7 (38.1 - 41.3) | 1.39 (1.24 - 1.56) | <.0001 |
| **Coagulopathy** |  |  |  |  |  |  |
| No (Reference) | 8,304 | 79.1% | 2,246 | 27.0 (26.1 - 28.0) | - | . |
| Yes | 2,192 | 20.9% | 887 | 40.5 (38.4 - 42.6) | 1.20 (1.05 - 1.37) | 0.008 |
| **Congestive Heart Failure** |  |  |  |  |  |  |
| No (Reference) | 7,982 | 76.0% | 2,210 | 27.7 (26.7 - 28.7) | - | . |
| Yes | 2,514 | 24.0% | 923 | 36.7 (34.8 - 38.6) | 1.03 (0.87 - 1.21) | 0.732 |
| **Deficiency Anemia** |  |  |  |  |  |  |
| No (Reference) | 10,055 | 95.8% | 3,006 | 29.9 (29.0 - 30.8) | - | . |
| Yes | 441 | 4.2% | 127 | 28.8 (24.7 - 33.3) | 0.65 (0.49 - 0.86) | 0.003 |
| **Depression** |  |  |  |  |  |  |
| No (Reference) | 9,211 | 87.8% | 2,783 | 30.2 (29.3 - 31.2) | - | . |
| Yes | 1,285 | 12.2% | 350 | 27.2 (24.8 - 29.8) | 0.99 (0.83 - 1.18) | 0.908 |
| **Diabetes** |  |  |  |  |  |  |
| No (Reference) | 5,766 | 54.9% | 1,617 | 28.0 (26.9 - 29.2) | - | . |
| Yes | 4,730 | 45.1% | 1,516 | 32.1 (30.7 - 33.4) | 0.95 (0.83 - 1.10) | 0.514 |
| **Fluid and Electrolyte Disorder** |  |  |  |  |  |  |
| No (Reference) | 3,580 | 34.1% | 596 | 16.6 (15.5 - 17.9) | - | . |
| Yes | 6,916 | 65.9% | 2,537 | 36.7 (35.5 - 37.8) | 1.73 (1.52 - 1.98) | <.0001 |
| **Hypertension (uncomplicated)** |  |  |  |  |  |  |
| No (Reference) | 5,811 | 55.4% | 1,900 | 32.7 (31.5 - 33.9) | - | . |
| Yes | 4,685 | 44.6% | 1,233 | 26.3 (25.1 - 27.6) | 0.89 (0.77 - 1.03) | 0.106 |
| **Hypertension (complicated)** |  |  |  |  |  |  |
| No (Reference) | 6,549 | 62.4% | 1,739 | 26.6 (25.5 - 27.6) | - | . |
| Yes | 3,947 | 37.6% | 1,394 | 35.3 (33.8 - 36.8) | 0.94 (0.79 - 1.12) | 0.485 |
| **Hypothyroidism** |  |  |  |  |  |  |
| No (Reference) | 8,351 | 79.6% | 2,484 | 29.7 (28.8 - 30.7) | - | . |
| Yes | 2,145 | 20.4% | 649 | 30.3 (28.3 - 32.3) | 1.21 (1.05 - 1.39) | 0.007 |
| **Liver Disease** |  |  |  |  |  |  |
| No (Reference) | 9,887 | 94.2% | 2,850 | 28.8 (27.9 - 29.7) | - | . |
| Yes | 609 | 5.8% | 283 | 46.5 (42.5 - 50.5) | 1.73 (1.36 - 2.19) | <.0001 |
| **Other Neurological Disorders** |  |  |  |  |  |  |
| No (Reference) | 7,760 | 73.9% | 1,987 | 25.6 (24.6 - 26.6) | - | . |
| Yes | 2,736 | 26.1% | 1,146 | 41.9 (40.0 - 43.8) | 1.33 (1.17 - 1.50) | <.0001 |
| **Paralysis** |  |  |  |  |  |  |
| No (Reference) | 10,337 | 98.5% | 3,063 | 29.6 (28.8 - 30.5) | - | . |
| Yes | 159 | 1.5% | 70 | 44.0 (36.2 - 52.1) | 0.98 (0.64 - 1.51) | 0.935 |
| **Pulmonary Circulation Disorder** |  |  |  |  |  |  |
| No (Reference) | 10,150 | 96.7% | 2,998 | 29.5 (28.7 - 30.4) | - | . |
| Yes | 346 | 3.3% | 135 | 39.0 (33.9 - 44.4) | 1.44 (1.05 - 1.98) | 0.022 |
| **Renal Failure** |  |  |  |  |  |  |
| No (Reference) | 8,459 | 80.6% | 2,368 | 28.0 (27.0 - 29.0) | - | . |
| Yes | 2,037 | 19.4% | 765 | 37.6 (35.5 - 39.7) | 1.15 (0.95 - 1.40) | 0.158 |
| **Weight Loss** |  |  |  |  |  |  |
| No (Reference) | 9,129 | 87.0% | 2,564 | 28.1 (27.2 - 29.0) | - | . |
| Yes | 1,367 | 13.0% | 569 | 41.6 (39.0 - 44.3) | 1.11 (0.94 - 1.30) | 0.209 |
| ^a^ Patients with missing values for ADI, Urban/Rural, Hospital Size and Ownership were dropped from the multivariate analysis. | | | | | | |
| ^b^ Patients have not had COVID-19 diagnosis recorded in the 30 days prior to admission. | | | | | | |
| **Notes:** Model evaluation metrics were ROC Area Under the Curve (0.8489); Pearson Chi Sq Goodness of Fit test p value (0.9073); Deviance Goodness of Fit test p value (1). | | | | | |  |
| Intravenous Immunoglobulin/Hyperimmune Globulin and Peptic Ulcer Disease Groups have been omitted to avoid identification due to low counts. | | | | | | |

| **Supplementary Table 8.** Inpatient (IP) Mortality Rates and Corresponding Adjusted Odds Ratios (ORs) by Patient and Hospital Characteristics Among Recipients of COVID-19 Convalescent Plasma (CCP) Residing in the Northeast US Census Region | | | | | | |
| --- | --- | --- | --- | --- | --- | --- |
| **Patient and Hospital Characteristics** | **CCP Transfusion Stays #** | **CCP Transfusion Stays  %** | **IP Mortality  #** | **Crude IP Mortality Rate per 100  (95% CIs)** | **Adjusted ORs (95% CIs)** | **P-value** |
| **Overall** | 7,645 | 100.0% | 2,037 | 26.6 (25.7 - 27.7) | - | . |
| **Age (continuous)** | 7,645 | 100.0% | 2,037 | 26.6 (25.7 - 27.7) | 1.07 (1.06 - 1.08) | <.0001 |
| **Sex** |  |  |  |  |  |  |
| Female (Reference) | 3,501 | 45.8% | 843 | 24.1 (22.7 - 25.5) | - | . |
| Male | 4,144 | 54.2% | 1,194 | 28.8 (27.4 - 30.2) | 1.15 (1.01 - 1.33) | 0.042 |
| **ADI^a^** |  |  |  |  |  |  |
| 81 - 100 (Reference) | 703 | 9.2% | 203 | 28.9 (25.6 - 32.4) | - | . |
| 61 – 80 | 688 | 9.0% | 210 | 30.5 (27.1 - 34.1) | 1.24 (0.92 - 1.67) | 0.159 |
| 41 – 60 | 1,363 | 17.8% | 368 | 27.0 (24.7 - 29.5) | 1.04 (0.79 - 1.35) | 0.793 |
| 21 – 40 | 1,960 | 25.6% | 498 | 25.4 (23.5 - 27.4) | 0.88 (0.69 - 1.14) | 0.340 |
| 0 – 20 | 2,498 | 32.7% | 651 | 26.1 (24.4 - 27.8) | 0.96 (0.75 - 1.22) | 0.723 |
| **Medicaid Eligibility** |  |  |  |  |  |  |
| No (Reference) | 5,784 | 75.7% | 1,475 | 25.5 (24.4 - 26.6) | - | . |
| Yes | 1,861 | 24.3% | 562 | 30.2 (28.1 - 32.4) | 1.26 (1.07 - 1.49) | 0.005 |
| **Sub-Region of Residence** |  |  |  |  |  |  |
| Northeast: New England (Reference) | 1,331 | 17.4% | 301 | 22.6 (20.4 - 25.0) | - | . |
| Northeast: Middle Atlantic | 6,314 | 82.6% | 1,736 | 27.5 (26.4 - 28.6) | 1.26 (1.05 - 1.52) | 0.013 |
| **Urban/Rural^a^** |  |  |  |  |  |  |
| Urban (Reference) | 6,667 | 87.2% | 1,780 | 26.7 (25.6 - 27.8) | - | . |
| Rural | 640 | 8.4% | 175 | 27.3 (24.0 - 31.0) | 1.27 (0.98 - 1.65) | 0.066 |
| **Immunocompromised Status** |  |  |  |  |  |  |
| No (Reference) | 6,806 | 89.0% | 1,797 | 26.4 (25.4 - 27.5) | - | . |
| Yes | 839 | 11.0% | 240 | 28.6 (25.6 - 31.8) | 1.16 (0.93 - 1.46) | 0.189 |
| **Nursing Home Stay** |  |  |  |  |  |  |
| No (Reference) | 6,986 | 91.4% | 1,798 | 25.7 (24.7 - 26.8) | - | . |
| Yes | 659 | 8.6% | 239 | 36.3 (32.6 - 40.1) | 1.50 (1.20 - 1.87) | 0.000 |
| **Emergency Room Stay** |  |  |  |  |  |  |
| No (Reference) | 424 | 5.5% | 141 | 33.3 (28.8 - 38.0) | - | . |
| Yes | 7,221 | 94.5% | 1,896 | 26.3 (25.3 - 27.3) | 0.71 (0.53 - 0.94) | 0.019 |
| **COVID-19 As a Reason for Admission** |  |  |  |  |  |  |
| No (Reference) | 3,861 | 50.5% | 1,026 | 26.6 (25.2 - 28.0) | - | . |
| Yes | 3,784 | 49.5% | 1,011 | 26.7 (25.3 - 28.2) | 1.14 (1.00 - 1.30) | 0.058 |
| **Time Since Most Recent COVID-19 Diagnosis in the 30 Days Prior to Admission** |  |  |  |  |  |  |
| 0 days (Reference)^b^ | 4,262 | 55.7% | 1,169 | 27.4 (26.1 - 28.8) | - | . |
| 1 - 3 days | 1,774 | 23.2% | 459 | 25.9 (23.9 - 28.0) | 0.91 (0.77 - 1.07) | 0.250 |
| 4 - 10 days | 1,237 | 16.2% | 320 | 25.9 (23.5 - 28.4) | 0.91 (0.75 - 1.10) | 0.331 |
| 11-30 days | 372 | 4.9% | 89 | 23.9 (19.7 - 28.7) | 0.65 (0.47 - 0.91) | 0.011 |
| **Thrombosis on Index** |  |  |  |  |  |  |
| No (Reference) | 6,532 | 85.4% | 1,552 | 23.8 (22.7 - 24.8) | - | . |
| Yes | 1,113 | 14.6% | 485 | 43.6 (40.6 - 46.6) | 1.51 (1.25 - 1.83) | <.0001 |
| **Hospital Ownership^a^** |  |  |  |  |  |  |
| Government (Reference) | 328 | 4.3% | 99 | 30.2 (25.3 - 35.5) | - | . |
| For Profit | 432 | 5.7% | 114 | 26.4 (22.4 - 30.9) | 0.75 (0.49 - 1.16) | 0.195 |
| Non-Profit | 6,841 | 89.5% | 1,812 | 26.5 (25.4 - 27.6) | 0.80 (0.59 - 1.10) | 0.175 |
| **Hospital Size^a^** |  |  |  |  |  |  |
| Small (< 100 beds) (Reference) | 426 | 5.6% | 94 | 22.1 (18.3 - 26.4) | - | . |
| Medium (100 - 500 beds) | 4,408 | 57.7% | 1,215 | 27.6 (26.3 - 28.9) | 1.50 (1.09 - 2.07) | 0.013 |
| Large (> 500 beds) | 2,591 | 33.9% | 670 | 25.9 (24.2 - 27.6) | 1.16 (0.83 - 1.62) | 0.382 |
| **ICU/CCU Stay** |  |  |  |  |  |  |
| No (Reference) | 3,647 | 47.7% | 471 | 12.9 (11.9 - 14.1) | - | . |
| Yes | 3,998 | 52.3% | 1,566 | 39.2 (37.7 - 40.7) | 2.23 (1.92 - 2.59) | <.0001 |
| **Mechanical Ventilation** |  |  |  |  |  |  |
| No (Reference) | 6,343 | 83.0% | 1,036 | 16.3 (15.4 - 17.3) | - | . |
| Less Than 96hrs | 447 | 5.8% | 343 | 76.7 (72.5 - 80.5) | 17.06 (13.03 - 22.33) | <.0001 |
| More Than 96hrs | 855 | 11.2% | 658 | 77.0 (74.0 - 79.7) | 14.06 (11.36 - 17.40) | <.0001 |
| **Tocilizumab** |  |  |  |  |  |  |
| No (Reference) | 7,282 | 95.3% | 1,868 | 25.7 (24.7 - 26.7) | - | . |
| Yes | 363 | 4.7% | 169 | 46.6 (41.4 - 51.8) | 2.29 (1.73 - 3.03) | <.0001 |
| **Baricitinib** |  |  |  |  |  |  |
| No (Reference) | 7,572 | 99.0% | 2,013 | 26.6 (25.6 - 27.6) | - | . |
| Yes | 73 | 1.0% | 24 | 32.9 (22.6 - 45.0) | 1.01 (0.50 - 2.03) | 0.976 |
| **Remdesivir** |  |  |  |  |  |  |
| No (Reference) | 1,547 | 20.2% | 470 | 30.4 (28.1 - 32.8) | - | . |
| Yes | 6,098 | 79.8% | 1,567 | 25.7 (24.6 - 26.8) | 0.83 (0.70 - 0.98) | 0.025 |
| **Charlson Comorbidity Index (CCI)** |  |  |  |  |  |  |
| 0 (Reference) | 1,304 | 17.1% | 241 | 18.5 (16.4 - 20.7) | - | . |
| 1 | 1,867 | 24.4% | 425 | 22.8 (20.9 - 24.8) | 1.24 (0.98 - 1.58) | 0.069 |
| 2 | 1,509 | 19.7% | 429 | 28.4 (26.2 - 30.8) | 1.50 (1.16 - 1.94) | 0.002 |
| 3 – 4 | 1,800 | 23.5% | 547 | 30.4 (28.3 - 32.6) | 1.49 (1.12 - 1.97) | 0.006 |
| 5+ | 1,165 | 15.2% | 395 | 33.9 (31.2 - 36.7) | 1.63 (1.15 - 2.32) | 0.006 |
| **90-Day Comorbidity History** |  |  |  |  |  |  |
| **Thrombosis History** |  |  |  |  |  |  |
| No (Reference) | 7,415 | 97.0% | 1,954 | 26.4 (25.4 - 27.4) | - | . |
| Yes | 230 | 3.0% | 83 | 36.1 (30.0 - 42.7) | 0.96 (0.66 - 1.40) | 0.833 |
| **Elixhauser Comorbidity History** |  |  |  |  |  |  |
| **Cardiac Arrhythmia** |  |  |  |  |  |  |
| No (Reference) | 4,655 | 60.9% | 970 | 20.8 (19.7 - 22.0) | - | . |
| Yes | 2,990 | 39.1% | 1,067 | 35.7 (34.0 - 37.4) | 1.36 (1.18 - 1.56) | <.0001 |
| **Coagulopathy** |  |  |  |  |  |  |
| No (Reference) | 6,191 | 81.0% | 1,490 | 24.1 (23.0 - 25.2) | - | . |
| Yes | 1,454 | 19.0% | 547 | 37.6 (35.1 - 40.2) | 1.22 (1.03 - 1.44) | 0.019 |
| **Congestive Heart Failure** |  |  |  |  |  |  |
| No (Reference) | 5,377 | 70.3% | 1,286 | 23.9 (22.8 - 25.1) | - | . |
| Yes | 2,268 | 29.7% | 751 | 33.1 (31.2 - 35.1) | 0.87 (0.72 - 1.05) | 0.139 |
| **Deficiency Anemia** |  |  |  |  |  |  |
| No (Reference) | 7,247 | 94.8% | 1,908 | 26.3 (25.3 - 27.4) | - | . |
| Yes | 398 | 5.2% | 129 | 32.4 (27.9 - 37.3) | 1.23 (0.93 - 1.62) | 0.154 |
| **Diabetes** |  |  |  |  |  |  |
| No (Reference) | 4,511 | 59.0% | 1,155 | 25.6 (24.3 - 26.9) | - | . |
| Yes | 3,134 | 41.0% | 882 | 28.1 (26.6 - 29.8) | 0.85 (0.72 - 1.00) | 0.056 |
| **Fluid and Electrolyte Disorder** |  |  |  |  |  |  |
| No (Reference) | 2,825 | 37.0% | 387 | 13.7 (12.5 - 15.0) | - | . |
| Yes | 4,820 | 63.0% | 1,650 | 34.2 (32.9 - 35.6) | 1.72 (1.48 - 2.01) | <.0001 |
| **Hypertension (uncomplicated)** |  |  |  |  |  |  |
| No (Reference) | 4,356 | 57.0% | 1,292 | 29.7 (28.3 - 31.0) | - | . |
| Yes | 3,289 | 43.0% | 745 | 22.7 (21.2 - 24.1) | 0.92 (0.76 - 1.10) | 0.348 |
| **Hypertension (complicated)** |  |  |  |  |  |  |
| No (Reference) | 4,319 | 56.5% | 961 | 22.3 (21.0 - 23.5) | - | . |
| Yes | 3,326 | 43.5% | 1,076 | 32.4 (30.8 - 34.0) | 1.16 (0.94 - 1.44) | 0.171 |
| **Hypothyroidism** |  |  |  |  |  |  |
| No (Reference) | 6,104 | 79.8% | 1,601 | 26.2 (25.1 - 27.4) | - | . |
| Yes | 1,541 | 20.2% | 436 | 28.3 (26.1 - 30.6) | 1.18 (1.01 - 1.40) | 0.043 |
| **Liver Disease** |  |  |  |  |  |  |
| No (Reference) | 7,261 | 95.0% | 1,886 | 26.0 (25.0 - 27.0) | - | . |
| Yes | 384 | 5.0% | 151 | 39.3 (34.4 - 44.4) | 1.22 (0.91 - 1.63) | 0.178 |
| **Other Neurological Disorders** |  |  |  |  |  |  |
| No (Reference) | 5,842 | 76.4% | 1,359 | 23.3 (22.2 - 24.4) | - | . |
| Yes | 1,803 | 23.6% | 678 | 37.6 (35.4 - 39.9) | 1.30 (1.11 - 1.51) | 0.001 |
| **Paralysis** |  |  |  |  |  |  |
| No (Reference) | 7,547 | 98.7% | 1,992 | 26.4 (25.4 - 27.4) | - | . |
| Yes | 98 | 1.3% | 45 | 45.9 (35.9 - 56.3) | 1.20 (0.68 - 2.13) | 0.527 |
| **Pulmonary Circulation Disorder** |  |  |  |  |  |  |
| No (Reference) | 7,413 | 97.0% | 1,955 | 26.4 (25.4 - 27.4) | - | . |
| Yes | 232 | 3.0% | 82 | 35.3 (29.3 - 41.9) | 0.84 (0.56 - 1.24) | 0.376 |
| **Renal Failure** |  |  |  |  |  |  |
| No (Reference) | 6,240 | 81.6% | 1,553 | 24.9 (23.8 - 26.0) | - | . |
| Yes | 1,405 | 18.4% | 484 | 34.4 (32.0 - 37.0) | 1.01 (0.81 - 1.25) | 0.944 |
| **Valvular Disease** |  |  |  |  |  |  |
| No (Reference) | 6,811 | 89.1% | 1,788 | 26.3 (25.2 - 27.3) | - | . |
| Yes | 834 | 10.9% | 249 | 29.9 (26.8 - 33.1) | 0.95 (0.77 - 1.17) | 0.654 |
| **Weight Loss** |  |  |  |  |  |  |
| No (Reference) | 6,792 | 88.8% | 1,695 | 25.0 (23.9 - 26.0) | - | . |
| Yes | 853 | 11.2% | 342 | 40.1 (36.8 - 43.5) | 1.22 (1.00 - 1.48) | 0.052 |
| ^a^ Patients with missing values for ADI, Urban/Rural, Hospital Size and Ownership were dropped from the multivariate analysis. | | | | | | |
| ^b^ Patients have not had COVID-19 diagnosis recorded in the 30 days prior to admission. | | | | | | |
| **Notes:** Model evaluation metrics were ROC Area Under the Curve (0.8564); Pearson Chi Sq Goodness of Fit test p value (0.9969); Deviance Goodness of Fit test p value (1). | | | | | | |
| Intravenous Immunoglobulin/Hyperimmune Globulin Group has been omitted to avoid identification due to low counts. | | | | | | |

| **Supplementary Table 9.** Inpatient (IP) Mortality Rates and Corresponding Adjusted Odds Ratios (ORs) by Patient and Hospital Characteristics Among Recipients of COVID-19 Convalescent Plasma (CCP) Aged 65-79 | | | | | |  |
| --- | --- | --- | --- | --- | --- | --- |
| **Patient and Hospital Characteristics** | **CCP Transfusion Stays #** | **CCP Transfusion Stays  %** | **IP Mortality  #** | **Crude IP Mortality Rate per 100  (95% CIs)** | **Adjusted ORs (95% CIs)** | **P-value** |
| **Overall** | 51,618 | 100.0% | 11,972 | 23.2 (22.8 - 23.6) | - | . |
| **Age (continuous)** | 51,618 | 100.0% | 11,972 | 23.2 (22.8 - 23.6) | 1.07 (1.06 - 1.08) | <.0001 |
| **Sex** |  |  |  |  |  |  |
| Female (Reference) | 22,741 | 44.1% | 4,694 | 20.6 (20.1 - 21.2) | - | . |
| Male | 28,877 | 55.9% | 7,278 | 25.2 (24.7 - 25.7) | 1.14 (1.08 - 1.21) | <.0001 |
| **ADI^a^** |  |  |  |  |  |  |
| 81 - 100 (Reference) | 8,183 | 15.9% | 2,075 | 25.4 (24.4 - 26.3) | - | . |
| 61 – 80 | 11,387 | 22.1% | 2,670 | 23.4 (22.7 - 24.2) | 0.99 (0.91 - 1.08) | 0.861 |
| 41 – 60 | 11,632 | 22.5% | 2,631 | 22.6 (21.9 - 23.4) | 0.98 (0.89 - 1.07) | 0.611 |
| 21 – 40 | 9,949 | 19.3% | 2,213 | 22.2 (21.4 - 23.1) | 0.88 (0.80 - 0.97) | 0.008 |
| 0 – 20 | 5,605 | 10.9% | 1,200 | 21.4 (20.3 - 22.5) | 0.83 (0.74 - 0.93) | 0.002 |
| **Medicaid Eligibility** |  |  |  |  |  |  |
| No (Reference) | 41,406 | 80.2% | 9,228 | 22.3 (21.9 - 22.7) | - | . |
| Yes | 10,212 | 19.8% | 2,744 | 26.9 (26.0 - 27.7) | 1.15 (1.06 - 1.24) | 0.001 |
| **Region of Residence^b^** |  |  |  |  |  |  |
| South (Reference) | 28,430 | 55.1% | 6,486 | 22.8 (22.3 - 23.3) | - | . |
| Midwest | 12,250 | 23.7% | 2,676 | 21.8 (21.1 - 22.6) | 0.98 (0.91 - 1.06) | 0.639 |
| West | 6,651 | 12.9% | 1,820 | 27.4 (26.3 - 28.5) | 1.13 (1.03 - 1.24) | 0.007 |
| Northeast | 4,287 | 8.3% | 990 | 23.1 (21.8 - 24.4) | 1.09 (0.98 - 1.22) | 0.128 |
| **Urban/Rural^a^** |  |  |  |  |  |  |
| Urban (Reference) | 34,446 | 66.7% | 8,025 | 23.3 (22.9 - 23.7) | - | . |
| Rural | 12,725 | 24.7% | 2,867 | 22.5 (21.8 - 23.3) | 1.07 (0.99 - 1.15) | 0.090 |
| **Immunocompromised Status** |  |  |  |  |  |  |
| No (Reference) | 45,745 | 88.6% | 10,391 | 22.7 (22.3 - 23.1) | - | . |
| Yes | 5,873 | 11.4% | 1,581 | 26.9 (25.8 - 28.1) | 1.12 (1.00 - 1.25) | 0.049 |
| **Nursing Home Stay** |  |  |  |  |  |  |
| No (Reference) | 48,243 | 93.5% | 11,006 | 22.8 (22.4 - 23.2) | - | . |
| Yes | 3,375 | 6.5% | 966 | 28.6 (27.1 - 30.2) | 1.17 (1.05 - 1.32) | 0.007 |
| **Emergency Room Stay** |  |  |  |  |  |  |
| No (Reference) | 5,881 | 11.4% | 1,499 | 25.5 (24.4 - 26.6) | - | . |
| Yes | 45,737 | 88.6% | 10,473 | 22.9 (22.5 - 23.3) | 1.00 (0.91 - 1.10) | 0.998 |
| **COVID-19 As a Reason for Admission** |  |  |  |  |  |  |
| No (Reference) | 24,419 | 47.3% | 6,011 | 24.6 (24.1 - 25.2) | - | . |
| Yes | 27,199 | 52.7% | 5,961 | 21.9 (21.4 - 22.4) | 1.08 (1.02 - 1.14) | 0.012 |
| **Time Since Most Recent COVID-19 Diagnosis in the 30 Days Prior to Admission** |  |  |  |  |  |  |
| 0 days (Reference)^c^ | 25,086 | 48.6% | 5,879 | 23.4 (22.9 - 24.0) | - | . |
| 1 - 3 days | 12,331 | 23.9% | 2,793 | 22.7 (21.9 - 23.4) | 0.99 (0.92 - 1.07) | 0.859 |
| 4 - 10 days | 11,433 | 22.1% | 2,695 | 23.6 (22.8 - 24.4) | 1.03 (0.96 - 1.11) | 0.397 |
| 11-30 days | 2,768 | 5.4% | 605 | 21.9 (20.3 - 23.5) | 0.79 (0.69 - 0.91) | 0.001 |
| **Thrombosis on Index** |  |  |  |  |  |  |
| No (Reference) | 44,667 | 86.5% | 9,087 | 20.3 (20.0 - 20.7) | - | . |
| Yes | 6,951 | 13.5% | 2,885 | 41.5 (40.3 - 42.7) | 1.30 (1.19 - 1.41) | <.0001 |
| **Hospital Ownership^a^** |  |  |  |  |  |  |
| Government (Reference) | 6,686 | 13.0% | 1,544 | 23.1 (22.1 - 24.1) | - | . |
| For Profit | 10,462 | 20.3% | 2,569 | 24.6 (23.7 - 25.4) | 0.80 (0.73 - 0.89) | <.0001 |
| Non-Profit | 34,362 | 66.6% | 7,839 | 22.8 (22.4 - 23.3) | 0.84 (0.77 - 0.92) | 0.000 |
| **Hospital Size^a^** |  |  |  |  |  |  |
| Small (< 100 beds) (Reference) | 6,211 | 12.0% | 1,053 | 17.0 (16.0 - 17.9) | - | . |
| Medium (100 - 500 beds) | 33,127 | 64.2% | 7,883 | 23.8 (23.3 - 24.3) | 1.18 (1.07 - 1.30) | 0.001 |
| Large (> 500 beds) | 9,822 | 19.0% | 2,491 | 25.4 (24.5 - 26.2) | 1.17 (1.04 - 1.31) | 0.007 |
| **ICU/CCU Stay** |  |  |  |  |  |  |
| No (Reference) | 20,261 | 39.3% | 1,184 | 5.8 (5.5 - 6.2) | - | . |
| Yes | 31,357 | 60.7% | 10,788 | 34.4 (33.9 - 34.9) | 2.91 (2.70 - 3.15) | <.0001 |
| **Mechanical Ventilation** |  |  |  |  |  |  |
| No (Reference) | 40,738 | 78.9% | 4,140 | 10.2 (9.9 - 10.5) | - | . |
| Less Than 96hrs | 3,425 | 6.6% | 2,489 | 72.7 (71.1 - 74.2) | 15.31 (13.93 - 16.84) | <.0001 |
| More Than 96hrs | 7,455 | 14.4% | 5,343 | 71.7 (70.6 - 72.7) | 12.75 (11.85 - 13.72) | <.0001 |
| **Tocilizumab** |  |  |  |  |  |  |
| No (Reference) | 49,644 | 96.2% | 11,212 | 22.6 (22.2 - 23.0) | - | . |
| Yes | 1,974 | 3.8% | 760 | 38.5 (36.4 - 40.7) | 1.49 (1.31 - 1.70) | <.0001 |
| **Baricitinib** |  |  |  |  |  |  |
| No (Reference) | 51,061 | 98.9% | 11,785 | 23.1 (22.7 - 23.4) | - | . |
| Yes | 557 | 1.1% | 187 | 33.6 (29.7 - 37.7) | 1.89 (1.47 - 2.43) | <.0001 |
| **Remdesivir** |  |  |  |  |  |  |
| No (Reference) | 11,810 | 22.9% | 2,883 | 24.4 (23.6 - 25.2) | - | . |
| Yes | 39,808 | 77.1% | 9,089 | 22.8 (22.4 - 23.2) | 0.96 (0.89 - 1.03) | 0.273 |
| **Charlson Comorbidity Index (CCI)** |  |  |  |  |  |  |
| 0 Reference) | 10,173 | 19.7% | 1,563 | 15.4 (14.7 - 16.1) | - | . |
| 1 | 13,239 | 25.6% | 2,570 | 19.4 (18.7 - 20.1) | 1.19 (1.07 - 1.32) | 0.001 |
| 2 | 9,686 | 18.8% | 2,348 | 24.2 (23.4 - 25.1) | 1.35 (1.20 - 1.53) | <.0001 |
| 3 – 4 | 10,915 | 21.1% | 3,041 | 27.9 (27.0 - 28.7) | 1.34 (1.17 - 1.54) | <.0001 |
| 5+ | 7,605 | 14.7% | 2,450 | 32.2 (31.2 - 33.3) | 1.58 (1.31 - 1.89) | <.0001 |
| **90-Day Comorbidity History** |  |  |  |  |  |  |
| **Thrombosis History** |  |  |  |  |  |  |
| No (Reference) | 50,309 | 97.5% | 11,564 | 23.0 (22.6 - 23.4) | - | . |
| Yes | 1,309 | 2.5% | 408 | 31.2 (28.7 - 33.8) | 0.98 (0.83 - 1.17) | 0.839 |
| **ESRD History** |  |  |  |  |  |  |
| No (Reference) | 49,751 | 96.4% | 11,438 | 23.0 (22.6 - 23.4) | - | . |
| Yes | 1,867 | 3.6% | 534 | 28.6 (26.6 - 30.7) | 0.92 (0.77 - 1.11) | 0.381 |
| **Elixhauser Comorbidity History** |  |  |  |  |  |  |
| **Alcohol Abuse** |  |  |  |  |  |  |
| No (Reference) | 50,744 | 98.3% | 11,724 | 23.1 (22.7 - 23.5) | - | . |
| Yes | 874 | 1.7% | 248 | 28.4 (25.4 - 31.5) | 0.91 (0.74 - 1.12) | 0.369 |
| **Blood Loss Anemia** |  |  |  |  |  |  |
| No (Reference) | 51,387 | 99.6% | 11,891 | 23.1 (22.8 - 23.5) | - | . |
| Yes | 231 | 0.4% | 81 | 35.1 (29.0 - 41.6) | 1.45 (1.00 - 2.12) | 0.052 |
| **Cardiac Arrhythmia** |  |  |  |  |  |  |
| No (Reference) | 34,676 | 67.2% | 6,255 | 18.0 (17.6 - 18.4) | - | . |
| Yes | 16,942 | 32.8% | 5,717 | 33.7 (33.0 - 34.5) | 1.42 (1.34 - 1.51) | <.0001 |
| **Chronic Pulmonary Disease** |  |  |  |  |  |  |
| No (Reference) | 34,927 | 67.7% | 7,915 | 22.7 (22.2 - 23.1) | - | . |
| Yes | 16,691 | 32.3% | 4,057 | 24.3 (23.7 - 25.0) | 1.03 (0.96 - 1.10) | 0.481 |
| **Coagulopathy** |  |  |  |  |  |  |
| No (Reference) | 42,695 | 82.7% | 8,647 | 20.3 (19.9 - 20.6) | - | . |
| Yes | 8,923 | 17.3% | 3,325 | 37.3 (36.3 - 38.3) | 1.33 (1.24 - 1.43) | <.0001 |
| **Congestive Heart Failure** |  |  |  |  |  |  |
| No (Reference) | 39,489 | 76.5% | 8,308 | 21.0 (20.6 - 21.4) | - | . |
| Yes | 12,129 | 23.5% | 3,664 | 30.2 (29.4 - 31.0) | 1.00 (0.92 - 1.09) | 0.982 |
| **Deficiency Anemia** |  |  |  |  |  |  |
| No (Reference) | 49,405 | 95.7% | 11,403 | 23.1 (22.7 - 23.5) | - | . |
| Yes | 2,213 | 4.3% | 569 | 25.7 (23.9 - 27.6) | 0.89 (0.78 - 1.02) | 0.091 |
| **Depression** |  |  |  |  |  |  |
| No (Reference) | 43,525 | 84.3% | 10,165 | 23.4 (23.0 - 23.8) | - | . |
| Yes | 8,093 | 15.7% | 1,807 | 22.3 (21.4 - 23.3) | 1.07 (0.99 - 1.16) | 0.094 |
| **Diabetes** |  |  |  |  |  |  |
| No (Reference) | 27,251 | 52.8% | 5,780 | 21.2 (20.7 - 21.7) | - | . |
| Yes | 24,367 | 47.2% | 6,192 | 25.4 (24.9 - 26.0) | 0.90 (0.83 - 0.97) | 0.006 |
| **Fluid and Electrolyte Disorder** |  |  |  |  |  |  |
| No (Reference) | 20,024 | 38.8% | 2,258 | 11.3 (10.8 - 11.7) | - | . |
| Yes | 31,594 | 61.2% | 9,714 | 30.7 (30.2 - 31.3) | 1.61 (1.51 - 1.73) | <.0001 |
| **Hypertension (uncomplicated)** |  |  |  |  |  |  |
| No (Reference) | 27,327 | 52.9% | 7,060 | 25.8 (25.3 - 26.4) | - | . |
| Yes | 24,291 | 47.1% | 4,912 | 20.2 (19.7 - 20.7) | 0.98 (0.91 - 1.05) | 0.563 |
| **Hypertension (complicated)** |  |  |  |  |  |  |
| No (Reference) | 32,758 | 63.5% | 6,534 | 19.9 (19.5 - 20.4) | - | . |
| Yes | 18,860 | 36.5% | 5,438 | 28.8 (28.2 - 29.5) | 1.09 (0.99 - 1.20) | 0.071 |
| **Liver Disease** |  |  |  |  |  |  |
| No (Reference) | 48,663 | 94.3% | 10,771 | 22.1 (21.8 - 22.5) | - | . |
| Yes | 2,955 | 5.7% | 1,201 | 40.6 (38.9 - 42.4) | 1.47 (1.31 - 1.64) | <.0001 |
| **Metastatic Cancer** |  |  |  |  |  |  |
| No (Reference) | 50,863 | 98.5% | 11,753 | 23.1 (22.7 - 23.5) | - | . |
| Yes | 755 | 1.5% | 219 | 29.0 (25.8 - 32.4) | 1.34 (1.03 - 1.73) | 0.027 |
| **Obesity** |  |  |  |  |  |  |
| No (Reference) | 35,347 | 68.5% | 7,893 | 22.3 (21.9 - 22.8) | - | . |
| Yes | 16,271 | 31.5% | 4,079 | 25.1 (24.4 - 25.7) | 1.04 (0.98 - 1.11) | 0.230 |
| **Other Neurological Disorders** |  |  |  |  |  |  |
| No (Reference) | 41,091 | 79.6% | 8,128 | 19.8 (19.4 - 20.2) | - | . |
| Yes | 10,527 | 20.4% | 3,844 | 36.5 (35.6 - 37.4) | 1.23 (1.15 - 1.32) | <.0001 |
| **Paralysis** |  |  |  |  |  |  |
| No (Reference) | 50,863 | 98.5% | 11,689 | 23.0 (22.6 - 23.3) | - | . |
| Yes | 755 | 1.5% | 283 | 37.5 (34.0 - 41.1) | 0.93 (0.75 - 1.16) | 0.537 |
| **Peptic Ulcer Disease** |  |  |  |  |  |  |
| No (Reference) | 51,494 | 99.8% | 11,936 | 23.2 (22.8 - 23.5) | - | . |
| Yes | 124 | 0.2% | 36 | 29.0 (21.4 - 38.0) | 0.55 (0.32 - 0.93) | 0.025 |
| **Peripheral Vascular Disorder** |  |  |  |  |  |  |
| No (Reference) | 47,850 | 92.7% | 10,961 | 22.9 (22.5 - 23.3) | - | . |
| Yes | 3,768 | 7.3% | 1,011 | 26.8 (25.4 - 28.3) | 0.98 (0.88 - 1.09) | 0.717 |
| **Pulmonary Circulation Disorder** |  |  |  |  |  |  |
| No (Reference) | 49,753 | 96.4% | 11,359 | 22.8 (22.5 - 23.2) | - | . |
| Yes | 1,865 | 3.6% | 613 | 32.9 (30.7 - 35.1) | 1.11 (0.96 - 1.30) | 0.163 |
| **Renal Failure** |  |  |  |  |  |  |
| No (Reference) | 43,051 | 83.4% | 9,319 | 21.6 (21.3 - 22.0) | - | . |
| Yes | 8,567 | 16.6% | 2,653 | 31.0 (30.0 - 32.0) | 1.04 (0.94 - 1.16) | 0.436 |
| **Rheumatoid Arthritis/Collagen Vascular Diseases (CVD)** |  |  |  |  |  |  |
| No (Reference) | 49,068 | 95.1% | 11,335 | 23.1 (22.7 - 23.5) | - | . |
| Yes | 2,550 | 4.9% | 637 | 25.0 (23.3 - 26.7) | 1.18 (1.03 - 1.34) | 0.014 |
| **Solid Tumor** |  |  |  |  |  |  |
| No (Reference) | 49,724 | 96.3% | 11,468 | 23.1 (22.7 - 23.4) | - | . |
| Yes | 1,894 | 3.7% | 504 | 26.6 (24.6 - 28.7) | 1.04 (0.88 - 1.23) | 0.645 |
| **Valvular Disease** |  |  |  |  |  |  |
| No (Reference) | 48,595 | 94.1% | 11,166 | 23.0 (22.6 - 23.4) | - | . |
| Yes | 3,023 | 5.9% | 806 | 26.7 (25.1 - 28.3) | 1.01 (0.90 - 1.14) | 0.809 |
| **Weight Loss** |  |  |  |  |  |  |
| No (Reference) | 46,699 | 90.5% | 10,064 | 21.6 (21.2 - 21.9) | - | . |
| Yes | 4,919 | 9.5% | 1,908 | 38.8 (37.4 - 40.2) | 1.18 (1.08 - 1.29) | 0.000 |
| ^a^ Patients with missing values for ADI, Urban/Rural, Hospital Size and Ownership were dropped from the multivariate analysis. | | | | | | |
| ^b^ The South US census region includes counts for "Other" territories to avoid identification due to low counts. | | | | | | |
| ^c^ Patients have not had COVID-19 diagnosis recorded in the 30 days prior to admission. | | | | | | |
| **Notes:** Model evaluation metrics were ROC Area Under the Curve (0.8775); Pearson Chi Sq Goodness of Fit test p value(1); Deviance Goodness of Fit test p value(1). | | | | | |  |
| Intravenous Immunoglobulin/Hyperimmune Globulin Group has been omitted to avoid identification due to low counts. | | | | | | |

| **Supplementary Table 10.** Inpatient (IP) Mortality Rates and Corresponding Adjusted Odds Ratios (ORs) by Patient and Hospital Characteristics Among Recipients of COVID-19 Convalescent Plasma (CCP) Aged 80+ | | | | | |  |
| --- | --- | --- | --- | --- | --- | --- |
| **Patient and Hospital Characteristics** | **CCP Transfusion Stays #** | **CCP Transfusion Stays  %** | **IP Mortality  #** | **Crude IP Mortality Rate per 100  (95% CIs)** | **Adjusted ORs (95% CIs)** | **P-value** |
| **Overall** | 30,133 | 100.0% | 8,904 | 29.5 (29.0 - 30.1) | - | . |
| **Age (continuous)** | 30,133 | 100.0% | 8,904 | 29.5 (29.0 - 30.1) | 1.05 (1.04 - 1.05) | <.0001 |
| **Sex** |  |  |  |  |  |  |
| Female (Reference) | 14,597 | 48.4% | 3,785 | 25.9 (25.2 - 26.7) | - | . |
| Male | 15,536 | 51.6% | 5,119 | 32.9 (32.2 - 33.7) | 1.26 (1.19 - 1.35) | <.0001 |
| **ADI^a^** |  |  |  |  |  |  |
| 81 - 100 (Reference) | 4,247 | 14.1% | 1,336 | 31.5 (30.1 - 32.9) | - | . |
| 61 – 80 | 6,196 | 20.6% | 1,857 | 30.0 (28.8 - 31.1) | 1.02 (0.92 - 1.13) | 0.720 |
| 41 – 60 | 6,582 | 21.8% | 1,836 | 27.9 (26.8 - 29.0) | 0.92 (0.83 - 1.02) | 0.097 |
| 21 – 40 | 6,094 | 20.2% | 1,711 | 28.1 (27.0 - 29.2) | 0.85 (0.77 - 0.95) | 0.003 |
| 0 – 20 | 4,446 | 14.8% | 1,317 | 29.6 (28.3 - 31.0) | 0.87 (0.78 - 0.98) | 0.022 |
| **Medicaid Eligibility** |  |  |  |  |  |  |
| No (Reference) | 23,508 | 78.0% | 6,665 | 28.4 (27.8 - 28.9) | - | . |
| Yes | 6,625 | 22.0% | 2,239 | 33.8 (32.7 - 35.0) | 1.12 (1.03 - 1.22) | 0.006 |
| **Region of Residence^b^** |  |  |  |  |  |  |
| South (Reference) | 15,738 | 52.2% | 4,568 | 29.0 (28.3 - 29.7) | - | . |
| Midwest | 7,192 | 23.9% | 1,976 | 27.5 (26.4 - 28.5) | 0.99 (0.91 - 1.07) | 0.821 |
| West | 3,845 | 12.8% | 1,313 | 34.1 (32.7 - 35.7) | 1.23 (1.11 - 1.35) | <.0001 |
| Northeast | 3,358 | 11.1% | 1,047 | 31.2 (29.6 - 32.8) | 1.29 (1.16 - 1.44) | <.0001 |
| **Urban/Rural^a^** |  |  |  |  |  |  |
| Urban (Reference) | 21,250 | 70.5% | 6,185 | 29.1 (28.5 - 29.7) | - | . |
| Rural | 6,574 | 21.8% | 1,957 | 29.8 (28.7 - 30.9) | 1.18 (1.09 - 1.29) | <.0001 |
| **Immunocompromised Status** |  |  |  |  |  |  |
| No (Reference) | 27,859 | 92.5% | 8,181 | 29.4 (28.8 - 29.9) | - | . |
| Yes | 2,274 | 7.5% | 723 | 31.8 (29.9 - 33.8) | 1.14 (0.99 - 1.31) | 0.079 |
| **Nursing Home Stay** |  |  |  |  |  |  |
| No (Reference) | 26,270 | 87.2% | 7,541 | 28.7 (28.2 - 29.3) | - | . |
| Yes | 3,863 | 12.8% | 1,363 | 35.3 (33.8 - 36.8) | 1.21 (1.10 - 1.33) | <.0001 |
| **Emergency Room Stay** |  |  |  |  |  |  |
| No (Reference) | 2,924 | 9.7% | 953 | 32.6 (30.9 - 34.3) | - | . |
| Yes | 27,209 | 90.3% | 7,951 | 29.2 (28.7 - 29.8) | 0.88 (0.78 - 0.98) | 0.019 |
| **COVID-19 As a Reason for Admission** |  |  |  |  |  |  |
| No (Reference) | 14,698 | 48.8% | 4,597 | 31.3 (30.5 - 32.0) | - | . |
| Yes | 15,435 | 51.2% | 4,307 | 27.9 (27.2 - 28.6) | 0.99 (0.93 - 1.05) | 0.793 |
| **Time Since Most Recent COVID-19 Diagnosis in the 30 Days Prior to Admission** |  |  |  |  |  |  |
| 0 days (Reference)^c^ | 15,896 | 52.8% | 4,625 | 29.1 (28.4 - 29.8) | - | . |
| 1 - 3 days | 7,066 | 23.4% | 1,998 | 28.3 (27.2 - 29.3) | 0.96 (0.89 - 1.04) | 0.286 |
| 4 - 10 days | 5,400 | 17.9% | 1,778 | 32.9 (31.7 - 34.2) | 1.13 (1.04 - 1.23) | 0.003 |
| 11-30 days | 1,771 | 5.9% | 503 | 28.4 (26.3 - 30.6) | 0.86 (0.75 - 0.98) | 0.028 |
| **Thrombosis on Index** |  |  |  |  |  |  |
| No (Reference) | 25,555 | 84.8% | 6,973 | 27.3 (26.7 - 27.8) | - | . |
| Yes | 4,578 | 15.2% | 1,931 | 42.2 (40.7 - 43.6) | 1.41 (1.29 - 1.54) | <.0001 |
| **Hospital Ownership^a^** |  |  |  |  |  |  |
| Government (Reference) | 3,775 | 12.5% | 1,109 | 29.4 (27.9 - 30.9) | - | . |
| For Profit | 6,036 | 20.0% | 1,785 | 29.6 (28.4 - 30.7) | 0.77 (0.69 - 0.86) | <.0001 |
| Non-Profit | 20,278 | 67.3% | 5,998 | 29.6 (29.0 - 30.2) | 0.88 (0.80 - 0.97) | 0.007 |
| **Hospital Size^a^** |  |  |  |  |  |  |
| Small (< 100 beds) (Reference) | 3,469 | 11.5% | 842 | 24.3 (22.9 - 25.7) | - | . |
| Medium (100 - 500 beds) | 19,378 | 64.3% | 5,857 | 30.2 (29.6 - 30.9) | 1.11 (1.00 - 1.24) | 0.047 |
| Large (> 500 beds) | 5,816 | 19.3% | 1,786 | 30.7 (29.5 - 31.9) | 1.04 (0.92 - 1.17) | 0.527 |
| **ICU/CCU Stay** |  |  |  |  |  |  |
| No (Reference) | 13,092 | 43.4% | 1,960 | 15.0 (14.4 - 15.6) | - | . |
| Yes | 17,041 | 56.6% | 6,944 | 40.7 (40.0 - 41.5) | 2.57 (2.40 - 2.75) | <.0001 |
| **Mechanical Ventilation** |  |  |  |  |  |  |
| No (Reference) | 26,746 | 88.8% | 6,190 | 23.1 (22.6 - 23.7) | - | . |
| Less Than 96hrs | 1,406 | 4.7% | 1,110 | 78.9 (76.7 - 81.0) | 8.97 (7.74 - 10.39) | <.0001 |
| More Than 96hrs | 1,981 | 6.6% | 1,604 | 81.0 (79.2 - 82.7) | 8.69 (7.62 - 9.91) | <.0001 |
| **Tocilizumab** |  |  |  |  |  |  |
| No (Reference) | 29,342 | 97.4% | 8,509 | 29.0 (28.5 - 29.5) | - | . |
| Yes | 791 | 2.6% | 395 | 49.9 (46.4 - 53.5) | 1.98 (1.67 - 2.36) | <.0001 |
| **Baricitinib** |  |  |  |  |  |  |
| No (Reference) | 29,908 | 99.3% | 8,837 | 29.5 (29.0 - 30.1) | - | . |
| Yes | 225 | 0.7% | 67 | 29.8 (24.0 - 36.3) | 0.97 (0.68 - 1.38) | 0.878 |
| **Remdesivir** |  |  |  |  |  |  |
| No (Reference) | 8,103 | 26.9% | 2,548 | 31.4 (30.4 - 32.5) | - | . |
| Yes | 22,030 | 73.1% | 6,356 | 28.9 (28.3 - 29.5) | 0.92 (0.86 - 0.98) | 0.016 |
| **Charlson Comorbidity Index (CCI)** |  |  |  |  |  |  |
| 0 (Reference) | 4,105 | 13.6% | 962 | 23.4 (22.2 - 24.8) | - | . |
| 1 | 7,264 | 24.1% | 1,851 | 25.5 (24.5 - 26.5) | 1.01 (0.90 - 1.13) | 0.876 |
| 2 | 6,254 | 20.8% | 1,825 | 29.2 (28.1 - 30.3) | 1.09 (0.97 - 1.23) | 0.168 |
| 3 – 4 | 7,860 | 26.1% | 2,595 | 33.0 (32.0 - 34.1) | 1.12 (0.98 - 1.27) | 0.099 |
| 5+ | 4,650 | 15.4% | 1,671 | 35.9 (34.6 - 37.3) | 1.06 (0.90 - 1.25) | 0.502 |
| **90-Day Comorbidity History** |  |  |  |  |  |  |
| **Thrombosis History** |  |  |  |  |  |  |
| No (Reference) | 29,125 | 96.7% | 8,527 | 29.3 (28.8 - 29.8) | - | . |
| Yes | 1,008 | 3.3% | 377 | 37.4 (34.4 - 40.5) | 1.01 (0.85 - 1.19) | 0.915 |
| **ESRD History** |  |  |  |  |  |  |
| No (Reference) | 29,555 | 98.1% | 8,720 | 29.5 (29.0 - 30.0) | - | . |
| Yes | 578 | 1.9% | 184 | 31.8 (28.1 - 35.8) | 0.76 (0.59 - 0.99) | 0.041 |
| **Elixhauser Comorbidity History** |  |  |  |  |  |  |
| **Cardiac Arrhythmia** |  |  |  |  |  |  |
| No (Reference) | 16,888 | 56.0% | 4,287 | 25.4 (24.7 - 26.0) | - | . |
| Yes | 13,245 | 44.0% | 4,617 | 34.9 (34.0 - 35.7) | 1.26 (1.18 - 1.34) | <.0001 |
| **Coagulopathy** |  |  |  |  |  |  |
| No (Reference) | 24,785 | 82.3% | 6,788 | 27.4 (26.8 - 27.9) | - | . |
| Yes | 5,348 | 17.7% | 2,116 | 39.6 (38.3 - 40.9) | 1.24 (1.15 - 1.34) | <.0001 |
| **Congestive Heart Failure** |  |  |  |  |  |  |
| No (Reference) | 19,689 | 65.3% | 5,401 | 27.4 (26.8 - 28.1) | - | . |
| Yes | 10,444 | 34.7% | 3,503 | 33.5 (32.6 - 34.5) | 1.03 (0.95 - 1.12) | 0.516 |
| **Depression** |  |  |  |  |  |  |
| No (Reference) | 25,756 | 85.5% | 7,691 | 29.9 (29.3 - 30.4) | - | . |
| Yes | 4,377 | 14.5% | 1,213 | 27.7 (26.4 - 29.1) | 0.98 (0.89 - 1.07) | 0.596 |
| **Diabetes** |  |  |  |  |  |  |
| No (Reference) | 18,771 | 62.3% | 5,288 | 28.2 (27.5 - 28.8) | - | . |
| Yes | 11,362 | 37.7% | 3,616 | 31.8 (31.0 - 32.7) | 1.01 (0.94 - 1.09) | 0.719 |
| **Fluid and Electrolyte Disorder** |  |  |  |  |  |  |
| No (Reference) | 10,878 | 36.1% | 2,156 | 19.8 (19.1 - 20.6) | - | . |
| Yes | 19,255 | 63.9% | 6,748 | 35.0 (34.4 - 35.7) | 1.49 (1.39 - 1.59) | <.0001 |
| **Hypertension (uncomplicated)** |  |  |  |  |  |  |
| No (Reference) | 18,013 | 59.8% | 5,756 | 32.0 (31.3 - 32.6) | - | . |
| Yes | 12,120 | 40.2% | 3,148 | 26.0 (25.2 - 26.8) | 0.92 (0.85 - 1.00) | 0.052 |
| **Hypertension (complicated)** |  |  |  |  |  |  |
| No (Reference) | 14,934 | 49.6% | 3,905 | 26.1 (25.4 - 26.9) | - | . |
| Yes | 15,199 | 50.4% | 4,999 | 32.9 (32.1 - 33.6) | 1.04 (0.94 - 1.14) | 0.426 |
| **Liver Disease** |  |  |  |  |  |  |
| No (Reference) | 29,260 | 97.1% | 8,519 | 29.1 (28.6 - 29.6) | - | . |
| Yes | 873 | 2.9% | 385 | 44.1 (40.8 - 47.5) | 1.30 (1.09 - 1.55) | 0.003 |
| **Lymphoma** |  |  |  |  |  |  |
| No (Reference) | 29,725 | 98.6% | 8,764 | 29.5 (29.0 - 30.0) | - | . |
| Yes | 408 | 1.4% | 140 | 34.3 (29.8 - 39.2) | 1.13 (0.85 - 1.49) | 0.401 |
| **Obesity** |  |  |  |  |  |  |
| No (Reference) | 25,934 | 86.1% | 7,592 | 29.3 (28.7 - 29.8) | - | . |
| Yes | 4,199 | 13.9% | 1,312 | 31.2 (29.8 - 32.7) | 1.05 (0.96 - 1.15) | 0.305 |
| **Other Neurological Disorders** |  |  |  |  |  |  |
| No (Reference) | 21,448 | 71.2% | 5,559 | 25.9 (25.3 - 26.5) | - | . |
| Yes | 8,685 | 28.8% | 3,345 | 38.5 (37.5 - 39.5) | 1.39 (1.30 - 1.49) | <.0001 |
| **Paralysis** |  |  |  |  |  |  |
| No (Reference) | 29,820 | 99.0% | 8,760 | 29.4 (28.9 - 29.9) | - | . |
| Yes | 313 | 1.0% | 144 | 46.0 (40.4 - 51.7) | 1.16 (0.87 - 1.54) | 0.322 |
| **Peptic Ulcer Disease** |  |  |  |  |  |  |
| No (Reference) | 30,077 | 99.8% | 8,893 | 29.6 (29.1 - 30.1) | - | . |
| Yes | 56 | 0.2% | 11 | 19.6 (10.7 - 32.8) | 0.37 (0.15 - 0.88) | 0.024 |
| **Pulmonary Circulation Disorder** |  |  |  |  |  |  |
| No (Reference) | 29,190 | 96.9% | 8,575 | 29.4 (28.9 - 29.9) | - | . |
| Yes | 943 | 3.1% | 329 | 34.9 (31.9 - 38.0) | 0.96 (0.80 - 1.15) | 0.636 |
| **Renal Failure** |  |  |  |  |  |  |
| No (Reference) | 23,869 | 79.2% | 6,682 | 28.0 (27.4 - 28.6) | - | . |
| Yes | 6,264 | 20.8% | 2,222 | 35.5 (34.3 - 36.7) | 1.09 (0.99 - 1.20) | 0.072 |
| **Valvular Disease** |  |  |  |  |  |  |
| No (Reference) | 26,743 | 88.7% | 7,850 | 29.4 (28.8 - 29.9) | - | . |
| Yes | 3,390 | 11.3% | 1,054 | 31.1 (29.5 - 32.7) | 0.91 (0.83 - 1.00) | 0.056 |
| **Weight Loss** |  |  |  |  |  |  |
| No (Reference) | 26,197 | 86.9% | 7,400 | 28.2 (27.7 - 28.8) | - | . |
| Yes | 3,936 | 13.1% | 1,504 | 38.2 (36.7 - 39.8) | 1.12 (1.02 - 1.22) | 0.014 |
| ^a^ Patients with missing values for ADI, Urban/Rural, Hospital Size and Ownership were dropped from the multivariate analysis. | | | | | | |
| ^b^ The South US census region includes counts for "Other" territories to avoid identification due to low counts. | | | | | | |
| ^c^ Patients have not had COVID-19 diagnosis recorded in the 30 days prior to admission. | | | | | | |
| **Notes:** Model evaluation metrics were ROC Area Under the Curve (0.7786); Pearson Chi Sq Goodness of Fit test p value (0.8398); Deviance Goodness of Fit test p value (0.9459). | | | | | | |
| Intravenous Immunoglobulin/Hyperimmune Globulin Group has been omitted to avoid identification due to low counts. | | | | |  |  |

| **Supplementary Table 11.** Inpatient (IP) Mortality Rates and Corresponding Adjusted Odds Ratios (ORs) by Patient and Hospital Characteristics Among Male Recipients of COVID-19 Convalescent Plasma (CCP) | | | | | |  |
| --- | --- | --- | --- | --- | --- | --- |
| **Patient and Hospital Characteristics** | **CCP Transfusion Stays #** | **CCP Transfusion Stays  %** | **IP Mortality  #** | **Crude IP Mortality Rate per 100  (95% CIs)** | **Adjusted ORs (95% CIs)** | **P-value** |
| **Overall** | 44,413 | 100.0% | 12,397 | 27.9 (27.5 - 28.3) | - | . |
| **Age (continuous)** | 44,413 | 100.0% | 12,397 | 27.9 (27.5 - 28.3) | 1.07 (1.06 - 1.07) | <.0001 |
| **ADI^a^** |  |  |  |  |  |  |
| 81 - 100 (Reference) | 6,373 | 14.3% | 1,945 | 30.5 (29.4 - 31.7) | - | . |
| 61 - 80 | 9,293 | 20.9% | 2,660 | 28.6 (27.7 - 29.6) | 1.00 (0.91 - 1.09) | 0.983 |
| 41 – 60 | 9,982 | 22.5% | 2,655 | 26.6 (25.7 - 27.5) | 0.92 (0.84 - 1.00) | 0.063 |
| 21 – 40 | 8,860 | 19.9% | 2,370 | 26.7 (25.8 - 27.7) | 0.86 (0.79 - 0.95) | 0.003 |
| 0 – 20 | 5,699 | 12.8% | 1,531 | 26.9 (25.7 - 28.0) | 0.79 (0.71 - 0.88) | <.0001 |
| **Medicaid Eligibility** |  |  |  |  |  |  |
| No (Reference) | 37,097 | 83.5% | 10,048 | 27.1 (26.6 - 27.5) | - | . |
| Yes | 7,316 | 16.5% | 2,349 | 32.1 (31.0 - 33.2) | 1.09 (1.00 - 1.19) | 0.041 |
| **Region of Residence^b^** |  |  |  |  |  |  |
| South (Reference) | 23,474 | 52.9% | 6,517 | 27.8 (27.2 - 28.3) | - | . |
| Midwest | 10,927 | 24.6% | 2,811 | 25.7 (24.9 - 26.6) | 0.93 (0.86 - 1.00) | 0.050 |
| West | 5,868 | 13.2% | 1,875 | 32.0 (30.8 - 33.2) | 1.10 (1.01 - 1.21) | 0.029 |
| Northeast | 4,144 | 9.3% | 1,194 | 28.8 (27.4 - 30.2) | 1.11 (1.00 - 1.23) | 0.053 |
| **Urban/Rural^a^** |  |  |  |  |  |  |
| Urban (Reference) | 29,920 | 67.4% | 8,382 | 28.0 (27.5 - 28.5) | - | . |
| Rural | 10,657 | 24.0% | 2,891 | 27.1 (26.3 - 28.0) | 1.09 (1.02 - 1.18) | 0.018 |
| **Immunocompromised Status** |  |  |  |  |  |  |
| No (Reference) | 39,804 | 89.6% | 11,015 | 27.7 (27.2 - 28.1) | - | . |
| Yes | 4,609 | 10.4% | 1,382 | 30.0 (28.7 - 31.3) | 1.11 (1.01 - 1.22) | 0.037 |
| **Nursing Home Stay** |  |  |  |  |  |  |
| No (Reference) | 41,052 | 92.4% | 11,235 | 27.4 (26.9 - 27.8) | - | . |
| Yes | 3,361 | 7.6% | 1,162 | 34.6 (33.0 - 36.2) | 1.16 (1.05 - 1.29) | 0.005 |
| **Emergency Room Stay** |  |  |  |  |  |  |
| No (Reference) | 5,020 | 11.3% | 1,523 | 30.3 (29.1 - 31.6) | - | . |
| Yes | 39,393 | 88.7% | 10,874 | 27.6 (27.2 - 28.0) | 0.94 (0.85 - 1.04) | 0.220 |
| **COVID-19 As a Reason for Admission** |  |  |  |  |  |  |
| No (Reference) | 21,313 | 48.0% | 6,301 | 29.6 (29.0 - 30.2) | - | . |
| Yes | 23,100 | 52.0% | 6,096 | 26.4 (25.8 - 27.0) | 1.01 (0.95 - 1.07) | 0.773 |
| **Time Since Most Recent COVID-19 Diagnosis in the 30 Days Prior to Admission** |  |  |  |  |  |  |
| 0 days (Reference)^c^ | 22,253 | 50.1% | 6,268 | 28.2 (27.6 - 28.8) | - | . |
| 1 - 3 days | 10,403 | 23.4% | 2,821 | 27.1 (26.3 - 28.0) | 0.99 (0.93 - 1.07) | 0.872 |
| 4 - 10 days | 9,253 | 20.8% | 2,648 | 28.6 (27.7 - 29.6) | 1.08 (1.00 - 1.16) | 0.049 |
| 11-30 days | 2,504 | 5.6% | 660 | 26.4 (24.6 - 28.1) | 0.83 (0.73 - 0.94) | 0.004 |
| **Thrombosis on Index** |  |  |  |  |  |  |
| No (Reference) | 37,496 | 84.4% | 9,347 | 24.9 (24.5 - 25.4) | - | . |
| Yes | 6,917 | 15.6% | 3,050 | 44.1 (42.9 - 45.3) | 1.34 (1.24 - 1.46) | <.0001 |
| **Hospital Ownership^a^** |  |  |  |  |  |  |
| Government (Reference) | 5,562 | 12.5% | 1,537 | 27.6 (26.5 - 28.8) | - | . |
| For Profit | 8,737 | 19.7% | 2,530 | 29.0 (28.0 - 29.9) | 0.80 (0.73 - 0.89) | <.0001 |
| Non-Profit | 30,019 | 67.6% | 8,310 | 27.7 (27.2 - 28.2) | 0.88 (0.81 - 0.96) | 0.006 |
| **Hospital Size^a^** |  |  |  |  |  |  |
| Small (< 100 beds) (Reference) | 5,226 | 11.8% | 1,098 | 21.0 (19.9 - 22.1) | - | . |
| Medium (100 - 500 beds) | 28,470 | 64.1% | 8,165 | 28.7 (28.2 - 29.2) | 1.18 (1.07 - 1.29) | 0.001 |
| Large (> 500 beds) | 8,579 | 19.3% | 2,566 | 29.9 (28.9 - 30.9) | 1.18 (1.05 - 1.32) | 0.004 |
| **ICU/CCU Stay** |  |  |  |  |  |  |
| No (Reference) | 16,959 | 38.2% | 1,692 | 10.0 (9.5 - 10.4) | - | . |
| Yes | 27,454 | 61.8% | 10,705 | 39.0 (38.4 - 39.6) | 2.71 (2.53 - 2.90) | <.0001 |
| **Mechanical Ventilation** |  |  |  |  |  |  |
| No (Reference) | 35,587 | 80.1% | 5,728 | 16.1 (15.7 - 16.5) | - | . |
| Less Than 96hrs | 3,040 | 6.8% | 2,297 | 75.6 (74.0 - 77.1) | 13.76 (12.42 - 15.25) | <.0001 |
| More Than 96hrs | 5,786 | 13.0% | 4,372 | 75.6 (74.4 - 76.7) | 12.42 (11.43 - 13.49) | <.0001 |
| **Tocilizumab** |  |  |  |  |  |  |
| No (Reference) | 42,812 | 96.4% | 11,719 | 27.4 (27.0 - 27.8) | - | . |
| Yes | 1,601 | 3.6% | 678 | 42.3 (39.9 - 44.8) | 1.52 (1.33 - 1.75) | <.0001 |
| **Baricitinib** |  |  |  |  |  |  |
| No (Reference) | 43,991 | 99.0% | 12,245 | 27.8 (27.4 - 28.3) | - | . |
| Yes | 422 | 1.0% | 152 | 36.0 (31.5 - 40.8) | 1.73 (1.33 - 2.25) | <.0001 |
| **Remdesivir** |  |  |  |  |  |  |
| No (Reference) | 10,564 | 23.8% | 3,139 | 29.7 (28.8 - 30.6) | - | . |
| Yes | 33,849 | 76.2% | 9,258 | 27.4 (26.9 - 27.8) | 0.95 (0.89 - 1.01) | 0.120 |
| **Charlson Comorbidity Index (CCI)** |  |  |  |  |  |  |
| 0 (Reference) | 7,485 | 16.9% | 1,493 | 19.9 (19.1 - 20.9) | - | . |
| 1 | 10,626 | 23.9% | 2,537 | 23.9 (23.1 - 24.7) | 1.11 (1.00 - 1.22) | 0.052 |
| 2 | 8,529 | 19.2% | 2,418 | 28.4 (27.4 - 29.3) | 1.30 (1.16 - 1.45) | <.0001 |
| 3 – 4 | 10,589 | 23.8% | 3,409 | 32.2 (31.3 - 33.1) | 1.30 (1.16 - 1.47) | <.0001 |
| 5+ | 7,184 | 16.2% | 2,540 | 35.4 (34.3 - 36.5) | 1.41 (1.21 - 1.64) | <.0001 |
| **90-Day Comorbidity History** |  |  |  |  |  |  |
| **Thrombosis History** |  |  |  |  |  |  |
| No (Reference) | 43,133 | 97.1% | 11,958 | 27.7 (27.3 - 28.1) | - | . |
| Yes | 1,280 | 2.9% | 439 | 34.3 (31.7 - 37.0) | 0.89 (0.76 - 1.05) | 0.175 |
| **Elixhauser Comorbidity History** |  |  |  |  |  |  |
| **Blood Loss Anemia** |  |  |  |  |  |  |
| No (Reference) | 44,218 | 99.6% | 12,319 | 27.9 (27.4 - 28.3) | - | . |
| Yes | 195 | 0.4% | 78 | 40.0 (33.1 - 47.3) | 1.46 (0.99 - 2.13) | 0.054 |
| **Cardiac Arrhythmia** |  |  |  |  |  |  |
| No (Reference) | 26,730 | 60.2% | 5,961 | 22.3 (21.8 - 22.8) | - | . |
| Yes | 17,683 | 39.8% | 6,436 | 36.4 (35.7 - 37.1) | 1.32 (1.25 - 1.40) | <.0001 |
| **Coagulopathy** |  |  |  |  |  |  |
| No (Reference) | 35,832 | 80.7% | 8,977 | 25.1 (24.6 - 25.5) | - | . |
| Yes | 8,581 | 19.3% | 3,420 | 39.9 (38.8 - 40.9) | 1.25 (1.17 - 1.34) | <.0001 |
| **Congestive Heart Failure** |  |  |  |  |  |  |
| No (Reference) | 31,944 | 71.9% | 8,169 | 25.6 (25.1 - 26.1) | - | . |
| Yes | 12,469 | 28.1% | 4,228 | 33.9 (33.1 - 34.7) | 1.02 (0.94 - 1.10) | 0.608 |
| **Deficiency Anemia** |  |  |  |  |  |  |
| No (Reference) | 42,497 | 95.7% | 11,797 | 27.8 (27.3 - 28.2) | - | . |
| Yes | 1,916 | 4.3% | 600 | 31.3 (29.3 - 33.5) | 0.97 (0.85 - 1.10) | 0.605 |
| **Diabetes** |  |  |  |  |  |  |
| No (Reference) | 24,168 | 54.4% | 6,412 | 26.5 (26.0 - 27.1) | - | . |
| Yes | 20,245 | 45.6% | 5,985 | 29.6 (28.9 - 30.2) | 0.93 (0.87 - 1.00) | 0.039 |
| **Fluid and Electrolyte Disorder** |  |  |  |  |  |  |
| No (Reference) | 17,032 | 38.3% | 2,588 | 15.2 (14.7 - 15.7) | - | . |
| Yes | 27,381 | 61.7% | 9,809 | 35.8 (35.3 - 36.4) | 1.58 (1.48 - 1.68) | <.0001 |
| **Hypertension (uncomplicated)** |  |  |  |  |  |  |
| No (Reference) | 25,440 | 57.3% | 7,788 | 30.6 (30.0 - 31.2) | - | . |
| Yes | 18,973 | 42.7% | 4,609 | 24.3 (23.7 - 24.9) | 0.94 (0.87 - 1.01) | 0.084 |
| **Hypertension (complicated)** |  |  |  |  |  |  |
| No (Reference) | 25,231 | 56.8% | 6,105 | 24.2 (23.7 - 24.7) | - | . |
| Yes | 19,182 | 43.2% | 6,292 | 32.8 (32.1 - 33.5) | 1.07 (0.98 - 1.16) | 0.159 |
| **Hypothyroidism** |  |  |  |  |  |  |
| No (Reference) | 37,967 | 85.5% | 10,498 | 27.7 (27.2 - 28.1) | - | . |
| Yes | 6,446 | 14.5% | 1,899 | 29.5 (28.4 - 30.6) | 1.01 (0.94 - 1.10) | 0.739 |
| **Liver Disease** |  |  |  |  |  |  |
| No (Reference) | 42,314 | 95.3% | 11,474 | 27.1 (26.7 - 27.5) | - | . |
| Yes | 2,099 | 4.7% | 923 | 44.0 (41.8 - 46.1) | 1.41 (1.25 - 1.60) | <.0001 |
| **Other Neurological Disorders** |  |  |  |  |  |  |
| No (Reference) | 33,816 | 76.1% | 8,090 | 23.9 (23.5 - 24.4) | - | . |
| Yes | 10,597 | 23.9% | 4,307 | 40.6 (39.7 - 41.6) | 1.36 (1.28 - 1.45) | <.0001 |
| **Paralysis** |  |  |  |  |  |  |
| No (Reference) | 43,781 | 98.6% | 12,137 | 27.7 (27.3 - 28.1) | - | . |
| Yes | 632 | 1.4% | 260 | 41.1 (37.3 - 45.1) | 0.92 (0.73 - 1.15) | 0.452 |
| **Peptic Ulcer Disease** |  |  |  |  |  |  |
| No (Reference) | 44,311 | 99.8% | 12,371 | 27.9 (27.5 - 28.3) | - | . |
| Yes | 102 | 0.2% | 26 | 25.5 (17.6 - 35.3) | 0.43 (0.24 - 0.78) | 0.005 |
| **Peripheral Vascular Disorder** |  |  |  |  |  |  |
| No (Reference) | 40,022 | 90.1% | 11,088 | 27.7 (27.3 - 28.1) | - | . |
| Yes | 4,391 | 9.9% | 1,309 | 29.8 (28.5 - 31.2) | 0.97 (0.89 - 1.07) | 0.587 |
| **Pulmonary Circulation Disorder** |  |  |  |  |  |  |
| No (Reference) | 42,829 | 96.4% | 11,826 | 27.6 (27.2 - 28.0) | - | . |
| Yes | 1,584 | 3.6% | 571 | 36.0 (33.7 - 38.5) | 1.05 (0.90 - 1.22) | 0.565 |
| **Renal Failure** |  |  |  |  |  |  |
| No (Reference) | 35,804 | 80.6% | 9,432 | 26.3 (25.9 - 26.8) | - | . |
| Yes | 8,609 | 19.4% | 2,965 | 34.4 (33.4 - 35.5) | 0.98 (0.90 - 1.08) | 0.723 |
| **Valvular Disease** |  |  |  |  |  |  |
| No (Reference) | 41,015 | 92.3% | 11,351 | 27.7 (27.2 - 28.1) | - | . |
| Yes | 3,398 | 7.7% | 1,046 | 30.8 (29.2 - 32.4) | 0.93 (0.84 - 1.03) | 0.188 |
| **Weight Loss** |  |  |  |  |  |  |
| No (Reference) | 39,576 | 89.1% | 10,376 | 26.2 (25.8 - 26.7) | - | . |
| Yes | 4,837 | 10.9% | 2,021 | 41.8 (40.4 - 43.2) | 1.13 (1.04 - 1.23) | 0.004 |
| ^a^ Patients with missing values for ADI, Urban/Rural, Hospital Size and Ownership were dropped from the multivariate analysis. | | | | | | |
| ^b^ The South US census region includes counts for "Other" territories to avoid identification due to low counts. | | | | | | |
| ^c^ Patients have not had COVID-19 diagnosis recorded in the 30 days prior to admission. | | | | | | |
| **Notes:** Model evaluation metrics were ROC Area Under the Curve (0.8544); Pearson Chi Sq Goodness of Fit test p value (1); Deviance Goodness of Fit test p value (1). | | | | | |  |
| Intravenous Immunoglobulin/Hyperimmune Globulin Group has been omitted to avoid identification due to low counts. | | | | |  |  |

| **Supplementary Table 12.** Inpatient (IP) Mortality Rates and Corresponding Adjusted Odds Ratios (ORs) by Patient and Hospital Characteristics Among Female Recipients of COVID-19 Convalescent Plasma (CCP) | | | | | |  |
| --- | --- | --- | --- | --- | --- | --- |
| **Patient and Hospital Characteristics** | **CCP Transfusion Stays #** | **CCP Transfusion Stays  %** | **IP Mortality  #** | **Crude IP Mortality Rate per 100  (95% CIs)** | **Adjusted ORs (95% CIs)** | **P-value** |
| **Overall** | 37,338 | 100.0% | 8,479 | 22.7 (22.3 - 23.1) | - | . |
| **Age (continuous)** | 37,338 | 100.0% | 8,479 | 22.7 (22.3 - 23.1) | 1.06 (1.05 - 1.06) | <.0001 |
| **ADI^a^** |  |  |  |  |  |  |
| 81 - 100 (Reference) | 6,057 | 16.2% | 1,466 | 24.2 (23.1 - 25.3) | - | . |
| 61 – 80 | 8,290 | 22.2% | 1,867 | 22.5 (21.6 - 23.4) | 1.01 (0.91 - 1.11) | 0.892 |
| 41 – 60 | 8,232 | 22.0% | 1,812 | 22.0 (21.1 - 22.9) | 0.99 (0.90 - 1.09) | 0.847 |
| 21 – 40 | 7,183 | 19.2% | 1,554 | 21.6 (20.7 - 22.6) | 0.87 (0.78 - 0.96) | 0.008 |
| 0 – 20 | 4,352 | 11.7% | 986 | 22.7 (21.4 - 23.9) | 0.94 (0.83 - 1.06) | 0.331 |
| **Medicaid Eligibility** |  |  |  |  |  |  |
| No (Reference) | 27,817 | 74.5% | 5,845 | 21.0 (20.5 - 21.5) | - | . |
| Yes | 9,521 | 25.5% | 2,634 | 27.7 (26.8 - 28.6) | 1.17 (1.08 - 1.26) | 0.000 |
| **Region of Residence^b^** |  |  |  |  |  |  |
| South (Reference) | 20,694 | 55.4% | 4,537 | 21.9 (21.4 - 22.5) | - | . |
| Midwest | 8,515 | 22.8% | 1,841 | 21.6 (20.8 - 22.5) | 1.07 (0.99 - 1.17) | 0.098 |
| West | 4,628 | 12.4% | 1,258 | 27.2 (25.9 - 28.5) | 1.26 (1.14 - 1.40) | <.0001 |
| Northeast | 3,501 | 9.4% | 843 | 24.1 (22.7 - 25.5) | 1.30 (1.16 - 1.46) | <.0001 |
| **Urban/Rural^a^** |  |  |  |  |  |  |
| Urban (Reference) | 25,776 | 69.0% | 5,828 | 22.6 (22.1 - 23.1) | - | . |
| Rural | 8,642 | 23.1% | 1,933 | 22.4 (21.5 - 23.3) | 1.15 (1.06 - 1.25) | 0.001 |
| **Immunocompromised Status** |  |  |  |  |  |  |
| No (Reference) | 33,800 | 90.5% | 7,557 | 22.4 (21.9 - 22.8) | - | . |
| Yes | 3,538 | 9.5% | 922 | 26.1 (24.6 - 27.5) | 1.13 (0.99 - 1.29) | 0.062 |
| **Nursing Home Stay** |  |  |  |  |  |  |
| No (Reference) | 33,461 | 89.6% | 7,312 | 21.9 (21.4 - 22.3) | - | . |
| Yes | 3,877 | 10.4% | 1,167 | 30.1 (28.7 - 31.6) | 1.22 (1.11 - 1.35) | <.0001 |
| **Emergency Room Stay** |  |  |  |  |  |  |
| No (Reference) | 3,785 | 10.1% | 929 | 24.5 (23.2 - 26.0) | - | . |
| Yes | 33,553 | 89.9% | 7,550 | 22.5 (22.1 - 23.0) | 0.95 (0.85 - 1.06) | 0.365 |
| **COVID-19 As a Reason for Admission** |  |  |  |  |  |  |
| No (Reference) | 17,804 | 47.7% | 4,307 | 24.2 (23.6 - 24.8) | - | . |
| Yes | 19,534 | 52.3% | 4,172 | 21.4 (20.8 - 21.9) | 1.06 (1.00 - 1.13) | 0.052 |
| **Time Since Most Recent COVID-19 Diagnosis in the 30 Days Prior to Admission** |  |  |  |  |  |  |
| 0 days (Reference)^c^ | 18,729 | 50.2% | 4,236 | 22.6 (22.0 - 23.2) | - | . |
| 1 - 3 days | 8,994 | 24.1% | 1,970 | 21.9 (21.1 - 22.8) | 0.96 (0.89 - 1.04) | 0.365 |
| 4 - 10 days | 7,580 | 20.3% | 1,825 | 24.1 (23.1 - 25.1) | 1.08 (0.99 - 1.17) | 0.072 |
| 11-30 days | 2,035 | 5.5% | 448 | 22.0 (20.2 - 23.9) | 0.83 (0.72 - 0.96) | 0.012 |
| **Thrombosis on Index** |  |  |  |  |  |  |
| No (Reference) | 32,726 | 87.6% | 6,713 | 20.5 (20.1 - 21.0) | - | . |
| Yes | 4,612 | 12.4% | 1,766 | 38.3 (36.9 - 39.7) | 1.34 (1.22 - 1.47) | <.0001 |
| **Hospital Ownership^a^** |  |  |  |  |  |  |
| Government (Reference) | 4,899 | 13.1% | 1,116 | 22.8 (21.6 - 24.0) | - | . |
| For Profit | 7,761 | 20.8% | 1,824 | 23.5 (22.6 - 24.5) | 0.77 (0.69 - 0.86) | <.0001 |
| Non-Profit | 24,621 | 65.9% | 5,527 | 22.4 (21.9 - 23.0) | 0.83 (0.75 - 0.91) | 0.000 |
| **Hospital Size^a^** |  |  |  |  |  |  |
| Small (< 100 beds) (Reference) | 4,454 | 11.9% | 797 | 17.9 (16.8 - 19.1) | - | . |
| Medium (100 - 500 beds) | 24,035 | 64.4% | 5,575 | 23.2 (22.7 - 23.7) | 1.09 (0.98 - 1.21) | 0.121 |
| Large (> 500 beds) | 7,059 | 18.9% | 1,711 | 24.2 (23.2 - 25.3) | 1.00 (0.88 - 1.13) | 0.987 |
| **ICU/CCU Stay** |  |  |  |  |  |  |
| No (Reference) | 16,394 | 43.9% | 1,452 | 8.9 (8.4 - 9.3) | - | . |
| Yes | 20,944 | 56.1% | 7,027 | 33.6 (32.9 - 34.2) | 2.72 (2.52 - 2.93) | <.0001 |
| **Mechanical Ventilation** |  |  |  |  |  |  |
| No (Reference) | 31,897 | 85.4% | 4,602 | 14.4 (14.0 - 14.8) | - | . |
| Less Than 96hrs | 1,791 | 4.8% | 1,302 | 72.7 (70.6 - 74.7) | 12.39 (10.91 - 14.06) | <.0001 |
| More Than 96hrs | 3,650 | 9.8% | 2,575 | 70.5 (69.0 - 72.0) | 10.59 (9.62 - 11.66) | <.0001 |
| **Tocilizumab** |  |  |  |  |  |  |
| No (Reference) | 36,174 | 96.9% | 8,002 | 22.1 (21.7 - 22.6) | - | . |
| Yes | 1,164 | 3.1% | 477 | 41.0 (38.1 - 43.9) | 1.85 (1.58 - 2.17) | <.0001 |
| **Baricitinib** |  |  |  |  |  |  |
| No (Reference) | 36,978 | 99.0% | 8,377 | 22.7 (22.2 - 23.1) | - | . |
| Yes | 360 | 1.0% | 102 | 28.3 (23.8 - 33.3) | 1.22 (0.89 - 1.66) | 0.216 |
| **Remdesivir** |  |  |  |  |  |  |
| No (Reference) | 9,349 | 25.0% | 2,292 | 24.5 (23.6 - 25.4) | - | . |
| Yes | 27,989 | 75.0% | 6,187 | 22.1 (21.6 - 22.6) | 0.94 (0.87 - 1.01) | 0.109 |
| **Charlson Comorbidity Index (CCI)** |  |  |  |  |  |  |
| 0 (Reference) | 6,793 | 18.2% | 1,032 | 15.2 (14.4 - 16.1) | - | . |
| 1 | 9,877 | 26.5% | 1,884 | 19.1 (18.3 - 19.9) | 1.12 (1.00 - 1.25) | 0.055 |
| 2 | 7,411 | 19.8% | 1,755 | 23.7 (22.7 - 24.7) | 1.17 (1.03 - 1.34) | 0.018 |
| 3 – 4 | 8,186 | 21.9% | 2,227 | 27.2 (26.2 - 28.2) | 1.19 (1.02 - 1.38) | 0.025 |
| 5+ | 5,071 | 13.6% | 1,581 | 31.2 (29.9 - 32.5) | 1.27 (1.04 - 1.55) | 0.022 |
| **90-Day Comorbidity History** |  |  |  |  |  |  |
| **Thrombosis History** |  |  |  |  |  |  |
| No (Reference) | 36,301 | 97.2% | 8,133 | 22.4 (22.0 - 22.8) | - | . |
| Yes | 1,037 | 2.8% | 346 | 33.4 (30.5 - 36.3) | 1.14 (0.96 - 1.37) | 0.136 |
| **ESRD History** |  |  |  |  |  |  |
| No (Reference) | 36,214 | 97.0% | 8,161 | 22.5 (22.1 - 23.0) | - | . |
| Yes | 1,124 | 3.0% | 318 | 28.3 (25.7 - 31.0) | 0.85 (0.68 - 1.07) | 0.160 |
| **Elixhauser Comorbidity History** |  |  |  |  |  |  |
| **Blood Loss Anemia** |  |  |  |  |  |  |
| No (Reference) | 37,151 | 99.5% | 8,425 | 22.7 (22.3 - 23.1) | - | . |
| Yes | 187 | 0.5% | 54 | 28.9 (22.6 - 36.0) | 0.96 (0.64 - 1.45) | 0.839 |
| **Cardiac Arrhythmia** |  |  |  |  |  |  |
| No (Reference) | 24,834 | 66.5% | 4,581 | 18.4 (18.0 - 18.9) | - | . |
| Yes | 12,504 | 33.5% | 3,898 | 31.2 (30.4 - 32.0) | 1.38 (1.29 - 1.48) | <.0001 |
| **Chronic Pulmonary Disease** |  |  |  |  |  |  |
| No (Reference) | 24,828 | 66.5% | 5,485 | 22.1 (21.6 - 22.6) | - | . |
| Yes | 12,510 | 33.5% | 2,994 | 23.9 (23.2 - 24.7) | 1.04 (0.96 - 1.12) | 0.359 |
| **Coagulopathy** |  |  |  |  |  |  |
| No (Reference) | 31,648 | 84.8% | 6,458 | 20.4 (20.0 - 20.9) | - | . |
| Yes | 5,690 | 15.2% | 2,021 | 35.5 (34.3 - 36.8) | 1.36 (1.26 - 1.48) | <.0001 |
| **Congestive Heart Failure** |  |  |  |  |  |  |
| No (Reference) | 27,234 | 72.9% | 5,540 | 20.3 (19.9 - 20.8) | - | . |
| Yes | 10,104 | 27.1% | 2,939 | 29.1 (28.2 - 30.0) | 0.99 (0.90 - 1.09) | 0.860 |
| **Deficiency Anemia** |  |  |  |  |  |  |
| No (Reference) | 35,347 | 94.7% | 7,988 | 22.6 (22.2 - 23.0) | - | . |
| Yes | 1,991 | 5.3% | 491 | 24.7 (22.8 - 26.6) | 0.94 (0.82 - 1.07) | 0.334 |
| **Diabetes** |  |  |  |  |  |  |
| No (Reference) | 21,854 | 58.5% | 4,656 | 21.3 (20.8 - 21.9) | - | . |
| Yes | 15,484 | 41.5% | 3,823 | 24.7 (24.0 - 25.4) | 0.98 (0.90 - 1.07) | 0.701 |
| **Fluid and Electrolyte Disorder** |  |  |  |  |  |  |
| No (Reference) | 13,870 | 37.1% | 1,826 | 13.2 (12.6 - 13.7) | - | . |
| Yes | 23,468 | 62.9% | 6,653 | 28.3 (27.8 - 28.9) | 1.53 (1.42 - 1.64) | <.0001 |
| **Hypertension (uncomplicated)** |  |  |  |  |  |  |
| No (Reference) | 19,900 | 53.3% | 5,028 | 25.3 (24.7 - 25.9) | - | . |
| Yes | 17,438 | 46.7% | 3,451 | 19.8 (19.2 - 20.4) | 0.98 (0.90 - 1.07) | 0.637 |
| **Hypertension (complicated)** |  |  |  |  |  |  |
| No (Reference) | 22,461 | 60.2% | 4,334 | 19.3 (18.8 - 19.8) | - | . |
| Yes | 14,877 | 39.8% | 4,145 | 27.9 (27.1 - 28.6) | 1.08 (0.97 - 1.19) | 0.160 |
| **Liver Disease** |  |  |  |  |  |  |
| No (Reference) | 35,609 | 95.4% | 7,816 | 21.9 (21.5 - 22.4) | - | . |
| Yes | 1,729 | 4.6% | 663 | 38.3 (36.1 - 40.7) | 1.45 (1.26 - 1.67) | <.0001 |
| **Metastatic Cancer** |  |  |  |  |  |  |
| No (Reference) | 36,848 | 98.7% | 8,343 | 22.6 (22.2 - 23.1) | - | . |
| Yes | 490 | 1.3% | 136 | 27.8 (23.9 - 32.0) | 1.31 (0.96 - 1.77) | 0.089 |
| **Obesity** |  |  |  |  |  |  |
| No (Reference) | 27,068 | 72.5% | 5,975 | 22.1 (21.6 - 22.6) | - | . |
| Yes | 10,270 | 27.5% | 2,504 | 24.4 (23.6 - 25.2) | 1.07 (0.99 - 1.15) | 0.073 |
| **Other Neurological Disorders** |  |  |  |  |  |  |
| No (Reference) | 28,723 | 76.9% | 5,597 | 19.5 (19.0 - 20.0) | - | . |
| Yes | 8,615 | 23.1% | 2,882 | 33.5 (32.5 - 34.5) | 1.25 (1.16 - 1.34) | <.0001 |
| **Paralysis** |  |  |  |  |  |  |
| No (Reference) | 36,902 | 98.8% | 8,312 | 22.5 (22.1 - 23.0) | - | . |
| Yes | 436 | 1.2% | 167 | 38.3 (33.7 - 43.1) | 1.16 (0.90 - 1.51) | 0.255 |
| **Peripheral Vascular Disorder** |  |  |  |  |  |  |
| No (Reference) | 34,940 | 93.6% | 7,839 | 22.4 (22.0 - 22.9) | - | . |
| Yes | 2,398 | 6.4% | 640 | 26.7 (24.9 - 28.5) | 0.98 (0.87 - 1.11) | 0.749 |
| **Pulmonary Circulation Disorder** |  |  |  |  |  |  |
| No (Reference) | 36,114 | 96.7% | 8,108 | 22.5 (22.0 - 22.9) | - | . |
| Yes | 1,224 | 3.3% | 371 | 30.3 (27.8 - 33.0) | 1.05 (0.88 - 1.25) | 0.606 |
| **Renal Failure** |  |  |  |  |  |  |
| No (Reference) | 31,116 | 83.3% | 6,569 | 21.1 (20.7 - 21.6) | - | . |
| Yes | 6,222 | 16.7% | 1,910 | 30.7 (29.6 - 31.9) | 1.16 (1.03 - 1.29) | 0.011 |
| **Rheumatoid Arthritis/Collagen Vascular Diseases (CVD)** |  |  |  |  |  |  |
| No (Reference) | 34,792 | 93.2% | 7,838 | 22.5 (22.1 - 23.0) | - | . |
| Yes | 2,546 | 6.8% | 641 | 25.2 (23.5 - 26.9) | 1.21 (1.07 - 1.36) | 0.003 |
| **Solid Tumor** |  |  |  |  |  |  |
| No (Reference) | 36,149 | 96.8% | 8,168 | 22.6 (22.2 - 23.0) | - | . |
| Yes | 1,189 | 3.2% | 311 | 26.2 (23.7 - 28.8) | 1.12 (0.91 - 1.37) | 0.276 |
| **Valvular Disease** |  |  |  |  |  |  |
| No (Reference) | 34,323 | 91.9% | 7,665 | 22.3 (21.9 - 22.8) | - | . |
| Yes | 3,015 | 8.1% | 814 | 27.0 (25.4 - 28.6) | 0.96 (0.86 - 1.07) | 0.430 |
| **Weight Loss** |  |  |  |  |  |  |
| No (Reference) | 33,320 | 89.2% | 7,088 | 21.3 (20.8 - 21.7) | - | . |
| Yes | 4,018 | 10.8% | 1,391 | 34.6 (33.2 - 36.1) | 1.17 (1.07 - 1.28) | 0.001 |
| ^a^ Patients with missing values for ADI, Urban/Rural, Hospital Size and Ownership were dropped from the multivariate analysis. | | | | | | |
| ^b^ The South US census region includes counts for "Other" territories to avoid identification due to low counts. | | | | | | |
| ^c^ Patients have not had COVID-19 diagnosis recorded in the 30 days prior to admission. | | | | | | |
| **Notes:** Model evaluation metrics were ROC Area Under the Curve (0.8376); Pearson Chi Sq Goodness of Fit test p value (1); Deviance Goodness of Fit test p value (1). | | | | | |  |
| Intravenous Immunoglobulin/Hyperimmune Globulin Group has been omitted to avoid identification due to low counts. | | | | |  |  |

| **Supplementary Table 13.** Inpatient (IP) Mortality Rates and Corresponding Adjusted Odds Ratios (ORs) by Patient and Hospital Characteristics Among Recipients of COVID-19 Convalescent Plasma (CCP) with no COVID-19 Diagnosis Recorded in the 30 Days Prior to Hospital Admission | | | | | | |
| --- | --- | --- | --- | --- | --- | --- |
| **Patient and Hospital Characteristics** | **CCP Transfusion Stays #** | **CCP Transfusion Stays  %** | **IP Mortality  #** | **Crude IP Mortality Rate per 100  (95% CIs)** | **Adjusted ORs (95% CIs)** | **P-value** |
| **Overall** | 40,982 | 100.0% | 10,504 | 25.6 (25.2 - 26.1) | - | . |
| **Age (continuous)** | 40,982 | 100.0% | 10,504 | 25.6 (25.2 - 26.1) | 1.06 (1.06 - 1.07) | <.0001 |
| **Sex** |  |  |  |  |  |  |
| Female (Reference) | 18,729 | 45.7% | 4,236 | 22.6 (22.0 - 23.2) | - | . |
| Male | 22,253 | 54.3% | 6,268 | 28.2 (27.6 - 28.8) | 1.19 (1.12 - 1.26) | <.0001 |
| **ADI^a^** |  |  |  |  |  |  |
| 81 - 100 (Reference) | 6,226 | 15.2% | 1,671 | 26.8 (25.7 - 28.0) | - | . |
| 61 – 80 | 8,661 | 21.1% | 2,204 | 25.4 (24.5 - 26.4) | 0.99 (0.91 - 1.09) | 0.907 |
| 41 – 60 | 9,116 | 22.2% | 2,250 | 24.7 (23.8 - 25.6) | 0.96 (0.88 - 1.06) | 0.447 |
| 21 – 40 | 8,121 | 19.8% | 2,065 | 25.4 (24.5 - 26.4) | 0.92 (0.84 - 1.02) | 0.104 |
| 0 – 20 | 5,339 | 13.0% | 1,403 | 26.3 (25.1 - 27.5) | 0.90 (0.81 - 1.01) | 0.067 |
| **Medicaid Eligibility** |  |  |  |  |  |  |
| No (Reference) | 32,247 | 78.7% | 7,954 | 24.7 (24.2 - 25.1) | - | . |
| Yes | 8,735 | 21.3% | 2,550 | 29.2 (28.2 - 30.2) | 1.12 (1.03 - 1.21) | 0.006 |
| **Region of Residence^b^** |  |  |  |  |  |  |
| South (Reference) | 21,329 | 52.0% | 5,310 | 24.9 (24.3 - 25.5) | - | . |
| Midwest | 9,630 | 23.5% | 2,271 | 23.6 (22.7 - 24.4) | 1.00 (0.93 - 1.09) | 0.907 |
| West | 5,761 | 14.1% | 1,754 | 30.4 (29.3 - 31.7) | 1.19 (1.09 - 1.31) | 0.000 |
| Northeast | 4,262 | 10.4% | 1,169 | 27.4 (26.1 - 28.8) | 1.23 (1.11 - 1.37) | <.0001 |
| **Urban/Rural^a^** |  |  |  |  |  |  |
| Urban (Reference) | 28,958 | 70.7% | 7,560 | 26.1 (25.6 - 26.6) | - | . |
| Rural | 8,829 | 21.5% | 2,117 | 24.0 (23.1 - 24.9) | 1.17 (1.08 - 1.26) | 0.000 |
| **Immunocompromised Status** |  |  |  |  |  |  |
| No (Reference) | 37,017 | 90.3% | 9,369 | 25.3 (24.9 - 25.8) | - | . |
| Yes | 3,965 | 9.7% | 1,135 | 28.6 (27.2 - 30.1) | 1.14 (1.01 - 1.29) | 0.029 |
| **Nursing Home Stay** |  |  |  |  |  |  |
| No (Reference) | 37,547 | 91.6% | 9,483 | 25.3 (24.8 - 25.7) | - | . |
| Yes | 3,435 | 8.4% | 1,021 | 29.7 (28.2 - 31.3) | 1.05 (0.94 - 1.17) | 0.378 |
| **Emergency Room Stay** |  |  |  |  |  |  |
| No (Reference) | 3,325 | 8.1% | 811 | 24.4 (22.9 - 25.9) | - | . |
| Yes | 37,657 | 91.9% | 9,693 | 25.7 (25.3 - 26.2) | 0.90 (0.80 - 1.02) | 0.092 |
| **COVID-19 As a Reason for Admission** |  |  |  |  |  |  |
| No (Reference) | 20,696 | 50.5% | 5,702 | 27.6 (26.9 - 28.2) | - | . |
| Yes | 20,286 | 49.5% | 4,802 | 23.7 (23.1 - 24.3) | 1.01 (0.95 - 1.07) | 0.819 |
| **Thrombosis on Index** |  |  |  |  |  |  |
| No (Reference) | 34,951 | 85.3% | 8,019 | 22.9 (22.5 - 23.4) | - | . |
| Yes | 6,031 | 14.7% | 2,485 | 41.2 (40.0 - 42.5) | 1.29 (1.18 - 1.40) | <.0001 |
| **Hospital Ownership^a^** |  |  |  |  |  |  |
| Government (Reference) | 5,151 | 12.6% | 1,296 | 25.2 (24.0 - 26.4) | - | . |
| For Profit | 8,422 | 20.6% | 2,212 | 26.3 (25.3 - 27.2) | 0.79 (0.71 - 0.88) | <.0001 |
| Non-Profit | 27,316 | 66.7% | 6,980 | 25.6 (25.0 - 26.1) | 0.88 (0.81 - 0.97) | 0.009 |
| **Hospital Size^a^** |  |  |  |  |  |  |
| Small (< 100 beds) (Reference) | 4,806 | 11.7% | 910 | 18.9 (17.8 - 20.1) | - | . |
| Medium (100 - 500 beds) | 26,473 | 64.6% | 6,985 | 26.4 (25.9 - 26.9) | 1.20 (1.08 - 1.33) | 0.001 |
| Large (> 500 beds) | 7,692 | 18.8% | 2,118 | 27.5 (26.5 - 28.6) | 1.15 (1.03 - 1.30) | 0.018 |
| **ICU/CCU Stay** |  |  |  |  |  |  |
| No (Reference) | 16,508 | 40.3% | 1,593 | 9.6 (9.2 - 10.1) | - | . |
| Yes | 24,474 | 59.7% | 8,911 | 36.4 (35.8 - 37.0) | 2.66 (2.48 - 2.86) | <.0001 |
| **Mechanical Ventilation** |  |  |  |  |  |  |
| No (Reference) | 33,982 | 82.9% | 5,306 | 15.6 (15.2 - 16.0) | - | . |
| Less Than 96hrs | 2,425 | 5.9% | 1,810 | 74.6 (72.8 - 76.4) | 13.20 (11.80 - 14.76) | <.0001 |
| More Than 96hrs | 4,575 | 11.2% | 3,388 | 74.1 (72.8 - 75.3) | 11.58 (10.59 - 12.65) | <.0001 |
| **Tocilizumab** |  |  |  |  |  |  |
| No (Reference) | 39,650 | 96.7% | 9,917 | 25.0 (24.6 - 25.4) | - | . |
| Yes | 1,332 | 3.3% | 587 | 44.1 (41.4 - 46.8) | 1.77 (1.53 - 2.05) | <.0001 |
| **Baricitinib** |  |  |  |  |  |  |
| No (Reference) | 40,594 | 99.1% | 10,372 | 25.6 (25.1 - 26.0) | - | . |
| Yes | 388 | 0.9% | 132 | 34.0 (29.4 - 39.0) | 1.68 (1.27 - 2.21) | 0.000 |
| **Remdesivir** |  |  |  |  |  |  |
| No (Reference) | 9,721 | 23.7% | 2,721 | 28.0 (27.1 - 28.9) | - | . |
| Yes | 31,261 | 76.3% | 7,783 | 24.9 (24.4 - 25.4) | 0.89 (0.83 - 0.95) | 0.001 |
| **Charlson Comorbidity Index (CCI)** |  |  |  |  |  |  |
| 0 (Reference) | 6,630 | 16.2% | 1,210 | 18.3 (17.3 - 19.2) | - | . |
| 1 | 10,076 | 24.6% | 2,151 | 21.3 (20.6 - 22.2) | 1.06 (0.95 - 1.18) | 0.296 |
| 2 | 8,036 | 19.6% | 2,102 | 26.2 (25.2 - 27.1) | 1.22 (1.08 - 1.37) | 0.001 |
| 3 – 4 | 9,801 | 23.9% | 2,895 | 29.5 (28.6 - 30.5) | 1.20 (1.05 - 1.37) | 0.006 |
| 5+ | 6,439 | 15.7% | 2,146 | 33.3 (32.2 - 34.5) | 1.26 (1.06 - 1.49) | 0.009 |
| **90-Day Comorbidity History** |  |  |  |  |  |  |
| **Thrombosis History** |  |  |  |  |  |  |
| No (Reference) | 39,981 | 97.6% | 10,173 | 25.4 (25.0 - 25.9) | - | . |
| Yes | 1,001 | 2.4% | 331 | 33.1 (30.2 - 36.1) | 1.05 (0.88 - 1.26) | 0.563 |
| **ESRD History** |  |  |  |  |  |  |
| No (Reference) | 39,686 | 96.8% | 10,123 | 25.5 (25.1 - 25.9) | - | . |
| Yes | 1,296 | 3.2% | 381 | 29.4 (26.9 - 32.0) | 0.83 (0.67 - 1.01) | 0.068 |
| **Elixhauser Comorbidity History** |  |  |  |  |  |  |
| **Cardiac Arrhythmia** |  |  |  |  |  |  |
| No (Reference) | 25,841 | 63.1% | 5,423 | 21.0 (20.5 - 21.5) | - | . |
| Yes | 15,141 | 36.9% | 5,081 | 33.6 (32.8 - 34.3) | 1.28 (1.21 - 1.36) | <.0001 |
| **Coagulopathy** |  |  |  |  |  |  |
| No (Reference) | 33,914 | 82.8% | 7,847 | 23.1 (22.7 - 23.6) | - | . |
| Yes | 7,068 | 17.2% | 2,657 | 37.6 (36.5 - 38.7) | 1.29 (1.20 - 1.38) | <.0001 |
| **Congestive Heart Failure** |  |  |  |  |  |  |
| No (Reference) | 28,962 | 70.7% | 6,767 | 23.4 (22.9 - 23.9) | - | . |
| Yes | 12,020 | 29.3% | 3,737 | 31.1 (30.3 - 31.9) | 1.01 (0.92 - 1.09) | 0.900 |
| **Deficiency Anemia** |  |  |  |  |  |  |
| No (Reference) | 39,008 | 95.2% | 9,950 | 25.5 (25.1 - 25.9) | - | . |
| Yes | 1,974 | 4.8% | 554 | 28.1 (26.1 - 30.1) | 0.97 (0.86 - 1.11) | 0.691 |
| **Depression** |  |  |  |  |  |  |
| No (Reference) | 35,011 | 85.4% | 9,100 | 26.0 (25.5 - 26.5) | - | . |
| Yes | 5,971 | 14.6% | 1,404 | 23.5 (22.4 - 24.6) | 1.02 (0.93 - 1.11) | 0.691 |
| **Diabetes** |  |  |  |  |  |  |
| No (Reference) | 23,011 | 56.1% | 5,598 | 24.3 (23.8 - 24.9) | - | . |
| Yes | 17,971 | 43.9% | 4,906 | 27.3 (26.7 - 28.0) | 0.98 (0.91 - 1.05) | 0.538 |
| **Fluid and Electrolyte Disorder** |  |  |  |  |  |  |
| No (Reference) | 15,407 | 37.6% | 2,185 | 14.2 (13.6 - 14.7) | - | . |
| Yes | 25,575 | 62.4% | 8,319 | 32.5 (32.0 - 33.1) | 1.62 (1.52 - 1.74) | <.0001 |
| **Hypertension (uncomplicated)** |  |  |  |  |  |  |
| No (Reference) | 23,649 | 57.7% | 6,748 | 28.5 (28.0 - 29.1) | - | . |
| Yes | 17,333 | 42.3% | 3,756 | 21.7 (21.1 - 22.3) | 0.89 (0.82 - 0.97) | 0.006 |
| **Hypertension (complicated)** |  |  |  |  |  |  |
| No (Reference) | 23,350 | 57.0% | 5,164 | 22.1 (21.6 - 22.7) | - | . |
| Yes | 17,632 | 43.0% | 5,340 | 30.3 (29.6 - 31.0) | 1.06 (0.96 - 1.16) | 0.233 |
| **Liver Disease** |  |  |  |  |  |  |
| No (Reference) | 39,013 | 95.2% | 9,652 | 24.7 (24.3 - 25.2) | - | . |
| Yes | 1,969 | 4.8% | 852 | 43.3 (41.1 - 45.5) | 1.48 (1.30 - 1.68) | <.0001 |
| **Metastatic Cancer** |  |  |  |  |  |  |
| No (Reference) | 40,377 | 98.5% | 10,316 | 25.5 (25.1 - 26.0) | - | . |
| Yes | 605 | 1.5% | 188 | 31.1 (27.4 - 35.0) | 1.44 (1.10 - 1.87) | 0.008 |
| **Other Neurological Disorders** |  |  |  |  |  |  |
| No (Reference) | 31,186 | 76.1% | 6,867 | 22.0 (21.6 - 22.5) | - | . |
| Yes | 9,796 | 23.9% | 3,637 | 37.1 (36.2 - 38.1) | 1.28 (1.20 - 1.37) | <.0001 |
| **Paralysis** |  |  |  |  |  |  |
| No (Reference) | 40,447 | 98.7% | 10,285 | 25.4 (25.0 - 25.9) | - | . |
| Yes | 535 | 1.3% | 219 | 40.9 (36.8 - 45.2) | 1.06 (0.84 - 1.36) | 0.612 |
| **Peripheral Vascular Disorder** |  |  |  |  |  |  |
| No (Reference) | 37,574 | 91.7% | 9,532 | 25.4 (24.9 - 25.8) | - | . |
| Yes | 3,408 | 8.3% | 972 | 28.5 (27.0 - 30.1) | 1.03 (0.93 - 1.14) | 0.614 |
| **Pulmonary Circulation Disorder** |  |  |  |  |  |  |
| No (Reference) | 39,618 | 96.7% | 10,033 | 25.3 (24.9 - 25.8) | - | . |
| Yes | 1,364 | 3.3% | 471 | 34.5 (32.0 - 37.1) | 1.11 (0.95 - 1.31) | 0.196 |
| **Renal Failure** |  |  |  |  |  |  |
| No (Reference) | 33,361 | 81.4% | 8,046 | 24.1 (23.7 - 24.6) | - | . |
| Yes | 7,621 | 18.6% | 2,458 | 32.3 (31.2 - 33.3) | 1.01 (0.91 - 1.12) | 0.852 |
| **Rheumatoid Arthritis/Collagen Vascular Diseases (CVD)** |  |  |  |  |  |  |
| No (Reference) | 39,206 | 95.7% | 10,047 | 25.6 (25.2 - 26.1) | - | . |
| Yes | 1,776 | 4.3% | 457 | 25.7 (23.7 - 27.8) | 1.19 (1.03 - 1.37) | 0.016 |
| **Solid Tumor** |  |  |  |  |  |  |
| No (Reference) | 39,361 | 96.0% | 10,050 | 25.5 (25.1 - 26.0) | - | . |
| Yes | 1,621 | 4.0% | 454 | 28.0 (25.8 - 30.3) | 0.99 (0.84 - 1.17) | 0.901 |
| **Valvular Disease** |  |  |  |  |  |  |
| No (Reference) | 37,638 | 91.8% | 9,553 | 25.4 (24.9 - 25.8) | - | . |
| Yes | 3,344 | 8.2% | 951 | 28.4 (26.9 - 30.0) | 0.92 (0.83 - 1.02) | 0.113 |
| **Weight Loss** |  |  |  |  |  |  |
| No (Reference) | 36,589 | 89.3% | 8,813 | 24.1 (23.6 - 24.5) | - | . |
| Yes | 4,393 | 10.7% | 1,691 | 38.5 (37.1 - 40.0) | 1.15 (1.05 - 1.26) | 0.002 |
| ^a^ Patients with missing values for ADI, Urban/Rural, Hospital Size and Ownership were dropped from the multivariate analysis. | | | | | | |
| ^b^ The South US census region includes counts for "Other" territories to avoid identification due to low counts. | | | | | | |
| **Notes:** Model evaluation metrics were ROC Area Under the Curve (0.8426); Pearson Chi Sq Goodness of Fit test p value (1); Deviance Goodness of Fit test p value (1). | | | | | |  |
| Intravenous Immunoglobulin/Hyperimmune Globulin and Peptic Ulcer Disease Groups have been omitted to avoid identification due to low counts. | | | | | | |

| **Supplementary Table 14.** Inpatient (IP) Mortality Rates and Corresponding Adjusted Odds Ratios (ORs) by Patient and Hospital Characteristics Among Recipients of COVID-19 Convalescent Plasma (CP) with Most Recent COVID-19 Diagnosis Recorded 1-10 Days Prior to Hospital Admission | | | | | | |
| --- | --- | --- | --- | --- | --- | --- |
| **Patient and Hospital Characteristics** | **CCP Transfusion Stays #** | **CCP Transfusion Stays  %** | **IP Mortality  #** | **Crude IP Mortality Rate per 100  (95% CIs)** | **Adjusted ORs (95% CIs)** | **P-value** |
| **Overall** | 36,230 | 100.0% | 9,264 | 25.6 (25.1 - 26.0) | - | . |
| **Age (continuous)** | 36,230 | 100.0% | 9,264 | 25.6 (25.1 - 26.0) | 1.06 (1.06 - 1.07) | <.0001 |
| **Sex** |  |  |  |  |  |  |
| Female (Reference) | 16,574 | 45.7% | 3,795 | 22.9 (22.3 - 23.5) | - | . |
| Male | 19,656 | 54.3% | 5,469 | 27.8 (27.2 - 28.5) | 1.22 (1.14 - 1.30) | <.0001 |
| **ADI^a^** |  |  |  |  |  |  |
| 81 - 100 (Reference) | 5,434 | 15.0% | 1,536 | 28.3 (27.1 - 29.5) | - | . |
| 61 – 80 | 7,967 | 22.0% | 2,083 | 26.1 (25.2 - 27.1) | 1.03 (0.93 - 1.14) | 0.525 |
| 41 – 60 | 8,114 | 22.4% | 1,992 | 24.6 (23.6 - 25.5) | 0.95 (0.86 - 1.06) | 0.374 |
| 21 – 40 | 7,085 | 19.6% | 1,683 | 23.8 (22.8 - 24.8) | 0.83 (0.75 - 0.93) | 0.001 |
| 0 – 20 | 4,188 | 11.6% | 976 | 23.3 (22.0 - 24.6) | 0.80 (0.70 - 0.91) | 0.001 |
| **Medicaid Eligibility** |  |  |  |  |  |  |
| No (Reference) | 29,390 | 81.1% | 7,156 | 24.3 (23.9 - 24.8) | - | . |
| Yes | 6,840 | 18.9% | 2,108 | 30.8 (29.7 - 31.9) | 1.20 (1.09 - 1.31) | <.0001 |
| **Region of Residence^b^** |  |  |  |  |  |  |
| South (Reference) | 20,073 | 55.4% | 5,099 | 25.4 (24.8 - 26.0) | - | . |
| Midwest | 8,969 | 24.8% | 2,193 | 24.5 (23.6 - 25.4) | 0.97 (0.90 - 1.06) | 0.505 |
| West | 4,177 | 11.5% | 1,193 | 28.6 (27.2 - 30.0) | 1.10 (0.99 - 1.23) | 0.072 |
| Northeast | 3,011 | 8.3% | 779 | 25.9 (24.3 - 27.5) | 1.12 (1.00 - 1.27) | 0.057 |
| **Urban/Rural^a^** |  |  |  |  |  |  |
| Urban (Reference) | 23,739 | 65.5% | 5,917 | 24.9 (24.4 - 25.5) | - | . |
| Rural | 9,353 | 25.8% | 2,446 | 26.2 (25.3 - 27.1) | 1.10 (1.02 - 1.20) | 0.017 |
| **Immunocompromised Status** |  |  |  |  |  |  |
| No (Reference) | 32,805 | 90.5% | 8,285 | 25.3 (24.8 - 25.7) | - | . |
| Yes | 3,425 | 9.5% | 979 | 28.6 (27.1 - 30.1) | 1.20 (1.05 - 1.38) | 0.008 |
| **Nursing Home Stay** |  |  |  |  |  |  |
| No (Reference) | 33,536 | 92.6% | 8,302 | 24.8 (24.3 - 25.2) | - | . |
| Yes | 2,694 | 7.4% | 962 | 35.7 (33.9 - 37.6) | 1.40 (1.25 - 1.57) | <.0001 |
| **Emergency Room Stay** |  |  |  |  |  |  |
| No (Reference) | 4,823 | 13.3% | 1,447 | 30.0 (28.7 - 31.3) | - | . |
| Yes | 31,407 | 86.7% | 7,817 | 24.9 (24.4 - 25.4) | 0.99 (0.90 - 1.10) | 0.885 |
| **COVID-19 As a Reason for Admission** |  |  |  |  |  |  |
| No (Reference) | 16,192 | 44.7% | 4,320 | 26.7 (26.0 - 27.4) | - | . |
| Yes | 20,038 | 55.3% | 4,944 | 24.7 (24.1 - 25.3) | 1.08 (1.01 - 1.15) | 0.018 |
| **Thrombosis on Index** |  |  |  |  |  |  |
| No (Reference) | 31,459 | 86.8% | 7,197 | 22.9 (22.4 - 23.3) | - | . |
| Yes | 4,771 | 13.2% | 2,067 | 43.3 (41.9 - 44.7) | 1.41 (1.28 - 1.55) | <.0001 |
| **Hospital Ownership^a^** |  |  |  |  |  |  |
| Government (Reference) | 4,710 | 13.0% | 1,210 | 25.7 (24.5 – 27.0) | - | . |
| For Profit | 6,985 | 19.3% | 1,870 | 26.8 (25.7 - 27.8) | 0.80 (0.71 - 0.90) | 0.000 |
| Non-Profit | 24,490 | 67.6% | 6,174 | 25.2 (24.7 – 25.8) | 0.83 (0.75 - 0.91) | 0.000 |
| **Hospital Size^a^** |  |  |  |  |  |  |
| Small (< 100 beds) (Reference) | 4,366 | 12.1% | 884 | 20.2 (19.1 - 21.5) | - | . |
| Medium (100 - 500 beds) | 23,071 | 63.7% | 6,015 | 26.1 (25.5 - 26.6) | 1.10 (0.99 - 1.22) | 0.077 |
| Large (> 500 beds) | 7,080 | 19.5% | 1,930 | 27.3 (26.2 - 28.3) | 1.05 (0.93 - 1.18) | 0.453 |
| **ICU/CCU Stay** |  |  |  |  |  |  |
| No (Reference) | 15,055 | 41.6% | 1,378 | 9.2 (8.7 - 9.6) | - | . |
| Yes | 21,175 | 58.4% | 7,886 | 37.2 (36.6 - 37.9) | 2.79 (2.58 - 3.01) | <.0001 |
| **Mechanical Ventilation** |  |  |  |  |  |  |
| No (Reference) | 29,646 | 81.8% | 4,404 | 14.9 (14.5 - 15.3) | - | . |
| Less Than 96hrs | 2,142 | 5.9% | 1,592 | 74.3 (72.4 - 76.2) | 13.10 (11.61 - 14.78) | <.0001 |
| More Than 96hrs | 4,442 | 12.3% | 3,268 | 73.6 (72.2 - 74.9) | 11.86 (10.80 - 13.02) | <.0001 |
| **Tocilizumab** |  |  |  |  |  |  |
| No (Reference) | 34,923 | 96.4% | 8,738 | 25.0 (24.6 - 25.5) | - | . |
| Yes | 1,307 | 3.6% | 526 | 40.2 (37.6 - 43.0) | 1.56 (1.33 - 1.82) | <.0001 |
| **Baricitinib** |  |  |  |  |  |  |
| No (Reference) | 35,866 | 99.0% | 9,153 | 25.5 (25.1 - 26.0) | - | . |
| Yes | 364 | 1.0% | 111 | 30.5 (25.9 - 35.5) | 1.22 (0.89 - 1.66) | 0.218 |
| **Remdesivir** |  |  |  |  |  |  |
| No (Reference) | 8,500 | 23.5% | 2,274 | 26.8 (25.8 - 27.7) | - | . |
| Yes | 27,730 | 76.5% | 6,990 | 25.2 (24.7 - 25.7) | 1.00 (0.93 - 1.09) | 0.907 |
| **Charlson Comorbidity Index (CCI)** |  |  |  |  |  |  |
| 0 (Reference) | 7,025 | 19.4% | 1,217 | 17.3 (16.4 - 18.2) | - | . |
| 1 | 9,414 | 26.0% | 2,068 | 22.0 (21.1 - 22.8) | 1.16 (1.03 - 1.30) | 0.011 |
| 2 | 6,988 | 19.3% | 1,840 | 26.3 (25.3 - 27.4) | 1.23 (1.08 - 1.40) | 0.002 |
| 3 – 4 | 7,814 | 21.6% | 2,417 | 30.9 (29.9 - 32.0) | 1.28 (1.10 - 1.49) | 0.001 |
| 5+ | 4,989 | 13.8% | 1,722 | 34.5 (33.2 - 35.9) | 1.37 (1.13 - 1.67) | 0.002 |
| **90-Day Comorbidity History** |  |  |  |  |  |  |
| **Thrombosis History** |  |  |  |  |  |  |
| No (Reference) | 35,240 | 97.3% | 8,922 | 25.3 (24.9 - 25.8) | - | . |
| Yes | 990 | 2.7% | 342 | 34.5 (31.6 - 37.6) | 0.88 (0.73 - 1.06) | 0.170 |
| **ESRD History** |  |  |  |  |  |  |
| No (Reference) | 35,273 | 97.4% | 8,970 | 25.4 (25.0 - 25.9) | - | . |
| Yes | 957 | 2.6% | 294 | 30.7 (27.8 - 33.8) | 0.91 (0.72 - 1.15) | 0.450 |
| **Elixhauser Comorbidity History** |  |  |  |  |  |  |
| **Alcohol Abuse** |  |  |  |  |  |  |
| No (Reference) | 35,842 | 98.9% | 9,142 | 25.5 (25.1 - 26.0) | - | . |
| Yes | 388 | 1.1% | 122 | 31.4 (26.9 - 36.4) | 0.95 (0.71 - 1.29) | 0.751 |
| **Blood Loss Anemia** |  |  |  |  |  |  |
| No (Reference) | 36,069 | 99.6% | 9,201 | 25.5 (25.1 - 26.0) | - | . |
| Yes | 161 | 0.4% | 63 | 39.1 (31.6 - 47.2) | 1.44 (0.94 - 2.21) | 0.090 |
| **Cardiac Arrhythmia** |  |  |  |  |  |  |
| No (Reference) | 23,059 | 63.6% | 4,612 | 20.0 (19.5 - 20.5) | - | . |
| Yes | 13,171 | 36.4% | 4,652 | 35.3 (34.5 - 36.1) | 1.44 (1.35 - 1.54) | <.0001 |
| **Chronic Pulmonary Disease** |  |  |  |  |  |  |
| No (Reference) | 25,281 | 69.8% | 6,340 | 25.1 (24.5 - 25.6) | - | . |
| Yes | 10,949 | 30.2% | 2,924 | 26.7 (25.9 - 27.5) | 1.01 (0.94 - 1.09) | 0.741 |
| **Coagulopathy** |  |  |  |  |  |  |
| No (Reference) | 29,962 | 82.7% | 6,834 | 22.8 (22.3 - 23.3) | - | . |
| Yes | 6,268 | 17.3% | 2,430 | 38.8 (37.6 - 40.0) | 1.28 (1.18 - 1.39) | <.0001 |
| **Congestive Heart Failure** |  |  |  |  |  |  |
| No (Reference) | 27,139 | 74.9% | 6,278 | 23.1 (22.6 - 23.6) | - | . |
| Yes | 9,091 | 25.1% | 2,986 | 32.8 (31.9 - 33.8) | 1.02 (0.93 - 1.12) | 0.637 |
| **Deficiency Anemia** |  |  |  |  |  |  |
| No (Reference) | 34,631 | 95.6% | 8,811 | 25.4 (25.0 - 25.9) | - | . |
| Yes | 1,599 | 4.4% | 453 | 28.3 (26.1 - 30.6) | 0.94 (0.81 - 1.09) | 0.443 |
| **Diabetes** |  |  |  |  |  |  |
| No (Reference) | 20,489 | 56.6% | 4,889 | 23.9 (23.3 - 24.5) | - | . |
| Yes | 15,741 | 43.4% | 4,375 | 27.8 (27.1 - 28.5) | 0.95 (0.87 - 1.04) | 0.254 |
| **Fluid and Electrolyte Disorder** |  |  |  |  |  |  |
| No (Reference) | 14,005 | 38.7% | 2,016 | 14.4 (13.8 - 15.0) | - | . |
| Yes | 22,225 | 61.3% | 7,248 | 32.6 (32.0 - 33.2) | 1.50 (1.40 - 1.61) | <.0001 |
| **Hypertension (uncomplicated)** |  |  |  |  |  |  |
| No (Reference) | 19,288 | 53.2% | 5,448 | 28.2 (27.6 - 28.9) | - | . |
| Yes | 16,942 | 46.8% | 3,816 | 22.5 (21.9 - 23.2) | 1.01 (0.93 - 1.10) | 0.798 |
| **Hypertension (complicated)** |  |  |  |  |  |  |
| No (Reference) | 21,952 | 60.6% | 4,789 | 21.8 (21.3 - 22.4) | - | . |
| Yes | 14,278 | 39.4% | 4,475 | 31.3 (30.6 - 32.1) | 1.06 (0.95 - 1.17) | 0.291 |
| **Liver Disease** |  |  |  |  |  |  |
| No (Reference) | 34,602 | 95.5% | 8,625 | 24.9 (24.5 - 25.4) | - | . |
| Yes | 1,628 | 4.5% | 639 | 39.3 (36.9 - 41.7) | 1.34 (1.16 - 1.56) | 0.000 |
| **Lymphoma** |  |  |  |  |  |  |
| No (Reference) | 35,734 | 98.6% | 9,142 | 25.6 (25.1 - 26.0) | - | . |
| Yes | 496 | 1.4% | 122 | 24.6 (20.9 - 28.7) | 0.73 (0.54 - 0.98) | 0.038 |
| **Obesity** |  |  |  |  |  |  |
| No (Reference) | 26,980 | 74.5% | 6,751 | 25.0 (24.5 - 25.5) | - | . |
| Yes | 9,250 | 25.5% | 2,513 | 27.2 (26.3 - 28.1) | 1.09 (1.01 - 1.17) | 0.033 |
| **Other Neurological Disorders** |  |  |  |  |  |  |
| No (Reference) | 28,243 | 78.0% | 6,197 | 21.9 (21.5 - 22.4) | - | . |
| Yes | 7,987 | 22.0% | 3,067 | 38.4 (37.3 - 39.5) | 1.33 (1.24 - 1.44) | <.0001 |
| **Paralysis** |  |  |  |  |  |  |
| No (Reference) | 35,794 | 98.8% | 9,088 | 25.4 (24.9 - 25.8) | - | . |
| Yes | 436 | 1.2% | 176 | 40.4 (35.8 - 45.2) | 0.98 (0.75 - 1.27) | 0.860 |
| **Peripheral Vascular Disorder** |  |  |  |  |  |  |
| No (Reference) | 33,351 | 92.1% | 8,423 | 25.3 (24.8 - 25.7) | - | . |
| Yes | 2,879 | 7.9% | 841 | 29.2 (27.6 - 30.9) | 0.95 (0.84 - 1.06) | 0.365 |
| **Pulmonary Circulation Disorder** |  |  |  |  |  |  |
| No (Reference) | 35,035 | 96.7% | 8,857 | 25.3 (24.8 - 25.7) | - | . |
| Yes | 1,195 | 3.3% | 407 | 34.1 (31.4 - 36.8) | 1.04 (0.87 - 1.24) | 0.696 |
| **Renal Failure** |  |  |  |  |  |  |
| No (Reference) | 30,086 | 83.0% | 7,162 | 23.8 (23.3 - 24.3) | - | . |
| Yes | 6,144 | 17.0% | 2,102 | 34.2 (33.0 - 35.4) | 1.15 (1.03 - 1.29) | 0.010 |
| **Rheumatoid Arthritis/Collagen Vascular Diseases (CVD)** |  |  |  |  |  |  |
| No (Reference) | 34,463 | 95.1% | 8,779 | 25.5 (25.0 - 25.9) | - | . |
| Yes | 1,767 | 4.9% | 485 | 27.4 (25.4 - 29.6) | 1.17 (1.01 - 1.36) | 0.035 |
| **Solid Tumor** |  |  |  |  |  |  |
| No (Reference) | 34,988 | 96.6% | 8,915 | 25.5 (25.0 - 25.9) | - | . |
| Yes | 1,242 | 3.4% | 349 | 28.1 (25.6 - 30.7) | 1.13 (0.95 - 1.35) | 0.165 |
| **Valvular Disease** |  |  |  |  |  |  |
| No (Reference) | 33,593 | 92.7% | 8,473 | 25.2 (24.8 - 25.7) | - | . |
| Yes | 2,637 | 7.3% | 791 | 30.0 (28.3 - 31.8) | 0.95 (0.85 - 1.07) | 0.424 |
| **Weight Loss** |  |  |  |  |  |  |
| No (Reference) | 32,482 | 89.7% | 7,795 | 24.0 (23.5 - 24.5) | - | . |
| Yes | 3,748 | 10.3% | 1,469 | 39.2 (37.6 - 40.8) | 1.15 (1.04 - 1.26) | 0.005 |
| ^a^ Patients with missing values for ADI, Urban/Rural, Hospital Size and Ownership were dropped from the multivariate analysis. | | | | | | |
| ^b^ The South US census region includes counts for "Other" territories to avoid identification due to low counts. | | | | | | |
| **Notes:** Model evaluation metrics were ROC Area Under the Curve (0.856); Pearson Chi Sq Goodness of Fit test p value (1); Deviance Goodness of Fit test p value (1). | | | | | | |
| Intravenous Immunoglobulin/Hyperimmune Globulin Group has been omitted to avoid identification due to low counts. | | | | | | |

| **Supplementary Table 15.** Inpatient (IP) Mortality Rates and Corresponding Adjusted Odds Ratios (ORs) by Patient and Hospital Characteristics Among Recipients of COVID-19 Convalescent Plasma (CCP) with most recent COVID-19 Diagnosis Recorded in the 11-30 Days Prior to Hospital Admission | | | | | | |
| --- | --- | --- | --- | --- | --- | --- |
| **Patient and Hospital Characteristics** | **CCP Transfusion Stays #** | **CCP Transfusion Stays  %** | **IP Mortality  #** | **Crude IP Mortality Rate per 100  (95% CIs)** | **Adjusted ORs (95% CIs)** | **P-value** |
| **Overall** | 4,539 | 100.0% | 1,108 | 24.4 (23.2 - 25.7) | - | . |
| **Age (continuous)** | 4,539 | 100.0% | 1,108 | 24.4 (23.2 - 25.7) | 1.06 (1.05 - 1.07) | <.0001 |
| **Sex** |  |  |  |  |  |  |
| Female (Reference) | 2,035 | 44.8% | 448 | 22.0 (20.2 - 23.9) | - | . |
| Male | 2,504 | 55.2% | 660 | 26.4 (24.6 - 28.1) | 1.10 (0.92 - 1.33) | 0.284 |
| **ADI^a^** |  |  |  |  |  |  |
| 81 - 100 (Reference) | 770 | 17.0% | 204 | 26.5 (23.4 - 29.8) | - | . |
| 61 - 80 | 955 | 21.0% | 240 | 25.1 (22.4 - 28.0) | 0.91 (0.69 - 1.18) | 0.468 |
| 41 - 60 | 984 | 21.7% | 225 | 22.9 (20.3 - 25.6) | 0.81 (0.62 - 1.07) | 0.147 |
| 21 - 40 | 837 | 18.4% | 176 | 21.0 (18.3 - 24.0) | 0.67 (0.50 - 0.90) | 0.008 |
| 0 - 20 | 524 | 11.5% | 138 | 26.3 (22.7 - 30.4) | 0.85 (0.61 - 1.19) | 0.353 |
| **Medicaid Eligibility** |  |  |  |  |  |  |
| No (Reference) | 3,277 | 72.2% | 783 | 23.9 (22.4 - 25.4) | - | . |
| Yes | 1,262 | 27.8% | 325 | 25.8 (23.4 - 28.3) | 0.84 (0.67 - 1.05) | 0.133 |
| **Region of Residence^b^** |  |  |  |  |  |  |
| South (Reference) | 2,766 | 60.9% | 645 | 23.3 (21.8 - 24.9) | - | . |
| Midwest | 843 | 18.6% | 188 | 22.3 (19.6 - 25.3) | 0.93 (0.72 - 1.20) | 0.577 |
| West | 558 | 12.3% | 186 | 33.3 (29.5 - 37.4) | 1.37 (1.04 - 1.81) | 0.025 |
| Northeast | 372 | 8.2% | 89 | 23.9 (19.7 - 28.7) | 1.07 (0.76 - 1.51) | 0.695 |
| **Urban/Rural^a^** |  |  |  |  |  |  |
| Urban (Reference) | 2,999 | 66.1% | 733 | 24.4 (22.9 - 26.0) | - | . |
| Rural | 1,117 | 24.6% | 261 | 23.4 (20.9 - 26.0) | 0.94 (0.74 - 1.18) | 0.582 |
| **Immunocompromised Status** |  |  |  |  |  |  |
| No (Reference) | 3,782 | 83.3% | 918 | 24.3 (22.9 - 25.7) | - | . |
| Yes | 757 | 16.7% | 190 | 25.1 (22.1 - 28.4) | 0.97 (0.75 - 1.27) | 0.830 |
| **Nursing Home Stay** |  |  |  |  |  |  |
| No (Reference) | 3,430 | 75.6% | 762 | 22.2 (20.8 - 23.7) | - | . |
| Yes | 1,109 | 24.4% | 346 | 31.2 (28.5 - 34.0) | 1.34 (1.08 - 1.67) | 0.009 |
| **Emergency Room Stay** |  |  |  |  |  |  |
| No (Reference) | 657 | 14.5% | 194 | 29.5 (26.1 - 33.2) | - | . |
| Yes | 3,882 | 85.5% | 914 | 23.5 (22.2 - 24.9) | 0.85 (0.64 - 1.12) | 0.252 |
| **COVID-19 As a Reason for Admission** |  |  |  |  |  |  |
| No (Reference) | 2,229 | 49.1% | 586 | 26.3 (24.5 - 28.2) | - | . |
| Yes | 2,310 | 50.9% | 522 | 22.6 (20.9 - 24.4) | 0.98 (0.82 - 1.17) | 0.831 |
| **Thrombosis on Index** |  |  |  |  |  |  |
| No (Reference) | 3,812 | 84.0% | 844 | 22.1 (20.8 - 23.5) | - | . |
| Yes | 727 | 16.0% | 264 | 36.3 (32.8 - 39.9) | 1.42 (1.13 - 1.79) | 0.002 |
| **Hospital Ownership^a^** |  |  |  |  |  |  |
| Government (Reference) | 600 | 13.2% | 147 | 24.5 (21.1 – 28.2) | - | . |
| For Profit | 1,091 | 24.0% | 272 | 24.9 (22.4 - 27.6) | 0.66 (0.49 - 0.90) | 0.009 |
| Non-Profit | 2,834 | 62.4% | 683 | 24.1 (22.5 - 25.7) | 0.85 (0.65 - 1.12) | 0.254 |
| **Hospital Size^a^** |  |  |  |  |  |  |
| Small (< 100 beds) (Reference) | 508 | 11.2% | 101 | 19.9 (16.6 - 23.7) | - | . |
| Medium (100 - 500 beds) | 2,961 | 65.2% | 740 | 25.0 (23.4 - 26.6) | 0.96 (0.70 - 1.33) | 0.822 |
| Large (> 500 beds) | 866 | 19.1% | 229 | 26.4 (23.6 - 29.5) | 0.94 (0.65 - 1.35) | 0.744 |
| **ICU/CCU Stay** |  |  |  |  |  |  |
| No (Reference) | 1,790 | 39.4% | 173 | 9.7 (8.4 - 11.2) | - | . |
| Yes | 2,749 | 60.6% | 935 | 34.0 (32.2 - 35.8) | 2.62 (2.10 - 3.27) | <.0001 |
| **Mechanical Ventilation** |  |  |  |  |  |  |
| No (Reference) | 3,856 | 85.0% | 620 | 16.1 (14.9 - 17.3) | - | . |
| Less Than 96hrs | 264 | 5.8% | 197 | 74.6 (68.8 - 79.7) | 13.77 (9.78 - 19.41) | <.0001 |
| More Than 96hrs | 419 | 9.2% | 291 | 69.5 (64.8 - 73.8) | 10.41 (7.85 - 13.80) | <.0001 |
| **Tocilizumab** |  |  |  |  |  |  |
| No (Reference) | 4,413 | 97.2% | 1,066 | 24.2 (22.9 - 25.5) | - | . |
| Yes | 126 | 2.8% | 42 | 33.3 (25.3 - 42.4) | 1.45 (0.88 - 2.39) | 0.149 |
| **Baricitinib** |  |  |  |  |  |  |
| No (Reference) | 4,509 | 99.3% | 1,097 | 24.3 (23.1 - 25.6) | - | . |
| Yes | 30 | 0.7% | 11 | 36.7 (20.5 - 56.1) | 2.60 (1.03 - 6.54) | 0.042 |
| **Remdesivir** |  |  |  |  |  |  |
| No (Reference) | 1,692 | 37.3% | 436 | 25.8 (23.7 - 27.9) | - | . |
| Yes | 2,847 | 62.7% | 672 | 23.6 (22.1 - 25.2) | 0.98 (0.81 - 1.18) | 0.838 |
| **Charlson Comorbidity Index (CCI)** |  |  |  |  |  |  |
| 0 (Reference) | 623 | 13.7% | 98 | 15.7 (13.0 - 18.9) | - | . |
| 1 | 1,013 | 22.3% | 202 | 19.9 (17.5 - 22.6) | 1.12 (0.79 - 1.59) | 0.520 |
| 2 | 916 | 20.2% | 231 | 25.2 (22.5 - 28.2) | 1.31 (0.91 - 1.89) | 0.147 |
| 3 - 4 | 1,160 | 25.6% | 324 | 27.9 (25.4 - 30.6) | 1.28 (0.87 - 1.89) | 0.206 |
| 5+ | 827 | 18.2% | 253 | 30.6 (27.5 - 33.9) | 1.45 (0.90 - 2.35) | 0.125 |
| **90-Day Comorbidity History** |  |  |  |  |  |  |
| **Thrombosis History** |  |  |  |  |  |  |
| No (Reference) | 4,213 | 92.8% | 996 | 23.6 (22.4 - 25.0) | - | . |
| Yes | 326 | 7.2% | 112 | 34.4 (29.3 - 39.8) | 1.26 (0.91 - 1.74) | 0.167 |
| **Elixhauser Comorbidity History** |  |  |  |  |  |  |
| **Alcohol Abuse** |  |  |  |  |  |  |
| No (Reference) | 4,446 | 98.0% | 1,075 | 24.2 (22.9 - 25.5) | - | . |
| Yes | 93 | 2.0% | 33 | 35.5 (26.0 - 46.2) | 1.49 (0.84 - 2.63) | 0.172 |
| **Cardiac Arrhythmia** |  |  |  |  |  |  |
| No (Reference) | 2,664 | 58.7% | 507 | 19.0 (17.6 - 20.6) | - | . |
| Yes | 1,875 | 41.3% | 601 | 32.1 (30.0 - 34.2) | 1.32 (1.09 - 1.58) | 0.003 |
| **Coagulopathy** |  |  |  |  |  |  |
| No (Reference) | 3,604 | 79.4% | 754 | 20.9 (19.6 - 22.3) | - | . |
| Yes | 935 | 20.6% | 354 | 37.9 (34.8 - 41.1) | 1.52 (1.24 - 1.87) | <.0001 |
| **Congestive Heart Failure** |  |  |  |  |  |  |
| No (Reference) | 3,077 | 67.8% | 664 | 21.6 (20.1 - 23.1) | - | . |
| Yes | 1,462 | 32.2% | 444 | 30.4 (28.0 - 32.8) | 0.95 (0.74 - 1.22) | 0.690 |
| **Diabetes** |  |  |  |  |  |  |
| No (Reference) | 2,522 | 55.6% | 581 | 23.0 (21.4 - 24.7) | - | . |
| Yes | 2,017 | 44.4% | 527 | 26.1 (24.2 - 28.1) | 0.82 (0.66 - 1.01) | 0.066 |
| **Fluid and Electrolyte Disorder** |  |  |  |  |  |  |
| No (Reference) | 1,490 | 32.8% | 213 | 14.3 (12.6 - 16.2) | - | . |
| Yes | 3,049 | 67.2% | 895 | 29.4 (27.7 - 31.0) | 1.34 (1.08 - 1.66) | 0.008 |
| **Hypertension (uncomplicated)** |  |  |  |  |  |  |
| No (Reference) | 2,403 | 52.9% | 620 | 25.8 (24.1 - 27.6) | - | . |
| Yes | 2,136 | 47.1% | 488 | 22.8 (21.1 - 24.7) | 1.03 (0.83 - 1.28) | 0.758 |
| **Hypertension (complicated)** |  |  |  |  |  |  |
| No (Reference) | 2,390 | 52.7% | 486 | 20.3 (18.7 - 22.0) | - | . |
| Yes | 2,149 | 47.3% | 622 | 28.9 (27.0 - 30.9) | 1.25 (0.95 - 1.65) | 0.110 |
| **Liver Disease** |  |  |  |  |  |  |
| No (Reference) | 4,308 | 94.9% | 1,013 | 23.5 (22.3 - 24.8) | - | . |
| Yes | 231 | 5.1% | 95 | 41.1 (34.8 - 47.8) | 1.51 (1.02 - 2.23) | 0.038 |
| **Metastatic Cancer** |  |  |  |  |  |  |
| No (Reference) | 4,470 | 98.5% | 1,083 | 24.2 (23.0 - 25.5) | - | . |
| Yes | 69 | 1.5% | 25 | 36.2 (25.3 - 48.8) | 1.58 (0.76 - 3.29) | 0.220 |
| **Other Neurological Disorders** |  |  |  |  |  |  |
| No (Reference) | 3,110 | 68.5% | 623 | 20.0 (18.6 - 21.5) | - | . |
| Yes | 1,429 | 31.5% | 485 | 33.9 (31.5 - 36.5) | 1.37 (1.13 - 1.66) | 0.001 |
| **Paralysis** |  |  |  |  |  |  |
| No (Reference) | 4,442 | 97.9% | 1,076 | 24.2 (23.0 - 25.5) | - | . |
| Yes | 97 | 2.1% | 32 | 33.0 (24.0 - 43.4) | 0.84 (0.46 - 1.54) | 0.566 |
| **Renal Failure** |  |  |  |  |  |  |
| No (Reference) | 3,473 | 76.5% | 793 | 22.8 (21.5 - 24.3) | - | . |
| Yes | 1,066 | 23.5% | 315 | 29.5 (26.8 - 32.4) | 0.95 (0.73 - 1.24) | 0.716 |
| **Solid Tumor** |  |  |  |  |  |  |
| No (Reference) | 4,348 | 95.8% | 1,047 | 24.1 (22.8 - 25.4) | - | . |
| Yes | 191 | 4.2% | 61 | 31.9 (25.5 - 39.1) | 0.92 (0.58 - 1.46) | 0.712 |
| **Weight Loss** |  |  |  |  |  |  |
| No (Reference) | 3,825 | 84.3% | 856 | 22.4 (21.1 - 23.7) | - | . |
| Yes | 714 | 15.7% | 252 | 35.3 (31.8 - 38.9) | 1.08 (0.86 - 1.36) | 0.495 |
| ^a^ Patients with missing values for ADI, Urban/Rural, Hospital Size and Ownership were dropped from the multivariate analysis. | | | | | | |
| ^b^ The South US census region includes counts for "Other" territories to avoid identification due to low counts. | | | | | | |
| **Notes:** Model evaluation metrics were ROC Area Under the Curve (0.8387); Pearson Chi Sq Goodness of Fit test p value (0.9894); Deviance Goodness of Fit test p value (1). | | | | | | |
| Intravenous Immunoglobulin/Hyperimmune Globulin Group has been omitted to avoid identification due to low counts. | | | | | | |

| **Supplementary Table 16.** Inpatient (IP) Mortality Rates and Corresponding Adjusted Odds Ratios (ORs) by Patient and Hospital Characteristics Among Medicaid Eligible Recipients of COVID-19 Convalescent Plasma (CCP) | | | | | |  |
| --- | --- | --- | --- | --- | --- | --- |
| **Patient and Hospital Characteristics** | **CCP Transfusion Stays #** | **CCP Transfusion Stays  %** | **IP Mortality  #** | **Crude IP Mortality Rate per 100  (95% CIs)** | **Adjusted ORs (95% CIs)** | **P-value** |
| **Overall** | 16,837 | 100.0% | 4,983 | 29.6 (28.9 - 30.3) | - | . |
| **Age (continuous)** | 16,837 | 100.0% | 4,983 | 29.6 (28.9 - 30.3) | 1.05 (1.05 - 1.06) | <.0001 |
| **Sex** |  |  |  |  |  |  |
| Female (Reference) | 9,521 | 56.5% | 2,634 | 27.7 (26.8 - 28.6) | - | . |
| Male | 7,316 | 43.5% | 2,349 | 32.1 (31.0 - 33.2) | 1.15 (1.05 - 1.25) | 0.002 |
| **ADI^a^** |  |  |  |  |  |  |
| 81 - 100 (Reference) | 4,116 | 24.4% | 1,204 | 29.3 (27.9 - 30.7) | - | . |
| 61 - 80 | 3,671 | 21.8% | 1,030 | 28.1 (26.6 - 29.6) | 0.99 (0.88 - 1.12) | 0.895 |
| 41 - 60 | 2,876 | 17.1% | 843 | 29.3 (27.7 - 31.0) | 1.06 (0.93 - 1.21) | 0.352 |
| 21 - 40 | 2,450 | 14.6% | 747 | 30.5 (28.7 - 32.4) | 0.92 (0.80 - 1.06) | 0.249 |
| 0 - 20 | 1,882 | 11.2% | 615 | 32.7 (30.6 - 34.9) | 0.93 (0.80 - 1.09) | 0.354 |
| **Region of Residence^b^** |  |  |  |  |  |  |
| South (Reference) | 8,464 | 50.3% | 2,345 | 27.7 (26.8 - 28.7) | - | . |
| Midwest | 3,033 | 18.0% | 799 | 26.3 (24.8 - 28.0) | 1.01 (0.89 - 1.15) | 0.859 |
| West | 3,479 | 20.7% | 1,277 | 36.7 (35.1 - 38.3) | 1.38 (1.22 - 1.56) | <.0001 |
| Northeast | 1,861 | 11.1% | 562 | 30.2 (28.1 - 32.4) | 1.42 (1.22 - 1.64) | <.0001 |
| **Urban/Rural^a^** |  |  |  |  |  |  |
| Urban (Reference) | 11,694 | 69.5% | 3,479 | 29.8 (28.9 - 30.6) | - | . |
| Rural | 3,524 | 20.9% | 1,017 | 28.9 (27.4 - 30.4) | 1.26 (1.12 - 1.42) | 0.000 |
| **Immunocompromised Status** |  |  |  |  |  |  |
| No (Reference) | 14,962 | 88.9% | 4,409 | 29.5 (28.7 - 30.2) | - | . |
| Yes | 1,875 | 11.1% | 574 | 30.6 (28.5 - 32.8) | 0.98 (0.85 - 1.14) | 0.819 |
| **Nursing Home Stay** |  |  |  |  |  |  |
| No (Reference) | 12,357 | 73.4% | 3,582 | 29.0 (28.2 - 29.8) | - | . |
| Yes | 4,480 | 26.6% | 1,401 | 31.3 (29.9 - 32.7) | 1.15 (1.04 - 1.27) | 0.008 |
| **Emergency Room Stay** |  |  |  |  |  |  |
| No (Reference) | 1,705 | 10.1% | 532 | 31.2 (29.0 - 33.5) | - | . |
| Yes | 15,132 | 89.9% | 4,451 | 29.4 (28.7 - 30.1) | 0.94 (0.80 - 1.09) | 0.411 |
| **COVID-19 As a Reason for Admission** |  |  |  |  |  |  |
| No (Reference) | 8,732 | 51.9% | 2,736 | 31.3 (30.4 - 32.3) | - | . |
| Yes | 8,105 | 48.1% | 2,247 | 27.7 (26.8 - 28.7) | 0.97 (0.89 - 1.06) | 0.482 |
| **Time Since Most Recent COVID-19 Diagnosis in the 30 Days Prior to Admission** |  |  |  |  |  |  |
| 0 days (Reference)^c^ | 8,735 | 51.9% | 2,550 | 29.2 (28.2 - 30.2) | - | . |
| 1 - 3 days | 3,827 | 22.7% | 1,096 | 28.6 (27.2 - 30.1) | 1.02 (0.91 - 1.14) | 0.738 |
| 4 - 10 days | 3,013 | 17.9% | 1,012 | 33.6 (31.9 - 35.3) | 1.27 (1.13 - 1.43) | <.0001 |
| 11-30 days | 1,262 | 7.5% | 325 | 25.8 (23.4 - 28.3) | 0.78 (0.66 - 0.93) | 0.006 |
| **Thrombosis on Index** |  |  |  |  |  |  |
| No (Reference) | 14,383 | 85.4% | 3,907 | 27.2 (26.4 - 27.9) | - | . |
| Yes | 2,454 | 14.6% | 1,076 | 43.8 (41.9 - 45.8) | 1.24 (1.09 - 1.40) | 0.001 |
| **Hospital Ownership^a^** |  |  |  |  |  |  |
| Government (Reference) | 2,144 | 12.7% | 602 | 28.1 (26.2 - 30.0) | - | . |
| For Profit | 3,982 | 23.7% | 1,237 | 31.1 (29.6 - 32.5) | 0.83 (0.71 - 0.96) | 0.015 |
| Non-Profit | 10,680 | 63.4% | 3,135 | 29.4 (28.5 - 30.2) | 0.90 (0.79 – 1.03) | 0.123 |
| **Hospital Size^a^** |  |  |  |  |  |  |
| Small (< 100 beds) (Reference) | 1,754 | 10.4% | 416 | 23.7 (21.8 - 25.8) | - | . |
| Medium (100 - 500 beds) | 11,198 | 66.5% | 3,379 | 30.2 (29.3 - 31.0) | 1.04 (0.89 - 1.21) | 0.642 |
| Large (> 500 beds) | 3,196 | 19.0% | 988 | 30.9 (29.3 - 32.6) | 1.02 (0.86 - 1.22) | 0.795 |
| **ICU/CCU Stay** |  |  |  |  |  |  |
| No (Reference) | 6,320 | 37.5% | 873 | 13.8 (13.0 - 14.7) | - | . |
| Yes | 10,517 | 62.5% | 4,110 | 39.1 (38.1 - 40.0) | 2.22 (2.00 - 2.46) | <.0001 |
| **Mechanical Ventilation** |  |  |  |  |  |  |
| No (Reference) | 13,619 | 80.9% | 2,643 | 19.4 (18.7 - 20.1) | - | . |
| Less Than 96hrs | 1,125 | 6.7% | 833 | 74.0 (71.4 - 76.6) | 11.09 (9.41 - 13.05) | <.0001 |
| More Than 96hrs | 2,093 | 12.4% | 1,507 | 72.0 (70.0 - 73.9) | 9.09 (7.99 - 10.35) | <.0001 |
| **Tocilizumab** |  |  |  |  |  |  |
| No (Reference) | 16,336 | 97.0% | 4,769 | 29.2 (28.5 - 29.9) | - | . |
| Yes | 501 | 3.0% | 214 | 42.7 (38.4 - 47.2) | 1.33 (1.05 - 1.68) | 0.016 |
| **Baricitinib** |  |  |  |  |  |  |
| No (Reference) | 16,714 | 99.3% | 4,934 | 29.5 (28.8 - 30.2) | - | . |
| Yes | 123 | 0.7% | 49 | 39.8 (31.2 - 49.1) | 1.52 (0.93 - 2.49) | 0.094 |
| **Remdesivir** |  |  |  |  |  |  |
| No (Reference) | 5,015 | 29.8% | 1,603 | 32.0 (30.7 - 33.3) | - | . |
| Yes | 11,822 | 70.2% | 3,380 | 28.6 (27.8 - 29.4) | 0.91 (0.83 - 1.00) | 0.059 |
| **Charlson Comorbidity Index (CCI)** |  |  |  |  |  |  |
| 0 (Reference) | 1,611 | 9.6% | 359 | 22.3 (20.3 - 24.4) | - | . |
| 1 | 3,639 | 21.6% | 931 | 25.6 (24.2 - 27.0) | 1.12 (0.93 - 1.34) | 0.229 |
| 2 | 3,317 | 19.7% | 953 | 28.7 (27.2 - 30.3) | 1.13 (0.94 - 1.37) | 0.194 |
| 3 - 4 | 4,533 | 26.9% | 1,438 | 31.7 (30.4 - 33.1) | 1.18 (0.97 - 1.45) | 0.099 |
| 5+ | 3,737 | 22.2% | 1,302 | 34.8 (33.3 - 36.4) | 1.18 (0.93 - 1.51) | 0.176 |
| **90-Day Comorbidity History** |  |  |  |  |  |  |
| **Thrombosis History** |  |  |  |  |  |  |
| No (Reference) | 16,311 | 96.9% | 4,807 | 29.5 (28.8 - 30.2) | - | . |
| Yes | 526 | 3.1% | 176 | 33.5 (29.5 - 37.7) | 0.99 (0.78 - 1.26) | 0.932 |
| **Elixhauser Comorbidity History** |  |  |  |  |  |  |
| **Cardiac Arrhythmia** |  |  |  |  |  |  |
| No (Reference) | 10,806 | 64.2% | 2,741 | 25.4 (24.5 - 26.2) | - | . |
| Yes | 6,031 | 35.8% | 2,242 | 37.2 (36.0 - 38.4) | 1.30 (1.19 - 1.42) | <.0001 |
| **Coagulopathy** |  |  |  |  |  |  |
| No (Reference) | 13,792 | 81.9% | 3,759 | 27.3 (26.5 - 28.0) | - | . |
| Yes | 3,045 | 18.1% | 1,224 | 40.2 (38.5 - 42.0) | 1.22 (1.09 - 1.36) | 0.000 |
| **Congestive Heart Failure** |  |  |  |  |  |  |
| No (Reference) | 11,150 | 66.2% | 3,082 | 27.6 (26.8 - 28.5) | - | . |
| Yes | 5,687 | 33.8% | 1,901 | 33.4 (32.2 - 34.7) | 1.05 (0.93 - 1.18) | 0.420 |
| **Deficiency Anemia** |  |  |  |  |  |  |
| No (Reference) | 15,869 | 94.3% | 4,725 | 29.8 (29.1 - 30.5) | - | . |
| Yes | 968 | 5.7% | 258 | 26.7 (23.9 - 29.6) | 0.77 (0.64 - 0.93) | 0.006 |
| **Depression** |  |  |  |  |  |  |
| No (Reference) | 13,694 | 81.3% | 4,151 | 30.3 (29.5 - 31.1) | - | . |
| Yes | 3,143 | 18.7% | 832 | 26.5 (24.9 - 28.1) | 0.97 (0.87 - 1.09) | 0.608 |
| **Diabetes** |  |  |  |  |  |  |
| No (Reference) | 7,986 | 47.4% | 2,272 | 28.4 (27.5 - 29.5) | - | . |
| Yes | 8,851 | 52.6% | 2,711 | 30.6 (29.7 - 31.6) | 1.01 (0.91 - 1.12) | 0.846 |
| **Fluid and Electrolyte Disorder** |  |  |  |  |  |  |
| No (Reference) | 5,457 | 32.4% | 1,027 | 18.8 (17.8 - 19.9) | - | . |
| Yes | 11,380 | 67.6% | 3,956 | 34.8 (33.9 - 35.6) | 1.42 (1.29 - 1.57) | <.0001 |
| **Hypertension (uncomplicated)** |  |  |  |  |  |  |
| No (Reference) | 10,151 | 60.3% | 3,244 | 32.0 (31.1 - 32.9) | - | . |
| Yes | 6,686 | 39.7% | 1,739 | 26.0 (25.0 - 27.1) | 0.82 (0.73 - 0.92) | 0.001 |
| **Hypertension (complicated)** |  |  |  |  |  |  |
| No (Reference) | 8,598 | 51.1% | 2,316 | 26.9 (26.0 - 27.9) | - | . |
| Yes | 8,239 | 48.9% | 2,667 | 32.4 (31.4 - 33.4) | 0.91 (0.79 - 1.05) | 0.185 |
| **Liver Disease** |  |  |  |  |  |  |
| No (Reference) | 15,857 | 94.2% | 4,548 | 28.7 (28.0 - 29.4) | - | . |
| Yes | 980 | 5.8% | 435 | 44.4 (41.3 - 47.6) | 1.38 (1.16 - 1.64) | 0.000 |
| **Obesity** |  |  |  |  |  |  |
| No (Reference) | 12,860 | 76.4% | 3,860 | 30.0 (29.2 - 30.8) | - | . |
| Yes | 3,977 | 23.6% | 1,123 | 28.2 (26.8 - 29.7) | 0.94 (0.84 - 1.05) | 0.253 |
| **Other Neurological Disorders** |  |  |  |  |  |  |
| No (Reference) | 11,146 | 66.2% | 2,944 | 26.4 (25.6 - 27.2) | - | . |
| Yes | 5,691 | 33.8% | 2,039 | 35.8 (34.6 - 37.1) | 1.18 (1.08 - 1.29) | 0.000 |
| **Pulmonary Circulation Disorder** |  |  |  |  |  |  |
| No (Reference) | 16,391 | 97.4% | 4,826 | 29.4 (28.7 - 30.1) | - | . |
| Yes | 446 | 2.6% | 157 | 35.2 (30.8 - 39.9) | 1.02 (0.77 - 1.33) | 0.910 |
| **Renal Failure** |  |  |  |  |  |  |
| No (Reference) | 12,655 | 75.2% | 3,531 | 27.9 (27.1 - 28.7) | - | . |
| Yes | 4,182 | 24.8% | 1,452 | 34.7 (33.3 - 36.2) | 1.08 (0.95 - 1.24) | 0.246 |
| **Rheumatoid Arthritis/Collagen Vascular Diseases (CVD)** |  |  |  |  |  |  |
| No (Reference) | 16,209 | 96.3% | 4,779 | 29.5 (28.8 - 30.2) | - | . |
| Yes | 628 | 3.7% | 204 | 32.5 (28.9 - 36.3) | 1.27 (1.02 - 1.58) | 0.032 |
| **Weight Loss** |  |  |  |  |  |  |
| No (Reference) | 14,587 | 86.6% | 4,116 | 28.2 (27.5 - 29.0) | - | . |
| Yes | 2,250 | 13.4% | 867 | 38.5 (36.5 - 40.6) | 1.05 (0.93 - 1.19) | 0.428 |
| ^a^ Patients with missing values for ADI, Urban/Rural, Hospital Size and Ownership were dropped from the multivariate analysis. | | | | | | |
| ^b^ The South US census region includes counts for "Other" territories to avoid identification due to low counts. | | | | | | |
| ^c^ Patients have not had COVID-19 diagnosis recorded in the 30 days prior to admission. | | | | | | |
| **Notes:** Model evaluation metrics were ROC Area Under the Curve (0.8106); Pearson Chi Sq Goodness of Fit test p value (0.8866); Deviance Goodness of Fit test p value (1). | | | | | | |
| Intravenous Immunoglobulin/Hyperimmune Globulin and Peptic Ulcer Disease Groups have been omitted to avoid identification due to low counts. | | | | | | |

| **Supplementary Table 17.** Inpatient (IP) Mortality Rates and Corresponding Adjusted Odds Ratios (ORs) by Patient and Hospital Characteristics Among Non-Medicaid Eligible Recipients of COVID-19 Convalescent Plasma (CCP) | | | | | |  |
| --- | --- | --- | --- | --- | --- | --- |
| **Patient and Hospital Characteristics** | **CCP Transfusion Stays #** | **CCP Transfusion Stays  %** | **IP Mortality  #** | **Crude IP Mortality Rate per 100  (95% CIs)** | **Adjusted ORs (95% CIs)** | **P-value** |
| **Overall** | 64,914 | 100.0% | 15,893 | 24.5 (24.2 - 24.8) | - | . |
| **Age (continuous)** | 64,914 | 100.0% | 15,893 | 24.5 (24.2 - 24.8) | 1.06 (1.06 - 1.07) | <.0001 |
| **Sex** |  |  |  |  |  |  |
| Female (Reference) | 27,817 | 42.9% | 5,845 | 21.0 (20.5 - 21.5) | - | . |
| Male | 37,097 | 57.1% | 10,048 | 27.1 (26.6 - 27.5) | 1.22 (1.16 - 1.28) | <.0001 |
| **ADI^a^** |  |  |  |  |  |  |
| 81 - 100 (Reference) | 8,314 | 12.8% | 2,207 | 26.5 (25.6 - 27.5) | - | . |
| 61 – 80 | 13,912 | 21.4% | 3,497 | 25.1 (24.4 - 25.9) | 1.00 (0.93 - 1.09) | 0.945 |
| 41 – 60 | 15,338 | 23.6% | 3,624 | 23.6 (23.0 - 24.3) | 0.92 (0.85 - 0.99) | 0.031 |
| 21 – 40 | 13,593 | 20.9% | 3,177 | 23.4 (22.7 - 24.1) | 0.84 (0.77 - 0.91) | <.0001 |
| 0 – 20 | 8,169 | 12.6% | 1,902 | 23.3 (22.4 - 24.2) | 0.83 (0.75 - 0.91) | <.0001 |
| **Region of Residence^b^** |  |  |  |  |  |  |
| South (Reference) | 35,704 | 55.0% | 8,709 | 24.4 (23.9 - 24.8) | - | . |
| Midwest | 16,409 | 25.3% | 3,853 | 23.5 (22.8 - 24.1) | 0.98 (0.92 - 1.04) | 0.508 |
| West | 7,017 | 10.8% | 1,856 | 26.5 (25.4 - 27.5) | 1.09 (1.01 - 1.19) | 0.032 |
| Northeast | 5,784 | 8.9% | 1,475 | 25.5 (24.4 - 26.6) | 1.11 (1.01 - 1.21) | 0.027 |
| **Urban/Rural^a^** |  |  |  |  |  |  |
| Urban (Reference) | 44,002 | 67.8% | 10,731 | 24.4 (24.0 - 24.8) | - | . |
| Rural | 15,775 | 24.3% | 3,807 | 24.1 (23.5 - 24.8) | 1.09 (1.02 - 1.16) | 0.008 |
| **Immunocompromised Status** |  |  |  |  |  |  |
| No (Reference) | 58,642 | 90.3% | 14,163 | 24.2 (23.8 - 24.5) | - | . |
| Yes | 6,272 | 9.7% | 1,730 | 27.6 (26.5 - 28.7) | 1.15 (1.05 - 1.27) | 0.003 |
| **Nursing Home Stay** |  |  |  |  |  |  |
| No (Reference) | 62,156 | 95.8% | 14,965 | 24.1 (23.7 - 24.4) | - | . |
| Yes | 2,758 | 4.2% | 928 | 33.6 (31.9 - 35.5) | 1.25 (1.13 - 1.39) | <.0001 |
| **Emergency Room Stay** |  |  |  |  |  |  |
| No (Reference) | 7,100 | 10.9% | 1,920 | 27.0 (26.0 - 28.1) | - | . |
| Yes | 57,814 | 89.1% | 13,973 | 24.2 (23.8 - 24.5) | 0.95 (0.87 - 1.03) | 0.236 |
| **COVID-19 As a Reason for Admission** |  |  |  |  |  |  |
| No (Reference) | 30,385 | 46.8% | 7,872 | 25.9 (25.4 - 26.4) | - | . |
| Yes | 34,529 | 53.2% | 8,021 | 23.2 (22.8 - 23.7) | 1.05 (1.00 - 1.11) | 0.031 |
| **Time Since Most Recent COVID-19 Diagnosis in the 30 Days Prior to Admission** |  |  |  |  |  |  |
| 0 days (Reference)^c^ | 32,247 | 49.7% | 7,954 | 24.7 (24.2 - 25.1) | - | . |
| 1 - 3 days | 15,570 | 24.0% | 3,695 | 23.7 (23.1 - 24.4) | 0.97 (0.91 - 1.03) | 0.318 |
| 4 - 10 days | 13,820 | 21.3% | 3,461 | 25.0 (24.3 - 25.8) | 1.03 (0.96 - 1.09) | 0.405 |
| 11-30 days | 3,277 | 5.0% | 783 | 23.9 (22.4 - 25.4) | 0.86 (0.76 - 0.96) | 0.007 |
| **Thrombosis on Index** |  |  |  |  |  |  |
| No (Reference) | 55,839 | 86.0% | 12,153 | 21.8 (21.4 - 22.1) | - | . |
| Yes | 9,075 | 14.0% | 3,740 | 41.2 (40.2 - 42.2) | 1.38 (1.29 - 1.49) | <.0001 |
| **Hospital Ownership^a^** |  |  |  |  |  |  |
| Government (Reference) | 8,317 | 12.8% | 2,051 | 24.7 (23.7 – 25.6) | - | . |
| For Profit | 12,516 | 19.3% | 3,117 | 24.9 (24.2 – 25.7) | 0.79 (0.72 - 0.86) | <.0001 |
| Non-Profit | 43,960 | 67.7% | 10,702 | 24.3 (23.9 – 24.7) | 0.85 (0.79 – 0.92) | <.0001 |
| **Hospital Size^a^** |  |  |  |  |  |  |
| Small (< 100 beds) (Reference) | 7,926 | 12.2% | 1,479 | 18.7 (17.8 - 19.5) | - | . |
| Medium (100 - 500 beds) | 41,307 | 63.6% | 10,361 | 25.1 (24.7 - 25.5) | 1.16 (1.07 - 1.26) | 0.000 |
| Large (> 500 beds) | 12,442 | 19.2% | 3,289 | 26.4 (25.7 - 27.2) | 1.11 (1.01 - 1.22) | 0.024 |
| **ICU/CCU Stay** |  |  |  |  |  |  |
| No (Reference) | 27,033 | 41.6% | 2,271 | 8.4 (8.1 - 8.7) | - | . |
| Yes | 37,881 | 58.4% | 13,622 | 36.0 (35.5 - 36.4) | 2.88 (2.72 - 3.06) | <.0001 |
| **Mechanical Ventilation** |  |  |  |  |  |  |
| No (Reference) | 53,865 | 83.0% | 7,687 | 14.3 (14.0 - 14.6) | - | . |
| Less Than 96hrs | 3,706 | 5.7% | 2,766 | 74.6 (73.2 - 76.0) | 13.82 (12.61 - 15.15) | <.0001 |
| More Than 96hrs | 7,343 | 11.3% | 5,440 | 74.1 (73.1 - 75.1) | 12.44 (11.57 - 13.36) | <.0001 |
| **Tocilizumab** |  |  |  |  |  |  |
| No (Reference) | 62,650 | 96.5% | 14,952 | 23.9 (23.5 - 24.2) | - | . |
| Yes | 2,264 | 3.5% | 941 | 41.6 (39.5 - 43.6) | 1.73 (1.54 - 1.95) | <.0001 |
| **Baricitinib** |  |  |  |  |  |  |
| No (Reference) | 64,255 | 99.0% | 15,688 | 24.4 (24.1 - 24.7) | - | . |
| Yes | 659 | 1.0% | 205 | 31.1 (27.6 - 34.8) | 1.49 (1.20 - 1.86) | 0.000 |
| **Remdesivir** |  |  |  |  |  |  |
| No (Reference) | 14,898 | 23.0% | 3,828 | 25.7 (25.0 - 26.4) | - | . |
| Yes | 50,016 | 77.0% | 12,065 | 24.1 (23.7 - 24.5) | 0.96 (0.90 - 1.02) | 0.148 |
| **Charlson Comorbidity Index (CCI)** |  |  |  |  |  |  |
| 0 (Reference) | 12,667 | 19.5% | 2,166 | 17.1 (16.4 - 17.8) | - | . |
| 1 | 16,864 | 26.0% | 3,490 | 20.7 (20.1 - 21.3) | 1.08 (1.00 - 1.18) | 0.063 |
| 2 | 12,623 | 19.4% | 3,220 | 25.5 (24.8 - 26.3) | 1.23 (1.11 - 1.36) | <.0001 |
| 3 – 4 | 14,242 | 21.9% | 4,198 | 29.5 (28.7 - 30.2) | 1.22 (1.09 - 1.37) | 0.001 |
| 5+ | 8,518 | 13.1% | 2,819 | 33.1 (32.1 - 34.1) | 1.34 (1.15 - 1.56) | 0.000 |
| **90-Day Comorbidity History** |  |  |  |  |  |  |
| **Thrombosis History** |  |  |  |  |  |  |
| No (Reference) | 63,123 | 97.2% | 15,284 | 24.2 (23.9 - 24.5) | - | . |
| Yes | 1,791 | 2.8% | 609 | 34.0 (31.8 - 36.3) | 0.99 (0.86 - 1.13) | 0.853 |
| **ESRD History** |  |  |  |  |  |  |
| No (Reference) | 63,519 | 97.9% | 15,482 | 24.4 (24.0 - 24.7) | - | . |
| Yes | 1,395 | 2.1% | 411 | 29.5 (27.1 - 31.9) | 0.92 (0.76 - 1.10) | 0.359 |
| **Elixhauser Comorbidity History** |  |  |  |  |  |  |
| **Alcohol Abuse** |  |  |  |  |  |  |
| No (Reference) | 64,051 | 98.7% | 15,649 | 24.4 (24.1 - 24.8) | - | . |
| Yes | 863 | 1.3% | 244 | 28.3 (25.3 - 31.4) | 0.86 (0.70 - 1.05) | 0.145 |
| **Blood Loss Anemia** |  |  |  |  |  |  |
| No (Reference) | 64,623 | 99.6% | 15,784 | 24.4 (24.1 - 24.8) | - | . |
| Yes | 291 | 0.4% | 109 | 37.5 (31.9 - 43.3) | 1.38 (1.01 - 1.89) | 0.044 |
| **Cardiac Arrhythmia** |  |  |  |  |  |  |
| No (Reference) | 40,758 | 62.8% | 7,801 | 19.1 (18.8 - 19.5) | - | . |
| Yes | 24,156 | 37.2% | 8,092 | 33.5 (32.9 - 34.1) | 1.36 (1.29 - 1.43) | <.0001 |
| **Chronic Pulmonary Disease** |  |  |  |  |  |  |
| No (Reference) | 44,896 | 69.2% | 10,816 | 24.1 (23.7 - 24.5) | - | . |
| Yes | 20,018 | 30.8% | 5,077 | 25.4 (24.8 - 26.0) | 1.04 (0.98 - 1.10) | 0.197 |
| **Coagulopathy** |  |  |  |  |  |  |
| No (Reference) | 53,688 | 82.7% | 11,676 | 21.7 (21.4 - 22.1) | - | . |
| Yes | 11,226 | 17.3% | 4,217 | 37.6 (36.7 - 38.5) | 1.31 (1.24 - 1.39) | <.0001 |
| **Congestive Heart Failure** |  |  |  |  |  |  |
| No (Reference) | 48,028 | 74.0% | 10,627 | 22.1 (21.8 - 22.5) | - | . |
| Yes | 16,886 | 26.0% | 5,266 | 31.2 (30.5 - 31.9) | 0.99 (0.93 - 1.07) | 0.874 |
| **Deficiency Anemia** |  |  |  |  |  |  |
| No (Reference) | 61,975 | 95.5% | 15,060 | 24.3 (24.0 - 24.6) | - | . |
| Yes | 2,939 | 4.5% | 833 | 28.3 (26.7 - 30.0) | 1.03 (0.92 - 1.14) | 0.612 |
| **Depression** |  |  |  |  |  |  |
| No (Reference) | 55,587 | 85.6% | 13,705 | 24.7 (24.3 - 25.0) | - | . |
| Yes | 9,327 | 14.4% | 2,188 | 23.5 (22.6 - 24.3) | 1.05 (0.98 - 1.12) | 0.165 |
| **Diabetes** |  |  |  |  |  |  |
| No (Reference) | 38,036 | 58.6% | 8,796 | 23.1 (22.7 - 23.6) | - | . |
| Yes | 26,878 | 41.4% | 7,097 | 26.4 (25.9 - 26.9) | 0.94 (0.88 - 1.00) | 0.064 |
| **Fluid and Electrolyte Disorder** |  |  |  |  |  |  |
| No (Reference) | 25,445 | 39.2% | 3,387 | 13.3 (12.9 - 13.7) | - | . |
| Yes | 39,469 | 60.8% | 12,506 | 31.7 (31.2 - 32.1) | 1.59 (1.51 - 1.68) | <.0001 |
| **Hypertension (uncomplicated)** |  |  |  |  |  |  |
| No (Reference) | 35,189 | 54.2% | 9,572 | 27.2 (26.7 - 27.7) | - | . |
| Yes | 29,725 | 45.8% | 6,321 | 21.3 (20.8 - 21.7) | 0.99 (0.93 - 1.05) | 0.681 |
| **Hypertension (complicated)** |  |  |  |  |  |  |
| No (Reference) | 39,094 | 60.2% | 8,123 | 20.8 (20.4 - 21.2) | - | . |
| Yes | 25,820 | 39.8% | 7,770 | 30.1 (29.5 - 30.7) | 1.11 (1.03 - 1.20) | 0.006 |
| **Liver Disease** |  |  |  |  |  |  |
| No (Reference) | 62,066 | 95.6% | 14,742 | 23.8 (23.4 - 24.1) | - | . |
| Yes | 2,848 | 4.4% | 1,151 | 40.4 (38.6 - 42.2) | 1.47 (1.32 - 1.64) | <.0001 |
| **Metastatic Cancer** |  |  |  |  |  |  |
| No (Reference) | 63,978 | 98.6% | 15,617 | 24.4 (24.1 - 24.7) | - | . |
| Yes | 936 | 1.4% | 276 | 29.5 (26.6 - 32.5) | 1.15 (0.93 - 1.43) | 0.208 |
| **Obesity** |  |  |  |  |  |  |
| No (Reference) | 48,421 | 74.6% | 11,625 | 24.0 (23.6 - 24.4) | - | . |
| Yes | 16,493 | 25.4% | 4,268 | 25.9 (25.2 - 26.6) | 1.07 (1.01 - 1.14) | 0.017 |
| **Other Neurological Disorders** |  |  |  |  |  |  |
| No (Reference) | 51,393 | 79.2% | 10,743 | 20.9 (20.6 - 21.3) | - | . |
| Yes | 13,521 | 20.8% | 5,150 | 38.1 (37.3 - 38.9) | 1.36 (1.28 - 1.44) | <.0001 |
| **Paralysis** |  |  |  |  |  |  |
| No (Reference) | 64,170 | 98.9% | 15,576 | 24.3 (23.9 - 24.6) | - | . |
| Yes | 744 | 1.1% | 317 | 42.6 (39.0 - 46.3) | 1.05 (0.86 - 1.29) | 0.640 |
| **Peripheral Vascular Disorder** |  |  |  |  |  |  |
| No (Reference) | 59,578 | 91.8% | 14,388 | 24.1 (23.8 - 24.5) | - | . |
| Yes | 5,336 | 8.2% | 1,505 | 28.2 (27.0 - 29.4) | 1.00 (0.92 - 1.09) | 0.951 |
| **Pulmonary Circulation Disorder** |  |  |  |  |  |  |
| No (Reference) | 62,552 | 96.4% | 15,108 | 24.2 (23.8 - 24.5) | - | . |
| Yes | 2,362 | 3.6% | 785 | 33.2 (31.3 - 35.2) | 1.05 (0.92 - 1.19) | 0.494 |
| **Renal Failure** |  |  |  |  |  |  |
| No (Reference) | 54,265 | 83.6% | 12,470 | 23.0 (22.6 - 23.3) | - | . |
| Yes | 10,649 | 16.4% | 3,423 | 32.1 (31.3 - 33.0) | 1.06 (0.97 - 1.15) | 0.201 |
| **Rheumatoid Arthritis/Collagen Vascular Diseases (CVD)** |  |  |  |  |  |  |
| No (Reference) | 61,741 | 95.1% | 15,094 | 24.4 (24.1 - 24.8) | - | . |
| Yes | 3,173 | 4.9% | 799 | 25.2 (23.7 - 26.7) | 1.16 (1.04 - 1.29) | 0.010 |
| **Solid Tumor** |  |  |  |  |  |  |
| No (Reference) | 62,364 | 96.1% | 15,186 | 24.4 (24.0 - 24.7) | - | . |
| Yes | 2,550 | 3.9% | 707 | 27.7 (26.0 - 29.5) | 1.08 (0.94 - 1.23) | 0.281 |
| **Valvular Disease** |  |  |  |  |  |  |
| No (Reference) | 59,658 | 91.9% | 14,403 | 24.1 (23.8 - 24.5) | - | . |
| Yes | 5,256 | 8.1% | 1,490 | 28.3 (27.1 - 29.6) | 0.95 (0.87 - 1.03) | 0.209 |
| **Weight Loss** |  |  |  |  |  |  |
| No (Reference) | 58,309 | 89.8% | 13,348 | 22.9 (22.6 - 23.2) | - | . |
| Yes | 6,605 | 10.2% | 2,545 | 38.5 (37.4 - 39.7) | 1.17 (1.09 - 1.26) | <.0001 |
| ^a^ Patients with missing values for ADI, Urban/Rural, Hospital Size and Ownership were dropped from the multivariate analysis. | | | | | | |
| ^b^ The South US census region includes counts for "Other" territories to avoid identification due to low counts. | | | | | | |
| ^c^ Patients have not had COVID-19 diagnosis recorded in the 30 days prior to admission. | | | | | | |
| **Notes:** Model evaluation metrics were ROC Area Under the Curve (0.8571); Pearson Chi Sq Goodness of Fit test p value (1); Deviance Goodness of Fit test p value (1). | | | | | |  |
| Intravenous Immunoglobulin/Hyperimmune Globulin and Peptic Ulcer Disease Groups have been omitted to avoid identification due to low counts. | | | | | |  |

**Supplementary Figure 1:** Monthly Inpatient (IP) Mortality Rates among Inpatient COVID-19 Convalescent Plasma (CCP) Recipients Residing in the Midwest, Overall and by IC Status

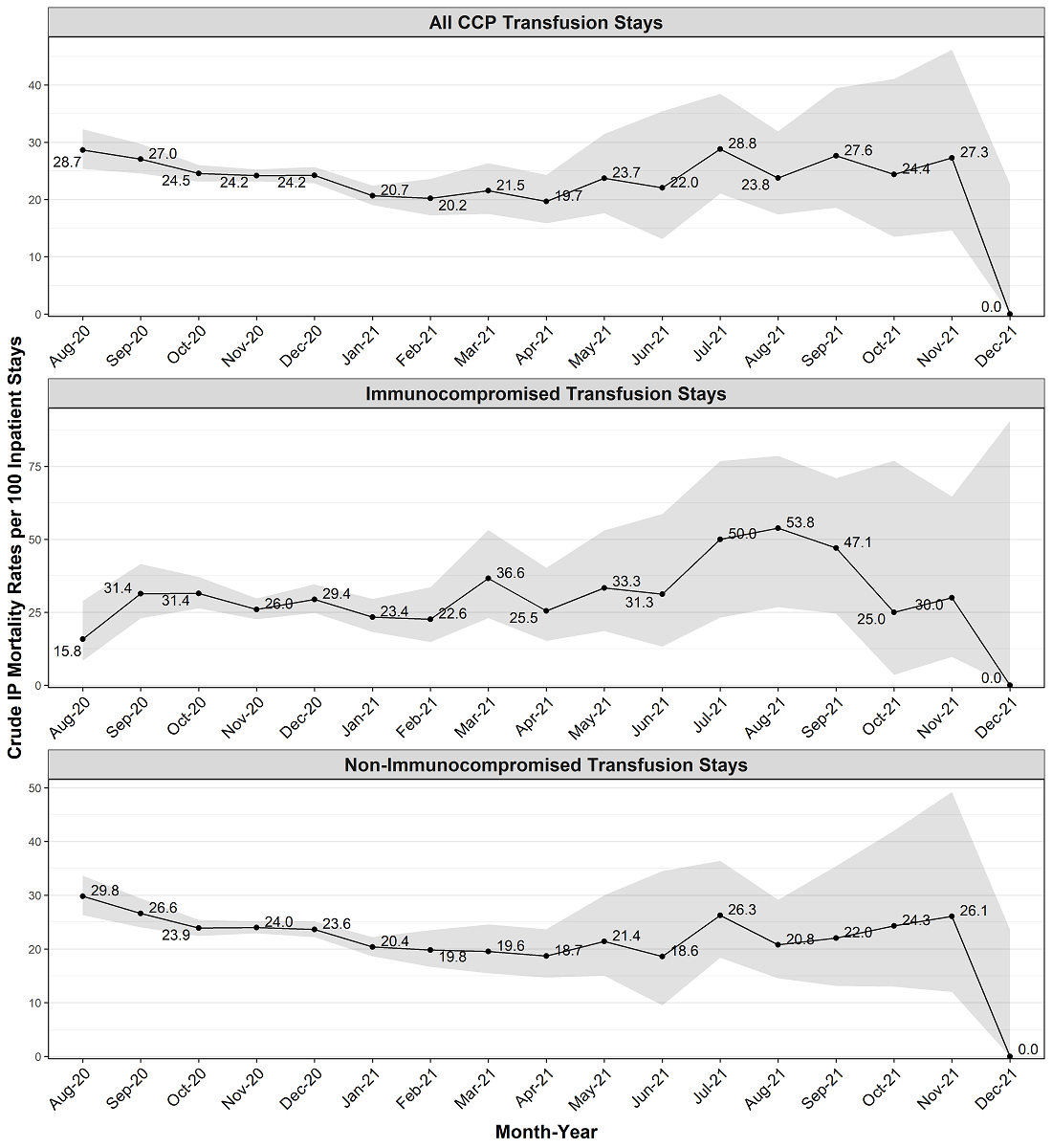

**Supplementary Figure 2:** Monthly Inpatient (IP) Mortality Rates among Inpatient COVID-19 Convalescent Plasma (CCP) Recipients Residing in the Northeast, Overall and by IC Status

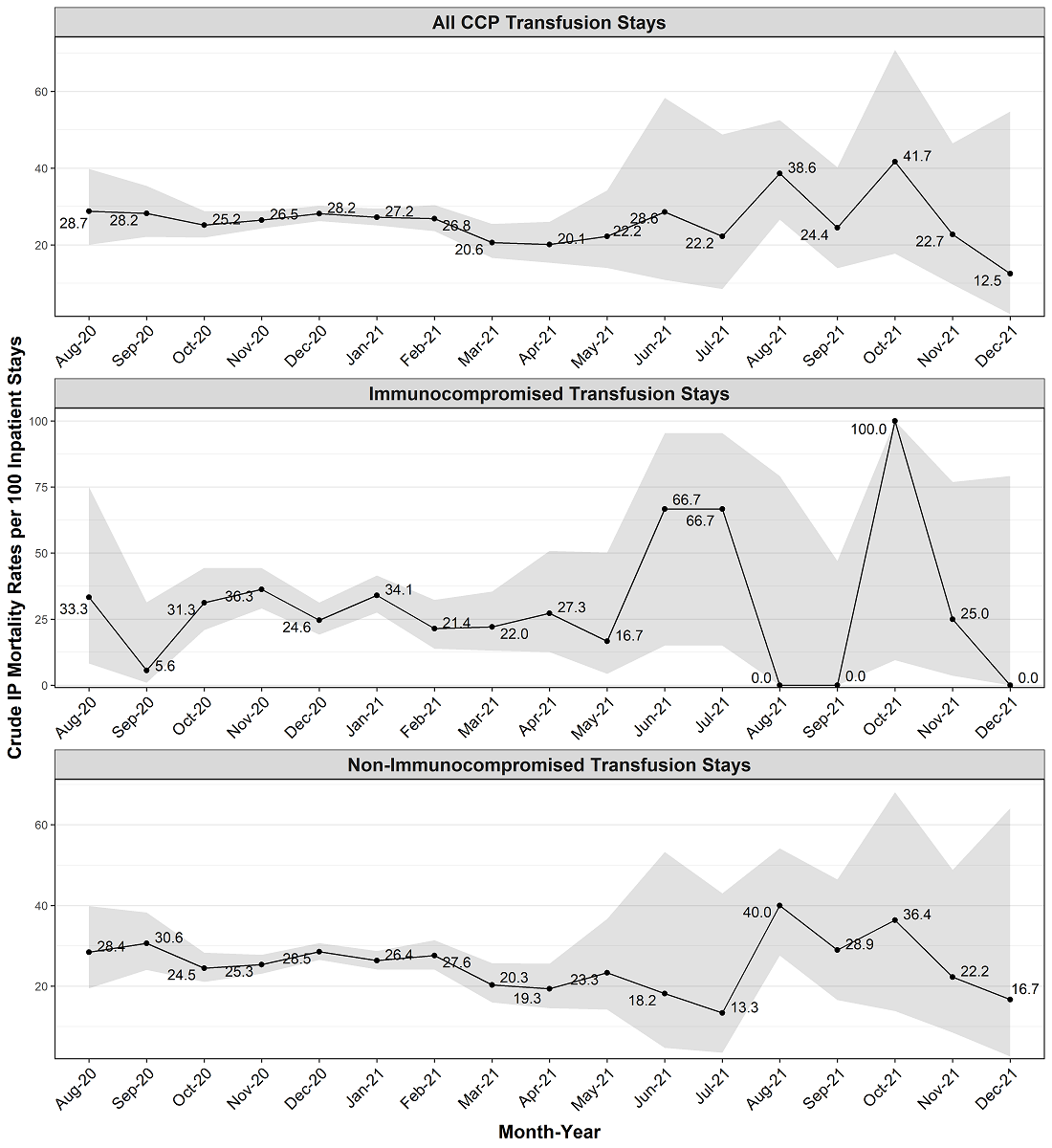

**Supplementary Figure 3:** Monthly Inpatient (IP) Mortality Rates among Inpatient COVID-19 Convalescent Plasma (CCP) Recipients Residing in the South, Overall and by IC Status

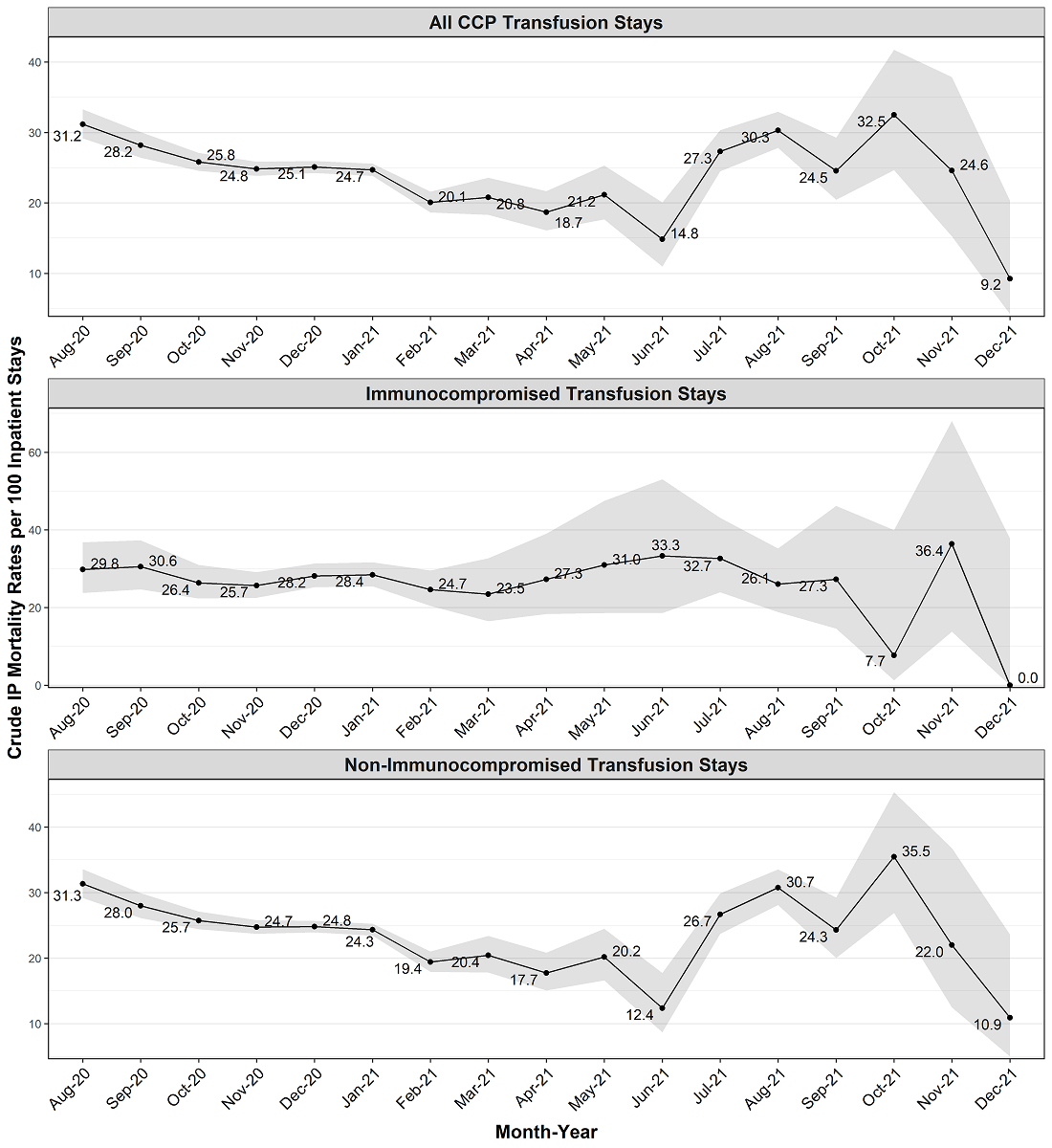

**Supplementary Figure 4:** Monthly Inpatient (IP) Mortality Rates among Inpatient COVID-19 Convalescent Plasma (CCP) Recipients Residing in the West, Overall and by IC Status

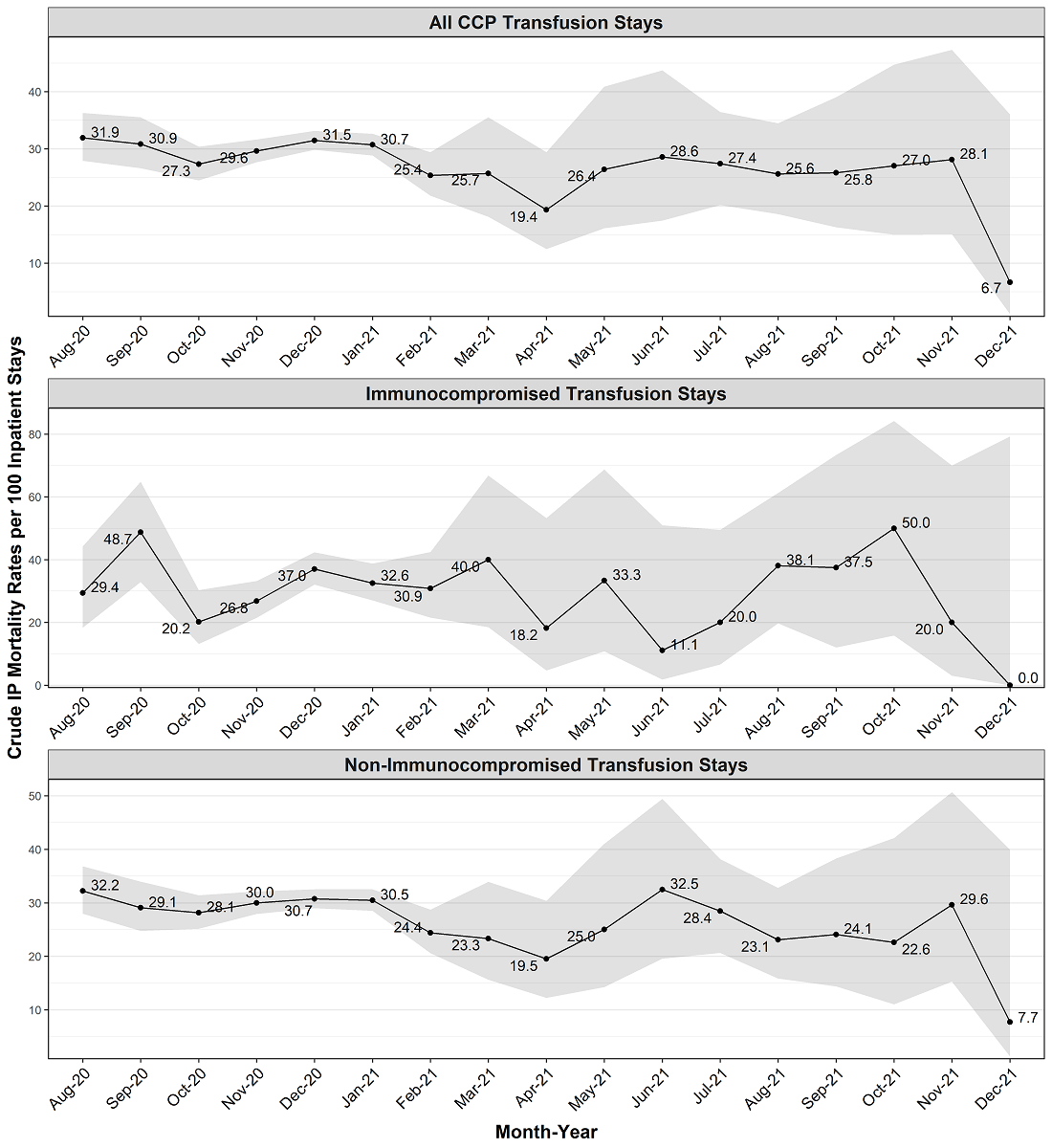

**Supplementary Figure 5:** Adjusted Odds of Inpatient (IP) Mortality by Age, While Controlling for Covariates in the Final Overall Regression Model

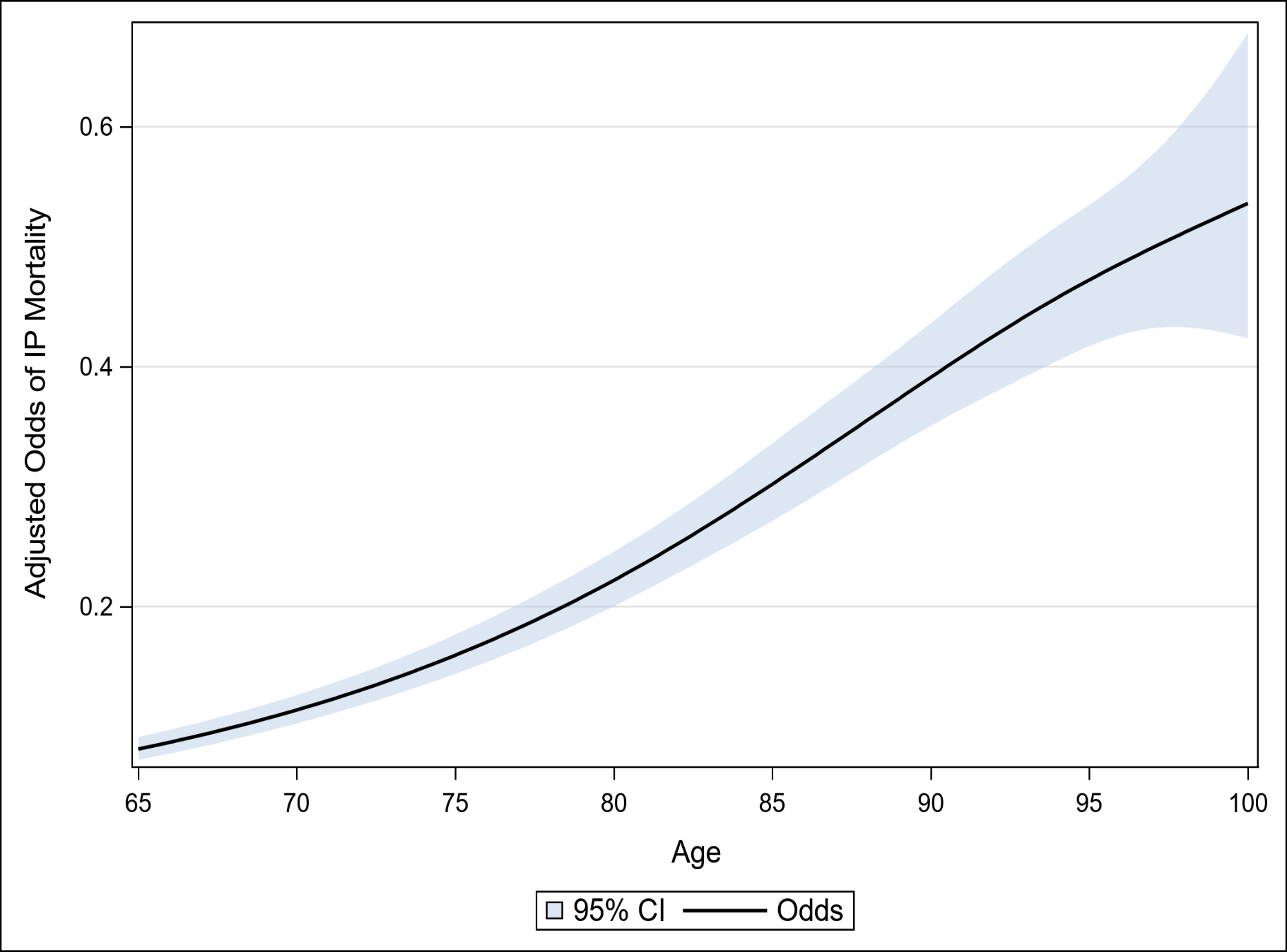

**Supplementary Figure 6:** Adjusted Odds of Inpatient (IP) Mortality by Area Deprivation Index, While Controlling for Covariates in the Final Overall Regression Model

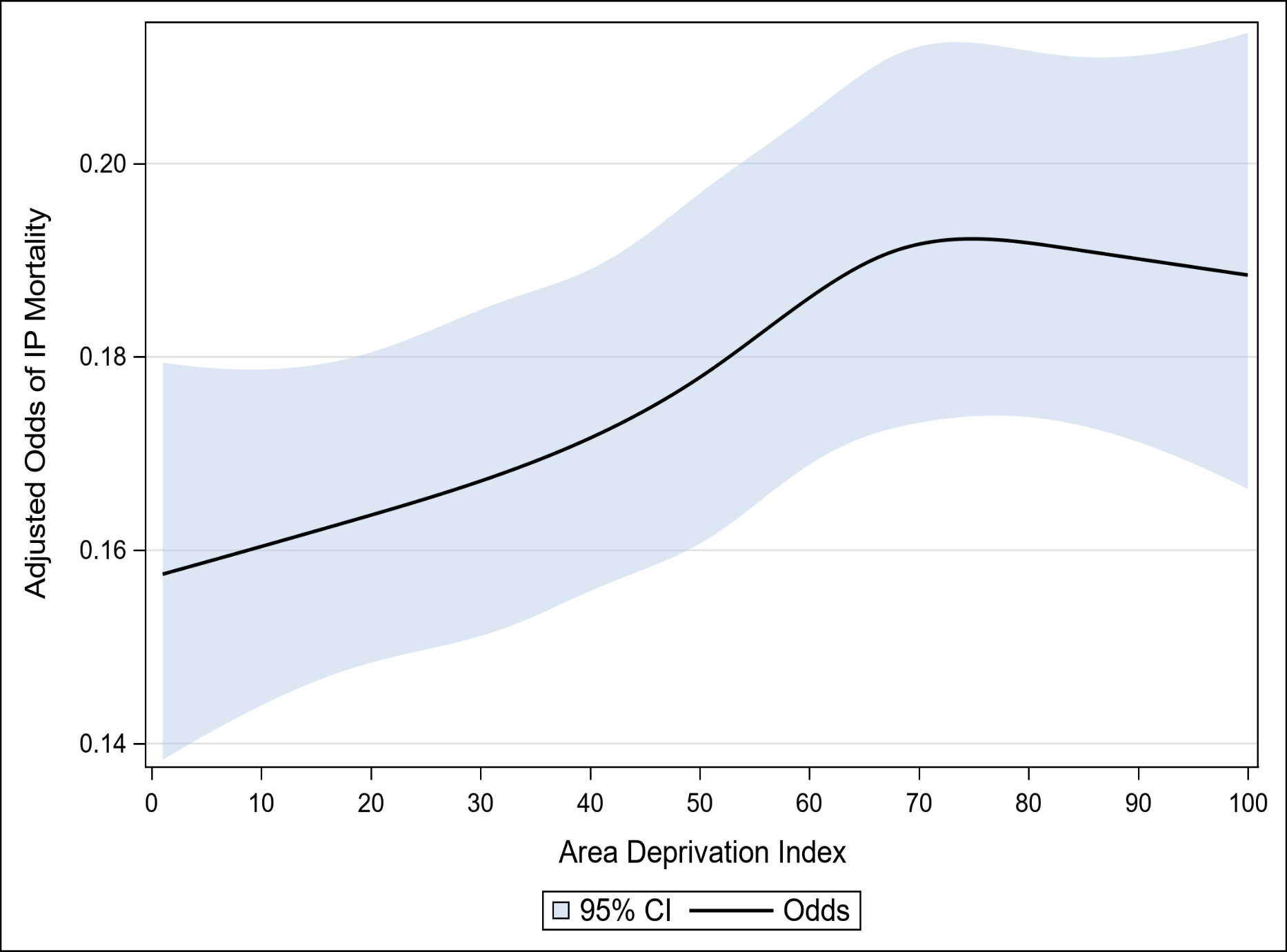

**Supplementary Figure 7:** Adjusted Odds of Inpatient (IP) Mortality by Charlson Comorbidity Score, While Controlling for Covariates in the Final Overall Regression Model

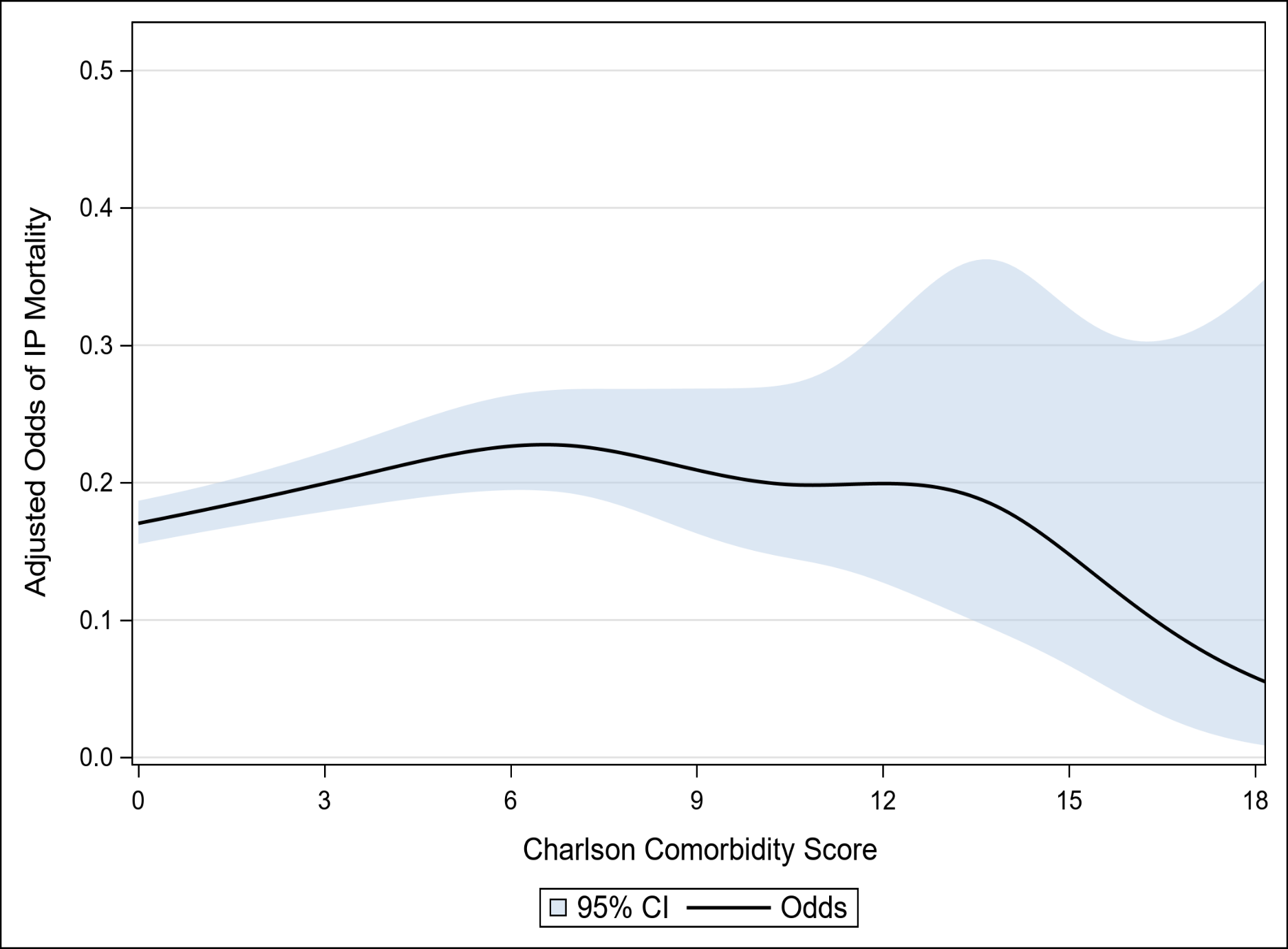

**Supplementary Figure 8:** Adjusted Odds of Inpatient (IP) Mortality by Time Since Most Recent COVID-19 Diagnosis Recorded in the 30 Day Prior to Hospital Admission, While Controlling for Covariates in the Final Overall Regression Model

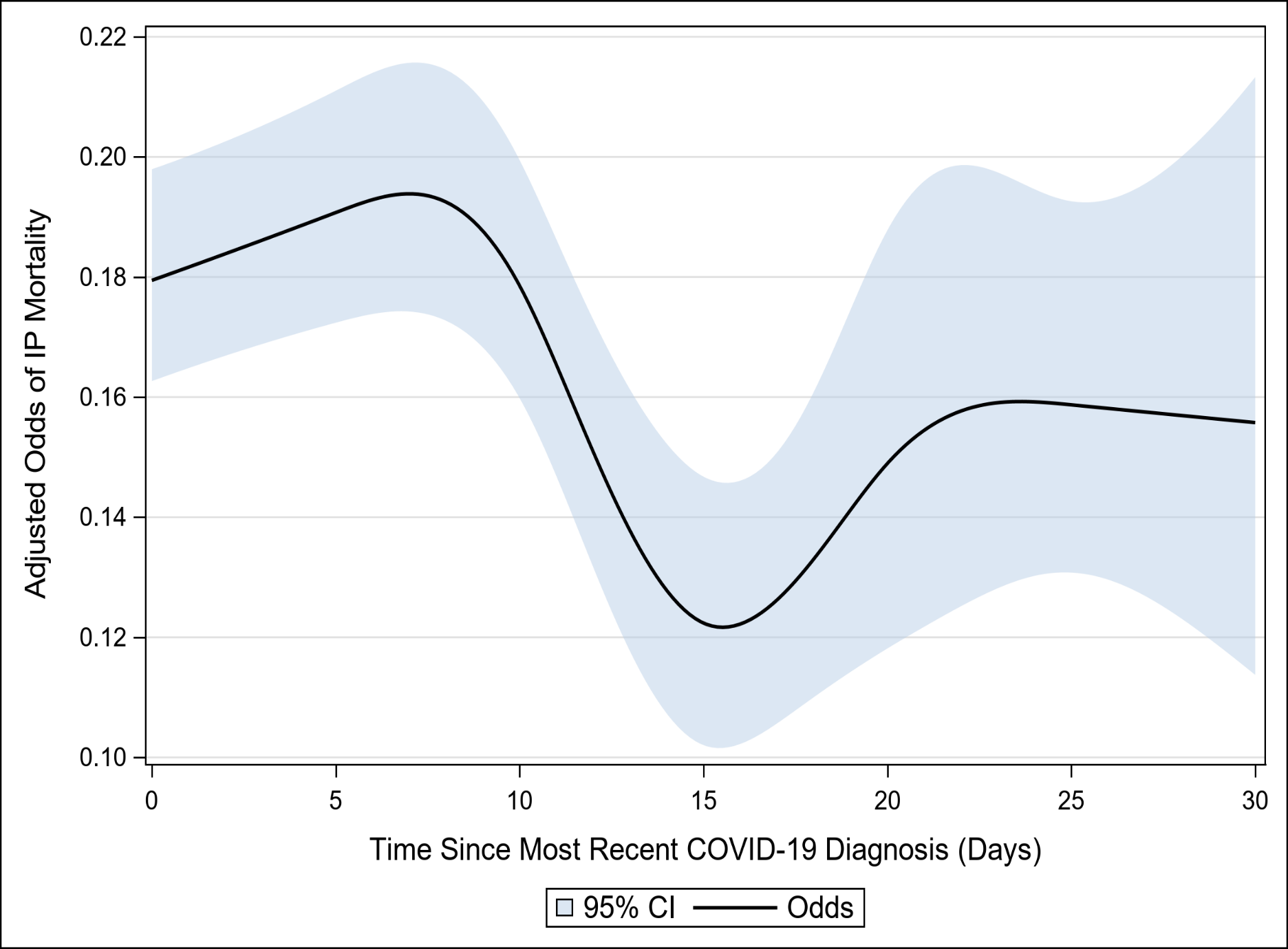

**Supplementary Figure 9:** Adjusted Odds of Inpatient (IP) Mortality by Hospital Admission Month, While Controlling for Covariates in the Final Overall Regression Model

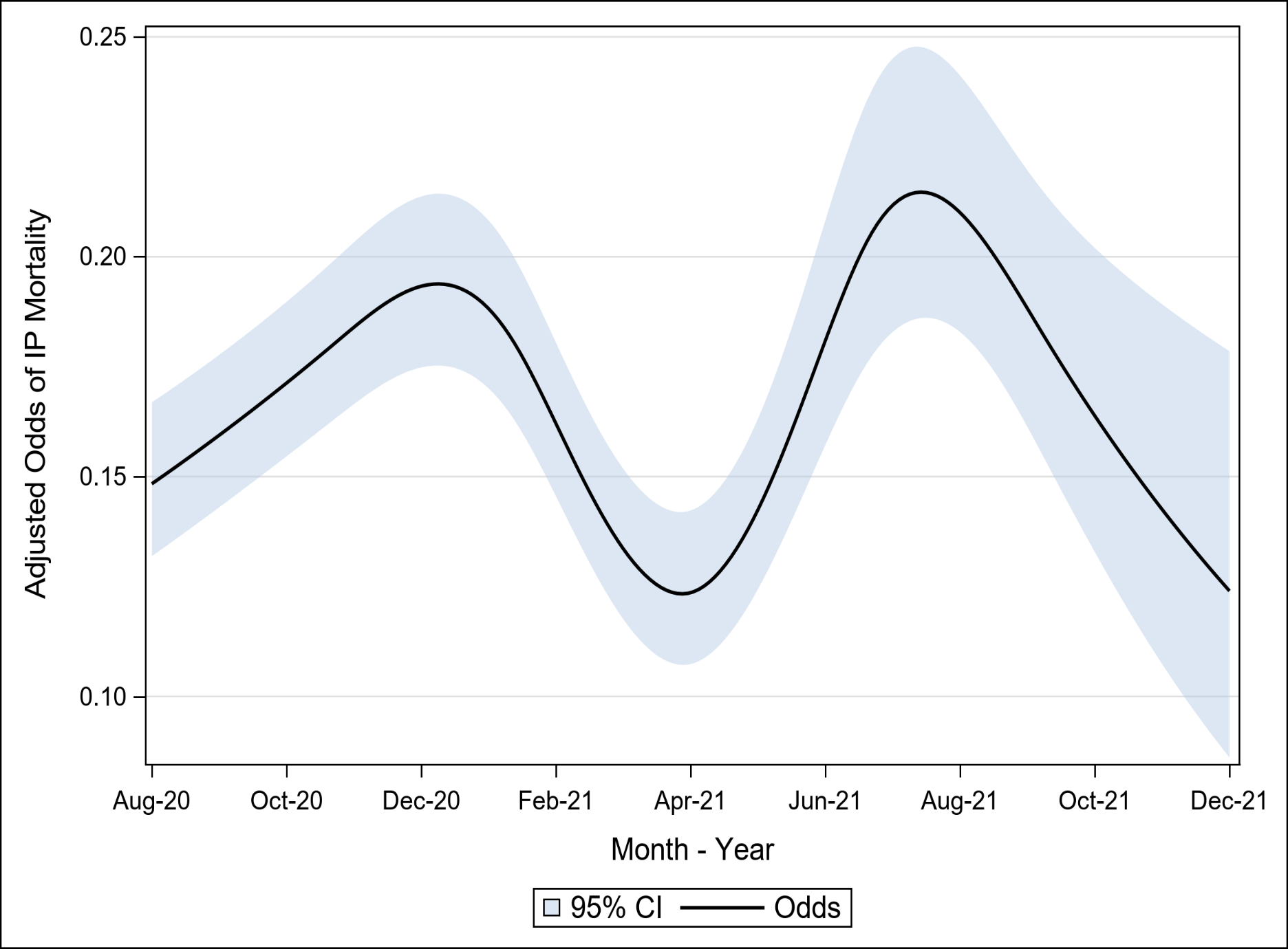

| **Supplementary Appendix Table. Immunocompromised Status Code List** | | |  |  |
| --- | --- | --- | --- | --- |
| **Code Type^a^** | **Code** | **Full Description** | **Code Group** | **Condition/Treatment Type^b,c^** |
| DGN10 | C882 | Heavy chain disease | Diagnosis | MDS/MPN/IPD |
| DGN10 | C883 | Immunoproliferative small intestinal disease | Diagnosis | MDS/MPN/IPD |
| DGN10 | C888 | Other malignant immunoproliferative diseases | Diagnosis | MDS/MPN/IPD |
| DGN10 | C889 | Malignant immunoproliferative disease, unspecified | Diagnosis | MDS/MPN/IPD |
| DGN10 | C9440 | Acute panmyelosis with myelofibrosis not having achieved remission | Diagnosis | MDS/MPN/IPD |
| DGN10 | C9441 | Acute panmyelosis with myelofibrosis, in remission | Diagnosis | MDS/MPN/IPD |
| DGN10 | C9442 | Acute panmyelosis with myelofibrosis, in relapse | Diagnosis | MDS/MPN/IPD |
| DGN10 | C946 | Myelodysplastic disease, not classified | Diagnosis | MDS/MPN/IPD |
| DGN10 | D45 | Polycythemia vera | Diagnosis | MDS/MPN/IPD |
| DGN10 | D460 | Refractory anemia without ring sideroblasts, so stated | Diagnosis | MDS/MPN/IPD |
| DGN10 | D461 | Refractory anemia with ring sideroblasts | Diagnosis | MDS/MPN/IPD |
| DGN10 | D4620 | Refractory anemia with excess of blasts, unspecified | Diagnosis | MDS/MPN/IPD |
| DGN10 | D4621 | Refractory anemia with excess of blasts 1 | Diagnosis | MDS/MPN/IPD |
| DGN10 | D4622 | Refractory anemia with excess of blasts 2 | Diagnosis | MDS/MPN/IPD |
| DGN10 | D464 | Refractory anemia, unspecified | Diagnosis | MDS/MPN/IPD |
| DGN10 | D469 | Myelodysplastic syndrome, unspecified | Diagnosis | MDS/MPN/IPD |
| DGN10 | D46A | Refractory cytopenia with multilineage dysplasia | Diagnosis | MDS/MPN/IPD |
| DGN10 | D46B | Refractory cytopenia with multilineage dysplasia and ring sideroblasts | Diagnosis | MDS/MPN/IPD |
| DGN10 | D46C | Myelodysplastic syndrome with isolated del(5q) chromosomal abnormality | Diagnosis | MDS/MPN/IPD |
| DGN10 | D46Z | Other myelodysplastic syndromes | Diagnosis | MDS/MPN/IPD |
| DGN10 | D4701 | Cutaneous mastocytosis | Diagnosis | MDS/MPN/IPD |
| DGN10 | D4702 | Systemic mastocytosis | Diagnosis | MDS/MPN/IPD |
| DGN10 | D4709 | Other mast cell neoplasms of uncertain behavior | Diagnosis | MDS/MPN/IPD |
| DGN10 | D471 | Chronic myeloproliferative disease | Diagnosis | MDS/MPN/IPD |
| DGN10 | D47Z2 | Castleman disease | Diagnosis | MDS/MPN/IPD |
| DGN10 | D7581 | Myelofibrosis | Diagnosis | MDS/MPN/IPD |
| DGN10 | D479 | Neoplasm of uncertain behavior of lymphoid, hematopoietic and related tissue, unspecified | Diagnosis | MDS/MPN/IPD |
| DGN10 | D47Z1 | Post-transplant lymphoproliferative disorder (PTLD) | Diagnosis | MDS/MPN/IPD |
| DGN10 | D47Z9 | Other specified neoplasms of uncertain behavior of lymphoid, hematopoietic and related tissue | Diagnosis | MDS/MPN/IPD |
| DGN10 | C802 | Malignant neoplasm associated with transplanted organ | Diagnosis | MDS/MPN/IPD |
| DGN10 | C960 | Multifocal and multisystemic (disseminated) Langerhans-cell histiocytosis | Diagnosis | MDS/MPN/IPD |
| DGN10 | C962 | Malignant mast cell tumor | Diagnosis | MDS/MPN/IPD |
| DGN10 | C965 | Multifocal and unisystemic Langerhans-cell histiocytosis | Diagnosis | MDS/MPN/IPD |
| DGN10 | C966 | Unifocal Langerhans-cell histiocytosis | Diagnosis | MDS/MPN/IPD |
| DGN10 | C96A | Histiocytic sarcoma | Diagnosis | MDS/MPN/IPD |
| DGN10 | D474 | Osteomyelofibrosis | Diagnosis | MDS/MPN/IPD |
| DGN10 | D6182 | Myelophthisis | Diagnosis | MDS/MPN/IPD |
| DGN10 | D8982 | Autoimmune lymphoproliferative syndrome [ALPS] | Diagnosis | MDS/MPN/IPD |
| DGN10 | D761 | Hemophagocytic lymphohistiocytosis | Diagnosis | MDS/MPN/IPD |
| DGN10 | D762 | Hemophagocytic syndrome, infection-associated | Diagnosis | MDS/MPN/IPD |
| DGN10 | D763 | Other histiocytosis syndromes | Diagnosis | MDS/MPN/IPD |
| DGN10 | C800 | Disseminated malignant neoplasm, unspecified | Diagnosis | MDS/MPN/IPD |
| DGN10 | D473 | Essential (hemorrhagic) thrombocythemia | Diagnosis | MDS/MPN/IPD |
| DGN10 | C9620 | Malignant mast cell neoplasm, unspecified | Diagnosis | MDS/MPN/IPD |
| DGN10 | C9621 | Aggressive systemic mastocytosis | Diagnosis | MDS/MPN/IPD |
| DGN10 | C9622 | Mast cell sarcoma | Diagnosis | MDS/MPN/IPD |
| DGN10 | C9629 | Other malignant mast cell neoplasm | Diagnosis | MDS/MPN/IPD |
| DGN10 | D470 | Mast cell neoplasms of uncertain behavior | Diagnosis | MDS/MPN/IPD |
| DGN10 | D6109 | Other constitutional aplastic anemia | Diagnosis | MDS/MPN/IPD |
| DGN10 | D640 | D64.0 | Diagnosis | MDS/MPN/IPD |
| DGN10 | D641 | Secondary sideroblastic anemia due to disease | Diagnosis | MDS/MPN/IPD |
| DGN10 | D642 | Secondary sideroblastic anemia due to drugs and toxins | Diagnosis | MDS/MPN/IPD |
| DGN10 | D643 | Other sideroblastic anemias | Diagnosis | MDS/MPN/IPD |
| DGN10 | D8941 | Monoclonal mast cell activation syndrome | Diagnosis | MDS/MPN/IPD |
| HCPCS | 3490F | History of AIDS-defining condition (HIV) | Diagnosis | HIV |
| DGN10 | B20 | Human immunodeficiency virus [HIV] disease | Diagnosis | HIV |
| DGN10 | B9735 | Human immunodeficiency virus, type 2 [HIV 2] as the cause of diseases classified elsewhere | Diagnosis | HIV |
| DGN10 | Z21 | Asymptomatic human immunodeficiency virus [HIV] infection status | Diagnosis | HIV |
| DGN10 | D893 | Immune reconstitution syndrome | Diagnosis | HIV |
| DGN10 | C8100 | Nodular lymphocyte predominant Hodgkin lymphoma, unspecified site | Diagnosis | Hodgkins Lymphoma |
| DGN10 | C8101 | Nodular lymphocyte predominant Hodgkin lymphoma, lymph nodes of head, face, and neck | Diagnosis | Hodgkins Lymphoma |
| DGN10 | C8102 | Nodular lymphocyte predominant Hodgkin lymphoma, intrathoracic lymph nodes | Diagnosis | Hodgkins Lymphoma |
| DGN10 | C8103 | Nodular lymphocyte predominant Hodgkin lymphoma, intra-abdominal lymph nodes | Diagnosis | Hodgkins Lymphoma |
| DGN10 | C8104 | Nodular lymphocyte predominant Hodgkin lymphoma, lymph nodes of axilla and upper limb | Diagnosis | Hodgkins Lymphoma |
| DGN10 | C8105 | Nodular lymphocyte predominant Hodgkin lymphoma, lymph nodes of inguinal region and lower limb | Diagnosis | Hodgkins Lymphoma |
| DGN10 | C8106 | Nodular lymphocyte predominant Hodgkin lymphoma, intrapelvic lymph nodes | Diagnosis | Hodgkins Lymphoma |
| DGN10 | C8107 | Nodular lymphocyte predominant Hodgkin lymphoma, spleen | Diagnosis | Hodgkins Lymphoma |
| DGN10 | C8108 | Nodular lymphocyte predominant Hodgkin lymphoma, lymph nodes of multiple sites | Diagnosis | Hodgkins Lymphoma |
| DGN10 | C8109 | Nodular lymphocyte predominant Hodgkin lymphoma, extranodal and solid organ sites | Diagnosis | Hodgkins Lymphoma |
| DGN10 | C8110 | Nodular sclerosis Hodgkin lymphoma, unspecified site | Diagnosis | Hodgkins Lymphoma |
| DGN10 | C8111 | Nodular sclerosis Hodgkin lymphoma, lymph nodes of head, face, and neck | Diagnosis | Hodgkins Lymphoma |
| DGN10 | C8112 | Nodular sclerosis Hodgkin lymphoma, intrathoracic lymph nodes | Diagnosis | Hodgkins Lymphoma |
| DGN10 | C8113 | Nodular sclerosis Hodgkin lymphoma, intra-abdominal lymph nodes | Diagnosis | Hodgkins Lymphoma |
| DGN10 | C8114 | Nodular sclerosis Hodgkin lymphoma, lymph nodes of axilla and upper limb | Diagnosis | Hodgkins Lymphoma |
| DGN10 | C8115 | Nodular sclerosis Hodgkin lymphoma, lymph nodes of inguinal region and lower limb | Diagnosis | Hodgkins Lymphoma |
| DGN10 | C8116 | Nodular sclerosis Hodgkin lymphoma, intrapelvic lymph nodes | Diagnosis | Hodgkins Lymphoma |
| DGN10 | C8117 | Nodular sclerosis Hodgkin lymphoma, spleen | Diagnosis | Hodgkins Lymphoma |
| DGN10 | C8118 | Nodular sclerosis Hodgkin lymphoma, lymph nodes of multiple sites | Diagnosis | Hodgkins Lymphoma |
| DGN10 | C8119 | Nodular sclerosis Hodgkin lymphoma, extranodal and solid organ sites | Diagnosis | Hodgkins Lymphoma |
| DGN10 | C8120 | Mixed cellularity Hodgkin lymphoma, unspecified site | Diagnosis | Hodgkins Lymphoma |
| DGN10 | C8121 | Mixed cellularity Hodgkin lymphoma, lymph nodes of head, face, and neck | Diagnosis | Hodgkins Lymphoma |
| DGN10 | C8122 | Mixed cellularity Hodgkin lymphoma, intrathoracic lymph nodes | Diagnosis | Hodgkins Lymphoma |
| DGN10 | C8123 | Mixed cellularity Hodgkin lymphoma, intra-abdominal lymph nodes | Diagnosis | Hodgkins Lymphoma |
| DGN10 | C8124 | Mixed cellularity Hodgkin lymphoma, lymph nodes of axilla and upper limb | Diagnosis | Hodgkins Lymphoma |
| DGN10 | C8125 | Mixed cellularity Hodgkin lymphoma, lymph nodes of inguinal region and lower limb | Diagnosis | Hodgkins Lymphoma |
| DGN10 | C8126 | Mixed cellularity Hodgkin lymphoma, intrapelvic lymph nodes | Diagnosis | Hodgkins Lymphoma |
| DGN10 | C8127 | Mixed cellularity Hodgkin lymphoma, spleen | Diagnosis | Hodgkins Lymphoma |
| DGN10 | C8128 | Mixed cellularity Hodgkin lymphoma, lymph nodes of multiple sites | Diagnosis | Hodgkins Lymphoma |
| DGN10 | C8129 | Mixed cellularity Hodgkin lymphoma, extranodal and solid organ sites | Diagnosis | Hodgkins Lymphoma |
| DGN10 | C8130 | Lymphocyte depleted Hodgkin lymphoma, unspecified site | Diagnosis | Hodgkins Lymphoma |
| DGN10 | C8131 | Lymphocyte depleted Hodgkin lymphoma, lymph nodes of head, face, and neck | Diagnosis | Hodgkins Lymphoma |
| DGN10 | C8132 | Lymphocyte depleted Hodgkin lymphoma, intrathoracic lymph nodes | Diagnosis | Hodgkins Lymphoma |
| DGN10 | C8133 | Lymphocyte depleted Hodgkin lymphoma, intra-abdominal lymph nodes | Diagnosis | Hodgkins Lymphoma |
| DGN10 | C8134 | Lymphocyte depleted Hodgkin lymphoma, lymph nodes of axilla and upper limb | Diagnosis | Hodgkins Lymphoma |
| DGN10 | C8135 | Lymphocyte depleted Hodgkin lymphoma, lymph nodes of inguinal region and lower limb | Diagnosis | Hodgkins Lymphoma |
| DGN10 | C8136 | Lymphocyte depleted Hodgkin lymphoma, intrapelvic lymph nodes | Diagnosis | Hodgkins Lymphoma |
| DGN10 | C8137 | Lymphocyte depleted Hodgkin lymphoma, spleen | Diagnosis | Hodgkins Lymphoma |
| DGN10 | C8138 | Lymphocyte depleted Hodgkin lymphoma, lymph nodes of multiple sites | Diagnosis | Hodgkins Lymphoma |
| DGN10 | C8139 | Lymphocyte depleted Hodgkin lymphoma, extranodal and solid organ sites | Diagnosis | Hodgkins Lymphoma |
| DGN10 | C8140 | Lymphocyte-rich Hodgkin lymphoma, unspecified site | Diagnosis | Hodgkins Lymphoma |
| DGN10 | C8141 | Lymphocyte-rich Hodgkin lymphoma, lymph nodes of head, face, and neck | Diagnosis | Hodgkins Lymphoma |
| DGN10 | C8142 | Lymphocyte-rich Hodgkin lymphoma, intrathoracic lymph nodes | Diagnosis | Hodgkins Lymphoma |
| DGN10 | C8143 | Lymphocyte-rich Hodgkin lymphoma, intra-abdominal lymph nodes | Diagnosis | Hodgkins Lymphoma |
| DGN10 | C8144 | Lymphocyte-rich Hodgkin lymphoma, lymph nodes of axilla and upper limb | Diagnosis | Hodgkins Lymphoma |
| DGN10 | C8145 | Lymphocyte-rich Hodgkin lymphoma, lymph nodes of inguinal region and lower limb | Diagnosis | Hodgkins Lymphoma |
| DGN10 | C8146 | Lymphocyte-rich Hodgkin lymphoma, intrapelvic lymph nodes | Diagnosis | Hodgkins Lymphoma |
| DGN10 | C8147 | Lymphocyte-rich Hodgkin lymphoma, spleen | Diagnosis | Hodgkins Lymphoma |
| DGN10 | C8148 | Lymphocyte-rich Hodgkin lymphoma, lymph nodes of multiple sites | Diagnosis | Hodgkins Lymphoma |
| DGN10 | C8149 | Lymphocyte-rich Hodgkin lymphoma, extranodal and solid organ sites | Diagnosis | Hodgkins Lymphoma |
| DGN10 | C8170 | Other Hodgkin lymphoma, unspecified site | Diagnosis | Hodgkins Lymphoma |
| DGN10 | C8171 | Other Hodgkin lymphoma, lymph nodes of head, face, and neck | Diagnosis | Hodgkins Lymphoma |
| DGN10 | C8172 | Other Hodgkin lymphoma, intrathoracic lymph nodes | Diagnosis | Hodgkins Lymphoma |
| DGN10 | C8173 | Other Hodgkin lymphoma, intra-abdominal lymph nodes | Diagnosis | Hodgkins Lymphoma |
| DGN10 | C8174 | Other Hodgkin lymphoma, lymph nodes of axilla and upper limb | Diagnosis | Hodgkins Lymphoma |
| DGN10 | C8175 | Other Hodgkin lymphoma, lymph nodes of inguinal region and lower limb | Diagnosis | Hodgkins Lymphoma |
| DGN10 | C8176 | Other Hodgkin lymphoma, intrapelvic lymph nodes | Diagnosis | Hodgkins Lymphoma |
| DGN10 | C8177 | Other Hodgkin lymphoma, spleen | Diagnosis | Hodgkins Lymphoma |
| DGN10 | C8178 | Other Hodgkin lymphoma, lymph nodes of multiple sites | Diagnosis | Hodgkins Lymphoma |
| DGN10 | C8179 | Other Hodgkin lymphoma, extranodal and solid organ sites | Diagnosis | Hodgkins Lymphoma |
| DGN10 | C8190 | Hodgkin lymphoma, unspecified, unspecified site | Diagnosis | Hodgkins Lymphoma |
| DGN10 | C8191 | Hodgkin lymphoma, unspecified, lymph nodes of head, face, and neck | Diagnosis | Hodgkins Lymphoma |
| DGN10 | C8192 | Hodgkin lymphoma, unspecified, intrathoracic lymph nodes | Diagnosis | Hodgkins Lymphoma |
| DGN10 | C8193 | Hodgkin lymphoma, unspecified, intra-abdominal lymph nodes | Diagnosis | Hodgkins Lymphoma |
| DGN10 | C8194 | Hodgkin lymphoma, unspecified, lymph nodes of axilla and upper limb | Diagnosis | Hodgkins Lymphoma |
| DGN10 | C8195 | Hodgkin lymphoma, unspecified, lymph nodes of inguinal region and lower limb | Diagnosis | Hodgkins Lymphoma |
| DGN10 | C8196 | Hodgkin lymphoma, unspecified, intrapelvic lymph nodes | Diagnosis | Hodgkins Lymphoma |
| DGN10 | C8197 | Hodgkin lymphoma, unspecified, spleen | Diagnosis | Hodgkins Lymphoma |
| DGN10 | C8198 | Hodgkin lymphoma, unspecified, lymph nodes of multiple sites | Diagnosis | Hodgkins Lymphoma |
| DGN10 | C8199 | Hodgkin lymphoma, unspecified, extranodal and solid organ sites | Diagnosis | Hodgkins Lymphoma |
| DGN10 | C8200 | Follicular lymphoma grade I, unspecified site | Diagnosis | Non-Hodgkins Lymphoma |
| DGN10 | C8201 | Follicular lymphoma grade I, lymph nodes of head, face, and neck | Diagnosis | Non-Hodgkins Lymphoma |
| DGN10 | C8202 | Follicular lymphoma grade I, intrathoracic lymph nodes | Diagnosis | Non-Hodgkins Lymphoma |
| DGN10 | C8203 | Follicular lymphoma grade I, intra-abdominal lymph nodes | Diagnosis | Non-Hodgkins Lymphoma |
| DGN10 | C8204 | Follicular lymphoma grade I, lymph nodes of axilla and upper limb | Diagnosis | Non-Hodgkins Lymphoma |
| DGN10 | C8205 | Follicular lymphoma grade I, lymph nodes of inguinal region and lower limb | Diagnosis | Non-Hodgkins Lymphoma |
| DGN10 | C8206 | Follicular lymphoma grade I, intrapelvic lymph nodes | Diagnosis | Non-Hodgkins Lymphoma |
| DGN10 | C8207 | Follicular lymphoma grade I, spleen | Diagnosis | Non-Hodgkins Lymphoma |
| DGN10 | C8208 | Follicular lymphoma grade I, lymph nodes of multiple sites | Diagnosis | Non-Hodgkins Lymphoma |
| DGN10 | C8209 | Follicular lymphoma grade I, extranodal and solid organ sites | Diagnosis | Non-Hodgkins Lymphoma |
| DGN10 | C8210 | Follicular lymphoma grade II, unspecified site | Diagnosis | Non-Hodgkins Lymphoma |
| DGN10 | C8211 | Follicular lymphoma grade II, lymph nodes of head, face, and neck | Diagnosis | Non-Hodgkins Lymphoma |
| DGN10 | C8212 | Follicular lymphoma grade II, intrathoracic lymph nodes | Diagnosis | Non-Hodgkins Lymphoma |
| DGN10 | C8213 | Follicular lymphoma grade II, intra-abdominal lymph nodes | Diagnosis | Non-Hodgkins Lymphoma |
| DGN10 | C8214 | Follicular lymphoma grade II, lymph nodes of axilla and upper limb | Diagnosis | Non-Hodgkins Lymphoma |
| DGN10 | C8215 | Follicular lymphoma grade II, lymph nodes of inguinal region and lower limb | Diagnosis | Non-Hodgkins Lymphoma |
| DGN10 | C8216 | Follicular lymphoma grade II, intrapelvic lymph nodes | Diagnosis | Non-Hodgkins Lymphoma |
| DGN10 | C8217 | Follicular lymphoma grade II, spleen | Diagnosis | Non-Hodgkins Lymphoma |
| DGN10 | C8218 | Follicular lymphoma grade II, lymph nodes of multiple sites | Diagnosis | Non-Hodgkins Lymphoma |
| DGN10 | C8219 | Follicular lymphoma grade II, extranodal and solid organ sites | Diagnosis | Non-Hodgkins Lymphoma |
| DGN10 | C8220 | Follicular lymphoma grade III, unspecified, unspecified site | Diagnosis | Non-Hodgkins Lymphoma |
| DGN10 | C8221 | Follicular lymphoma grade III, unspecified, lymph nodes of head, face, and neck | Diagnosis | Non-Hodgkins Lymphoma |
| DGN10 | C8222 | Follicular lymphoma grade III, unspecified, intrathoracic lymph nodes | Diagnosis | Non-Hodgkins Lymphoma |
| DGN10 | C8223 | Follicular lymphoma grade III, unspecified, intra-abdominal lymph nodes | Diagnosis | Non-Hodgkins Lymphoma |
| DGN10 | C8224 | Follicular lymphoma grade III, unspecified, lymph nodes of axilla and upper limb | Diagnosis | Non-Hodgkins Lymphoma |
| DGN10 | C8225 | Follicular lymphoma grade III, unspecified, lymph nodes of inguinal region and lower limb | Diagnosis | Non-Hodgkins Lymphoma |
| DGN10 | C8226 | Follicular lymphoma grade III, unspecified, intrapelvic lymph nodes | Diagnosis | Non-Hodgkins Lymphoma |
| DGN10 | C8227 | Follicular lymphoma grade III, unspecified, spleen | Diagnosis | Non-Hodgkins Lymphoma |
| DGN10 | C8228 | Follicular lymphoma grade III, unspecified, lymph nodes of multiple sites | Diagnosis | Non-Hodgkins Lymphoma |
| DGN10 | C8229 | Follicular lymphoma grade III, unspecified, extranodal and solid organ sites | Diagnosis | Non-Hodgkins Lymphoma |
| DGN10 | C8230 | Follicular lymphoma grade IIIa, unspecified site | Diagnosis | Non-Hodgkins Lymphoma |
| DGN10 | C8231 | Follicular lymphoma grade IIIa, lymph nodes of head, face, and neck | Diagnosis | Non-Hodgkins Lymphoma |
| DGN10 | C8232 | Follicular lymphoma grade IIIa, intrathoracic lymph nodes | Diagnosis | Non-Hodgkins Lymphoma |
| DGN10 | C8233 | Follicular lymphoma grade IIIa, intra-abdominal lymph nodes | Diagnosis | Non-Hodgkins Lymphoma |
| DGN10 | C8234 | Follicular lymphoma grade IIIa, lymph nodes of axilla and upper limb | Diagnosis | Non-Hodgkins Lymphoma |
| DGN10 | C8235 | Follicular lymphoma grade IIIa, lymph nodes of inguinal region and lower limb | Diagnosis | Non-Hodgkins Lymphoma |
| DGN10 | C8236 | Follicular lymphoma grade IIIa, intrapelvic lymph nodes | Diagnosis | Non-Hodgkins Lymphoma |
| DGN10 | C8237 | Follicular lymphoma grade IIIa, spleen | Diagnosis | Non-Hodgkins Lymphoma |
| DGN10 | C8238 | Follicular lymphoma grade IIIa, lymph nodes of multiple sites | Diagnosis | Non-Hodgkins Lymphoma |
| DGN10 | C8239 | Follicular lymphoma grade IIIa, extranodal and solid organ sites | Diagnosis | Non-Hodgkins Lymphoma |
| DGN10 | C8240 | Follicular lymphoma grade IIIb, unspecified site | Diagnosis | Non-Hodgkins Lymphoma |
| DGN10 | C8241 | Follicular lymphoma grade IIIb, lymph nodes of head, face, and neck | Diagnosis | Non-Hodgkins Lymphoma |
| DGN10 | C8242 | Follicular lymphoma grade IIIb, intrathoracic lymph nodes | Diagnosis | Non-Hodgkins Lymphoma |
| DGN10 | C8243 | Follicular lymphoma grade IIIb, intra-abdominal lymph nodes | Diagnosis | Non-Hodgkins Lymphoma |
| DGN10 | C8244 | Follicular lymphoma grade IIIb, lymph nodes of axilla and upper limb | Diagnosis | Non-Hodgkins Lymphoma |
| DGN10 | C8245 | Follicular lymphoma grade IIIb, lymph nodes of inguinal region and lower limb | Diagnosis | Non-Hodgkins Lymphoma |
| DGN10 | C8246 | Follicular lymphoma grade IIIb, intrapelvic lymph nodes | Diagnosis | Non-Hodgkins Lymphoma |
| DGN10 | C8247 | Follicular lymphoma grade IIIb, spleen | Diagnosis | Non-Hodgkins Lymphoma |
| DGN10 | C8248 | Follicular lymphoma grade IIIb, lymph nodes of multiple sites | Diagnosis | Non-Hodgkins Lymphoma |
| DGN10 | C8249 | Follicular lymphoma grade IIIb, extranodal and solid organ sites | Diagnosis | Non-Hodgkins Lymphoma |
| DGN10 | C8250 | Diffuse follicle center lymphoma, unspecified site | Diagnosis | Non-Hodgkins Lymphoma |
| DGN10 | C8251 | Diffuse follicle center lymphoma, lymph nodes of head, face, and neck | Diagnosis | Non-Hodgkins Lymphoma |
| DGN10 | C8252 | Diffuse follicle center lymphoma, intrathoracic lymph nodes | Diagnosis | Non-Hodgkins Lymphoma |
| DGN10 | C8253 | Diffuse follicle center lymphoma, intra-abdominal lymph nodes | Diagnosis | Non-Hodgkins Lymphoma |
| DGN10 | C8254 | Diffuse follicle center lymphoma, lymph nodes of axilla and upper limb | Diagnosis | Non-Hodgkins Lymphoma |
| DGN10 | C8255 | Diffuse follicle center lymphoma, lymph nodes of inguinal region and lower limb | Diagnosis | Non-Hodgkins Lymphoma |
| DGN10 | C8256 | Diffuse follicle center lymphoma, intrapelvic lymph nodes | Diagnosis | Non-Hodgkins Lymphoma |
| DGN10 | C8257 | Diffuse follicle center lymphoma, spleen | Diagnosis | Non-Hodgkins Lymphoma |
| DGN10 | C8258 | Diffuse follicle center lymphoma, lymph nodes of multiple sites | Diagnosis | Non-Hodgkins Lymphoma |
| DGN10 | C8259 | Diffuse follicle center lymphoma, extranodal and solid organ sites | Diagnosis | Non-Hodgkins Lymphoma |
| DGN10 | C8260 | Cutaneous follicle center lymphoma, unspecified site | Diagnosis | Non-Hodgkins Lymphoma |
| DGN10 | C8261 | Cutaneous follicle center lymphoma, lymph nodes of head, face, and neck | Diagnosis | Non-Hodgkins Lymphoma |
| DGN10 | C8262 | Cutaneous follicle center lymphoma, intrathoracic lymph nodes | Diagnosis | Non-Hodgkins Lymphoma |
| DGN10 | C8263 | Cutaneous follicle center lymphoma, intra-abdominal lymph nodes | Diagnosis | Non-Hodgkins Lymphoma |
| DGN10 | C8264 | Cutaneous follicle center lymphoma, lymph nodes of axilla and upper limb | Diagnosis | Non-Hodgkins Lymphoma |
| DGN10 | C8265 | Cutaneous follicle center lymphoma, lymph nodes of inguinal region and lower limb | Diagnosis | Non-Hodgkins Lymphoma |
| DGN10 | C8266 | Cutaneous follicle center lymphoma, intrapelvic lymph nodes | Diagnosis | Non-Hodgkins Lymphoma |
| DGN10 | C8267 | Cutaneous follicle center lymphoma, spleen | Diagnosis | Non-Hodgkins Lymphoma |
| DGN10 | C8268 | Cutaneous follicle center lymphoma, lymph nodes of multiple sites | Diagnosis | Non-Hodgkins Lymphoma |
| DGN10 | C8269 | Cutaneous follicle center lymphoma, extranodal and solid organ sites | Diagnosis | Non-Hodgkins Lymphoma |
| DGN10 | C8280 | Other types of follicular lymphoma, unspecified site | Diagnosis | Non-Hodgkins Lymphoma |
| DGN10 | C8281 | Other types of follicular lymphoma, lymph nodes of head, face, and neck | Diagnosis | Non-Hodgkins Lymphoma |
| DGN10 | C8282 | Other types of follicular lymphoma, intrathoracic lymph nodes | Diagnosis | Non-Hodgkins Lymphoma |
| DGN10 | C8283 | Other types of follicular lymphoma, intra-abdominal lymph nodes | Diagnosis | Non-Hodgkins Lymphoma |
| DGN10 | C8284 | Other types of follicular lymphoma, lymph nodes of axilla and upper limb | Diagnosis | Non-Hodgkins Lymphoma |
| DGN10 | C8285 | Other types of follicular lymphoma, lymph nodes of inguinal region and lower limb | Diagnosis | Non-Hodgkins Lymphoma |
| DGN10 | C8286 | Other types of follicular lymphoma, intrapelvic lymph nodes | Diagnosis | Non-Hodgkins Lymphoma |
| DGN10 | C8287 | Other types of follicular lymphoma, spleen | Diagnosis | Non-Hodgkins Lymphoma |
| DGN10 | C8288 | Other types of follicular lymphoma, lymph nodes of multiple sites | Diagnosis | Non-Hodgkins Lymphoma |
| DGN10 | C8289 | Other types of follicular lymphoma, extranodal and solid organ sites | Diagnosis | Non-Hodgkins Lymphoma |
| DGN10 | C8290 | Follicular lymphoma, unspecified, unspecified site | Diagnosis | Non-Hodgkins Lymphoma |
| DGN10 | C8291 | Follicular lymphoma, unspecified, lymph nodes of head, face, and neck | Diagnosis | Non-Hodgkins Lymphoma |
| DGN10 | C8292 | Follicular lymphoma, unspecified, intrathoracic lymph nodes | Diagnosis | Non-Hodgkins Lymphoma |
| DGN10 | C8293 | Follicular lymphoma, unspecified, intra-abdominal lymph nodes | Diagnosis | Non-Hodgkins Lymphoma |
| DGN10 | C8294 | Follicular lymphoma, unspecified, lymph nodes of axilla and upper limb | Diagnosis | Non-Hodgkins Lymphoma |
| DGN10 | C8295 | Follicular lymphoma, unspecified, lymph nodes of inguinal region and lower limb | Diagnosis | Non-Hodgkins Lymphoma |
| DGN10 | C8296 | Follicular lymphoma, unspecified, intrapelvic lymph nodes | Diagnosis | Non-Hodgkins Lymphoma |
| DGN10 | C8297 | Follicular lymphoma, unspecified, spleen | Diagnosis | Non-Hodgkins Lymphoma |
| DGN10 | C8298 | Follicular lymphoma, unspecified, lymph nodes of multiple sites | Diagnosis | Non-Hodgkins Lymphoma |
| DGN10 | C8299 | Follicular lymphoma, unspecified, extranodal and solid organ sites | Diagnosis | Non-Hodgkins Lymphoma |
| DGN10 | C8300 | Small cell B-cell lymphoma, unspecified site | Diagnosis | Non-Hodgkins Lymphoma |
| DGN10 | C8301 | Small cell B-cell lymphoma, lymph nodes of head, face, and neck | Diagnosis | Non-Hodgkins Lymphoma |
| DGN10 | C8302 | Small cell B-cell lymphoma, intrathoracic lymph nodes | Diagnosis | Non-Hodgkins Lymphoma |
| DGN10 | C8303 | Small cell B-cell lymphoma, intra-abdominal lymph nodes | Diagnosis | Non-Hodgkins Lymphoma |
| DGN10 | C8304 | Small cell B-cell lymphoma, lymph nodes of axilla and upper limb | Diagnosis | Non-Hodgkins Lymphoma |
| DGN10 | C8305 | Small cell B-cell lymphoma, lymph nodes of inguinal region and lower limb | Diagnosis | Non-Hodgkins Lymphoma |
| DGN10 | C8306 | Small cell B-cell lymphoma, intrapelvic lymph nodes | Diagnosis | Non-Hodgkins Lymphoma |
| DGN10 | C8307 | Small cell B-cell lymphoma, spleen | Diagnosis | Non-Hodgkins Lymphoma |
| DGN10 | C8308 | Small cell B-cell lymphoma, lymph nodes of multiple sites | Diagnosis | Non-Hodgkins Lymphoma |
| DGN10 | C8309 | Small cell B-cell lymphoma, extranodal and solid organ sites | Diagnosis | Non-Hodgkins Lymphoma |
| DGN10 | C8310 | Mantle cell lymphoma, unspecified site | Diagnosis | Non-Hodgkins Lymphoma |
| DGN10 | C8311 | Mantle cell lymphoma, lymph nodes of head, face, and neck | Diagnosis | Non-Hodgkins Lymphoma |
| DGN10 | C8312 | Mantle cell lymphoma, intrathoracic lymph nodes | Diagnosis | Non-Hodgkins Lymphoma |
| DGN10 | C8313 | Mantle cell lymphoma, intra-abdominal lymph nodes | Diagnosis | Non-Hodgkins Lymphoma |
| DGN10 | C8314 | Mantle cell lymphoma, lymph nodes of axilla and upper limb | Diagnosis | Non-Hodgkins Lymphoma |
| DGN10 | C8315 | Mantle cell lymphoma, lymph nodes of inguinal region and lower limb | Diagnosis | Non-Hodgkins Lymphoma |
| DGN10 | C8316 | Mantle cell lymphoma, intrapelvic lymph nodes | Diagnosis | Non-Hodgkins Lymphoma |
| DGN10 | C8317 | Mantle cell lymphoma, spleen | Diagnosis | Non-Hodgkins Lymphoma |
| DGN10 | C8318 | Mantle cell lymphoma, lymph nodes of multiple sites | Diagnosis | Non-Hodgkins Lymphoma |
| DGN10 | C8319 | Mantle cell lymphoma, extranodal and solid organ sites | Diagnosis | Non-Hodgkins Lymphoma |
| DGN10 | C8330 | Diffuse large B-cell lymphoma, unspecified site | Diagnosis | Non-Hodgkins Lymphoma |
| DGN10 | C8331 | Diffuse large B-cell lymphoma, lymph nodes of head, face, and neck | Diagnosis | Non-Hodgkins Lymphoma |
| DGN10 | C8332 | Diffuse large B-cell lymphoma, intrathoracic lymph nodes | Diagnosis | Non-Hodgkins Lymphoma |
| DGN10 | C8333 | Diffuse large B-cell lymphoma, intra-abdominal lymph nodes | Diagnosis | Non-Hodgkins Lymphoma |
| DGN10 | C8334 | Diffuse large B-cell lymphoma, lymph nodes of axilla and upper limb | Diagnosis | Non-Hodgkins Lymphoma |
| DGN10 | C8335 | Diffuse large B-cell lymphoma, lymph nodes of inguinal region and lower limb | Diagnosis | Non-Hodgkins Lymphoma |
| DGN10 | C8336 | Diffuse large B-cell lymphoma, intrapelvic lymph nodes | Diagnosis | Non-Hodgkins Lymphoma |
| DGN10 | C8337 | Diffuse large B-cell lymphoma, spleen | Diagnosis | Non-Hodgkins Lymphoma |
| DGN10 | C8338 | Diffuse large B-cell lymphoma, lymph nodes of multiple sites | Diagnosis | Non-Hodgkins Lymphoma |
| DGN10 | C8339 | Diffuse large B-cell lymphoma, extranodal and solid organ sites | Diagnosis | Non-Hodgkins Lymphoma |
| DGN10 | C8350 | Lymphoblastic (diffuse) lymphoma, unspecified site | Diagnosis | Non-Hodgkins Lymphoma |
| DGN10 | C8351 | Lymphoblastic (diffuse) lymphoma, lymph nodes of head, face, and neck | Diagnosis | Non-Hodgkins Lymphoma |
| DGN10 | C8352 | Lymphoblastic (diffuse) lymphoma, intrathoracic lymph nodes | Diagnosis | Non-Hodgkins Lymphoma |
| DGN10 | C8353 | Lymphoblastic (diffuse) lymphoma, intra-abdominal lymph nodes | Diagnosis | Non-Hodgkins Lymphoma |
| DGN10 | C8354 | Lymphoblastic (diffuse) lymphoma, lymph nodes of axilla and upper limb | Diagnosis | Non-Hodgkins Lymphoma |
| DGN10 | C8355 | Lymphoblastic (diffuse) lymphoma, lymph nodes of inguinal region and lower limb | Diagnosis | Non-Hodgkins Lymphoma |
| DGN10 | C8356 | Lymphoblastic (diffuse) lymphoma, intrapelvic lymph nodes | Diagnosis | Non-Hodgkins Lymphoma |
| DGN10 | C8357 | Lymphoblastic (diffuse) lymphoma, spleen | Diagnosis | Non-Hodgkins Lymphoma |
| DGN10 | C8358 | Lymphoblastic (diffuse) lymphoma, lymph nodes of multiple sites | Diagnosis | Non-Hodgkins Lymphoma |
| DGN10 | C8359 | Lymphoblastic (diffuse) lymphoma, extranodal and solid organ sites | Diagnosis | Non-Hodgkins Lymphoma |
| DGN10 | C8370 | Burkitt lymphoma, unspecified site | Diagnosis | Non-Hodgkins Lymphoma |
| DGN10 | C8371 | Burkitt lymphoma, lymph nodes of head, face, and neck | Diagnosis | Non-Hodgkins Lymphoma |
| DGN10 | C8372 | Burkitt lymphoma, intrathoracic lymph nodes | Diagnosis | Non-Hodgkins Lymphoma |
| DGN10 | C8373 | Burkitt lymphoma, intra-abdominal lymph nodes | Diagnosis | Non-Hodgkins Lymphoma |
| DGN10 | C8374 | Burkitt lymphoma, lymph nodes of axilla and upper limb | Diagnosis | Non-Hodgkins Lymphoma |
| DGN10 | C8375 | Burkitt lymphoma, lymph nodes of inguinal region and lower limb | Diagnosis | Non-Hodgkins Lymphoma |
| DGN10 | C8376 | Burkitt lymphoma, intrapelvic lymph nodes | Diagnosis | Non-Hodgkins Lymphoma |
| DGN10 | C8377 | Burkitt lymphoma, spleen | Diagnosis | Non-Hodgkins Lymphoma |
| DGN10 | C8378 | Burkitt lymphoma, lymph nodes of multiple sites | Diagnosis | Non-Hodgkins Lymphoma |
| DGN10 | C8379 | Burkitt lymphoma, extranodal and solid organ sites | Diagnosis | Non-Hodgkins Lymphoma |
| DGN10 | C8380 | Other non-follicular lymphoma, unspecified site | Diagnosis | Non-Hodgkins Lymphoma |
| DGN10 | C8381 | Other non-follicular lymphoma, lymph nodes of head, face, and neck | Diagnosis | Non-Hodgkins Lymphoma |
| DGN10 | C8382 | Other non-follicular lymphoma, intrathoracic lymph nodes | Diagnosis | Non-Hodgkins Lymphoma |
| DGN10 | C8383 | Other non-follicular lymphoma, intra-abdominal lymph nodes | Diagnosis | Non-Hodgkins Lymphoma |
| DGN10 | C8384 | Other non-follicular lymphoma, lymph nodes of axilla and upper limb | Diagnosis | Non-Hodgkins Lymphoma |
| DGN10 | C8385 | Other non-follicular lymphoma, lymph nodes of inguinal region and lower limb | Diagnosis | Non-Hodgkins Lymphoma |
| DGN10 | C8386 | Other non-follicular lymphoma, intrapelvic lymph nodes | Diagnosis | Non-Hodgkins Lymphoma |
| DGN10 | C8387 | Other non-follicular lymphoma, spleen | Diagnosis | Non-Hodgkins Lymphoma |
| DGN10 | C8388 | Other non-follicular lymphoma, lymph nodes of multiple sites | Diagnosis | Non-Hodgkins Lymphoma |
| DGN10 | C8389 | Other non-follicular lymphoma, extranodal and solid organ sites | Diagnosis | Non-Hodgkins Lymphoma |
| DGN10 | C8390 | Non-follicular (diffuse) lymphoma, unspecified, unspecified site | Diagnosis | Non-Hodgkins Lymphoma |
| DGN10 | C8391 | Non-follicular (diffuse) lymphoma, unspecified, lymph nodes of head, face, and neck | Diagnosis | Non-Hodgkins Lymphoma |
| DGN10 | C8392 | Non-follicular (diffuse) lymphoma, unspecified, intrathoracic lymph nodes | Diagnosis | Non-Hodgkins Lymphoma |
| DGN10 | C8393 | Non-follicular (diffuse) lymphoma, unspecified, intra-abdominal lymph nodes | Diagnosis | Non-Hodgkins Lymphoma |
| DGN10 | C8394 | Non-follicular (diffuse) lymphoma, unspecified, lymph nodes of axilla and upper limb | Diagnosis | Non-Hodgkins Lymphoma |
| DGN10 | C8395 | Non-follicular (diffuse) lymphoma, unspecified, lymph nodes of inguinal region and lower limb | Diagnosis | Non-Hodgkins Lymphoma |
| DGN10 | C8396 | Non-follicular (diffuse) lymphoma, unspecified, intrapelvic lymph nodes | Diagnosis | Non-Hodgkins Lymphoma |
| DGN10 | C8397 | Non-follicular (diffuse) lymphoma, unspecified, spleen | Diagnosis | Non-Hodgkins Lymphoma |
| DGN10 | C8398 | Non-follicular (diffuse) lymphoma, unspecified, lymph nodes of multiple sites | Diagnosis | Non-Hodgkins Lymphoma |
| DGN10 | C8399 | Non-follicular (diffuse) lymphoma, unspecified, extranodal and solid organ sites | Diagnosis | Non-Hodgkins Lymphoma |
| DGN10 | C8400 | Mycosis fungoides, unspecified site | Diagnosis | Non-Hodgkins Lymphoma |
| DGN10 | C8401 | Mycosis fungoides, lymph nodes of head, face, and neck | Diagnosis | Non-Hodgkins Lymphoma |
| DGN10 | C8402 | Mycosis fungoides, intrathoracic lymph nodes | Diagnosis | Non-Hodgkins Lymphoma |
| DGN10 | C8403 | Mycosis fungoides, intra-abdominal lymph nodes | Diagnosis | Non-Hodgkins Lymphoma |
| DGN10 | C8404 | Mycosis fungoides, lymph nodes of axilla and upper limb | Diagnosis | Non-Hodgkins Lymphoma |
| DGN10 | C8405 | Mycosis fungoides, lymph nodes of inguinal region and lower limb | Diagnosis | Non-Hodgkins Lymphoma |
| DGN10 | C8406 | Mycosis fungoides, intrapelvic lymph nodes | Diagnosis | Non-Hodgkins Lymphoma |
| DGN10 | C8407 | Mycosis fungoides, spleen | Diagnosis | Non-Hodgkins Lymphoma |
| DGN10 | C8408 | Mycosis fungoides, lymph nodes of multiple sites | Diagnosis | Non-Hodgkins Lymphoma |
| DGN10 | C8409 | Mycosis fungoides, extranodal and solid organ sites | Diagnosis | Non-Hodgkins Lymphoma |
| DGN10 | C8410 | Sezary disease, unspecified site | Diagnosis | Non-Hodgkins Lymphoma |
| DGN10 | C8411 | Sezary disease, lymph nodes of head, face, and neck | Diagnosis | Non-Hodgkins Lymphoma |
| DGN10 | C8412 | Sezary disease, intrathoracic lymph nodes | Diagnosis | Non-Hodgkins Lymphoma |
| DGN10 | C8413 | Sezary disease, intra-abdominal lymph nodes | Diagnosis | Non-Hodgkins Lymphoma |
| DGN10 | C8414 | Sezary disease, lymph nodes of axilla and upper limb | Diagnosis | Non-Hodgkins Lymphoma |
| DGN10 | C8415 | Sezary disease, lymph nodes of inguinal region and lower limb | Diagnosis | Non-Hodgkins Lymphoma |
| DGN10 | C8416 | Sezary disease, intrapelvic lymph nodes | Diagnosis | Non-Hodgkins Lymphoma |
| DGN10 | C8417 | Sezary disease, spleen | Diagnosis | Non-Hodgkins Lymphoma |
| DGN10 | C8418 | Sezary disease, lymph nodes of multiple sites | Diagnosis | Non-Hodgkins Lymphoma |
| DGN10 | C8419 | Sezary disease, extranodal and solid organ sites | Diagnosis | Non-Hodgkins Lymphoma |
| DGN10 | C8460 | Anaplastic large cell lymphoma, ALK-positive, unspecified site | Diagnosis | Non-Hodgkins Lymphoma |
| DGN10 | C8461 | Anaplastic large cell lymphoma, ALK-positive, lymph nodes of head, face, and neck | Diagnosis | Non-Hodgkins Lymphoma |
| DGN10 | C8462 | Anaplastic large cell lymphoma, ALK-positive, intrathoracic lymph nodes | Diagnosis | Non-Hodgkins Lymphoma |
| DGN10 | C8463 | Anaplastic large cell lymphoma, ALK-positive, intra-abdominal lymph nodes | Diagnosis | Non-Hodgkins Lymphoma |
| DGN10 | C8464 | Anaplastic large cell lymphoma, ALK-positive, lymph nodes of axilla and upper limb | Diagnosis | Non-Hodgkins Lymphoma |
| DGN10 | C8465 | Anaplastic large cell lymphoma, ALK-positive, lymph nodes of inguinal region and lower limb | Diagnosis | Non-Hodgkins Lymphoma |
| DGN10 | C8466 | Anaplastic large cell lymphoma, ALK-positive, intrapelvic lymph nodes | Diagnosis | Non-Hodgkins Lymphoma |
| DGN10 | C8467 | Anaplastic large cell lymphoma, ALK-positive, spleen | Diagnosis | Non-Hodgkins Lymphoma |
| DGN10 | C8468 | Anaplastic large cell lymphoma, ALK-positive, lymph nodes of multiple sites | Diagnosis | Non-Hodgkins Lymphoma |
| DGN10 | C8469 | Anaplastic large cell lymphoma, ALK-positive, extranodal and solid organ sites | Diagnosis | Non-Hodgkins Lymphoma |
| DGN10 | C8470 | Anaplastic large cell lymphoma, ALK-negative, unspecified site | Diagnosis | Non-Hodgkins Lymphoma |
| DGN10 | C8471 | Anaplastic large cell lymphoma, ALK-negative, lymph nodes of head, face, and neck | Diagnosis | Non-Hodgkins Lymphoma |
| DGN10 | C8472 | Anaplastic large cell lymphoma, ALK-negative, intrathoracic lymph nodes | Diagnosis | Non-Hodgkins Lymphoma |
| DGN10 | C8473 | Anaplastic large cell lymphoma, ALK-negative, intra-abdominal lymph nodes | Diagnosis | Non-Hodgkins Lymphoma |
| DGN10 | C8474 | Anaplastic large cell lymphoma, ALK-negative, lymph nodes of axilla and upper limb | Diagnosis | Non-Hodgkins Lymphoma |
| DGN10 | C8475 | Anaplastic large cell lymphoma, ALK-negative, lymph nodes of inguinal region and lower limb | Diagnosis | Non-Hodgkins Lymphoma |
| DGN10 | C8476 | Anaplastic large cell lymphoma, ALK-negative, intrapelvic lymph nodes | Diagnosis | Non-Hodgkins Lymphoma |
| DGN10 | C8477 | Anaplastic large cell lymphoma, ALK-negative, spleen | Diagnosis | Non-Hodgkins Lymphoma |
| DGN10 | C8478 | Anaplastic large cell lymphoma, ALK-negative, lymph nodes of multiple sites | Diagnosis | Non-Hodgkins Lymphoma |
| DGN10 | C8479 | Anaplastic large cell lymphoma, ALK-negative, extranodal and solid organ sites | Diagnosis | Non-Hodgkins Lymphoma |
| DGN10 | C8490 | Mature T/NK-cell lymphomas, unspecified, unspecified site | Diagnosis | Non-Hodgkins Lymphoma |
| DGN10 | C8491 | Mature T/NK-cell lymphomas, unspecified, lymph nodes of head, face, and neck | Diagnosis | Non-Hodgkins Lymphoma |
| DGN10 | C8492 | Mature T/NK-cell lymphomas, unspecified, intrathoracic lymph nodes | Diagnosis | Non-Hodgkins Lymphoma |
| DGN10 | C8493 | Mature T/NK-cell lymphomas, unspecified, intra-abdominal lymph nodes | Diagnosis | Non-Hodgkins Lymphoma |
| DGN10 | C8494 | Mature T/NK-cell lymphomas, unspecified, lymph nodes of axilla and upper limb | Diagnosis | Non-Hodgkins Lymphoma |
| DGN10 | C8495 | Mature T/NK-cell lymphomas, unspecified, lymph nodes of inguinal region and lower limb | Diagnosis | Non-Hodgkins Lymphoma |
| DGN10 | C8496 | Mature T/NK-cell lymphomas, unspecified, intrapelvic lymph nodes | Diagnosis | Non-Hodgkins Lymphoma |
| DGN10 | C8497 | Mature T/NK-cell lymphomas, unspecified, spleen | Diagnosis | Non-Hodgkins Lymphoma |
| DGN10 | C8498 | Mature T/NK-cell lymphomas, unspecified, lymph nodes of multiple sites | Diagnosis | Non-Hodgkins Lymphoma |
| DGN10 | C8499 | Mature T/NK-cell lymphomas, unspecified, extranodal and solid organ sites | Diagnosis | Non-Hodgkins Lymphoma |
| DGN10 | C84A0 | Cutaneous T-cell lymphoma, unspecified, unspecified site | Diagnosis | Non-Hodgkins Lymphoma |
| DGN10 | C84A1 | Cutaneous T-cell lymphoma, unspecified lymph nodes of head, face, and neck | Diagnosis | Non-Hodgkins Lymphoma |
| DGN10 | C84A2 | Cutaneous T-cell lymphoma, unspecified, intrathoracic lymph nodes | Diagnosis | Non-Hodgkins Lymphoma |
| DGN10 | C84A3 | Cutaneous T-cell lymphoma, unspecified, intra-abdominal lymph nodes | Diagnosis | Non-Hodgkins Lymphoma |
| DGN10 | C84A4 | Cutaneous T-cell lymphoma, unspecified, lymph nodes of axilla and upper limb | Diagnosis | Non-Hodgkins Lymphoma |
| DGN10 | C84A5 | Cutaneous T-cell lymphoma, unspecified, lymph nodes of inguinal region and lower limb | Diagnosis | Non-Hodgkins Lymphoma |
| DGN10 | C84A6 | Cutaneous T-cell lymphoma, unspecified, intrapelvic lymph nodes | Diagnosis | Non-Hodgkins Lymphoma |
| DGN10 | C84A7 | Cutaneous T-cell lymphoma, unspecified, spleen | Diagnosis | Non-Hodgkins Lymphoma |
| DGN10 | C84A8 | Cutaneous T-cell lymphoma, unspecified, lymph nodes of multiple sites | Diagnosis | Non-Hodgkins Lymphoma |
| DGN10 | C84A9 | Cutaneous T-cell lymphoma, unspecified, extranodal and solid organ sites | Diagnosis | Non-Hodgkins Lymphoma |
| DGN10 | C84Z0 | Other mature T/NK-cell lymphomas, unspecified site | Diagnosis | Non-Hodgkins Lymphoma |
| DGN10 | C84Z1 | Other mature T/NK-cell lymphomas, lymph nodes of head, face, and neck | Diagnosis | Non-Hodgkins Lymphoma |
| DGN10 | C84Z2 | Other mature T/NK-cell lymphomas, intrathoracic lymph nodes | Diagnosis | Non-Hodgkins Lymphoma |
| DGN10 | C84Z3 | Other mature T/NK-cell lymphomas, intra-abdominal lymph nodes | Diagnosis | Non-Hodgkins Lymphoma |
| DGN10 | C84Z4 | Other mature T/NK-cell lymphomas, lymph nodes of axilla and upper limb | Diagnosis | Non-Hodgkins Lymphoma |
| DGN10 | C84Z5 | Other mature T/NK-cell lymphomas, lymph nodes of inguinal region and lower limb | Diagnosis | Non-Hodgkins Lymphoma |
| DGN10 | C84Z6 | Other mature T/NK-cell lymphomas, intrapelvic lymph nodes | Diagnosis | Non-Hodgkins Lymphoma |
| DGN10 | C84Z7 | Other mature T/NK-cell lymphomas, spleen | Diagnosis | Non-Hodgkins Lymphoma |
| DGN10 | C84Z8 | Other mature T/NK-cell lymphomas, lymph nodes of multiple sites | Diagnosis | Non-Hodgkins Lymphoma |
| DGN10 | C84Z9 | Other mature T/NK-cell lymphomas, extranodal and solid organ sites | Diagnosis | Non-Hodgkins Lymphoma |
| DGN10 | C8510 | Unspecified B-cell lymphoma, unspecified site | Diagnosis | Non-Hodgkins Lymphoma |
| DGN10 | C8511 | Unspecified B-cell lymphoma, lymph nodes of head, face, and neck | Diagnosis | Non-Hodgkins Lymphoma |
| DGN10 | C8512 | Unspecified B-cell lymphoma, intrathoracic lymph nodes | Diagnosis | Non-Hodgkins Lymphoma |
| DGN10 | C8513 | Unspecified B-cell lymphoma, intra-abdominal lymph nodes | Diagnosis | Non-Hodgkins Lymphoma |
| DGN10 | C8514 | Unspecified B-cell lymphoma, lymph nodes of axilla and upper limb | Diagnosis | Non-Hodgkins Lymphoma |
| DGN10 | C8515 | Unspecified B-cell lymphoma, lymph nodes of inguinal region and lower limb | Diagnosis | Non-Hodgkins Lymphoma |
| DGN10 | C8516 | Unspecified B-cell lymphoma, intrapelvic lymph nodes | Diagnosis | Non-Hodgkins Lymphoma |
| DGN10 | C8517 | Unspecified B-cell lymphoma, spleen | Diagnosis | Non-Hodgkins Lymphoma |
| DGN10 | C8518 | Unspecified B-cell lymphoma, lymph nodes of multiple sites | Diagnosis | Non-Hodgkins Lymphoma |
| DGN10 | C8519 | Unspecified B-cell lymphoma, extranodal and solid organ sites | Diagnosis | Non-Hodgkins Lymphoma |
| DGN10 | C8520 | Mediastinal (thymic) large B-cell lymphoma, unspecified site | Diagnosis | Non-Hodgkins Lymphoma |
| DGN10 | C8521 | Mediastinal (thymic) large B-cell lymphoma, lymph nodes of head, face, and neck | Diagnosis | Non-Hodgkins Lymphoma |
| DGN10 | C8522 | Mediastinal (thymic) large B-cell lymphoma, intrathoracic lymph nodes | Diagnosis | Non-Hodgkins Lymphoma |
| DGN10 | C8523 | Mediastinal (thymic) large B-cell lymphoma, intra-abdominal lymph nodes | Diagnosis | Non-Hodgkins Lymphoma |
| DGN10 | C8524 | Mediastinal (thymic) large B-cell lymphoma, lymph nodes of axilla and upper limb | Diagnosis | Non-Hodgkins Lymphoma |
| DGN10 | C8525 | Mediastinal (thymic) large B-cell lymphoma, lymph nodes of inguinal region and lower limb | Diagnosis | Non-Hodgkins Lymphoma |
| DGN10 | C8526 | Mediastinal (thymic) large B-cell lymphoma, intrapelvic lymph nodes | Diagnosis | Non-Hodgkins Lymphoma |
| DGN10 | C8527 | Mediastinal (thymic) large B-cell lymphoma, spleen | Diagnosis | Non-Hodgkins Lymphoma |
| DGN10 | C8528 | Mediastinal (thymic) large B-cell lymphoma, lymph nodes of multiple sites | Diagnosis | Non-Hodgkins Lymphoma |
| DGN10 | C8529 | Mediastinal (thymic) large B-cell lymphoma, extranodal and solid organ sites | Diagnosis | Non-Hodgkins Lymphoma |
| DGN10 | C8580 | Other specified types of non-Hodgkin lymphoma, unspecified site | Diagnosis | Non-Hodgkins Lymphoma |
| DGN10 | C8581 | Other specified types of non-Hodgkin lymphoma, lymph nodes of head, face, and neck | Diagnosis | Non-Hodgkins Lymphoma |
| DGN10 | C8582 | Other specified types of non-Hodgkin lymphoma, intrathoracic lymph nodes | Diagnosis | Non-Hodgkins Lymphoma |
| DGN10 | C8583 | Other specified types of non-Hodgkin lymphoma, intra-abdominal lymph nodes | Diagnosis | Non-Hodgkins Lymphoma |
| DGN10 | C8584 | Other specified types of non-Hodgkin lymphoma, lymph nodes of axilla and upper limb | Diagnosis | Non-Hodgkins Lymphoma |
| DGN10 | C8585 | Other specified types of non-Hodgkin lymphoma, lymph nodes of inguinal region and lower limb | Diagnosis | Non-Hodgkins Lymphoma |
| DGN10 | C8586 | Other specified types of non-Hodgkin lymphoma, intrapelvic lymph nodes | Diagnosis | Non-Hodgkins Lymphoma |
| DGN10 | C8587 | Other specified types of non-Hodgkin lymphoma, spleen | Diagnosis | Non-Hodgkins Lymphoma |
| DGN10 | C8588 | Other specified types of non-Hodgkin lymphoma, lymph nodes of multiple sites | Diagnosis | Non-Hodgkins Lymphoma |
| DGN10 | C8589 | Other specified types of non-Hodgkin lymphoma, extranodal and solid organ sites | Diagnosis | Non-Hodgkins Lymphoma |
| DGN10 | C8590 | Non-Hodgkin lymphoma, unspecified, unspecified site | Diagnosis | Non-Hodgkins Lymphoma |
| DGN10 | C8591 | Non-Hodgkin lymphoma, unspecified, lymph nodes of head, face, and neck | Diagnosis | Non-Hodgkins Lymphoma |
| DGN10 | C8592 | Non-Hodgkin lymphoma, unspecified, intrathoracic lymph nodes | Diagnosis | Non-Hodgkins Lymphoma |
| DGN10 | C8593 | Non-Hodgkin lymphoma, unspecified, intra-abdominal lymph nodes | Diagnosis | Non-Hodgkins Lymphoma |
| DGN10 | C8594 | Non-Hodgkin lymphoma, unspecified, lymph nodes of axilla and upper limb | Diagnosis | Non-Hodgkins Lymphoma |
| DGN10 | C8595 | Non-Hodgkin lymphoma, unspecified, lymph nodes of inguinal region and lower limb | Diagnosis | Non-Hodgkins Lymphoma |
| DGN10 | C8596 | Non-Hodgkin lymphoma, unspecified, intrapelvic lymph nodes | Diagnosis | Non-Hodgkins Lymphoma |
| DGN10 | C8597 | Non-Hodgkin lymphoma, unspecified, spleen | Diagnosis | Non-Hodgkins Lymphoma |
| DGN10 | C8598 | Non-Hodgkin lymphoma, unspecified, lymph nodes of multiple sites | Diagnosis | Non-Hodgkins Lymphoma |
| DGN10 | C8599 | Non-Hodgkin lymphoma, unspecified, extranodal and solid organ sites | Diagnosis | Non-Hodgkins Lymphoma |
| DGN10 | C860 | Extranodal NK/T-cell lymphoma, nasal type | Diagnosis | Non-Hodgkins Lymphoma |
| DGN10 | C861 | Hepatosplenic T-cell lymphoma | Diagnosis | Non-Hodgkins Lymphoma |
| DGN10 | C862 | Enteropathy-type (intestinal) T-cell lymphoma | Diagnosis | Non-Hodgkins Lymphoma |
| DGN10 | C863 | Subcutaneous panniculitis-like T-cell lymphoma | Diagnosis | Non-Hodgkins Lymphoma |
| DGN10 | C864 | Blastic NK-cell lymphoma | Diagnosis | Non-Hodgkins Lymphoma |
| DGN10 | C865 | Angioimmunoblastic T-cell lymphoma | Diagnosis | Non-Hodgkins Lymphoma |
| DGN10 | C866 | Primary cutaneous CD30-positive T-cell proliferations | Diagnosis | Non-Hodgkins Lymphoma |
| DGN10 | C884 | Extranodal marginal zone B-cell lymphoma of mucosa-associated lymphoid tissue [MALT-lymphoma] | Diagnosis | Non-Hodgkins Lymphoma |
| DGN10 | C964 | Sarcoma of dendritic cells (accessory cells) | Diagnosis | Non-Hodgkins Lymphoma |
| DGN10 | C969 | Malignant neoplasm of lymphoid, hematopoietic and related tissue, unspecified | Diagnosis | Non-Hodgkins Lymphoma |
| DGN10 | C96Z | Other specified malignant neoplasms of lymphoid, hematopoietic and related tissue | Diagnosis | Non-Hodgkins Lymphoma |
| DGN10 | C9140 | Hairy cell leukemia not having achieved remission | Diagnosis | Non-Hodgkins Lymphoma |
| DGN10 | C9141 | Hairy cell leukemia, in remission | Diagnosis | Non-Hodgkins Lymphoma |
| DGN10 | C9142 | Hairy cell leukemia, in relapse | Diagnosis | Non-Hodgkins Lymphoma |
| DGN10 | C8440 | Peripheral T-cell lymphoma, not classified, unspecified site | Diagnosis | Non-Hodgkins Lymphoma |
| DGN10 | C8441 | Peripheral T-cell lymphoma, not classified, lymph nodes of head, face, and neck | Diagnosis | Non-Hodgkins Lymphoma |
| DGN10 | C8442 | Peripheral T-cell lymphoma, not classified, intrathoracic lymph nodes | Diagnosis | Non-Hodgkins Lymphoma |
| DGN10 | C8443 | Peripheral T-cell lymphoma, not classified, intra-abdominal lymph nodes | Diagnosis | Non-Hodgkins Lymphoma |
| DGN10 | C8444 | Peripheral T-cell lymphoma, not classified, lymph nodes of axilla and upper limb | Diagnosis | Non-Hodgkins Lymphoma |
| DGN10 | C8445 | Peripheral T-cell lymphoma, not classified, lymph nodes of inguinal region and lower limb | Diagnosis | Non-Hodgkins Lymphoma |
| DGN10 | C8446 | Peripheral T-cell lymphoma, not classified, intrapelvic lymph nodes | Diagnosis | Non-Hodgkins Lymphoma |
| DGN10 | C8447 | Peripheral T-cell lymphoma, not classified, spleen | Diagnosis | Non-Hodgkins Lymphoma |
| DGN10 | C8448 | Peripheral T-cell lymphoma, not classified, lymph nodes of multiple sites | Diagnosis | Non-Hodgkins Lymphoma |
| DGN10 | C8449 | Peripheral T-cell lymphoma, not classified, extranodal and solid organ sites | Diagnosis | Non-Hodgkins Lymphoma |
| DGN10 | C880 | Waldenstrom macroglobulinemia | Diagnosis | Non-Hodgkins Lymphoma |
| DGN10 | C91A2 | Mature B-cell leukemia Burkitt-type, in relapse | Diagnosis | Non-Hodgkins Lymphoma |
| DGN10 | C9010 | Plasma cell leukemia not having achieved remission | Diagnosis | Leukemia |
| DGN10 | C9011 | Plasma cell leukemia in remission | Diagnosis | Leukemia |
| DGN10 | C9100 | Acute lymphoblastic leukemia not having achieved remission | Diagnosis | Leukemia |
| DGN10 | C9101 | Acute lymphoblastic leukemia, in remission | Diagnosis | Leukemia |
| DGN10 | C9110 | Chronic lymphocytic leukemia of B-cell type not having achieved remission | Diagnosis | Leukemia |
| DGN10 | C9111 | Chronic lymphocytic leukemia of B-cell type in remission | Diagnosis | Leukemia |
| DGN10 | C9130 | Prolymphocytic leukemia of B-cell type not having achieved remission | Diagnosis | Leukemia |
| DGN10 | C9131 | Prolymphocytic leukemia of B-cell type, in remission | Diagnosis | Leukemia |
| DGN10 | C9150 | Adult T-cell lymphoma/leukemia (HTLV-1-associated) not having achieved remission | Diagnosis | Leukemia |
| DGN10 | C9151 | Adult T-cell lymphoma/leukemia (HTLV-1-associated), in remission | Diagnosis | Leukemia |
| DGN10 | C9160 | Prolymphocytic leukemia of T-cell type not having achieved remission | Diagnosis | Leukemia |
| DGN10 | C9161 | Prolymphocytic leukemia of T-cell type, in remission | Diagnosis | Leukemia |
| DGN10 | C9190 | Lymphoid leukemia, unspecified not having achieved remission | Diagnosis | Leukemia |
| DGN10 | C9191 | Lymphoid leukemia, unspecified, in remission | Diagnosis | Leukemia |
| DGN10 | C91A0 | Mature B-cell leukemia Burkitt-type not having achieved remission | Diagnosis | Leukemia |
| DGN10 | C91A1 | Mature B-cell leukemia Burkitt-type, in remission | Diagnosis | Leukemia |
| DGN10 | C91Z0 | Other lymphoid leukemia not having achieved remission | Diagnosis | Leukemia |
| DGN10 | C91Z1 | Other lymphoid leukemia, in remission | Diagnosis | Leukemia |
| DGN10 | C9200 | Acute myeloblastic leukemia, not having achieved remission | Diagnosis | Leukemia |
| DGN10 | C9201 | Acute myeloblastic leukemia, in remission | Diagnosis | Leukemia |
| DGN10 | C9210 | Chronic myeloid leukemia, BCR/ABL-positive, not having achieved remission | Diagnosis | Leukemia |
| DGN10 | C9211 | Chronic myeloid leukemia, BCR/ABL-positive, in remission | Diagnosis | Leukemia |
| DGN10 | C9220 | Atypical chronic myeloid leukemia, BCR/ABL-negative, not having achieved remission | Diagnosis | Leukemia |
| DGN10 | C9221 | Atypical chronic myeloid leukemia, BCR/ABL-negative, in remission | Diagnosis | Leukemia |
| DGN10 | C9230 | Myeloid sarcoma, not having achieved remission | Diagnosis | Leukemia |
| DGN10 | C9231 | Myeloid sarcoma, in remission | Diagnosis | Leukemia |
| DGN10 | C9240 | Acute promyelocytic leukemia, not having achieved remission | Diagnosis | Leukemia |
| DGN10 | C9241 | Acute promyelocytic leukemia, in remission | Diagnosis | Leukemia |
| DGN10 | C9250 | Acute myelomonocytic leukemia, not having achieved remission | Diagnosis | Leukemia |
| DGN10 | C9251 | Acute myelomonocytic leukemia, in remission | Diagnosis | Leukemia |
| DGN10 | C9260 | Acute myeloid leukemia with 11q23-abnormality not having achieved remission | Diagnosis | Leukemia |
| DGN10 | C9261 | Acute myeloid leukemia with 11q23-abnormality in remission | Diagnosis | Leukemia |
| DGN10 | C9290 | Myeloid leukemia, unspecified, not having achieved remission | Diagnosis | Leukemia |
| DGN10 | C9291 | Myeloid leukemia, unspecified in remission | Diagnosis | Leukemia |
| DGN10 | C92A0 | Acute myeloid leukemia with multilineage dysplasia, not having achieved remission | Diagnosis | Leukemia |
| DGN10 | C92A1 | Acute myeloid leukemia with multilineage dysplasia, in remission | Diagnosis | Leukemia |
| DGN10 | C92Z0 | Other myeloid leukemia not having achieved remission | Diagnosis | Leukemia |
| DGN10 | C92Z1 | Other myeloid leukemia, in remission | Diagnosis | Leukemia |
| DGN10 | C9300 | Acute monoblastic/monocytic leukemia, not having achieved remission | Diagnosis | Leukemia |
| DGN10 | C9301 | Acute monoblastic/monocytic leukemia, in remission | Diagnosis | Leukemia |
| DGN10 | C9310 | Chronic myelomonocytic leukemia not having achieved remission | Diagnosis | Leukemia |
| DGN10 | C9311 | Chronic myelomonocytic leukemia, in remission | Diagnosis | Leukemia |
| DGN10 | C9330 | Juvenile myelomonocytic leukemia, not having achieved remission | Diagnosis | Leukemia |
| DGN10 | C9331 | Juvenile myelomonocytic leukemia, in remission | Diagnosis | Leukemia |
| DGN10 | C9390 | Monocytic leukemia, unspecified, not having achieved remission | Diagnosis | Leukemia |
| DGN10 | C9391 | Monocytic leukemia, unspecified in remission | Diagnosis | Leukemia |
| DGN10 | C93Z0 | Other monocytic leukemia, not having achieved remission | Diagnosis | Leukemia |
| DGN10 | C93Z1 | Other monocytic leukemia, in remission | Diagnosis | Leukemia |
| DGN10 | C9400 | Acute erythroid leukemia, not having achieved remission | Diagnosis | Leukemia |
| DGN10 | C9401 | Acute erythroid leukemia, in remission | Diagnosis | Leukemia |
| DGN10 | C9420 | Acute megakaryoblastic leukemia not having achieved remission | Diagnosis | Leukemia |
| DGN10 | C9421 | Acute megakaryoblastic leukemia, in remission | Diagnosis | Leukemia |
| DGN10 | C9430 | Mast cell leukemia not having achieved remission | Diagnosis | Leukemia |
| DGN10 | C9431 | Mast cell leukemia, in remission | Diagnosis | Leukemia |
| DGN10 | C9480 | Other specified leukemias not having achieved remission | Diagnosis | Leukemia |
| DGN10 | C9481 | Other specified leukemias, in remission | Diagnosis | Leukemia |
| DGN10 | C9500 | Acute leukemia of unspecified cell type not having achieved remission | Diagnosis | Leukemia |
| DGN10 | C9501 | Acute leukemia of unspecified cell type, in remission | Diagnosis | Leukemia |
| DGN10 | C9510 | Chronic leukemia of unspecified cell type not having achieved remission | Diagnosis | Leukemia |
| DGN10 | C9511 | Chronic leukemia of unspecified cell type, in remission | Diagnosis | Leukemia |
| DGN10 | C9590 | Leukemia, unspecified not having achieved remission | Diagnosis | Leukemia |
| DGN10 | C9591 | Leukemia, unspecified, in remission | Diagnosis | Leukemia |
| DGN10 | C9302 | Acute monoblastic/monocytic leukemia, in relapse | Diagnosis | Leukemia |
| DGN10 | C9312 | Chronic myelomonocytic leukemia, in relapse | Diagnosis | Leukemia |
| DGN10 | C9392 | Monocytic leukemia, unspecified in relapse | Diagnosis | Leukemia |
| DGN10 | C93Z2 | Other monocytic leukemia, in relapse | Diagnosis | Leukemia |
| DGN10 | C9402 | Acute erythroid leukemia, in relapse | Diagnosis | Leukemia |
| DGN10 | C9482 | Other specified leukemias, in relapse | Diagnosis | Leukemia |
| DGN10 | C9502 | Acute leukemia of unspecified cell type, in relapse | Diagnosis | Leukemia |
| DGN10 | C9512 | Chronic leukemia of unspecified cell type, in relapse | Diagnosis | Leukemia |
| DGN10 | C9592 | Leukemia, unspecified, in relapse | Diagnosis | Leukemia |
| DGN10 | C9012 | Plasma cell leukemia in relapse | Diagnosis | Leukemia |
| DGN10 | C9102 | Acute lymphoblastic leukemia [ALL] in relapse | Diagnosis | Leukemia |
| DGN10 | C9112 | Chronic lymphocytic leukemia of B-cell type in relapse | Diagnosis | Leukemia |
| DGN10 | C9132 | Prolymphocytic leukemia of B-cell type, in relapse | Diagnosis | Leukemia |
| DGN10 | C9152 | Adult T-cell lymphoma/leukemia (HTLV-1-associated), in relapse | Diagnosis | Leukemia |
| DGN10 | C9162 | Prolymphocytic leukemia of T-cell type, in relapse | Diagnosis | Leukemia |
| DGN10 | C9192 | Lymphoid leukemia, unspecified in relapse | Diagnosis | Leukemia |
| DGN10 | C91Z2 | Other lymphoid leukemia, in relapse | Diagnosis | Leukemia |
| DGN10 | C9202 | Acute myeloblastic leukemia in relapse | Diagnosis | Leukemia |
| DGN10 | C9212 | Chronic myeloid leukemia, BCR/ABL-positive in relapse | Diagnosis | Leukemia |
| DGN10 | C9222 | Atypical chronic myeloid leukemia, BCR/ABL-negative in relapse | Diagnosis | Leukemia |
| DGN10 | C9232 | Myeloid sarcoma in relapse | Diagnosis | Leukemia |
| DGN10 | C9242 | Acute promyelocytic leukemia in relapse | Diagnosis | Leukemia |
| DGN10 | C9252 | Acute myelomonocytic leukemia in relapse | Diagnosis | Leukemia |
| DGN10 | C9262 | Acute myeloid leukemia with 11q23-abnormality in relapse | Diagnosis | Leukemia |
| DGN10 | C9292 | Myeloid leukemia, unspecified in relapse | Diagnosis | Leukemia |
| DGN10 | C92A2 | Acute myeloid leukemia with multilineage dysplasia, in relapse | Diagnosis | Leukemia |
| DGN10 | C92Z2 | Other myeloid leukemia in relapse | Diagnosis | Leukemia |
| DGN10 | C9332 | Juvenile myelomonocytic leukemia, in relapse | Diagnosis | Leukemia |
| DGN10 | C9422 | Acute megakaryoblastic leukemia, in relapse | Diagnosis | Leukemia |
| DGN10 | C9432 | Mast cell leukemia, in relapse | Diagnosis | Leukemia |
| DGN10 | C9000 | Multiple myeloma not having achieved remission | Diagnosis | Myeloma/Amyloid |
| DGN10 | C9001 | Multiple myeloma in remission | Diagnosis | Myeloma/Amyloid |
| DGN10 | C9002 | Multiple myeloma in relapse | Diagnosis | Myeloma/Amyloid |
| DGN10 | E851 | Neuropathic heredofamilial amyloidosis | Diagnosis | Myeloma/Amyloid |
| DGN10 | E852 | Heredofamilial amyloidosis, unspecified | Diagnosis | Myeloma/Amyloid |
| DGN10 | E853 | Secondary systemic amyloidosis | Diagnosis | Myeloma/Amyloid |
| DGN10 | E854 | Organ-limited amyloidosis | Diagnosis | Myeloma/Amyloid |
| DGN10 | E8581 | Light chain (AL) amyloidosis | Diagnosis | Myeloma/Amyloid |
| DGN10 | E8582 | Wild-type transthyretin-related (ATTR) amyloidosis | Diagnosis | Myeloma/Amyloid |
| DGN10 | E8589 | Other amyloidosis | Diagnosis | Myeloma/Amyloid |
| DGN10 | E859 | Amyloidosis, unspecified | Diagnosis | Myeloma/Amyloid |
| DGN10 | E850 | Non-neuropathic heredofamilial amyloidosis | Diagnosis | Myeloma/Amyloid |
| DGN10 | C9020 | Extramedullary plasmacytoma not having achieved remission | Diagnosis | Myeloma/Amyloid |
| DGN10 | C9022 | Extramedullary plasmacytoma in relapse | Diagnosis | Myeloma/Amyloid |
| DGN10 | C9030 | Solitary plasmacytoma not having achieved remission | Diagnosis | Myeloma/Amyloid |
| DGN10 | C9032 | Solitary plasmacytoma in relapse | Diagnosis | Myeloma/Amyloid |
| DGN10 | E858 | Other amyloidosis | Diagnosis | Myeloma/Amyloid |
| DGN10 | D808 | Other immunodeficiencies with predominantly antibody defects | Diagnosis | Immune Disorders |
| DGN10 | D809 | Immunodeficiency with predominantly antibody defects, unspecified | Diagnosis | Immune Disorders |
| DGN10 | D810 | Severe combined immunodeficiency [SCID] with reticular dysgenesis | Diagnosis | Immune Disorders |
| DGN10 | D811 | Severe combined immunodeficiency [SCID] with low T- and B-cell numbers | Diagnosis | Immune Disorders |
| DGN10 | D812 | Severe combined immunodeficiency [SCID] with low or normal B-cell numbers | Diagnosis | Immune Disorders |
| DGN10 | D816 | Major histocompatibility complex class I deficiency | Diagnosis | Immune Disorders |
| DGN10 | D817 | Major histocompatibility complex class II deficiency | Diagnosis | Immune Disorders |
| DGN10 | D8189 | Other combined immunodeficiencies | Diagnosis | Immune Disorders |
| DGN10 | D819 | Combined immunodeficiency, unspecified | Diagnosis | Immune Disorders |
| DGN10 | D821 | Di George's syndrome | Diagnosis | Immune Disorders |
| DGN10 | D830 | Common variable immunodeficiency with predominant abnormalities of B-cell numbers and function | Diagnosis | Immune Disorders |
| DGN10 | D831 | Common variable immunodeficiency with predominant immunoregulatory T-cell disorders | Diagnosis | Immune Disorders |
| DGN10 | D832 | Common variable immunodeficiency with autoantibodies to B- or T-cells | Diagnosis | Immune Disorders |
| DGN10 | D838 | Other common variable immunodeficiencies | Diagnosis | Immune Disorders |
| DGN10 | D839 | Common variable immunodeficiency, unspecified | Diagnosis | Immune Disorders |
| DGN10 | D848 | Other specified immunodeficiencies | Diagnosis | Immune Disorders |
| DGN10 | D849 | Immunodeficiency, unspecified | Diagnosis | Immune Disorders |
| DGN10 | D89810 | Acute graft-versus-host disease | Diagnosis | Immune Disorders |
| DGN10 | D89811 | Chronic graft-versus-host disease | Diagnosis | Immune Disorders |
| DGN10 | D89812 | Acute on chronic graft-versus-host disease | Diagnosis | Immune Disorders |
| DGN10 | D89813 | Graft-versus-host disease, unspecified | Diagnosis | Immune Disorders |
| DGN10 | D800 | Hereditary hypogammaglobulinemia | Diagnosis | Immune Disorders |
| DGN10 | D801 | Nonfamilial hypogammaglobulinemia | Diagnosis | Immune Disorders |
| DGN10 | D802 | Selective deficiency of immunoglobulin A [IgA] | Diagnosis | Immune Disorders |
| DGN10 | D803 | Selective deficiency of immunoglobulin G [IgG] subclasses | Diagnosis | Immune Disorders |
| DGN10 | D804 | Selective deficiency of immunoglobulin M [IgM] | Diagnosis | Immune Disorders |
| DGN10 | D805 | Immunodeficiency with increased immunoglobulin M [IgM] | Diagnosis | Immune Disorders |
| DGN10 | D611 | Drug-induced aplastic anemia | Diagnosis | Immune Disorders |
| DGN10 | D612 | Aplastic anemia due to other external agents | Diagnosis | Immune Disorders |
| DGN10 | D613 | Idiopathic aplastic anemia | Diagnosis | Immune Disorders |
| DGN10 | D6189 | Other specified aplastic anemias and other bone marrow failure syndromes | Diagnosis | Immune Disorders |
| DGN10 | D700 | Congenital agranulocytosis | Diagnosis | Immune Disorders |
| DGN10 | D704 | Cyclic neutropenia | Diagnosis | Immune Disorders |
| DGN10 | D71 | Functional disorders of polymorphonuclear neutrophils | Diagnosis | Immune Disorders |
| DGN10 | D720 | Genetic anomalies of leukocytes | Diagnosis | Immune Disorders |
| DGN10 | D814 | Nezelof's syndrome | Diagnosis | Immune Disorders |
| DGN10 | D820 | Wiskott-Aldrich syndrome | Diagnosis | Immune Disorders |
| DGN10 | D600 | Chronic acquired pure red cell aplasia | Diagnosis | Immune Disorders |
| DGN10 | D601 | Transient acquired pure red cell aplasia | Diagnosis | Immune Disorders |
| DGN10 | D608 | Other acquired pure red cell aplasias | Diagnosis | Immune Disorders |
| DGN10 | D609 | Acquired pure red cell aplasia, unspecified | Diagnosis | Immune Disorders |
| DGN10 | D6101 | Constitutional (pure) red blood cell aplasia | Diagnosis | Immune Disorders |
| DGN10 | D619 | Aplastic anemia, unspecified | Diagnosis | Immune Disorders |
| DGN10 | D6481 | Anemia due to antineoplastic chemotherapy | Diagnosis | Immune Disorders |
| DGN10 | E803 | Defects of catalase and peroxidase | Diagnosis | Immune Disorders |
| DGN10 | [D61810](https://www.icd10data.com/ICD10CM/Codes/D50-D89/D60-D64/D61-/D61.810) | Antineoplastic chemotherapy induced pancytopenia | Diagnosis | Immune Disorders |
| DGN10 | [T451X1](https://www.icd10data.com/ICD10CM/Codes/S00-T88/T36-T50/T45-/T45.1X1) | Poisoning by antineoplastic and immunosuppressive drugs, accidental (unintentional) | Diagnosis | Immune Disorders |
| DGN10 | [T451X1A](https://www.icd10data.com/ICD10CM/Codes/S00-T88/T36-T50/T45-/T45.1X1A) | Poisoning by antineoplastic and immunosuppressive drugs, accidental (unintentional) initial encounter | Diagnosis | Immune Disorders |
| DGN10 | [T451X1D](https://www.icd10data.com/ICD10CM/Codes/S00-T88/T36-T50/T45-/T45.1X1D) | Poisoning by antineoplastic and immunosuppressive drugs, accidental (unintentional) subsequent encounter | Diagnosis | Immune Disorders |
| DGN10 | [T451X1S](https://www.icd10data.com/ICD10CM/Codes/S00-T88/T36-T50/T45-/T45.1X1S) | Poisoning by antineoplastic and immunosuppressive drugs, accidental (unintentional) sequela | Diagnosis | Immune Disorders |
| DGN10 | [T451X2](https://www.icd10data.com/ICD10CM/Codes/S00-T88/T36-T50/T45-/T45.1X2) | Poisoning by antineoplastic and immunosuppressive drugs, intentional self-harm | Diagnosis | Immune Disorders |
| DGN10 | [T451X2A](https://www.icd10data.com/ICD10CM/Codes/S00-T88/T36-T50/T45-/T45.1X2A) | Poisoning by antineoplastic and immunosuppressive drugs, intentional self-harm initial encounter | Diagnosis | Immune Disorders |
| DGN10 | [T451X2D](https://www.icd10data.com/ICD10CM/Codes/S00-T88/T36-T50/T45-/T45.1X2D) | Poisoning by antineoplastic and immunosuppressive drugs, intentional self-harm subsequent encounter | Diagnosis | Immune Disorders |
| DGN10 | [T451X2S](https://www.icd10data.com/ICD10CM/Codes/S00-T88/T36-T50/T45-/T45.1X2S) | Poisoning by antineoplastic and immunosuppressive drugs, intentional self-harm sequela | Diagnosis | Immune Disorders |
| DGN10 | [T451X3](https://www.icd10data.com/ICD10CM/Codes/S00-T88/T36-T50/T45-/T45.1X3) | Poisoning by antineoplastic and immunosuppressive drugs, assault | Diagnosis | Immune Disorders |
| DGN10 | [T451X3A](https://www.icd10data.com/ICD10CM/Codes/S00-T88/T36-T50/T45-/T45.1X3A) | Poisoning by antineoplastic and immunosuppressive drugs, assault initial encounter | Diagnosis | Immune Disorders |
| DGN10 | [T451X3D](https://www.icd10data.com/ICD10CM/Codes/S00-T88/T36-T50/T45-/T45.1X3D) | Poisoning by antineoplastic and immunosuppressive drugs, assault subsequent encounter | Diagnosis | Immune Disorders |
| DGN10 | [T451X3S](https://www.icd10data.com/ICD10CM/Codes/S00-T88/T36-T50/T45-/T45.1X3S) | Poisoning by antineoplastic and immunosuppressive drugs, assault sequela | Diagnosis | Immune Disorders |
| DGN10 | [T451X4](https://www.icd10data.com/ICD10CM/Codes/S00-T88/T36-T50/T45-/T45.1X4) | Poisoning by antineoplastic and immunosuppressive drugs, undetermined | Diagnosis | Immune Disorders |
| DGN10 | [T451X4A](https://www.icd10data.com/ICD10CM/Codes/S00-T88/T36-T50/T45-/T45.1X4A) | Poisoning by antineoplastic and immunosuppressive drugs, undetermined initial encounter | Diagnosis | Immune Disorders |
| DGN10 | [T451X4D](https://www.icd10data.com/ICD10CM/Codes/S00-T88/T36-T50/T45-/T45.1X4D) | Poisoning by antineoplastic and immunosuppressive drugs, undetermined subsequent encounter | Diagnosis | Immune Disorders |
| DGN10 | [T451X4S](https://www.icd10data.com/ICD10CM/Codes/S00-T88/T36-T50/T45-/T45.1X4S) | Poisoning by antineoplastic and immunosuppressive drugs, undetermined sequela | Diagnosis | Immune Disorders |
| DGN10 | [T451X5](https://www.icd10data.com/ICD10CM/Codes/S00-T88/T36-T50/T45-/T45.1X5) | Adverse effect of antineoplastic and immunosuppressive drugs | Diagnosis | Immune Disorders |
| DGN10 | [T451X5A](https://www.icd10data.com/ICD10CM/Codes/S00-T88/T36-T50/T45-/T45.1X5A) | Adverse effect of antineoplastic and immunosuppressive drugs initial encounter | Diagnosis | Immune Disorders |
| DGN10 | [T451X5D](https://www.icd10data.com/ICD10CM/Codes/S00-T88/T36-T50/T45-/T45.1X5D) | Adverse effect of antineoplastic and immunosuppressive drugs subsequent encounter | Diagnosis | Immune Disorders |
| DGN10 | [T451X5S](https://www.icd10data.com/ICD10CM/Codes/S00-T88/T36-T50/T45-/T45.1X5S) | Adverse effect of antineoplastic and immunosuppressive drugs sequela | Diagnosis | Immune Disorders |
| DGN10 | D701 | Agranulocytosis secondary to cancer chemotherapy | Diagnosis | Immune Disorders |
| DGN10 | D813 | Adenosine deaminase [ADA] deficiency | Diagnosis | Immune Disorders |
| DGN10 | D815 | Purine nucleoside phosphorylase [PNP] deficiency | Diagnosis | Immune Disorders |
| DGN10 | D822 | Immunodeficiency with short-limbed stature | Diagnosis | Immune Disorders |
| DGN10 | D823 | Immunodeficiency following hereditary defective response to Epstein-Barr virus | Diagnosis | Immune Disorders |
| DGN10 | D824 | Hyperimmunoglobulin E [IgE] syndrome | Diagnosis | Immune Disorders |
| DGN10 | D828 | Immunodeficiency associated with other specified major defects | Diagnosis | Immune Disorders |
| DGN10 | D829 | Immunodeficiency associated with major defect, unspecified | Diagnosis | Immune Disorders |
| DGN10 | D840 | Lymphocyte function antigen-1 [LFA-1] defect | Diagnosis | Immune Disorders |
| DGN10 | D841 | Defects in the complement system | Diagnosis | Immune Disorders |
| DGN10 | D8989 | Other specified disorders involving the immune mechanism, not elsewhere classified | Diagnosis | Immune Disorders |
| DGN10 | D899 | Disorder involving the immune mechanism, unspecified | Diagnosis | Immune Disorders |
| DGN10 | D806 | Antibody deficiency with near-normal immunoglobulins or with hyperimmunoglobulinemia | Diagnosis | Immune Disorders |
| HCPCS | 32851 | Lung transplant, single; without cardiopulmonary bypass | Diagnosis | Transplant/Dialysis |
| HCPCS | 32852 | Lung transplant, single; with cardiopulmonary bypass | Diagnosis | Transplant/Dialysis |
| HCPCS | 32853 | Lung transplant, double (bilateral sequential or en bloc); without cardiopulmonary bypass | Diagnosis | Transplant/Dialysis |
| HCPCS | 32854 | Lung transplant, double (bilateral sequential or en bloc); with cardiopulmonary bypass | Diagnosis | Transplant/Dialysis |
| HCPCS | 33935 | Heart-lung transplant with recipient cardiectomy-pneumonectomy | Diagnosis | Transplant/Dialysis |
| HCPCS | 33945 | Heart transplant, with or without recipient cardiectomy | Diagnosis | Transplant/Dialysis |
| HCPCS | 47135 | Liver allotransplantation; orthotopic, partial or whole, from cadaver or living donor, any age | Diagnosis | Transplant/Dialysis |
| HCPCS | 48554 | Transplantation of pancreatic allograft | Diagnosis | Transplant/Dialysis |
| HCPCS | 50360 | Renal allotransplantation, implantation of graft; without recipient nephrectomy | Diagnosis | Transplant/Dialysis |
| HCPCS | 50365 | Renal allotransplantation, implantation of graft; with recipient nephrectomy | Diagnosis | Transplant/Dialysis |
| HCPCS | 50366 | Renal Homotransplantation, Implantation Of Graft | Diagnosis | Transplant/Dialysis |
| HCPCS | 50370 | Removal of transplanted renal allograft | Diagnosis | Transplant/Dialysis |
| HCPCS | 50380 | Renal autotransplantation, reimplantation of kidney | Diagnosis | Transplant/Dialysis |
| HCPCS | S2053 | Transplantation of small intestine and liver allografts | Diagnosis | Transplant/Dialysis |
| HCPCS | S2054 | Transplantation of multivisceral organs | Diagnosis | Transplant/Dialysis |
| HCPCS | S2060 | Lobar lung transplantation | Diagnosis | Transplant/Dialysis |
| HCPCS | S2065 | Simultaneous pancreas kidney transplantation | Diagnosis | Transplant/Dialysis |
| HCPCS | S2142 | Cord blood-derived stem-cell transplantation, allogeneic | Diagnosis | Transplant/Dialysis |
| PRC10 | 02YA0Z0 | Transplantation of Heart, Allogeneic, Open Approach | Diagnosis | Transplant/Dialysis |
| PRC10 | 02YA0Z1 | Transplantation of Heart, Syngeneic, Open Approach | Diagnosis | Transplant/Dialysis |
| PRC10 | 02YA0Z2 | Transplantation of Heart, Zooplastic, Open Approach | Diagnosis | Transplant/Dialysis |
| PRC10 | 0BYM0Z0 | Transplantation of Bilateral Lungs, Allogeneic, Open Approach | Diagnosis | Transplant/Dialysis |
| PRC10 | 0BYM0Z1 | Transplantation of Bilateral Lungs, Syngeneic, Open Approach | Diagnosis | Transplant/Dialysis |
| PRC10 | 0BYM0Z2 | Transplantation of Bilateral Lungs, Zooplastic, Open Approach | Diagnosis | Transplant/Dialysis |
| PRC10 | 0FY00Z0 | Transplantation of Liver, Allogeneic, Open Approach | Diagnosis | Transplant/Dialysis |
| PRC10 | 0FY00Z1 | Transplantation of Liver, Syngeneic, Open Approach | Diagnosis | Transplant/Dialysis |
| PRC10 | 0FY00Z2 | Transplantation of Liver, Zooplastic, Open Approach | Diagnosis | Transplant/Dialysis |
| PRC10 | 0TY00Z0 | Transplantation of Right Kidney, Allogeneic, Open Approach | Diagnosis | Transplant/Dialysis |
| PRC10 | 0TY00Z1 | Transplantation of Right Kidney, Syngeneic, Open Approach | Diagnosis | Transplant/Dialysis |
| PRC10 | 0TY00Z2 | Transplantation of Right Kidney, Zooplastic, Open Approach | Diagnosis | Transplant/Dialysis |
| PRC10 | 0TY10Z0 | Transplantation of Left Kidney, Allogeneic, Open Approach | Diagnosis | Transplant/Dialysis |
| PRC10 | 0TY10Z1 | Transplantation of Left Kidney, Syngeneic, Open Approach | Diagnosis | Transplant/Dialysis |
| PRC10 | 0TY10Z2 | Transplantation of Left Kidney, Zooplastic, Open Approach | Diagnosis | Transplant/Dialysis |
| DGN10 | T8610 | Unspecified complication of kidney transplant | Diagnosis | Transplant/Dialysis |
| DGN10 | T8611 | Kidney transplant rejection | Diagnosis | Transplant/Dialysis |
| DGN10 | T8612 | Kidney transplant failure | Diagnosis | Transplant/Dialysis |
| DGN10 | T8613 | Kidney transplant infection | Diagnosis | Transplant/Dialysis |
| DGN10 | T8619 | Other complication of kidney transplant | Diagnosis | Transplant/Dialysis |
| DGN10 | T8620 | Unspecified complication of heart transplant | Diagnosis | Transplant/Dialysis |
| DGN10 | T8621 | Heart transplant rejection | Diagnosis | Transplant/Dialysis |
| DGN10 | T8622 | Heart transplant failure | Diagnosis | Transplant/Dialysis |
| DGN10 | T8623 | Heart transplant infection | Diagnosis | Transplant/Dialysis |
| DGN10 | T86290 | Cardiac allograft vasculopathy | Diagnosis | Transplant/Dialysis |
| DGN10 | T86298 | Other complications of heart transplant | Diagnosis | Transplant/Dialysis |
| DGN10 | T8630 | Unspecified complication of heart-lung transplant | Diagnosis | Transplant/Dialysis |
| DGN10 | T8631 | Heart-lung transplant rejection | Diagnosis | Transplant/Dialysis |
| DGN10 | T8632 | Heart-lung transplant failure | Diagnosis | Transplant/Dialysis |
| DGN10 | T8633 | Heart-lung transplant infection | Diagnosis | Transplant/Dialysis |
| DGN10 | T8639 | Other complications of heart-lung transplant | Diagnosis | Transplant/Dialysis |
| DGN10 | T8640 | Unspecified complication of liver transplant | Diagnosis | Transplant/Dialysis |
| DGN10 | T8641 | Liver transplant rejection | Diagnosis | Transplant/Dialysis |
| DGN10 | T8642 | Liver transplant failure | Diagnosis | Transplant/Dialysis |
| DGN10 | T8643 | Liver transplant infection | Diagnosis | Transplant/Dialysis |
| DGN10 | T8649 | Other complications of liver transplant | Diagnosis | Transplant/Dialysis |
| DGN10 | Z4821 | Encounter for aftercare following heart transplant | Diagnosis | Transplant/Dialysis |
| DGN10 | Z4822 | Encounter for aftercare following kidney transplant | Diagnosis | Transplant/Dialysis |
| DGN10 | Z4823 | Encounter for aftercare following liver transplant | Diagnosis | Transplant/Dialysis |
| DGN10 | Z4824 | Encounter for aftercare following lung transplant | Diagnosis | Transplant/Dialysis |
| DGN10 | Z48280 | Encounter for aftercare following heart-lung transplant | Diagnosis | Transplant/Dialysis |
| DGN10 | Z48290 | Encounter for aftercare following bone marrow transplant | Diagnosis | Transplant/Dialysis |
| DGN10 | Z940 | Kidney transplant status | Diagnosis | Transplant/Dialysis |
| DGN10 | Z941 | Heart transplant status | Diagnosis | Transplant/Dialysis |
| DGN10 | Z942 | Lung transplant status | Diagnosis | Transplant/Dialysis |
| DGN10 | Z943 | Heart and lungs transplant status | Diagnosis | Transplant/Dialysis |
| DGN10 | Z944 | Liver transplant status | Diagnosis | Transplant/Dialysis |
| DGN10 | Z9481 | Bone marrow transplant status | Diagnosis | Transplant/Dialysis |
| DGN10 | Z9482 | Intestine transplant status | Diagnosis | Transplant/Dialysis |
| DGN10 | Z9483 | Pancreas transplant status | Diagnosis | Transplant/Dialysis |
| DGN10 | Z9484 | Stem cells transplant status | Diagnosis | Transplant/Dialysis |
| HCPCS | 38240 | Bone marrow or blood-derived peripheral stem cell transplantation; allogenic | Diagnosis | Transplant/Dialysis |
| HCPCS | 38241 | Bone marrow or blood-derived peripheral stem cell transplantation; autologous | Diagnosis | Transplant/Dialysis |
| HCPCS | 38242 | Bone marrow or blood-derived peripheral stem cell transplantation; allogeneic donor lymphocyte infusions | Diagnosis | Transplant/Dialysis |
| HCPCS | 44135 | Intestinal allotransplantation; from cadaver donor | Diagnosis | Transplant/Dialysis |
| HCPCS | 44136 | Intestinal allotransplantation; from living donor | Diagnosis | Transplant/Dialysis |
| HCPCS | 47136 | Liver allotransplantation; heterotopic, partial or whole, from cadaver or living donor, any age | Diagnosis | Transplant/Dialysis |
| PRC10 | 07YP0Z0 | Transplantation of Spleen, Allogeneic, Open Approach | Diagnosis | Transplant/Dialysis |
| PRC10 | 07YP0Z1 | Transplantation of Spleen, Syngeneic, Open Approach | Diagnosis | Transplant/Dialysis |
| PRC10 | 07YP0Z2 | Transplantation of Spleen, Zooplastic, Open Approach | Diagnosis | Transplant/Dialysis |
| PRC10 | 0BYC0Z0 | Transplantation of Right Upper Lung Lobe, Allogeneic, Open Approach | Diagnosis | Transplant/Dialysis |
| PRC10 | 0BYC0Z1 | Transplantation of Right Upper Lung Lobe, Syngeneic, Open Approach | Diagnosis | Transplant/Dialysis |
| PRC10 | 0BYC0Z2 | Transplantation of Right Upper Lung Lobe, Zooplastic, Open Approach | Diagnosis | Transplant/Dialysis |
| PRC10 | 0BYD0Z0 | Transplantation of Right Middle Lung Lobe, Allogeneic, Open Approach | Diagnosis | Transplant/Dialysis |
| PRC10 | 0BYD0Z1 | Transplantation of Right Middle Lung Lobe, Syngeneic, Open Approach | Diagnosis | Transplant/Dialysis |
| PRC10 | 0BYD0Z2 | Transplantation of Right Middle Lung Lobe, Zooplastic, Open Approach | Diagnosis | Transplant/Dialysis |
| PRC10 | 0BYF0Z0 | Transplantation of Right Lower Lung Lobe, Allogeneic, Open Approach | Diagnosis | Transplant/Dialysis |
| PRC10 | 0BYF0Z1 | Transplantation of Right Lower Lung Lobe, Syngeneic, Open Approach | Diagnosis | Transplant/Dialysis |
| PRC10 | 0BYF0Z2 | Transplantation of Right Lower Lung Lobe, Zooplastic, Open Approach | Diagnosis | Transplant/Dialysis |
| PRC10 | 0BYG0Z0 | Transplantation of Left Upper Lung Lobe, Allogeneic, Open Approach | Diagnosis | Transplant/Dialysis |
| PRC10 | 0BYG0Z1 | Transplantation of Left Upper Lung Lobe, Syngeneic, Open Approach | Diagnosis | Transplant/Dialysis |
| PRC10 | 0BYG0Z2 | Transplantation of Left Upper Lung Lobe, Zooplastic, Open Approach | Diagnosis | Transplant/Dialysis |
| PRC10 | 0BYH0Z0 | Transplantation of Lung Lingula, Allogeneic, Open Approach | Diagnosis | Transplant/Dialysis |
| PRC10 | 0BYH0Z1 | Transplantation of Lung Lingula, Syngeneic, Open Approach | Diagnosis | Transplant/Dialysis |
| PRC10 | 0BYH0Z2 | Transplantation of Lung Lingula, Zooplastic, Open Approach | Diagnosis | Transplant/Dialysis |
| PRC10 | 0BYJ0Z0 | Transplantation of Left Lower Lung Lobe, Allogeneic, Open Approach | Diagnosis | Transplant/Dialysis |
| PRC10 | 0BYJ0Z1 | Transplantation of Left Lower Lung Lobe, Syngeneic, Open Approach | Diagnosis | Transplant/Dialysis |
| PRC10 | 0BYJ0Z2 | Transplantation of Left Lower Lung Lobe, Zooplastic, Open Approach | Diagnosis | Transplant/Dialysis |
| PRC10 | 0BYK0Z0 | Transplantation of Right Lung, Allogeneic, Open Approach | Diagnosis | Transplant/Dialysis |
| PRC10 | 0BYK0Z1 | Transplantation of Right Lung, Syngeneic, Open Approach | Diagnosis | Transplant/Dialysis |
| PRC10 | 0BYK0Z2 | Transplantation of Right Lung, Zooplastic, Open Approach | Diagnosis | Transplant/Dialysis |
| PRC10 | 0BYL0Z0 | Transplantation of Left Lung, Allogeneic, Open Approach | Diagnosis | Transplant/Dialysis |
| PRC10 | 0BYL0Z1 | Transplantation of Left Lung, Syngeneic, Open Approach | Diagnosis | Transplant/Dialysis |
| PRC10 | 0BYL0Z2 | Transplantation of Left Lung, Zooplastic, Open Approach | Diagnosis | Transplant/Dialysis |
| PRC10 | 0DY80Z0 | Transplantation of Small Intestine, Allogeneic, Open Approach | Diagnosis | Transplant/Dialysis |
| PRC10 | 0DY80Z1 | Transplantation of Small Intestine, Syngeneic, Open Approach | Diagnosis | Transplant/Dialysis |
| PRC10 | 0DY80Z2 | Transplantation of Small Intestine, Zooplastic, Open Approach | Diagnosis | Transplant/Dialysis |
| PRC10 | 0DYE0Z0 | Transplantation of Large Intestine, Allogeneic, Open Approach | Diagnosis | Transplant/Dialysis |
| PRC10 | 0DYE0Z1 | Transplantation of Large Intestine, Syngeneic, Open Approach | Diagnosis | Transplant/Dialysis |
| PRC10 | 0DYE0Z2 | Transplantation of Large Intestine, Zooplastic, Open Approach | Diagnosis | Transplant/Dialysis |
| PRC10 | 0FYG0Z0 | Transplantation of Pancreas, Allogeneic, Open Approach | Diagnosis | Transplant/Dialysis |
| PRC10 | 0FYG0Z1 | Transplantation of Pancreas, Syngeneic, Open Approach | Diagnosis | Transplant/Dialysis |
| PRC10 | 0FYG0Z2 | Transplantation of Pancreas, Zooplastic, Open Approach | Diagnosis | Transplant/Dialysis |
| PRC10 | 30230AZ | Transfusion of Embryonic Stem Cells into Peripheral Vein, Open Approach | Diagnosis | Transplant/Dialysis |
| PRC10 | 30230G0 | Transfusion of Autologous Bone Marrow into Peripheral Vein, Open Approach | Diagnosis | Transplant/Dialysis |
| PRC10 | 30230G1 | Transfusion of Nonautologous Bone Marrow into Peripheral Vein, Open Approach | Diagnosis | Transplant/Dialysis |
| PRC10 | 30230X0 | Transfusion of Autologous Cord Blood Stem Cells into Peripheral Vein, Open Approach | Diagnosis | Transplant/Dialysis |
| PRC10 | 30230X1 | Transfusion of Nonautologous Cord Blood Stem Cells into Peripheral Vein, Open Approach | Diagnosis | Transplant/Dialysis |
| PRC10 | 30230Y0 | Transfusion of Autologous Hematopoietic Stem Cells into Peripheral Vein, Open Approach | Diagnosis | Transplant/Dialysis |
| PRC10 | 30230Y1 | Transfusion of Nonautologous Hematopoietic Stem Cells into Peripheral Vein, Open Approach | Diagnosis | Transplant/Dialysis |
| PRC10 | 30233AZ | Transfusion of Embryonic Stem Cells into Peripheral Vein, Percutaneous Approach | Diagnosis | Transplant/Dialysis |
| PRC10 | 30233G0 | Transfusion of Autologous Bone Marrow into Peripheral Vein, Percutaneous Approach | Diagnosis | Transplant/Dialysis |
| PRC10 | 30233G1 | Transfusion of Nonautologous Bone Marrow into Peripheral Vein, Percutaneous Approach | Diagnosis | Transplant/Dialysis |
| PRC10 | 30233X0 | Transfusion of Autologous Cord Blood Stem Cells into Peripheral Vein, Percutaneous Approach | Diagnosis | Transplant/Dialysis |
| PRC10 | 30233X1 | Transfusion of Nonautologous Cord Blood Stem Cells into Peripheral Vein, Percutaneous Approach | Diagnosis | Transplant/Dialysis |
| PRC10 | 30233Y0 | Transfusion of Autologous Hematopoietic Stem Cells into Peripheral Vein, Percutaneous Approach | Diagnosis | Transplant/Dialysis |
| PRC10 | 30233Y1 | Transfusion of Nonautologous Hematopoietic Stem Cells into Peripheral Vein, Percutaneous Approach | Diagnosis | Transplant/Dialysis |
| PRC10 | 30240AZ | Transfusion of Embryonic Stem Cells into Central Vein, Open Approach | Diagnosis | Transplant/Dialysis |
| PRC10 | 30240G0 | Transfusion of Autologous Bone Marrow into Central Vein, Open Approach | Diagnosis | Transplant/Dialysis |
| PRC10 | 30240G1 | Transfusion of Nonautologous Bone Marrow into Central Vein, Open Approach | Diagnosis | Transplant/Dialysis |
| PRC10 | 30240X0 | Transfusion of Autologous Cord Blood Stem Cells into Central Vein, Open Approach | Diagnosis | Transplant/Dialysis |
| PRC10 | 30240X1 | Transfusion of Nonautologous Cord Blood Stem Cells into Central Vein, Open Approach | Diagnosis | Transplant/Dialysis |
| PRC10 | 30240Y0 | Transfusion of Autologous Hematopoietic Stem Cells into Central Vein, Open Approach | Diagnosis | Transplant/Dialysis |
| PRC10 | 30240Y1 | Transfusion of Nonautologous Hematopoietic Stem Cells into Central Vein, Open Approach | Diagnosis | Transplant/Dialysis |
| PRC10 | 30243AZ | Transfusion of Embryonic Stem Cells into Central Vein, Percutaneous Approach | Diagnosis | Transplant/Dialysis |
| PRC10 | 30243G0 | Transfusion of Autologous Bone Marrow into Central Vein, Percutaneous Approach | Diagnosis | Transplant/Dialysis |
| PRC10 | 30243G1 | Transfusion of Nonautologous Bone Marrow into Central Vein, Percutaneous Approach | Diagnosis | Transplant/Dialysis |
| PRC10 | 30243X0 | Transfusion of Autologous Cord Blood Stem Cells into Central Vein, Percutaneous Approach | Diagnosis | Transplant/Dialysis |
| PRC10 | 30243X1 | Transfusion of Nonautologous Cord Blood Stem Cells into Central Vein, Percutaneous Approach | Diagnosis | Transplant/Dialysis |
| PRC10 | 30243Y0 | Transfusion of Autologous Hematopoietic Stem Cells into Central Vein, Percutaneous Approach | Diagnosis | Transplant/Dialysis |
| PRC10 | 30243Y1 | Transfusion of Nonautologous Hematopoietic Stem Cells into Central Vein, Percutaneous Approach | Diagnosis | Transplant/Dialysis |
| PRC10 | 30250G0 | Transfusion of Autologous Bone Marrow into Peripheral Artery, Open Approach | Diagnosis | Transplant/Dialysis |
| PRC10 | 30250G1 | Transfusion of Nonautologous Bone Marrow into Peripheral Artery, Open Approach | Diagnosis | Transplant/Dialysis |
| PRC10 | 30250X0 | Transfusion of Autologous Cord Blood Stem Cells into Peripheral Artery, Open Approach | Diagnosis | Transplant/Dialysis |
| PRC10 | 30250X1 | Transfusion of Nonautologous Cord Blood Stem Cells into Peripheral Artery, Open Approach | Diagnosis | Transplant/Dialysis |
| PRC10 | 30250Y0 | Transfusion of Autologous Hematopoietic Stem Cells into Peripheral Artery, Open Approach | Diagnosis | Transplant/Dialysis |
| PRC10 | 30250Y1 | Transfusion of Nonautologous Hematopoietic Stem Cells into Peripheral Artery, Open Approach | Diagnosis | Transplant/Dialysis |
| PRC10 | 30253G0 | Transfusion of Autologous Bone Marrow into Peripheral Artery, Percutaneous Approach | Diagnosis | Transplant/Dialysis |
| PRC10 | 30253G1 | Transfusion of Nonautologous Bone Marrow into Peripheral Artery, Percutaneous Approach | Diagnosis | Transplant/Dialysis |
| PRC10 | 30253X0 | Transfusion of Autologous Cord Blood Stem Cells into Peripheral Artery, Percutaneous Approach | Diagnosis | Transplant/Dialysis |
| PRC10 | 30253X1 | Transfusion of Nonautologous Cord Blood Stem Cells into Peripheral Artery, Percutaneous Approach | Diagnosis | Transplant/Dialysis |
| PRC10 | 30253Y0 | Transfusion of Autologous Hematopoietic Stem Cells into Peripheral Artery, Percutaneous Approach | Diagnosis | Transplant/Dialysis |
| PRC10 | 30253Y1 | Transfusion of Nonautologous Hematopoietic Stem Cells into Peripheral Artery, Percutaneous Approach | Diagnosis | Transplant/Dialysis |
| PRC10 | 30260G0 | Transfusion of Autologous Bone Marrow into Central Artery, Open Approach | Diagnosis | Transplant/Dialysis |
| PRC10 | 30260G1 | Transfusion of Nonautologous Bone Marrow into Central Artery, Open Approach | Diagnosis | Transplant/Dialysis |
| PRC10 | 30260X0 | Transfusion of Autologous Cord Blood Stem Cells into Central Artery, Open Approach | Diagnosis | Transplant/Dialysis |
| PRC10 | 30260X1 | Transfusion of Nonautologous Cord Blood Stem Cells into Central Artery, Open Approach | Diagnosis | Transplant/Dialysis |
| PRC10 | 30260Y0 | Transfusion of Autologous Hematopoietic Stem Cells into Central Artery, Open Approach | Diagnosis | Transplant/Dialysis |
| PRC10 | 30260Y1 | Transfusion of Nonautologous Hematopoietic Stem Cells into Central Artery, Open Approach | Diagnosis | Transplant/Dialysis |
| PRC10 | 30263G0 | Transfusion of Autologous Bone Marrow into Central Artery, Percutaneous Approach | Diagnosis | Transplant/Dialysis |
| PRC10 | 30263G1 | Transfusion of Nonautologous Bone Marrow into Central Artery, Percutaneous Approach | Diagnosis | Transplant/Dialysis |
| PRC10 | 30263X0 | Transfusion of Autologous Cord Blood Stem Cells into Central Artery, Percutaneous Approach | Diagnosis | Transplant/Dialysis |
| PRC10 | 30263X1 | Transfusion of Nonautologous Cord Blood Stem Cells into Central Artery, Percutaneous Approach | Diagnosis | Transplant/Dialysis |
| PRC10 | 30263Y0 | Transfusion of Autologous Hematopoietic Stem Cells into Central Artery, Percutaneous Approach | Diagnosis | Transplant/Dialysis |
| PRC10 | 30263Y1 | Transfusion of Nonautologous Hematopoietic Stem Cells into Central Artery, Percutaneous Approach | Diagnosis | Transplant/Dialysis |
| PRC10 | 3E030U1 | Introduction of Nonautologous Pancreatic Islet Cells into Peripheral Vein, Open Approach | Diagnosis | Transplant/Dialysis |
| PRC10 | 3E033U1 | Introduction of Nonautologous Pancreatic Islet Cells into Peripheral Vein, Percutaneous Approach | Diagnosis | Transplant/Dialysis |
| PRC10 | 3E0J3U1 | Introduction of Nonautologous Pancreatic Islet Cells into Biliary and Pancreatic Tract, Percutaneous Approach | Diagnosis | Transplant/Dialysis |
| PRC10 | 3E0J7U1 | Introduction of Nonautologous Pancreatic Islet Cells into Biliary and Pancreatic Tract, Via Natural or Artificial Opening | Diagnosis | Transplant/Dialysis |
| PRC10 | 3E0J8U1 | Introduction of Nonautologous Pancreatic Islet Cells into Biliary and Pancreatic Tract, Via Natural or Artificial Opening Endoscopic | Diagnosis | Transplant/Dialysis |
| HCPCS | S2102 | Islet cell tissue transplant from pancreas; allogeneic | Diagnosis | Transplant/Dialysis |
| HCPCS | S2150 | Bone marrow or blood-derived stem cells (peripheral or umbilical), allogeneic or autologous, harvesting, transplantation, and related complications including pheresis and cell preparation/storage | Diagnosis | Transplant/Dialysis |
| HCPCS | S2152 | Solid organ(s), complete or segmental, single organ or combination of organs; deceased or living donor (s), procurement, transplantation, and related complications | Diagnosis | Transplant/Dialysis |
| DGN10 | Z992 | Dependence on renal dialysis | Diagnosis | Transplant/Dialysis |
| DGN10 | I25750 | Atherosclerosis of native coronary artery of transplanted heart with unstable angina | Diagnosis | Transplant/Dialysis |
| DGN10 | I25751 | Atherosclerosis of native coronary artery of transplanted heart with angina pectoris with documented spasm | Diagnosis | Transplant/Dialysis |
| DGN10 | I25758 | Atherosclerosis of native coronary artery of transplanted heart with other forms of angina pectoris | Diagnosis | Transplant/Dialysis |
| DGN10 | I25759 | Atherosclerosis of native coronary artery of transplanted heart with unspecified angina pectoris | Diagnosis | Transplant/Dialysis |
| DGN10 | I25760 | Atherosclerosis of bypass graft of coronary artery of transplanted heart with unstable angina | Diagnosis | Transplant/Dialysis |
| DGN10 | I25761 | Atherosclerosis of bypass graft of coronary artery of transplanted heart with angina pectoris with documented spasm | Diagnosis | Transplant/Dialysis |
| DGN10 | I25768 | Atherosclerosis of bypass graft of coronary artery of transplanted heart with other forms of angina pectoris | Diagnosis | Transplant/Dialysis |
| DGN10 | I25769 | Atherosclerosis of bypass graft of coronary artery of transplanted heart with unspecified angina pectoris | Diagnosis | Transplant/Dialysis |
| DGN10 | I25811 | Atherosclerosis of native coronary artery of transplanted heart without angina pectoris | Diagnosis | Transplant/Dialysis |
| DGN10 | I25812 | Atherosclerosis of bypass graft of coronary artery of transplanted heart without angina pectoris | Diagnosis | Transplant/Dialysis |
| DGN10 | T8600 | Unspecified complication of bone marrow transplant | Diagnosis | Transplant/Dialysis |
| DGN10 | T8601 | Bone marrow transplant rejection | Diagnosis | Transplant/Dialysis |
| DGN10 | T8602 | Bone marrow transplant failure | Diagnosis | Transplant/Dialysis |
| DGN10 | T8603 | Bone marrow transplant infection | Diagnosis | Transplant/Dialysis |
| DGN10 | T8609 | Other complications of bone marrow transplant | Diagnosis | Transplant/Dialysis |
| DGN10 | T865 | Complications of stem cell transplant | Diagnosis | Transplant/Dialysis |
| DGN10 | T86810 | Lung transplant rejection | Diagnosis | Transplant/Dialysis |
| DGN10 | T86811 | Lung transplant failure | Diagnosis | Transplant/Dialysis |
| DGN10 | T86812 | Lung transplant infection | Diagnosis | Transplant/Dialysis |
| DGN10 | T86818 | Other complications of lung transplant | Diagnosis | Transplant/Dialysis |
| DGN10 | T86819 | Unspecified complication of lung transplant | Diagnosis | Transplant/Dialysis |
| DGN10 | T86850 | Intestine transplant rejection | Diagnosis | Transplant/Dialysis |
| DGN10 | T86851 | Intestine transplant failure | Diagnosis | Transplant/Dialysis |
| DGN10 | T86852 | Intestine transplant infection | Diagnosis | Transplant/Dialysis |
| DGN10 | T86858 | Other complications of intestine transplant | Diagnosis | Transplant/Dialysis |
| DGN10 | T86859 | Unspecified complication of intestine transplant | Diagnosis | Transplant/Dialysis |
| DGN10 | T8690 | Unspecified complication of unspecified transplanted organ and tissue | Diagnosis | Transplant/Dialysis |
| DGN10 | T8691 | Unspecified transplanted organ and tissue rejection | Diagnosis | Transplant/Dialysis |
| DGN10 | T8692 | Unspecified transplanted organ and tissue failure | Diagnosis | Transplant/Dialysis |
| DGN10 | T8693 | Unspecified transplanted organ and tissue infection | Diagnosis | Transplant/Dialysis |
| DGN10 | T8699 | Other complications of unspecified transplanted organ and tissue | Diagnosis | Transplant/Dialysis |
| DGN10 | Y830 | Surgical operation with transplant of whole organ as the cause of abnormal reaction of the patient, or of later complication, without mention of misadventure at the time of the procedure | Diagnosis | Transplant/Dialysis |
| DGN10 | Z48288 | Encounter for aftercare following multiple organ transplant | Diagnosis | Transplant/Dialysis |
| DGN10 | Z48298 | Encounter for aftercare following other organ transplant | Diagnosis | Transplant/Dialysis |
| DGN10 | Z9489 | Other transplanted organ and tissue status | Diagnosis | Transplant/Dialysis |
| DGN10 | Z949 | Transplanted organ and tissue status, unspecified | Diagnosis | Transplant/Dialysis |
| DGN10 | T860 | Complications of bone marrow transplant | Diagnosis | Transplant/Dialysis |
| DGN10 | T861 | Complications of kidney transplant | Diagnosis | Transplant/Dialysis |
| DGN10 | T862 | Complications of heart transplant | Diagnosis | Transplant/Dialysis |
| DGN10 | T8629 | Other complications of heart transplant | Diagnosis | Transplant/Dialysis |
| DGN10 | T863 | Complications of heart-lung transplant | Diagnosis | Transplant/Dialysis |
| DGN10 | T864 | Complications of liver transplant | Diagnosis | Transplant/Dialysis |
| DGN10 | T868 | Complications of other transplanted organs and tissues | Diagnosis | Transplant/Dialysis |
| DGN10 | T8681 | Complications of lung transplant | Diagnosis | Transplant/Dialysis |
| DGN10 | T8685 | Complication of intestine transplant | Diagnosis | Transplant/Dialysis |
| DGN10 | Z482 | Encounter for aftercare following organ transplant | Diagnosis | Transplant/Dialysis |
| DGN10 | Z4828 | Encounter for aftercare following multiple organ transplant | Diagnosis | Transplant/Dialysis |
| DGN10 | Z4829 | Encounter for aftercare following other organ transplant | Diagnosis | Transplant/Dialysis |
| DGN10 | Z94 | Transplanted organ and tissue status | Diagnosis | Transplant/Dialysis |
| DGN10 | Z948 | Other transplanted organ and tissue status | Diagnosis | Transplant/Dialysis |
| DGN10 | Z49 | Encounter for care involving renal dialysis | Diagnosis | Transplant/Dialysis |
| DGN10 | Z490 | Preparatory care for renal dialysis | Diagnosis | Transplant/Dialysis |
| DGN10 | Z4901 | Encounter for fitting and adjustment of extracorporeal dialysis catheter | Diagnosis | Transplant/Dialysis |
| DGN10 | Z4902 | Encounter for fitting and adjustment of peritoneal dialysis catheter | Diagnosis | Transplant/Dialysis |
| DGN10 | Z493 | Encounter for adequacy testing for dialysis | Diagnosis | Transplant/Dialysis |
| DGN10 | Z4931 | Encounter for adequacy testing for hemodialysis | Diagnosis | Transplant/Dialysis |
| DGN10 | Z4932 | Encounter for adequacy testing for peritoneal dialysis | Diagnosis | Transplant/Dialysis |
| PRC10 | BT29 | Computerized Tomography (CT Scan) / Kidney Transplant | Diagnosis | Transplant/Dialysis |
| PRC10 | BT29ZZZ | Computerized Tomography (CT Scan) of Kidney Transplant | Diagnosis | Transplant/Dialysis |
| PRC10 | BT290ZZ | Computerized Tomography (CT Scan) of Kidney Transplant using High Osmolar Contrast | Diagnosis | Transplant/Dialysis |
| PRC10 | BT2900Z | Computerized Tomography (CT Scan) of Kidney Transplant using High Osmolar Contrast, Unenhanced and Enhanced | Diagnosis | Transplant/Dialysis |
| PRC10 | BT291ZZ | Computerized Tomography (CT Scan) of Kidney Transplant using Low Osmolar Contrast | Diagnosis | Transplant/Dialysis |
| PRC10 | BT2910Z | Computerized Tomography (CT Scan) of Kidney Transplant using Low Osmolar Contrast, Unenhanced and Enhanced | Diagnosis | Transplant/Dialysis |
| PRC10 | BT29YZZ | Computerized Tomography (CT Scan) of Kidney Transplant using Other Contrast | Diagnosis | Transplant/Dialysis |
| PRC10 | BT29Y0Z | Computerized Tomography (CT Scan) of Kidney Transplant using Other Contrast, Unenhanced and Enhanced | Diagnosis | Transplant/Dialysis |
| PRC10 | 0DY | Gastrointestinal System, Transplantation | Diagnosis | Transplant/Dialysis |
| PRC10 | 02Y | Heart and Great Vessels, Transplantation | Diagnosis | Transplant/Dialysis |
| PRC10 | 0FY | Hepatobiliary System and Pancreas, Transplantation | Diagnosis | Transplant/Dialysis |
| PRC10 | 07Y | Lymphatic and Hemic Systems, Transplantation | Diagnosis | Transplant/Dialysis |
| PRC10 | BT39 | Magnetic Resonance Imaging (MRI) / Kidney Transplant | Diagnosis | Transplant/Dialysis |
| PRC10 | BT39ZZZ | Magnetic Resonance Imaging (MRI) of Kidney Transplant | Diagnosis | Transplant/Dialysis |
| PRC10 | BT39YZZ | Magnetic Resonance Imaging (MRI) of Kidney Transplant using Other Contrast | Diagnosis | Transplant/Dialysis |
| PRC10 | BT39Y0Z | Magnetic Resonance Imaging (MRI) of Kidney Transplant using Other Contrast, Unenhanced and Enhanced | Diagnosis | Transplant/Dialysis |
| PRC10 | 0BY | Respiratory System, Transplantation | Diagnosis | Transplant/Dialysis |
| PRC10 | 0DY5 | Transplantation / Esophagus | Diagnosis | Transplant/Dialysis |
| PRC10 | 02YA | Transplantation / Heart | Diagnosis | Transplant/Dialysis |
| PRC10 | 0TY1 | Transplantation / Kidney, Left | Diagnosis | Transplant/Dialysis |
| PRC10 | 0TY0 | Transplantation / Kidney, Right | Diagnosis | Transplant/Dialysis |
| PRC10 | 0DYE | Transplantation / Large Intestine | Diagnosis | Transplant/Dialysis |
| PRC10 | 0FY0 | Transplantation / Liver | Diagnosis | Transplant/Dialysis |
| PRC10 | 0BYJ | Transplantation / Lower Lung Lobe, Left | Diagnosis | Transplant/Dialysis |
| PRC10 | 0BYF | Transplantation / Lower Lung Lobe, Right | Diagnosis | Transplant/Dialysis |
| PRC10 | 0BYH | Transplantation / Lung Lingula | Diagnosis | Transplant/Dialysis |
| PRC10 | 0BYL | Transplantation / Lung, Left | Diagnosis | Transplant/Dialysis |
| PRC10 | 0BYK | Transplantation / Lung, Right | Diagnosis | Transplant/Dialysis |
| PRC10 | 0BYM | Transplantation / Lungs, Bilateral | Diagnosis | Transplant/Dialysis |
| PRC10 | 0BYD | Transplantation / Middle Lung Lobe, Right | Diagnosis | Transplant/Dialysis |
| PRC10 | 0FYG | Transplantation / Pancreas | Diagnosis | Transplant/Dialysis |
| PRC10 | 0DY8 | Transplantation / Small Intestine | Diagnosis | Transplant/Dialysis |
| PRC10 | 07YP | Transplantation / Spleen | Diagnosis | Transplant/Dialysis |
| PRC10 | 0DY6 | Transplantation / Stomach | Diagnosis | Transplant/Dialysis |
| PRC10 | 0BYG | Transplantation / Upper Lung Lobe, Left | Diagnosis | Transplant/Dialysis |
| PRC10 | 0BYC | Transplantation / Upper Lung Lobe, Right | Diagnosis | Transplant/Dialysis |
| PRC10 | 0DY50Z0 | Transplantation of Esophagus, Allogeneic, Open Approach | Diagnosis | Transplant/Dialysis |
| PRC10 | 0DY50Z1 | Transplantation of Esophagus, Syngeneic, Open Approach | Diagnosis | Transplant/Dialysis |
| PRC10 | 0DY50Z2 | Transplantation of Esophagus, Zooplastic, Open Approach | Diagnosis | Transplant/Dialysis |
| PRC10 | 0DY60Z0 | Transplantation of Stomach, Allogeneic, Open Approach | Diagnosis | Transplant/Dialysis |
| PRC10 | 0DY60Z1 | Transplantation of Stomach, Syngeneic, Open Approach | Diagnosis | Transplant/Dialysis |
| PRC10 | 0DY60Z2 | Transplantation of Stomach, Zooplastic, Open Approach | Diagnosis | Transplant/Dialysis |
| PRC10 | BT49 | Ultrasonography / Kidney Transplant | Diagnosis | Transplant/Dialysis |
| PRC10 | BT49ZZZ | Ultrasonography of Kidney Transplant | Diagnosis | Transplant/Dialysis |
| PRC10 | 0TY | Urinary System, Transplantation | Diagnosis | Transplant/Dialysis |
| HCPCS | 55859 | Transperineal placement of needles or catheters into prostate for interstitial radioelement application, with or without cystoscopy | Treatment | Radiation |
| HCPCS | 55860 | Exposure of prostate, any approach, for insertion of radioactive substance; | Treatment | Radiation |
| HCPCS | 55862 | Exposure of prostate, any approach, for insertion of radioactive substance; with lymph node biopsy(s) (limited pelvic lymphadenectomy) | Treatment | Radiation |
| HCPCS | 55865 | Exposure of prostate, any approach, for insertion of radioactive substance; with bilateral pelvic lymphadenectomy, including external iliac, hypogastric and obturator nodes | Treatment | Radiation |
| HCPCS | 55875 | Transperineal placement of needles or catheters into prostate for interstitial radioelement application, with or without cystoscopy | Treatment | Radiation |
| HCPCS | 76950 | Ultrasonic guidance for placement of radiation therapy fields | Treatment | Radiation |
| HCPCS | 76960 | Ultrasonic guidance for placement of radiation therapy fields, except for B-scan echography | Treatment | Radiation |
| HCPCS | 77280 | Therapeutic radiology simulation-aided field setting; simple | Treatment | Radiation |
| HCPCS | 77285 | Therapeutic radiology simulation-aided field setting; intermediate | Treatment | Radiation |
| HCPCS | 77290 | Therapeutic radiology simulation-aided field setting; complex | Treatment | Radiation |
| HCPCS | 77295 | 3-dimensional radiotherapy plan, including dose-volume histograms | Treatment | Radiation |
| HCPCS | 77299 | Unlisted procedure, therapeutic radiology clinical treatment planning | Treatment | Radiation |
| HCPCS | 77300 | Basic radiation dosimetry calculation, central axis depth dose calculation, TDF, NSD, gap calculation, off axis factor, tissue inhomogeneity factors, calculation of non-ionizing radiation surface and depth dose, as required during course of treatment, only when prescribed by the treating physician | Treatment | Radiation |
| HCPCS | 77305 | Teletherapy, isodose plan (whether hand or computer calculated); simple (1 or 2 parallel opposed unmodified ports directed to a single area of interest) | Treatment | Radiation |
| HCPCS | 77310 | Teletherapy, isodose plan (whether hand or computer calculated); intermediate (3 or more treatment ports directed to a single area of interest) | Treatment | Radiation |
| HCPCS | 77315 | Teletherapy, isodose plan (whether hand or computer calculated); complex (mantle or inverted Y, tangential ports, the use of wedges, compensators, complex blocking, rotational beam, or special beam considerations) | Treatment | Radiation |
| HCPCS | 77321 | Special teletherapy port plan, particles, hemibody, total body | Treatment | Radiation |
| HCPCS | 77326 | Brachytherapy isodose plan; simple (calculation made from single plane, 1 to 4 sources/ribbon application, remote afterloading brachytherapy, 1 to 8 sources) | Treatment | Radiation |
| HCPCS | 77327 | Brachytherapy isodose plan; intermediate (multiplane dosage calculations, application involving 5 to 10 sources/ribbons, remote afterloading brachytherapy, 9 to 12 sources) | Treatment | Radiation |
| HCPCS | 77328 | Brachytherapy isodose plan; complex (multiplane isodose plan, volume implant calculations, over 10 sources/ribbons used, special spatial reconstruction, remote afterloading brachytherapy, over 12 sources) | Treatment | Radiation |
| HCPCS | 77401 | Radiation treatment delivery, superficial and/or ortho voltage, per day | Treatment | Radiation |
| HCPCS | 77402 | Radiation treatment delivery,=>1 MeV; simple | Treatment | Radiation |
| HCPCS | 77403 | Radiation treatment delivery, single treatment area, single port or parallel opposed ports, simple blocks or no blocks; 6-10 MeV | Treatment | Radiation |
| HCPCS | 77404 | Radiation treatment delivery, single treatment area, single port or parallel opposed ports, simple blocks or no blocks; 11-19 MeV | Treatment | Radiation |
| HCPCS | 77406 | Radiation treatment delivery, single treatment area, single port or parallel opposed ports, simple blocks or no blocks; 20 MeV or greater | Treatment | Radiation |
| HCPCS | 77407 | Radiation treatment delivery, =>1 MeV; intermediate | Treatment | Radiation |
| HCPCS | 77408 | Radiation treatment delivery, 2 separate treatment areas, 3 or more ports on a single treatment area, use of multiple blocks; 6-10 MeV | Treatment | Radiation |
| HCPCS | 77409 | Radiation treatment delivery, 2 separate treatment areas, 3 or more ports on a single treatment area, use of multiple blocks; 11-19 MeV | Treatment | Radiation |
| HCPCS | 77411 | Radiation treatment delivery, 2 separate treatment areas, 3 or more ports on a single treatment area, use of multiple blocks; 20 MeV or greater | Treatment | Radiation |
| HCPCS | 77412 | Radiation treatment delivery, =>1 MeV; complex | Treatment | Radiation |
| HCPCS | 77413 | Radiation treatment delivery, 3 or more separate treatment areas, custom blocking, tangential ports, wedges, rotational beam, compensators, electron beam; 6-10 MeV | Treatment | Radiation |
| HCPCS | 77414 | Radiation treatment delivery, 3 or more separate treatment areas, custom blocking, tangential ports, wedges, rotational beam, compensators, electron beam; 11-19 MeV | Treatment | Radiation |
| HCPCS | 77416 | Radiation treatment delivery, 3 or more separate treatment areas, custom blocking, tangential ports, wedges, rotational beam, compensators, electron beam; 20 MeV or greater | Treatment | Radiation |
| HCPCS | 77427 | Radiation treatment management, 5 treatments | Treatment | Radiation |
| HCPCS | 77432 | Stereotactic radiation treatment management of cranial lesion(s) (complete course of treatment consisting of 1 session) | Treatment | Radiation |
| HCPCS | 77469 | Intraoperative radiation treatment management | Treatment | Radiation |
| HCPCS | 77470 | Special treatment procedure (eg, total body irradiation, hemibody radiation, per oral or endocavitary irradiation) | Treatment | Radiation |
| HCPCS | 77499 | Unlisted procedure, therapeutic radiology treatment management | Treatment | Radiation |
| HCPCS | 77750 | Infusion or instillation of radioelement solution (includes 3-month follow-up care) | Treatment | Radiation |
| HCPCS | 77761 | Intracavitary radiation source application; simple | Treatment | Radiation |
| HCPCS | 77762 | Intracavitary radiation source application; intermediate | Treatment | Radiation |
| HCPCS | 77763 | Intracavitary radiation source application; complex | Treatment | Radiation |
| HCPCS | 77776 | Interstitial radiation source application; simple | Treatment | Radiation |
| HCPCS | 77777 | Interstitial radiation source application; intermediate | Treatment | Radiation |
| HCPCS | 77778 | Interstitial radiation source application; complex | Treatment | Radiation |
| HCPCS | 77781 | Remote afterloading high intensity brachytherapy; 1-4 source positions or catheters | Treatment | Radiation |
| HCPCS | 77782 | Remote afterloading high intensity brachytherapy; 5-8 source positions or catheters | Treatment | Radiation |
| HCPCS | 77783 | Remote afterloading high intensity brachytherapy; 9-12 source positions or catheters | Treatment | Radiation |
| HCPCS | 77784 | Remote afterloading high intensity brachytherapy; over 12 source positions or catheters | Treatment | Radiation |
| HCPCS | 77785 | Remote afterloading high dose rate radionuclide brachytherapy; 1 channel | Treatment | Radiation |
| HCPCS | 77786 | Remote afterloading high dose rate radionuclide brachytherapy; 2-12 channels | Treatment | Radiation |
| HCPCS | 77787 | Remote afterloading high dose rate radionuclide brachytherapy; over 12 channels | Treatment | Radiation |
| HCPCS | 77789 | Surface application of low dose rate radionuclide source | Treatment | Radiation |
| HCPCS | 77799 | Unlisted procedure, clinical brachytherapy | Treatment | Radiation |
| HCPCS | 79005 | Radiopharmaceutical therapy, by oral administration | Treatment | Radiation |
| HCPCS | 79101 | Radiopharmaceutical therapy, by intravenous administration | Treatment | Radiation |
| HCPCS | 79200 | Radiopharmaceutical therapy, by intracavitary administration | Treatment | Radiation |
| HCPCS | 79300 | Radiopharmaceutical therapy, by interstitial radioactive colloid administration | Treatment | Radiation |
| HCPCS | 79400 | Radiopharmaceutical therapy, nonthyroid, nonhematologic by intervenous injection | Treatment | Radiation |
| HCPCS | 79403 | Radiopharmaceutical therapy, radiolabeled monoclonal antibody by intravenous infusion | Treatment | Radiation |
| HCPCS | 79420 | Intravascular radiopharmaceutical therapy, particulate | Treatment | Radiation |
| HCPCS | 79440 | Radiopharmaceutical therapy, by intra-articular administration | Treatment | Radiation |
| HCPCS | 79900 | Provision of therapeutic radiopharmaceutical(s) | Treatment | Radiation |
| HCPCS | 79999 | Radiopharmaceutical therapy, unlisted procedure | Treatment | Radiation |
| PRC10 | 0BH001Z | Insertion of Radioactive Element into Tracheobronchial Tree, Open Approach | Treatment | Radiation |
| PRC10 | 0BH031Z | Insertion of Radioactive Element into Tracheobronchial Tree, Percutaneous Approach | Treatment | Radiation |
| PRC10 | 0BH041Z | Insertion of Radioactive Element into Tracheobronchial Tree, Percutaneous Endoscopic Approach | Treatment | Radiation |
| PRC10 | 0BH071Z | Insertion of Radioactive Element into Tracheobronchial Tree, Via Natural or Artificial Opening | Treatment | Radiation |
| PRC10 | 0BH081Z | Insertion of Radioactive Element into Tracheobronchial Tree, Via Natural or Artificial Opening Endoscopic | Treatment | Radiation |
| PRC10 | 0BHK01Z | Insertion of Radioactive Element into Right Lung, Open Approach | Treatment | Radiation |
| PRC10 | 0BHK31Z | Insertion of Radioactive Element into Right Lung, Percutaneous Approach | Treatment | Radiation |
| PRC10 | 0BHK41Z | Insertion of Radioactive Element into Right Lung, Percutaneous Endoscopic Approach | Treatment | Radiation |
| PRC10 | 0BHK71Z | Insertion of Radioactive Element into Right Lung, Via Natural or Artificial Opening | Treatment | Radiation |
| PRC10 | 0BHK81Z | Insertion of Radioactive Element into Right Lung, Via Natural or Artificial Opening Endoscopic | Treatment | Radiation |
| PRC10 | 0BHL01Z | Insertion of Radioactive Element into Left Lung, Open Approach | Treatment | Radiation |
| PRC10 | 0BHL31Z | Insertion of Radioactive Element into Left Lung, Percutaneous Approach | Treatment | Radiation |
| PRC10 | 0BHL41Z | Insertion of Radioactive Element into Left Lung, Percutaneous Endoscopic Approach | Treatment | Radiation |
| PRC10 | 0BHL71Z | Insertion of Radioactive Element into Left Lung, Via Natural or Artificial Opening | Treatment | Radiation |
| PRC10 | 0BHL81Z | Insertion of Radioactive Element into Left Lung, Via Natural or Artificial Opening Endoscopic | Treatment | Radiation |
| PRC10 | 0CH701Z | Insertion of Radioactive Element into Tongue, Open Approach | Treatment | Radiation |
| PRC10 | 0CH731Z | Insertion of Radioactive Element into Tongue, Percutaneous Approach | Treatment | Radiation |
| PRC10 | 0CH7X1Z | Insertion of Radioactive Element into Tongue, External Approach | Treatment | Radiation |
| PRC10 | 0DH501Z | Insertion of Radioactive Element into Esophagus, Open Approach | Treatment | Radiation |
| PRC10 | 0DH531Z | Insertion of Radioactive Element into Esophagus, Percutaneous Approach | Treatment | Radiation |
| PRC10 | 0DH541Z | Insertion of Radioactive Element into Esophagus, Percutaneous Endoscopic Approach | Treatment | Radiation |
| PRC10 | 0DH571Z | Insertion of Radioactive Element into Esophagus, Via Natural or Artificial Opening | Treatment | Radiation |
| PRC10 | 0DH581Z | Insertion of Radioactive Element into Esophagus, Via Natural or Artificial Opening Endoscopic | Treatment | Radiation |
| PRC10 | 0DHP01Z | Insertion of Radioactive Element into Rectum, Open Approach | Treatment | Radiation |
| PRC10 | 0DHP31Z | Insertion of Radioactive Element into Rectum, Percutaneous Approach | Treatment | Radiation |
| PRC10 | 0DHP41Z | Insertion of Radioactive Element into Rectum, Percutaneous Endoscopic Approach | Treatment | Radiation |
| PRC10 | 0DHP71Z | Insertion of Radioactive Element into Rectum, Via Natural or Artificial Opening | Treatment | Radiation |
| PRC10 | 0DHP81Z | Insertion of Radioactive Element into Rectum, Via Natural or Artificial Opening Endoscopic | Treatment | Radiation |
| PRC10 | 0FHB01Z | Insertion of Radioactive Element into Hepatobiliary Duct, Open Approach | Treatment | Radiation |
| PRC10 | 0FHB31Z | Insertion of Radioactive Element into Hepatobiliary Duct, Percutaneous Approach | Treatment | Radiation |
| PRC10 | 0FHB41Z | Insertion of Radioactive Element into Hepatobiliary Duct, Percutaneous Endoscopic Approach | Treatment | Radiation |
| PRC10 | 0FHB71Z | Insertion of Radioactive Element into Hepatobiliary Duct, Via Natural or Artificial Opening | Treatment | Radiation |
| PRC10 | 0FHB81Z | Insertion of Radioactive Element into Hepatobiliary Duct, Via Natural or Artificial Opening Endoscopic | Treatment | Radiation |
| PRC10 | 0FHD01Z | Insertion of Radioactive Element into Pancreatic Duct, Open Approach | Treatment | Radiation |
| PRC10 | 0FHD31Z | Insertion of Radioactive Element into Pancreatic Duct, Percutaneous Approach | Treatment | Radiation |
| PRC10 | 0FHD41Z | Insertion of Radioactive Element into Pancreatic Duct, Percutaneous Endoscopic Approach | Treatment | Radiation |
| PRC10 | 0FHD71Z | Insertion of Radioactive Element into Pancreatic Duct, Via Natural or Artificial Opening | Treatment | Radiation |
| PRC10 | 0FHD81Z | Insertion of Radioactive Element into Pancreatic Duct, Via Natural or Artificial Opening Endoscopic | Treatment | Radiation |
| PRC10 | 0HHT01Z | Insertion of Radioactive Element into Right Breast, Open Approach | Treatment | Radiation |
| PRC10 | 0HHT31Z | Insertion of Radioactive Element into Right Breast, Percutaneous Approach | Treatment | Radiation |
| PRC10 | 0HHT71Z | Insertion of Radioactive Element into Right Breast, Via Natural or Artificial Opening | Treatment | Radiation |
| PRC10 | 0HHT81Z | Insertion of Radioactive Element into Right Breast, Via Natural or Artificial Opening Endoscopic | Treatment | Radiation |
| PRC10 | 0HHTX1Z | Insertion of Radioactive Element into Right Breast, External Approach | Treatment | Radiation |
| PRC10 | 0HHU01Z | Insertion of Radioactive Element into Left Breast, Open Approach | Treatment | Radiation |
| PRC10 | 0HHU31Z | Insertion of Radioactive Element into Left Breast, Percutaneous Approach | Treatment | Radiation |
| PRC10 | 0HHU71Z | Insertion of Radioactive Element into Left Breast, Via Natural or Artificial Opening | Treatment | Radiation |
| PRC10 | 0HHU81Z | Insertion of Radioactive Element into Left Breast, Via Natural or Artificial Opening Endoscopic | Treatment | Radiation |
| PRC10 | 0HHUX1Z | Insertion of Radioactive Element into Left Breast, External Approach | Treatment | Radiation |
| PRC10 | 0HHV01Z | Insertion of Radioactive Element into Bilateral Breast, Open Approach | Treatment | Radiation |
| PRC10 | 0HHV31Z | Insertion of Radioactive Element into Bilateral Breast, Percutaneous Approach | Treatment | Radiation |
| PRC10 | 0HHV71Z | Insertion of Radioactive Element into Bilateral Breast, Via Natural or Artificial Opening | Treatment | Radiation |
| PRC10 | 0HHV81Z | Insertion of Radioactive Element into Bilateral Breast, Via Natural or Artificial Opening Endoscopic | Treatment | Radiation |
| PRC10 | 0HHVX1Z | Insertion of Radioactive Element into Bilateral Breast, External Approach | Treatment | Radiation |
| PRC10 | 0HHW01Z | Insertion of Radioactive Element into Right Nipple, Open Approach | Treatment | Radiation |
| PRC10 | 0HHW31Z | Insertion of Radioactive Element into Right Nipple, Percutaneous Approach | Treatment | Radiation |
| PRC10 | 0HHW71Z | Insertion of Radioactive Element into Right Nipple, Via Natural or Artificial Opening | Treatment | Radiation |
| PRC10 | 0HHW81Z | Insertion of Radioactive Element into Right Nipple, Via Natural or Artificial Opening Endoscopic | Treatment | Radiation |
| PRC10 | 0HHWX1Z | Insertion of Radioactive Element into Right Nipple, External Approach | Treatment | Radiation |
| PRC10 | 0HHX01Z | Insertion of Radioactive Element into Left Nipple, Open Approach | Treatment | Radiation |
| PRC10 | 0HHX31Z | Insertion of Radioactive Element into Left Nipple, Percutaneous Approach | Treatment | Radiation |
| PRC10 | 0HHX71Z | Insertion of Radioactive Element into Left Nipple, Via Natural or Artificial Opening | Treatment | Radiation |
| PRC10 | 0HHX81Z | Insertion of Radioactive Element into Left Nipple, Via Natural or Artificial Opening Endoscopic | Treatment | Radiation |
| PRC10 | 0HHXX1Z | Insertion of Radioactive Element into Left Nipple, External Approach | Treatment | Radiation |
| PRC10 | 0JHS01Z | Insertion of Radioactive Element into Head and Neck Subcutaneous Tissue and Fascia, Open Approach | Treatment | Radiation |
| PRC10 | 0JHS31Z | Insertion of Radioactive Element into Head and Neck Subcutaneous Tissue and Fascia, Percutaneous Approach | Treatment | Radiation |
| PRC10 | 0JHT01Z | Insertion of Radioactive Element into Trunk Subcutaneous Tissue and Fascia, Open Approach | Treatment | Radiation |
| PRC10 | 0JHT31Z | Insertion of Radioactive Element into Trunk Subcutaneous Tissue and Fascia, Percutaneous Approach | Treatment | Radiation |
| PRC10 | 0JHV01Z | Insertion of Radioactive Element into Upper Extremity Subcutaneous Tissue and Fascia, Open Approach | Treatment | Radiation |
| PRC10 | 0JHV31Z | Insertion of Radioactive Element into Upper Extremity Subcutaneous Tissue and Fascia, Percutaneous Approach | Treatment | Radiation |
| PRC10 | 0JHW01Z | Insertion of Radioactive Element into Lower Extremity Subcutaneous Tissue and Fascia, Open Approach | Treatment | Radiation |
| PRC10 | 0JHW31Z | Insertion of Radioactive Element into Lower Extremity Subcutaneous Tissue and Fascia, Percutaneous Approach | Treatment | Radiation |
| PRC10 | 0UHC01Z | Insertion of Radioactive Element into Cervix, Open Approach | Treatment | Radiation |
| PRC10 | 0UHC31Z | Insertion of Radioactive Element into Cervix, Percutaneous Approach | Treatment | Radiation |
| PRC10 | 0UHC41Z | Insertion of Radioactive Element into Cervix, Percutaneous Endoscopic Approach | Treatment | Radiation |
| PRC10 | 0UHC71Z | Insertion of Radioactive Element into Cervix, Via Natural or Artificial Opening | Treatment | Radiation |
| PRC10 | 0UHC81Z | Insertion of Radioactive Element into Cervix, Via Natural or Artificial Opening Endoscopic | Treatment | Radiation |
| PRC10 | 0UHG01Z | Insertion of Radioactive Element into Vagina, Open Approach | Treatment | Radiation |
| PRC10 | 0UHG31Z | Insertion of Radioactive Element into Vagina, Percutaneous Approach | Treatment | Radiation |
| PRC10 | 0UHG41Z | Insertion of Radioactive Element into Vagina, Percutaneous Endoscopic Approach | Treatment | Radiation |
| PRC10 | 0UHG71Z | Insertion of Radioactive Element into Vagina, Via Natural or Artificial Opening | Treatment | Radiation |
| PRC10 | 0UHG81Z | Insertion of Radioactive Element into Vagina, Via Natural or Artificial Opening Endoscopic | Treatment | Radiation |
| PRC10 | 0UHGX1Z | Insertion of Radioactive Element into Vagina, External Approach | Treatment | Radiation |
| PRC10 | 0VH001Z | Insertion of Radioactive Element into Prostate, Open Approach | Treatment | Radiation |
| PRC10 | 0VH031Z | Insertion of Radioactive Element into Prostate, Percutaneous Approach | Treatment | Radiation |
| PRC10 | 0VH041Z | Insertion of Radioactive Element into Prostate, Percutaneous Endoscopic Approach | Treatment | Radiation |
| PRC10 | 0VH071Z | Insertion of Radioactive Element into Prostate, Via Natural or Artificial Opening | Treatment | Radiation |
| PRC10 | 0VH081Z | Insertion of Radioactive Element into Prostate, Via Natural or Artificial Opening Endoscopic | Treatment | Radiation |
| PRC10 | 0WH001Z | Insertion of Radioactive Element into Head, Open Approach | Treatment | Radiation |
| PRC10 | 0WH031Z | Insertion of Radioactive Element into Head, Percutaneous Approach | Treatment | Radiation |
| PRC10 | 0WH041Z | Insertion of Radioactive Element into Head, Percutaneous Endoscopic Approach | Treatment | Radiation |
| PRC10 | 0WH101Z | Insertion of Radioactive Element into Cranial Cavity, Open Approach | Treatment | Radiation |
| PRC10 | 0WH131Z | Insertion of Radioactive Element into Cranial Cavity, Percutaneous Approach | Treatment | Radiation |
| PRC10 | 0WH141Z | Insertion of Radioactive Element into Cranial Cavity, Percutaneous Endoscopic Approach | Treatment | Radiation |
| PRC10 | 0WH201Z | Insertion of Radioactive Element into Face, Open Approach | Treatment | Radiation |
| PRC10 | 0WH231Z | Insertion of Radioactive Element into Face, Percutaneous Approach | Treatment | Radiation |
| PRC10 | 0WH241Z | Insertion of Radioactive Element into Face, Percutaneous Endoscopic Approach | Treatment | Radiation |
| PRC10 | 0WH301Z | Insertion of Radioactive Element into Oral Cavity and Throat, Open Approach | Treatment | Radiation |
| PRC10 | 0WH331Z | Insertion of Radioactive Element into Oral Cavity and Throat, Percutaneous Approach | Treatment | Radiation |
| PRC10 | 0WH341Z | Insertion of Radioactive Element into Oral Cavity and Throat, Percutaneous Endoscopic Approach | Treatment | Radiation |
| PRC10 | 0WH401Z | Insertion of Radioactive Element into Upper Jaw, Open Approach | Treatment | Radiation |
| PRC10 | 0WH431Z | Insertion of Radioactive Element into Upper Jaw, Percutaneous Approach | Treatment | Radiation |
| PRC10 | 0WH441Z | Insertion of Radioactive Element into Upper Jaw, Percutaneous Endoscopic Approach | Treatment | Radiation |
| PRC10 | 0WH501Z | Insertion of Radioactive Element into Lower Jaw, Open Approach | Treatment | Radiation |
| PRC10 | 0WH531Z | Insertion of Radioactive Element into Lower Jaw, Percutaneous Approach | Treatment | Radiation |
| PRC10 | 0WH541Z | Insertion of Radioactive Element into Lower Jaw, Percutaneous Endoscopic Approach | Treatment | Radiation |
| PRC10 | 0WH601Z | Insertion of Radioactive Element into Neck, Open Approach | Treatment | Radiation |
| PRC10 | 0WH631Z | Insertion of Radioactive Element into Neck, Percutaneous Approach | Treatment | Radiation |
| PRC10 | 0WH641Z | Insertion of Radioactive Element into Neck, Percutaneous Endoscopic Approach | Treatment | Radiation |
| PRC10 | 0WH801Z | Insertion of Radioactive Element into Chest Wall, Open Approach | Treatment | Radiation |
| PRC10 | 0WH831Z | Insertion of Radioactive Element into Chest Wall, Percutaneous Approach | Treatment | Radiation |
| PRC10 | 0WH841Z | Insertion of Radioactive Element into Chest Wall, Percutaneous Endoscopic Approach | Treatment | Radiation |
| PRC10 | 0WH901Z | Insertion of Radioactive Element into Right Pleural Cavity, Open Approach | Treatment | Radiation |
| PRC10 | 0WH931Z | Insertion of Radioactive Element into Right Pleural Cavity, Percutaneous Approach | Treatment | Radiation |
| PRC10 | 0WH941Z | Insertion of Radioactive Element into Right Pleural Cavity, Percutaneous Endoscopic Approach | Treatment | Radiation |
| PRC10 | 0WHB01Z | Insertion of Radioactive Element into Left Pleural Cavity, Open Approach | Treatment | Radiation |
| PRC10 | 0WHB31Z | Insertion of Radioactive Element into Left Pleural Cavity, Percutaneous Approach | Treatment | Radiation |
| PRC10 | 0WHB41Z | Insertion of Radioactive Element into Left Pleural Cavity, Percutaneous Endoscopic Approach | Treatment | Radiation |
| PRC10 | 0WHC01Z | Insertion of Radioactive Element into Mediastinum, Open Approach | Treatment | Radiation |
| PRC10 | 0WHC31Z | Insertion of Radioactive Element into Mediastinum, Percutaneous Approach | Treatment | Radiation |
| PRC10 | 0WHC41Z | Insertion of Radioactive Element into Mediastinum, Percutaneous Endoscopic Approach | Treatment | Radiation |
| PRC10 | 0WHD01Z | Insertion of Radioactive Element into Pericardial Cavity, Open Approach | Treatment | Radiation |
| PRC10 | 0WHD31Z | Insertion of Radioactive Element into Pericardial Cavity, Percutaneous Approach | Treatment | Radiation |
| PRC10 | 0WHD41Z | Insertion of Radioactive Element into Pericardial Cavity, Percutaneous Endoscopic Approach | Treatment | Radiation |
| PRC10 | 0WHF01Z | Insertion of Radioactive Element into Abdominal Wall, Open Approach | Treatment | Radiation |
| PRC10 | 0WHF31Z | Insertion of Radioactive Element into Abdominal Wall, Percutaneous Approach | Treatment | Radiation |
| PRC10 | 0WHF41Z | Insertion of Radioactive Element into Abdominal Wall, Percutaneous Endoscopic Approach | Treatment | Radiation |
| PRC10 | 0WHG01Z | Insertion of Radioactive Element into Peritoneal Cavity, Open Approach | Treatment | Radiation |
| PRC10 | 0WHG31Z | Insertion of Radioactive Element into Peritoneal Cavity, Percutaneous Approach | Treatment | Radiation |
| PRC10 | 0WHG41Z | Insertion of Radioactive Element into Peritoneal Cavity, Percutaneous Endoscopic Approach | Treatment | Radiation |
| PRC10 | 0WHH01Z | Insertion of Radioactive Element into Retroperitoneum, Open Approach | Treatment | Radiation |
| PRC10 | 0WHH31Z | Insertion of Radioactive Element into Retroperitoneum, Percutaneous Approach | Treatment | Radiation |
| PRC10 | 0WHH41Z | Insertion of Radioactive Element into Retroperitoneum, Percutaneous Endoscopic Approach | Treatment | Radiation |
| PRC10 | 0WHJ01Z | Insertion of Radioactive Element into Pelvic Cavity, Open Approach | Treatment | Radiation |
| PRC10 | 0WHJ31Z | Insertion of Radioactive Element into Pelvic Cavity, Percutaneous Approach | Treatment | Radiation |
| PRC10 | 0WHJ41Z | Insertion of Radioactive Element into Pelvic Cavity, Percutaneous Endoscopic Approach | Treatment | Radiation |
| PRC10 | 0WHK01Z | Insertion of Radioactive Element into Upper Back, Open Approach | Treatment | Radiation |
| PRC10 | 0WHK31Z | Insertion of Radioactive Element into Upper Back, Percutaneous Approach | Treatment | Radiation |
| PRC10 | 0WHK41Z | Insertion of Radioactive Element into Upper Back, Percutaneous Endoscopic Approach | Treatment | Radiation |
| PRC10 | 0WHL01Z | Insertion of Radioactive Element into Lower Back, Open Approach | Treatment | Radiation |
| PRC10 | 0WHL31Z | Insertion of Radioactive Element into Lower Back, Percutaneous Approach | Treatment | Radiation |
| PRC10 | 0WHL41Z | Insertion of Radioactive Element into Lower Back, Percutaneous Endoscopic Approach | Treatment | Radiation |
| PRC10 | 0WHM01Z | Insertion of Radioactive Element into Male Perineum, Open Approach | Treatment | Radiation |
| PRC10 | 0WHM31Z | Insertion of Radioactive Element into Male Perineum, Percutaneous Approach | Treatment | Radiation |
| PRC10 | 0WHM41Z | Insertion of Radioactive Element into Male Perineum, Percutaneous Endoscopic Approach | Treatment | Radiation |
| PRC10 | 0WHN01Z | Insertion of Radioactive Element into Female Perineum, Open Approach | Treatment | Radiation |
| PRC10 | 0WHN31Z | Insertion of Radioactive Element into Female Perineum, Percutaneous Approach | Treatment | Radiation |
| PRC10 | 0WHN41Z | Insertion of Radioactive Element into Female Perineum, Percutaneous Endoscopic Approach | Treatment | Radiation |
| PRC10 | 0WHP01Z | Insertion of Radioactive Element into Gastrointestinal Tract, Open Approach | Treatment | Radiation |
| PRC10 | 0WHP31Z | Insertion of Radioactive Element into Gastrointestinal Tract, Percutaneous Approach | Treatment | Radiation |
| PRC10 | 0WHP41Z | Insertion of Radioactive Element into Gastrointestinal Tract, Percutaneous Endoscopic Approach | Treatment | Radiation |
| PRC10 | 0WHP71Z | Insertion of Radioactive Element into Gastrointestinal Tract, Via Natural or Artificial Opening | Treatment | Radiation |
| PRC10 | 0WHP81Z | Insertion of Radioactive Element into Gastrointestinal Tract, Via Natural or Artificial Opening Endoscopic | Treatment | Radiation |
| PRC10 | 0WHQ01Z | Insertion of Radioactive Element into Respiratory Tract, Open Approach | Treatment | Radiation |
| PRC10 | 0WHQ31Z | Insertion of Radioactive Element into Respiratory Tract, Percutaneous Approach | Treatment | Radiation |
| PRC10 | 0WHQ41Z | Insertion of Radioactive Element into Respiratory Tract, Percutaneous Endoscopic Approach | Treatment | Radiation |
| PRC10 | 0WHQ71Z | Insertion of Radioactive Element into Respiratory Tract, Via Natural or Artificial Opening | Treatment | Radiation |
| PRC10 | 0WHQ81Z | Insertion of Radioactive Element into Respiratory Tract, Via Natural or Artificial Opening Endoscopic | Treatment | Radiation |
| PRC10 | 0WHR01Z | Insertion of Radioactive Element into Genitourinary Tract, Open Approach | Treatment | Radiation |
| PRC10 | 0WHR31Z | Insertion of Radioactive Element into Genitourinary Tract, Percutaneous Approach | Treatment | Radiation |
| PRC10 | 0WHR41Z | Insertion of Radioactive Element into Genitourinary Tract, Percutaneous Endoscopic Approach | Treatment | Radiation |
| PRC10 | 0WHR71Z | Insertion of Radioactive Element into Genitourinary Tract, Via Natural or Artificial Opening | Treatment | Radiation |
| PRC10 | 0WHR81Z | Insertion of Radioactive Element into Genitourinary Tract, Via Natural or Artificial Opening Endoscopic | Treatment | Radiation |
| PRC10 | 0XH201Z | Insertion of Radioactive Element into Right Shoulder Region, Open Approach | Treatment | Radiation |
| PRC10 | 0XH231Z | Insertion of Radioactive Element into Right Shoulder Region, Percutaneous Approach | Treatment | Radiation |
| PRC10 | 0XH241Z | Insertion of Radioactive Element into Right Shoulder Region, Percutaneous Endoscopic Approach | Treatment | Radiation |
| PRC10 | 0XH301Z | Insertion of Radioactive Element into Left Shoulder Region, Open Approach | Treatment | Radiation |
| PRC10 | 0XH331Z | Insertion of Radioactive Element into Left Shoulder Region, Percutaneous Approach | Treatment | Radiation |
| PRC10 | 0XH341Z | Insertion of Radioactive Element into Left Shoulder Region, Percutaneous Endoscopic Approach | Treatment | Radiation |
| PRC10 | 0XH401Z | Insertion of Radioactive Element into Right Axilla, Open Approach | Treatment | Radiation |
| PRC10 | 0XH431Z | Insertion of Radioactive Element into Right Axilla, Percutaneous Approach | Treatment | Radiation |
| PRC10 | 0XH441Z | Insertion of Radioactive Element into Right Axilla, Percutaneous Endoscopic Approach | Treatment | Radiation |
| PRC10 | 0XH501Z | Insertion of Radioactive Element into Left Axilla, Open Approach | Treatment | Radiation |
| PRC10 | 0XH531Z | Insertion of Radioactive Element into Left Axilla, Percutaneous Approach | Treatment | Radiation |
| PRC10 | 0XH541Z | Insertion of Radioactive Element into Left Axilla, Percutaneous Endoscopic Approach | Treatment | Radiation |
| PRC10 | 0XH601Z | Insertion of Radioactive Element into Right Upper Extremity, Open Approach | Treatment | Radiation |
| PRC10 | 0XH631Z | Insertion of Radioactive Element into Right Upper Extremity, Percutaneous Approach | Treatment | Radiation |
| PRC10 | 0XH641Z | Insertion of Radioactive Element into Right Upper Extremity, Percutaneous Endoscopic Approach | Treatment | Radiation |
| PRC10 | 0XH701Z | Insertion of Radioactive Element into Left Upper Extremity, Open Approach | Treatment | Radiation |
| PRC10 | 0XH731Z | Insertion of Radioactive Element into Left Upper Extremity, Percutaneous Approach | Treatment | Radiation |
| PRC10 | 0XH741Z | Insertion of Radioactive Element into Left Upper Extremity, Percutaneous Endoscopic Approach | Treatment | Radiation |
| PRC10 | 0XH801Z | Insertion of Radioactive Element into Right Upper Arm, Open Approach | Treatment | Radiation |
| PRC10 | 0XH831Z | Insertion of Radioactive Element into Right Upper Arm, Percutaneous Approach | Treatment | Radiation |
| PRC10 | 0XH841Z | Insertion of Radioactive Element into Right Upper Arm, Percutaneous Endoscopic Approach | Treatment | Radiation |
| PRC10 | 0XH901Z | Insertion of Radioactive Element into Left Upper Arm, Open Approach | Treatment | Radiation |
| PRC10 | 0XH931Z | Insertion of Radioactive Element into Left Upper Arm, Percutaneous Approach | Treatment | Radiation |
| PRC10 | 0XH941Z | Insertion of Radioactive Element into Left Upper Arm, Percutaneous Endoscopic Approach | Treatment | Radiation |
| PRC10 | 0XHB01Z | Insertion of Radioactive Element into Right Elbow Region, Open Approach | Treatment | Radiation |
| PRC10 | 0XHB31Z | Insertion of Radioactive Element into Right Elbow Region, Percutaneous Approach | Treatment | Radiation |
| PRC10 | 0XHB41Z | Insertion of Radioactive Element into Right Elbow Region, Percutaneous Endoscopic Approach | Treatment | Radiation |
| PRC10 | 0XHC01Z | Insertion of Radioactive Element into Left Elbow Region, Open Approach | Treatment | Radiation |
| PRC10 | 0XHC31Z | Insertion of Radioactive Element into Left Elbow Region, Percutaneous Approach | Treatment | Radiation |
| PRC10 | 0XHC41Z | Insertion of Radioactive Element into Left Elbow Region, Percutaneous Endoscopic Approach | Treatment | Radiation |
| PRC10 | 0XHD01Z | Insertion of Radioactive Element into Right Lower Arm, Open Approach | Treatment | Radiation |
| PRC10 | 0XHD31Z | Insertion of Radioactive Element into Right Lower Arm, Percutaneous Approach | Treatment | Radiation |
| PRC10 | 0XHD41Z | Insertion of Radioactive Element into Right Lower Arm, Percutaneous Endoscopic Approach | Treatment | Radiation |
| PRC10 | 0XHF01Z | Insertion of Radioactive Element into Left Lower Arm, Open Approach | Treatment | Radiation |
| PRC10 | 0XHF31Z | Insertion of Radioactive Element into Left Lower Arm, Percutaneous Approach | Treatment | Radiation |
| PRC10 | 0XHF41Z | Insertion of Radioactive Element into Left Lower Arm, Percutaneous Endoscopic Approach | Treatment | Radiation |
| PRC10 | 0XHG01Z | Insertion of Radioactive Element into Right Wrist Region, Open Approach | Treatment | Radiation |
| PRC10 | 0XHG31Z | Insertion of Radioactive Element into Right Wrist Region, Percutaneous Approach | Treatment | Radiation |
| PRC10 | 0XHG41Z | Insertion of Radioactive Element into Right Wrist Region, Percutaneous Endoscopic Approach | Treatment | Radiation |
| PRC10 | 0XHH01Z | Insertion of Radioactive Element into Left Wrist Region, Open Approach | Treatment | Radiation |
| PRC10 | 0XHH31Z | Insertion of Radioactive Element into Left Wrist Region, Percutaneous Approach | Treatment | Radiation |
| PRC10 | 0XHH41Z | Insertion of Radioactive Element into Left Wrist Region, Percutaneous Endoscopic Approach | Treatment | Radiation |
| PRC10 | 0XHJ01Z | Insertion of Radioactive Element into Right Hand, Open Approach | Treatment | Radiation |
| PRC10 | 0XHJ31Z | Insertion of Radioactive Element into Right Hand, Percutaneous Approach | Treatment | Radiation |
| PRC10 | 0XHJ41Z | Insertion of Radioactive Element into Right Hand, Percutaneous Endoscopic Approach | Treatment | Radiation |
| PRC10 | 0XHK01Z | Insertion of Radioactive Element into Left Hand, Open Approach | Treatment | Radiation |
| PRC10 | 0XHK31Z | Insertion of Radioactive Element into Left Hand, Percutaneous Approach | Treatment | Radiation |
| PRC10 | 0XHK41Z | Insertion of Radioactive Element into Left Hand, Percutaneous Endoscopic Approach | Treatment | Radiation |
| PRC10 | 0YH001Z | Insertion of Radioactive Element into Right Buttock, Open Approach | Treatment | Radiation |
| PRC10 | 0YH031Z | Insertion of Radioactive Element into Right Buttock, Percutaneous Approach | Treatment | Radiation |
| PRC10 | 0YH041Z | Insertion of Radioactive Element into Right Buttock, Percutaneous Endoscopic Approach | Treatment | Radiation |
| PRC10 | 0YH101Z | Insertion of Radioactive Element into Left Buttock, Open Approach | Treatment | Radiation |
| PRC10 | 0YH131Z | Insertion of Radioactive Element into Left Buttock, Percutaneous Approach | Treatment | Radiation |
| PRC10 | 0YH141Z | Insertion of Radioactive Element into Left Buttock, Percutaneous Endoscopic Approach | Treatment | Radiation |
| PRC10 | 0YH501Z | Insertion of Radioactive Element into Right Inguinal Region, Open Approach | Treatment | Radiation |
| PRC10 | 0YH531Z | Insertion of Radioactive Element into Right Inguinal Region, Percutaneous Approach | Treatment | Radiation |
| PRC10 | 0YH541Z | Insertion of Radioactive Element into Right Inguinal Region, Percutaneous Endoscopic Approach | Treatment | Radiation |
| PRC10 | 0YH601Z | Insertion of Radioactive Element into Left Inguinal Region, Open Approach | Treatment | Radiation |
| PRC10 | 0YH631Z | Insertion of Radioactive Element into Left Inguinal Region, Percutaneous Approach | Treatment | Radiation |
| PRC10 | 0YH641Z | Insertion of Radioactive Element into Left Inguinal Region, Percutaneous Endoscopic Approach | Treatment | Radiation |
| PRC10 | 0YH701Z | Insertion of Radioactive Element into Right Femoral Region, Open Approach | Treatment | Radiation |
| PRC10 | 0YH731Z | Insertion of Radioactive Element into Right Femoral Region, Percutaneous Approach | Treatment | Radiation |
| PRC10 | 0YH741Z | Insertion of Radioactive Element into Right Femoral Region, Percutaneous Endoscopic Approach | Treatment | Radiation |
| PRC10 | 0YH801Z | Insertion of Radioactive Element into Left Femoral Region, Open Approach | Treatment | Radiation |
| PRC10 | 0YH831Z | Insertion of Radioactive Element into Left Femoral Region, Percutaneous Approach | Treatment | Radiation |
| PRC10 | 0YH841Z | Insertion of Radioactive Element into Left Femoral Region, Percutaneous Endoscopic Approach | Treatment | Radiation |
| PRC10 | 0YH901Z | Insertion of Radioactive Element into Right Lower Extremity, Open Approach | Treatment | Radiation |
| PRC10 | 0YH931Z | Insertion of Radioactive Element into Right Lower Extremity, Percutaneous Approach | Treatment | Radiation |
| PRC10 | 0YH941Z | Insertion of Radioactive Element into Right Lower Extremity, Percutaneous Endoscopic Approach | Treatment | Radiation |
| PRC10 | 0YHB01Z | Insertion of Radioactive Element into Left Lower Extremity, Open Approach | Treatment | Radiation |
| PRC10 | 0YHB31Z | Insertion of Radioactive Element into Left Lower Extremity, Percutaneous Approach | Treatment | Radiation |
| PRC10 | 0YHB41Z | Insertion of Radioactive Element into Left Lower Extremity, Percutaneous Endoscopic Approach | Treatment | Radiation |
| PRC10 | 0YHC01Z | Insertion of Radioactive Element into Right Upper Leg, Open Approach | Treatment | Radiation |
| PRC10 | 0YHC31Z | Insertion of Radioactive Element into Right Upper Leg, Percutaneous Approach | Treatment | Radiation |
| PRC10 | 0YHC41Z | Insertion of Radioactive Element into Right Upper Leg, Percutaneous Endoscopic Approach | Treatment | Radiation |
| PRC10 | 0YHD01Z | Insertion of Radioactive Element into Left Upper Leg, Open Approach | Treatment | Radiation |
| PRC10 | 0YHD31Z | Insertion of Radioactive Element into Left Upper Leg, Percutaneous Approach | Treatment | Radiation |
| PRC10 | 0YHD41Z | Insertion of Radioactive Element into Left Upper Leg, Percutaneous Endoscopic Approach | Treatment | Radiation |
| PRC10 | 0YHF01Z | Insertion of Radioactive Element into Right Knee Region, Open Approach | Treatment | Radiation |
| PRC10 | 0YHF31Z | Insertion of Radioactive Element into Right Knee Region, Percutaneous Approach | Treatment | Radiation |
| PRC10 | 0YHF41Z | Insertion of Radioactive Element into Right Knee Region, Percutaneous Endoscopic Approach | Treatment | Radiation |
| PRC10 | 0YHG01Z | Insertion of Radioactive Element into Left Knee Region, Open Approach | Treatment | Radiation |
| PRC10 | 0YHG31Z | Insertion of Radioactive Element into Left Knee Region, Percutaneous Approach | Treatment | Radiation |
| PRC10 | 0YHG41Z | Insertion of Radioactive Element into Left Knee Region, Percutaneous Endoscopic Approach | Treatment | Radiation |
| PRC10 | 0YHH01Z | Insertion of Radioactive Element into Right Lower Leg, Open Approach | Treatment | Radiation |
| PRC10 | 0YHH31Z | Insertion of Radioactive Element into Right Lower Leg, Percutaneous Approach | Treatment | Radiation |
| PRC10 | 0YHH41Z | Insertion of Radioactive Element into Right Lower Leg, Percutaneous Endoscopic Approach | Treatment | Radiation |
| PRC10 | 0YHJ01Z | Insertion of Radioactive Element into Left Lower Leg, Open Approach | Treatment | Radiation |
| PRC10 | 0YHJ31Z | Insertion of Radioactive Element into Left Lower Leg, Percutaneous Approach | Treatment | Radiation |
| PRC10 | 0YHJ41Z | Insertion of Radioactive Element into Left Lower Leg, Percutaneous Endoscopic Approach | Treatment | Radiation |
| PRC10 | 0YHK01Z | Insertion of Radioactive Element into Right Ankle Region, Open Approach | Treatment | Radiation |
| PRC10 | 0YHK31Z | Insertion of Radioactive Element into Right Ankle Region, Percutaneous Approach | Treatment | Radiation |
| PRC10 | 0YHK41Z | Insertion of Radioactive Element into Right Ankle Region, Percutaneous Endoscopic Approach | Treatment | Radiation |
| PRC10 | 0YHL01Z | Insertion of Radioactive Element into Left Ankle Region, Open Approach | Treatment | Radiation |
| PRC10 | 0YHL31Z | Insertion of Radioactive Element into Left Ankle Region, Percutaneous Approach | Treatment | Radiation |
| PRC10 | 0YHL41Z | Insertion of Radioactive Element into Left Ankle Region, Percutaneous Endoscopic Approach | Treatment | Radiation |
| PRC10 | 0YHM01Z | Insertion of Radioactive Element into Right Foot, Open Approach | Treatment | Radiation |
| PRC10 | 0YHM31Z | Insertion of Radioactive Element into Right Foot, Percutaneous Approach | Treatment | Radiation |
| PRC10 | 0YHM41Z | Insertion of Radioactive Element into Right Foot, Percutaneous Endoscopic Approach | Treatment | Radiation |
| PRC10 | 0YHN01Z | Insertion of Radioactive Element into Left Foot, Open Approach | Treatment | Radiation |
| PRC10 | 0YHN31Z | Insertion of Radioactive Element into Left Foot, Percutaneous Approach | Treatment | Radiation |
| PRC10 | 0YHN41Z | Insertion of Radioactive Element into Left Foot, Percutaneous Endoscopic Approach | Treatment | Radiation |
| PRC10 | 3E0B304 | Introduction of Liquid Brachytherapy Radioisotope into Ear, Percutaneous Approach | Treatment | Radiation |
| PRC10 | 3E0B704 | Introduction of Liquid Brachytherapy Radioisotope into Ear, Via Natural or Artificial Opening | Treatment | Radiation |
| PRC10 | 3E0BX04 | Introduction of Liquid Brachytherapy Radioisotope into Ear, External Approach | Treatment | Radiation |
| PRC10 | 3E0C304 | Introduction of Liquid Brachytherapy Radioisotope into Eye, Percutaneous Approach | Treatment | Radiation |
| PRC10 | 3E0C704 | Introduction of Liquid Brachytherapy Radioisotope into Eye, Via Natural or Artificial Opening | Treatment | Radiation |
| PRC10 | 3E0CX04 | Introduction of Liquid Brachytherapy Radioisotope into Eye, External Approach | Treatment | Radiation |
| PRC10 | 3E0D304 | Introduction of Liquid Brachytherapy Radioisotope into Mouth and Pharynx, Percutaneous Approach | Treatment | Radiation |
| PRC10 | 3E0D704 | Introduction of Liquid Brachytherapy Radioisotope into Mouth and Pharynx, Via Natural or Artificial Opening | Treatment | Radiation |
| PRC10 | 3E0DX04 | Introduction of Liquid Brachytherapy Radioisotope into Mouth and Pharynx, External Approach | Treatment | Radiation |
| PRC10 | 3E0E304 | Introduction of Liquid Brachytherapy Radioisotope into Products of Conception, Percutaneous Approach | Treatment | Radiation |
| PRC10 | 3E0E704 | Introduction of Liquid Brachytherapy Radioisotope into Products of Conception, Via Natural or Artificial Opening | Treatment | Radiation |
| PRC10 | 3E0E804 | Introduction of Liquid Brachytherapy Radioisotope into Products of Conception, Via Natural or Artificial Opening Endoscopic | Treatment | Radiation |
| PRC10 | 3E0F304 | Introduction of Liquid Brachytherapy Radioisotope into Respiratory Tract, Percutaneous Approach | Treatment | Radiation |
| PRC10 | 3E0F704 | Introduction of Liquid Brachytherapy Radioisotope into Respiratory Tract, Via Natural or Artificial Opening | Treatment | Radiation |
| PRC10 | 3E0F804 | Introduction of Liquid Brachytherapy Radioisotope into Respiratory Tract, Via Natural or Artificial Opening Endoscopic | Treatment | Radiation |
| PRC10 | 3E0G304 | Introduction of Liquid Brachytherapy Radioisotope into Upper GI, Percutaneous Approach | Treatment | Radiation |
| PRC10 | 3E0G704 | Introduction of Liquid Brachytherapy Radioisotope into Upper GI, Via Natural or Artificial Opening | Treatment | Radiation |
| PRC10 | 3E0G804 | Introduction of Liquid Brachytherapy Radioisotope into Upper GI, Via Natural or Artificial Opening Endoscopic | Treatment | Radiation |
| PRC10 | 3E0H304 | Introduction of Liquid Brachytherapy Radioisotope into Lower GI, Percutaneous Approach | Treatment | Radiation |
| PRC10 | 3E0H704 | Introduction of Liquid Brachytherapy Radioisotope into Lower GI, Via Natural or Artificial Opening | Treatment | Radiation |
| PRC10 | 3E0H804 | Introduction of Liquid Brachytherapy Radioisotope into Lower GI, Via Natural or Artificial Opening Endoscopic | Treatment | Radiation |
| PRC10 | 3E0J304 | Introduction of Liquid Brachytherapy Radioisotope into Biliary and Pancreatic Tract, Percutaneous Approach | Treatment | Radiation |
| PRC10 | 3E0J704 | Introduction of Liquid Brachytherapy Radioisotope into Biliary and Pancreatic Tract, Via Natural or Artificial Opening | Treatment | Radiation |
| PRC10 | 3E0J804 | Introduction of Liquid Brachytherapy Radioisotope into Biliary and Pancreatic Tract, Via Natural or Artificial Opening Endoscopic | Treatment | Radiation |
| PRC10 | 3E0K304 | Introduction of Liquid Brachytherapy Radioisotope into Genitourinary Tract, Percutaneous Approach | Treatment | Radiation |
| PRC10 | 3E0K704 | Introduction of Liquid Brachytherapy Radioisotope into Genitourinary Tract, Via Natural or Artificial Opening | Treatment | Radiation |
| PRC10 | 3E0K804 | Introduction of Liquid Brachytherapy Radioisotope into Genitourinary Tract, Via Natural or Artificial Opening Endoscopic | Treatment | Radiation |
| PRC10 | 3E0L304 | Introduction of Liquid Brachytherapy Radioisotope into Pleural Cavity, Percutaneous Approach | Treatment | Radiation |
| PRC10 | 3E0L704 | Introduction of Liquid Brachytherapy Radioisotope into Pleural Cavity, Via Natural or Artificial Opening | Treatment | Radiation |
| PRC10 | 3E0M304 | Introduction of Liquid Brachytherapy Radioisotope into Peritoneal Cavity, Percutaneous Approach | Treatment | Radiation |
| PRC10 | 3E0M704 | Introduction of Liquid Brachytherapy Radioisotope into Peritoneal Cavity, Via Natural or Artificial Opening | Treatment | Radiation |
| PRC10 | 3E0N304 | Introduction of Liquid Brachytherapy Radioisotope into Male Reproductive, Percutaneous Approach | Treatment | Radiation |
| PRC10 | 3E0N704 | Introduction of Liquid Brachytherapy Radioisotope into Male Reproductive, Via Natural or Artificial Opening | Treatment | Radiation |
| PRC10 | 3E0N804 | Introduction of Liquid Brachytherapy Radioisotope into Male Reproductive, Via Natural or Artificial Opening Endoscopic | Treatment | Radiation |
| PRC10 | 3E0P304 | Introduction of Liquid Brachytherapy Radioisotope into Female Reproductive, Percutaneous Approach | Treatment | Radiation |
| PRC10 | 3E0P704 | Introduction of Liquid Brachytherapy Radioisotope into Female Reproductive, Via Natural or Artificial Opening | Treatment | Radiation |
| PRC10 | 3E0P804 | Introduction of Liquid Brachytherapy Radioisotope into Female Reproductive, Via Natural or Artificial Opening Endoscopic | Treatment | Radiation |
| PRC10 | 3E0Q304 | Introduction of Liquid Brachytherapy Radioisotope into Cranial Cavity and Brain, Percutaneous Approach | Treatment | Radiation |
| PRC10 | 3E0Q704 | Introduction of Liquid Brachytherapy Radioisotope into Cranial Cavity and Brain, Via Natural or Artificial Opening | Treatment | Radiation |
| PRC10 | 3E0R304 | Introduction of Liquid Brachytherapy Radioisotope into Spinal Canal, Percutaneous Approach | Treatment | Radiation |
| PRC10 | 3E0S304 | Introduction of Liquid Brachytherapy Radioisotope into Epidural Space, Percutaneous Approach | Treatment | Radiation |
| PRC10 | 3E0U304 | Introduction of Liquid Brachytherapy Radioisotope into Joints, Percutaneous Approach | Treatment | Radiation |
| PRC10 | 3E0Y304 | Introduction of Liquid Brachytherapy Radioisotope into Pericardial Cavity, Percutaneous Approach | Treatment | Radiation |
| PRC10 | 3E0Y704 | Introduction of Liquid Brachytherapy Radioisotope into Pericardial Cavity, Via Natural or Artificial Opening | Treatment | Radiation |
| PRC10 | CW70NZZ | Systemic Nuclear Medicine Therapy of Abdomen using Phosphorus 32 (P-32) | Treatment | Radiation |
| PRC10 | CW70YZZ | Systemic Nuclear Medicine Therapy of Abdomen using Other Radionuclide | Treatment | Radiation |
| PRC10 | CW73NZZ | Systemic Nuclear Medicine Therapy of Chest using Phosphorus 32 (P-32) | Treatment | Radiation |
| PRC10 | CW73YZZ | Systemic Nuclear Medicine Therapy of Chest using Other Radionuclide | Treatment | Radiation |
| PRC10 | CW7GYZZ | Systemic Nuclear Medicine Therapy of Thyroid using Other Radionuclide | Treatment | Radiation |
| PRC10 | CW7N8ZZ | Systemic Nuclear Medicine Therapy of Whole Body using Samarium 153 (Sm-153) | Treatment | Radiation |
| PRC10 | CW7NNZZ | Systemic Nuclear Medicine Therapy of Whole Body using Phosphorus 32 (P-32) | Treatment | Radiation |
| PRC10 | CW7NPZZ | Systemic Nuclear Medicine Therapy of Whole Body using Strontium 89 (Sr-89) | Treatment | Radiation |
| PRC10 | CW7NYZZ | Systemic Nuclear Medicine Therapy of Whole Body using Other Radionuclide | Treatment | Radiation |
| PRC10 | CW7YYZZ | Systemic Nuclear Medicine Therapy of Multiple Anatomical Regions using Other Radionuclide | Treatment | Radiation |
| PRC10 | D0000ZZ | Beam Radiation of Brain using Photons <1 MeV | Treatment | Radiation |
| PRC10 | D0003ZZ | Beam Radiation of Brain using Electrons | Treatment | Radiation |
| PRC10 | D0004ZZ | Beam Radiation of Brain using Heavy Particles (Protons,Ions) | Treatment | Radiation |
| PRC10 | D0005ZZ | Beam Radiation of Brain using Neutrons | Treatment | Radiation |
| PRC10 | D0010ZZ | Beam Radiation of Brain Stem using Photons <1 MeV | Treatment | Radiation |
| PRC10 | D0013ZZ | Beam Radiation of Brain Stem using Electrons | Treatment | Radiation |
| PRC10 | D0014ZZ | Beam Radiation of Brain Stem using Heavy Particles (Protons,Ions) | Treatment | Radiation |
| PRC10 | D0015ZZ | Beam Radiation of Brain Stem using Neutrons | Treatment | Radiation |
| PRC10 | D0060ZZ | Beam Radiation of Spinal Cord using Photons <1 MeV | Treatment | Radiation |
| PRC10 | D0063ZZ | Beam Radiation of Spinal Cord using Electrons | Treatment | Radiation |
| PRC10 | D0064ZZ | Beam Radiation of Spinal Cord using Heavy Particles (Protons,Ions) | Treatment | Radiation |
| PRC10 | D0065ZZ | Beam Radiation of Spinal Cord using Neutrons | Treatment | Radiation |
| PRC10 | D0070ZZ | Beam Radiation of Peripheral Nerve using Photons <1 MeV | Treatment | Radiation |
| PRC10 | D0073ZZ | Beam Radiation of Peripheral Nerve using Electrons | Treatment | Radiation |
| PRC10 | D0074ZZ | Beam Radiation of Peripheral Nerve using Heavy Particles (Protons,Ions) | Treatment | Radiation |
| PRC10 | D0075ZZ | Beam Radiation of Peripheral Nerve using Neutrons | Treatment | Radiation |
| PRC10 | D01097Z | High Dose Rate (HDR) Brachytherapy of Brain using Cesium 137 (Cs-137) | Treatment | Radiation |
| PRC10 | D01098Z | High Dose Rate (HDR) Brachytherapy of Brain using Iridium 192 (Ir-192) | Treatment | Radiation |
| PRC10 | D01099Z | High Dose Rate (HDR) Brachytherapy of Brain using Iodine 125 (I-125) | Treatment | Radiation |
| PRC10 | D0109BZ | High Dose Rate (HDR) Brachytherapy of Brain using Palladium 103 (Pd-103) | Treatment | Radiation |
| PRC10 | D0109CZ | High Dose Rate (HDR) Brachytherapy of Brain using Californium 252 (Cf-252) | Treatment | Radiation |
| PRC10 | D0109YZ | High Dose Rate (HDR) Brachytherapy of Brain using Other Isotope | Treatment | Radiation |
| PRC10 | D010B7Z | Low Dose Rate (LDR) Brachytherapy of Brain using Cesium 137 (Cs-137) | Treatment | Radiation |
| PRC10 | D010B8Z | Low Dose Rate (LDR) Brachytherapy of Brain using Iridium 192 (Ir-192) | Treatment | Radiation |
| PRC10 | D010B9Z | Low Dose Rate (LDR) Brachytherapy of Brain using Iodine 125 (I-125) | Treatment | Radiation |
| PRC10 | D010BBZ | Low Dose Rate (LDR) Brachytherapy of Brain using Palladium 103 (Pd-103) | Treatment | Radiation |
| PRC10 | D010BCZ | Low Dose Rate (LDR) Brachytherapy of Brain using Californium 252 (Cf-252) | Treatment | Radiation |
| PRC10 | D010BYZ | Low Dose Rate (LDR) Brachytherapy of Brain using Other Isotope | Treatment | Radiation |
| PRC10 | D01197Z | High Dose Rate (HDR) Brachytherapy of Brain Stem using Cesium 137 (Cs-137) | Treatment | Radiation |
| PRC10 | D01198Z | High Dose Rate (HDR) Brachytherapy of Brain Stem using Iridium 192 (Ir-192) | Treatment | Radiation |
| PRC10 | D01199Z | High Dose Rate (HDR) Brachytherapy of Brain Stem using Iodine 125 (I-125) | Treatment | Radiation |
| PRC10 | D0119BZ | High Dose Rate (HDR) Brachytherapy of Brain Stem using Palladium 103 (Pd-103) | Treatment | Radiation |
| PRC10 | D0119CZ | High Dose Rate (HDR) Brachytherapy of Brain Stem using Californium 252 (Cf-252) | Treatment | Radiation |
| PRC10 | D0119YZ | High Dose Rate (HDR) Brachytherapy of Brain Stem using Other Isotope | Treatment | Radiation |
| PRC10 | D011B7Z | Low Dose Rate (LDR) Brachytherapy of Brain Stem using Cesium 137 (Cs-137) | Treatment | Radiation |
| PRC10 | D011B8Z | Low Dose Rate (LDR) Brachytherapy of Brain Stem using Iridium 192 (Ir-192) | Treatment | Radiation |
| PRC10 | D011B9Z | Low Dose Rate (LDR) Brachytherapy of Brain Stem using Iodine 125 (I-125) | Treatment | Radiation |
| PRC10 | D011BBZ | Low Dose Rate (LDR) Brachytherapy of Brain Stem using Palladium 103 (Pd-103) | Treatment | Radiation |
| PRC10 | D011BCZ | Low Dose Rate (LDR) Brachytherapy of Brain Stem using Californium 252 (Cf-252) | Treatment | Radiation |
| PRC10 | D011BYZ | Low Dose Rate (LDR) Brachytherapy of Brain Stem using Other Isotope | Treatment | Radiation |
| PRC10 | D01697Z | High Dose Rate (HDR) Brachytherapy of Spinal Cord using Cesium 137 (Cs-137) | Treatment | Radiation |
| PRC10 | D01698Z | High Dose Rate (HDR) Brachytherapy of Spinal Cord using Iridium 192 (Ir-192) | Treatment | Radiation |
| PRC10 | D01699Z | High Dose Rate (HDR) Brachytherapy of Spinal Cord using Iodine 125 (I-125) | Treatment | Radiation |
| PRC10 | D0169BZ | High Dose Rate (HDR) Brachytherapy of Spinal Cord using Palladium 103 (Pd-103) | Treatment | Radiation |
| PRC10 | D0169CZ | High Dose Rate (HDR) Brachytherapy of Spinal Cord using Californium 252 (Cf-252) | Treatment | Radiation |
| PRC10 | D0169YZ | High Dose Rate (HDR) Brachytherapy of Spinal Cord using Other Isotope | Treatment | Radiation |
| PRC10 | D016B7Z | Low Dose Rate (LDR) Brachytherapy of Spinal Cord using Cesium 137 (Cs-137) | Treatment | Radiation |
| PRC10 | D016B8Z | Low Dose Rate (LDR) Brachytherapy of Spinal Cord using Iridium 192 (Ir-192) | Treatment | Radiation |
| PRC10 | D016B9Z | Low Dose Rate (LDR) Brachytherapy of Spinal Cord using Iodine 125 (I-125) | Treatment | Radiation |
| PRC10 | D016BBZ | Low Dose Rate (LDR) Brachytherapy of Spinal Cord using Palladium 103 (Pd-103) | Treatment | Radiation |
| PRC10 | D016BCZ | Low Dose Rate (LDR) Brachytherapy of Spinal Cord using Californium 252 (Cf-252) | Treatment | Radiation |
| PRC10 | D016BYZ | Low Dose Rate (LDR) Brachytherapy of Spinal Cord using Other Isotope | Treatment | Radiation |
| PRC10 | D01797Z | High Dose Rate (HDR) Brachytherapy of Peripheral Nerve using Cesium 137 (Cs-137) | Treatment | Radiation |
| PRC10 | D01798Z | High Dose Rate (HDR) Brachytherapy of Peripheral Nerve using Iridium 192 (Ir-192) | Treatment | Radiation |
| PRC10 | D01799Z | High Dose Rate (HDR) Brachytherapy of Peripheral Nerve using Iodine 125 (I-125) | Treatment | Radiation |
| PRC10 | D0179BZ | High Dose Rate (HDR) Brachytherapy of Peripheral Nerve using Palladium 103 (Pd-103) | Treatment | Radiation |
| PRC10 | D0179CZ | High Dose Rate (HDR) Brachytherapy of Peripheral Nerve using Californium 252 (Cf-252) | Treatment | Radiation |
| PRC10 | D0179YZ | High Dose Rate (HDR) Brachytherapy of Peripheral Nerve using Other Isotope | Treatment | Radiation |
| PRC10 | D017B7Z | Low Dose Rate (LDR) Brachytherapy of Peripheral Nerve using Cesium 137 (Cs-137) | Treatment | Radiation |
| PRC10 | D017B8Z | Low Dose Rate (LDR) Brachytherapy of Peripheral Nerve using Iridium 192 (Ir-192) | Treatment | Radiation |
| PRC10 | D017B9Z | Low Dose Rate (LDR) Brachytherapy of Peripheral Nerve using Iodine 125 (I-125) | Treatment | Radiation |
| PRC10 | D017BBZ | Low Dose Rate (LDR) Brachytherapy of Peripheral Nerve using Palladium 103 (Pd-103) | Treatment | Radiation |
| PRC10 | D017BCZ | Low Dose Rate (LDR) Brachytherapy of Peripheral Nerve using Californium 252 (Cf-252) | Treatment | Radiation |
| PRC10 | D017BYZ | Low Dose Rate (LDR) Brachytherapy of Peripheral Nerve using Other Isotope | Treatment | Radiation |
| PRC10 | D0Y07ZZ | Contact Radiation of Brain | Treatment | Radiation |
| PRC10 | D0Y0FZZ | Plaque Radiation of Brain | Treatment | Radiation |
| PRC10 | D0Y17ZZ | Contact Radiation of Brain Stem | Treatment | Radiation |
| PRC10 | D0Y1FZZ | Plaque Radiation of Brain Stem | Treatment | Radiation |
| PRC10 | D0Y67ZZ | Contact Radiation of Spinal Cord | Treatment | Radiation |
| PRC10 | D0Y6FZZ | Plaque Radiation of Spinal Cord | Treatment | Radiation |
| PRC10 | D0Y77ZZ | Contact Radiation of Peripheral Nerve | Treatment | Radiation |
| PRC10 | D0Y7FZZ | Plaque Radiation of Peripheral Nerve | Treatment | Radiation |
| PRC10 | D7000ZZ | Beam Radiation of Bone Marrow using Photons <1 MeV | Treatment | Radiation |
| PRC10 | D7003ZZ | Beam Radiation of Bone Marrow using Electrons | Treatment | Radiation |
| PRC10 | D7004ZZ | Beam Radiation of Bone Marrow using Heavy Particles (Protons,Ions) | Treatment | Radiation |
| PRC10 | D7005ZZ | Beam Radiation of Bone Marrow using Neutrons | Treatment | Radiation |
| PRC10 | D7010ZZ | Beam Radiation of Thymus using Photons <1 MeV | Treatment | Radiation |
| PRC10 | D7013ZZ | Beam Radiation of Thymus using Electrons | Treatment | Radiation |
| PRC10 | D7014ZZ | Beam Radiation of Thymus using Heavy Particles (Protons,Ions) | Treatment | Radiation |
| PRC10 | D7015ZZ | Beam Radiation of Thymus using Neutrons | Treatment | Radiation |
| PRC10 | D7020ZZ | Beam Radiation of Spleen using Photons <1 MeV | Treatment | Radiation |
| PRC10 | D7023ZZ | Beam Radiation of Spleen using Electrons | Treatment | Radiation |
| PRC10 | D7024ZZ | Beam Radiation of Spleen using Heavy Particles (Protons,Ions) | Treatment | Radiation |
| PRC10 | D7025ZZ | Beam Radiation of Spleen using Neutrons | Treatment | Radiation |
| PRC10 | D7030ZZ | Beam Radiation of Neck Lymphatics using Photons <1 MeV | Treatment | Radiation |
| PRC10 | D7033ZZ | Beam Radiation of Neck Lymphatics using Electrons | Treatment | Radiation |
| PRC10 | D7034ZZ | Beam Radiation of Neck Lymphatics using Heavy Particles (Protons,Ions) | Treatment | Radiation |
| PRC10 | D7035ZZ | Beam Radiation of Neck Lymphatics using Neutrons | Treatment | Radiation |
| PRC10 | D7040ZZ | Beam Radiation of Axillary Lymphatics using Photons <1 MeV | Treatment | Radiation |
| PRC10 | D7043ZZ | Beam Radiation of Axillary Lymphatics using Electrons | Treatment | Radiation |
| PRC10 | D7044ZZ | Beam Radiation of Axillary Lymphatics using Heavy Particles (Protons,Ions) | Treatment | Radiation |
| PRC10 | D7045ZZ | Beam Radiation of Axillary Lymphatics using Neutrons | Treatment | Radiation |
| PRC10 | D7050ZZ | Beam Radiation of Thorax Lymphatics using Photons <1 MeV | Treatment | Radiation |
| PRC10 | D7053ZZ | Beam Radiation of Thorax Lymphatics using Electrons | Treatment | Radiation |
| PRC10 | D7054ZZ | Beam Radiation of Thorax Lymphatics using Heavy Particles (Protons,Ions) | Treatment | Radiation |
| PRC10 | D7055ZZ | Beam Radiation of Thorax Lymphatics using Neutrons | Treatment | Radiation |
| PRC10 | D7060ZZ | Beam Radiation of Abdomen Lymphatics using Photons <1 MeV | Treatment | Radiation |
| PRC10 | D7063ZZ | Beam Radiation of Abdomen Lymphatics using Electrons | Treatment | Radiation |
| PRC10 | D7064ZZ | Beam Radiation of Abdomen Lymphatics using Heavy Particles (Protons,Ions) | Treatment | Radiation |
| PRC10 | D7065ZZ | Beam Radiation of Abdomen Lymphatics using Neutrons | Treatment | Radiation |
| PRC10 | D7070ZZ | Beam Radiation of Pelvis Lymphatics using Photons <1 MeV | Treatment | Radiation |
| PRC10 | D7073ZZ | Beam Radiation of Pelvis Lymphatics using Electrons | Treatment | Radiation |
| PRC10 | D7074ZZ | Beam Radiation of Pelvis Lymphatics using Heavy Particles (Protons,Ions) | Treatment | Radiation |
| PRC10 | D7075ZZ | Beam Radiation of Pelvis Lymphatics using Neutrons | Treatment | Radiation |
| PRC10 | D7080ZZ | Beam Radiation of Inguinal Lymphatics using Photons <1 MeV | Treatment | Radiation |
| PRC10 | D7083ZZ | Beam Radiation of Inguinal Lymphatics using Electrons | Treatment | Radiation |
| PRC10 | D7084ZZ | Beam Radiation of Inguinal Lymphatics using Heavy Particles (Protons,Ions) | Treatment | Radiation |
| PRC10 | D7085ZZ | Beam Radiation of Inguinal Lymphatics using Neutrons | Treatment | Radiation |
| PRC10 | D71097Z | High Dose Rate (HDR) Brachytherapy of Bone Marrow using Cesium 137 (Cs-137) | Treatment | Radiation |
| PRC10 | D71098Z | High Dose Rate (HDR) Brachytherapy of Bone Marrow using Iridium 192 (Ir-192) | Treatment | Radiation |
| PRC10 | D71099Z | High Dose Rate (HDR) Brachytherapy of Bone Marrow using Iodine 125 (I-125) | Treatment | Radiation |
| PRC10 | D7109BZ | High Dose Rate (HDR) Brachytherapy of Bone Marrow using Palladium 103 (Pd-103) | Treatment | Radiation |
| PRC10 | D7109CZ | High Dose Rate (HDR) Brachytherapy of Bone Marrow using Californium 252 (Cf-252) | Treatment | Radiation |
| PRC10 | D7109YZ | High Dose Rate (HDR) Brachytherapy of Bone Marrow using Other Isotope | Treatment | Radiation |
| PRC10 | D710B7Z | Low Dose Rate (LDR) Brachytherapy of Bone Marrow using Cesium 137 (Cs-137) | Treatment | Radiation |
| PRC10 | D710B8Z | Low Dose Rate (LDR) Brachytherapy of Bone Marrow using Iridium 192 (Ir-192) | Treatment | Radiation |
| PRC10 | D710B9Z | Low Dose Rate (LDR) Brachytherapy of Bone Marrow using Iodine 125 (I-125) | Treatment | Radiation |
| PRC10 | D710BBZ | Low Dose Rate (LDR) Brachytherapy of Bone Marrow using Palladium 103 (Pd-103) | Treatment | Radiation |
| PRC10 | D710BCZ | Low Dose Rate (LDR) Brachytherapy of Bone Marrow using Californium 252 (Cf-252) | Treatment | Radiation |
| PRC10 | D710BYZ | Low Dose Rate (LDR) Brachytherapy of Bone Marrow using Other Isotope | Treatment | Radiation |
| PRC10 | D71197Z | High Dose Rate (HDR) Brachytherapy of Thymus using Cesium 137 (Cs-137) | Treatment | Radiation |
| PRC10 | D71198Z | High Dose Rate (HDR) Brachytherapy of Thymus using Iridium 192 (Ir-192) | Treatment | Radiation |
| PRC10 | D71199Z | High Dose Rate (HDR) Brachytherapy of Thymus using Iodine 125 (I-125) | Treatment | Radiation |
| PRC10 | D7119BZ | High Dose Rate (HDR) Brachytherapy of Thymus using Palladium 103 (Pd-103) | Treatment | Radiation |
| PRC10 | D7119CZ | High Dose Rate (HDR) Brachytherapy of Thymus using Californium 252 (Cf-252) | Treatment | Radiation |
| PRC10 | D7119YZ | High Dose Rate (HDR) Brachytherapy of Thymus using Other Isotope | Treatment | Radiation |
| PRC10 | D711B7Z | Low Dose Rate (LDR) Brachytherapy of Thymus using Cesium 137 (Cs-137) | Treatment | Radiation |
| PRC10 | D711B8Z | Low Dose Rate (LDR) Brachytherapy of Thymus using Iridium 192 (Ir-192) | Treatment | Radiation |
| PRC10 | D711B9Z | Low Dose Rate (LDR) Brachytherapy of Thymus using Iodine 125 (I-125) | Treatment | Radiation |
| PRC10 | D711BBZ | Low Dose Rate (LDR) Brachytherapy of Thymus using Palladium 103 (Pd-103) | Treatment | Radiation |
| PRC10 | D711BCZ | Low Dose Rate (LDR) Brachytherapy of Thymus using Californium 252 (Cf-252) | Treatment | Radiation |
| PRC10 | D711BYZ | Low Dose Rate (LDR) Brachytherapy of Thymus using Other Isotope | Treatment | Radiation |
| PRC10 | D71297Z | High Dose Rate (HDR) Brachytherapy of Spleen using Cesium 137 (Cs-137) | Treatment | Radiation |
| PRC10 | D71298Z | High Dose Rate (HDR) Brachytherapy of Spleen using Iridium 192 (Ir-192) | Treatment | Radiation |
| PRC10 | D71299Z | High Dose Rate (HDR) Brachytherapy of Spleen using Iodine 125 (I-125) | Treatment | Radiation |
| PRC10 | D7129BZ | High Dose Rate (HDR) Brachytherapy of Spleen using Palladium 103 (Pd-103) | Treatment | Radiation |
| PRC10 | D7129CZ | High Dose Rate (HDR) Brachytherapy of Spleen using Californium 252 (Cf-252) | Treatment | Radiation |
| PRC10 | D7129YZ | High Dose Rate (HDR) Brachytherapy of Spleen using Other Isotope | Treatment | Radiation |
| PRC10 | D712B7Z | Low Dose Rate (LDR) Brachytherapy of Spleen using Cesium 137 (Cs-137) | Treatment | Radiation |
| PRC10 | D712B8Z | Low Dose Rate (LDR) Brachytherapy of Spleen using Iridium 192 (Ir-192) | Treatment | Radiation |
| PRC10 | D712B9Z | Low Dose Rate (LDR) Brachytherapy of Spleen using Iodine 125 (I-125) | Treatment | Radiation |
| PRC10 | D712BBZ | Low Dose Rate (LDR) Brachytherapy of Spleen using Palladium 103 (Pd-103) | Treatment | Radiation |
| PRC10 | D712BCZ | Low Dose Rate (LDR) Brachytherapy of Spleen using Californium 252 (Cf-252) | Treatment | Radiation |
| PRC10 | D712BYZ | Low Dose Rate (LDR) Brachytherapy of Spleen using Other Isotope | Treatment | Radiation |
| PRC10 | D71397Z | High Dose Rate (HDR) Brachytherapy of Neck Lymphatics using Cesium 137 (Cs-137) | Treatment | Radiation |
| PRC10 | D71398Z | High Dose Rate (HDR) Brachytherapy of Neck Lymphatics using Iridium 192 (Ir-192) | Treatment | Radiation |
| PRC10 | D71399Z | High Dose Rate (HDR) Brachytherapy of Neck Lymphatics using Iodine 125 (I-125) | Treatment | Radiation |
| PRC10 | D7139BZ | High Dose Rate (HDR) Brachytherapy of Neck Lymphatics using Palladium 103 (Pd-103) | Treatment | Radiation |
| PRC10 | D7139CZ | High Dose Rate (HDR) Brachytherapy of Neck Lymphatics using Californium 252 (Cf-252) | Treatment | Radiation |
| PRC10 | D7139YZ | High Dose Rate (HDR) Brachytherapy of Neck Lymphatics using Other Isotope | Treatment | Radiation |
| PRC10 | D713B7Z | Low Dose Rate (LDR) Brachytherapy of Neck Lymphatics using Cesium 137 (Cs-137) | Treatment | Radiation |
| PRC10 | D713B8Z | Low Dose Rate (LDR) Brachytherapy of Neck Lymphatics using Iridium 192 (Ir-192) | Treatment | Radiation |
| PRC10 | D713B9Z | Low Dose Rate (LDR) Brachytherapy of Neck Lymphatics using Iodine 125 (I-125) | Treatment | Radiation |
| PRC10 | D713BBZ | Low Dose Rate (LDR) Brachytherapy of Neck Lymphatics using Palladium 103 (Pd-103) | Treatment | Radiation |
| PRC10 | D713BCZ | Low Dose Rate (LDR) Brachytherapy of Neck Lymphatics using Californium 252 (Cf-252) | Treatment | Radiation |
| PRC10 | D713BYZ | Low Dose Rate (LDR) Brachytherapy of Neck Lymphatics using Other Isotope | Treatment | Radiation |
| PRC10 | D71497Z | High Dose Rate (HDR) Brachytherapy of Axillary Lymphatics using Cesium 137 (Cs-137) | Treatment | Radiation |
| PRC10 | D71498Z | High Dose Rate (HDR) Brachytherapy of Axillary Lymphatics using Iridium 192 (Ir-192) | Treatment | Radiation |
| PRC10 | D71499Z | High Dose Rate (HDR) Brachytherapy of Axillary Lymphatics using Iodine 125 (I-125) | Treatment | Radiation |
| PRC10 | D7149BZ | High Dose Rate (HDR) Brachytherapy of Axillary Lymphatics using Palladium 103 (Pd-103) | Treatment | Radiation |
| PRC10 | D7149CZ | High Dose Rate (HDR) Brachytherapy of Axillary Lymphatics using Californium 252 (Cf-252) | Treatment | Radiation |
| PRC10 | D7149YZ | High Dose Rate (HDR) Brachytherapy of Axillary Lymphatics using Other Isotope | Treatment | Radiation |
| PRC10 | D714B7Z | Low Dose Rate (LDR) Brachytherapy of Axillary Lymphatics using Cesium 137 (Cs-137) | Treatment | Radiation |
| PRC10 | D714B8Z | Low Dose Rate (LDR) Brachytherapy of Axillary Lymphatics using Iridium 192 (Ir-192) | Treatment | Radiation |
| PRC10 | D714B9Z | Low Dose Rate (LDR) Brachytherapy of Axillary Lymphatics using Iodine 125 (I-125) | Treatment | Radiation |
| PRC10 | D714BBZ | Low Dose Rate (LDR) Brachytherapy of Axillary Lymphatics using Palladium 103 (Pd-103) | Treatment | Radiation |
| PRC10 | D714BCZ | Low Dose Rate (LDR) Brachytherapy of Axillary Lymphatics using Californium 252 (Cf-252) | Treatment | Radiation |
| PRC10 | D714BYZ | Low Dose Rate (LDR) Brachytherapy of Axillary Lymphatics using Other Isotope | Treatment | Radiation |
| PRC10 | D71597Z | High Dose Rate (HDR) Brachytherapy of Thorax Lymphatics using Cesium 137 (Cs-137) | Treatment | Radiation |
| PRC10 | D71598Z | High Dose Rate (HDR) Brachytherapy of Thorax Lymphatics using Iridium 192 (Ir-192) | Treatment | Radiation |
| PRC10 | D71599Z | High Dose Rate (HDR) Brachytherapy of Thorax Lymphatics using Iodine 125 (I-125) | Treatment | Radiation |
| PRC10 | D7159BZ | High Dose Rate (HDR) Brachytherapy of Thorax Lymphatics using Palladium 103 (Pd-103) | Treatment | Radiation |
| PRC10 | D7159CZ | High Dose Rate (HDR) Brachytherapy of Thorax Lymphatics using Californium 252 (Cf-252) | Treatment | Radiation |
| PRC10 | D7159YZ | High Dose Rate (HDR) Brachytherapy of Thorax Lymphatics using Other Isotope | Treatment | Radiation |
| PRC10 | D715B7Z | Low Dose Rate (LDR) Brachytherapy of Thorax Lymphatics using Cesium 137 (Cs-137) | Treatment | Radiation |
| PRC10 | D715B8Z | Low Dose Rate (LDR) Brachytherapy of Thorax Lymphatics using Iridium 192 (Ir-192) | Treatment | Radiation |
| PRC10 | D715B9Z | Low Dose Rate (LDR) Brachytherapy of Thorax Lymphatics using Iodine 125 (I-125) | Treatment | Radiation |
| PRC10 | D715BBZ | Low Dose Rate (LDR) Brachytherapy of Thorax Lymphatics using Palladium 103 (Pd-103) | Treatment | Radiation |
| PRC10 | D715BCZ | Low Dose Rate (LDR) Brachytherapy of Thorax Lymphatics using Californium 252 (Cf-252) | Treatment | Radiation |
| PRC10 | D715BYZ | Low Dose Rate (LDR) Brachytherapy of Thorax Lymphatics using Other Isotope | Treatment | Radiation |
| PRC10 | D71697Z | High Dose Rate (HDR) Brachytherapy of Abdomen Lymphatics using Cesium 137 (Cs-137) | Treatment | Radiation |
| PRC10 | D71698Z | High Dose Rate (HDR) Brachytherapy of Abdomen Lymphatics using Iridium 192 (Ir-192) | Treatment | Radiation |
| PRC10 | D71699Z | High Dose Rate (HDR) Brachytherapy of Abdomen Lymphatics using Iodine 125 (I-125) | Treatment | Radiation |
| PRC10 | D7169BZ | High Dose Rate (HDR) Brachytherapy of Abdomen Lymphatics using Palladium 103 (Pd-103) | Treatment | Radiation |
| PRC10 | D7169CZ | High Dose Rate (HDR) Brachytherapy of Abdomen Lymphatics using Californium 252 (Cf-252) | Treatment | Radiation |
| PRC10 | D7169YZ | High Dose Rate (HDR) Brachytherapy of Abdomen Lymphatics using Other Isotope | Treatment | Radiation |
| PRC10 | D716B7Z | Low Dose Rate (LDR) Brachytherapy of Abdomen Lymphatics using Cesium 137 (Cs-137) | Treatment | Radiation |
| PRC10 | D716B8Z | Low Dose Rate (LDR) Brachytherapy of Abdomen Lymphatics using Iridium 192 (Ir-192) | Treatment | Radiation |
| PRC10 | D716B9Z | Low Dose Rate (LDR) Brachytherapy of Abdomen Lymphatics using Iodine 125 (I-125) | Treatment | Radiation |
| PRC10 | D716BBZ | Low Dose Rate (LDR) Brachytherapy of Abdomen Lymphatics using Palladium 103 (Pd-103) | Treatment | Radiation |
| PRC10 | D716BCZ | Low Dose Rate (LDR) Brachytherapy of Abdomen Lymphatics using Californium 252 (Cf-252) | Treatment | Radiation |
| PRC10 | D716BYZ | Low Dose Rate (LDR) Brachytherapy of Abdomen Lymphatics using Other Isotope | Treatment | Radiation |
| PRC10 | D71797Z | High Dose Rate (HDR) Brachytherapy of Pelvis Lymphatics using Cesium 137 (Cs-137) | Treatment | Radiation |
| PRC10 | D71798Z | High Dose Rate (HDR) Brachytherapy of Pelvis Lymphatics using Iridium 192 (Ir-192) | Treatment | Radiation |
| PRC10 | D71799Z | High Dose Rate (HDR) Brachytherapy of Pelvis Lymphatics using Iodine 125 (I-125) | Treatment | Radiation |
| PRC10 | D7179BZ | High Dose Rate (HDR) Brachytherapy of Pelvis Lymphatics using Palladium 103 (Pd-103) | Treatment | Radiation |
| PRC10 | D7179CZ | High Dose Rate (HDR) Brachytherapy of Pelvis Lymphatics using Californium 252 (Cf-252) | Treatment | Radiation |
| PRC10 | D7179YZ | High Dose Rate (HDR) Brachytherapy of Pelvis Lymphatics using Other Isotope | Treatment | Radiation |
| PRC10 | D717B7Z | Low Dose Rate (LDR) Brachytherapy of Pelvis Lymphatics using Cesium 137 (Cs-137) | Treatment | Radiation |
| PRC10 | D717B8Z | Low Dose Rate (LDR) Brachytherapy of Pelvis Lymphatics using Iridium 192 (Ir-192) | Treatment | Radiation |
| PRC10 | D717B9Z | Low Dose Rate (LDR) Brachytherapy of Pelvis Lymphatics using Iodine 125 (I-125) | Treatment | Radiation |
| PRC10 | D717BBZ | Low Dose Rate (LDR) Brachytherapy of Pelvis Lymphatics using Palladium 103 (Pd-103) | Treatment | Radiation |
| PRC10 | D717BCZ | Low Dose Rate (LDR) Brachytherapy of Pelvis Lymphatics using Californium 252 (Cf-252) | Treatment | Radiation |
| PRC10 | D717BYZ | Low Dose Rate (LDR) Brachytherapy of Pelvis Lymphatics using Other Isotope | Treatment | Radiation |
| PRC10 | D71897Z | High Dose Rate (HDR) Brachytherapy of Inguinal Lymphatics using Cesium 137 (Cs-137) | Treatment | Radiation |
| PRC10 | D71898Z | High Dose Rate (HDR) Brachytherapy of Inguinal Lymphatics using Iridium 192 (Ir-192) | Treatment | Radiation |
| PRC10 | D71899Z | High Dose Rate (HDR) Brachytherapy of Inguinal Lymphatics using Iodine 125 (I-125) | Treatment | Radiation |
| PRC10 | D7189BZ | High Dose Rate (HDR) Brachytherapy of Inguinal Lymphatics using Palladium 103 (Pd-103) | Treatment | Radiation |
| PRC10 | D7189CZ | High Dose Rate (HDR) Brachytherapy of Inguinal Lymphatics using Californium 252 (Cf-252) | Treatment | Radiation |
| PRC10 | D7189YZ | High Dose Rate (HDR) Brachytherapy of Inguinal Lymphatics using Other Isotope | Treatment | Radiation |
| PRC10 | D718B7Z | Low Dose Rate (LDR) Brachytherapy of Inguinal Lymphatics using Cesium 137 (Cs-137) | Treatment | Radiation |
| PRC10 | D718B8Z | Low Dose Rate (LDR) Brachytherapy of Inguinal Lymphatics using Iridium 192 (Ir-192) | Treatment | Radiation |
| PRC10 | D718B9Z | Low Dose Rate (LDR) Brachytherapy of Inguinal Lymphatics using Iodine 125 (I-125) | Treatment | Radiation |
| PRC10 | D718BBZ | Low Dose Rate (LDR) Brachytherapy of Inguinal Lymphatics using Palladium 103 (Pd-103) | Treatment | Radiation |
| PRC10 | D718BCZ | Low Dose Rate (LDR) Brachytherapy of Inguinal Lymphatics using Californium 252 (Cf-252) | Treatment | Radiation |
| PRC10 | D718BYZ | Low Dose Rate (LDR) Brachytherapy of Inguinal Lymphatics using Other Isotope | Treatment | Radiation |
| PRC10 | D7Y0FZZ | Plaque Radiation of Bone Marrow | Treatment | Radiation |
| PRC10 | D7Y1FZZ | Plaque Radiation of Thymus | Treatment | Radiation |
| PRC10 | D7Y2FZZ | Plaque Radiation of Spleen | Treatment | Radiation |
| PRC10 | D7Y3FZZ | Plaque Radiation of Neck Lymphatics | Treatment | Radiation |
| PRC10 | D7Y4FZZ | Plaque Radiation of Axillary Lymphatics | Treatment | Radiation |
| PRC10 | D7Y5FZZ | Plaque Radiation of Thorax Lymphatics | Treatment | Radiation |
| PRC10 | D7Y6FZZ | Plaque Radiation of Abdomen Lymphatics | Treatment | Radiation |
| PRC10 | D7Y7FZZ | Plaque Radiation of Pelvis Lymphatics | Treatment | Radiation |
| PRC10 | D7Y8FZZ | Plaque Radiation of Inguinal Lymphatics | Treatment | Radiation |
| PRC10 | D8000ZZ | Beam Radiation of Eye using Photons <1 MeV | Treatment | Radiation |
| PRC10 | D8003ZZ | Beam Radiation of Eye using Electrons | Treatment | Radiation |
| PRC10 | D8004ZZ | Beam Radiation of Eye using Heavy Particles (Protons,Ions) | Treatment | Radiation |
| PRC10 | D8005ZZ | Beam Radiation of Eye using Neutrons | Treatment | Radiation |
| PRC10 | D81097Z | High Dose Rate (HDR) Brachytherapy of Eye using Cesium 137 (Cs-137) | Treatment | Radiation |
| PRC10 | D81098Z | High Dose Rate (HDR) Brachytherapy of Eye using Iridium 192 (Ir-192) | Treatment | Radiation |
| PRC10 | D81099Z | High Dose Rate (HDR) Brachytherapy of Eye using Iodine 125 (I-125) | Treatment | Radiation |
| PRC10 | D8109BZ | High Dose Rate (HDR) Brachytherapy of Eye using Palladium 103 (Pd-103) | Treatment | Radiation |
| PRC10 | D8109CZ | High Dose Rate (HDR) Brachytherapy of Eye using Californium 252 (Cf-252) | Treatment | Radiation |
| PRC10 | D8109YZ | High Dose Rate (HDR) Brachytherapy of Eye using Other Isotope | Treatment | Radiation |
| PRC10 | D810B7Z | Low Dose Rate (LDR) Brachytherapy of Eye using Cesium 137 (Cs-137) | Treatment | Radiation |
| PRC10 | D810B8Z | Low Dose Rate (LDR) Brachytherapy of Eye using Iridium 192 (Ir-192) | Treatment | Radiation |
| PRC10 | D810B9Z | Low Dose Rate (LDR) Brachytherapy of Eye using Iodine 125 (I-125) | Treatment | Radiation |
| PRC10 | D810BBZ | Low Dose Rate (LDR) Brachytherapy of Eye using Palladium 103 (Pd-103) | Treatment | Radiation |
| PRC10 | D810BCZ | Low Dose Rate (LDR) Brachytherapy of Eye using Californium 252 (Cf-252) | Treatment | Radiation |
| PRC10 | D810BYZ | Low Dose Rate (LDR) Brachytherapy of Eye using Other Isotope | Treatment | Radiation |
| PRC10 | D8Y07ZZ | Contact Radiation of Eye | Treatment | Radiation |
| PRC10 | D8Y0FZZ | Plaque Radiation of Eye | Treatment | Radiation |
| PRC10 | D9000ZZ | Beam Radiation of Ear using Photons <1 MeV | Treatment | Radiation |
| PRC10 | D9003ZZ | Beam Radiation of Ear using Electrons | Treatment | Radiation |
| PRC10 | D9004ZZ | Beam Radiation of Ear using Heavy Particles (Protons,Ions) | Treatment | Radiation |
| PRC10 | D9005ZZ | Beam Radiation of Ear using Neutrons | Treatment | Radiation |
| PRC10 | D9010ZZ | Beam Radiation of Nose using Photons <1 MeV | Treatment | Radiation |
| PRC10 | D9013ZZ | Beam Radiation of Nose using Electrons | Treatment | Radiation |
| PRC10 | D9014ZZ | Beam Radiation of Nose using Heavy Particles (Protons,Ions) | Treatment | Radiation |
| PRC10 | D9015ZZ | Beam Radiation of Nose using Neutrons | Treatment | Radiation |
| PRC10 | D9030ZZ | Beam Radiation of Hypopharynx using Photons <1 MeV | Treatment | Radiation |
| PRC10 | D9033ZZ | Beam Radiation of Hypopharynx using Electrons | Treatment | Radiation |
| PRC10 | D9034ZZ | Beam Radiation of Hypopharynx using Heavy Particles (Protons,Ions) | Treatment | Radiation |
| PRC10 | D9035ZZ | Beam Radiation of Hypopharynx using Neutrons | Treatment | Radiation |
| PRC10 | D9040ZZ | Beam Radiation of Mouth using Photons <1 MeV | Treatment | Radiation |
| PRC10 | D9043ZZ | Beam Radiation of Mouth using Electrons | Treatment | Radiation |
| PRC10 | D9044ZZ | Beam Radiation of Mouth using Heavy Particles (Protons,Ions) | Treatment | Radiation |
| PRC10 | D9045ZZ | Beam Radiation of Mouth using Neutrons | Treatment | Radiation |
| PRC10 | D9050ZZ | Beam Radiation of Tongue using Photons <1 MeV | Treatment | Radiation |
| PRC10 | D9053ZZ | Beam Radiation of Tongue using Electrons | Treatment | Radiation |
| PRC10 | D9054ZZ | Beam Radiation of Tongue using Heavy Particles (Protons,Ions) | Treatment | Radiation |
| PRC10 | D9055ZZ | Beam Radiation of Tongue using Neutrons | Treatment | Radiation |
| PRC10 | D9060ZZ | Beam Radiation of Salivary Glands using Photons <1 MeV | Treatment | Radiation |
| PRC10 | D9063ZZ | Beam Radiation of Salivary Glands using Electrons | Treatment | Radiation |
| PRC10 | D9064ZZ | Beam Radiation of Salivary Glands using Heavy Particles (Protons,Ions) | Treatment | Radiation |
| PRC10 | D9065ZZ | Beam Radiation of Salivary Glands using Neutrons | Treatment | Radiation |
| PRC10 | D9070ZZ | Beam Radiation of Sinuses using Photons <1 MeV | Treatment | Radiation |
| PRC10 | D9073ZZ | Beam Radiation of Sinuses using Electrons | Treatment | Radiation |
| PRC10 | D9074ZZ | Beam Radiation of Sinuses using Heavy Particles (Protons,Ions) | Treatment | Radiation |
| PRC10 | D9075ZZ | Beam Radiation of Sinuses using Neutrons | Treatment | Radiation |
| PRC10 | D9080ZZ | Beam Radiation of Hard Palate using Photons <1 MeV | Treatment | Radiation |
| PRC10 | D9083ZZ | Beam Radiation of Hard Palate using Electrons | Treatment | Radiation |
| PRC10 | D9084ZZ | Beam Radiation of Hard Palate using Heavy Particles (Protons,Ions) | Treatment | Radiation |
| PRC10 | D9085ZZ | Beam Radiation of Hard Palate using Neutrons | Treatment | Radiation |
| PRC10 | D9090ZZ | Beam Radiation of Soft Palate using Photons <1 MeV | Treatment | Radiation |
| PRC10 | D9093ZZ | Beam Radiation of Soft Palate using Electrons | Treatment | Radiation |
| PRC10 | D9094ZZ | Beam Radiation of Soft Palate using Heavy Particles (Protons,Ions) | Treatment | Radiation |
| PRC10 | D9095ZZ | Beam Radiation of Soft Palate using Neutrons | Treatment | Radiation |
| PRC10 | D90B0ZZ | Beam Radiation of Larynx using Photons <1 MeV | Treatment | Radiation |
| PRC10 | D90B3ZZ | Beam Radiation of Larynx using Electrons | Treatment | Radiation |
| PRC10 | D90B4ZZ | Beam Radiation of Larynx using Heavy Particles (Protons,Ions) | Treatment | Radiation |
| PRC10 | D90B5ZZ | Beam Radiation of Larynx using Neutrons | Treatment | Radiation |
| PRC10 | D90D0ZZ | Beam Radiation of Nasopharynx using Photons <1 MeV | Treatment | Radiation |
| PRC10 | D90D3ZZ | Beam Radiation of Nasopharynx using Electrons | Treatment | Radiation |
| PRC10 | D90D4ZZ | Beam Radiation of Nasopharynx using Heavy Particles (Protons,Ions) | Treatment | Radiation |
| PRC10 | D90D5ZZ | Beam Radiation of Nasopharynx using Neutrons | Treatment | Radiation |
| PRC10 | D90F0ZZ | Beam Radiation of Oropharynx using Photons <1 MeV | Treatment | Radiation |
| PRC10 | D90F3ZZ | Beam Radiation of Oropharynx using Electrons | Treatment | Radiation |
| PRC10 | D90F4ZZ | Beam Radiation of Oropharynx using Heavy Particles (Protons,Ions) | Treatment | Radiation |
| PRC10 | D90F5ZZ | Beam Radiation of Oropharynx using Neutrons | Treatment | Radiation |
| PRC10 | D91097Z | High Dose Rate (HDR) Brachytherapy of Ear using Cesium 137 (Cs-137) | Treatment | Radiation |
| PRC10 | D91098Z | High Dose Rate (HDR) Brachytherapy of Ear using Iridium 192 (Ir-192) | Treatment | Radiation |
| PRC10 | D91099Z | High Dose Rate (HDR) Brachytherapy of Ear using Iodine 125 (I-125) | Treatment | Radiation |
| PRC10 | D9109BZ | High Dose Rate (HDR) Brachytherapy of Ear using Palladium 103 (Pd-103) | Treatment | Radiation |
| PRC10 | D9109CZ | High Dose Rate (HDR) Brachytherapy of Ear using Californium 252 (Cf-252) | Treatment | Radiation |
| PRC10 | D9109YZ | High Dose Rate (HDR) Brachytherapy of Ear using Other Isotope | Treatment | Radiation |
| PRC10 | D910B7Z | Low Dose Rate (LDR) Brachytherapy of Ear using Cesium 137 (Cs-137) | Treatment | Radiation |
| PRC10 | D910B8Z | Low Dose Rate (LDR) Brachytherapy of Ear using Iridium 192 (Ir-192) | Treatment | Radiation |
| PRC10 | D910B9Z | Low Dose Rate (LDR) Brachytherapy of Ear using Iodine 125 (I-125) | Treatment | Radiation |
| PRC10 | D910BBZ | Low Dose Rate (LDR) Brachytherapy of Ear using Palladium 103 (Pd-103) | Treatment | Radiation |
| PRC10 | D910BCZ | Low Dose Rate (LDR) Brachytherapy of Ear using Californium 252 (Cf-252) | Treatment | Radiation |
| PRC10 | D910BYZ | Low Dose Rate (LDR) Brachytherapy of Ear using Other Isotope | Treatment | Radiation |
| PRC10 | D91197Z | High Dose Rate (HDR) Brachytherapy of Nose using Cesium 137 (Cs-137) | Treatment | Radiation |
| PRC10 | D91198Z | High Dose Rate (HDR) Brachytherapy of Nose using Iridium 192 (Ir-192) | Treatment | Radiation |
| PRC10 | D91199Z | High Dose Rate (HDR) Brachytherapy of Nose using Iodine 125 (I-125) | Treatment | Radiation |
| PRC10 | D9119BZ | High Dose Rate (HDR) Brachytherapy of Nose using Palladium 103 (Pd-103) | Treatment | Radiation |
| PRC10 | D9119CZ | High Dose Rate (HDR) Brachytherapy of Nose using Californium 252 (Cf-252) | Treatment | Radiation |
| PRC10 | D9119YZ | High Dose Rate (HDR) Brachytherapy of Nose using Other Isotope | Treatment | Radiation |
| PRC10 | D911B7Z | Low Dose Rate (LDR) Brachytherapy of Nose using Cesium 137 (Cs-137) | Treatment | Radiation |
| PRC10 | D911B8Z | Low Dose Rate (LDR) Brachytherapy of Nose using Iridium 192 (Ir-192) | Treatment | Radiation |
| PRC10 | D911B9Z | Low Dose Rate (LDR) Brachytherapy of Nose using Iodine 125 (I-125) | Treatment | Radiation |
| PRC10 | D911BBZ | Low Dose Rate (LDR) Brachytherapy of Nose using Palladium 103 (Pd-103) | Treatment | Radiation |
| PRC10 | D911BCZ | Low Dose Rate (LDR) Brachytherapy of Nose using Californium 252 (Cf-252) | Treatment | Radiation |
| PRC10 | D911BYZ | Low Dose Rate (LDR) Brachytherapy of Nose using Other Isotope | Treatment | Radiation |
| PRC10 | D91397Z | High Dose Rate (HDR) Brachytherapy of Hypopharynx using Cesium 137 (Cs-137) | Treatment | Radiation |
| PRC10 | D91398Z | High Dose Rate (HDR) Brachytherapy of Hypopharynx using Iridium 192 (Ir-192) | Treatment | Radiation |
| PRC10 | D91399Z | High Dose Rate (HDR) Brachytherapy of Hypopharynx using Iodine 125 (I-125) | Treatment | Radiation |
| PRC10 | D9139BZ | High Dose Rate (HDR) Brachytherapy of Hypopharynx using Palladium 103 (Pd-103) | Treatment | Radiation |
| PRC10 | D9139CZ | High Dose Rate (HDR) Brachytherapy of Hypopharynx using Californium 252 (Cf-252) | Treatment | Radiation |
| PRC10 | D9139YZ | High Dose Rate (HDR) Brachytherapy of Hypopharynx using Other Isotope | Treatment | Radiation |
| PRC10 | D913B7Z | Low Dose Rate (LDR) Brachytherapy of Hypopharynx using Cesium 137 (Cs-137) | Treatment | Radiation |
| PRC10 | D913B8Z | Low Dose Rate (LDR) Brachytherapy of Hypopharynx using Iridium 192 (Ir-192) | Treatment | Radiation |
| PRC10 | D913B9Z | Low Dose Rate (LDR) Brachytherapy of Hypopharynx using Iodine 125 (I-125) | Treatment | Radiation |
| PRC10 | D913BBZ | Low Dose Rate (LDR) Brachytherapy of Hypopharynx using Palladium 103 (Pd-103) | Treatment | Radiation |
| PRC10 | D913BCZ | Low Dose Rate (LDR) Brachytherapy of Hypopharynx using Californium 252 (Cf-252) | Treatment | Radiation |
| PRC10 | D913BYZ | Low Dose Rate (LDR) Brachytherapy of Hypopharynx using Other Isotope | Treatment | Radiation |
| PRC10 | D91497Z | High Dose Rate (HDR) Brachytherapy of Mouth using Cesium 137 (Cs-137) | Treatment | Radiation |
| PRC10 | D91498Z | High Dose Rate (HDR) Brachytherapy of Mouth using Iridium 192 (Ir-192) | Treatment | Radiation |
| PRC10 | D91499Z | High Dose Rate (HDR) Brachytherapy of Mouth using Iodine 125 (I-125) | Treatment | Radiation |
| PRC10 | D9149BZ | High Dose Rate (HDR) Brachytherapy of Mouth using Palladium 103 (Pd-103) | Treatment | Radiation |
| PRC10 | D9149CZ | High Dose Rate (HDR) Brachytherapy of Mouth using Californium 252 (Cf-252) | Treatment | Radiation |
| PRC10 | D9149YZ | High Dose Rate (HDR) Brachytherapy of Mouth using Other Isotope | Treatment | Radiation |
| PRC10 | D914B7Z | Low Dose Rate (LDR) Brachytherapy of Mouth using Cesium 137 (Cs-137) | Treatment | Radiation |
| PRC10 | D914B8Z | Low Dose Rate (LDR) Brachytherapy of Mouth using Iridium 192 (Ir-192) | Treatment | Radiation |
| PRC10 | D914B9Z | Low Dose Rate (LDR) Brachytherapy of Mouth using Iodine 125 (I-125) | Treatment | Radiation |
| PRC10 | D914BBZ | Low Dose Rate (LDR) Brachytherapy of Mouth using Palladium 103 (Pd-103) | Treatment | Radiation |
| PRC10 | D914BCZ | Low Dose Rate (LDR) Brachytherapy of Mouth using Californium 252 (Cf-252) | Treatment | Radiation |
| PRC10 | D914BYZ | Low Dose Rate (LDR) Brachytherapy of Mouth using Other Isotope | Treatment | Radiation |
| PRC10 | D91597Z | High Dose Rate (HDR) Brachytherapy of Tongue using Cesium 137 (Cs-137) | Treatment | Radiation |
| PRC10 | D91598Z | High Dose Rate (HDR) Brachytherapy of Tongue using Iridium 192 (Ir-192) | Treatment | Radiation |
| PRC10 | D91599Z | High Dose Rate (HDR) Brachytherapy of Tongue using Iodine 125 (I-125) | Treatment | Radiation |
| PRC10 | D9159BZ | High Dose Rate (HDR) Brachytherapy of Tongue using Palladium 103 (Pd-103) | Treatment | Radiation |
| PRC10 | D9159CZ | High Dose Rate (HDR) Brachytherapy of Tongue using Californium 252 (Cf-252) | Treatment | Radiation |
| PRC10 | D9159YZ | High Dose Rate (HDR) Brachytherapy of Tongue using Other Isotope | Treatment | Radiation |
| PRC10 | D915B7Z | Low Dose Rate (LDR) Brachytherapy of Tongue using Cesium 137 (Cs-137) | Treatment | Radiation |
| PRC10 | D915B8Z | Low Dose Rate (LDR) Brachytherapy of Tongue using Iridium 192 (Ir-192) | Treatment | Radiation |
| PRC10 | D915B9Z | Low Dose Rate (LDR) Brachytherapy of Tongue using Iodine 125 (I-125) | Treatment | Radiation |
| PRC10 | D915BBZ | Low Dose Rate (LDR) Brachytherapy of Tongue using Palladium 103 (Pd-103) | Treatment | Radiation |
| PRC10 | D915BCZ | Low Dose Rate (LDR) Brachytherapy of Tongue using Californium 252 (Cf-252) | Treatment | Radiation |
| PRC10 | D915BYZ | Low Dose Rate (LDR) Brachytherapy of Tongue using Other Isotope | Treatment | Radiation |
| PRC10 | D91697Z | High Dose Rate (HDR) Brachytherapy of Salivary Glands using Cesium 137 (Cs-137) | Treatment | Radiation |
| PRC10 | D91698Z | High Dose Rate (HDR) Brachytherapy of Salivary Glands using Iridium 192 (Ir-192) | Treatment | Radiation |
| PRC10 | D91699Z | High Dose Rate (HDR) Brachytherapy of Salivary Glands using Iodine 125 (I-125) | Treatment | Radiation |
| PRC10 | D9169BZ | High Dose Rate (HDR) Brachytherapy of Salivary Glands using Palladium 103 (Pd-103) | Treatment | Radiation |
| PRC10 | D9169CZ | High Dose Rate (HDR) Brachytherapy of Salivary Glands using Californium 252 (Cf-252) | Treatment | Radiation |
| PRC10 | D9169YZ | High Dose Rate (HDR) Brachytherapy of Salivary Glands using Other Isotope | Treatment | Radiation |
| PRC10 | D916B7Z | Low Dose Rate (LDR) Brachytherapy of Salivary Glands using Cesium 137 (Cs-137) | Treatment | Radiation |
| PRC10 | D916B8Z | Low Dose Rate (LDR) Brachytherapy of Salivary Glands using Iridium 192 (Ir-192) | Treatment | Radiation |
| PRC10 | D916B9Z | Low Dose Rate (LDR) Brachytherapy of Salivary Glands using Iodine 125 (I-125) | Treatment | Radiation |
| PRC10 | D916BBZ | Low Dose Rate (LDR) Brachytherapy of Salivary Glands using Palladium 103 (Pd-103) | Treatment | Radiation |
| PRC10 | D916BCZ | Low Dose Rate (LDR) Brachytherapy of Salivary Glands using Californium 252 (Cf-252) | Treatment | Radiation |
| PRC10 | D916BYZ | Low Dose Rate (LDR) Brachytherapy of Salivary Glands using Other Isotope | Treatment | Radiation |
| PRC10 | D91797Z | High Dose Rate (HDR) Brachytherapy of Sinuses using Cesium 137 (Cs-137) | Treatment | Radiation |
| PRC10 | D91798Z | High Dose Rate (HDR) Brachytherapy of Sinuses using Iridium 192 (Ir-192) | Treatment | Radiation |
| PRC10 | D91799Z | High Dose Rate (HDR) Brachytherapy of Sinuses using Iodine 125 (I-125) | Treatment | Radiation |
| PRC10 | D9179BZ | High Dose Rate (HDR) Brachytherapy of Sinuses using Palladium 103 (Pd-103) | Treatment | Radiation |
| PRC10 | D9179CZ | High Dose Rate (HDR) Brachytherapy of Sinuses using Californium 252 (Cf-252) | Treatment | Radiation |
| PRC10 | D9179YZ | High Dose Rate (HDR) Brachytherapy of Sinuses using Other Isotope | Treatment | Radiation |
| PRC10 | D917B7Z | Low Dose Rate (LDR) Brachytherapy of Sinuses using Cesium 137 (Cs-137) | Treatment | Radiation |
| PRC10 | D917B8Z | Low Dose Rate (LDR) Brachytherapy of Sinuses using Iridium 192 (Ir-192) | Treatment | Radiation |
| PRC10 | D917B9Z | Low Dose Rate (LDR) Brachytherapy of Sinuses using Iodine 125 (I-125) | Treatment | Radiation |
| PRC10 | D917BBZ | Low Dose Rate (LDR) Brachytherapy of Sinuses using Palladium 103 (Pd-103) | Treatment | Radiation |
| PRC10 | D917BCZ | Low Dose Rate (LDR) Brachytherapy of Sinuses using Californium 252 (Cf-252) | Treatment | Radiation |
| PRC10 | D917BYZ | Low Dose Rate (LDR) Brachytherapy of Sinuses using Other Isotope | Treatment | Radiation |
| PRC10 | D91897Z | High Dose Rate (HDR) Brachytherapy of Hard Palate using Cesium 137 (Cs-137) | Treatment | Radiation |
| PRC10 | D91898Z | High Dose Rate (HDR) Brachytherapy of Hard Palate using Iridium 192 (Ir-192) | Treatment | Radiation |
| PRC10 | D91899Z | High Dose Rate (HDR) Brachytherapy of Hard Palate using Iodine 125 (I-125) | Treatment | Radiation |
| PRC10 | D9189BZ | High Dose Rate (HDR) Brachytherapy of Hard Palate using Palladium 103 (Pd-103) | Treatment | Radiation |
| PRC10 | D9189CZ | High Dose Rate (HDR) Brachytherapy of Hard Palate using Californium 252 (Cf-252) | Treatment | Radiation |
| PRC10 | D9189YZ | High Dose Rate (HDR) Brachytherapy of Hard Palate using Other Isotope | Treatment | Radiation |
| PRC10 | D918B7Z | Low Dose Rate (LDR) Brachytherapy of Hard Palate using Cesium 137 (Cs-137) | Treatment | Radiation |
| PRC10 | D918B8Z | Low Dose Rate (LDR) Brachytherapy of Hard Palate using Iridium 192 (Ir-192) | Treatment | Radiation |
| PRC10 | D918B9Z | Low Dose Rate (LDR) Brachytherapy of Hard Palate using Iodine 125 (I-125) | Treatment | Radiation |
| PRC10 | D918BBZ | Low Dose Rate (LDR) Brachytherapy of Hard Palate using Palladium 103 (Pd-103) | Treatment | Radiation |
| PRC10 | D918BCZ | Low Dose Rate (LDR) Brachytherapy of Hard Palate using Californium 252 (Cf-252) | Treatment | Radiation |
| PRC10 | D918BYZ | Low Dose Rate (LDR) Brachytherapy of Hard Palate using Other Isotope | Treatment | Radiation |
| PRC10 | D91997Z | High Dose Rate (HDR) Brachytherapy of Soft Palate using Cesium 137 (Cs-137) | Treatment | Radiation |
| PRC10 | D91998Z | High Dose Rate (HDR) Brachytherapy of Soft Palate using Iridium 192 (Ir-192) | Treatment | Radiation |
| PRC10 | D91999Z | High Dose Rate (HDR) Brachytherapy of Soft Palate using Iodine 125 (I-125) | Treatment | Radiation |
| PRC10 | D9199BZ | High Dose Rate (HDR) Brachytherapy of Soft Palate using Palladium 103 (Pd-103) | Treatment | Radiation |
| PRC10 | D9199CZ | High Dose Rate (HDR) Brachytherapy of Soft Palate using Californium 252 (Cf-252) | Treatment | Radiation |
| PRC10 | D9199YZ | High Dose Rate (HDR) Brachytherapy of Soft Palate using Other Isotope | Treatment | Radiation |
| PRC10 | D919B7Z | Low Dose Rate (LDR) Brachytherapy of Soft Palate using Cesium 137 (Cs-137) | Treatment | Radiation |
| PRC10 | D919B8Z | Low Dose Rate (LDR) Brachytherapy of Soft Palate using Iridium 192 (Ir-192) | Treatment | Radiation |
| PRC10 | D919B9Z | Low Dose Rate (LDR) Brachytherapy of Soft Palate using Iodine 125 (I-125) | Treatment | Radiation |
| PRC10 | D919BBZ | Low Dose Rate (LDR) Brachytherapy of Soft Palate using Palladium 103 (Pd-103) | Treatment | Radiation |
| PRC10 | D919BCZ | Low Dose Rate (LDR) Brachytherapy of Soft Palate using Californium 252 (Cf-252) | Treatment | Radiation |
| PRC10 | D919BYZ | Low Dose Rate (LDR) Brachytherapy of Soft Palate using Other Isotope | Treatment | Radiation |
| PRC10 | D91B97Z | High Dose Rate (HDR) Brachytherapy of Larynx using Cesium 137 (Cs-137) | Treatment | Radiation |
| PRC10 | D91B98Z | High Dose Rate (HDR) Brachytherapy of Larynx using Iridium 192 (Ir-192) | Treatment | Radiation |
| PRC10 | D91B99Z | High Dose Rate (HDR) Brachytherapy of Larynx using Iodine 125 (I-125) | Treatment | Radiation |
| PRC10 | D91B9BZ | High Dose Rate (HDR) Brachytherapy of Larynx using Palladium 103 (Pd-103) | Treatment | Radiation |
| PRC10 | D91B9CZ | High Dose Rate (HDR) Brachytherapy of Larynx using Californium 252 (Cf-252) | Treatment | Radiation |
| PRC10 | D91B9YZ | High Dose Rate (HDR) Brachytherapy of Larynx using Other Isotope | Treatment | Radiation |
| PRC10 | D91BB7Z | Low Dose Rate (LDR) Brachytherapy of Larynx using Cesium 137 (Cs-137) | Treatment | Radiation |
| PRC10 | D91BB8Z | Low Dose Rate (LDR) Brachytherapy of Larynx using Iridium 192 (Ir-192) | Treatment | Radiation |
| PRC10 | D91BB9Z | Low Dose Rate (LDR) Brachytherapy of Larynx using Iodine 125 (I-125) | Treatment | Radiation |
| PRC10 | D91BBBZ | Low Dose Rate (LDR) Brachytherapy of Larynx using Palladium 103 (Pd-103) | Treatment | Radiation |
| PRC10 | D91BBCZ | Low Dose Rate (LDR) Brachytherapy of Larynx using Californium 252 (Cf-252) | Treatment | Radiation |
| PRC10 | D91BBYZ | Low Dose Rate (LDR) Brachytherapy of Larynx using Other Isotope | Treatment | Radiation |
| PRC10 | D91D97Z | High Dose Rate (HDR) Brachytherapy of Nasopharynx using Cesium 137 (Cs-137) | Treatment | Radiation |
| PRC10 | D91D98Z | High Dose Rate (HDR) Brachytherapy of Nasopharynx using Iridium 192 (Ir-192) | Treatment | Radiation |
| PRC10 | D91D99Z | High Dose Rate (HDR) Brachytherapy of Nasopharynx using Iodine 125 (I-125) | Treatment | Radiation |
| PRC10 | D91D9BZ | High Dose Rate (HDR) Brachytherapy of Nasopharynx using Palladium 103 (Pd-103) | Treatment | Radiation |
| PRC10 | D91D9CZ | High Dose Rate (HDR) Brachytherapy of Nasopharynx using Californium 252 (Cf-252) | Treatment | Radiation |
| PRC10 | D91D9YZ | High Dose Rate (HDR) Brachytherapy of Nasopharynx using Other Isotope | Treatment | Radiation |
| PRC10 | D91DB7Z | Low Dose Rate (LDR) Brachytherapy of Nasopharynx using Cesium 137 (Cs-137) | Treatment | Radiation |
| PRC10 | D91DB8Z | Low Dose Rate (LDR) Brachytherapy of Nasopharynx using Iridium 192 (Ir-192) | Treatment | Radiation |
| PRC10 | D91DB9Z | Low Dose Rate (LDR) Brachytherapy of Nasopharynx using Iodine 125 (I-125) | Treatment | Radiation |
| PRC10 | D91DBBZ | Low Dose Rate (LDR) Brachytherapy of Nasopharynx using Palladium 103 (Pd-103) | Treatment | Radiation |
| PRC10 | D91DBCZ | Low Dose Rate (LDR) Brachytherapy of Nasopharynx using Californium 252 (Cf-252) | Treatment | Radiation |
| PRC10 | D91DBYZ | Low Dose Rate (LDR) Brachytherapy of Nasopharynx using Other Isotope | Treatment | Radiation |
| PRC10 | D91F97Z | High Dose Rate (HDR) Brachytherapy of Oropharynx using Cesium 137 (Cs-137) | Treatment | Radiation |
| PRC10 | D91F98Z | High Dose Rate (HDR) Brachytherapy of Oropharynx using Iridium 192 (Ir-192) | Treatment | Radiation |
| PRC10 | D91F99Z | High Dose Rate (HDR) Brachytherapy of Oropharynx using Iodine 125 (I-125) | Treatment | Radiation |
| PRC10 | D91F9BZ | High Dose Rate (HDR) Brachytherapy of Oropharynx using Palladium 103 (Pd-103) | Treatment | Radiation |
| PRC10 | D91F9CZ | High Dose Rate (HDR) Brachytherapy of Oropharynx using Californium 252 (Cf-252) | Treatment | Radiation |
| PRC10 | D91F9YZ | High Dose Rate (HDR) Brachytherapy of Oropharynx using Other Isotope | Treatment | Radiation |
| PRC10 | D91FB7Z | Low Dose Rate (LDR) Brachytherapy of Oropharynx using Cesium 137 (Cs-137) | Treatment | Radiation |
| PRC10 | D91FB8Z | Low Dose Rate (LDR) Brachytherapy of Oropharynx using Iridium 192 (Ir-192) | Treatment | Radiation |
| PRC10 | D91FB9Z | Low Dose Rate (LDR) Brachytherapy of Oropharynx using Iodine 125 (I-125) | Treatment | Radiation |
| PRC10 | D91FBBZ | Low Dose Rate (LDR) Brachytherapy of Oropharynx using Palladium 103 (Pd-103) | Treatment | Radiation |
| PRC10 | D91FBCZ | Low Dose Rate (LDR) Brachytherapy of Oropharynx using Californium 252 (Cf-252) | Treatment | Radiation |
| PRC10 | D91FBYZ | Low Dose Rate (LDR) Brachytherapy of Oropharynx using Other Isotope | Treatment | Radiation |
| PRC10 | D9Y07ZZ | Contact Radiation of Ear | Treatment | Radiation |
| PRC10 | D9Y0FZZ | Plaque Radiation of Ear | Treatment | Radiation |
| PRC10 | D9Y17ZZ | Contact Radiation of Nose | Treatment | Radiation |
| PRC10 | D9Y1FZZ | Plaque Radiation of Nose | Treatment | Radiation |
| PRC10 | D9Y37ZZ | Contact Radiation of Hypopharynx | Treatment | Radiation |
| PRC10 | D9Y47ZZ | Contact Radiation of Mouth | Treatment | Radiation |
| PRC10 | D9Y4CZZ | Intraoperative Radiation Therapy (IORT) of Mouth | Treatment | Radiation |
| PRC10 | D9Y4FZZ | Plaque Radiation of Mouth | Treatment | Radiation |
| PRC10 | D9Y57ZZ | Contact Radiation of Tongue | Treatment | Radiation |
| PRC10 | D9Y5FZZ | Plaque Radiation of Tongue | Treatment | Radiation |
| PRC10 | D9Y67ZZ | Contact Radiation of Salivary Glands | Treatment | Radiation |
| PRC10 | D9Y6FZZ | Plaque Radiation of Salivary Glands | Treatment | Radiation |
| PRC10 | D9Y77ZZ | Contact Radiation of Sinuses | Treatment | Radiation |
| PRC10 | D9Y7FZZ | Plaque Radiation of Sinuses | Treatment | Radiation |
| PRC10 | D9Y87ZZ | Contact Radiation of Hard Palate | Treatment | Radiation |
| PRC10 | D9Y8FZZ | Plaque Radiation of Hard Palate | Treatment | Radiation |
| PRC10 | D9Y97ZZ | Contact Radiation of Soft Palate | Treatment | Radiation |
| PRC10 | D9Y9FZZ | Plaque Radiation of Soft Palate | Treatment | Radiation |
| PRC10 | D9YB7ZZ | Contact Radiation of Larynx | Treatment | Radiation |
| PRC10 | D9YBCZZ | Intraoperative Radiation Therapy (IORT) of Larynx | Treatment | Radiation |
| PRC10 | D9YBFZZ | Plaque Radiation of Larynx | Treatment | Radiation |
| PRC10 | D9YCCZZ | Intraoperative Radiation Therapy (IORT) of Pharynx | Treatment | Radiation |
| PRC10 | D9YCFZZ | Plaque Radiation of Pharynx | Treatment | Radiation |
| PRC10 | D9YD7ZZ | Contact Radiation of Nasopharynx | Treatment | Radiation |
| PRC10 | D9YDCZZ | Intraoperative Radiation Therapy (IORT) of Nasopharynx | Treatment | Radiation |
| PRC10 | D9YDFZZ | Plaque Radiation of Nasopharynx | Treatment | Radiation |
| PRC10 | D9YF7ZZ | Contact Radiation of Oropharynx | Treatment | Radiation |
| PRC10 | DB000ZZ | Beam Radiation of Trachea using Photons <1 MeV | Treatment | Radiation |
| PRC10 | DB003ZZ | Beam Radiation of Trachea using Electrons | Treatment | Radiation |
| PRC10 | DB004ZZ | Beam Radiation of Trachea using Heavy Particles (Protons,Ions) | Treatment | Radiation |
| PRC10 | DB005ZZ | Beam Radiation of Trachea using Neutrons | Treatment | Radiation |
| PRC10 | DB010ZZ | Beam Radiation of Bronchus using Photons <1 MeV | Treatment | Radiation |
| PRC10 | DB013ZZ | Beam Radiation of Bronchus using Electrons | Treatment | Radiation |
| PRC10 | DB014ZZ | Beam Radiation of Bronchus using Heavy Particles (Protons,Ions) | Treatment | Radiation |
| PRC10 | DB015ZZ | Beam Radiation of Bronchus using Neutrons | Treatment | Radiation |
| PRC10 | DB020ZZ | Beam Radiation of Lung using Photons <1 MeV | Treatment | Radiation |
| PRC10 | DB023ZZ | Beam Radiation of Lung using Electrons | Treatment | Radiation |
| PRC10 | DB024ZZ | Beam Radiation of Lung using Heavy Particles (Protons,Ions) | Treatment | Radiation |
| PRC10 | DB025ZZ | Beam Radiation of Lung using Neutrons | Treatment | Radiation |
| PRC10 | DB050ZZ | Beam Radiation of Pleura using Photons <1 MeV | Treatment | Radiation |
| PRC10 | DB053ZZ | Beam Radiation of Pleura using Electrons | Treatment | Radiation |
| PRC10 | DB054ZZ | Beam Radiation of Pleura using Heavy Particles (Protons,Ions) | Treatment | Radiation |
| PRC10 | DB055ZZ | Beam Radiation of Pleura using Neutrons | Treatment | Radiation |
| PRC10 | DB060ZZ | Beam Radiation of Mediastinum using Photons <1 MeV | Treatment | Radiation |
| PRC10 | DB063ZZ | Beam Radiation of Mediastinum using Electrons | Treatment | Radiation |
| PRC10 | DB064ZZ | Beam Radiation of Mediastinum using Heavy Particles (Protons,Ions) | Treatment | Radiation |
| PRC10 | DB065ZZ | Beam Radiation of Mediastinum using Neutrons | Treatment | Radiation |
| PRC10 | DB070ZZ | Beam Radiation of Chest Wall using Photons <1 MeV | Treatment | Radiation |
| PRC10 | DB073ZZ | Beam Radiation of Chest Wall using Electrons | Treatment | Radiation |
| PRC10 | DB074ZZ | Beam Radiation of Chest Wall using Heavy Particles (Protons,Ions) | Treatment | Radiation |
| PRC10 | DB075ZZ | Beam Radiation of Chest Wall using Neutrons | Treatment | Radiation |
| PRC10 | DB080ZZ | Beam Radiation of Diaphragm using Photons <1 MeV | Treatment | Radiation |
| PRC10 | DB083ZZ | Beam Radiation of Diaphragm using Electrons | Treatment | Radiation |
| PRC10 | DB084ZZ | Beam Radiation of Diaphragm using Heavy Particles (Protons,Ions) | Treatment | Radiation |
| PRC10 | DB085ZZ | Beam Radiation of Diaphragm using Neutrons | Treatment | Radiation |
| PRC10 | DB1097Z | High Dose Rate (HDR) Brachytherapy of Trachea using Cesium 137 (Cs-137) | Treatment | Radiation |
| PRC10 | DB1098Z | High Dose Rate (HDR) Brachytherapy of Trachea using Iridium 192 (Ir-192) | Treatment | Radiation |
| PRC10 | DB1099Z | High Dose Rate (HDR) Brachytherapy of Trachea using Iodine 125 (I-125) | Treatment | Radiation |
| PRC10 | DB109BZ | High Dose Rate (HDR) Brachytherapy of Trachea using Palladium 103 (Pd-103) | Treatment | Radiation |
| PRC10 | DB109CZ | High Dose Rate (HDR) Brachytherapy of Trachea using Californium 252 (Cf-252) | Treatment | Radiation |
| PRC10 | DB109YZ | High Dose Rate (HDR) Brachytherapy of Trachea using Other Isotope | Treatment | Radiation |
| PRC10 | DB10B7Z | Low Dose Rate (LDR) Brachytherapy of Trachea using Cesium 137 (Cs-137) | Treatment | Radiation |
| PRC10 | DB10B8Z | Low Dose Rate (LDR) Brachytherapy of Trachea using Iridium 192 (Ir-192) | Treatment | Radiation |
| PRC10 | DB10B9Z | Low Dose Rate (LDR) Brachytherapy of Trachea using Iodine 125 (I-125) | Treatment | Radiation |
| PRC10 | DB10BBZ | Low Dose Rate (LDR) Brachytherapy of Trachea using Palladium 103 (Pd-103) | Treatment | Radiation |
| PRC10 | DB10BCZ | Low Dose Rate (LDR) Brachytherapy of Trachea using Californium 252 (Cf-252) | Treatment | Radiation |
| PRC10 | DB10BYZ | Low Dose Rate (LDR) Brachytherapy of Trachea using Other Isotope | Treatment | Radiation |
| PRC10 | DB1197Z | High Dose Rate (HDR) Brachytherapy of Bronchus using Cesium 137 (Cs-137) | Treatment | Radiation |
| PRC10 | DB1198Z | High Dose Rate (HDR) Brachytherapy of Bronchus using Iridium 192 (Ir-192) | Treatment | Radiation |
| PRC10 | DB1199Z | High Dose Rate (HDR) Brachytherapy of Bronchus using Iodine 125 (I-125) | Treatment | Radiation |
| PRC10 | DB119BZ | High Dose Rate (HDR) Brachytherapy of Bronchus using Palladium 103 (Pd-103) | Treatment | Radiation |
| PRC10 | DB119CZ | High Dose Rate (HDR) Brachytherapy of Bronchus using Californium 252 (Cf-252) | Treatment | Radiation |
| PRC10 | DB119YZ | High Dose Rate (HDR) Brachytherapy of Bronchus using Other Isotope | Treatment | Radiation |
| PRC10 | DB11B7Z | Low Dose Rate (LDR) Brachytherapy of Bronchus using Cesium 137 (Cs-137) | Treatment | Radiation |
| PRC10 | DB11B8Z | Low Dose Rate (LDR) Brachytherapy of Bronchus using Iridium 192 (Ir-192) | Treatment | Radiation |
| PRC10 | DB11B9Z | Low Dose Rate (LDR) Brachytherapy of Bronchus using Iodine 125 (I-125) | Treatment | Radiation |
| PRC10 | DB11BBZ | Low Dose Rate (LDR) Brachytherapy of Bronchus using Palladium 103 (Pd-103) | Treatment | Radiation |
| PRC10 | DB11BCZ | Low Dose Rate (LDR) Brachytherapy of Bronchus using Californium 252 (Cf-252) | Treatment | Radiation |
| PRC10 | DB11BYZ | Low Dose Rate (LDR) Brachytherapy of Bronchus using Other Isotope | Treatment | Radiation |
| PRC10 | DB1297Z | High Dose Rate (HDR) Brachytherapy of Lung using Cesium 137 (Cs-137) | Treatment | Radiation |
| PRC10 | DB1298Z | High Dose Rate (HDR) Brachytherapy of Lung using Iridium 192 (Ir-192) | Treatment | Radiation |
| PRC10 | DB1299Z | High Dose Rate (HDR) Brachytherapy of Lung using Iodine 125 (I-125) | Treatment | Radiation |
| PRC10 | DB129BZ | High Dose Rate (HDR) Brachytherapy of Lung using Palladium 103 (Pd-103) | Treatment | Radiation |
| PRC10 | DB129CZ | High Dose Rate (HDR) Brachytherapy of Lung using Californium 252 (Cf-252) | Treatment | Radiation |
| PRC10 | DB129YZ | High Dose Rate (HDR) Brachytherapy of Lung using Other Isotope | Treatment | Radiation |
| PRC10 | DB12B7Z | Low Dose Rate (LDR) Brachytherapy of Lung using Cesium 137 (Cs-137) | Treatment | Radiation |
| PRC10 | DB12B8Z | Low Dose Rate (LDR) Brachytherapy of Lung using Iridium 192 (Ir-192) | Treatment | Radiation |
| PRC10 | DB12B9Z | Low Dose Rate (LDR) Brachytherapy of Lung using Iodine 125 (I-125) | Treatment | Radiation |
| PRC10 | DB12BBZ | Low Dose Rate (LDR) Brachytherapy of Lung using Palladium 103 (Pd-103) | Treatment | Radiation |
| PRC10 | DB12BCZ | Low Dose Rate (LDR) Brachytherapy of Lung using Californium 252 (Cf-252) | Treatment | Radiation |
| PRC10 | DB12BYZ | Low Dose Rate (LDR) Brachytherapy of Lung using Other Isotope | Treatment | Radiation |
| PRC10 | DB1597Z | High Dose Rate (HDR) Brachytherapy of Pleura using Cesium 137 (Cs-137) | Treatment | Radiation |
| PRC10 | DB1598Z | High Dose Rate (HDR) Brachytherapy of Pleura using Iridium 192 (Ir-192) | Treatment | Radiation |
| PRC10 | DB1599Z | High Dose Rate (HDR) Brachytherapy of Pleura using Iodine 125 (I-125) | Treatment | Radiation |
| PRC10 | DB159BZ | High Dose Rate (HDR) Brachytherapy of Pleura using Palladium 103 (Pd-103) | Treatment | Radiation |
| PRC10 | DB159CZ | High Dose Rate (HDR) Brachytherapy of Pleura using Californium 252 (Cf-252) | Treatment | Radiation |
| PRC10 | DB159YZ | High Dose Rate (HDR) Brachytherapy of Pleura using Other Isotope | Treatment | Radiation |
| PRC10 | DB15B7Z | Low Dose Rate (LDR) Brachytherapy of Pleura using Cesium 137 (Cs-137) | Treatment | Radiation |
| PRC10 | DB15B8Z | Low Dose Rate (LDR) Brachytherapy of Pleura using Iridium 192 (Ir-192) | Treatment | Radiation |
| PRC10 | DB15B9Z | Low Dose Rate (LDR) Brachytherapy of Pleura using Iodine 125 (I-125) | Treatment | Radiation |
| PRC10 | DB15BBZ | Low Dose Rate (LDR) Brachytherapy of Pleura using Palladium 103 (Pd-103) | Treatment | Radiation |
| PRC10 | DB15BCZ | Low Dose Rate (LDR) Brachytherapy of Pleura using Californium 252 (Cf-252) | Treatment | Radiation |
| PRC10 | DB15BYZ | Low Dose Rate (LDR) Brachytherapy of Pleura using Other Isotope | Treatment | Radiation |
| PRC10 | DB1697Z | High Dose Rate (HDR) Brachytherapy of Mediastinum using Cesium 137 (Cs-137) | Treatment | Radiation |
| PRC10 | DB1698Z | High Dose Rate (HDR) Brachytherapy of Mediastinum using Iridium 192 (Ir-192) | Treatment | Radiation |
| PRC10 | DB1699Z | High Dose Rate (HDR) Brachytherapy of Mediastinum using Iodine 125 (I-125) | Treatment | Radiation |
| PRC10 | DB169BZ | High Dose Rate (HDR) Brachytherapy of Mediastinum using Palladium 103 (Pd-103) | Treatment | Radiation |
| PRC10 | DB169CZ | High Dose Rate (HDR) Brachytherapy of Mediastinum using Californium 252 (Cf-252) | Treatment | Radiation |
| PRC10 | DB169YZ | High Dose Rate (HDR) Brachytherapy of Mediastinum using Other Isotope | Treatment | Radiation |
| PRC10 | DB16B7Z | Low Dose Rate (LDR) Brachytherapy of Mediastinum using Cesium 137 (Cs-137) | Treatment | Radiation |
| PRC10 | DB16B8Z | Low Dose Rate (LDR) Brachytherapy of Mediastinum using Iridium 192 (Ir-192) | Treatment | Radiation |
| PRC10 | DB16B9Z | Low Dose Rate (LDR) Brachytherapy of Mediastinum using Iodine 125 (I-125) | Treatment | Radiation |
| PRC10 | DB16BBZ | Low Dose Rate (LDR) Brachytherapy of Mediastinum using Palladium 103 (Pd-103) | Treatment | Radiation |
| PRC10 | DB16BCZ | Low Dose Rate (LDR) Brachytherapy of Mediastinum using Californium 252 (Cf-252) | Treatment | Radiation |
| PRC10 | DB16BYZ | Low Dose Rate (LDR) Brachytherapy of Mediastinum using Other Isotope | Treatment | Radiation |
| PRC10 | DB1797Z | High Dose Rate (HDR) Brachytherapy of Chest Wall using Cesium 137 (Cs-137) | Treatment | Radiation |
| PRC10 | DB1798Z | High Dose Rate (HDR) Brachytherapy of Chest Wall using Iridium 192 (Ir-192) | Treatment | Radiation |
| PRC10 | DB1799Z | High Dose Rate (HDR) Brachytherapy of Chest Wall using Iodine 125 (I-125) | Treatment | Radiation |
| PRC10 | DB179BZ | High Dose Rate (HDR) Brachytherapy of Chest Wall using Palladium 103 (Pd-103) | Treatment | Radiation |
| PRC10 | DB179CZ | High Dose Rate (HDR) Brachytherapy of Chest Wall using Californium 252 (Cf-252) | Treatment | Radiation |
| PRC10 | DB179YZ | High Dose Rate (HDR) Brachytherapy of Chest Wall using Other Isotope | Treatment | Radiation |
| PRC10 | DB17B7Z | Low Dose Rate (LDR) Brachytherapy of Chest Wall using Cesium 137 (Cs-137) | Treatment | Radiation |
| PRC10 | DB17B8Z | Low Dose Rate (LDR) Brachytherapy of Chest Wall using Iridium 192 (Ir-192) | Treatment | Radiation |
| PRC10 | DB17B9Z | Low Dose Rate (LDR) Brachytherapy of Chest Wall using Iodine 125 (I-125) | Treatment | Radiation |
| PRC10 | DB17BBZ | Low Dose Rate (LDR) Brachytherapy of Chest Wall using Palladium 103 (Pd-103) | Treatment | Radiation |
| PRC10 | DB17BCZ | Low Dose Rate (LDR) Brachytherapy of Chest Wall using Californium 252 (Cf-252) | Treatment | Radiation |
| PRC10 | DB17BYZ | Low Dose Rate (LDR) Brachytherapy of Chest Wall using Other Isotope | Treatment | Radiation |
| PRC10 | DB1897Z | High Dose Rate (HDR) Brachytherapy of Diaphragm using Cesium 137 (Cs-137) | Treatment | Radiation |
| PRC10 | DB1898Z | High Dose Rate (HDR) Brachytherapy of Diaphragm using Iridium 192 (Ir-192) | Treatment | Radiation |
| PRC10 | DB1899Z | High Dose Rate (HDR) Brachytherapy of Diaphragm using Iodine 125 (I-125) | Treatment | Radiation |
| PRC10 | DB189BZ | High Dose Rate (HDR) Brachytherapy of Diaphragm using Palladium 103 (Pd-103) | Treatment | Radiation |
| PRC10 | DB189CZ | High Dose Rate (HDR) Brachytherapy of Diaphragm using Californium 252 (Cf-252) | Treatment | Radiation |
| PRC10 | DB189YZ | High Dose Rate (HDR) Brachytherapy of Diaphragm using Other Isotope | Treatment | Radiation |
| PRC10 | DB18B7Z | Low Dose Rate (LDR) Brachytherapy of Diaphragm using Cesium 137 (Cs-137) | Treatment | Radiation |
| PRC10 | DB18B8Z | Low Dose Rate (LDR) Brachytherapy of Diaphragm using Iridium 192 (Ir-192) | Treatment | Radiation |
| PRC10 | DB18B9Z | Low Dose Rate (LDR) Brachytherapy of Diaphragm using Iodine 125 (I-125) | Treatment | Radiation |
| PRC10 | DB18BBZ | Low Dose Rate (LDR) Brachytherapy of Diaphragm using Palladium 103 (Pd-103) | Treatment | Radiation |
| PRC10 | DB18BCZ | Low Dose Rate (LDR) Brachytherapy of Diaphragm using Californium 252 (Cf-252) | Treatment | Radiation |
| PRC10 | DB18BYZ | Low Dose Rate (LDR) Brachytherapy of Diaphragm using Other Isotope | Treatment | Radiation |
| PRC10 | DBY07ZZ | Contact Radiation of Trachea | Treatment | Radiation |
| PRC10 | DBY0FZZ | Plaque Radiation of Trachea | Treatment | Radiation |
| PRC10 | DBY17ZZ | Contact Radiation of Bronchus | Treatment | Radiation |
| PRC10 | DBY1FZZ | Plaque Radiation of Bronchus | Treatment | Radiation |
| PRC10 | DBY27ZZ | Contact Radiation of Lung | Treatment | Radiation |
| PRC10 | DBY2FZZ | Plaque Radiation of Lung | Treatment | Radiation |
| PRC10 | DBY57ZZ | Contact Radiation of Pleura | Treatment | Radiation |
| PRC10 | DBY5FZZ | Plaque Radiation of Pleura | Treatment | Radiation |
| PRC10 | DBY67ZZ | Contact Radiation of Mediastinum | Treatment | Radiation |
| PRC10 | DBY6FZZ | Plaque Radiation of Mediastinum | Treatment | Radiation |
| PRC10 | DBY77ZZ | Contact Radiation of Chest Wall | Treatment | Radiation |
| PRC10 | DBY7FZZ | Plaque Radiation of Chest Wall | Treatment | Radiation |
| PRC10 | DBY87ZZ | Contact Radiation of Diaphragm | Treatment | Radiation |
| PRC10 | DBY8FZZ | Plaque Radiation of Diaphragm | Treatment | Radiation |
| PRC10 | DD000ZZ | Beam Radiation of Esophagus using Photons <1 MeV | Treatment | Radiation |
| PRC10 | DD003ZZ | Beam Radiation of Esophagus using Electrons | Treatment | Radiation |
| PRC10 | DD004ZZ | Beam Radiation of Esophagus using Heavy Particles (Protons,Ions) | Treatment | Radiation |
| PRC10 | DD005ZZ | Beam Radiation of Esophagus using Neutrons | Treatment | Radiation |
| PRC10 | DD010ZZ | Beam Radiation of Stomach using Photons <1 MeV | Treatment | Radiation |
| PRC10 | DD013ZZ | Beam Radiation of Stomach using Electrons | Treatment | Radiation |
| PRC10 | DD014ZZ | Beam Radiation of Stomach using Heavy Particles (Protons,Ions) | Treatment | Radiation |
| PRC10 | DD015ZZ | Beam Radiation of Stomach using Neutrons | Treatment | Radiation |
| PRC10 | DD020ZZ | Beam Radiation of Duodenum using Photons <1 MeV | Treatment | Radiation |
| PRC10 | DD023ZZ | Beam Radiation of Duodenum using Electrons | Treatment | Radiation |
| PRC10 | DD024ZZ | Beam Radiation of Duodenum using Heavy Particles (Protons,Ions) | Treatment | Radiation |
| PRC10 | DD025ZZ | Beam Radiation of Duodenum using Neutrons | Treatment | Radiation |
| PRC10 | DD030ZZ | Beam Radiation of Jejunum using Photons <1 MeV | Treatment | Radiation |
| PRC10 | DD033ZZ | Beam Radiation of Jejunum using Electrons | Treatment | Radiation |
| PRC10 | DD034ZZ | Beam Radiation of Jejunum using Heavy Particles (Protons,Ions) | Treatment | Radiation |
| PRC10 | DD035ZZ | Beam Radiation of Jejunum using Neutrons | Treatment | Radiation |
| PRC10 | DD040ZZ | Beam Radiation of Ileum using Photons <1 MeV | Treatment | Radiation |
| PRC10 | DD043ZZ | Beam Radiation of Ileum using Electrons | Treatment | Radiation |
| PRC10 | DD044ZZ | Beam Radiation of Ileum using Heavy Particles (Protons,Ions) | Treatment | Radiation |
| PRC10 | DD045ZZ | Beam Radiation of Ileum using Neutrons | Treatment | Radiation |
| PRC10 | DD050ZZ | Beam Radiation of Colon using Photons <1 MeV | Treatment | Radiation |
| PRC10 | DD053ZZ | Beam Radiation of Colon using Electrons | Treatment | Radiation |
| PRC10 | DD054ZZ | Beam Radiation of Colon using Heavy Particles (Protons,Ions) | Treatment | Radiation |
| PRC10 | DD055ZZ | Beam Radiation of Colon using Neutrons | Treatment | Radiation |
| PRC10 | DD070ZZ | Beam Radiation of Rectum using Photons <1 MeV | Treatment | Radiation |
| PRC10 | DD073ZZ | Beam Radiation of Rectum using Electrons | Treatment | Radiation |
| PRC10 | DD074ZZ | Beam Radiation of Rectum using Heavy Particles (Protons,Ions) | Treatment | Radiation |
| PRC10 | DD075ZZ | Beam Radiation of Rectum using Neutrons | Treatment | Radiation |
| PRC10 | DD1097Z | High Dose Rate (HDR) Brachytherapy of Esophagus using Cesium 137 (Cs-137) | Treatment | Radiation |
| PRC10 | DD1098Z | High Dose Rate (HDR) Brachytherapy of Esophagus using Iridium 192 (Ir-192) | Treatment | Radiation |
| PRC10 | DD1099Z | High Dose Rate (HDR) Brachytherapy of Esophagus using Iodine 125 (I-125) | Treatment | Radiation |
| PRC10 | DD109BZ | High Dose Rate (HDR) Brachytherapy of Esophagus using Palladium 103 (Pd-103) | Treatment | Radiation |
| PRC10 | DD109CZ | High Dose Rate (HDR) Brachytherapy of Esophagus using Californium 252 (Cf-252) | Treatment | Radiation |
| PRC10 | DD109YZ | High Dose Rate (HDR) Brachytherapy of Esophagus using Other Isotope | Treatment | Radiation |
| PRC10 | DD10B7Z | Low Dose Rate (LDR) Brachytherapy of Esophagus using Cesium 137 (Cs-137) | Treatment | Radiation |
| PRC10 | DD10B8Z | Low Dose Rate (LDR) Brachytherapy of Esophagus using Iridium 192 (Ir-192) | Treatment | Radiation |
| PRC10 | DD10B9Z | Low Dose Rate (LDR) Brachytherapy of Esophagus using Iodine 125 (I-125) | Treatment | Radiation |
| PRC10 | DD10BBZ | Low Dose Rate (LDR) Brachytherapy of Esophagus using Palladium 103 (Pd-103) | Treatment | Radiation |
| PRC10 | DD10BCZ | Low Dose Rate (LDR) Brachytherapy of Esophagus using Californium 252 (Cf-252) | Treatment | Radiation |
| PRC10 | DD10BYZ | Low Dose Rate (LDR) Brachytherapy of Esophagus using Other Isotope | Treatment | Radiation |
| PRC10 | DD1197Z | High Dose Rate (HDR) Brachytherapy of Stomach using Cesium 137 (Cs-137) | Treatment | Radiation |
| PRC10 | DD1198Z | High Dose Rate (HDR) Brachytherapy of Stomach using Iridium 192 (Ir-192) | Treatment | Radiation |
| PRC10 | DD1199Z | High Dose Rate (HDR) Brachytherapy of Stomach using Iodine 125 (I-125) | Treatment | Radiation |
| PRC10 | DD119BZ | High Dose Rate (HDR) Brachytherapy of Stomach using Palladium 103 (Pd-103) | Treatment | Radiation |
| PRC10 | DD119CZ | High Dose Rate (HDR) Brachytherapy of Stomach using Californium 252 (Cf-252) | Treatment | Radiation |
| PRC10 | DD119YZ | High Dose Rate (HDR) Brachytherapy of Stomach using Other Isotope | Treatment | Radiation |
| PRC10 | DD11B7Z | Low Dose Rate (LDR) Brachytherapy of Stomach using Cesium 137 (Cs-137) | Treatment | Radiation |
| PRC10 | DD11B8Z | Low Dose Rate (LDR) Brachytherapy of Stomach using Iridium 192 (Ir-192) | Treatment | Radiation |
| PRC10 | DD11B9Z | Low Dose Rate (LDR) Brachytherapy of Stomach using Iodine 125 (I-125) | Treatment | Radiation |
| PRC10 | DD11BBZ | Low Dose Rate (LDR) Brachytherapy of Stomach using Palladium 103 (Pd-103) | Treatment | Radiation |
| PRC10 | DD11BCZ | Low Dose Rate (LDR) Brachytherapy of Stomach using Californium 252 (Cf-252) | Treatment | Radiation |
| PRC10 | DD11BYZ | Low Dose Rate (LDR) Brachytherapy of Stomach using Other Isotope | Treatment | Radiation |
| PRC10 | DD1297Z | High Dose Rate (HDR) Brachytherapy of Duodenum using Cesium 137 (Cs-137) | Treatment | Radiation |
| PRC10 | DD1298Z | High Dose Rate (HDR) Brachytherapy of Duodenum using Iridium 192 (Ir-192) | Treatment | Radiation |
| PRC10 | DD1299Z | High Dose Rate (HDR) Brachytherapy of Duodenum using Iodine 125 (I-125) | Treatment | Radiation |
| PRC10 | DD129BZ | High Dose Rate (HDR) Brachytherapy of Duodenum using Palladium 103 (Pd-103) | Treatment | Radiation |
| PRC10 | DD129CZ | High Dose Rate (HDR) Brachytherapy of Duodenum using Californium 252 (Cf-252) | Treatment | Radiation |
| PRC10 | DD129YZ | High Dose Rate (HDR) Brachytherapy of Duodenum using Other Isotope | Treatment | Radiation |
| PRC10 | DD12B7Z | Low Dose Rate (LDR) Brachytherapy of Duodenum using Cesium 137 (Cs-137) | Treatment | Radiation |
| PRC10 | DD12B8Z | Low Dose Rate (LDR) Brachytherapy of Duodenum using Iridium 192 (Ir-192) | Treatment | Radiation |
| PRC10 | DD12B9Z | Low Dose Rate (LDR) Brachytherapy of Duodenum using Iodine 125 (I-125) | Treatment | Radiation |
| PRC10 | DD12BBZ | Low Dose Rate (LDR) Brachytherapy of Duodenum using Palladium 103 (Pd-103) | Treatment | Radiation |
| PRC10 | DD12BCZ | Low Dose Rate (LDR) Brachytherapy of Duodenum using Californium 252 (Cf-252) | Treatment | Radiation |
| PRC10 | DD12BYZ | Low Dose Rate (LDR) Brachytherapy of Duodenum using Other Isotope | Treatment | Radiation |
| PRC10 | DD1397Z | High Dose Rate (HDR) Brachytherapy of Jejunum using Cesium 137 (Cs-137) | Treatment | Radiation |
| PRC10 | DD1398Z | High Dose Rate (HDR) Brachytherapy of Jejunum using Iridium 192 (Ir-192) | Treatment | Radiation |
| PRC10 | DD1399Z | High Dose Rate (HDR) Brachytherapy of Jejunum using Iodine 125 (I-125) | Treatment | Radiation |
| PRC10 | DD139BZ | High Dose Rate (HDR) Brachytherapy of Jejunum using Palladium 103 (Pd-103) | Treatment | Radiation |
| PRC10 | DD139CZ | High Dose Rate (HDR) Brachytherapy of Jejunum using Californium 252 (Cf-252) | Treatment | Radiation |
| PRC10 | DD139YZ | High Dose Rate (HDR) Brachytherapy of Jejunum using Other Isotope | Treatment | Radiation |
| PRC10 | DD13B7Z | Low Dose Rate (LDR) Brachytherapy of Jejunum using Cesium 137 (Cs-137) | Treatment | Radiation |
| PRC10 | DD13B8Z | Low Dose Rate (LDR) Brachytherapy of Jejunum using Iridium 192 (Ir-192) | Treatment | Radiation |
| PRC10 | DD13B9Z | Low Dose Rate (LDR) Brachytherapy of Jejunum using Iodine 125 (I-125) | Treatment | Radiation |
| PRC10 | DD13BBZ | Low Dose Rate (LDR) Brachytherapy of Jejunum using Palladium 103 (Pd-103) | Treatment | Radiation |
| PRC10 | DD13BCZ | Low Dose Rate (LDR) Brachytherapy of Jejunum using Californium 252 (Cf-252) | Treatment | Radiation |
| PRC10 | DD13BYZ | Low Dose Rate (LDR) Brachytherapy of Jejunum using Other Isotope | Treatment | Radiation |
| PRC10 | DD1497Z | High Dose Rate (HDR) Brachytherapy of Ileum using Cesium 137 (Cs-137) | Treatment | Radiation |
| PRC10 | DD1498Z | High Dose Rate (HDR) Brachytherapy of Ileum using Iridium 192 (Ir-192) | Treatment | Radiation |
| PRC10 | DD1499Z | High Dose Rate (HDR) Brachytherapy of Ileum using Iodine 125 (I-125) | Treatment | Radiation |
| PRC10 | DD149BZ | High Dose Rate (HDR) Brachytherapy of Ileum using Palladium 103 (Pd-103) | Treatment | Radiation |
| PRC10 | DD149CZ | High Dose Rate (HDR) Brachytherapy of Ileum using Californium 252 (Cf-252) | Treatment | Radiation |
| PRC10 | DD149YZ | High Dose Rate (HDR) Brachytherapy of Ileum using Other Isotope | Treatment | Radiation |
| PRC10 | DD14B7Z | Low Dose Rate (LDR) Brachytherapy of Ileum using Cesium 137 (Cs-137) | Treatment | Radiation |
| PRC10 | DD14B8Z | Low Dose Rate (LDR) Brachytherapy of Ileum using Iridium 192 (Ir-192) | Treatment | Radiation |
| PRC10 | DD14B9Z | Low Dose Rate (LDR) Brachytherapy of Ileum using Iodine 125 (I-125) | Treatment | Radiation |
| PRC10 | DD14BBZ | Low Dose Rate (LDR) Brachytherapy of Ileum using Palladium 103 (Pd-103) | Treatment | Radiation |
| PRC10 | DD14BCZ | Low Dose Rate (LDR) Brachytherapy of Ileum using Californium 252 (Cf-252) | Treatment | Radiation |
| PRC10 | DD14BYZ | Low Dose Rate (LDR) Brachytherapy of Ileum using Other Isotope | Treatment | Radiation |
| PRC10 | DD1597Z | High Dose Rate (HDR) Brachytherapy of Colon using Cesium 137 (Cs-137) | Treatment | Radiation |
| PRC10 | DD1598Z | High Dose Rate (HDR) Brachytherapy of Colon using Iridium 192 (Ir-192) | Treatment | Radiation |
| PRC10 | DD1599Z | High Dose Rate (HDR) Brachytherapy of Colon using Iodine 125 (I-125) | Treatment | Radiation |
| PRC10 | DD159BZ | High Dose Rate (HDR) Brachytherapy of Colon using Palladium 103 (Pd-103) | Treatment | Radiation |
| PRC10 | DD159CZ | High Dose Rate (HDR) Brachytherapy of Colon using Californium 252 (Cf-252) | Treatment | Radiation |
| PRC10 | DD159YZ | High Dose Rate (HDR) Brachytherapy of Colon using Other Isotope | Treatment | Radiation |
| PRC10 | DD15B7Z | Low Dose Rate (LDR) Brachytherapy of Colon using Cesium 137 (Cs-137) | Treatment | Radiation |
| PRC10 | DD15B8Z | Low Dose Rate (LDR) Brachytherapy of Colon using Iridium 192 (Ir-192) | Treatment | Radiation |
| PRC10 | DD15B9Z | Low Dose Rate (LDR) Brachytherapy of Colon using Iodine 125 (I-125) | Treatment | Radiation |
| PRC10 | DD15BBZ | Low Dose Rate (LDR) Brachytherapy of Colon using Palladium 103 (Pd-103) | Treatment | Radiation |
| PRC10 | DD15BCZ | Low Dose Rate (LDR) Brachytherapy of Colon using Californium 252 (Cf-252) | Treatment | Radiation |
| PRC10 | DD15BYZ | Low Dose Rate (LDR) Brachytherapy of Colon using Other Isotope | Treatment | Radiation |
| PRC10 | DD1797Z | High Dose Rate (HDR) Brachytherapy of Rectum using Cesium 137 (Cs-137) | Treatment | Radiation |
| PRC10 | DD1798Z | High Dose Rate (HDR) Brachytherapy of Rectum using Iridium 192 (Ir-192) | Treatment | Radiation |
| PRC10 | DD1799Z | High Dose Rate (HDR) Brachytherapy of Rectum using Iodine 125 (I-125) | Treatment | Radiation |
| PRC10 | DD179BZ | High Dose Rate (HDR) Brachytherapy of Rectum using Palladium 103 (Pd-103) | Treatment | Radiation |
| PRC10 | DD179CZ | High Dose Rate (HDR) Brachytherapy of Rectum using Californium 252 (Cf-252) | Treatment | Radiation |
| PRC10 | DD179YZ | High Dose Rate (HDR) Brachytherapy of Rectum using Other Isotope | Treatment | Radiation |
| PRC10 | DD17B7Z | Low Dose Rate (LDR) Brachytherapy of Rectum using Cesium 137 (Cs-137) | Treatment | Radiation |
| PRC10 | DD17B8Z | Low Dose Rate (LDR) Brachytherapy of Rectum using Iridium 192 (Ir-192) | Treatment | Radiation |
| PRC10 | DD17B9Z | Low Dose Rate (LDR) Brachytherapy of Rectum using Iodine 125 (I-125) | Treatment | Radiation |
| PRC10 | DD17BBZ | Low Dose Rate (LDR) Brachytherapy of Rectum using Palladium 103 (Pd-103) | Treatment | Radiation |
| PRC10 | DD17BCZ | Low Dose Rate (LDR) Brachytherapy of Rectum using Californium 252 (Cf-252) | Treatment | Radiation |
| PRC10 | DD17BYZ | Low Dose Rate (LDR) Brachytherapy of Rectum using Other Isotope | Treatment | Radiation |
| PRC10 | DDY07ZZ | Contact Radiation of Esophagus | Treatment | Radiation |
| PRC10 | DDY0FZZ | Plaque Radiation of Esophagus | Treatment | Radiation |
| PRC10 | DDY17ZZ | Contact Radiation of Stomach | Treatment | Radiation |
| PRC10 | DDY1CZZ | Intraoperative Radiation Therapy (IORT) of Stomach | Treatment | Radiation |
| PRC10 | DDY1FZZ | Plaque Radiation of Stomach | Treatment | Radiation |
| PRC10 | DDY27ZZ | Contact Radiation of Duodenum | Treatment | Radiation |
| PRC10 | DDY2CZZ | Intraoperative Radiation Therapy (IORT) of Duodenum | Treatment | Radiation |
| PRC10 | DDY2FZZ | Plaque Radiation of Duodenum | Treatment | Radiation |
| PRC10 | DDY37ZZ | Contact Radiation of Jejunum | Treatment | Radiation |
| PRC10 | DDY3CZZ | Intraoperative Radiation Therapy (IORT) of Jejunum | Treatment | Radiation |
| PRC10 | DDY3FZZ | Plaque Radiation of Jejunum | Treatment | Radiation |
| PRC10 | DDY47ZZ | Contact Radiation of Ileum | Treatment | Radiation |
| PRC10 | DDY4CZZ | Intraoperative Radiation Therapy (IORT) of Ileum | Treatment | Radiation |
| PRC10 | DDY4FZZ | Plaque Radiation of Ileum | Treatment | Radiation |
| PRC10 | DDY57ZZ | Contact Radiation of Colon | Treatment | Radiation |
| PRC10 | DDY5CZZ | Intraoperative Radiation Therapy (IORT) of Colon | Treatment | Radiation |
| PRC10 | DDY5FZZ | Plaque Radiation of Colon | Treatment | Radiation |
| PRC10 | DDY77ZZ | Contact Radiation of Rectum | Treatment | Radiation |
| PRC10 | DDY7CZZ | Intraoperative Radiation Therapy (IORT) of Rectum | Treatment | Radiation |
| PRC10 | DDY7FZZ | Plaque Radiation of Rectum | Treatment | Radiation |
| PRC10 | DDY8CZZ | Intraoperative Radiation Therapy (IORT) of Anus | Treatment | Radiation |
| PRC10 | DDY8FZZ | Plaque Radiation of Anus | Treatment | Radiation |
| PRC10 | DF000ZZ | Beam Radiation of Liver using Photons <1 MeV | Treatment | Radiation |
| PRC10 | DF003ZZ | Beam Radiation of Liver using Electrons | Treatment | Radiation |
| PRC10 | DF004ZZ | Beam Radiation of Liver using Heavy Particles (Protons,Ions) | Treatment | Radiation |
| PRC10 | DF005ZZ | Beam Radiation of Liver using Neutrons | Treatment | Radiation |
| PRC10 | DF010ZZ | Beam Radiation of Gallbladder using Photons <1 MeV | Treatment | Radiation |
| PRC10 | DF013ZZ | Beam Radiation of Gallbladder using Electrons | Treatment | Radiation |
| PRC10 | DF014ZZ | Beam Radiation of Gallbladder using Heavy Particles (Protons,Ions) | Treatment | Radiation |
| PRC10 | DF015ZZ | Beam Radiation of Gallbladder using Neutrons | Treatment | Radiation |
| PRC10 | DF020ZZ | Beam Radiation of Bile Ducts using Photons <1 MeV | Treatment | Radiation |
| PRC10 | DF023ZZ | Beam Radiation of Bile Ducts using Electrons | Treatment | Radiation |
| PRC10 | DF024ZZ | Beam Radiation of Bile Ducts using Heavy Particles (Protons,Ions) | Treatment | Radiation |
| PRC10 | DF025ZZ | Beam Radiation of Bile Ducts using Neutrons | Treatment | Radiation |
| PRC10 | DF030ZZ | Beam Radiation of Pancreas using Photons <1 MeV | Treatment | Radiation |
| PRC10 | DF033ZZ | Beam Radiation of Pancreas using Electrons | Treatment | Radiation |
| PRC10 | DF034ZZ | Beam Radiation of Pancreas using Heavy Particles (Protons,Ions) | Treatment | Radiation |
| PRC10 | DF035ZZ | Beam Radiation of Pancreas using Neutrons | Treatment | Radiation |
| PRC10 | DF1097Z | High Dose Rate (HDR) Brachytherapy of Liver using Cesium 137 (Cs-137) | Treatment | Radiation |
| PRC10 | DF1098Z | High Dose Rate (HDR) Brachytherapy of Liver using Iridium 192 (Ir-192) | Treatment | Radiation |
| PRC10 | DF1099Z | High Dose Rate (HDR) Brachytherapy of Liver using Iodine 125 (I-125) | Treatment | Radiation |
| PRC10 | DF109BZ | High Dose Rate (HDR) Brachytherapy of Liver using Palladium 103 (Pd-103) | Treatment | Radiation |
| PRC10 | DF109CZ | High Dose Rate (HDR) Brachytherapy of Liver using Californium 252 (Cf-252) | Treatment | Radiation |
| PRC10 | DF109YZ | High Dose Rate (HDR) Brachytherapy of Liver using Other Isotope | Treatment | Radiation |
| PRC10 | DF10B7Z | Low Dose Rate (LDR) Brachytherapy of Liver using Cesium 137 (Cs-137) | Treatment | Radiation |
| PRC10 | DF10B8Z | Low Dose Rate (LDR) Brachytherapy of Liver using Iridium 192 (Ir-192) | Treatment | Radiation |
| PRC10 | DF10B9Z | Low Dose Rate (LDR) Brachytherapy of Liver using Iodine 125 (I-125) | Treatment | Radiation |
| PRC10 | DF10BBZ | Low Dose Rate (LDR) Brachytherapy of Liver using Palladium 103 (Pd-103) | Treatment | Radiation |
| PRC10 | DF10BCZ | Low Dose Rate (LDR) Brachytherapy of Liver using Californium 252 (Cf-252) | Treatment | Radiation |
| PRC10 | DF10BYZ | Low Dose Rate (LDR) Brachytherapy of Liver using Other Isotope | Treatment | Radiation |
| PRC10 | DF1197Z | High Dose Rate (HDR) Brachytherapy of Gallbladder using Cesium 137 (Cs-137) | Treatment | Radiation |
| PRC10 | DF1198Z | High Dose Rate (HDR) Brachytherapy of Gallbladder using Iridium 192 (Ir-192) | Treatment | Radiation |
| PRC10 | DF1199Z | High Dose Rate (HDR) Brachytherapy of Gallbladder using Iodine 125 (I-125) | Treatment | Radiation |
| PRC10 | DF119BZ | High Dose Rate (HDR) Brachytherapy of Gallbladder using Palladium 103 (Pd-103) | Treatment | Radiation |
| PRC10 | DF119CZ | High Dose Rate (HDR) Brachytherapy of Gallbladder using Californium 252 (Cf-252) | Treatment | Radiation |
| PRC10 | DF119YZ | High Dose Rate (HDR) Brachytherapy of Gallbladder using Other Isotope | Treatment | Radiation |
| PRC10 | DF11B7Z | Low Dose Rate (LDR) Brachytherapy of Gallbladder using Cesium 137 (Cs-137) | Treatment | Radiation |
| PRC10 | DF11B8Z | Low Dose Rate (LDR) Brachytherapy of Gallbladder using Iridium 192 (Ir-192) | Treatment | Radiation |
| PRC10 | DF11B9Z | Low Dose Rate (LDR) Brachytherapy of Gallbladder using Iodine 125 (I-125) | Treatment | Radiation |
| PRC10 | DF11BBZ | Low Dose Rate (LDR) Brachytherapy of Gallbladder using Palladium 103 (Pd-103) | Treatment | Radiation |
| PRC10 | DF11BCZ | Low Dose Rate (LDR) Brachytherapy of Gallbladder using Californium 252 (Cf-252) | Treatment | Radiation |
| PRC10 | DF11BYZ | Low Dose Rate (LDR) Brachytherapy of Gallbladder using Other Isotope | Treatment | Radiation |
| PRC10 | DF1297Z | High Dose Rate (HDR) Brachytherapy of Bile Ducts using Cesium 137 (Cs-137) | Treatment | Radiation |
| PRC10 | DF1298Z | High Dose Rate (HDR) Brachytherapy of Bile Ducts using Iridium 192 (Ir-192) | Treatment | Radiation |
| PRC10 | DF1299Z | High Dose Rate (HDR) Brachytherapy of Bile Ducts using Iodine 125 (I-125) | Treatment | Radiation |
| PRC10 | DF129BZ | High Dose Rate (HDR) Brachytherapy of Bile Ducts using Palladium 103 (Pd-103) | Treatment | Radiation |
| PRC10 | DF129CZ | High Dose Rate (HDR) Brachytherapy of Bile Ducts using Californium 252 (Cf-252) | Treatment | Radiation |
| PRC10 | DF129YZ | High Dose Rate (HDR) Brachytherapy of Bile Ducts using Other Isotope | Treatment | Radiation |
| PRC10 | DF12B7Z | Low Dose Rate (LDR) Brachytherapy of Bile Ducts using Cesium 137 (Cs-137) | Treatment | Radiation |
| PRC10 | DF12B8Z | Low Dose Rate (LDR) Brachytherapy of Bile Ducts using Iridium 192 (Ir-192) | Treatment | Radiation |
| PRC10 | DF12B9Z | Low Dose Rate (LDR) Brachytherapy of Bile Ducts using Iodine 125 (I-125) | Treatment | Radiation |
| PRC10 | DF12BBZ | Low Dose Rate (LDR) Brachytherapy of Bile Ducts using Palladium 103 (Pd-103) | Treatment | Radiation |
| PRC10 | DF12BCZ | Low Dose Rate (LDR) Brachytherapy of Bile Ducts using Californium 252 (Cf-252) | Treatment | Radiation |
| PRC10 | DF12BYZ | Low Dose Rate (LDR) Brachytherapy of Bile Ducts using Other Isotope | Treatment | Radiation |
| PRC10 | DF1397Z | High Dose Rate (HDR) Brachytherapy of Pancreas using Cesium 137 (Cs-137) | Treatment | Radiation |
| PRC10 | DF1398Z | High Dose Rate (HDR) Brachytherapy of Pancreas using Iridium 192 (Ir-192) | Treatment | Radiation |
| PRC10 | DF1399Z | High Dose Rate (HDR) Brachytherapy of Pancreas using Iodine 125 (I-125) | Treatment | Radiation |
| PRC10 | DF139BZ | High Dose Rate (HDR) Brachytherapy of Pancreas using Palladium 103 (Pd-103) | Treatment | Radiation |
| PRC10 | DF139CZ | High Dose Rate (HDR) Brachytherapy of Pancreas using Californium 252 (Cf-252) | Treatment | Radiation |
| PRC10 | DF139YZ | High Dose Rate (HDR) Brachytherapy of Pancreas using Other Isotope | Treatment | Radiation |
| PRC10 | DF13B7Z | Low Dose Rate (LDR) Brachytherapy of Pancreas using Cesium 137 (Cs-137) | Treatment | Radiation |
| PRC10 | DF13B8Z | Low Dose Rate (LDR) Brachytherapy of Pancreas using Iridium 192 (Ir-192) | Treatment | Radiation |
| PRC10 | DF13B9Z | Low Dose Rate (LDR) Brachytherapy of Pancreas using Iodine 125 (I-125) | Treatment | Radiation |
| PRC10 | DF13BBZ | Low Dose Rate (LDR) Brachytherapy of Pancreas using Palladium 103 (Pd-103) | Treatment | Radiation |
| PRC10 | DF13BCZ | Low Dose Rate (LDR) Brachytherapy of Pancreas using Californium 252 (Cf-252) | Treatment | Radiation |
| PRC10 | DF13BYZ | Low Dose Rate (LDR) Brachytherapy of Pancreas using Other Isotope | Treatment | Radiation |
| PRC10 | DFY07ZZ | Contact Radiation of Liver | Treatment | Radiation |
| PRC10 | DFY0CZZ | Intraoperative Radiation Therapy (IORT) of Liver | Treatment | Radiation |
| PRC10 | DFY0FZZ | Plaque Radiation of Liver | Treatment | Radiation |
| PRC10 | DFY17ZZ | Contact Radiation of Gallbladder | Treatment | Radiation |
| PRC10 | DFY1CZZ | Intraoperative Radiation Therapy (IORT) of Gallbladder | Treatment | Radiation |
| PRC10 | DFY1FZZ | Plaque Radiation of Gallbladder | Treatment | Radiation |
| PRC10 | DFY27ZZ | Contact Radiation of Bile Ducts | Treatment | Radiation |
| PRC10 | DFY2CZZ | Intraoperative Radiation Therapy (IORT) of Bile Ducts | Treatment | Radiation |
| PRC10 | DFY2FZZ | Plaque Radiation of Bile Ducts | Treatment | Radiation |
| PRC10 | DFY37ZZ | Contact Radiation of Pancreas | Treatment | Radiation |
| PRC10 | DFY3CZZ | Intraoperative Radiation Therapy (IORT) of Pancreas | Treatment | Radiation |
| PRC10 | DFY3FZZ | Plaque Radiation of Pancreas | Treatment | Radiation |
| PRC10 | DG000ZZ | Beam Radiation of Pituitary Gland using Photons <1 MeV | Treatment | Radiation |
| PRC10 | DG003ZZ | Beam Radiation of Pituitary Gland using Electrons | Treatment | Radiation |
| PRC10 | DG005ZZ | Beam Radiation of Pituitary Gland using Neutrons | Treatment | Radiation |
| PRC10 | DG010ZZ | Beam Radiation of Pineal Body using Photons <1 MeV | Treatment | Radiation |
| PRC10 | DG013ZZ | Beam Radiation of Pineal Body using Electrons | Treatment | Radiation |
| PRC10 | DG015ZZ | Beam Radiation of Pineal Body using Neutrons | Treatment | Radiation |
| PRC10 | DG020ZZ | Beam Radiation of Adrenal Glands using Photons <1 MeV | Treatment | Radiation |
| PRC10 | DG023ZZ | Beam Radiation of Adrenal Glands using Electrons | Treatment | Radiation |
| PRC10 | DG025ZZ | Beam Radiation of Adrenal Glands using Neutrons | Treatment | Radiation |
| PRC10 | DG040ZZ | Beam Radiation of Parathyroid Glands using Photons <1 MeV | Treatment | Radiation |
| PRC10 | DG043ZZ | Beam Radiation of Parathyroid Glands using Electrons | Treatment | Radiation |
| PRC10 | DG045ZZ | Beam Radiation of Parathyroid Glands using Neutrons | Treatment | Radiation |
| PRC10 | DG050ZZ | Beam Radiation of Thyroid using Photons <1 MeV | Treatment | Radiation |
| PRC10 | DG053ZZ | Beam Radiation of Thyroid using Electrons | Treatment | Radiation |
| PRC10 | DG055ZZ | Beam Radiation of Thyroid using Neutrons | Treatment | Radiation |
| PRC10 | DG1097Z | High Dose Rate (HDR) Brachytherapy of Pituitary Gland using Cesium 137 (Cs-137) | Treatment | Radiation |
| PRC10 | DG1098Z | High Dose Rate (HDR) Brachytherapy of Pituitary Gland using Iridium 192 (Ir-192) | Treatment | Radiation |
| PRC10 | DG1099Z | High Dose Rate (HDR) Brachytherapy of Pituitary Gland using Iodine 125 (I-125) | Treatment | Radiation |
| PRC10 | DG109BZ | High Dose Rate (HDR) Brachytherapy of Pituitary Gland using Palladium 103 (Pd-103) | Treatment | Radiation |
| PRC10 | DG109CZ | High Dose Rate (HDR) Brachytherapy of Pituitary Gland using Californium 252 (Cf-252) | Treatment | Radiation |
| PRC10 | DG109YZ | High Dose Rate (HDR) Brachytherapy of Pituitary Gland using Other Isotope | Treatment | Radiation |
| PRC10 | DG10B7Z | Low Dose Rate (LDR) Brachytherapy of Pituitary Gland using Cesium 137 (Cs-137) | Treatment | Radiation |
| PRC10 | DG10B8Z | Low Dose Rate (LDR) Brachytherapy of Pituitary Gland using Iridium 192 (Ir-192) | Treatment | Radiation |
| PRC10 | DG10B9Z | Low Dose Rate (LDR) Brachytherapy of Pituitary Gland using Iodine 125 (I-125) | Treatment | Radiation |
| PRC10 | DG10BBZ | Low Dose Rate (LDR) Brachytherapy of Pituitary Gland using Palladium 103 (Pd-103) | Treatment | Radiation |
| PRC10 | DG10BCZ | Low Dose Rate (LDR) Brachytherapy of Pituitary Gland using Californium 252 (Cf-252) | Treatment | Radiation |
| PRC10 | DG10BYZ | Low Dose Rate (LDR) Brachytherapy of Pituitary Gland using Other Isotope | Treatment | Radiation |
| PRC10 | DG1197Z | High Dose Rate (HDR) Brachytherapy of Pineal Body using Cesium 137 (Cs-137) | Treatment | Radiation |
| PRC10 | DG1198Z | High Dose Rate (HDR) Brachytherapy of Pineal Body using Iridium 192 (Ir-192) | Treatment | Radiation |
| PRC10 | DG1199Z | High Dose Rate (HDR) Brachytherapy of Pineal Body using Iodine 125 (I-125) | Treatment | Radiation |
| PRC10 | DG119BZ | High Dose Rate (HDR) Brachytherapy of Pineal Body using Palladium 103 (Pd-103) | Treatment | Radiation |
| PRC10 | DG119CZ | High Dose Rate (HDR) Brachytherapy of Pineal Body using Californium 252 (Cf-252) | Treatment | Radiation |
| PRC10 | DG119YZ | High Dose Rate (HDR) Brachytherapy of Pineal Body using Other Isotope | Treatment | Radiation |
| PRC10 | DG11B7Z | Low Dose Rate (LDR) Brachytherapy of Pineal Body using Cesium 137 (Cs-137) | Treatment | Radiation |
| PRC10 | DG11B8Z | Low Dose Rate (LDR) Brachytherapy of Pineal Body using Iridium 192 (Ir-192) | Treatment | Radiation |
| PRC10 | DG11B9Z | Low Dose Rate (LDR) Brachytherapy of Pineal Body using Iodine 125 (I-125) | Treatment | Radiation |
| PRC10 | DG11BBZ | Low Dose Rate (LDR) Brachytherapy of Pineal Body using Palladium 103 (Pd-103) | Treatment | Radiation |
| PRC10 | DG11BCZ | Low Dose Rate (LDR) Brachytherapy of Pineal Body using Californium 252 (Cf-252) | Treatment | Radiation |
| PRC10 | DG11BYZ | Low Dose Rate (LDR) Brachytherapy of Pineal Body using Other Isotope | Treatment | Radiation |
| PRC10 | DG1297Z | High Dose Rate (HDR) Brachytherapy of Adrenal Glands using Cesium 137 (Cs-137) | Treatment | Radiation |
| PRC10 | DG1298Z | High Dose Rate (HDR) Brachytherapy of Adrenal Glands using Iridium 192 (Ir-192) | Treatment | Radiation |
| PRC10 | DG1299Z | High Dose Rate (HDR) Brachytherapy of Adrenal Glands using Iodine 125 (I-125) | Treatment | Radiation |
| PRC10 | DG129BZ | High Dose Rate (HDR) Brachytherapy of Adrenal Glands using Palladium 103 (Pd-103) | Treatment | Radiation |
| PRC10 | DG129CZ | High Dose Rate (HDR) Brachytherapy of Adrenal Glands using Californium 252 (Cf-252) | Treatment | Radiation |
| PRC10 | DG129YZ | High Dose Rate (HDR) Brachytherapy of Adrenal Glands using Other Isotope | Treatment | Radiation |
| PRC10 | DG12B7Z | Low Dose Rate (LDR) Brachytherapy of Adrenal Glands using Cesium 137 (Cs-137) | Treatment | Radiation |
| PRC10 | DG12B8Z | Low Dose Rate (LDR) Brachytherapy of Adrenal Glands using Iridium 192 (Ir-192) | Treatment | Radiation |
| PRC10 | DG12B9Z | Low Dose Rate (LDR) Brachytherapy of Adrenal Glands using Iodine 125 (I-125) | Treatment | Radiation |
| PRC10 | DG12BBZ | Low Dose Rate (LDR) Brachytherapy of Adrenal Glands using Palladium 103 (Pd-103) | Treatment | Radiation |
| PRC10 | DG12BCZ | Low Dose Rate (LDR) Brachytherapy of Adrenal Glands using Californium 252 (Cf-252) | Treatment | Radiation |
| PRC10 | DG12BYZ | Low Dose Rate (LDR) Brachytherapy of Adrenal Glands using Other Isotope | Treatment | Radiation |
| PRC10 | DG1497Z | High Dose Rate (HDR) Brachytherapy of Parathyroid Glands using Cesium 137 (Cs-137) | Treatment | Radiation |
| PRC10 | DG1498Z | High Dose Rate (HDR) Brachytherapy of Parathyroid Glands using Iridium 192 (Ir-192) | Treatment | Radiation |
| PRC10 | DG1499Z | High Dose Rate (HDR) Brachytherapy of Parathyroid Glands using Iodine 125 (I-125) | Treatment | Radiation |
| PRC10 | DG149BZ | High Dose Rate (HDR) Brachytherapy of Parathyroid Glands using Palladium 103 (Pd-103) | Treatment | Radiation |
| PRC10 | DG149CZ | High Dose Rate (HDR) Brachytherapy of Parathyroid Glands using Californium 252 (Cf-252) | Treatment | Radiation |
| PRC10 | DG149YZ | High Dose Rate (HDR) Brachytherapy of Parathyroid Glands using Other Isotope | Treatment | Radiation |
| PRC10 | DG14B7Z | Low Dose Rate (LDR) Brachytherapy of Parathyroid Glands using Cesium 137 (Cs-137) | Treatment | Radiation |
| PRC10 | DG14B8Z | Low Dose Rate (LDR) Brachytherapy of Parathyroid Glands using Iridium 192 (Ir-192) | Treatment | Radiation |
| PRC10 | DG14B9Z | Low Dose Rate (LDR) Brachytherapy of Parathyroid Glands using Iodine 125 (I-125) | Treatment | Radiation |
| PRC10 | DG14BBZ | Low Dose Rate (LDR) Brachytherapy of Parathyroid Glands using Palladium 103 (Pd-103) | Treatment | Radiation |
| PRC10 | DG14BCZ | Low Dose Rate (LDR) Brachytherapy of Parathyroid Glands using Californium 252 (Cf-252) | Treatment | Radiation |
| PRC10 | DG14BYZ | Low Dose Rate (LDR) Brachytherapy of Parathyroid Glands using Other Isotope | Treatment | Radiation |
| PRC10 | DG1597Z | High Dose Rate (HDR) Brachytherapy of Thyroid using Cesium 137 (Cs-137) | Treatment | Radiation |
| PRC10 | DG1598Z | High Dose Rate (HDR) Brachytherapy of Thyroid using Iridium 192 (Ir-192) | Treatment | Radiation |
| PRC10 | DG1599Z | High Dose Rate (HDR) Brachytherapy of Thyroid using Iodine 125 (I-125) | Treatment | Radiation |
| PRC10 | DG159BZ | High Dose Rate (HDR) Brachytherapy of Thyroid using Palladium 103 (Pd-103) | Treatment | Radiation |
| PRC10 | DG159CZ | High Dose Rate (HDR) Brachytherapy of Thyroid using Californium 252 (Cf-252) | Treatment | Radiation |
| PRC10 | DG159YZ | High Dose Rate (HDR) Brachytherapy of Thyroid using Other Isotope | Treatment | Radiation |
| PRC10 | DG15B7Z | Low Dose Rate (LDR) Brachytherapy of Thyroid using Cesium 137 (Cs-137) | Treatment | Radiation |
| PRC10 | DG15B8Z | Low Dose Rate (LDR) Brachytherapy of Thyroid using Iridium 192 (Ir-192) | Treatment | Radiation |
| PRC10 | DG15B9Z | Low Dose Rate (LDR) Brachytherapy of Thyroid using Iodine 125 (I-125) | Treatment | Radiation |
| PRC10 | DG15BBZ | Low Dose Rate (LDR) Brachytherapy of Thyroid using Palladium 103 (Pd-103) | Treatment | Radiation |
| PRC10 | DG15BCZ | Low Dose Rate (LDR) Brachytherapy of Thyroid using Californium 252 (Cf-252) | Treatment | Radiation |
| PRC10 | DG15BYZ | Low Dose Rate (LDR) Brachytherapy of Thyroid using Other Isotope | Treatment | Radiation |
| PRC10 | DGY07ZZ | Contact Radiation of Pituitary Gland | Treatment | Radiation |
| PRC10 | DGY0FZZ | Plaque Radiation of Pituitary Gland | Treatment | Radiation |
| PRC10 | DGY17ZZ | Contact Radiation of Pineal Body | Treatment | Radiation |
| PRC10 | DGY1FZZ | Plaque Radiation of Pineal Body | Treatment | Radiation |
| PRC10 | DGY27ZZ | Contact Radiation of Adrenal Glands | Treatment | Radiation |
| PRC10 | DGY2FZZ | Plaque Radiation of Adrenal Glands | Treatment | Radiation |
| PRC10 | DGY47ZZ | Contact Radiation of Parathyroid Glands | Treatment | Radiation |
| PRC10 | DGY4FZZ | Plaque Radiation of Parathyroid Glands | Treatment | Radiation |
| PRC10 | DGY57ZZ | Contact Radiation of Thyroid | Treatment | Radiation |
| PRC10 | DGY5FZZ | Plaque Radiation of Thyroid | Treatment | Radiation |
| PRC10 | DM000ZZ | Beam Radiation of Left Breast using Photons <1 MeV | Treatment | Radiation |
| PRC10 | DM003ZZ | Beam Radiation of Left Breast using Electrons | Treatment | Radiation |
| PRC10 | DM004ZZ | Beam Radiation of Left Breast using Heavy Particles (Protons,Ions) | Treatment | Radiation |
| PRC10 | DM005ZZ | Beam Radiation of Left Breast using Neutrons | Treatment | Radiation |
| PRC10 | DM010ZZ | Beam Radiation of Right Breast using Photons <1 MeV | Treatment | Radiation |
| PRC10 | DM013ZZ | Beam Radiation of Right Breast using Electrons | Treatment | Radiation |
| PRC10 | DM014ZZ | Beam Radiation of Right Breast using Heavy Particles (Protons,Ions) | Treatment | Radiation |
| PRC10 | DM015ZZ | Beam Radiation of Right Breast using Neutrons | Treatment | Radiation |
| PRC10 | DM1097Z | High Dose Rate (HDR) Brachytherapy of Left Breast using Cesium 137 (Cs-137) | Treatment | Radiation |
| PRC10 | DM1098Z | High Dose Rate (HDR) Brachytherapy of Left Breast using Iridium 192 (Ir-192) | Treatment | Radiation |
| PRC10 | DM1099Z | High Dose Rate (HDR) Brachytherapy of Left Breast using Iodine 125 (I-125) | Treatment | Radiation |
| PRC10 | DM109BZ | High Dose Rate (HDR) Brachytherapy of Left Breast using Palladium 103 (Pd-103) | Treatment | Radiation |
| PRC10 | DM109CZ | High Dose Rate (HDR) Brachytherapy of Left Breast using Californium 252 (Cf-252) | Treatment | Radiation |
| PRC10 | DM109YZ | High Dose Rate (HDR) Brachytherapy of Left Breast using Other Isotope | Treatment | Radiation |
| PRC10 | DM10B7Z | Low Dose Rate (LDR) Brachytherapy of Left Breast using Cesium 137 (Cs-137) | Treatment | Radiation |
| PRC10 | DM10B8Z | Low Dose Rate (LDR) Brachytherapy of Left Breast using Iridium 192 (Ir-192) | Treatment | Radiation |
| PRC10 | DM10B9Z | Low Dose Rate (LDR) Brachytherapy of Left Breast using Iodine 125 (I-125) | Treatment | Radiation |
| PRC10 | DM10BBZ | Low Dose Rate (LDR) Brachytherapy of Left Breast using Palladium 103 (Pd-103) | Treatment | Radiation |
| PRC10 | DM10BCZ | Low Dose Rate (LDR) Brachytherapy of Left Breast using Californium 252 (Cf-252) | Treatment | Radiation |
| PRC10 | DM10BYZ | Low Dose Rate (LDR) Brachytherapy of Left Breast using Other Isotope | Treatment | Radiation |
| PRC10 | DM1197Z | High Dose Rate (HDR) Brachytherapy of Right Breast using Cesium 137 (Cs-137) | Treatment | Radiation |
| PRC10 | DM1198Z | High Dose Rate (HDR) Brachytherapy of Right Breast using Iridium 192 (Ir-192) | Treatment | Radiation |
| PRC10 | DM1199Z | High Dose Rate (HDR) Brachytherapy of Right Breast using Iodine 125 (I-125) | Treatment | Radiation |
| PRC10 | DM119BZ | High Dose Rate (HDR) Brachytherapy of Right Breast using Palladium 103 (Pd-103) | Treatment | Radiation |
| PRC10 | DM119CZ | High Dose Rate (HDR) Brachytherapy of Right Breast using Californium 252 (Cf-252) | Treatment | Radiation |
| PRC10 | DM119YZ | High Dose Rate (HDR) Brachytherapy of Right Breast using Other Isotope | Treatment | Radiation |
| PRC10 | DM11B7Z | Low Dose Rate (LDR) Brachytherapy of Right Breast using Cesium 137 (Cs-137) | Treatment | Radiation |
| PRC10 | DM11B8Z | Low Dose Rate (LDR) Brachytherapy of Right Breast using Iridium 192 (Ir-192) | Treatment | Radiation |
| PRC10 | DM11B9Z | Low Dose Rate (LDR) Brachytherapy of Right Breast using Iodine 125 (I-125) | Treatment | Radiation |
| PRC10 | DM11BBZ | Low Dose Rate (LDR) Brachytherapy of Right Breast using Palladium 103 (Pd-103) | Treatment | Radiation |
| PRC10 | DM11BCZ | Low Dose Rate (LDR) Brachytherapy of Right Breast using Californium 252 (Cf-252) | Treatment | Radiation |
| PRC10 | DM11BYZ | Low Dose Rate (LDR) Brachytherapy of Right Breast using Other Isotope | Treatment | Radiation |
| PRC10 | DMY07ZZ | Contact Radiation of Left Breast | Treatment | Radiation |
| PRC10 | DMY0FZZ | Plaque Radiation of Left Breast | Treatment | Radiation |
| PRC10 | DMY17ZZ | Contact Radiation of Right Breast | Treatment | Radiation |
| PRC10 | DMY1FZZ | Plaque Radiation of Right Breast | Treatment | Radiation |
| PRC10 | DP000ZZ | Beam Radiation of Skull using Photons <1 MeV | Treatment | Radiation |
| PRC10 | DP003ZZ | Beam Radiation of Skull using Electrons | Treatment | Radiation |
| PRC10 | DP004ZZ | Beam Radiation of Skull using Heavy Particles (Protons,Ions) | Treatment | Radiation |
| PRC10 | DP005ZZ | Beam Radiation of Skull using Neutrons | Treatment | Radiation |
| PRC10 | DP020ZZ | Beam Radiation of Maxilla using Photons <1 MeV | Treatment | Radiation |
| PRC10 | DP023ZZ | Beam Radiation of Maxilla using Electrons | Treatment | Radiation |
| PRC10 | DP024ZZ | Beam Radiation of Maxilla using Heavy Particles (Protons,Ions) | Treatment | Radiation |
| PRC10 | DP025ZZ | Beam Radiation of Maxilla using Neutrons | Treatment | Radiation |
| PRC10 | DP030ZZ | Beam Radiation of Mandible using Photons <1 MeV | Treatment | Radiation |
| PRC10 | DP033ZZ | Beam Radiation of Mandible using Electrons | Treatment | Radiation |
| PRC10 | DP034ZZ | Beam Radiation of Mandible using Heavy Particles (Protons,Ions) | Treatment | Radiation |
| PRC10 | DP035ZZ | Beam Radiation of Mandible using Neutrons | Treatment | Radiation |
| PRC10 | DP040ZZ | Beam Radiation of Sternum using Photons <1 MeV | Treatment | Radiation |
| PRC10 | DP043ZZ | Beam Radiation of Sternum using Electrons | Treatment | Radiation |
| PRC10 | DP044ZZ | Beam Radiation of Sternum using Heavy Particles (Protons,Ions) | Treatment | Radiation |
| PRC10 | DP045ZZ | Beam Radiation of Sternum using Neutrons | Treatment | Radiation |
| PRC10 | DP050ZZ | Beam Radiation of Rib(s) using Photons <1 MeV | Treatment | Radiation |
| PRC10 | DP053ZZ | Beam Radiation of Rib(s) using Electrons | Treatment | Radiation |
| PRC10 | DP054ZZ | Beam Radiation of Rib(s) using Heavy Particles (Protons,Ions) | Treatment | Radiation |
| PRC10 | DP055ZZ | Beam Radiation of Rib(s) using Neutrons | Treatment | Radiation |
| PRC10 | DP060ZZ | Beam Radiation of Humerus using Photons <1 MeV | Treatment | Radiation |
| PRC10 | DP063ZZ | Beam Radiation of Humerus using Electrons | Treatment | Radiation |
| PRC10 | DP064ZZ | Beam Radiation of Humerus using Heavy Particles (Protons,Ions) | Treatment | Radiation |
| PRC10 | DP065ZZ | Beam Radiation of Humerus using Neutrons | Treatment | Radiation |
| PRC10 | DP070ZZ | Beam Radiation of Radius/Ulna using Photons <1 MeV | Treatment | Radiation |
| PRC10 | DP073ZZ | Beam Radiation of Radius/Ulna using Electrons | Treatment | Radiation |
| PRC10 | DP074ZZ | Beam Radiation of Radius/Ulna using Heavy Particles (Protons,Ions) | Treatment | Radiation |
| PRC10 | DP075ZZ | Beam Radiation of Radius/Ulna using Neutrons | Treatment | Radiation |
| PRC10 | DP080ZZ | Beam Radiation of Pelvic Bones using Photons <1 MeV | Treatment | Radiation |
| PRC10 | DP083ZZ | Beam Radiation of Pelvic Bones using Electrons | Treatment | Radiation |
| PRC10 | DP084ZZ | Beam Radiation of Pelvic Bones using Heavy Particles (Protons,Ions) | Treatment | Radiation |
| PRC10 | DP085ZZ | Beam Radiation of Pelvic Bones using Neutrons | Treatment | Radiation |
| PRC10 | DP090ZZ | Beam Radiation of Femur using Photons <1 MeV | Treatment | Radiation |
| PRC10 | DP093ZZ | Beam Radiation of Femur using Electrons | Treatment | Radiation |
| PRC10 | DP094ZZ | Beam Radiation of Femur using Heavy Particles (Protons,Ions) | Treatment | Radiation |
| PRC10 | DP095ZZ | Beam Radiation of Femur using Neutrons | Treatment | Radiation |
| PRC10 | DP0B0ZZ | Beam Radiation of Tibia/Fibula using Photons <1 MeV | Treatment | Radiation |
| PRC10 | DP0B3ZZ | Beam Radiation of Tibia/Fibula using Electrons | Treatment | Radiation |
| PRC10 | DP0B4ZZ | Beam Radiation of Tibia/Fibula using Heavy Particles (Protons,Ions) | Treatment | Radiation |
| PRC10 | DP0B5ZZ | Beam Radiation of Tibia/Fibula using Neutrons | Treatment | Radiation |
| PRC10 | DP0C0ZZ | Beam Radiation of Other Bone using Photons <1 MeV | Treatment | Radiation |
| PRC10 | DP0C3ZZ | Beam Radiation of Other Bone using Electrons | Treatment | Radiation |
| PRC10 | DP0C4ZZ | Beam Radiation of Other Bone using Heavy Particles (Protons,Ions) | Treatment | Radiation |
| PRC10 | DP0C5ZZ | Beam Radiation of Other Bone using Neutrons | Treatment | Radiation |
| PRC10 | DPY07ZZ | Contact Radiation of Skull | Treatment | Radiation |
| PRC10 | DPY0FZZ | Plaque Radiation of Skull | Treatment | Radiation |
| PRC10 | DPY27ZZ | Contact Radiation of Maxilla | Treatment | Radiation |
| PRC10 | DPY2FZZ | Plaque Radiation of Maxilla | Treatment | Radiation |
| PRC10 | DPY37ZZ | Contact Radiation of Mandible | Treatment | Radiation |
| PRC10 | DPY3FZZ | Plaque Radiation of Mandible | Treatment | Radiation |
| PRC10 | DPY47ZZ | Contact Radiation of Sternum | Treatment | Radiation |
| PRC10 | DPY4FZZ | Plaque Radiation of Sternum | Treatment | Radiation |
| PRC10 | DPY57ZZ | Contact Radiation of Rib(s) | Treatment | Radiation |
| PRC10 | DPY5FZZ | Plaque Radiation of Rib(s) | Treatment | Radiation |
| PRC10 | DPY67ZZ | Contact Radiation of Humerus | Treatment | Radiation |
| PRC10 | DPY6FZZ | Plaque Radiation of Humerus | Treatment | Radiation |
| PRC10 | DPY77ZZ | Contact Radiation of Radius/Ulna | Treatment | Radiation |
| PRC10 | DPY7FZZ | Plaque Radiation of Radius/Ulna | Treatment | Radiation |
| PRC10 | DPY87ZZ | Contact Radiation of Pelvic Bones | Treatment | Radiation |
| PRC10 | DPY8FZZ | Plaque Radiation of Pelvic Bones | Treatment | Radiation |
| PRC10 | DPY97ZZ | Contact Radiation of Femur | Treatment | Radiation |
| PRC10 | DPY9FZZ | Plaque Radiation of Femur | Treatment | Radiation |
| PRC10 | DPYB7ZZ | Contact Radiation of Tibia/Fibula | Treatment | Radiation |
| PRC10 | DPYBFZZ | Plaque Radiation of Tibia/Fibula | Treatment | Radiation |
| PRC10 | DPYC7ZZ | Contact Radiation of Other Bone | Treatment | Radiation |
| PRC10 | DPYCFZZ | Plaque Radiation of Other Bone | Treatment | Radiation |
| PRC10 | DT000ZZ | Beam Radiation of Kidney using Photons <1 MeV | Treatment | Radiation |
| PRC10 | DT003ZZ | Beam Radiation of Kidney using Electrons | Treatment | Radiation |
| PRC10 | DT004ZZ | Beam Radiation of Kidney using Heavy Particles (Protons,Ions) | Treatment | Radiation |
| PRC10 | DT005ZZ | Beam Radiation of Kidney using Neutrons | Treatment | Radiation |
| PRC10 | DT010ZZ | Beam Radiation of Ureter using Photons <1 MeV | Treatment | Radiation |
| PRC10 | DT013ZZ | Beam Radiation of Ureter using Electrons | Treatment | Radiation |
| PRC10 | DT014ZZ | Beam Radiation of Ureter using Heavy Particles (Protons,Ions) | Treatment | Radiation |
| PRC10 | DT015ZZ | Beam Radiation of Ureter using Neutrons | Treatment | Radiation |
| PRC10 | DT020ZZ | Beam Radiation of Bladder using Photons <1 MeV | Treatment | Radiation |
| PRC10 | DT023ZZ | Beam Radiation of Bladder using Electrons | Treatment | Radiation |
| PRC10 | DT024ZZ | Beam Radiation of Bladder using Heavy Particles (Protons,Ions) | Treatment | Radiation |
| PRC10 | DT025ZZ | Beam Radiation of Bladder using Neutrons | Treatment | Radiation |
| PRC10 | DT030ZZ | Beam Radiation of Urethra using Photons <1 MeV | Treatment | Radiation |
| PRC10 | DT033ZZ | Beam Radiation of Urethra using Electrons | Treatment | Radiation |
| PRC10 | DT034ZZ | Beam Radiation of Urethra using Heavy Particles (Protons,Ions) | Treatment | Radiation |
| PRC10 | DT035ZZ | Beam Radiation of Urethra using Neutrons | Treatment | Radiation |
| PRC10 | DT1097Z | High Dose Rate (HDR) Brachytherapy of Kidney using Cesium 137 (Cs-137) | Treatment | Radiation |
| PRC10 | DT1098Z | High Dose Rate (HDR) Brachytherapy of Kidney using Iridium 192 (Ir-192) | Treatment | Radiation |
| PRC10 | DT1099Z | High Dose Rate (HDR) Brachytherapy of Kidney using Iodine 125 (I-125) | Treatment | Radiation |
| PRC10 | DT109BZ | High Dose Rate (HDR) Brachytherapy of Kidney using Palladium 103 (Pd-103) | Treatment | Radiation |
| PRC10 | DT109CZ | High Dose Rate (HDR) Brachytherapy of Kidney using Californium 252 (Cf-252) | Treatment | Radiation |
| PRC10 | DT109YZ | High Dose Rate (HDR) Brachytherapy of Kidney using Other Isotope | Treatment | Radiation |
| PRC10 | DT10B7Z | Low Dose Rate (LDR) Brachytherapy of Kidney using Cesium 137 (Cs-137) | Treatment | Radiation |
| PRC10 | DT10B8Z | Low Dose Rate (LDR) Brachytherapy of Kidney using Iridium 192 (Ir-192) | Treatment | Radiation |
| PRC10 | DT10B9Z | Low Dose Rate (LDR) Brachytherapy of Kidney using Iodine 125 (I-125) | Treatment | Radiation |
| PRC10 | DT10BBZ | Low Dose Rate (LDR) Brachytherapy of Kidney using Palladium 103 (Pd-103) | Treatment | Radiation |
| PRC10 | DT10BCZ | Low Dose Rate (LDR) Brachytherapy of Kidney using Californium 252 (Cf-252) | Treatment | Radiation |
| PRC10 | DT10BYZ | Low Dose Rate (LDR) Brachytherapy of Kidney using Other Isotope | Treatment | Radiation |
| PRC10 | DT1197Z | High Dose Rate (HDR) Brachytherapy of Ureter using Cesium 137 (Cs-137) | Treatment | Radiation |
| PRC10 | DT1198Z | High Dose Rate (HDR) Brachytherapy of Ureter using Iridium 192 (Ir-192) | Treatment | Radiation |
| PRC10 | DT1199Z | High Dose Rate (HDR) Brachytherapy of Ureter using Iodine 125 (I-125) | Treatment | Radiation |
| PRC10 | DT119BZ | High Dose Rate (HDR) Brachytherapy of Ureter using Palladium 103 (Pd-103) | Treatment | Radiation |
| PRC10 | DT119CZ | High Dose Rate (HDR) Brachytherapy of Ureter using Californium 252 (Cf-252) | Treatment | Radiation |
| PRC10 | DT119YZ | High Dose Rate (HDR) Brachytherapy of Ureter using Other Isotope | Treatment | Radiation |
| PRC10 | DT11B7Z | Low Dose Rate (LDR) Brachytherapy of Ureter using Cesium 137 (Cs-137) | Treatment | Radiation |
| PRC10 | DT11B8Z | Low Dose Rate (LDR) Brachytherapy of Ureter using Iridium 192 (Ir-192) | Treatment | Radiation |
| PRC10 | DT11B9Z | Low Dose Rate (LDR) Brachytherapy of Ureter using Iodine 125 (I-125) | Treatment | Radiation |
| PRC10 | DT11BBZ | Low Dose Rate (LDR) Brachytherapy of Ureter using Palladium 103 (Pd-103) | Treatment | Radiation |
| PRC10 | DT11BCZ | Low Dose Rate (LDR) Brachytherapy of Ureter using Californium 252 (Cf-252) | Treatment | Radiation |
| PRC10 | DT11BYZ | Low Dose Rate (LDR) Brachytherapy of Ureter using Other Isotope | Treatment | Radiation |
| PRC10 | DT1297Z | High Dose Rate (HDR) Brachytherapy of Bladder using Cesium 137 (Cs-137) | Treatment | Radiation |
| PRC10 | DT1298Z | High Dose Rate (HDR) Brachytherapy of Bladder using Iridium 192 (Ir-192) | Treatment | Radiation |
| PRC10 | DT1299Z | High Dose Rate (HDR) Brachytherapy of Bladder using Iodine 125 (I-125) | Treatment | Radiation |
| PRC10 | DT129BZ | High Dose Rate (HDR) Brachytherapy of Bladder using Palladium 103 (Pd-103) | Treatment | Radiation |
| PRC10 | DT129CZ | High Dose Rate (HDR) Brachytherapy of Bladder using Californium 252 (Cf-252) | Treatment | Radiation |
| PRC10 | DT129YZ | High Dose Rate (HDR) Brachytherapy of Bladder using Other Isotope | Treatment | Radiation |
| PRC10 | DT12B7Z | Low Dose Rate (LDR) Brachytherapy of Bladder using Cesium 137 (Cs-137) | Treatment | Radiation |
| PRC10 | DT12B8Z | Low Dose Rate (LDR) Brachytherapy of Bladder using Iridium 192 (Ir-192) | Treatment | Radiation |
| PRC10 | DT12B9Z | Low Dose Rate (LDR) Brachytherapy of Bladder using Iodine 125 (I-125) | Treatment | Radiation |
| PRC10 | DT12BBZ | Low Dose Rate (LDR) Brachytherapy of Bladder using Palladium 103 (Pd-103) | Treatment | Radiation |
| PRC10 | DT12BCZ | Low Dose Rate (LDR) Brachytherapy of Bladder using Californium 252 (Cf-252) | Treatment | Radiation |
| PRC10 | DT12BYZ | Low Dose Rate (LDR) Brachytherapy of Bladder using Other Isotope | Treatment | Radiation |
| PRC10 | DT1397Z | High Dose Rate (HDR) Brachytherapy of Urethra using Cesium 137 (Cs-137) | Treatment | Radiation |
| PRC10 | DT1398Z | High Dose Rate (HDR) Brachytherapy of Urethra using Iridium 192 (Ir-192) | Treatment | Radiation |
| PRC10 | DT1399Z | High Dose Rate (HDR) Brachytherapy of Urethra using Iodine 125 (I-125) | Treatment | Radiation |
| PRC10 | DT139BZ | High Dose Rate (HDR) Brachytherapy of Urethra using Palladium 103 (Pd-103) | Treatment | Radiation |
| PRC10 | DT139CZ | High Dose Rate (HDR) Brachytherapy of Urethra using Californium 252 (Cf-252) | Treatment | Radiation |
| PRC10 | DT139YZ | High Dose Rate (HDR) Brachytherapy of Urethra using Other Isotope | Treatment | Radiation |
| PRC10 | DT13B7Z | Low Dose Rate (LDR) Brachytherapy of Urethra using Cesium 137 (Cs-137) | Treatment | Radiation |
| PRC10 | DT13B8Z | Low Dose Rate (LDR) Brachytherapy of Urethra using Iridium 192 (Ir-192) | Treatment | Radiation |
| PRC10 | DT13B9Z | Low Dose Rate (LDR) Brachytherapy of Urethra using Iodine 125 (I-125) | Treatment | Radiation |
| PRC10 | DT13BBZ | Low Dose Rate (LDR) Brachytherapy of Urethra using Palladium 103 (Pd-103) | Treatment | Radiation |
| PRC10 | DT13BCZ | Low Dose Rate (LDR) Brachytherapy of Urethra using Californium 252 (Cf-252) | Treatment | Radiation |
| PRC10 | DT13BYZ | Low Dose Rate (LDR) Brachytherapy of Urethra using Other Isotope | Treatment | Radiation |
| PRC10 | DTY07ZZ | Contact Radiation of Kidney | Treatment | Radiation |
| PRC10 | DTY0CZZ | Intraoperative Radiation Therapy (IORT) of Kidney | Treatment | Radiation |
| PRC10 | DTY0FZZ | Plaque Radiation of Kidney | Treatment | Radiation |
| PRC10 | DTY17ZZ | Contact Radiation of Ureter | Treatment | Radiation |
| PRC10 | DTY1CZZ | Intraoperative Radiation Therapy (IORT) of Ureter | Treatment | Radiation |
| PRC10 | DTY1FZZ | Plaque Radiation of Ureter | Treatment | Radiation |
| PRC10 | DTY27ZZ | Contact Radiation of Bladder | Treatment | Radiation |
| PRC10 | DTY2CZZ | Intraoperative Radiation Therapy (IORT) of Bladder | Treatment | Radiation |
| PRC10 | DTY2FZZ | Plaque Radiation of Bladder | Treatment | Radiation |
| PRC10 | DTY37ZZ | Contact Radiation of Urethra | Treatment | Radiation |
| PRC10 | DTY3CZZ | Intraoperative Radiation Therapy (IORT) of Urethra | Treatment | Radiation |
| PRC10 | DTY3FZZ | Plaque Radiation of Urethra | Treatment | Radiation |
| PRC10 | DU000ZZ | Beam Radiation of Ovary using Photons <1 MeV | Treatment | Radiation |
| PRC10 | DU003ZZ | Beam Radiation of Ovary using Electrons | Treatment | Radiation |
| PRC10 | DU004ZZ | Beam Radiation of Ovary using Heavy Particles (Protons,Ions) | Treatment | Radiation |
| PRC10 | DU005ZZ | Beam Radiation of Ovary using Neutrons | Treatment | Radiation |
| PRC10 | DU010ZZ | Beam Radiation of Cervix using Photons <1 MeV | Treatment | Radiation |
| PRC10 | DU013ZZ | Beam Radiation of Cervix using Electrons | Treatment | Radiation |
| PRC10 | DU014ZZ | Beam Radiation of Cervix using Heavy Particles (Protons,Ions) | Treatment | Radiation |
| PRC10 | DU015ZZ | Beam Radiation of Cervix using Neutrons | Treatment | Radiation |
| PRC10 | DU020ZZ | Beam Radiation of Uterus using Photons <1 MeV | Treatment | Radiation |
| PRC10 | DU023ZZ | Beam Radiation of Uterus using Electrons | Treatment | Radiation |
| PRC10 | DU024ZZ | Beam Radiation of Uterus using Heavy Particles (Protons,Ions) | Treatment | Radiation |
| PRC10 | DU025ZZ | Beam Radiation of Uterus using Neutrons | Treatment | Radiation |
| PRC10 | DU1097Z | High Dose Rate (HDR) Brachytherapy of Ovary using Cesium 137 (Cs-137) | Treatment | Radiation |
| PRC10 | DU1098Z | High Dose Rate (HDR) Brachytherapy of Ovary using Iridium 192 (Ir-192) | Treatment | Radiation |
| PRC10 | DU1099Z | High Dose Rate (HDR) Brachytherapy of Ovary using Iodine 125 (I-125) | Treatment | Radiation |
| PRC10 | DU109BZ | High Dose Rate (HDR) Brachytherapy of Ovary using Palladium 103 (Pd-103) | Treatment | Radiation |
| PRC10 | DU109CZ | High Dose Rate (HDR) Brachytherapy of Ovary using Californium 252 (Cf-252) | Treatment | Radiation |
| PRC10 | DU109YZ | High Dose Rate (HDR) Brachytherapy of Ovary using Other Isotope | Treatment | Radiation |
| PRC10 | DU10B7Z | Low Dose Rate (LDR) Brachytherapy of Ovary using Cesium 137 (Cs-137) | Treatment | Radiation |
| PRC10 | DU10B8Z | Low Dose Rate (LDR) Brachytherapy of Ovary using Iridium 192 (Ir-192) | Treatment | Radiation |
| PRC10 | DU10B9Z | Low Dose Rate (LDR) Brachytherapy of Ovary using Iodine 125 (I-125) | Treatment | Radiation |
| PRC10 | DU10BBZ | Low Dose Rate (LDR) Brachytherapy of Ovary using Palladium 103 (Pd-103) | Treatment | Radiation |
| PRC10 | DU10BCZ | Low Dose Rate (LDR) Brachytherapy of Ovary using Californium 252 (Cf-252) | Treatment | Radiation |
| PRC10 | DU10BYZ | Low Dose Rate (LDR) Brachytherapy of Ovary using Other Isotope | Treatment | Radiation |
| PRC10 | DU1197Z | High Dose Rate (HDR) Brachytherapy of Cervix using Cesium 137 (Cs-137) | Treatment | Radiation |
| PRC10 | DU1198Z | High Dose Rate (HDR) Brachytherapy of Cervix using Iridium 192 (Ir-192) | Treatment | Radiation |
| PRC10 | DU1199Z | High Dose Rate (HDR) Brachytherapy of Cervix using Iodine 125 (I-125) | Treatment | Radiation |
| PRC10 | DU119BZ | High Dose Rate (HDR) Brachytherapy of Cervix using Palladium 103 (Pd-103) | Treatment | Radiation |
| PRC10 | DU119CZ | High Dose Rate (HDR) Brachytherapy of Cervix using Californium 252 (Cf-252) | Treatment | Radiation |
| PRC10 | DU119YZ | High Dose Rate (HDR) Brachytherapy of Cervix using Other Isotope | Treatment | Radiation |
| PRC10 | DU11B7Z | Low Dose Rate (LDR) Brachytherapy of Cervix using Cesium 137 (Cs-137) | Treatment | Radiation |
| PRC10 | DU11B8Z | Low Dose Rate (LDR) Brachytherapy of Cervix using Iridium 192 (Ir-192) | Treatment | Radiation |
| PRC10 | DU11B9Z | Low Dose Rate (LDR) Brachytherapy of Cervix using Iodine 125 (I-125) | Treatment | Radiation |
| PRC10 | DU11BBZ | Low Dose Rate (LDR) Brachytherapy of Cervix using Palladium 103 (Pd-103) | Treatment | Radiation |
| PRC10 | DU11BCZ | Low Dose Rate (LDR) Brachytherapy of Cervix using Californium 252 (Cf-252) | Treatment | Radiation |
| PRC10 | DU11BYZ | Low Dose Rate (LDR) Brachytherapy of Cervix using Other Isotope | Treatment | Radiation |
| PRC10 | DU1297Z | High Dose Rate (HDR) Brachytherapy of Uterus using Cesium 137 (Cs-137) | Treatment | Radiation |
| PRC10 | DU1298Z | High Dose Rate (HDR) Brachytherapy of Uterus using Iridium 192 (Ir-192) | Treatment | Radiation |
| PRC10 | DU1299Z | High Dose Rate (HDR) Brachytherapy of Uterus using Iodine 125 (I-125) | Treatment | Radiation |
| PRC10 | DU129BZ | High Dose Rate (HDR) Brachytherapy of Uterus using Palladium 103 (Pd-103) | Treatment | Radiation |
| PRC10 | DU129CZ | High Dose Rate (HDR) Brachytherapy of Uterus using Californium 252 (Cf-252) | Treatment | Radiation |
| PRC10 | DU129YZ | High Dose Rate (HDR) Brachytherapy of Uterus using Other Isotope | Treatment | Radiation |
| PRC10 | DU12B7Z | Low Dose Rate (LDR) Brachytherapy of Uterus using Cesium 137 (Cs-137) | Treatment | Radiation |
| PRC10 | DU12B8Z | Low Dose Rate (LDR) Brachytherapy of Uterus using Iridium 192 (Ir-192) | Treatment | Radiation |
| PRC10 | DU12B9Z | Low Dose Rate (LDR) Brachytherapy of Uterus using Iodine 125 (I-125) | Treatment | Radiation |
| PRC10 | DU12BBZ | Low Dose Rate (LDR) Brachytherapy of Uterus using Palladium 103 (Pd-103) | Treatment | Radiation |
| PRC10 | DU12BCZ | Low Dose Rate (LDR) Brachytherapy of Uterus using Californium 252 (Cf-252) | Treatment | Radiation |
| PRC10 | DU12BYZ | Low Dose Rate (LDR) Brachytherapy of Uterus using Other Isotope | Treatment | Radiation |
| PRC10 | DUY07ZZ | Contact Radiation of Ovary | Treatment | Radiation |
| PRC10 | DUY0CZZ | Intraoperative Radiation Therapy (IORT) of Ovary | Treatment | Radiation |
| PRC10 | DUY0FZZ | Plaque Radiation of Ovary | Treatment | Radiation |
| PRC10 | DUY17ZZ | Contact Radiation of Cervix | Treatment | Radiation |
| PRC10 | DUY1CZZ | Intraoperative Radiation Therapy (IORT) of Cervix | Treatment | Radiation |
| PRC10 | DUY1FZZ | Plaque Radiation of Cervix | Treatment | Radiation |
| PRC10 | DUY27ZZ | Contact Radiation of Uterus | Treatment | Radiation |
| PRC10 | DUY2CZZ | Intraoperative Radiation Therapy (IORT) of Uterus | Treatment | Radiation |
| PRC10 | DUY2FZZ | Plaque Radiation of Uterus | Treatment | Radiation |
| PRC10 | DV000ZZ | Beam Radiation of Prostate using Photons <1 MeV | Treatment | Radiation |
| PRC10 | DV003ZZ | Beam Radiation of Prostate using Electrons | Treatment | Radiation |
| PRC10 | DV004ZZ | Beam Radiation of Prostate using Heavy Particles (Protons,Ions) | Treatment | Radiation |
| PRC10 | DV005ZZ | Beam Radiation of Prostate using Neutrons | Treatment | Radiation |
| PRC10 | DV010ZZ | Beam Radiation of Testis using Photons <1 MeV | Treatment | Radiation |
| PRC10 | DV013ZZ | Beam Radiation of Testis using Electrons | Treatment | Radiation |
| PRC10 | DV014ZZ | Beam Radiation of Testis using Heavy Particles (Protons,Ions) | Treatment | Radiation |
| PRC10 | DV015ZZ | Beam Radiation of Testis using Neutrons | Treatment | Radiation |
| PRC10 | DV1097Z | High Dose Rate (HDR) Brachytherapy of Prostate using Cesium 137 (Cs-137) | Treatment | Radiation |
| PRC10 | DV1098Z | High Dose Rate (HDR) Brachytherapy of Prostate using Iridium 192 (Ir-192) | Treatment | Radiation |
| PRC10 | DV1099Z | High Dose Rate (HDR) Brachytherapy of Prostate using Iodine 125 (I-125) | Treatment | Radiation |
| PRC10 | DV109BZ | High Dose Rate (HDR) Brachytherapy of Prostate using Palladium 103 (Pd-103) | Treatment | Radiation |
| PRC10 | DV109CZ | High Dose Rate (HDR) Brachytherapy of Prostate using Californium 252 (Cf-252) | Treatment | Radiation |
| PRC10 | DV109YZ | High Dose Rate (HDR) Brachytherapy of Prostate using Other Isotope | Treatment | Radiation |
| PRC10 | DV10B7Z | Low Dose Rate (LDR) Brachytherapy of Prostate using Cesium 137 (Cs-137) | Treatment | Radiation |
| PRC10 | DV10B8Z | Low Dose Rate (LDR) Brachytherapy of Prostate using Iridium 192 (Ir-192) | Treatment | Radiation |
| PRC10 | DV10B9Z | Low Dose Rate (LDR) Brachytherapy of Prostate using Iodine 125 (I-125) | Treatment | Radiation |
| PRC10 | DV10BBZ | Low Dose Rate (LDR) Brachytherapy of Prostate using Palladium 103 (Pd-103) | Treatment | Radiation |
| PRC10 | DV10BCZ | Low Dose Rate (LDR) Brachytherapy of Prostate using Californium 252 (Cf-252) | Treatment | Radiation |
| PRC10 | DV10BYZ | Low Dose Rate (LDR) Brachytherapy of Prostate using Other Isotope | Treatment | Radiation |
| PRC10 | DV1197Z | High Dose Rate (HDR) Brachytherapy of Testis using Cesium 137 (Cs-137) | Treatment | Radiation |
| PRC10 | DV1198Z | High Dose Rate (HDR) Brachytherapy of Testis using Iridium 192 (Ir-192) | Treatment | Radiation |
| PRC10 | DV1199Z | High Dose Rate (HDR) Brachytherapy of Testis using Iodine 125 (I-125) | Treatment | Radiation |
| PRC10 | DV119BZ | High Dose Rate (HDR) Brachytherapy of Testis using Palladium 103 (Pd-103) | Treatment | Radiation |
| PRC10 | DV119CZ | High Dose Rate (HDR) Brachytherapy of Testis using Californium 252 (Cf-252) | Treatment | Radiation |
| PRC10 | DV119YZ | High Dose Rate (HDR) Brachytherapy of Testis using Other Isotope | Treatment | Radiation |
| PRC10 | DV11B7Z | Low Dose Rate (LDR) Brachytherapy of Testis using Cesium 137 (Cs-137) | Treatment | Radiation |
| PRC10 | DV11B8Z | Low Dose Rate (LDR) Brachytherapy of Testis using Iridium 192 (Ir-192) | Treatment | Radiation |
| PRC10 | DV11B9Z | Low Dose Rate (LDR) Brachytherapy of Testis using Iodine 125 (I-125) | Treatment | Radiation |
| PRC10 | DV11BBZ | Low Dose Rate (LDR) Brachytherapy of Testis using Palladium 103 (Pd-103) | Treatment | Radiation |
| PRC10 | DV11BCZ | Low Dose Rate (LDR) Brachytherapy of Testis using Californium 252 (Cf-252) | Treatment | Radiation |
| PRC10 | DV11BYZ | Low Dose Rate (LDR) Brachytherapy of Testis using Other Isotope | Treatment | Radiation |
| PRC10 | DVY07ZZ | Contact Radiation of Prostate | Treatment | Radiation |
| PRC10 | DVY0CZZ | Intraoperative Radiation Therapy (IORT) of Prostate | Treatment | Radiation |
| PRC10 | DVY0FZZ | Plaque Radiation of Prostate | Treatment | Radiation |
| PRC10 | DVY17ZZ | Contact Radiation of Testis | Treatment | Radiation |
| PRC10 | DVY1FZZ | Plaque Radiation of Testis | Treatment | Radiation |
| PRC10 | DW010ZZ | Beam Radiation of Head and Neck using Photons <1 MeV | Treatment | Radiation |
| PRC10 | DW013ZZ | Beam Radiation of Head and Neck using Electrons | Treatment | Radiation |
| PRC10 | DW014ZZ | Beam Radiation of Head and Neck using Heavy Particles (Protons,Ions) | Treatment | Radiation |
| PRC10 | DW015ZZ | Beam Radiation of Head and Neck using Neutrons | Treatment | Radiation |
| PRC10 | DW020ZZ | Beam Radiation of Chest using Photons <1 MeV | Treatment | Radiation |
| PRC10 | DW023ZZ | Beam Radiation of Chest using Electrons | Treatment | Radiation |
| PRC10 | DW024ZZ | Beam Radiation of Chest using Heavy Particles (Protons,Ions) | Treatment | Radiation |
| PRC10 | DW025ZZ | Beam Radiation of Chest using Neutrons | Treatment | Radiation |
| PRC10 | DW030ZZ | Beam Radiation of Abdomen using Photons <1 MeV | Treatment | Radiation |
| PRC10 | DW033ZZ | Beam Radiation of Abdomen using Electrons | Treatment | Radiation |
| PRC10 | DW034ZZ | Beam Radiation of Abdomen using Heavy Particles (Protons,Ions) | Treatment | Radiation |
| PRC10 | DW035ZZ | Beam Radiation of Abdomen using Neutrons | Treatment | Radiation |
| PRC10 | DW040ZZ | Beam Radiation of Hemibody using Photons <1 MeV | Treatment | Radiation |
| PRC10 | DW043ZZ | Beam Radiation of Hemibody using Electrons | Treatment | Radiation |
| PRC10 | DW044ZZ | Beam Radiation of Hemibody using Heavy Particles (Protons,Ions) | Treatment | Radiation |
| PRC10 | DW045ZZ | Beam Radiation of Hemibody using Neutrons | Treatment | Radiation |
| PRC10 | DW050ZZ | Beam Radiation of Whole Body using Photons <1 MeV | Treatment | Radiation |
| PRC10 | DW053ZZ | Beam Radiation of Whole Body using Electrons | Treatment | Radiation |
| PRC10 | DW054ZZ | Beam Radiation of Whole Body using Heavy Particles (Protons,Ions) | Treatment | Radiation |
| PRC10 | DW055ZZ | Beam Radiation of Whole Body using Neutrons | Treatment | Radiation |
| PRC10 | DW060ZZ | Beam Radiation of Pelvic Region using Photons <1 MeV | Treatment | Radiation |
| PRC10 | DW063ZZ | Beam Radiation of Pelvic Region using Electrons | Treatment | Radiation |
| PRC10 | DW064ZZ | Beam Radiation of Pelvic Region using Heavy Particles (Protons,Ions) | Treatment | Radiation |
| PRC10 | DW065ZZ | Beam Radiation of Pelvic Region using Neutrons | Treatment | Radiation |
| PRC10 | DW1197Z | High Dose Rate (HDR) Brachytherapy of Head and Neck using Cesium 137 (Cs-137) | Treatment | Radiation |
| PRC10 | DW1198Z | High Dose Rate (HDR) Brachytherapy of Head and Neck using Iridium 192 (Ir-192) | Treatment | Radiation |
| PRC10 | DW1199Z | High Dose Rate (HDR) Brachytherapy of Head and Neck using Iodine 125 (I-125) | Treatment | Radiation |
| PRC10 | DW119BZ | High Dose Rate (HDR) Brachytherapy of Head and Neck using Palladium 103 (Pd-103) | Treatment | Radiation |
| PRC10 | DW119CZ | High Dose Rate (HDR) Brachytherapy of Head and Neck using Californium 252 (Cf-252) | Treatment | Radiation |
| PRC10 | DW119YZ | High Dose Rate (HDR) Brachytherapy of Head and Neck using Other Isotope | Treatment | Radiation |
| PRC10 | DW11B7Z | Low Dose Rate (LDR) Brachytherapy of Head and Neck using Cesium 137 (Cs-137) | Treatment | Radiation |
| PRC10 | DW11B8Z | Low Dose Rate (LDR) Brachytherapy of Head and Neck using Iridium 192 (Ir-192) | Treatment | Radiation |
| PRC10 | DW11B9Z | Low Dose Rate (LDR) Brachytherapy of Head and Neck using Iodine 125 (I-125) | Treatment | Radiation |
| PRC10 | DW11BBZ | Low Dose Rate (LDR) Brachytherapy of Head and Neck using Palladium 103 (Pd-103) | Treatment | Radiation |
| PRC10 | DW11BCZ | Low Dose Rate (LDR) Brachytherapy of Head and Neck using Californium 252 (Cf-252) | Treatment | Radiation |
| PRC10 | DW11BYZ | Low Dose Rate (LDR) Brachytherapy of Head and Neck using Other Isotope | Treatment | Radiation |
| PRC10 | DW1297Z | High Dose Rate (HDR) Brachytherapy of Chest using Cesium 137 (Cs-137) | Treatment | Radiation |
| PRC10 | DW1298Z | High Dose Rate (HDR) Brachytherapy of Chest using Iridium 192 (Ir-192) | Treatment | Radiation |
| PRC10 | DW1299Z | High Dose Rate (HDR) Brachytherapy of Chest using Iodine 125 (I-125) | Treatment | Radiation |
| PRC10 | DW129BZ | High Dose Rate (HDR) Brachytherapy of Chest using Palladium 103 (Pd-103) | Treatment | Radiation |
| PRC10 | DW129CZ | High Dose Rate (HDR) Brachytherapy of Chest using Californium 252 (Cf-252) | Treatment | Radiation |
| PRC10 | DW129YZ | High Dose Rate (HDR) Brachytherapy of Chest using Other Isotope | Treatment | Radiation |
| PRC10 | DW12B7Z | Low Dose Rate (LDR) Brachytherapy of Chest using Cesium 137 (Cs-137) | Treatment | Radiation |
| PRC10 | DW12B8Z | Low Dose Rate (LDR) Brachytherapy of Chest using Iridium 192 (Ir-192) | Treatment | Radiation |
| PRC10 | DW12B9Z | Low Dose Rate (LDR) Brachytherapy of Chest using Iodine 125 (I-125) | Treatment | Radiation |
| PRC10 | DW12BBZ | Low Dose Rate (LDR) Brachytherapy of Chest using Palladium 103 (Pd-103) | Treatment | Radiation |
| PRC10 | DW12BCZ | Low Dose Rate (LDR) Brachytherapy of Chest using Californium 252 (Cf-252) | Treatment | Radiation |
| PRC10 | DW12BYZ | Low Dose Rate (LDR) Brachytherapy of Chest using Other Isotope | Treatment | Radiation |
| PRC10 | DW1397Z | High Dose Rate (HDR) Brachytherapy of Abdomen using Cesium 137 (Cs-137) | Treatment | Radiation |
| PRC10 | DW1398Z | High Dose Rate (HDR) Brachytherapy of Abdomen using Iridium 192 (Ir-192) | Treatment | Radiation |
| PRC10 | DW1399Z | High Dose Rate (HDR) Brachytherapy of Abdomen using Iodine 125 (I-125) | Treatment | Radiation |
| PRC10 | DW139BZ | High Dose Rate (HDR) Brachytherapy of Abdomen using Palladium 103 (Pd-103) | Treatment | Radiation |
| PRC10 | DW139CZ | High Dose Rate (HDR) Brachytherapy of Abdomen using Californium 252 (Cf-252) | Treatment | Radiation |
| PRC10 | DW139YZ | High Dose Rate (HDR) Brachytherapy of Abdomen using Other Isotope | Treatment | Radiation |
| PRC10 | DW13B7Z | Low Dose Rate (LDR) Brachytherapy of Abdomen using Cesium 137 (Cs-137) | Treatment | Radiation |
| PRC10 | DW13B8Z | Low Dose Rate (LDR) Brachytherapy of Abdomen using Iridium 192 (Ir-192) | Treatment | Radiation |
| PRC10 | DW13B9Z | Low Dose Rate (LDR) Brachytherapy of Abdomen using Iodine 125 (I-125) | Treatment | Radiation |
| PRC10 | DW13BBZ | Low Dose Rate (LDR) Brachytherapy of Abdomen using Palladium 103 (Pd-103) | Treatment | Radiation |
| PRC10 | DW13BCZ | Low Dose Rate (LDR) Brachytherapy of Abdomen using Californium 252 (Cf-252) | Treatment | Radiation |
| PRC10 | DW13BYZ | Low Dose Rate (LDR) Brachytherapy of Abdomen using Other Isotope | Treatment | Radiation |
| PRC10 | DW1697Z | High Dose Rate (HDR) Brachytherapy of Pelvic Region using Cesium 137 (Cs-137) | Treatment | Radiation |
| PRC10 | DW1698Z | High Dose Rate (HDR) Brachytherapy of Pelvic Region using Iridium 192 (Ir-192) | Treatment | Radiation |
| PRC10 | DW1699Z | High Dose Rate (HDR) Brachytherapy of Pelvic Region using Iodine 125 (I-125) | Treatment | Radiation |
| PRC10 | DW169BZ | High Dose Rate (HDR) Brachytherapy of Pelvic Region using Palladium 103 (Pd-103) | Treatment | Radiation |
| PRC10 | DW169CZ | High Dose Rate (HDR) Brachytherapy of Pelvic Region using Californium 252 (Cf-252) | Treatment | Radiation |
| PRC10 | DW169YZ | High Dose Rate (HDR) Brachytherapy of Pelvic Region using Other Isotope | Treatment | Radiation |
| PRC10 | DW16B7Z | Low Dose Rate (LDR) Brachytherapy of Pelvic Region using Cesium 137 (Cs-137) | Treatment | Radiation |
| PRC10 | DW16B8Z | Low Dose Rate (LDR) Brachytherapy of Pelvic Region using Iridium 192 (Ir-192) | Treatment | Radiation |
| PRC10 | DW16B9Z | Low Dose Rate (LDR) Brachytherapy of Pelvic Region using Iodine 125 (I-125) | Treatment | Radiation |
| PRC10 | DW16BBZ | Low Dose Rate (LDR) Brachytherapy of Pelvic Region using Palladium 103 (Pd-103) | Treatment | Radiation |
| PRC10 | DW16BCZ | Low Dose Rate (LDR) Brachytherapy of Pelvic Region using Californium 252 (Cf-252) | Treatment | Radiation |
| PRC10 | DW16BYZ | Low Dose Rate (LDR) Brachytherapy of Pelvic Region using Other Isotope | Treatment | Radiation |
| PRC10 | DWY17ZZ | Contact Radiation of Head and Neck | Treatment | Radiation |
| PRC10 | DWY1FZZ | Plaque Radiation of Head and Neck | Treatment | Radiation |
| PRC10 | DWY27ZZ | Contact Radiation of Chest | Treatment | Radiation |
| PRC10 | DWY2FZZ | Plaque Radiation of Chest | Treatment | Radiation |
| PRC10 | DWY37ZZ | Contact Radiation of Abdomen | Treatment | Radiation |
| PRC10 | DWY3FZZ | Plaque Radiation of Abdomen | Treatment | Radiation |
| PRC10 | DWY47ZZ | Contact Radiation of Hemibody | Treatment | Radiation |
| PRC10 | DWY4FZZ | Plaque Radiation of Hemibody | Treatment | Radiation |
| PRC10 | DWY57ZZ | Contact Radiation of Whole Body | Treatment | Radiation |
| PRC10 | DWY5FZZ | Plaque Radiation of Whole Body | Treatment | Radiation |
| PRC10 | DWY5GFZ | Isotope Administration to Whole Body using Phosphorus 32 (P-32) | Treatment | Radiation |
| PRC10 | DWY5GGZ | Isotope Administration to Whole Body using Strontium 89 (Sr-89) | Treatment | Radiation |
| PRC10 | DWY5GHZ | Isotope Administration to Whole Body using Strontium 90 (Sr-90) | Treatment | Radiation |
| PRC10 | DWY5GYZ | Isotope Administration to Whole Body using Other Isotope | Treatment | Radiation |
| PRC10 | DWY67ZZ | Contact Radiation of Pelvic Region | Treatment | Radiation |
| PRC10 | DWY6FZZ | Plaque Radiation of Pelvic Region | Treatment | Radiation |
| DGN10 | Z510 | Encounter for antineoplastic radiation therapy | Treatment | Radiation |
| PRC10 | 00H004Z | Insertion of Radioactive Element, Cesium-131 Collagen Implant into Brain, Open Approach | Treatment | Radiation |
| PRC10 | 3E0Q004 | Introduction of Liquid Brachytherapy Radioisotope into Cranial Cavity and Brain, Open Approach | Treatment | Radiation |
| PRC10 | D0006ZZ | Beam Radiation of Brain using Neutron Capture | Treatment | Radiation |
| PRC10 | D0016ZZ | Beam Radiation of Brain Stem using Neutron Capture | Treatment | Radiation |
| PRC10 | D0066ZZ | Beam Radiation of Spinal Cord using Neutron Capture | Treatment | Radiation |
| PRC10 | D7006ZZ | Beam Radiation of Bone Marrow using Neutron Capture | Treatment | Radiation |
| PRC10 | D7016ZZ | Beam Radiation of Thymus using Neutron Capture | Treatment | Radiation |
| PRC10 | D7026ZZ | Beam Radiation of Spleen using Neutron Capture | Treatment | Radiation |
| PRC10 | D7036ZZ | Beam Radiation of Neck Lymphatics using Neutron Capture | Treatment | Radiation |
| PRC10 | D7046ZZ | Beam Radiation of Axillary Lymphatics using Neutron Capture | Treatment | Radiation |
| PRC10 | D7056ZZ | Beam Radiation of Thorax Lymphatics using Neutron Capture | Treatment | Radiation |
| PRC10 | D7066ZZ | Beam Radiation of Abdomen Lymphatics using Neutron Capture | Treatment | Radiation |
| PRC10 | D7076ZZ | Beam Radiation of Pelvis Lymphatics using Neutron Capture | Treatment | Radiation |
| PRC10 | D7086ZZ | Beam Radiation of Inguinal Lymphatics using Neutron Capture | Treatment | Radiation |
| PRC10 | D9016ZZ | Beam Radiation of Nose using Neutron Capture | Treatment | Radiation |
| PRC10 | D9036ZZ | Beam Radiation of Hypopharynx using Neutron Capture | Treatment | Radiation |
| PRC10 | D9046ZZ | Beam Radiation of Mouth using Neutron Capture | Treatment | Radiation |
| PRC10 | D9056ZZ | Beam Radiation of Tongue using Neutron Capture | Treatment | Radiation |
| PRC10 | D9066ZZ | Beam Radiation of Salivary Glands using Neutron Capture | Treatment | Radiation |
| PRC10 | D9076ZZ | Beam Radiation of Sinuses using Neutron Capture | Treatment | Radiation |
| PRC10 | D9086ZZ | Beam Radiation of Hard Palate using Neutron Capture | Treatment | Radiation |
| PRC10 | D9096ZZ | Beam Radiation of Soft Palate using Neutron Capture | Treatment | Radiation |
| PRC10 | D90B6ZZ | Beam Radiation of Larynx using Neutron Capture | Treatment | Radiation |
| PRC10 | D90D6ZZ | Beam Radiation of Nasopharynx using Neutron Capture | Treatment | Radiation |
| PRC10 | D90F6ZZ | Beam Radiation of Oropharynx using Neutron Capture | Treatment | Radiation |
| PRC10 | DB006ZZ | Beam Radiation of Trachea using Neutron Capture | Treatment | Radiation |
| PRC10 | DB016ZZ | Beam Radiation of Bronchus using Neutron Capture | Treatment | Radiation |
| PRC10 | DB026ZZ | Beam Radiation of Lung using Neutron Capture | Treatment | Radiation |
| PRC10 | DB056ZZ | Beam Radiation of Pleura using Neutron Capture | Treatment | Radiation |
| PRC10 | DB066ZZ | Beam Radiation of Mediastinum using Neutron Capture | Treatment | Radiation |
| PRC10 | DB076ZZ | Beam Radiation of Chest Wall using Neutron Capture | Treatment | Radiation |
| PRC10 | DB086ZZ | Beam Radiation of Diaphragm using Neutron Capture | Treatment | Radiation |
| PRC10 | DD006ZZ | Beam Radiation of Esophagus using Neutron Capture | Treatment | Radiation |
| PRC10 | DD016ZZ | Beam Radiation of Stomach using Neutron Capture | Treatment | Radiation |
| PRC10 | DD026ZZ | Beam Radiation of Duodenum using Neutron Capture | Treatment | Radiation |
| PRC10 | DD036ZZ | Beam Radiation of Jejunum using Neutron Capture | Treatment | Radiation |
| PRC10 | DD046ZZ | Beam Radiation of Ileum using Neutron Capture | Treatment | Radiation |
| PRC10 | DD056ZZ | Beam Radiation of Colon using Neutron Capture | Treatment | Radiation |
| PRC10 | DD076ZZ | Beam Radiation of Rectum using Neutron Capture | Treatment | Radiation |
| PRC10 | DF006ZZ | Beam Radiation of Liver using Neutron Capture | Treatment | Radiation |
| PRC10 | DF016ZZ | Beam Radiation of Gallbladder using Neutron Capture | Treatment | Radiation |
| PRC10 | DF026ZZ | Beam Radiation of Bile Ducts using Neutron Capture | Treatment | Radiation |
| PRC10 | DF036ZZ | Beam Radiation of Pancreas using Neutron Capture | Treatment | Radiation |
| PRC10 | DG006ZZ | Beam Radiation of Pituitary Gland using Neutron Capture | Treatment | Radiation |
| PRC10 | DG016ZZ | Beam Radiation of Pineal Body using Neutron Capture | Treatment | Radiation |
| PRC10 | DG026ZZ | Beam Radiation of Adrenal Glands using Neutron Capture | Treatment | Radiation |
| PRC10 | DG046ZZ | Beam Radiation of Parathyroid Glands using Neutron Capture | Treatment | Radiation |
| PRC10 | DG056ZZ | Beam Radiation of Thyroid using Neutron Capture | Treatment | Radiation |
| PRC10 | DM006ZZ | Beam Radiation of Left Breast using Neutron Capture | Treatment | Radiation |
| PRC10 | DM016ZZ | Beam Radiation of Right Breast using Neutron Capture | Treatment | Radiation |
| PRC10 | DP006ZZ | Beam Radiation of Skull using Neutron Capture | Treatment | Radiation |
| PRC10 | DP026ZZ | Beam Radiation of Maxilla using Neutron Capture | Treatment | Radiation |
| PRC10 | DP036ZZ | Beam Radiation of Mandible using Neutron Capture | Treatment | Radiation |
| PRC10 | DP046ZZ | Beam Radiation of Sternum using Neutron Capture | Treatment | Radiation |
| PRC10 | DP056ZZ | Beam Radiation of Rib(s) using Neutron Capture | Treatment | Radiation |
| PRC10 | DP066ZZ | Beam Radiation of Humerus using Neutron Capture | Treatment | Radiation |
| PRC10 | DP076ZZ | Beam Radiation of Radius/Ulna using Neutron Capture | Treatment | Radiation |
| PRC10 | DP086ZZ | Beam Radiation of Pelvic Bones using Neutron Capture | Treatment | Radiation |
| PRC10 | DP096ZZ | Beam Radiation of Femur using Neutron Capture | Treatment | Radiation |
| PRC10 | DP0B6ZZ | Beam Radiation of Tibia/Fibula using Neutron Capture | Treatment | Radiation |
| PRC10 | DP0C6ZZ | Beam Radiation of Other Bone using Neutron Capture | Treatment | Radiation |
| PRC10 | DT006ZZ | Beam Radiation of Kidney using Neutron Capture | Treatment | Radiation |
| PRC10 | DT016ZZ | Beam Radiation of Ureter using Neutron Capture | Treatment | Radiation |
| PRC10 | DT026ZZ | Beam Radiation of Bladder using Neutron Capture | Treatment | Radiation |
| PRC10 | DT036ZZ | Beam Radiation of Urethra using Neutron Capture | Treatment | Radiation |
| PRC10 | DU006ZZ | Beam Radiation of Ovary using Neutron Capture | Treatment | Radiation |
| PRC10 | DU016ZZ | Beam Radiation of Cervix using Neutron Capture | Treatment | Radiation |
| PRC10 | DU026ZZ | Beam Radiation of Uterus using Neutron Capture | Treatment | Radiation |
| PRC10 | DV006ZZ | Beam Radiation of Prostate using Neutron Capture | Treatment | Radiation |
| PRC10 | DV016ZZ | Beam Radiation of Testis using Neutron Capture | Treatment | Radiation |
| PRC10 | DW016ZZ | Beam Radiation of Head and Neck using Neutron Capture | Treatment | Radiation |
| PRC10 | DW026ZZ | Beam Radiation of Chest using Neutron Capture | Treatment | Radiation |
| PRC10 | DW036ZZ | Beam Radiation of Abdomen using Neutron Capture | Treatment | Radiation |
| PRC10 | DW046ZZ | Beam Radiation of Hemibody using Neutron Capture | Treatment | Radiation |
| PRC10 | DW056ZZ | Beam Radiation of Whole Body using Neutron Capture | Treatment | Radiation |
| PRC10 | DW066ZZ | Beam Radiation of Pelvic Region using Neutron Capture | Treatment | Radiation |
| PRC10 | D0001ZZ | Beam Radiation of Brain using Photons 1 - 10 MeV | Treatment | Radiation |
| PRC10 | D0002ZZ | Beam Radiation of Brain using Photons >10 MeV | Treatment | Radiation |
| PRC10 | D0011ZZ | Beam Radiation of Brain Stem using Photons 1 - 10 MeV | Treatment | Radiation |
| PRC10 | D0012ZZ | Beam Radiation of Brain Stem using Photons >10 MeV | Treatment | Radiation |
| PRC10 | D0061ZZ | Beam Radiation of Spinal Cord using Photons 1 - 10 MeV | Treatment | Radiation |
| PRC10 | D0062ZZ | Beam Radiation of Spinal Cord using Photons >10 MeV | Treatment | Radiation |
| PRC10 | D7001ZZ | Beam Radiation of Bone Marrow using Photons 1 - 10 MeV | Treatment | Radiation |
| PRC10 | D7002ZZ | Beam Radiation of Bone Marrow using Photons >10 MeV | Treatment | Radiation |
| PRC10 | D7011ZZ | Beam Radiation of Thymus using Photons 1 - 10 MeV | Treatment | Radiation |
| PRC10 | D7012ZZ | Beam Radiation of Thymus using Photons >10 MeV | Treatment | Radiation |
| PRC10 | D7021ZZ | Beam Radiation of Spleen using Photons 1 - 10 MeV | Treatment | Radiation |
| PRC10 | D7022ZZ | Beam Radiation of Spleen using Photons >10 MeV | Treatment | Radiation |
| PRC10 | D7031ZZ | Beam Radiation of Neck Lymphatics using Photons 1 - 10 MeV | Treatment | Radiation |
| PRC10 | D7032ZZ | Beam Radiation of Neck Lymphatics using Photons >10 MeV | Treatment | Radiation |
| PRC10 | D7041ZZ | Beam Radiation of Axillary Lymphatics using Photons 1 - 10 MeV | Treatment | Radiation |
| PRC10 | D7042ZZ | Beam Radiation of Axillary Lymphatics using Photons >10 MeV | Treatment | Radiation |
| PRC10 | D7051ZZ | Beam Radiation of Thorax Lymphatics using Photons 1 - 10 MeV | Treatment | Radiation |
| PRC10 | D7052ZZ | Beam Radiation of Thorax Lymphatics using Photons >10 MeV | Treatment | Radiation |
| PRC10 | D7061ZZ | Beam Radiation of Abdomen Lymphatics using Photons 1 - 10 MeV | Treatment | Radiation |
| PRC10 | D7062ZZ | Beam Radiation of Abdomen Lymphatics using Photons >10 MeV | Treatment | Radiation |
| PRC10 | D7071ZZ | Beam Radiation of Pelvis Lymphatics using Photons 1 - 10 MeV | Treatment | Radiation |
| PRC10 | D7072ZZ | Beam Radiation of Pelvis Lymphatics using Photons >10 MeV | Treatment | Radiation |
| PRC10 | D7081ZZ | Beam Radiation of Inguinal Lymphatics using Photons 1 - 10 MeV | Treatment | Radiation |
| PRC10 | D7082ZZ | Beam Radiation of Inguinal Lymphatics using Photons >10 MeV | Treatment | Radiation |
| PRC10 | D9011ZZ | Beam Radiation of Nose using Photons 1 - 10 MeV | Treatment | Radiation |
| PRC10 | D9012ZZ | Beam Radiation of Nose using Photons >10 MeV | Treatment | Radiation |
| PRC10 | D9031ZZ | Beam Radiation of Hypopharynx using Photons 1 - 10 MeV | Treatment | Radiation |
| PRC10 | D9032ZZ | Beam Radiation of Hypopharynx using Photons >10 MeV | Treatment | Radiation |
| PRC10 | D9041ZZ | Beam Radiation of Mouth using Photons 1 - 10 MeV | Treatment | Radiation |
| PRC10 | D9042ZZ | Beam Radiation of Mouth using Photons >10 MeV | Treatment | Radiation |
| PRC10 | D9051ZZ | Beam Radiation of Tongue using Photons 1 - 10 MeV | Treatment | Radiation |
| PRC10 | D9052ZZ | Beam Radiation of Tongue using Photons >10 MeV | Treatment | Radiation |
| PRC10 | D9061ZZ | Beam Radiation of Salivary Glands using Photons 1 - 10 MeV | Treatment | Radiation |
| PRC10 | D9062ZZ | Beam Radiation of Salivary Glands using Photons >10 MeV | Treatment | Radiation |
| PRC10 | D9071ZZ | Beam Radiation of Sinuses using Photons 1 - 10 MeV | Treatment | Radiation |
| PRC10 | D9072ZZ | Beam Radiation of Sinuses using Photons >10 MeV | Treatment | Radiation |
| PRC10 | D9081ZZ | Beam Radiation of Hard Palate using Photons 1 - 10 MeV | Treatment | Radiation |
| PRC10 | D9082ZZ | Beam Radiation of Hard Palate using Photons >10 MeV | Treatment | Radiation |
| PRC10 | D9091ZZ | Beam Radiation of Soft Palate using Photons 1 - 10 MeV | Treatment | Radiation |
| PRC10 | D9092ZZ | Beam Radiation of Soft Palate using Photons >10 MeV | Treatment | Radiation |
| PRC10 | D90B1ZZ | Beam Radiation of Larynx using Photons 1 - 10 MeV | Treatment | Radiation |
| PRC10 | D90B2ZZ | Beam Radiation of Larynx using Photons >10 MeV | Treatment | Radiation |
| PRC10 | D90D1ZZ | Beam Radiation of Nasopharynx using Photons 1 - 10 MeV | Treatment | Radiation |
| PRC10 | D90D2ZZ | Beam Radiation of Nasopharynx using Photons >10 MeV | Treatment | Radiation |
| PRC10 | D90F1ZZ | Beam Radiation of Oropharynx using Photons 1 - 10 MeV | Treatment | Radiation |
| PRC10 | D90F2ZZ | Beam Radiation of Oropharynx using Photons >10 MeV | Treatment | Radiation |
| PRC10 | DB001ZZ | Beam Radiation of Trachea using Photons 1 - 10 MeV | Treatment | Radiation |
| PRC10 | DB002ZZ | Beam Radiation of Trachea using Photons >10 MeV | Treatment | Radiation |
| PRC10 | DB011ZZ | Beam Radiation of Bronchus using Photons 1 - 10 MeV | Treatment | Radiation |
| PRC10 | DB012ZZ | Beam Radiation of Bronchus using Photons >10 MeV | Treatment | Radiation |
| PRC10 | DB021ZZ | Beam Radiation of Lung using Photons 1 - 10 MeV | Treatment | Radiation |
| PRC10 | DB022ZZ | Beam Radiation of Lung using Photons >10 MeV | Treatment | Radiation |
| PRC10 | DB051ZZ | Beam Radiation of Pleura using Photons 1 - 10 MeV | Treatment | Radiation |
| PRC10 | DB052ZZ | Beam Radiation of Pleura using Photons >10 MeV | Treatment | Radiation |
| PRC10 | DB061ZZ | Beam Radiation of Mediastinum using Photons 1 - 10 MeV | Treatment | Radiation |
| PRC10 | DB062ZZ | Beam Radiation of Mediastinum using Photons >10 MeV | Treatment | Radiation |
| PRC10 | DB071ZZ | Beam Radiation of Chest Wall using Photons 1 - 10 MeV | Treatment | Radiation |
| PRC10 | DB072ZZ | Beam Radiation of Chest Wall using Photons >10 MeV | Treatment | Radiation |
| PRC10 | DB081ZZ | Beam Radiation of Diaphragm using Photons 1 - 10 MeV | Treatment | Radiation |
| PRC10 | DB082ZZ | Beam Radiation of Diaphragm using Photons >10 MeV | Treatment | Radiation |
| PRC10 | DD001ZZ | Beam Radiation of Esophagus using Photons 1 - 10 MeV | Treatment | Radiation |
| PRC10 | DD002ZZ | Beam Radiation of Esophagus using Photons >10 MeV | Treatment | Radiation |
| PRC10 | DD011ZZ | Beam Radiation of Stomach using Photons 1 - 10 MeV | Treatment | Radiation |
| PRC10 | DD012ZZ | Beam Radiation of Stomach using Photons >10 MeV | Treatment | Radiation |
| PRC10 | DD021ZZ | Beam Radiation of Duodenum using Photons 1 - 10 MeV | Treatment | Radiation |
| PRC10 | DD022ZZ | Beam Radiation of Duodenum using Photons >10 MeV | Treatment | Radiation |
| PRC10 | DD031ZZ | Beam Radiation of Jejunum using Photons 1 - 10 MeV | Treatment | Radiation |
| PRC10 | DD032ZZ | Beam Radiation of Jejunum using Photons >10 MeV | Treatment | Radiation |
| PRC10 | DD041ZZ | Beam Radiation of Ileum using Photons 1 - 10 MeV | Treatment | Radiation |
| PRC10 | DD042ZZ | Beam Radiation of Ileum using Photons >10 MeV | Treatment | Radiation |
| PRC10 | DD051ZZ | Beam Radiation of Colon using Photons 1 - 10 MeV | Treatment | Radiation |
| PRC10 | DD052ZZ | Beam Radiation of Colon using Photons >10 MeV | Treatment | Radiation |
| PRC10 | DD071ZZ | Beam Radiation of Rectum using Photons 1 - 10 MeV | Treatment | Radiation |
| PRC10 | DD072ZZ | Beam Radiation of Rectum using Photons >10 MeV | Treatment | Radiation |
| PRC10 | DF001ZZ | Beam Radiation of Liver using Photons 1 - 10 MeV | Treatment | Radiation |
| PRC10 | DF002ZZ | Beam Radiation of Liver using Photons >10 MeV | Treatment | Radiation |
| PRC10 | DF011ZZ | Beam Radiation of Gallbladder using Photons 1 - 10 MeV | Treatment | Radiation |
| PRC10 | DF012ZZ | Beam Radiation of Gallbladder using Photons >10 MeV | Treatment | Radiation |
| PRC10 | DF021ZZ | Beam Radiation of Bile Ducts using Photons 1 - 10 MeV | Treatment | Radiation |
| PRC10 | DF022ZZ | Beam Radiation of Bile Ducts using Photons >10 MeV | Treatment | Radiation |
| PRC10 | DF031ZZ | Beam Radiation of Pancreas using Photons 1 - 10 MeV | Treatment | Radiation |
| PRC10 | DF032ZZ | Beam Radiation of Pancreas using Photons >10 MeV | Treatment | Radiation |
| PRC10 | DG001ZZ | Beam Radiation of Pituitary Gland using Photons 1 - 10 MeV | Treatment | Radiation |
| PRC10 | DG002ZZ | Beam Radiation of Pituitary Gland using Photons >10 MeV | Treatment | Radiation |
| PRC10 | DG011ZZ | Beam Radiation of Pineal Body using Photons 1 - 10 MeV | Treatment | Radiation |
| PRC10 | DG012ZZ | Beam Radiation of Pineal Body using Photons >10 MeV | Treatment | Radiation |
| PRC10 | DG021ZZ | Beam Radiation of Adrenal Glands using Photons 1 - 10 MeV | Treatment | Radiation |
| PRC10 | DG022ZZ | Beam Radiation of Adrenal Glands using Photons >10 MeV | Treatment | Radiation |
| PRC10 | DG041ZZ | Beam Radiation of Parathyroid Glands using Photons 1 - 10 MeV | Treatment | Radiation |
| PRC10 | DG042ZZ | Beam Radiation of Parathyroid Glands using Photons >10 MeV | Treatment | Radiation |
| PRC10 | DG051ZZ | Beam Radiation of Thyroid using Photons 1 - 10 MeV | Treatment | Radiation |
| PRC10 | DG052ZZ | Beam Radiation of Thyroid using Photons >10 MeV | Treatment | Radiation |
| PRC10 | DM001ZZ | Beam Radiation of Left Breast using Photons 1 - 10 MeV | Treatment | Radiation |
| PRC10 | DM002ZZ | Beam Radiation of Left Breast using Photons >10 MeV | Treatment | Radiation |
| PRC10 | DM011ZZ | Beam Radiation of Right Breast using Photons 1 - 10 MeV | Treatment | Radiation |
| PRC10 | DM012ZZ | Beam Radiation of Right Breast using Photons >10 MeV | Treatment | Radiation |
| PRC10 | DP001ZZ | Beam Radiation of Skull using Photons 1 - 10 MeV | Treatment | Radiation |
| PRC10 | DP002ZZ | Beam Radiation of Skull using Photons >10 MeV | Treatment | Radiation |
| PRC10 | DP021ZZ | Beam Radiation of Maxilla using Photons 1 - 10 MeV | Treatment | Radiation |
| PRC10 | DP022ZZ | Beam Radiation of Maxilla using Photons >10 MeV | Treatment | Radiation |
| PRC10 | DP031ZZ | Beam Radiation of Mandible using Photons 1 - 10 MeV | Treatment | Radiation |
| PRC10 | DP032ZZ | Beam Radiation of Mandible using Photons >10 MeV | Treatment | Radiation |
| PRC10 | DP041ZZ | Beam Radiation of Sternum using Photons 1 - 10 MeV | Treatment | Radiation |
| PRC10 | DP042ZZ | Beam Radiation of Sternum using Photons >10 MeV | Treatment | Radiation |
| PRC10 | DP051ZZ | Beam Radiation of Rib(s) using Photons 1 - 10 MeV | Treatment | Radiation |
| PRC10 | DP052ZZ | Beam Radiation of Rib(s) using Photons >10 MeV | Treatment | Radiation |
| PRC10 | DP061ZZ | Beam Radiation of Humerus using Photons 1 - 10 MeV | Treatment | Radiation |
| PRC10 | DP062ZZ | Beam Radiation of Humerus using Photons >10 MeV | Treatment | Radiation |
| PRC10 | DP071ZZ | Beam Radiation of Radius/Ulna using Photons 1 - 10 MeV | Treatment | Radiation |
| PRC10 | DP072ZZ | Beam Radiation of Radius/Ulna using Photons >10 MeV | Treatment | Radiation |
| PRC10 | DP081ZZ | Beam Radiation of Pelvic Bones using Photons 1 - 10 MeV | Treatment | Radiation |
| PRC10 | DP082ZZ | Beam Radiation of Pelvic Bones using Photons >10 MeV | Treatment | Radiation |
| PRC10 | DP091ZZ | Beam Radiation of Femur using Photons 1 - 10 MeV | Treatment | Radiation |
| PRC10 | DP092ZZ | Beam Radiation of Femur using Photons >10 MeV | Treatment | Radiation |
| PRC10 | DP0B1ZZ | Beam Radiation of Tibia/Fibula using Photons 1 - 10 MeV | Treatment | Radiation |
| PRC10 | DP0B2ZZ | Beam Radiation of Tibia/Fibula using Photons >10 MeV | Treatment | Radiation |
| PRC10 | DP0C1ZZ | Beam Radiation of Other Bone using Photons 1 - 10 MeV | Treatment | Radiation |
| PRC10 | DP0C2ZZ | Beam Radiation of Other Bone using Photons >10 MeV | Treatment | Radiation |
| PRC10 | DT001ZZ | Beam Radiation of Kidney using Photons 1 - 10 MeV | Treatment | Radiation |
| PRC10 | DT002ZZ | Beam Radiation of Kidney using Photons >10 MeV | Treatment | Radiation |
| PRC10 | DT011ZZ | Beam Radiation of Ureter using Photons 1 - 10 MeV | Treatment | Radiation |
| PRC10 | DT012ZZ | Beam Radiation of Ureter using Photons >10 MeV | Treatment | Radiation |
| PRC10 | DT021ZZ | Beam Radiation of Bladder using Photons 1 - 10 MeV | Treatment | Radiation |
| PRC10 | DT022ZZ | Beam Radiation of Bladder using Photons >10 MeV | Treatment | Radiation |
| PRC10 | DT031ZZ | Beam Radiation of Urethra using Photons 1 - 10 MeV | Treatment | Radiation |
| PRC10 | DT032ZZ | Beam Radiation of Urethra using Photons >10 MeV | Treatment | Radiation |
| PRC10 | DU001ZZ | Beam Radiation of Ovary using Photons 1 - 10 MeV | Treatment | Radiation |
| PRC10 | DU002ZZ | Beam Radiation of Ovary using Photons >10 MeV | Treatment | Radiation |
| PRC10 | DU011ZZ | Beam Radiation of Cervix using Photons 1 - 10 MeV | Treatment | Radiation |
| PRC10 | DU012ZZ | Beam Radiation of Cervix using Photons >10 MeV | Treatment | Radiation |
| PRC10 | DU021ZZ | Beam Radiation of Uterus using Photons 1 - 10 MeV | Treatment | Radiation |
| PRC10 | DU022ZZ | Beam Radiation of Uterus using Photons >10 MeV | Treatment | Radiation |
| PRC10 | DV001ZZ | Beam Radiation of Prostate using Photons 1 - 10 MeV | Treatment | Radiation |
| PRC10 | DV002ZZ | Beam Radiation of Prostate using Photons >10 MeV | Treatment | Radiation |
| PRC10 | DV011ZZ | Beam Radiation of Testis using Photons 1 - 10 MeV | Treatment | Radiation |
| PRC10 | DV012ZZ | Beam Radiation of Testis using Photons >10 MeV | Treatment | Radiation |
| PRC10 | DW011ZZ | Beam Radiation of Head and Neck using Photons 1 - 10 MeV | Treatment | Radiation |
| PRC10 | DW012ZZ | Beam Radiation of Head and Neck using Photons >10 MeV | Treatment | Radiation |
| PRC10 | DW021ZZ | Beam Radiation of Chest using Photons 1 - 10 MeV | Treatment | Radiation |
| PRC10 | DW022ZZ | Beam Radiation of Chest using Photons >10 MeV | Treatment | Radiation |
| PRC10 | DW031ZZ | Beam Radiation of Abdomen using Photons 1 - 10 MeV | Treatment | Radiation |
| PRC10 | DW032ZZ | Beam Radiation of Abdomen using Photons >10 MeV | Treatment | Radiation |
| PRC10 | DW041ZZ | Beam Radiation of Hemibody using Photons 1 - 10 MeV | Treatment | Radiation |
| PRC10 | DW042ZZ | Beam Radiation of Hemibody using Photons >10 MeV | Treatment | Radiation |
| PRC10 | DW051ZZ | Beam Radiation of Whole Body using Photons 1 - 10 MeV | Treatment | Radiation |
| PRC10 | DW052ZZ | Beam Radiation of Whole Body using Photons >10 MeV | Treatment | Radiation |
| PRC10 | DW061ZZ | Beam Radiation of Pelvic Region using Photons 1 - 10 MeV | Treatment | Radiation |
| PRC10 | DW062ZZ | Beam Radiation of Pelvic Region using Photons >10 MeV | Treatment | Radiation |
| PRC10 | D0076ZZ | Beam Radiation of Peripheral Nerve using Neutron Capture | Treatment | Radiation |
| REV | 0331 | Radiology therapeutic-chemotherapy injected | Treatment | Chemo/Radiation Revenue Codes |
| REV | 0332 | Radiology therapeutic-chemotherapy oral | Treatment | Chemo/Radiation Revenue Codes |
| REV | 0333 | Radiology therapeutic-radiation therapy | Treatment | Chemo/Radiation Revenue Codes |
| REV | 0335 | Radiology therapeutic-chemotherapy IV | Treatment | Chemo/Radiation Revenue Codes |
| HCPCS | J9015 | Injection, aldesleukin, per single use vial | Treatment | Chemotherapy |
| HCPCS | C9110 | Injection, alemtuzumab, per 10 mg/ ml | Treatment | Chemotherapy |
| HCPCS | J0202 | Injection, alemtuzumab, 1 mg | Treatment | Chemotherapy |
| HCPCS | Q9979 | Injection, Alemtuzumab, 1 mg | Treatment | Chemotherapy |
| HCPCS | J9010 | Injection, alemtuzumab, 10 mg | Treatment | Chemotherapy |
| HCPCS | S0087 | Injection, alemtuzumab, 30 mg | Treatment | Chemotherapy |
| HCPCS | C9012 | Injection, arsenic trioxide, per 1 mg/kg | Treatment | Chemotherapy |
| HCPCS | J9017 | Injection, arsenic trioxide, 1 mg | Treatment | Chemotherapy |
| HCPCS | C9289 | Injection, asparaginase Erwinia chrysanthemi, 1,000 IU | Treatment | Chemotherapy |
| HCPCS | J9019 | Injection, asparaginase (Erwinaze), 1,000 IU | Treatment | Chemotherapy |
| HCPCS | J9020 | Injection, asparaginase, not otherwise specified, 10,000 units | Treatment | Chemotherapy |
| HCPCS | C9218 | Injection, azacitidine, per 1 mg | Treatment | Chemotherapy |
| HCPCS | J9025 | Injection, azacitidine, 1 mg | Treatment | Chemotherapy |
| HCPCS | S0168 | Injection, azacitidine, 100 mg | Treatment | Chemotherapy |
| HCPCS | C9243 | Injection, bendamustine hcl, 1 mg | Treatment | Chemotherapy |
| HCPCS | J9033 | Injection, bendamustine HCl (Treanda), 1 mg | Treatment | Chemotherapy |
| HCPCS | J9034 | Injection, bendamustine HCl (Bendeka), 1 mg | Treatment | Chemotherapy |
| HCPCS | C9214 | Injection, bevacizumab, per 10 mg | Treatment | Chemotherapy |
| HCPCS | J9035 | Injection, bevacizumab, 10 mg | Treatment | Chemotherapy |
| HCPCS | S0116 | Bevacizumab, 100 mg | Treatment | Chemotherapy |
| HCPCS | C9417 | Bleomycin sulfate, 15 units, brand name | Treatment | Chemotherapy |
| HCPCS | J9040 | Injection, bleomycin sulfate, 15 units | Treatment | Chemotherapy |
| HCPCS | C9207 | Injection, bortezomib, per 3.5 mg | Treatment | Chemotherapy |
| HCPCS | J9041 | Injection, bortezomib, 0.1 mg | Treatment | Chemotherapy |
| HCPCS | S0115 | Bortezomib, 3.5 mg | Treatment | Chemotherapy |
| HCPCS | C9287 | Injection, brentuximab vedotin, 1 mg | Treatment | Chemotherapy |
| HCPCS | J9042 | Injection, brentuximab vedotin, 1 mg | Treatment | Chemotherapy |
| HCPCS | C1178 | Injection, busulfan, per 6 mg | Treatment | Chemotherapy |
| HCPCS | J0594 | Injection, busulfan, 1 mg | Treatment | Chemotherapy |
| HCPCS | J8510 | Busulfan, oral, 2 mg | Treatment | Chemotherapy |
| HCPCS | C9276 | Injection, cabazitaxel, 1 mg | Treatment | Chemotherapy |
| HCPCS | J9043 | Injection, cabazitaxel, 1 mg | Treatment | Chemotherapy |
| HCPCS | J8520 | Capecitabine, oral, 150 mg | Treatment | Chemotherapy |
| HCPCS | J8521 | Capecitabine, oral, 500 mg | Treatment | Chemotherapy |
| HCPCS | J9045 | Injection, carboplatin, 50 mg | Treatment | Chemotherapy |
| HCPCS | C9295 | Injection, carfilzomib, 1 mg | Treatment | Chemotherapy |
| HCPCS | J9047 | Injection, carfilzomib, 1 mg | Treatment | Chemotherapy |
| HCPCS | C9437 | Carmustine, brand name, 100 mg | Treatment | Chemotherapy |
| HCPCS | J9050 | Injection, carmustine, 100 mg | Treatment | Chemotherapy |
| HCPCS | C9215 | Injection, cetuximab, per 10 mg | Treatment | Chemotherapy |
| HCPCS | J9055 | Injection, cetuximab, 10 mg | Treatment | Chemotherapy |
| HCPCS | S0172 | Chlorambucil, oral, 2 mg | Treatment | Chemotherapy |
| HCPCS | C9418 | Cisplatin, powder or solution, per 10 mg, brand name | Treatment | Chemotherapy |
| HCPCS | J9060 | Injection, cisplatin, powder or solution, 10 mg | Treatment | Chemotherapy |
| HCPCS | J9062 | Cisplatin, 50 mg | Treatment | Chemotherapy |
| HCPCS | C9419 | Injection, cladribine, per 1 mg, brand name | Treatment | Chemotherapy |
| HCPCS | J9065 | Injection, cladribine, per 1 mg | Treatment | Chemotherapy |
| HCPCS | J9027 | Injection, clofarabine, 1 mg | Treatment | Chemotherapy |
| HCPCS | C9129 | Injection, clofarabine, per 1 mg | Treatment | Chemotherapy |
| HCPCS | J9095 | Cyclophosphamide, lyophilized, 500 mg | Treatment | Chemotherapy |
| HCPCS | C9421 | Cyclophosphamide, lyophilized, 100 mg, brand name | Treatment | Chemotherapy |
| HCPCS | J9096 | Cyclophosphamide, lyophilized, 1 g | Treatment | Chemotherapy |
| HCPCS | J9080 | Cyclophosphamide, 200 mg | Treatment | Chemotherapy |
| HCPCS | J9090 | Cyclophosphamide, 500 mg | Treatment | Chemotherapy |
| HCPCS | J9094 | Cyclophosphamide, lyophilized, 200 mg | Treatment | Chemotherapy |
| HCPCS | J9097 | Cyclophosphamide, lyophilized, 2 g | Treatment | Chemotherapy |
| HCPCS | J8530 | Cyclophosphamide, oral, 25 mg | Treatment | Chemotherapy |
| HCPCS | J9092 | Cyclophosphamide, 2 g | Treatment | Chemotherapy |
| HCPCS | J9091 | Cyclophosphamide, 1 g | Treatment | Chemotherapy |
| HCPCS | C9420 | Cyclophosphamide, 100 mg, brand name | Treatment | Chemotherapy |
| HCPCS | J9070 | Cyclophosphamide, 100 mg | Treatment | Chemotherapy |
| HCPCS | J9093 | Cyclophosphamide, lyophilized, 100 mg | Treatment | Chemotherapy |
| HCPCS | C1166 | Injection, cytarabine liposome, per 10 mg | Treatment | Chemotherapy |
| HCPCS | J9100 | Injection, cytarabine, 100 mg | Treatment | Chemotherapy |
| HCPCS | J9098 | Injection, cytarabine liposome, 10 mg | Treatment | Chemotherapy |
| HCPCS | J9110 | Injection, cytarabine, 500 mg | Treatment | Chemotherapy |
| HCPCS | C9422 | Cytarabine, 100 mg, brand name | Treatment | Chemotherapy |
| HCPCS | C9423 | Dacarbazine, 100 mg, brand name | Treatment | Chemotherapy |
| HCPCS | J9130 | Dacarbazine, 100 mg | Treatment | Chemotherapy |
| HCPCS | J9140 | Dacarbazine, 200 mg | Treatment | Chemotherapy |
| HCPCS | J9120 | Injection, dactinomycin, 0.5 mg | Treatment | Chemotherapy |
| HCPCS | C9024 | Injection, liposomal, 1 mg daunorubicin and 2.27 mg cytarabine | Treatment | Chemotherapy |
| HCPCS | C9424 | Daunorubicin, 10 mg | Treatment | Chemotherapy |
| HCPCS | J9150 | Injection, daunorubicin, 10 mg | Treatment | Chemotherapy |
| HCPCS | J9151 | Injection, daunorubicin citrate, liposomal formulation, 10 mg | Treatment | Chemotherapy |
| HCPCS | C9231 | Injection, decitabine, per 1 mg | Treatment | Chemotherapy |
| HCPCS | J0894 | Injection, decitabine, 1 mg | Treatment | Chemotherapy |
| HCPCS | C1084 | Denileukin diftitox, 300 mcg, ontak iv | Treatment | Chemotherapy |
| HCPCS | J9160 | Injection, denileukin diftitox, 300 mcg | Treatment | Chemotherapy |
| HCPCS | J9170 | Injection, docetaxel, 20 mg | Treatment | Chemotherapy |
| HCPCS | J9171 | Injection, docetaxel, 1 mg | Treatment | Chemotherapy |
| HCPCS | C9415 | Doxorubicin HCl, 10 mg, brand name | Treatment | Chemotherapy |
| HCPCS | J9000 | Injection, doxorubicin HCl, 10 mg | Treatment | Chemotherapy |
| HCPCS | J9001 | Injection, doxorubicin HCl, all lipid formulations, 10 mg | Treatment | Chemotherapy |
| HCPCS | J9002 | Injection, doxorubicin hydrochloride, liposomal, Doxil, 10 mg | Treatment | Chemotherapy |
| HCPCS | Q2048 | Injection, doxorubicin hydrochloride, liposomal, DOXIL, 10 mg | Treatment | Chemotherapy |
| HCPCS | Q2049 | Injection, doxorubicin hydrochloride, liposomal, imported Lipodox, 10 mg | Treatment | Chemotherapy |
| HCPCS | Q2050 | Injection, doxorubicin hydrochloride, liposomal, not otherwise specified, 10 mg | Treatment | Chemotherapy |
| HCPCS | C1167 | Injection, epirubicin HCl, 2 mg | Treatment | Chemotherapy |
| HCPCS | J9178 | Injection, epirubicin HCl, 2 mg | Treatment | Chemotherapy |
| HCPCS | J9180 | Epirubicin HCl, 50 mg | Treatment | Chemotherapy |
| HCPCS | C9280 | Injection, eribulin mesylate, 1 mg | Treatment | Chemotherapy |
| HCPCS | J9179 | Injection, eribulin mesylate, 0.1 mg | Treatment | Chemotherapy |
| HCPCS | C9414 | Etoposide; oral, 50 mg, brand name | Treatment | Chemotherapy |
| HCPCS | C9425 | Etoposide, 10 mg, brand name | Treatment | Chemotherapy |
| HCPCS | J8560 | Etoposide, oral, 50 mg | Treatment | Chemotherapy |
| HCPCS | J9181 | Injection, etoposide, 10 mg | Treatment | Chemotherapy |
| HCPCS | J9182 | Etoposide, 100 mg | Treatment | Chemotherapy |
| HCPCS | 80169 | Everolimus | Treatment | Chemotherapy |
| HCPCS | J7527 | Everolimus, oral, 0.25 mg | Treatment | Chemotherapy |
| HCPCS | J8561 | Everolimus, oral, 0.25 mg | Treatment | Chemotherapy |
| HCPCS | C9426 | Floxuridine, 500 mg, brand name | Treatment | Chemotherapy |
| HCPCS | J9200 | Injection, floxuridine, 500 mg | Treatment | Chemotherapy |
| HCPCS | C9262 | Fludarabine phosphate, oral, 1 mg | Treatment | Chemotherapy |
| HCPCS | J8562 | Fludarabine phosphate, oral, 10 mg | Treatment | Chemotherapy |
| HCPCS | J9185 | Injection, fludarabine phosphate, 50 mg | Treatment | Chemotherapy |
| HCPCS | Q2025 | Fludarabine phosphate, oral, 1 mg | Treatment | Chemotherapy |
| HCPCS | J9190 | Injection, fluorouracil, 500 mg | Treatment | Chemotherapy |
| HCPCS | J8565 | Gefitinib, oral, 250 mg | Treatment | Chemotherapy |
| HCPCS | J9201 | Injection, gemcitabine HCl, 200 mg | Treatment | Chemotherapy |
| HCPCS | C9004 | Injection, gemtuzumab ozogamicin, per 5 mg | Treatment | Chemotherapy |
| HCPCS | J9203 | Injection, gemtuzumab ozogamicin, 0.1 mg | Treatment | Chemotherapy |
| HCPCS | J9300 | Injection, gemtuzumab ozogamicin, 5 mg | Treatment | Chemotherapy |
| HCPCS | C9429 | Idarubicin HCl, 5 mg, brand name | Treatment | Chemotherapy |
| HCPCS | J9211 | Injection, idarubicin HCl, 5 mg | Treatment | Chemotherapy |
| HCPCS | C9427 | Ifosfamide, 1 gm, brand name | Treatment | Chemotherapy |
| HCPCS | J9208 | Injection, ifosfamide, 1 g | Treatment | Chemotherapy |
| HCPCS | S0088 | Imatinib, 100 mg | Treatment | Chemotherapy |
| HCPCS | C9284 | Injection, ipilimumab, 1 mg | Treatment | Chemotherapy |
| HCPCS | J9228 | Injection, ipilimumab, 1 mg | Treatment | Chemotherapy |
| HCPCS | C9474 | Injection, irinotecan liposome, 1 mg | Treatment | Chemotherapy |
| HCPCS | J9205 | Injection, irinotecan liposome, 1 mg | Treatment | Chemotherapy |
| HCPCS | J9206 | Injection, irinotecan, 20 mg | Treatment | Chemotherapy |
| HCPCS | C9240 | Injection, ixabepilone, 1 mg | Treatment | Chemotherapy |
| HCPCS | J9207 | Injection, ixabepilone, 1 mg | Treatment | Chemotherapy |
| HCPCS | C9017 | Lomustine, 10 mg | Treatment | Chemotherapy |
| HCPCS | S0178 | Lomustine, oral, 10 mg | Treatment | Chemotherapy |
| HCPCS | J9230 | Injection, mechlorethamine HCl, (nitrogen mustard), 10 mg | Treatment | Chemotherapy |
| HCPCS | J8600 | Melphalan, oral, 2 mg | Treatment | Chemotherapy |
| HCPCS | J9245 | Injection, melphalan HCl, 50 mg | Treatment | Chemotherapy |
| HCPCS | S0108 | Mercaptopurine, oral, 50 mg | Treatment | Chemotherapy |
| HCPCS | J8610 | Methotrexate, oral, 2.5 mg | Treatment | Chemotherapy |
| HCPCS | J9250 | Methotrexate sodium, 5 mg | Treatment | Chemotherapy |
| HCPCS | J9260 | Methotrexate sodium, 50 mg | Treatment | Chemotherapy |
| HCPCS | C9432 | Mitomycin, 5 mg, brand name | Treatment | Chemotherapy |
| HCPCS | J9280 | Injection, mitomycin, 5 mg | Treatment | Chemotherapy |
| HCPCS | J9290 | Mitomycin, 20 mg | Treatment | Chemotherapy |
| HCPCS | J9291 | Mitomycin, 40 mg | Treatment | Chemotherapy |
| HCPCS | J9293 | Injection, mitoxantrone HCl, per 5 mg | Treatment | Chemotherapy |
| HCPCS | J9261 | Injection, nelarabine, 50 mg | Treatment | Chemotherapy |
| HCPCS | C9021 | Injection, obinutuzumab, 10 mg | Treatment | Chemotherapy |
| HCPCS | J9301 | Injection, obinutuzumab, 10 mg | Treatment | Chemotherapy |
| HCPCS | C9260 | Injection, ofatumumab, 10 mg | Treatment | Chemotherapy |
| HCPCS | J9302 | Injection, ofatumumab, 10 mg | Treatment | Chemotherapy |
| HCPCS | C9297 | Injection, omacetaxine mepesuccinate, 0.01 mg | Treatment | Chemotherapy |
| HCPCS | J9262 | Injection, omacetaxine mepesuccinate, 0.01 mg | Treatment | Chemotherapy |
| HCPCS | C9205 | Injection, oxaliplatin, per 5 mg | Treatment | Chemotherapy |
| HCPCS | J9263 | Injection, oxaliplatin, 0.5 mg | Treatment | Chemotherapy |
| HCPCS | C9127 | Injection, paclitaxel protein-bound particles, per 1 mg | Treatment | Chemotherapy |
| HCPCS | C9431 | Paclitaxel, 30 mg, brand name | Treatment | Chemotherapy |
| HCPCS | J9264 | Injection, paclitaxel protein-bound particles, 1 mg | Treatment | Chemotherapy |
| HCPCS | J9265 | Injection, paclitaxel, 30 mg | Treatment | Chemotherapy |
| HCPCS | J9267 | Injection, paclitaxel, 1 mg | Treatment | Chemotherapy |
| HCPCS | C9235 | Injection, panitumumab, 10 mg | Treatment | Chemotherapy |
| HCPCS | J9303 | Injection, panitumumab, 10 mg | Treatment | Chemotherapy |
| HCPCS | J9266 | Injection, pegaspargase, per single dose vial | Treatment | Chemotherapy |
| HCPCS | C9213 | Injection, pemetrexed, per 10 mg | Treatment | Chemotherapy |
| HCPCS | J9305 | Injection, pemetrexed, 10 mg | Treatment | Chemotherapy |
| HCPCS | J9268 | Injection, pentostatin, 10 mg | Treatment | Chemotherapy |
| HCPCS | C9292 | Injection, pertuzumab, 10 mg | Treatment | Chemotherapy |
| HCPCS | J9306 | Injection, pertuzumab, 1 mg | Treatment | Chemotherapy |
| HCPCS | J9270 | Injection, plicamycin, 2.5 mg | Treatment | Chemotherapy |
| HCPCS | C9259 | Injection, pralatrexate, 1 mg | Treatment | Chemotherapy |
| HCPCS | J9307 | Injection, pralatrexate, 1 mg | Treatment | Chemotherapy |
| HCPCS | S0182 | Procarbazine HCl, oral, 50 mg | Treatment | Chemotherapy |
| HCPCS | J9310 | Injection, rituximab, 100 mg | Treatment | Chemotherapy |
| HCPCS | C9265 | Injection, romidepsin, 1 mg | Treatment | Chemotherapy |
| HCPCS | J9315 | Injection, romidepsin, 1 mg | Treatment | Chemotherapy |
| HCPCS | C9273 | Sipuleucel-T, minimum of 50 million autologous CD54+ cells activated with PAP-GM-CSF, including leukapheresis and all other preparatory procedures, per infusion | Treatment | Chemotherapy |
| HCPCS | Q2043 | Sipuleucel-T, minimum of 50 million autologous cd54+ cells activated with PAP-GM-CSF, including leukapheresis and all other preparatory procedures, per infusion | Treatment | Chemotherapy |
| HCPCS | J9320 | Injection, streptozocin, 1 g | Treatment | Chemotherapy |
| HCPCS | C1086 | Temozolomide, 5 mg, temodar | Treatment | Chemotherapy |
| HCPCS | C9253 | Injection, temozolomide, 1mg | Treatment | Chemotherapy |
| HCPCS | J8700 | Temozolomide, oral, 5 mg | Treatment | Chemotherapy |
| HCPCS | J9328 | Injection, temozolomide, 1 mg | Treatment | Chemotherapy |
| HCPCS | C9239 | Injection, temsirolimus, 1 mg | Treatment | Chemotherapy |
| HCPCS | J9330 | Injection, temsirolimus, 1 mg | Treatment | Chemotherapy |
| HCPCS | C9433 | Thiotepa, 15 mg, brand name | Treatment | Chemotherapy |
| HCPCS | J9340 | Injection, thiotepa, 15 mg | Treatment | Chemotherapy |
| HCPCS | J8705 | Topotecan, oral, 0.25 mg | Treatment | Chemotherapy |
| HCPCS | J9350 | Injection, topotecan, 4 mg | Treatment | Chemotherapy |
| HCPCS | J9351 | Injection, topotecan, 0.1 mg | Treatment | Chemotherapy |
| HCPCS | A9545 | Iodine I-131 tositumomab, therapeutic, per treatment dose | Treatment | Chemotherapy |
| HCPCS | G3001 | Administration and supply of tositumomab, 450 mg | Treatment | Chemotherapy |
| HCPCS | G9835 | Trastuzumab administered within 12 months of diagnosis | Treatment | Chemotherapy |
| HCPCS | G9837 | Trastuzumab not administered within 12 months of diagnosis | Treatment | Chemotherapy |
| HCPCS | J9355 | Injection, trastuzumab, 10 mg | Treatment | Chemotherapy |
| HCPCS | J9360 | Injection, vinblastine sulfate, 1 mg | Treatment | Chemotherapy |
| HCPCS | J9370 | Vincristine sulfate, 1 mg | Treatment | Chemotherapy |
| HCPCS | J9371 | Injection, vincristine sulfate liposome, 1 mg | Treatment | Chemotherapy |
| HCPCS | J9375 | Vincristine sulfate, 2 mg | Treatment | Chemotherapy |
| HCPCS | J9380 | Vincristine sulfate, 5 mg | Treatment | Chemotherapy |
| HCPCS | C9440 | Vinorelbine tartrate, brand name, per 10 mg | Treatment | Chemotherapy |
| HCPCS | J9390 | Injection, vinorelbine tartrate, 10 mg | Treatment | Chemotherapy |
| HCPCS | J9395 | fulvestrant 25mg | Treatment | Chemotherapy |
| PRC10 | 3E03005 | Introduction of Other Antineoplastic into Peripheral Vein, Open Approach | Treatment | Chemotherapy |
| PRC10 | 3E03305 | Introduction of Other Antineoplastic into Peripheral Vein, Percutaneous Approach | Treatment | Chemotherapy |
| PRC10 | 3E04005 | Introduction of Other Antineoplastic into Central Vein, Open Approach | Treatment | Chemotherapy |
| PRC10 | 3E04305 | Introduction of Other Antineoplastic into Central Vein, Percutaneous Approach | Treatment | Chemotherapy |
| PRC10 | 3E05005 | Introduction of Other Antineoplastic into Peripheral Artery, Open Approach | Treatment | Chemotherapy |
| PRC10 | 3E05305 | Introduction of Other Antineoplastic into Peripheral Artery, Percutaneous Approach | Treatment | Chemotherapy |
| PRC10 | 3E06005 | Introduction of Other Antineoplastic into Central Artery, Open Approach | Treatment | Chemotherapy |
| PRC10 | 3E06305 | Introduction of Other Antineoplastic into Central Artery, Percutaneous Approach | Treatment | Chemotherapy |
| HCPCS | J8999 | Prescription drug, oral, chemotherapeutic, NOS | Treatment | Chemotherapy |
| HCPCS | J9175 | Injection, Elliotts' B solution, 1 ml | Treatment | Chemotherapy |
| HCPCS | J9999 | Not otherwise classified, antineoplastic drugs | Treatment | Chemotherapy |
| HCPCS | J0207 | Amifostine | Treatment | Chemotherapy |
| HCPCS | J1190 | Dexrazoxane | Treatment | Chemotherapy |
| HCPCS | J9209 | Mesna | Treatment | Chemotherapy |
| DGN10 | [T80810](https://www.icd10data.com/ICD10CM/Codes/S00-T88/T80-T88/T80-/T80.810) | Extravasation of vesicant antineoplastic chemotherapy | Treatment | Chemotherapy |
| DGN10 | [T80810A](https://www.icd10data.com/ICD10CM/Codes/S00-T88/T80-T88/T80-/T80.810A) | …… initial encounter | Treatment | Chemotherapy |
| DGN10 | [T80810D](https://www.icd10data.com/ICD10CM/Codes/S00-T88/T80-T88/T80-/T80.810D) | …… subsequent encounter | Treatment | Chemotherapy |
| DGN10 | [T80810S](https://www.icd10data.com/ICD10CM/Codes/S00-T88/T80-T88/T80-/T80.810S) | …… sequela | Treatment | Chemotherapy |
| HCPCS | 96401 | Chemotherapy administration, subcutaneous or intramuscular | Treatment | Chemotherapy |
| HCPCS | 96402 | Chemotherapy administration, subcutaneous or intramuscular | Treatment | Chemotherapy |
| HCPCS | 96413 | Chemotherapy administration, IV infusion; up to one hour | Treatment | Chemotherapy |
| HCPCS | 96415 | Chemotherapy administration, IV infusion; each additional hour | Treatment | Chemotherapy |
| HCPCS | 96416 | Chemotherapy administration, IV infusion; initiation of prolonged | Treatment | Chemotherapy |
| HCPCS | 96417 | Chemotherapy administration, IV infusion; each additional | Treatment | Chemotherapy |
| HCPCS | G0498 | Chemotherapy administration, intravenous infusion technique; | Treatment | Chemotherapy |
| HCPCS | 96420 | Chemotherapy administration intra-arterial | Treatment | Chemotherapy |
| HCPCS | 96422 | Chemotherapy administration intra-arterial | Treatment | Chemotherapy |
| HCPCS | 96423 | Chemotherapy administration intra-arterial | Treatment | Chemotherapy |
| HCPCS | 96425 | Chemotherapy administration intra-arterial | Treatment | Chemotherapy |
| HCPCS | 96440 | Chemotherapy administration intrathecal/Peritoneal | Treatment | Chemotherapy |
| HCPCS | 96446 | Chemotherapy administration intrathecal/Peritoneal | Treatment | Chemotherapy |
| HCPCS | 96450 | Chemotherapy administration intrathecal/Peritoneal | Treatment | Chemotherapy |
| HCPCS | 96542 | Chemo administration into the brain | Treatment | Chemotherapy |
| HCPCS | 96549 | Chemo administration into the brain-unlisted chemo procedure | Treatment | Chemotherapy |
| PRC10 | XW03351 | Introduction of Blinatumomab Antineoplastic Immunotherapy into Peripheral Vein, Percutaneous Approach, New Technology Group 1 | Treatment | Chemotherapy |
| PRC10 | XW04351 | Introduction of Blinatumomab Antineoplastic Immunotherapy into Central Vein, Percutaneous Approach, New Technology Group 1 | Treatment | Chemotherapy |
| DGN10 | Z5111 | Encounter for antineoplastic chemotherapy | Treatment | Chemotherapy |
| DGN10 | Z5112 | Encounter for antineoplastic immunotherapy | Treatment | Chemotherapy |
| PRC10 | 3E0R305 | Introduction of Other Antineoplastic into Spinal Canal, Percutaneous Approach | Treatment | Chemotherapy |
| PRC10 | 3E0S305 | Introduction of Other Antineoplastic into Epidural Space, Percutaneous Approach | Treatment | Chemotherapy |
| PRC10 | XW033B3 | Introduction of Cytarabine and Daunorubicin Liposome Antineoplastic into Peripheral Vein, Percutaneous Approach, New Technology Group 3 | Treatment | Chemotherapy |
| PRC10 | XW033C3 | Introduction of Engineered Autologous Chimeric Antigen Receptor T-cell Immunotherapy into Peripheral Vein, Percutaneous Approach, New Technology Group 3 | Treatment | Chemotherapy |
| PRC10 | XW043B3 | Introduction of Cytarabine and Daunorubicin Liposome Antineoplastic into Central Vein, Percutaneous Approach, New Technology Group 3 | Treatment | Chemotherapy |
| PRC10 | XW043C3 | Introduction of Engineered Autologous Chimeric Antigen Receptor T-cell Immunotherapy into Central Vein, Percutaneous Approach, New Technology Group 3 | Treatment | Chemotherapy |
| PRC10 | 3E0A305 | Introduction of Other Antineoplastic into Bone Marrow, Percutaneous Approach | Treatment | Chemotherapy |
| PRC10 | 3E0Q005 | Introduction of Other Antineoplastic into Cranial Cavity and Brain, Open Approach | Treatment | Chemotherapy |
| PRC10 | 3E0Q305 | Introduction of Other Antineoplastic into Cranial Cavity and Brain, Percutaneous Approach | Treatment | Chemotherapy |
| PRC10 | 3E0Q705 | Introduction of Other Antineoplastic into Cranial Cavity and Brain, Via Natural or Artificial Opening | Treatment | Chemotherapy |
| PRC10 | 3E0300P | Introduction of Clofarabine into Peripheral Vein, Open Approach | Treatment | Chemotherapy |
| PRC10 | 3E0400P | Introduction of Clofarabine into Central Vein, Open Approach | Treatment | Chemotherapy |
| PRC10 | 3E0500P | Introduction of Clofarabine into Peripheral Artery, Open Approach | Treatment | Chemotherapy |
| PRC10 | 3E0600P | Introduction of Clofarabine into Central Artery, Open Approach | Treatment | Chemotherapy |
| HCPCS | C9230 | Injection, abatacept, per 10 mg | Treatment | Other Treatment |
| HCPCS | J0129 | Injection, abatacept, 10 mg (code may be used for Medicare when drug administered under the direct supervision of a physician, not for use when drug is self-administered) | Treatment | Other Treatment |
| HCPCS | J0135 | Injection, adalimumab, 20 mg | Treatment | Other Treatment |
| HCPCS | C9211 | Injection, alefacept, for intravenous use, per 7.5 mg | Treatment | Other Treatment |
| HCPCS | C9212 | Injection, alefacept, for intramuscular use, per 7.5 mg | Treatment | Other Treatment |
| HCPCS | J0215 | Injection, alefacept, 0.5 mg | Treatment | Other Treatment |
| HCPCS | S0193 | Injection alefacept, 7.5 mg (includes dose packaging) | Treatment | Other Treatment |
| HCPCS | J2910 | Injection, aurothioglucose, up to 50 mg | Treatment | Other Treatment |
| HCPCS | C9436 | Azathioprine, parenteral, brand name, per 100 mg | Treatment | Other Treatment |
| HCPCS | J7500 | Azathioprine, oral, 50 mg | Treatment | Other Treatment |
| HCPCS | J7501 | Azathioprine, parenteral, 100 mg | Treatment | Other Treatment |
| HCPCS | K0119 | Azathioprine, oral, tab, 50 mg | Treatment | Other Treatment |
| HCPCS | K0120 | Azathioprine, parenteral, 100 mg | Treatment | Other Treatment |
| HCPCS | J0480 | Injection, basiliximab, 20 mg | Treatment | Other Treatment |
| HCPCS | Q2019 | Injection, basiliximab, 20 mg | Treatment | Other Treatment |
| HCPCS | C9286 | Injection, belatacept, 1 mg | Treatment | Other Treatment |
| HCPCS | J0485 | Injection, belatacept, 1 mg | Treatment | Other Treatment |
| HCPCS | J0490 | Injection, belimumab, 10 mg | Treatment | Other Treatment |
| HCPCS | Q2044 | Injection, belimumab, 10 mg | Treatment | Other Treatment |
| HCPCS | J0638 | Injection, canakinumab, 1 mg | Treatment | Other Treatment |
| HCPCS | C9249 | Injection, certolizumab pegol, 1 mg | Treatment | Other Treatment |
| HCPCS | J0717 | Injection, certolizumab pegol, 1 mg (code may be used for Medicare when drug administered under the direct supervision of a physician, not for use when drug is self administered) | Treatment | Other Treatment |
| HCPCS | J0718 | Injection, certolizumab pegol, 1 mg | Treatment | Other Treatment |
| HCPCS | C9438 | Cyclosporine, oral, 100 mg, brand name | Treatment | Other Treatment |
| HCPCS | J7502 | Cyclosporine, oral, 100 mg | Treatment | Other Treatment |
| HCPCS | J7515 | Cyclosporine, oral, 25 mg | Treatment | Other Treatment |
| HCPCS | J7516 | Cyclosporine, parenteral, 250 mg | Treatment | Other Treatment |
| HCPCS | K0122 | Cyclosporine, parenteral, 250 mg | Treatment | Other Treatment |
| HCPCS | J7513 | Daclizumab, parenteral, 25 mg | Treatment | Other Treatment |
| HCPCS | C9236 | Injection, eculizumab, 10 mg | Treatment | Other Treatment |
| HCPCS | J1300 | Injection, eculizumab, 10 mg | Treatment | Other Treatment |
| HCPCS | S0162 | Injection, efalizumab, 125 mg | Treatment | Other Treatment |
| HCPCS | J1438 | Injection, etanercept, 25 mg (code may be used for Medicare when drug administered under the direct supervision of a physician, not for use when drug is self-administered) | Treatment | Other Treatment |
| HCPCS | J1595 | Injection, glatiramer acetate, 20 mg | Treatment | Other Treatment |
| HCPCS | Q2010 | Injection, glatiramer acetate, per dose | Treatment | Other Treatment |
| HCPCS | J1600 | Injection, gold sodium thiomalate, up to 50 mg | Treatment | Other Treatment |
| HCPCS | J1602 | Injection, golimumab, 1 mg, for intravenous use | Treatment | Other Treatment |
| HCPCS | S0176 | Hydroxyurea, oral, 500 mg | Treatment | Other Treatment |
| HCPCS | J1745 | Injection, infliximab, excludes biosimilar, 10 mg | Treatment | Other Treatment |
| HCPCS | Q5102 | Injection, infliximab, biosimilar, 10 mg | Treatment | Other Treatment |
| HCPCS | J9213 | Injection, interferon, alfa-2a, recombinant, 3 million units | Treatment | Other Treatment |
| HCPCS | S0145 | Injection, pegylated interferon alfa-2a, 180 mcg per ml | Treatment | Other Treatment |
| HCPCS | J9214 | Injection, interferon, alfa-2b, recombinant, 1 million units | Treatment | Other Treatment |
| HCPCS | S0146 | Injection, pegylated interferon alfa-2b, 10 mcg per 0.5 ml | Treatment | Other Treatment |
| HCPCS | S0148 | Injection, pegylated interferon alfa-2B, 10 mcg | Treatment | Other Treatment |
| HCPCS | J9212 | Injection, interferon alfacon-1, recombinant, 1 mcg | Treatment | Other Treatment |
| HCPCS | J9215 | Injection, interferon, alfa-N3, (human leukocyte derived), 250,000 IU | Treatment | Other Treatment |
| HCPCS | J1825 | Injection, interferon beta-1a, 33 mcg | Treatment | Other Treatment |
| HCPCS | J1826 | Injection, interferon beta-1a, 30 mcg | Treatment | Other Treatment |
| HCPCS | Q3025 | Injection, interferon beta-1a, 11 mcg for intramuscular use | Treatment | Other Treatment |
| HCPCS | Q3026 | Injection, interferon beta-1a, 11 mcg for subcutaneous use | Treatment | Other Treatment |
| HCPCS | Q3027 | Injection, interferon beta-1a, 1 mcg for intramuscular use | Treatment | Other Treatment |
| HCPCS | Q3028 | Injection, interferon beta-1a, 1 mcg for subcutaneous use | Treatment | Other Treatment |
| HCPCS | J1830 | Injection interferon beta-1b, 0.25 mg (code may be used for Medicare when drug administered under the direct supervision of a physician, not for use when drug is self-administered) | Treatment | Other Treatment |
| HCPCS | J9216 | Injection, interferon, gamma 1-b, 3 million units | Treatment | Other Treatment |
| HCPCS | J7505 | Muromonab-CD3, parenteral, 5 mg | Treatment | Other Treatment |
| HCPCS | J7517 | Mycophenolate mofetil, oral, 250 mg | Treatment | Other Treatment |
| HCPCS | K0412 | Mycophenolate mofetil, oral, 250 mg (CellCept) | Treatment | Other Treatment |
| HCPCS | C9126 | Injection natalizumab per 5 mg | Treatment | Other Treatment |
| HCPCS | J2323 | Injection, natalizumab, 1 mg | Treatment | Other Treatment |
| HCPCS | Q4079 | Injection, natalizumab, 1 mg | Treatment | Other Treatment |
| HCPCS | J2504 | Injection, pegademase bovine, 25 IU | Treatment | Other Treatment |
| HCPCS | Q2012 | Injection, pegademase bovine, 25 IU | Treatment | Other Treatment |
| HCPCS | 4153F | Combination peginterferon and ribavirin therapy prescribed (HEP-C) | Treatment | Other Treatment |
| HCPCS | J2793 | Injection, rilonacept, 1 mg | Treatment | Other Treatment |
| HCPCS | C9020 | Sirolimus tablet, 1 mg | Treatment | Other Treatment |
| HCPCS | C9106 | Sirolimus, per 1 mg/ml | Treatment | Other Treatment |
| HCPCS | J7520 | Sirolimus, oral, 1 mg | Treatment | Other Treatment |
| HCPCS | C9006 | Injection, tacrolimus, per 5 mg (1 amp) | Treatment | Other Treatment |
| HCPCS | J7503 | Tacrolimus, extended release, oral, 0.25 mg | Treatment | Other Treatment |
| HCPCS | J7507 | Tacrolimus, immediate release, oral, 1 mg | Treatment | Other Treatment |
| HCPCS | J7508 | Tacrolimus, extended release, (Astagraf XL), oral, 0.1 mg | Treatment | Other Treatment |
| HCPCS | J7525 | Tacrolimus, parenteral, 5 mg | Treatment | Other Treatment |
| HCPCS | C9264 | Injection, tocilizumab, 1 mg | Treatment | Other Treatment |
| HCPCS | J3262 | Injection, tocilizumab, 1 mg | Treatment | Other Treatment |
| HCPCS | C9261 | Injection, ustekinumab, 1 mg | Treatment | Other Treatment |
| HCPCS | C9487 | Ustekinumab, for intravenous injection, 1 mg | Treatment | Other Treatment |
| HCPCS | J3357 | Ustekinumab, for subcutaneous injection, 1 mg | Treatment | Other Treatment |
| HCPCS | J3358 | Ustekinumab, for intravenous injection, 1 mg | Treatment | Other Treatment |
| HCPCS | Q9989 | Ustekinumab, for intravenous injection, 1 mg | Treatment | Other Treatment |
| HCPCS | J7599 | Immunosuppressive drug, not otherwise classified | Treatment | Other Treatment |
| PRC10 | 3E040WL | Introduction of Immunosuppressive into Central Vein, Open | Treatment | Other Treatment |
| PRC10 | 3E030WL | Introduction of Immunosuppressive into Peripheral Vein, Open | Treatment | Other Treatment |
| PRC10 | 3E033WL | Introduction of Immunosuppressive into Peripheral Vein, Percutaneous | Treatment | Other Treatment |
| PRC10 | 3E043WL | Introduction of Immunosuppressive into Central Vein, Percutaneous | Treatment | Other Treatment |
| PRC10 | 3E050WL | Introduction of Immunosuppressive into Peripheral Artery, Open | Treatment | Other Treatment |
| PRC10 | 3E053WL | Introduction of Immunosuppressive into Peripheral Artery, Percutaneous | Treatment | Other Treatment |
| PRC10 | 3E060WL | Introduction of Immunosuppressive into Central Artery, Open | Treatment | Other Treatment |
| PRC10 | 3E063WL | Introduction of Immunosuppressive into Central Artery, Percutaneous | Treatment | Other Treatment |
| PRC10 | 3E0230M | Introduction of Monoclonal Antibody into Muscle, Percutaneous Approach | Treatment | Other Treatment |
| PRC10 | 3E0300M | Introduction of Monoclonal Antibody into Peripheral Vein, Open Approach | Treatment | Other Treatment |
| PRC10 | 3E0330M | Introduction of Monoclonal Antibody into Peripheral Vein, Percutaneous Approach | Treatment | Other Treatment |
| PRC10 | 3E0400M | Introduction of Monoclonal Antibody into Central Vein, Open Approach | Treatment | Other Treatment |
| PRC10 | 3E0430M | Introduction of Monoclonal Antibody into Central Vein, Percutaneous Approach | Treatment | Other Treatment |
| PRC10 | 3E0500M | Introduction of Monoclonal Antibody into Peripheral Artery, Open Approach | Treatment | Other Treatment |
| PRC10 | 3E0530M | Introduction of Monoclonal Antibody into Peripheral Artery, Percutaneous Approach | Treatment | Other Treatment |
| PRC10 | 3E0600M | Introduction of Monoclonal Antibody into Central Artery, Open Approach | Treatment | Other Treatment |
| PRC10 | 3E0630M | Introduction of Monoclonal Antibody into Central Artery, Percutaneous Approach | Treatment | Other Treatment |
| PRC10 | 3E0A30M | Introduction of Monoclonal Antibody into Bone Marrow, Percutaneous Approach | Treatment | Other Treatment |
| PRC10 | 3E0Q00M | Introduction of Monoclonal Antibody into Cranial Cavity and Brain, Open Approach | Treatment | Other Treatment |
| PRC10 | 3E0Q30M | Introduction of Monoclonal Antibody into Cranial Cavity and Brain, Percutaneous Approach | Treatment | Other Treatment |
| PRC10 | 3E0Q70M | Introduction of Monoclonal Antibody into Cranial Cavity and Brain, Via Natural or Artificial Opening | Treatment | Other Treatment |
| PRC10 | 3E0R30M | Introduction of Monoclonal Antibody into Spinal Canal, Percutaneous Approach | Treatment | Other Treatment |
| PRC10 | 3E0S30M | Introduction of Monoclonal Antibody into Epidural Space, Percutaneous Approach | Treatment | Other Treatment |
| PRC10 | 3E03002 | Introduction of High-dose Interleukin-2 into Peripheral Vein, Open Approach | Treatment | Other Treatment |
| PRC10 | 3E03003 | Introduction of Low-dose Interleukin-2 into Peripheral Vein, Open Approach | Treatment | Other Treatment |
| PRC10 | 3E04002 | Introduction of High-dose Interleukin-2 into Central Vein, Open Approach | Treatment | Other Treatment |
| PRC10 | 3E04003 | Introduction of Low-dose Interleukin-2 into Central Vein, Open Approach | Treatment | Other Treatment |
| PRC10 | 3E05002 | Introduction of High-dose Interleukin-2 into Peripheral Artery, Open Approach | Treatment | Other Treatment |
| PRC10 | 3E05003 | Introduction of Low-dose Interleukin-2 into Peripheral Artery, Open Approach | Treatment | Other Treatment |
| PRC10 | 3E06002 | Introduction of High-dose Interleukin-2 into Central Artery, Open Approach | Treatment | Other Treatment |
| PRC10 | 3E06003 | Introduction of Low-dose Interleukin-2 into Central Artery, Open Approach | Treatment | Other Treatment |
| PRC10 | 3E0R302 | Introduction of High-dose Interleukin-2 into Spinal Canal, Percutaneous Approach | Treatment | Other Treatment |
| PRC10 | 3E0R303 | Introduction of Low-dose Interleukin-2 into Spinal Canal, Percutaneous Approach | Treatment | Other Treatment |
| PRC10 | 3E0S302 | Introduction of High-dose Interleukin-2 into Epidural Space, Percutaneous Approach | Treatment | Other Treatment |
| PRC10 | 3E0S303 | Introduction of Low-dose Interleukin-2 into Epidural Space, Percutaneous Approach | Treatment | Other Treatment |
| ^a^ DGN10 indicates diagnosis codes and PRC10 indicates procedure codes sourced from International Classification of Diseases, Tenth Revision, Clinical Modification. HCPCS indicates codes sourced from Healthcare Common Procedure Coding System. REV indicates revenue codes. | | | | |
| ^b^ MDS/MPN/IPD includes Myelodysplastic Syndromes, Myeloproliferative disorder, Immunoproliferative diseases and other related conditions.  ^c^ Patients were determined to be immunocompromised if at least two condition or treatment codes were captured on separate days in the 3 months prior to hospitalization. | | | | |
